# Evaluating whole genome sequencing for rare diseases in newborn screening: evidence synthesis from a series of systematic reviews

**DOI:** 10.1101/2024.09.03.24312979

**Authors:** Karoline Freeman, Jacqueline Dinnes, Bethany Shinkins, Corinna Clark, Inès Kander, Katie Scandrett, Shivashri Chockalingam, Aziza Osman, Naila Dracup, Rachel Court, Furqan Butt, Cristina Visintin, James R Bonham, David Elliman, Graham Shortland, Anne Mackie, Zosia Miedzybrodzka, Sian Morgan, Felicity Boardman, Yemisi Takwoingi, Sian Taylor-Phillips

## Abstract

**Background:** Assessment of newborn screening using whole genome sequencing (WGS) presents considerable challenges for policy advisors, not least given the logistics of simultaneously evaluating the evidence for 200 rare genetic conditions. The ‘genotype first’ approach has the potential for harms, and benefits are uncertain.

**Objective:** To assess different approaches to evaluating WGS for newborn screening to inform the development of a robust method for informing policy decisions.

**Methods:** We undertook ‘traditional’ reviews of five conditions using standard systematic review methods (considering gene penetrance, expressivity, and prevalence, the accuracy and effectiveness of WGS, and effect of earlier treatment) (search inception to November 2023), evaluated the NIH Clinical Genome Resource (ClinGen) for evidence on the five conditions, reviewed genomic studies of paediatric screening cohorts reporting penetrance for pathogenic variants (search inception to February 2024) and undertook a methodological review of economic evaluations of WGS/ whole exome sequencing (WES) (search inception to January 2024). We explored public views on evaluating WGS.

**Data sources:** MEDLINE (Ovid), Embase (Ovid), Web of Science, Science Citation Index (via Clarivate), the Cochrane Library (via Wiley), CEA registry and Econlit.

Actionability reports and scores were downloaded from the ClinGen website on 30^th^ April 2024.

**Results:** The traditional review approach identified 221 studies that either reported on the genetic spectrum of individuals with the five conditions or provided limited evidence about the benefits of earlier treatment. No evidence about penetrance and expressivity or the accuracy or effectiveness of WGS in newborns was identified. ClinGen reviews were available for four of the five conditions. The ClinGen ‘actionability’ ratings for all four conditions disagreed with the findings of our traditional reviews. Our review of 14 genomic studies of newborn screening cohorts found insufficient information to allow individual highly penetrant pathogenic variants for any condition to be identified for consideration in a screening programme. None of the 86 economic evaluations of WGS or WES were set in a screening context. Some micro-costing studies are available that could help understand the resource use and costs associated with WGS. Following a series of PPI meetings, attendees appreciated the uncertainties of WGS and suggested that a wider stakeholder perspective was needed to inform policy decisions.

**Limitations:** Although we only examined five conditions in depth, the consistency in lack of data suggests our conclusions are robust.

**Conclusions:** The traditional systematic review approach for evaluating WGS of newborns identified a paucity of high-quality evidence. Extending the review to all 200 conditions is not feasible and is unlikely to yield the level of evidence required by policy advisors. The use of existing genome resources and review of genomic studies of newborn screening cohorts were not found to be viable alternatives. The cost-effectiveness of WGS in a newborn screening context is unknown.

**Future work:** Large-scale collaborative research is required to evaluate the short- and long-term harms, benefits and economic implications of WGS for screening newborns. We propose a staged approach to evaluation considering only conditions with pathogenic variants with very high penetrance to minimise harm from overdiagnosis.

**Study registration:** The protocol for this study is registered on PROSPERO: CRD42023475529

**Funding details:** This study/project is funded by the NIHR Evidence Synthesis Programme (ESG_HTA_NIHR159928). The views expressed are those of the author(s) and not necessarily those of the NIHR or the Department of Health and Social Care.

## Plain language summary

The government has funded a project that uses whole genome sequencing (WGS) to look for over 200 rare conditions in newborns’ DNA, before babies appear to be ill. This could be a new screening programme. We need to find a way to assess whether it improves lives and is good value for the NHS.

WGS detects variations in our DNA. We all have many variations. They make us unique and only a few cause harm. It is difficult to predict true disease from a genetic finding. A baby may have a harmful variant but that does not necessarily mean they will have the associated disease. Using WGS to detect harmful variants could provide health benefits. WGS may also cause uncertainty, anxiety and perhaps harm, by giving babies and children unnecessary treatment.

We explored three approaches:

1. a traditional approach to assess WGS for five rare conditions
2. an approach of measuring uncertainty when predicting disease from genetic findings
3. a free online resource with information on rare genetic diseases

We also looked for studies that have weighed up the cost and health implications of implementing WGS.

We met five times with a group of parents, expectant parents and charity representatives to explore challenges of WGS. Challenges include communication, consent, data security, privacy and uncertainty.

We found, that:

- there is insufficient evidence for the five conditions to inform policy
- available studies using WGS only report the number of variants detected, not how well these predict disease
- the free online resource does not have good quality evidence so is not useful for policy makers

We need to find new ways of collecting important information about how WGS could help babies and their families without causing too much harm before we can assess its value.

## Scientific summary

### Background

In 2021, Genomics England Limited (GEL) launched its Generation Study of whole genome sequencing (WGS) to screen for over 200 rare diseases in 100,000 newborns to explore its potential for an expanded UK newborn screening programme. This presents a number of new challenges for policy advisors.

Multiplex testing has been available for years. Tandem mass spectrometry, currently used in the newborn bloodspot (NBS) screening programme, can detect dozens of statistical abnormalities in the blood spot. Each condition included on the NBS programme, and any potential candidate conditions, have been assessed in turn. However, there is pressure to assess all the conditions that might be found with WGS at once.

The genotype first approach has the potential for harms and in some cases, may be more uncertain than more traditional methods. Not everyone with a pathogenic variant will develop symptomatic disease (incomplete penetrance) and symptoms caused by the same genetic variant can vary in severity among affected people (expressivity).

A cost-effectiveness analysis of WGS in newborn screening will be needed for a policy decision, but screening for potentially hundreds of conditions with a single test will require a different methodological approach than one that focuses on a single condition.

Finally, the use of WGS for newborn screening presents several ethical challenges. The majority are common to all screening programmes (anxiety, informed choice, penetrance) but there are some that are more pressing or likely in this programme. For example, some of the genetic variants might only be of significance later in life, there are implications for the relatives if a variant is found and there is considerable commercial interest in secondary uses of the data which will not benefit participants directly.

We, therefore, aimed to 1) assess different evidence sources and approaches to evidence synthesis, 2) review methods for evaluating cost-effectiveness, and 3) collate views of the public on the main challenges of WGS to inform an approach to assessing WGS for newborn screening in the future.

### Objectives

1. To undertake a series of five systematic reviews covering a stratified (by burden and cost of the intervention) random sample of rare diseases to establish the evidence base per condition and to provide a reference case for comparison with alternative review approaches. The reviews addressed six questions mapped to the UK NSC criteria on penetrance and expressivity, the proportion of children with disease who carry gene variants, test accuracy, effectiveness of earlier treatment, effectiveness, and benefits and harms of WGS.
2. To explore the utility of the existing online resource Clinical Genome Resource (ClinGen) as a shortcut into the evidence on actionability of rare paediatric genetic diseases in order to evaluate it as a potential evidence source for the UK NSC.
3. To undertake a review of genomic studies of paediatric screening cohorts reporting penetrance of pathogenic variants to explore the feasibility of identifying highly penetrant pathogenic variants that could be considered for a screening programme.
4. To produce a methodological overview of existing published economic evaluations and costing studies of WGS or whole exome sequencing (WES).
5. To explore patient and public views about the introduction of WGS for newborn screening.

## Methods

### Review of five conditions

A stratified random sample of five conditions was reviewed. Stratification was based on a range of scenarios that might reasonably have an impact on the UK NSC’s recommendations relating to WGS for newborn screening. The five conditions were:

a. Pyridoxine dependent epilepsy (PDE)
b. Heritable retinoblastoma (hRB)
c. X-linked hypophosphataemic rickets (XLHR)
d. Familial hemophagocytic lymphotistiocytosis (fHLH)
e. Medium Chain Acyl-CoA Dehydrogenase Deficiency (MCADD)

Data sources: MEDLINE (via Ovid), Embase (via Ovid), Science Citation Index (via Clarivate) and the Cochrane Library (via Wiley) from inception to November 2023.

Study eligibility criteria were defined for each review question and included:

#### Population

Studies of newborn screening cohorts or

Studies of newborns and children (≤18 years) with clinical, or biochemical and clinical features of the five conditions

#### Exposure / Intervention

Presence of pathogenic variants in the relevant gene(s) detected by sequencing

Eligible interventions relevant to the screening context with ‘early’ intervention defined separately for each condition

#### Target condition

Clinically or clinically and biochemically defined disease

#### Outcomes

Measures of disease-specific morbidity and mortality

Any health-related health outcomes that could be measured across conditions

Any harms or other benefits from WGS

We produced a narrative synthesis of studies.

### Exploring ClinGen as an evidence source

We searched the ClinGen database on 19th February 2024 for each of the genes included in the review of five conditions and tabulated actionability scores and evidence levels comparatively against the evaluation from GEL and against our assessment using the UK NSC criteria.

### Review of genomic studies of paediatric screening cohorts reporting penetrance of pathogenic variants

Data Source: MEDLINE (via Ovid), Embase (via Ovid), Science Citation Index (via Clarivate) and the Cochrane Library (via Wiley) from inception to January 2024

Study eligibility criteria: Studies of unselected newborns sequenced for any rare condition with outcomes of penetrance or an approximation

We produced a narrative synthesis of our findings.

### Methods for review of cost-effectiveness evaluations of WGS and WES

Data source: MEDLINE (Ovid), Embase (Ovid), CEA (Cost-Effectiveness Analysis) registry, Web of Science and Econlit from inception to February 2024 and hand searches of identified systematic reviews

Study eligibility criteria: Economic evaluations, clinical trials and health technology assessments reporting costs of WGS or WES in human healthcare

Evidence synthesis: a general narrative synthesis of the methodological approaches adopted will be reported. We will also focus on two specific methodological questions:

1. How were the costs associated with WGS and WES estimated?
2. What comparators were included in each study?

#### Consideration of the public voice in the evaluation of WGS

Eight members of the public attended five 2-hour virtual meetings between 15th January and 21st May 2024. Meetings were deliberative and explored pre-defined topics related to WGS: harms and benefits, genetic uncertainties, systematic review findings, and the role of PPIE in future reviews.

Themes of participants’ views were narratively synthesised.

## Results

### Review of five conditions

#### Extrapolating the traditional approach to 200 conditions

We screened 19,689 titles and abstracts for the five traditional reviews, of which 1,348 were selected for full text assessment (range 55 to 449 per condition). A total of 221 studies were eligible for inclusion across the five reviews (range 31 to 78). No evidence was identified for the four review questions that required studies to be conducted in newborns. Overall, the five traditional reviews yielded very little of the evidence required by the UK NSC. Considering the time taken to identify and select the evidence, and extrapolating to a review of 200 conditions, we could expect as many as 787,560 unique records, 53,920 full texts to be screened, and 8,840 studies to be reviewed and synthesised which is estimated to take a team of five reviewers 23 years.

#### Evidence on the genetic spectrum in children with disease

260 studies (range 26 to 73) were included that reported the genetic spectrum in children with the five conditions. The proportion of children testing positive on sequencing varied for each condition by

1. definition of disease from broadest (symptomatically defined) to narrowest (genetically defined) category,
2. the testing strategy (type of test, number of genes and extent of gene sequencing, additional genetic testing to supplement sequencing) and
3. the extent of ‘pre-screening’ using biochemical and clinical markers.

At variant level, studies provided data on the proportion of novel variants and type of variants but very little information on severity of disease for specific variants. The large number of novel variants present a challenge to sequencing newborns as their pathogenicity is difficult to ascertain.

#### Evidence on early versus late treatment

22 studies (range 1 to 9) reported outcomes of early versus late treatment. No study was designed to compare treatment effectiveness in screen detected versus symptomatically detected children. Definitions of early and late treatment varied and relied on study authors’ definitions. The evidence-base pointed towards some benefit in early treatment. However, the quality and volume of the evidence was low because of the definition of early vs late, the type of study, the number of participants, and the number of studies available. Therefore, there was insufficient evidence to clearly judge the effect.

### Learnings from the five traditional reviews

A single approach to reviewing five conditions was not feasible due to differences in the conditions’ characteristics, treatment and aim of screening. For instance, each search was developed individually. Disease specific categories were needed to organise studies by population subtype because of differences in the availability of biochemical tests, the number of disease groups with overlapping symptoms and whether conditions could only be defined genetically. The definition of early versus late depended on whether the relevant intervention was preventative, curative or for symptom management, whether an early intervention phase could be defined and whether conditions were progressive or presented following a trigger. A review of 200 conditions would require 200 individual reviews, however, some learning may be transferable between reviews of similar conditions, which we could not explore with the five conditions.

### Exploring ClinGen as an evidence source for the UK NSC

Four of the five conditions reviewed (PDE, hRB, XLHR and MCADD) had a paediatric actionability report available on ClinGen in March 2024. However, no information on variant classification in terms of pathogenicity was available for any of the genes.

Comparison of our assessment of the five conditions using the UK NSC criteria with the ClinGen scores of actionability alongside GEL’s decisions to include genes on their gene list was complicated. The overall decision on actionability differed for 4/4 conditions between ClinGen and our assessment using the UK NSC criteria and for 5/5 conditions between GEL’s assessment and our assessment.

It would be inappropriate for the UK NSC to base decisions on potential screening programs on the actionability reported in ClinGen without further assessment.

### Review of genomic studies of paediatric screening cohorts reporting penetrance of pathogenic variants

Fourteen studies reported experiences with gene sequencing in newborns of which five provided information that approximated penetrance by reporting some clinical follow-up after a sequence positive test. The number of included genes ranged from 134 to 954 across the five studies and the number of newborns sequenced ranged from 127 to 29,989. Gene Selection and variant interpretation varied across studies.

The proportion of babies designated screen positive from these studies ranged from 1.7% to 9.7%. Half of the positive screens were for conditions not included on conventional newborn screening panels in the study countries (US and China). However, the clinical significance of between 83.3% to 100% of these sequencing only positives was unknown, so we do not know if detecting and reporting these was overdiagnosis of clinically insignificant disease, misdiagnosis of disease or early detection of late onset disease.

Follow-up ranged from 2 months to >5 years. Penetrance was approximated by the number of confirmed cases after clinical follow-up. For all genes considered together, penetrance ranged from 1.6% after follow-up of 24-48 months to 50.4% after a median follow-up of 1.2 years. The studies did not provide sufficient evidence to understand penetrance for any genetic variant because:

- The number of infants with a specific condition displaying a range of variants was too low.
- Infants with confirmed genetic disease received management which precludes estimation of penetrance and expressivity for cases without symptomatic confirmation of disease.
- Clinical follow-up was not long enough to include all childhood onset cases.

Overall, there was little agreement on what genes should be considered in newborn screening, no indication of how to interpret discordant results from NBS programmes and genetic screening, and evidence of overdiagnosis. The studies demonstrated unequivocally that if WGS was to be introduced without further research, it would cause significant problems.

### Review of cost-effectiveness evaluations of WGS and WES

Eighty-six studies were included in the review. None of them focused on the use of WGS or WES in a screening context. Under half (n=39/86, 45%) were full economic evaluations, of which only 10 were cost utility studies i.e. studies which estimated cost per quality-adjusted life year. Most evaluations focused only on costs and outcomes associated with the diagnostic pathway, avoiding the complexity of capturing the impact of a diagnosis on patient management. Two thirds of the included studies reported a costing perspective; of which one third (29/86 (36%) adopted a broad health care system perspective, 15 a specific health system perspective, eight a patient perspective, and five a societal perspective. Only seven studies (8%) adopted a lifetime horizon. Of the studies that included a comparator (78/86, 91%), forty-four (56%) explicitly stated that the comparator was current standard of care testing consisting of a broad range of tests. Different assumptions were made in terms of which tests would no longer be needed following the incorporation of WES or WGS in the diagnostic pathway.

### Consideration of the public voice in the evaluation of WGS

The group largely supported WGS for newborn screening. As meetings progressed and the complexities were explored, however, views became more nuanced, for example, one participant mentioned that they now were “sifting on the fence a bit”.

Participants identified a wide range of benefits and harms, and broadly felt that the benefits outweigh the harms. Key harms they were concerned about ranged from personal (anxiety) to societal (strain on health services). Key benefits included saving lives and avoidance of a diagnostic odyssey.

The process would have benefited from having more time to develop and discuss ideas. For a future review, it would be beneficial to increase the diversity of viewpoints.

#### Conclusions

A traditional systematic review approach to evaluating WGS of newborns is unfeasible and we were unable to identify an acceptable alternative way to evaluate WGS for newborn screening in a single mechanism. Cost-effectiveness evidence for WGS has only focused on symptomatic populations to date. Our review highlights the main evidence gaps and informs the direction of future research efforts.

We propose a series of possible research approaches undertaken in large joint-up collaborations to produce the evidence that is needed for policy advisors before an evaluation of WGS is feasible. This may include a coordinated International approach to collecting penetrance data for pathogenic variants with a clear treatment plan. This could be followed by a staged approach of evaluation considering only those of the 200 conditions for screening that have pathogenic variants with very high penetrance.

Funding: This study/project is funded by the NIHR Evidence Synthesis Programme (ESG_HTA_NIHR159928). The views expressed are those of the author(s) and not necessarily those of the NIHR or the Department of Health and Social Care.

Study registration: The protocol for this study is registered on PROSPERO: CRD42023475529

## Introduction

Rare diseases are a group of disorders that are characterised by their relatively low prevalence in the population and are typically defined in the UK as affecting less than 1 in 2,000 individuals.^1^ There are approximately 7,000 rare disorders, which, combined, affect around 6% of the population in the Western world.^2^ About 80% of rare disorders are thought to have a genetic cause.^2^ If a disorder is caused by a variation in a single gene, the disorder is termed a monogenic disorder. For only half of the estimated 5,000 monogenic disorders, the underlying genes are known.^3^ The Identification of disease-causing variation in the known genes is challenging. Every human individual is believed to have up to 5 million variants compared to the reference genome.^4^ This variation is responsible for human diversity and only a small fraction of variants affects human health.

Variants in genes associated with human health can be benign or pathogenic or be of unknown significance meaning that the link between genetic variation and clinical phenotype is unknown. For pathogenic variants the link between genotype and phenotype is known but can be incomplete such that only a proportion of individuals with the pathogenic variation may develop symptoms (incomplete penetrance). These symptoms can further vary in severity even among affected family members (expressivity). This makes it difficult to predict disease even if the underlying genetic cause is well characterised.

Disorders caused by genetic variants can present at any time, from birth (e.g., phenylketonuria) to much later in life (e.g., Huntington’s disease). It is estimated that there are about 600 childhood onset conditions for which there is a potential intervention^4, 5^ and newborn screening aims to identify the disorders that benefit from pre-symptomatic detection. Early diagnosis enables surveillance and early intervention when available, which can significantly improve outcomes, and is particularly important in conditions with rapid progression or that cause irreversible damage. Current approaches to screening for rare disorders as part of the UK national newborn bloodspot (NBS) screening programme have been successful in both their capacity for detection of infants with rare conditions, and in the wide uptake of the screening programme by the public. However, the number of diseases currently screened for in the UK is limited to nine conditions and screening is based on biochemical markers. In recent years, whole genome sequencing (WGS) as a first-line screening test has emerged as a possible tool for an expansion of the NBS screening programme to identify genetic disease.

Genetic testing aims to identify changes in the genes to help confirm or rule out genetic disorders (diagnosis) or establish the likelihood of a person developing and passing on a genetic disorder in the future (screening). Genetic testing can target single variants or single genes either alone or as part of a gene panel, or can consist of whole exome sequencing (WES, looking at all coding portions of the DNA) and WGS (looking at both coding and non-coding regions of the DNA). WGS can potentially detect every variation in a genome, testing for hundreds of genetic diseases at the same time.

In 2021, Genomics England launched its Generation Study using WGS to sequence 100,000 newborns to screen for over 200 rare diseases to test its potential for an expanded UK NBS programme.^6^ Recruiting of pregnant women began in the first half of 2024. This poses several challenges to the UK National Screening Committee (UK NSC), who advise the four UK governments on screening related questions including the addition of new conditions to the NBS screening programme.

No methods exist for the evaluation of hundreds of conditions identified by one test. multiplex testing has been available for years and tandem mass spectrometry (MS/MS) currently used in the NBS programme can detect dozens of statistical abnormalities in the blood spot. The approach to assessing each of the conditions that could be detected on the bloodspot has been to look at each condition in turn allowing a thorough evaluation of the evidence around testing, early treatment and harms and benefits of screening and an assessment of the evidence against the 20 UK NSC screening criteria.^7^. However, there is pressure to assess all the conditions that might be found with WGS at once but a condition-by-condition approach for 200 conditions, with many thousands of gene variants, may not be feasible or effective considering the limited evidence base for rare genetic diseases.

The ‘genotype first’ approach has the potential for harms in the form of overdiagnosis and overmedicalisation, particularly for conditions in which penetrance is low and for conditions with a high number of variants of unknown significance. The wider psychological and societal impact of genetic testing on such a large scale is also unknown. Most healthy, adult, UK Biobank participants were found to have one or more rare non-synonymous variants when a panel to look at more than 500 disease genes was used.^8^ In healthy adults, these can be assumed to be benign. Similar findings in asymptomatic newborns, would be concerning demonstrating the difficulties in variant interpretation. To assess pathogenicity (e.g. >99% probability of pathogenicity for pathogenic variants and 90%–99% probability of pathogenicity for likely pathogenic variants), variants need do undergo a complex interpretation process considering prevalence of the variant, inheritance pattern, variant type and their predicted effects as well as observed gene-disease associations.^9^ However, not all pathogenic variants cause disease in all individuals who carry the variant. Penetrance is a measurement of the relationship between a genotype and phenotype. For rare diseases, the existing evidence and context for variant interpretation comes mainly from family and clinical studies, which means penetrance is generally overestimated and reported pathogenicity does not equate with penetrance.^10^ Moving to the screening context, knowledge of penetrance of known pathogenic variants in the general population is key for understanding the proportion of newborns who are likely to benefit from detection at screening and those who are likely overdiagnosed.^11^ An evaluation of WGS for newborn screening should be able to select variants with high penetrance for a low-risk screening programme or identify a variant annotation approach that is effective in filtering out harmless variants. We do not know how informative a review focused on penetrance outcomes in newborn sequencing would be.

A comprehensive health economic evaluation of the impact of screening for 200 conditions simultaneously is unlikely to be feasible, while addressing the question on a condition-by-condition basis is unlikely to be cost-effective. WGS may detect conditions for which high-risk, expensive treatments are available or for which no treatment is available. If more diseases are included in the newborn screening programme, adequately resourced referral pathways must be in place which will present a challenge for resource allocation in health care systems. There is a resource trade-off between early genetic diagnosis and intervention for less sick or asymptomatic individuals and resource-intensive diagnostic odysseys and later treatment when a rare disease presents symptomatically.^12^ Typically, an Initial step in a cost-effectiveness evaluation is to map out the respective clinical pathways for the intervention and the comparator(s), which is likely to consist of current clinical practice. Costs and health outcomes are then estimated based on the resources used and outcomes of each pathway. Mapping the full clinical pathways, i.e. diagnosis and treatment, for such a wide range of conditions where the patient is asymptomatic, is unlikely to be feasible. A different methodological approach is needed for the economic evaluation screening for potentially hundreds of conditions with a single test.

Finally, the use of WGS for newborn screening presents several ethical challenges. These include consent, data ownership and psychological implications. Since newborns cannot provide consent, the NBS screening programme in the UK relies on obtaining informed consent from parent(s)/guardian(s). However, obtaining fully informed consent for WGS could be challenging and parents may not feel that they can turn down screening.^13^ As such, seeking consent for WGS, where large panels of conditions are screened for simultaneously, in the same manner may be inappropriate. Beyond the consent issues, the possibility of retaining genomic data, linking results to health records, and re-evaluating data throughout a person’s life raises important questions about privacy and confidentiality, data ownership and secure data storage.^14^ This may have psychological implications and an understanding of how the public and new parents feel about WGS, making decisions about taking part, data security, communication of results, access to treatment and impact on the child and the whole family need to be understood.

We aimed to assess different evidence sources and approaches to evidence synthesis, review methods of costing, and collate views of parents with children with rare diseases and expectant parents on the main challenges of WGS to inform an approach to assess WGS for newborn screening in the future. To that effect, we undertook five traditional evidence reviews covering five conditions to establish the evidence base and to provide a reference case for comparison with alternative review approaches. We evaluated the use of the existing resource ClinGen as a shortcut into the evidence on actionability for the genes associated with the five selected conditions, where ‘actionability’ refers to the level of evidence about pathogenicity, penetrance and expressivity of a genetic variant and the extent to which interventions can be used to mitigate the effect of the disease.^15^ We undertook a focused review on genomic studies in paediartric screening populations reporting penetrance that may allow the Identification of gene variants with high penetrance and expressivity for a screening program that maximises benefits and minimise harms. We conducted a review of economic evaluations of WGS and WES to better understand the methodology employed to date to evaluate these tests. Finally, we explored patient and public views on questions relating to evaluating and communicating WGS in newborns.

## Methods

The methods for conducting this review were predetermined and published on PROSPERO (registration number CRD42023475529).

### 1. Review of five conditions

The traditional approach to evaluating the evidence for a new screening programme is to conduct a thorough systematic review and cost-effectiveness analysis of the test and condition to inform discussions around the extent to which UK NSC screening criteria are met. This traditional review approach was not considered to be feasible for each of the 200 rare diseases included in the Genomics England Generation Study.^6^ Instead, a series of five systematic reviews covering five of the 200 rare diseases was undertaken to establish the evidence base per condition and to provide a reference case for comparison with alternative review approaches.

#### Objectives

Six key Objectives aligned with evidencing UK NSC criteria were identified:

1. To identify the penetrance and expressivity of different gene variants associated with each condition in untreated infants/young people up to 18 years old,
2. To identify the proportion of infants/young people up to 18 years with biochemical or biochemical and clinical features of each condition carrying the genetic variants known for the conditions,
3. To evaluate the diagnostic accuracy (clinical validity) of gene sequencing for each condition,
4. To evaluate the effectiveness of earlier intervention (treatment or surveillance) for each condition, or, if comparative data on early vs late intervention is unavailable, to separately evaluate the effectiveness of treatment in screen detected cases and following clinical presentation,
5. To evaluate the effectiveness of WGS for newborn screening for each condition in terms of disease-related morbidity and mortality,
6. To identify any harms of WGS for newborn screening for each condition, and to identify any additional benefits beyond those afforded by earlier intervention.

The relevant UK NSC criteria per question are outlined in Table 1.

**Table 1.**
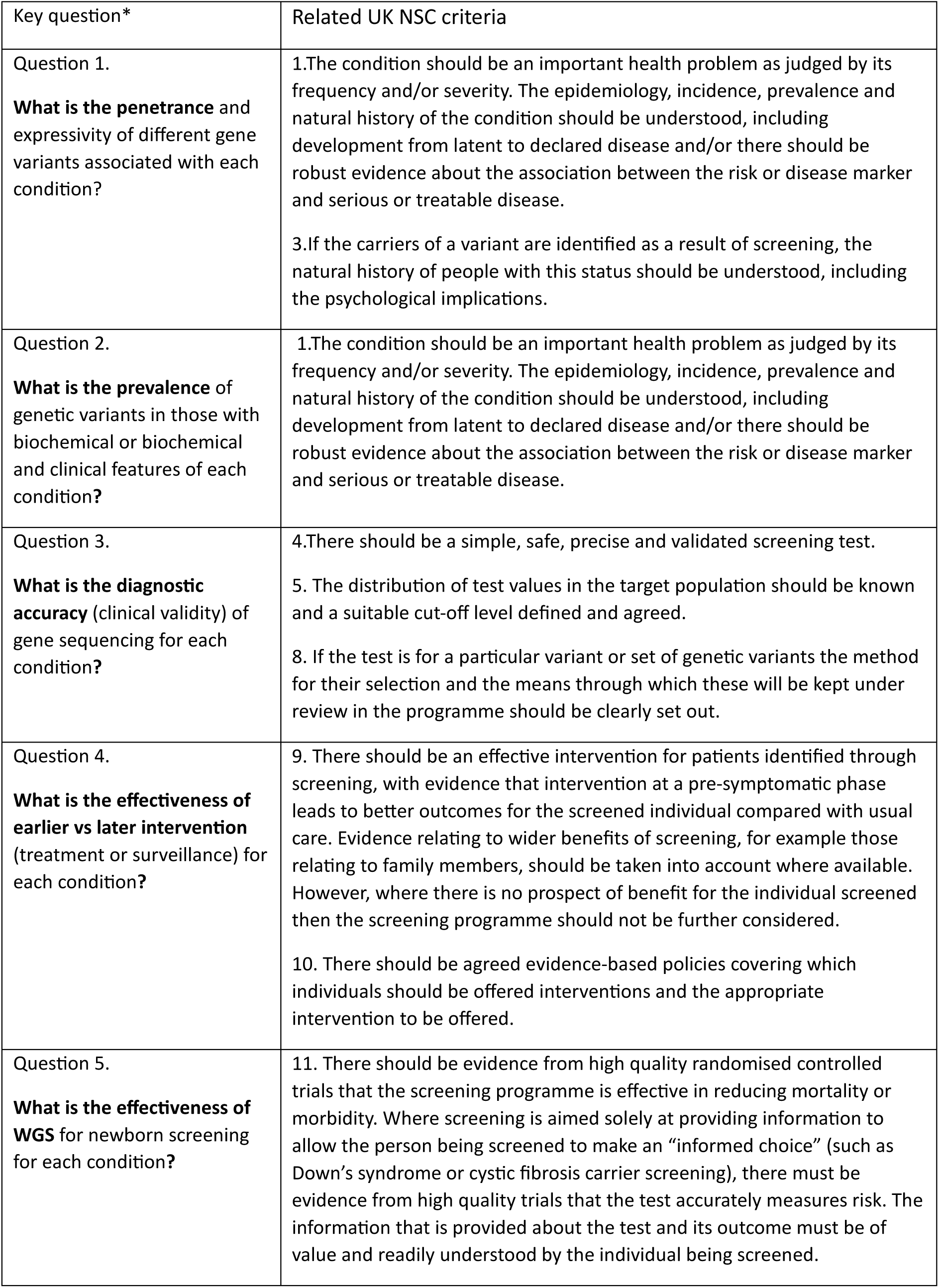
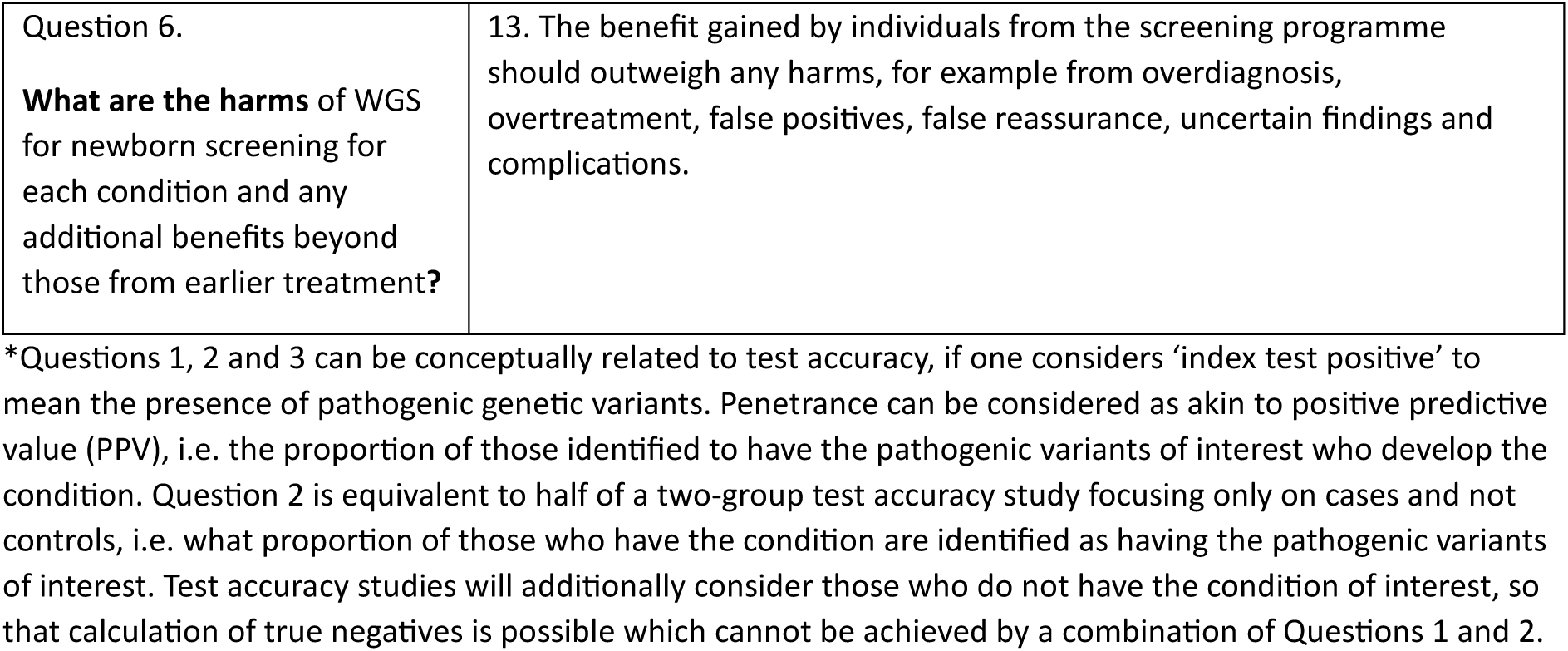
Key questions for the evidence summary, and relationship to UK NSC criteria.

#### Selection of five conditions for review

A range of scenarios that would be likely to impact the UK NSC’s advice on implementation of WGS for newborn screening were identified. These scenarios were primarily driven by the nature of the intervention(s) that could be introduced on detection of the condition, i.e.:

i. widely available and relatively low-cost treatment that carries a low risk of harm (e.g. adverse effects) for the patient and wider family, e.g. vitamin therapy
ii. intervention centred around long-term surveillance to allow earlier detection of the clinical manifestation of the condition with the associated anxiety and costs both in terms of resource use and time required to attend appointments, e.g. regular outpatient appointments to assess biochemical or developmental changes
iii. ongoing long-term high-cost treatment, potentially with more significant side-effects and requiring hospital visits and long-term monitoring, e.g. high cost or new drugs with unpleasant or unknown side effects
iv. potentially curative interventions carrying high short-term risks and costs to NHS but long-term lower impact on both NHS, patients and their families, e.g. stem cell transplantation
v. where existing screening and treatment pathways exist such that the impact of WGS would be incremental, e.g. hearing screening using Automated Auditory Brainstem Response (AABR test) and WGS

Genomics England Limited (GEL) provided a shortlist of 27 monogenic conditions which a) were considered to meet their four principles (GEL score 1, judgement in July 2023, see Table 12)^16^ and b) have a relatively high prevalence, within the context of rare disease. Following exclusion of conditions previously reviewed and not recommended for screening by the UK NSC, one condition with an intervention falling under each of the five scenarios above was selected at random. The final list consisted of the following five conditions (Table 9):

a. Pyridoxine dependent epilepsy (PDE): PDE is a neurological condition resulting from an enzyme deficiency which causes the accumulation of metabolites which inactivate pyridoxine. This pyridoxine depletion causes intractable neonatal seizures which become recurrent and prolonged if left untreated. Treatment with high dose pyridoxine (vitamin B6 supplementation) reduces the incidence and severity of seizures.
b. Heritable retinoblastoma (hRB): RB is cancer of the eye caused by disrupted function of the tumour suppressor protein RB. hRB occurs where both copies of the faulty gene *RB1* are present on conception (either inherited or occurring sporadically, referred to as ‘germline’ or ‘constitutional’ variants). Early intervention for hRB centres around regular ophthalmologic surveillance from birth to allow earlier Identification and treatment.
c. X-linked hypophosphataemic rickets (XLHR): XLHR is an endocrine condition caused by loss of function in the *PHEX* protein which ultimately leads to hypophosphatemia (low phosphate levels) manifesting as rickets. Calcitriol and oral phosphate can be used as preventive treatments; the only available curative treatment being monoclonal antibody burosumab (currently not licensed for <1-year olds)
d. Familial hemophagocytic lymphotistiocytosis (fHLH): fHLH is an immune deficiency caused by malfunction of the perforin/granzyme cytotoxic pathway leading to a proliferation of lymphocytes and overactive macrophages which ultimately lead to infiltration and damage of organs including the bone marrow, liver, spleen, and brain. fHLH is usually activated by infection and presents as an acute illness. Active disease can initially be managed using chemoimmunotherapy with allogenic haematopoietic stem cell transplantation (HSCT) providing curative treatment.
e. Medium Chain Acyl-CoA Dehydrogenase Deficiency (MCADD): MCADD is a metabolic condition, currently included in the UK newborn blood spot screening programme. MCADD is caused by inactivity or deficiency of the MCAD protein which affects proper functioning of the liver and can cause metabolic crises during periods of prolonged fasting or increased energy demands. MCADD is not curable but is managed preventively through dietary advice to avoid fasting and strict feeding regimens, and provision of an emergency regimen (glucose polymer feed) to be used during illnesses.

See Supplement 1 for full details of the five conditions.

The evidence was reviewed by condition with all genetic variants considered, rather than pre-specifying gene variants to be reviewed. This approach is not quite the same as WGS which aims to detect particular variants and therefore does not screen for conditions.

Reviews were undertaken using a rapid evidence assessment approach producing an Evidence Summary as described in the UK NSC guidance on the evidence review process.^7^

#### Identification and Selection of studies

##### Search strategy

Search strategies were developed for each condition by an Information Specialist. The searches were developed in a test database (MEDLINE (Ovid)) and were informed and refined through a series of scoping searches, checks of a proportion of results from these searches and iterative discussions between the Information Specialist (ND), project lead (KF) and members of the reviewing team (IK, JD, SC). The full process of search development is documented in Appendix 1.

The following databases were searched from inception to November 2023 (see Appendix 2 for exact dates and full search details): MEDLINE (via Ovid), Embase (via Ovid), Science Citation Index (via Clarivate) and the Cochrane Library (via Wiley). No date, language or study type filters were applied. Search results were managed using EndNote 20 and systematically de-duplicated using the University of Leeds method.^17^

##### Inclusion criteria

Study eligibility criteria were defined for each review question.

#### Population

For the evaluation of penetrance and expressivity (question 1), diagnostic accuracy of gene sequencing (question 3) and the effectiveness and harms and benefits from WGS (question 5 and 6), studies of newborn babies with no symptoms or known family history of the five selected conditions (i.e. newborn screening cohorts) were eligible for inclusion. Where evidence in newborns was limited, sibling or family studies or other approximations were considered.

For the evaluation of the prevalence of different genetic variants (question 2) and the evaluation of the effectiveness of earlier intervention (question 4), studies conducted in newborns, children and young people up to age 18 years with clinical, biochemical or genetic indicators of the five conditions were eligible. The clinical and biochemical definitions for each condition, where applicable, are provided in Table 2. Intervention studies (question 4) ideally included participants who were asymptomatic for the condition of interest (‘early’ intervention) and those treated following clinical presentation (‘later’ intervention).

**Table 2.**
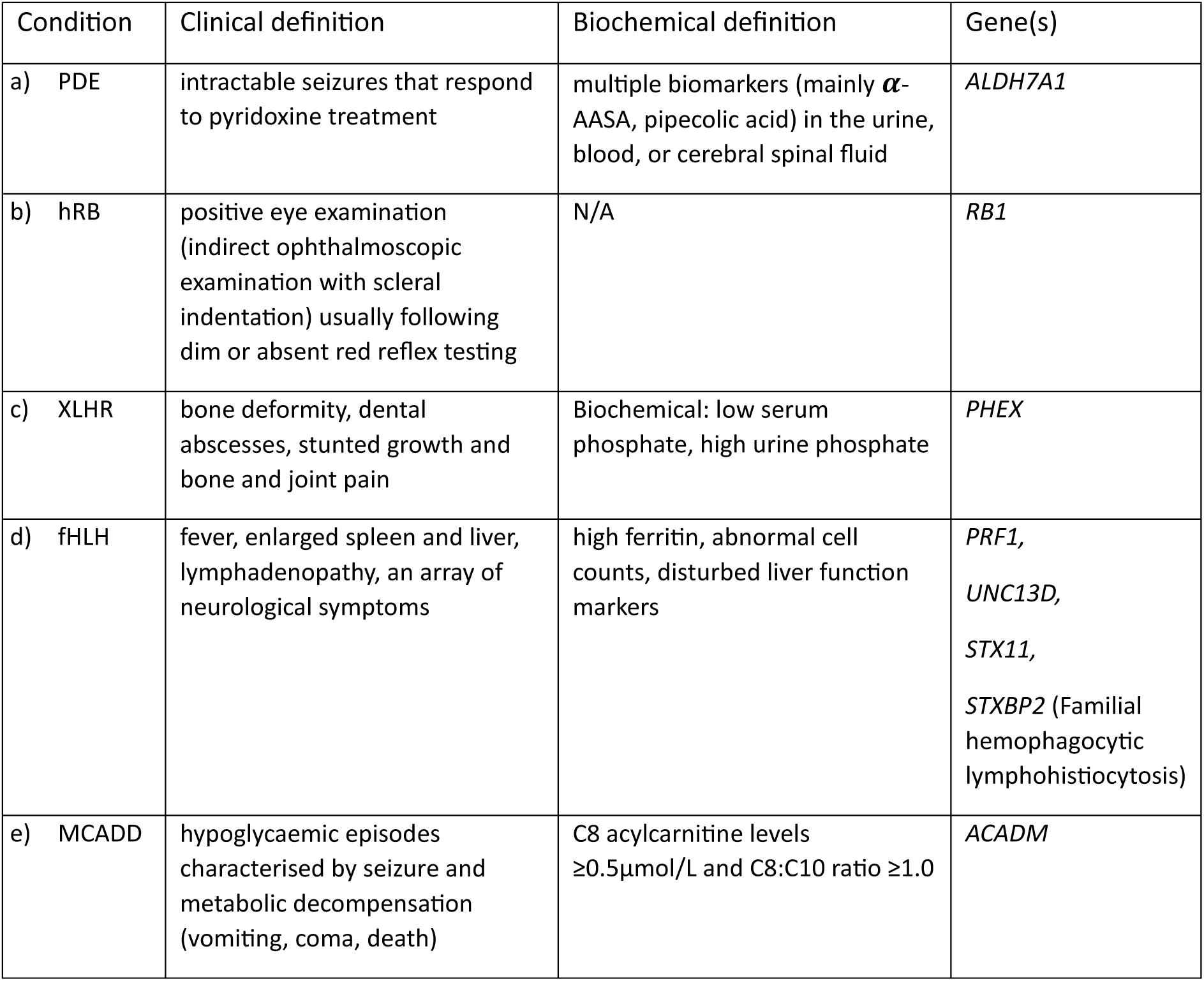
Clinical and biochemical definitions and genes associated with each selected condition.

#### Exposure / Intervention

For review questions 1, 2 and 3 the exposure was defined as the presence of pathogenic variants in the relevant gene(s) for each of the five conditions detected by direct sequencing using any technology (WGS, WES, next Generation sequencing (NGS), Sanger sequencing etc). The Identification of a genetic variant as pathogenic or ‘likely pathogenic’ is a complex process, however established classification systems, such as the one developed by the American Board of Medical Genetics,^9^ are often used. We relied on study authors’ classification of pathogenicity.

For review question 4 (effectiveness of earlier intervention), eligible interventions were defined separately for each condition as follows:

a. PDE: supplements of pyridoxine or vitamin B6; lysine reduction therapy (LRT)
b. hRB: regular ophthalmoscopic examination usually accompanied by red reflex testing
c. XLHR: supplements of oral phosphate, active vitamin D, monoclonal antibody Burosumab
d. fHLH: chemoimmunotherapy and allogeneic Hematopoietic stem-cell transplantation (HSCT)
e. MCADD: dietary advice

‘Early’ intervention was ideally defined as treatment or surveillance of newborns identified as positive for pathogenic variants of the relevant gene(s) (any test) for each condition or an approximation thereof, for example, treatment or surveillance of second or third siblings of the original proband who carried the same pathogenic variants. ‘Late’ intervention was considered to occur in symptomatically detected (without screening) newborns, children or young people up to the age of 18 for each of the five conditions or an approximation (anything else available). Where evidence was limited, any author-defined ‘early’ versus ‘late’ intervention was accepted.

For review questions 5 and 6 (effectiveness and harms from WGS), the intervention was screening using WGS.

#### Comparator

No comparators were considered for review questions 1 to 3.

For review question 4, the comparators were the same as the interventions per condition, introduced at a later time point in the disease, i.e. following symptomatic detection of the conditions (‘late’ as defined by the study authors).

For review questions 5 and 6, the comparator strategy was no screening using WGS, comparison with current practice (which in the UK is newborn blood spot screening for nine different rare diseases using methods such as tandem mass spectrometry), or no comparator.

#### Target condition (and reference standard)

For review question 1 (penetrance and expressivity) and question 3 (diagnostic accuracy of sequencing) the target conditions were clinically or clinically and biochemically defined conditions. For review question 2 (proportion of different genetic variants in those with the conditions of interest), the population and target conditions are the same (i.e. those with the conditions of interest). Each combination of biochemical and clinical features should be considered the ‘reference standard’ for presence of disease and calculation of diagnostic accuracy (review question 3).

#### Outcomes

For review questions 4 (effectiveness of earlier intervention) eligible outcomes were measures of disease-specific morbidity and mortality.

For review question 5 (effectiveness of WGS), any health-related health outcomes that could be measured across conditions (e.g. quality of life, time to diagnosis and intervention, mortality) were eligible.

For review question 6 (harms or other benefits from WGS in newborns), harms included effects associated with false-positive results, overdiagnosis (including Identification of variants of uncertain significance), ethical issues, parental or proband anxiety, referral to surveillance pathway, missing management pathways, data storage, adverse events from treating asymptomatic newborns. Potential benefits included greater certainty (doctors and patients), reduced anxiety, fewer investigations, appropriate surveillance or management plan (therapeutic yield), earlier diagnosis, earlier treatment.

#### Study designs

All the conditions selected for this review are considered rare, therefore we expected a relative lack of evidence for each of the six review questions. We did however rank study designs in order of priority for each review question. Systematic reviews were eligible for all six review questions.

For review questions 1 and 3, we considered the following types of primary study:

- observational studies of newborn screening population without treatment and follow up to disease,
- observational studies of any screening population with treatment and follow up to disease with matched comparator (or with no comparator),
- observational studies of screening of which only gene variant positives are included,
- case-control studies (review question 3 only)
- sibling studies,
- case series (i.e. more than one case or family)

For review question 2, any observational studies reporting sequencing results for individuals with the conditions of interest were eligible. A minimum of four cases with the condition was required.

For review questions 4, 5 and 6, randomised controlled trials, non-randomised studies, before-and-after studies, and other cohort studies were eligible.

##### Exclusion criteria

Papers that fulfil the following criteria were excluded:

- studies of people older than 18 years at diagnosis,
- studies of non-hereditary forms of the five conditions,
- qualitative studies,
- studies that provided insufficient information for assessment of methodological quality/risk of bias,
- studies reporting outcomes not relevant to our review questions,
- studies where more than 10% of the sample did not meet our inclusion criteria and are not reported separately,
- study reports not available in the English language,
- single case studies (studies of one case or one family; however, we reported the number of case studies per condition),
- letters, reviews, editorials, communications, conference abstracts, and other grey literature, publications that contained no numerical outcomes data.

##### Selection of studies

Titles and abstracts of records identified by the searches were screened by one reviewer. A random 20% sample of records were screened independently in duplicate by a second reviewer, and any records with any uncertainty over inclusion (coded ‘Maybe’ by either reviewer) were discussed and a consensus decision was reached. The full publications of all records selected were obtained and assessed by one reviewer, with a random 20% sample assessed independently by a second reviewer. Disagreements were resolved by consensus, or through discussion with a third reviewer.

#### Data collection and analysis

##### Data extraction

All data extraction were extracted into a piloted electronic data collection form. Data were extracted by one reviewer and checked by a second reviewer. Any disagreements were resolved by consensus or discussion with a third reviewer.

##### Assessment of methodological quality

a. At the outset we planned to carry out quality appraisal using dedicated tools for each study type, for example using the Risk of Bias in Systematic Reviews (ROBIS) tool for systematic reviews,^18^ the Quality in Prognostic Studies (QUIPS) tool^19^ or tools developed by the Joanna Briggs Institute.^20^
b. Ultimately, the majority of reports included were of relatively small series of patients with the conditions of interest. The decision was, therefore, taken to use a single tool designed for appraisal of case series and case reports, tailored to the review question.^21^ Use of a single tool allowed an overall picture of study quality across study types and conditions.

Quality assessment was conducted by one reviewer with all assessments checked by a second reviewer.

##### Methods for synthesis

We planned to employ an order of priority approach to synthesis, providing a narrative synthesis of studies that employ the highest priority design available with an accompanying tabulation of key details from studies using lower priority designs (e.g. study design, countries in which the studies took place, sample sizes, and key outcomes) (see Protocol for full details).

Due to the paucity of studies using higher priority study designs and the lack of available data for review questions 1 (penetrance and expressivity), question 3 (diagnostic accuracy of sequencing) and questions 5 and 6 (effectiveness and harms or other benefits from WGS in newborns), an alternative approach to synthesis was adopted.

Studies providing data for review question 2 (prevalence of different genetic variants in those with the condition of interest) were classified into subgroups according to the definition of disease using clinical and/or biochemical measures (broadest to narrowest) (Table 8) and the largest most representative study from each subgroup was synthesised. Data concerning study design, country in which the study took place, definition of the condition, number of participants, genes tested, and genetic tests used, gene frequency and types of variant identified were presented. Available data on expressivity in those with the condition was also presented. A subset of data items was reported for the remaining studies considered relevant for review question 2 (Appendix 3, Table 15).

Studies providing data for review question 4 (evidence on earlier versus later treatment) were classified into subgroups according to the definition of ‘early’ versus ‘late’ treatment that was used (Table 11). Studies using definitions most closely related to the review question were synthesised. Data concerning study design, country in which the study took place, definition of the condition, number of participants, definition of early and late treatment, outcome measures, time point of measurement and results were presented. A subset of data items was reported for the remaining studies considered relevant for review question 4 (Appendix 3, Table 18).

### 2. Exploring ClinGen as an evidence source

The existing online research Clinical Genome Resource, or ClinGen, was explored as a shortcut into the evidence base on the clinical actionability of rare paediatric genetic diseases (i.e. the risk of variants is known to be high, and intervening will prevent/mitigate disease) and findings were compared to the traditional reviews of the five conditions PDE, hRB, XLHR, fHLH and MCADD. The main aim was to assess the resource as a potential evidence source for the UK NSC considering that traditional reviews for over 200 conditions are unlikely to be feasible.

**Research question:** What is the utility of ClinGen as a shortcut into the evidence on clinical actionability for the genes associated with the five selected conditions and scaled up to 200 conditions?

#### Background ClinGen

The Clinical Genome Resource, or ClinGen, is a National Institutes of Health funded, open-access and centralised resource to define the clinical relevance and actionability of genomic variants. ClinGen’s action group developed practical methods to identify genetic disorders with the greatest clinical utility when detected in previously undiagnosed adults^22^ and adapted the methods to the paediatric context.^23^ Working groups use a standardised protocol to produce summary reports and semi quantitative metric scores for childhood onset rare genetic disorders. The methods to produce summary reports are adapted from work by Goddard et al. (2013) to guide decisions about returning incidental findings.^24^ The method provides a transparent, systematic, evidence-based process for identifying and quality rating of evidence. The evidence is reviewed by an expert panel which applies a semi-quantitative metric based on Berg et al. (2016),^25^ to score the overall clinical actionability of gene variants. Each topic is scored independently by multiple members and the scores are then discussed using consensus for assigning a single actionability score.

ClinGen focuses on four main curation activities: gene-disease validity (pathogenic variants in the gene clearly cause disease), dosage sensitivity (loss or gain of the gene results in disease), variant pathogenicity (categorisation of variants in the gene into benign, uncertain, pathogenic) and clinical actionability. As of March 2024, the database reports 6357 curated variants across 100 genes (not restricted to paediatric onset diseases) and includes 144 paediatric actionability reports.^23^ Conditions/genes have to meet a set of minimum requirements to be reviewed. These include:

1. Guidelines on an intervention relevant to an undiagnosed paediatric population exist (focus on disease prevention, lowering the clinical burden and improve clinical outcomes and not including ‘personal utility’, reproductive decision making, and ‘ending the diagnostic odyssey’).
2. At least one variant in the gene should have moderate to high penetrance (40% penetrance, relative risk=2) or no information on penetrance is available.
3. The health condition is significant.

Actionability reports determine the clinical actionability of secondary findings in paediatric patients undergoing clinically indicated diagnostic testing. Although the paediatric protocol states that elements relevant for population-based screening decisions may be captured, there is a lack of Consideration of systems-based practice and availability of population-scale follow-up. Actionability ratings from ClinGen are therefore insufficient for recommending screening in asymptomatic cohorts.^23^

Paediatric summary reports summarise the evidence on gene-condition pairs under four dimensions: severity of disease, likelihood of disease (similar to penetrance), effectiveness of intervention and nature of intervention. The dimensions are scored from 0-3 (3 being best for actionability) based on the evidence and the evidence is rated for the likelihood and effectiveness dimensions (poor evidence to substantial evidence). The scores are summarised across the four dimensions to provide an overall score for each gene-intervention pair. The scores and the evidence are taken into Consideration for the final assertion on actionability. The evidence review allows for a pragmatic approach allowing non-systematic and expert based references because it is recognised that this is the most commonly available evidence for rare genetic disorders.

#### ClinGen information for the five conditions reviewed

We searched the ClinGen database on 19^th^ February 2024 for each of the genes included in our review of five conditions and identified reports from the paediatric actionability working group. For each of the five conditions, we extracted the scores for condition-intervention pairs where available, the overall assertion, the evidence provided underlying the scores and the references cited. We tabulated scores and evidence level comparatively against the evaluation from Genomics England and against our assessment against the UK NSC criteria. We used the upper quintile of the ClinGen score as the cut-off for actionability as recommended by the Locus-Variant Binning Committee who developed the transparent semiquantitative metric for evaluating clinical actionability for pathogenic variants.^25^

We performed a narrative synthesis of our assessment.

### 3. Review of genomic studies of paediatric screening cohorts reporting penetrance for pathogenic variants

Knowledge of penetrance is important for the decision about which pathogenic variants of genes associated with childhood diseases should be reported for action following detection by sequencing. This is particularly important in the screening context to maximise the number of babies who are likely to benefit from detection at screening and those who are likely overdiagnosed (detection rate of pathogenic variants is not sufficient). The main aim of this focused review was to identify studies reporting penetrance as an outcome following sequencing in the paediatric screening setting for any paediatric condition to explore the feasibility of identifying highly penetrant pathogenic variants that could be considered for a screening programme (i.e. minimising harm from reporting variants of unknown or uncertain significance).

**Research question:** What is the penetrance or actionability of pathogenic/likely pathogenic variants of rare genetic child-onset diseases identified in newborn screening populations using WGS?

#### Identification and Selection of studies

Searches were developed iteratively in a single database (MEDLINE via Ovid). The final search combines the concept of newborn screening with either WGS, WES, penetrance, actionability, sequencing or allele frequency.

The following databases were searched from inception to January 2024 (Appendix 2 for exact dates and full search details): MEDLINE (via Ovid), Embase (via Ovid), Science Citation Index (via Clarivate) and the Cochrane Library (via Wiley). No date, language or study type filters were applied. Records were exported to EndNote and systematically de-duplicated using a process based on the University of Leeds method.^17^

#### Eligibility criteria

This approach relies on the availability of information on penetrance / expressivity and generalisability of findings to the screening context. We, therefore, only included studies of newborn screening populations that report as a minimum the penetrance (or an approximation) of gene variants linked to rare genetic diseases with childhood onset.

Studies that satisfied the criteria listed in Table 3 were included.

**Table 3.** Study eligibility criteria.

|  |  |
| --- | --- |
| **Population** | Newborn babies without symptoms or known family history of rare genetic diseases |
| **Target condition** | Any rare genetic condition with childhood onset of symptoms |
| **Exposure / Intervention** | Screening using WGS or WES<br>Step down: direct sequencing using panel tests or single gene testing in newborns without symptoms or known family history |
| **Comparator** | Order of priority (comparator is only needed for approximation of penetrance using allele frequency):<br>1. No comparator necessary<br>2. Randomisation WGS vs standard care<br>3. Contemporaneous cohort of matched / random newborns without genetic screening<br>4. General population of newborns without genetic screening<br>5. General population of healthy adults |
| **Outcomes** | For comparator 1:<br>a) follow-up without treatment to clinical presentation,<br>Step down:<br>b) follow-up to / comparison to biochemical tests (e.g. conventional NBS test) indicating presence of disease processes<br>c) categorisation into levels of penetrance based on existing literature<br>d) categorisation into pathogenic / likely pathogenic based on existing classification systems<br>For comparator 2 to 5:<br>Variant frequency matching the inheritance pattern of the relevant condition in screened and comparator to approximate penetrance |
| **Study designs** | Order of priority:<br>Observational study of WGS of asymptomatic newborns without reporting to parents (no treatment) and follow-up to clinical features (or reporting results but no treatment until symptomatic or reporting results and then later stopping treatment to determine whether necessary).<br>Observational study of WGS with follow-up testing (e.g. biochemical test)<br>Observational study of WGS follow-up to symptom onset despite treatment<br>Randomised trial of WGS (with results reported and treatment) vs standard care to determine variant frequency and approximate gene penetrance<br>Comparative studies using comparators detailed above to determine variant frequency and approximate gene penetrance |
| **Language** | English language |

Papers that fulfilled the following criteria were excluded:

Studies of populations other than newborns, studies on populations at risk or with symptoms, studies where WGS is second tier test, qualitative studies, studies only reporting variant frequency without an estimation of penetrance, studies only reporting carrier frequency, studies that provide insufficient information for assessment of methodological quality/risk of bias, studies reporting outcomes not listed in our inclusion criteria, studies where more than 10% of the sample did not meet our inclusion criteria and were not reported separately, articles not available in the English language, single case studies, letters, reviews, editorials, communications, conference abstracts, and other grey literature, publications that contain no numerical outcomes data.

#### Review strategy

Titles and abstracts of records identified by the searches were screened by one reviewer. A second reviewer independently assessed a random 20% sample of the titles/abstracts plus records labelled as unclear by the first reviewer. Disagreements were resolved by consensus. Full text articles were independently assessed against the inclusion/exclusion criteria by two reviewers. Disagreements were resolved by consensus. Records rejected at full text stage are listed in Supplement 2 with reasons for exclusion.

#### Data extraction strategy

Data were extracted into a piloted electronic data collection form by one reviewer and checked by a second reviewer.

#### Assessment of methodological quality

Methodological quality of each study was assessed based on study design as no appropriate tool was identified to assess bias in single arm cohort studies.

#### Methods for analysis/synthesis

We tabulated and narratively synthesised information on the proportion of sequencing positive cases, agreement with conventional screening results and the proportion with confirmed disease on clinical follow-up (penetrance) for each condition separately and for all conditions combined per study. We considered positive genetic screening outcomes as unconfirmed if within the studies’ type and length of follow-up the condition could not be confirmed with confirmatory testing and/or clinical follow-up. We provided our learning from this review for the context of WGS in newborn screening.

### 4. Review of cost-effectiveness evaluations of WGS and WES

Based on a quick search, we could not find any cost-effectiveness evaluations of WGS specifically for newborn screening. To help inform a future cost-effectiveness evaluation, we wanted to better understand the methodological approaches employed to date to evaluate the cost-effectiveness or cost of WGS. We also included studies which evaluated WES, as the methodological approach used in this context may also be relevant to an evaluation of WGS. We were particularly interested in the approaches taken to map and cost WGS/WES pathways and, where included, comparator pathways. We kept our search strategy and inclusion criteria broad to ensure that any WGS or WES studies conducted in a screening context would be identified.

Objective: To produce a methodological overview of published economic evaluations and costing studies of WGS or WES in any context or population.

#### Search strategy

Searches for cost or economic evaluations were conducted in the following databases in February 2024:

- MEDLINE (Ovid)
- Embase (Ovid)
- CEA registry
- Web of Science
- Econlit

Pre-print sources were not searched.

Our search terms were broadly based on those used in a previously published systematic review of WGS/WES cost-effectiveness studies.^26^ Searches combined concepts relating to sequencing analyses including, but not limited to, terms for WGS and WES and a) costing studies, b) budget impact studies, c) economic evaluations, and d) economic models. A previous scoping review Nurchis et al. (2022) was identified which focused specifically on health technology assessments (HTAs) of WGS.^27^ Targeted searches for HTAs were not included in the Schwarze et al. (2018) review, so we added targeted searches of Medline, Embase and International HTA from 2022 (i.e. post the searches conducted by Nurchis et al.(2022)^27^ to ensure Identification of any additional HTA reports that our original search may have missed. Searches were restricted to English language and humans. The search development methods can be found in Appendix 1 and the full search details are reported in Appendix 2.

Studies included in the Schwarze et al. (2018) systematic review^26^ and the Nurchis et al. (2022) HTA scoping review^27^ were assessed against our inclusion criteria. References of relevant systematic reviews were checked for any additional primary studies that were not identified by our search.

#### Inclusion and exclusion criteria

The eligibility criteria for the review (Table 3) broadly replicate those used in the Schwarze et al. (2018) review, apart from the outcome inclusion criteria.^26^ This review had a broader scope than our review in that it also included eight studies which only focused on health outcomes (even though the searches targeted economic evaluations). Because our inclusion criteria were restricted to studies which included costs as an outcome, we excluded these eight studies. We also excluded studies which focused on using WGS or WES to diagnose or monitor communicable diseases (e.g. bacterial infections). The reason for this was that the role of sequencing in this context was often very different compared to using it to diagnose a rare disease in an individual. For example, the motivation for sequencing was often to monitor for outbreaks and/or detect treatment resistance – potentially independent of the health of the individual concerned. The timing of the test is crucial in this context, and samples from multiple individuals were often batched together. We also excluded conference abstracts because word count restrictions meant that the studies could not be reported in sufficient methodological detail to facilitate meaningful evidence synthesis.

**Table 4.** Inclusion and exclusion criteria.

|  | Inclusion Criteria | Exclusion Criteria |
| --- | --- | --- |
| Population | Studies in Human Healthcare | Studies which focused on populations with suspected communicable diseases |
| Intervention | Whole-genome and whole-exome sequencing (any platform) | Targeted sequencing |
| Comparator | Any comparator or no comparator |  |
| Outcomes | Costs | Included health outcomes only |
| Study Design / publication type | Health technology assessments, costing studies, budget impact analyses, economic evaluations (partial and full), evidence-based guidelines, clinical trials | Conference abstracts |

Data were not extracted from included literature reviews, but the studies included in the reviews were checked against our database search results to ensure that we added any eligible studies that had been missed.

#### Screening

Initial screening of titles and abstracts, followed by full text screening was carried out using Rayyan.^28^ The titles and abstracts of records identified by the searches were independently screened by two reviewers (BS and AO). Where any disagreements occurred, the record was taken through to full text screening. Full texts were assessed against inclusion/exclusion criteria by one reviewer (BS or AO), with 20% independently checked by the other reviewer. Disagreements at this stage were resolved through discussion. There were some studies where it was unclear whether the test being evaluated was WGS or WES. These were shared with our genetics advisors for confirmation. The reasons for excluding records at the full text stage were documented.

#### Data extraction

An electronic data collection form was developed in Excel, piloted and refined. The form included the same study characteristics extracted in the Schwarze et al. (2018) systematic review (for continuity), but with additional components to capture additional methodological issues relating to costing of WGS/WES pathways and comparator pathways.^26^ Studies which produced a comparative analysis of costs and health outcomes were categorised as full economic evaluations. This included cost utility studies, where the outcome is defined as the cost per quality-adjusted life year, or cost-effectiveness studies, where the outcome is defined as the cost per change in a particular outcome. Partial economic evaluations, such as cost-consequence analyses, were defined as studies where costs and outcomes are reported, but they are reported in isolation. Costing studies are those which did not evaluate patient outcomes, and just reported costs.

Data were extracted by one reviewer (BS or AO), with a random 20% checked by the other reviewer. Disagreements were resolved by consensus or discussion with a third reviewer.

#### Critical appraisal

Given that the main focus of this cost-effectiveness review was to develop an understanding of the methodology used, rather than the results of the studies identified, we did not appraise the methodological quality of included studies.

#### Evidence Synthesis

For the studies meeting our eligibility criteria, we first provide a tabular/graphical overview of the key study characteristics, including number of publications by year, continent/country, the type of economic analysis conducted, which sequencing approaches were evaluated (WES, WGS or both), and the target population (grouped broadly by age).

We provide a narrative overview of the included studies, structured around the PICO:

- Population: who were the target population i.e. age, disease area
- Intervention: specifics about the timing of the test or aspects relating to how the test was conducted and integrated into the diagnostic pathway
- Comparator(s): what were the comparator(s) in the studies, if any, and what types of data were used to inform the comparator pathways and associated resource use
- Outcomes: what was the main outcome of the study e.g. cost per patient, cost per diagnosis, cost per quality-adjusted life year

We then focus on the methodological approaches adopted in the studies, including a narrative overview of the costing perspectives, time horizon, discounting, and whether any sensitivity or scenario analyses were conducted. A more in depth narrative description of the studies is then presented, focusing especially on the cost utility studies, given cost per quality-adjusted life year (QALY) is the standard outcome for UK NSC cost-effectiveness evaluations, and the costing studies which adopted a bottom-up approach to costing WGS as the resource use breakdown could be useful for future evaluations of screening newborns using WGS.

### 5. Consideration of the public voice in the evaluation of WGS

The broad aims of the Patient and Public Involvement and Engagement (PPIE) were to:

1. build an understanding of the perspectives and experiences of members of the rare disease community around the key challenges and opportunities of WGS in newborn screening
2. explore views towards, and understandings of, screening programmes and the role of the UK NSC
3. discuss methodological challenges identified during the review process
4. note views on limitations in the evidence base
5. contribute meaningfully to the development of future PPIE in this area.

Recruitment targeted people with differing lived experiences of rare genetic conditions (including parents and adults living with rare conditions), third sector representatives, and prospective parents from the public. Recruitment took place between October and December 2023. Due to the restricted timeframe, recruitment was initially through a targeted approach via known contacts at several third sector organisations and through Genomics England’s PPIE groups. Through these contacts, we recruited the charity representatives (including an adult living with a genetic condition). Four of the parent representatives were recruited through a post on a social media page for families living with rare genetic conditions and one approached the research team after reading a press release about the study. The public representative had previously been involved in PPIE work related to screening for a rare genetic condition, so they were able to meaningfully contribute without needing extensive guidance/input on genomics, screening and healthcare.

The Group consisted initially of eight members, five women and three men:

- A member of the public, who was also an expectant, and later a new, parent
- An adult living with a rare genetic condition, who was also a Diversity, Equity, Inclusion, and Belonging (DEIB) consultant and representative for a charity organisation supporting people living with a genetic condition
- Representative from a charity advocating for and supporting people living with genetic conditions
- Five parents of children living with a rare genetic condition (ages of their children ranged from 5 years old to young adults)

One of the parents withdrew from the group very early in the process due to their caring commitments.

Five (2-hour) virtual meetings took place between 15^th^ January and 21^st^ May 2024 (For discussion topics, see Table 5). Meetings 1 and 4 included the involvement of a member of the review team (KF) to present the review and its progress, with the remaining three meetings being independent of the review process. Meetings were recorded to facilitate note taking. To maximise inclusion participants who were unable to join any of the meetings were offered the opportunity to catch-up by watching recordings of missed meetings (with the permission of the participants present at the meeting) and sending feedback via email.

Meetings were deliberative, to create the knowledge space required for discussion, drawing on relevant evidence and the experience of the participants. Ahead of each meeting, participants were informed of the topic for discussion and given up to an hour of preparatory work in the form of reading from a variety of sources and/or formulating ideas/questions.

Following each meeting, participants were asked to complete a short evaluation questionnaire. This had two main purposes, (1) to evaluate the format and content of the meeting, (2) to allow participants time to reflect on the content of the meeting and provide any short summary points on this. In the final meeting, which brought together the group’s thinking on all of the topics covered, participants were asked to contribute to an online whiteboard (before, during and after the meeting) with their responses to prompts: what do you see as the main or most important (1) benefits and (2) harms to WGS for screening in newborns, (3) how should potential harms be prevented/ dealt with, (4) how should benefits and harms be balanced against each other, (6) what kinds of evidence should the UK NSC prioritise and (7) what should happen where the evidence is not available, and finally (8) how should PPIE contribute to this process (including when and who)?

**Table 5.** Schedule of PPIE Meeting Discussion Topics.

| Meeting | Topic |
| --- | --- |
| 1 | Introduction to the group and project<br>Team and PPIE group introductions. Presentation on the purpose and process of the review. Answering questions from the PPIE group on this. |
| 2 | Harms and Benefits<br>Pre-reading & presentation on: background/context to newborn screening, concept of harms and benefits, the role of the UK NSC, how WGS differs from current screening, and that different stakeholder groups may have different views. Open discussion on "What do you think are the key harms and benefits of screening that policy makers should be taking into account". |
| 3 | Genetic Uncertainties<br>Pre-reading & presentation: current understanding of genetic uncertainties, including defining terms such as penetrance, expressivity, variants of uncertain significance. Open discussion on the impact of this information on balancing harms and benefits and how to approach these complexities when explaining genetic screening to parents. |
| 4 | Systematic Review Findings<br>Pre-reading: executive summary of draft review and open discussion on the methodological challenges and evidence gaps identified during the review process and the conclusions reached. |
| 5 | Conclusions & Role of PPIE in Future Reviews<br>Open discussion on topics including: the most important harms and benefits; how these can be captured and traded off; how these harms and benefits can be transformed into evidence to support the decision-making of the UK NSC; how can PPIE be best incorporated into future reviews and who needs to be involved in these conversations |

## Results

### 1 Using a traditional review approach

This traditional review approach covers a stratified random sample of five of the 200 rare diseases included in Genomics England’s Generation Study^6^ to establish the evidence base per condition and to provide a reference case for comparison with alternative review approaches.

#### Workload and implications for scaling up to more than five conditions

Extensive scoping, refinement, testing and running of the electronic searches for the five conditions took a total of six weeks. We cannot envisage any way in which this could be simplified or shortened in duration. A traditional review for 200 conditions would require 200 individual searches to be developed.

The average time required to sift 100 abstracts was 40 minutes across five reviewers with varying levels of systematic review experience. A single full text sift took an average of 5 to 10 minutes depending on the question and the condition. Table 6 reports the approximate time required for searches and sifting activities for the five conditions, with extrapolation to 200 reviews of 200 conditions. This does not include time for double sifting, data extraction and quality assessment, discussion of disagreements, and categorisation of studies for synthesis purposes, i.e. by population for genetic testing (review question 2) and by timing of treatment initiation (review question 4). These time estimates, therefore, only illustrate the extrapolation of two reviewing tasks and do not reflect the time needed for a complete review process. The complete review process took a team of three full time and two part time reviewers seven months. We anticipate that a single similarly sized review team would need about 280 months or 23 years to review and synthesise the evidence for 200 conditions. The approximate time estimates demonstrate the scale of the effort required to evaluate WGS on a condition-by-condition basis, although some learning may be transferable to conditions similar to those reviewed here.

**Table 6.** Approximate time (in hours) needed for searches and sifting for a traditional review of 200 conditions extrapolated from five individual reviews.

|  | Approx time taken (hour) to develop searches | Approx time (hour) taken to run searches | Approx time (hour) to sift titles/abstract | Approx time (hour) to sift full texts |
| --- | --- | --- | --- | --- |
| 5 conditions | 47.5 | 21 | 131 | 170 |
| 200* conditions | 1900 (253 full time working days) | 840 (112 full time working days) | 5241 (699 full time working days) | 6795 (906 full time working days) |

\*Numbers multiplied by 40 to give estimated number for 200 conditions

#### Volume of evidence for the six review questions for five individual reviews

We sifted a total of 19,689 title and abstracts, of which 1,348 were selected for full text assessment (range 55 to 449 per condition). A total of 221 studies were eligible for inclusion across the five reviews (range 31 to 78). Table 7 summarises the search and eligibility results across conditions, with flow diagrams per condition presented in Appendix 4 (Figure 2 to Figure 6). The excluded studies and reasons for exclusion are listed in Supplement 2. Extrapolating to a review of 200 conditions, we could expect as many as 787,560 titles and abstracts and 53,920 full texts to be sifted.

**Figure 1.**
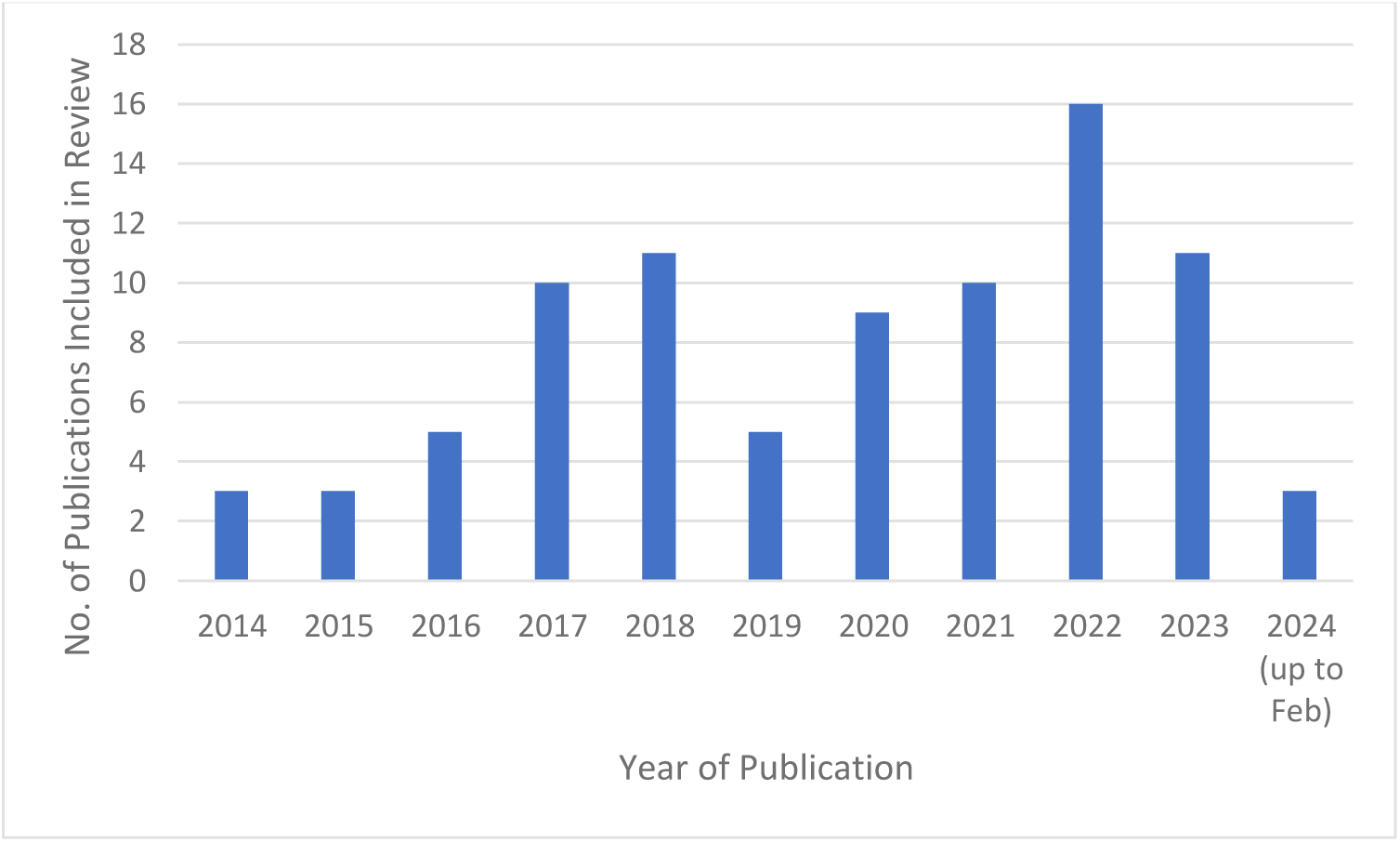
Number of studies included in our review by publication year.

**Figure 2.**
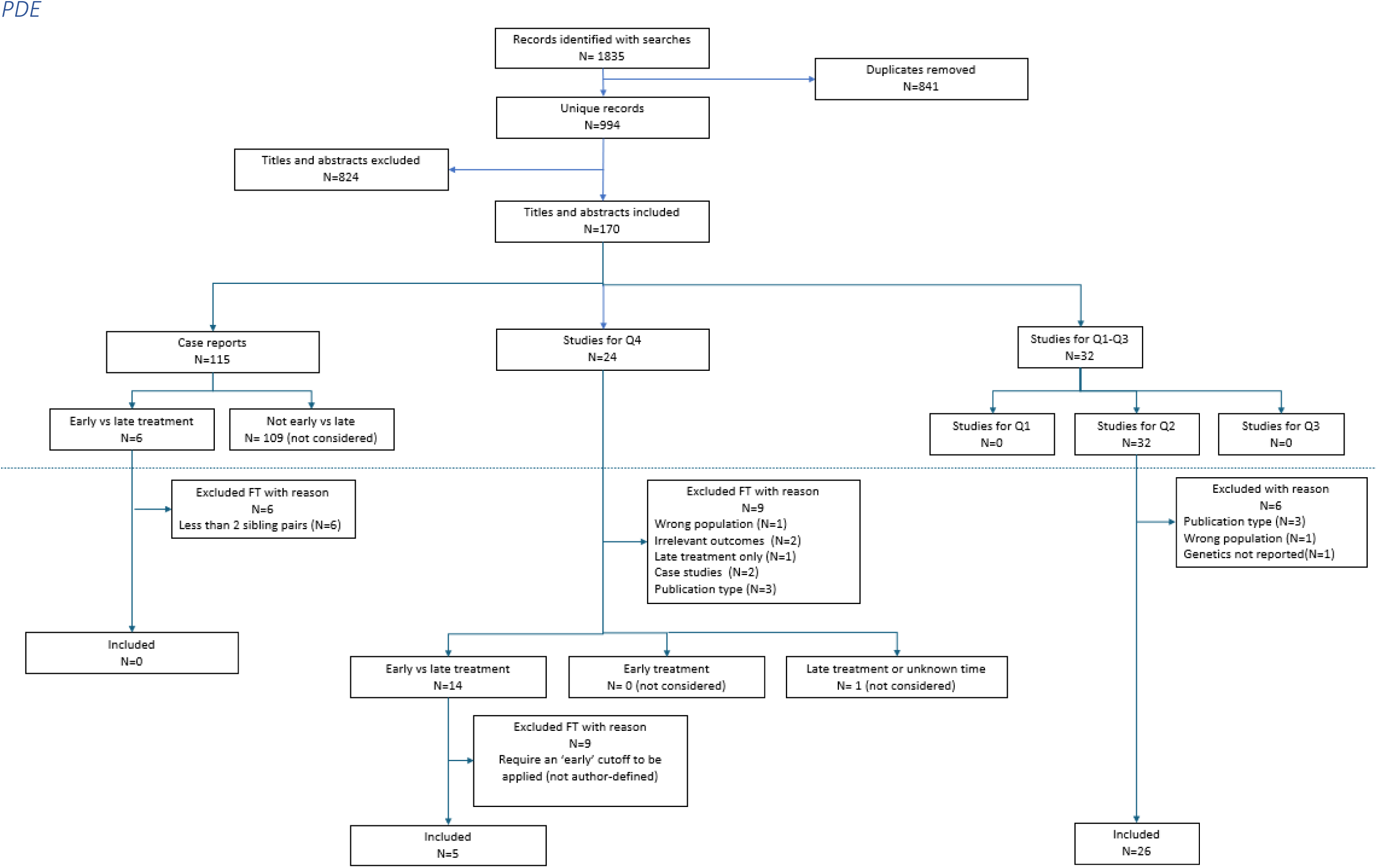
PRIMA flowchart for the review of PDE.

**Figure 3.**
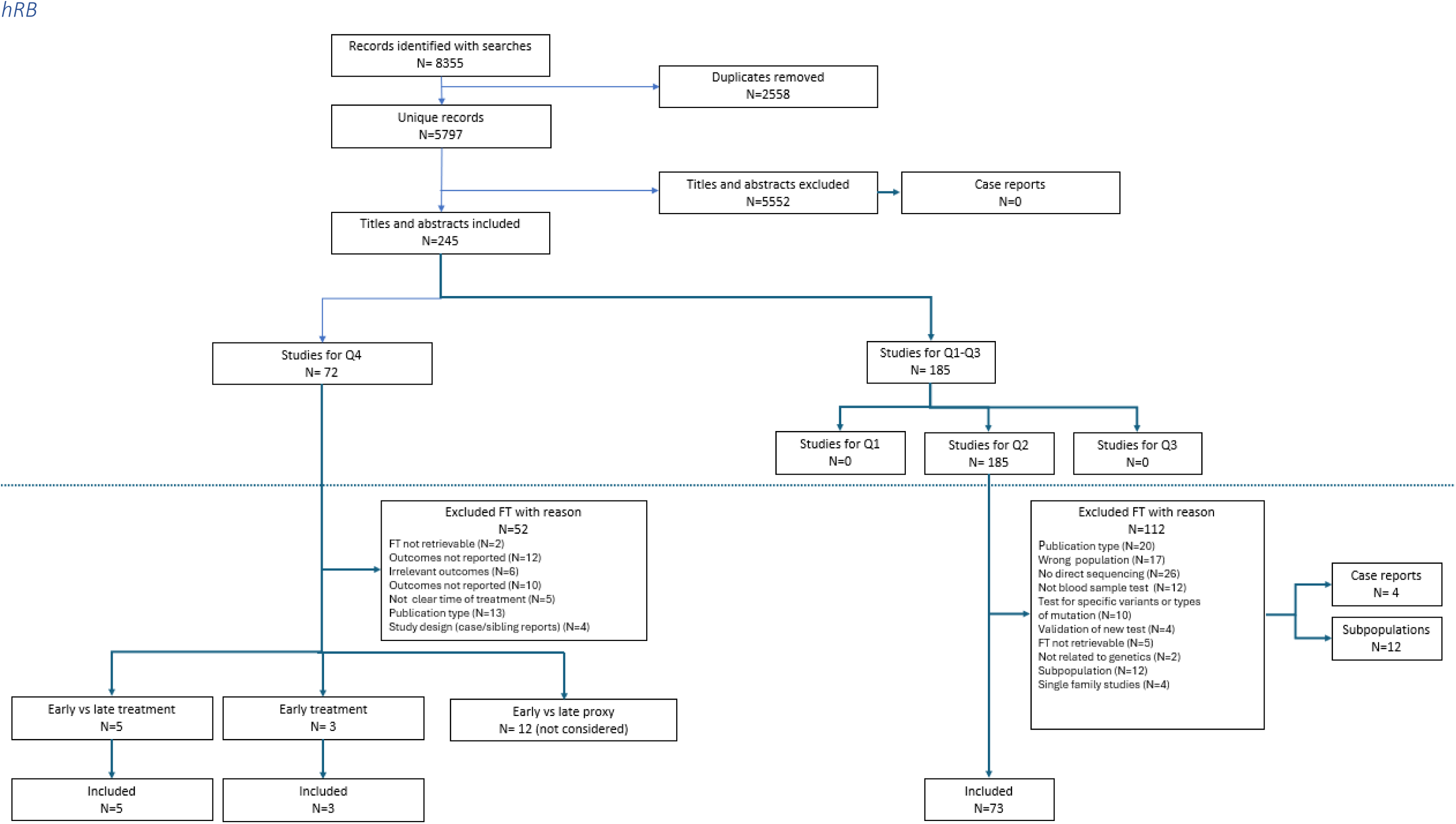
PRISMA flowchart for the review of hRB.

**Figure 4.**
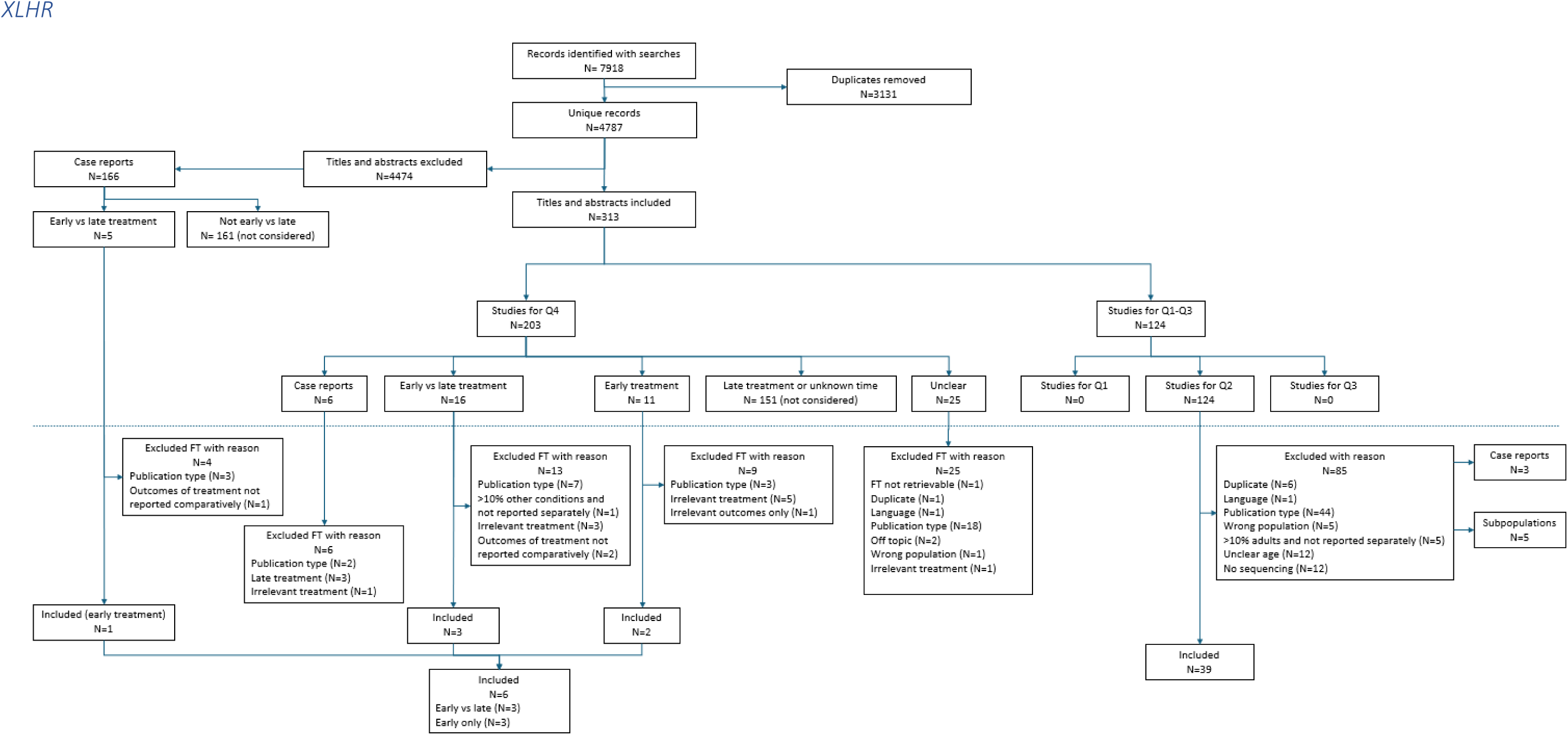
PRISMA flowchart for the review of XLHR.

**Figure 5.**
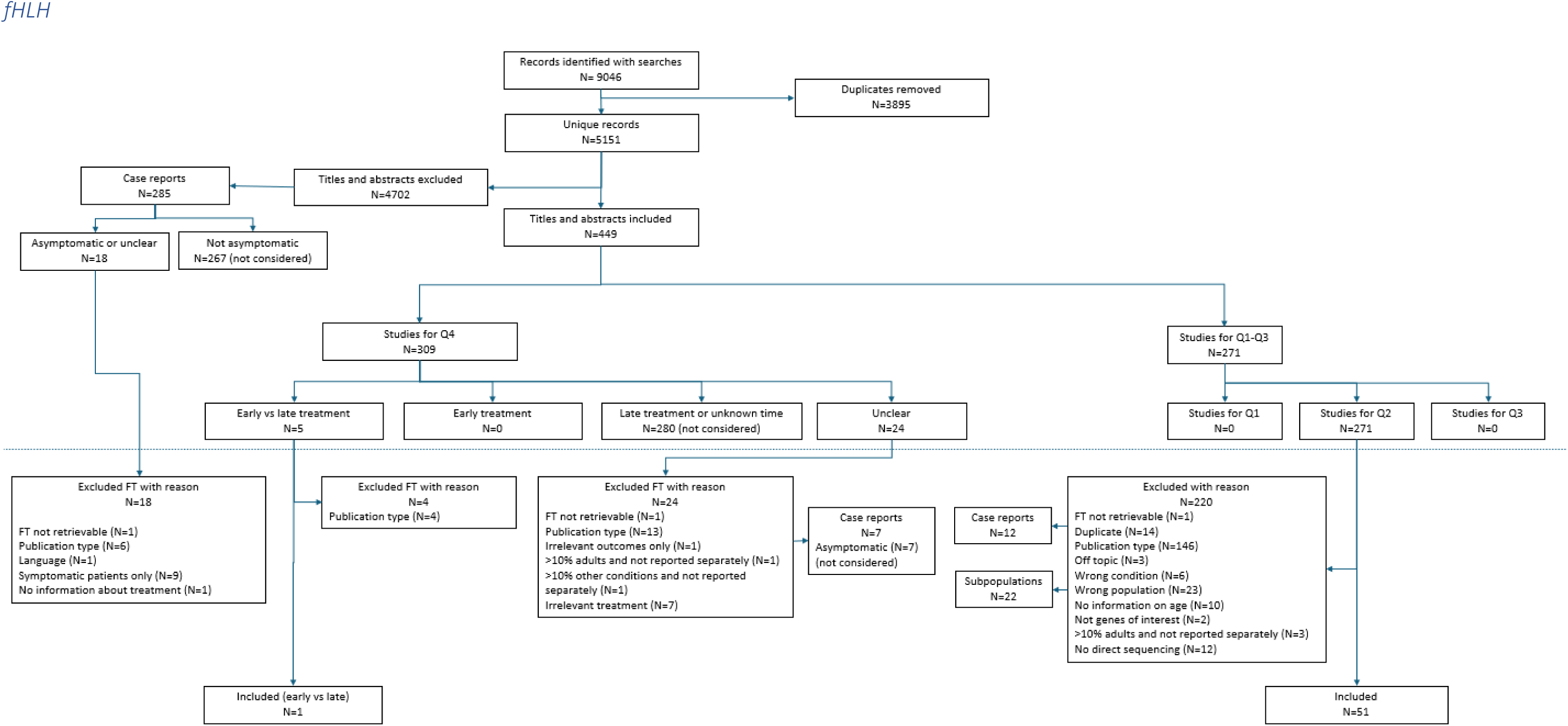
PRISMA flowchart for the review of fHLH.

**Figure 6.**
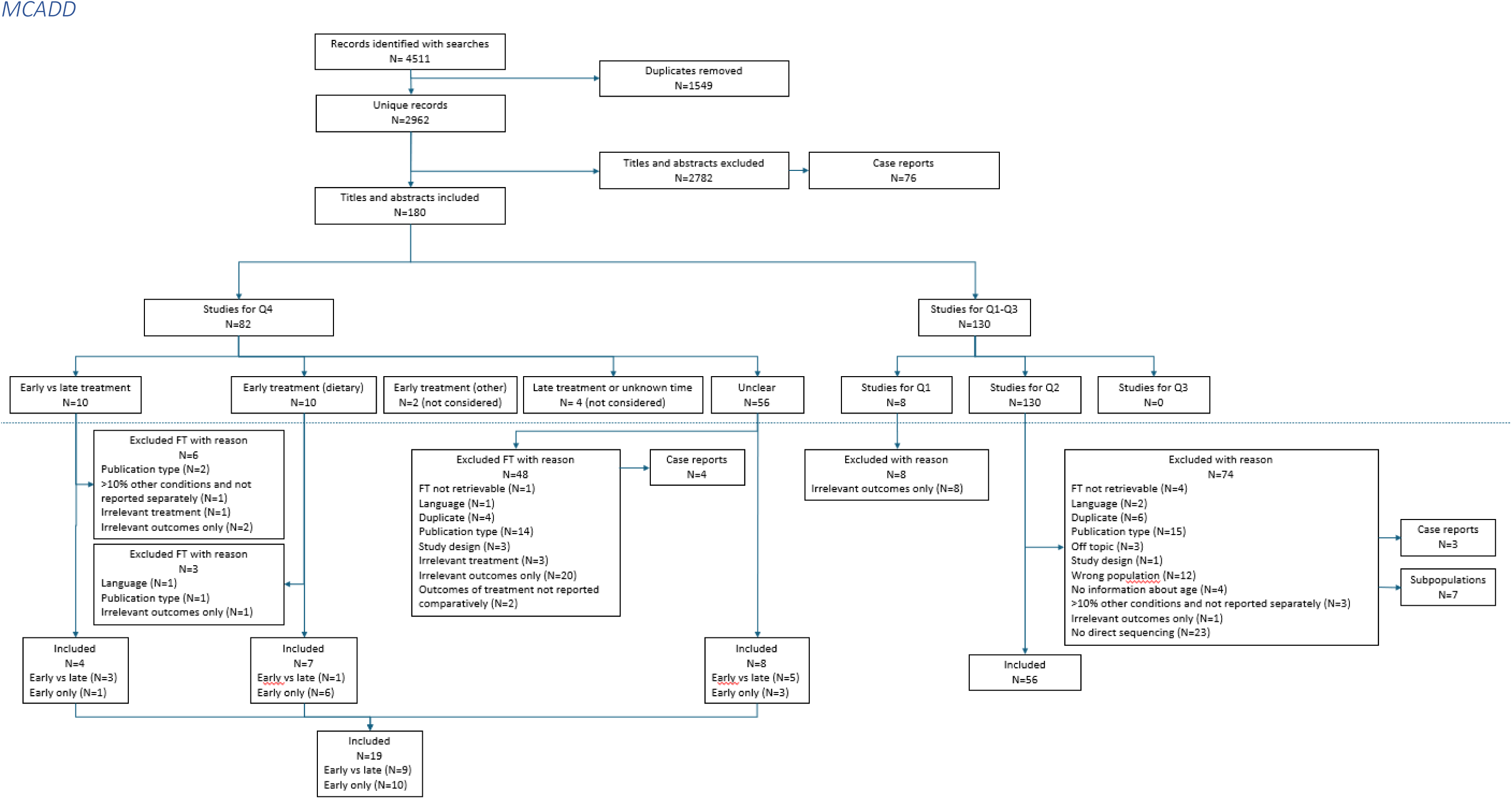
PRISMA flowchart for the review of MCADD.

No evidence was identified for four of the pre-specified review questions for any of the five selected conditions (Table 7). All four questions were those for which studies were required to be conducted in newborns:

- penetrance of the condition in a newborn cohort where sequencing was the first line screening test (Q1),
- test accuracy of WGS in the newborn screening setting (Q3),
- clinical effectiveness of WGS in newborns (Q5),
- harms or additional benefits from WGS in newborns (Q6).

Evidence was identified only for the two review questions focused on individuals with the conditions of interest (Table 7), i.e.:

- the prevalence of genetic variants and genetic spectrum in a paediatric population with the condition of interest (range 31-78 studies) (Q2), and
- evidence for early vs late treatment, where ‘early’ was the closest approximation to management of screen detected cases that was identified (range 1-9 studies).

Considering that the five conditions are rare diseases, the number of identified records was larger than expected. For some conditions such as PDE, data for the same study participants is likely to have been represented in multiple publications by different authors but this was not always clearly identifiable within the study reports. Overall, however, the five individual reviews yielded very little evidence of the sort required by the UK NSC.

**Table 7.** Volume of evidence for five conditions taking the traditional review approach and an extrapolation to a review of 200 conditions.

|  | **PDE** | **hRB** | **XLHR** | **fHLH** | **MCADD** | **Total** | **200 conditions\*** |
| --- | --- | --- | --- | --- | --- | --- | --- |
| **Titles and abstracts sifted** | 992 | 5,797 | 4,787 | 5151 | 2,962 | 19,689 | 787,560 |
| **Included on title and abstract** | 170 | 245 | 313 | 449 | 180 | 1,357 | 54,280 |
| **Included at full text** | 31 | 78 | 42 | 52 | 65 | 221 | 8,840 |
| **Number included per review question (brackets indicate n records selected for extraction)** |  |  |  |  |  |  |  |
| **Question 1:<sup>1</sup> Penetrance** | 0 | 0 | 0 | 0 | 0 | 0 | 0 |
| **Question 2:<sup>2</sup> Genetic spectrum in patients** | 26 | 73 | 39 | 51 | 56 | 260 | 10,400 |
| **Question 3:<sup>3</sup> Test accuracy** | 0 | 0 | 0 | 0 | 0 | 0 | 0 |
| **Question 4:<sup>4</sup> Early vs late treatment** | 5 | 5 | 3 | 1 | 9 | 22 | 880 |
| **Question 5:<sup>5</sup> Effectiveness of WGS** | 0 | 0 | 0 | 0 | 0 | 0 | 0 |
| **Question 6:<sup>6</sup> Harms of WGS** | 0 | 0 | 0 | 0 | 0 | 0 | 0 |
| <sup>1</sup> What is the penetrance and expressivity of different gene variants associated with the five conditions in untreated infants/young people up to 18 years?<br><sup>2</sup> What proportion of infants/young people up to 18 years with i. biochemical, and ii. biochemical and clinical features of the five conditions carry the gene variants known for the conditions?<br><sup>3</sup> What is the diagnostic accuracy (clinical validity) of gene sequencing for each of the five conditions?<br><sup>4</sup> What is the evidence on early (following screen detection or sibling detection (cascade testing)) versus late (following clinical presentation) treatment?<br><sup>5</sup> What is the effectiveness of WGS for newborn screening for each condition?<br><sup>6</sup> What are the harms of WGS for newborn screening for each condition and any additional benefits beyond those from earlier treatment? |  |  |  |  |  |  |  |

\*Numbers multiplied by 40 to give estimated number for 200 conditions

#### Findings from the traditional review approach for the 5 conditions

##### Gene/ variant frequency in patients with condition(s) of interest

Review question 2 looks to identify the proportion of infants/young people up to 18 years with either i. biochemical or ii. biochemical and clinical features of the five conditions (PDE, hRB, XLHR, fHLH, and MCADD) that carry the gene variants known for the conditions. This question can be considered in multiple parts. First and foremost, the idea was to compare the genetic spectrum in children with disease to that in sequencing positive newborns (Question 1) to assess the difference of clinical and genetic disease. Secondly, to identify the proportion of individuals with the conditions who have an underlying genetic cause (that can be identified by WGS) and the proportion of patients who may be missed because of non-genetic cause of the condition or because of non-specific symptoms caused by variation in a different gene. Thirdly, Consideration of the genetic spectrum in those with the condition to identify whether recurrent (or ‘reported’) genetic variants are responsible for large proportions of cases or whether novel variants that can be more analytically intensive to identify are frequently responsible. Finally, any reported patterns of expressivity associated with different variants or types of variants was considered.

The extent to which identified studies can inform these questions depends on the type of genetic test used and the inclusion of specific genes or genetic variants in the studies, as well as the nature of the included target population. The target populations identified did not easily fit into the two prespecified categories of clinically defined and clinically/biochemically defined disease and extended to genetically defined disease. Disease specific categories were therefore used that ranged from clinical symptoms, with or without biochemical markers, to study inclusion based on the presence of a genetic variant (Table 8). The categories were based on the expectation that the differences in the definition of disease may have an impact on the genetic spectrum reported.

**Table 8.** Categorisation of disease per condition and number of studies identified per disease category based on study populations.

|  | **PDE** | **hRB** | **XLHR** | **fHLH** | **MCADD** |
| --- | --- | --- | --- | --- | --- |
|  | **Disease definitions per condition (n studies)\*** |  |  |  |  |
| **Disease definition from broadest (top) to narrowest (bottom)** | Childhood onset pharmaco-resistant seizures<br>(n=6) | Any RB<br>(n=55) | Hereditary rickets (n=1) | HLH (n=38) | Inborn Errors of Immunity / Inherited Metabolic Disease (IEI/IEM) (n=2) |
|  | Clinical-biochemical PDE<br>(n=9) | Sporadic RB<br>(n=6) | Hypophosphatemic rickets<br>(n=20) | fHLH (two or more of *PRF1*, *UNC13D*, *STXBP2* and *STX11*) (n=2) | Fatty Acid Oxidation Disorders (FAOD)<br>(n=1) |
|  | Gene (*ALDH7A1*) positive PDE<br>(n=11) | Familial/germline / bilateral RB<br>(n=12) | Hereditary hypophosphatemic rickets (n=5) | Gene (*PRF1*) positive (n=6) | Acyl CoA Dehydrogenase Deficiencies (ACAD) (n=1) |
|  | - | - | x-linked hypophosphatemic rickets<br>(n=13) | Gene (*UNC13D*) positive (n=3) | Clinical-biochemical MCADD (n=51) |
|  | - | - | - | - | Gene (*ACADM*) positive MCADD<br>(n=1) |

\*the number of disease categories varies from three to five between conditions

Table 8 summarises the volume of evidence by disease definition for the five selected conditions. These definitions were condition-specific according to the availability of biochemical markers (not available for hRB), the number of umbrella terms of disease with overlapping symptoms (XLHR, hereditary hypophosphatemia and hypophosphatemia) and whether conditions can only be defined by genetic information (four conditions of fHLH).

Below we report the largest most representative study from each subgroup per condition; some subgroups have more than one study where results are complementary. Details per study regarding gene/ gene variant frequency are reported in Appendix 5. An overview of all included studies is included in Supplement 3. The detailed quality assessment of extracted studies is presented in Appendix 3 (Table 16, Table 17 and Table 19). Given the differences in populations and variant frequencies in the selected studies resulting from the different disease definitions, synthesising across studies was challenging. We therefore describe results per disease definition beginning with the broadest, with a particular emphasis on the disease category that is most relevant to WGS in newborns. Then, a short summary of results across the disease definitions is provided and quality assessment results are briefly discussed. Because the frequency of genes and variants identified in a population does not depend on the sequencing method, we did not prioritise studies based on sequencing method used.

#### Pyridoxine dependent epilepsy

The three subgroups and studies for PDE were: childhood-onset, pharmaco-resistant seizures^29^ clinically or biochemically defined PDE,^30^ and genetically-confirmed PDE (all with variants in *ALDH7A1*).^31, 32^ Details per study are provided in Appendix 5, Table 20. Three of the four studies were retrospective,^29–31^ including one multi-centre registry-based study.^31^ Two studies exclusively used sequencing to identifyvariants, two used targeted panels^30, 33^ and one used WES followed by short- or long-read genome sequencing for cases not solved by WES.^29^ The registry-based study did not report the genetic tests used to identify variants.^31^

##### Childhood-onset pharmaco-resistant seizures

The broadest category defined for PDE is a clinically defined population of children with early onset (neonatal or at latest infantile-onset) seizures that have been shown to be resistant to one or more standard pharmacological treatments for epilepsy (i.e. refractory or pharmaco-resistant). This is the group in whom PDE might ultimately be suspected on clinical presentation. Potentially hundreds of genes can be associated with epilepsy, however only a relatively small number (including *ALDH7A1*) have therapeutic implications (i.e. Identification of the gene directly informs treatment options). Other (treatable) pyridoxine responsive seizures include pyridoxal phosphate-responsive seizures (resulting from variants in *PNPO*) or pyridoxal phosphate-binding protein (PLPB) deficiency (resulting from variants in *PLPB*).

Boonsimma et al. (2023) included 103 cases with infantile-onset (age ≤12 months) pharmaco-resistant epilepsy seen or referred for genetic testing at a Thai tertiary care centre.^29^ Genes associated with Genetic Epilepsy Syndrome (n=728) were initially targeted using WES, followed by sequencing for additional selected candidate pathogenic variants in unsolved cases (only one heterozygous variant identified).

Of 103 cases, 6 (5.8%) were identified as having biallelic variants in *ALDH7A1*; 4 (66%) were identified on Initial WES and 2 (33%) required short-(n=1) and long-read (n=1) sequencing to identify two novel variants (all cases and both parents underwent sequencing). Two additional patients were identified as having ‘treatable’ disorders, one with a biallelic *PNPO* variant and one with a *BTD* variant. An additional 36 patients were identified as having genetic variants that could ‘inform’ treatment decisions.^34^

A total of eight *ALDH7A1* variants were identified in the six patients, five recurrent and three novel variants (2/3 were Copy number variants (CNVs)). Of the five recurrent variants, two occurred in more than one patient within the study sample, suggesting a possible founder effect (one was identified in four patients, and one in two patients).

##### Clinical-biochemically defined PDE

Traditionally PDE was clinically defined based on seizure recurrence (increase in number or severity) following pyridoxine withdrawal. Increasingly urinary or blood-based biomarkers (elevated α-AASA, piperidine-6-carboxylic acid, and pipecolic acid concentrations) are used to define PDE and may help to distinguish it from other pyridoxine responsive seizures such as pyridoxal phosphate-responsive seizures (*PNPO* gene) or pyridoxal phosphate-binding protein (PLPB) deficiency (*PLPB* gene). As PDE is an autosomal recessive disorder, those affected are usually children of unaffected carriers of the *ALDH7A1* variant, such that family history is not a strong indicator of the likelihood of PDE, although more than one sibling in a Generation can be affected. Further, there is an increased risk in consanguineous families. It is important to note that this category includes those who have either clinically, or biochemically defined PDE, or both, and this difference in disease definition may have an impact on results obtained from sequencing.

Koul et al. (2019) included all children (n=35) from a single centre with refractory neonatal or infantile seizures that did not respond to antiepileptic drugs, but later responded to pyridoxine.^30^ Only 7.9% (3/35) had elevated biochemical markers for PDE (pipecolic acid and AASA). All probands received ‘targeted variant testing in *ALDH7A1*’, but no further detail was given to describe the nature of this test. If the Initial test was negative, WES was conducted on multiple genes including *PLPBP, PRRT2* and *ALDH7A1*. The full spectrum of genes considered for WES was not reported.

Of the 35 cases, four (11%) were identified as having a variant in *ALDH7A1*, including the three with biochemical indicators of PDE, but it is unclear whether these variants were detected by the first line testing strategy or by WES. The number of patients with homozygous and compound heterozygous variants was not reported. Twelve (34%) patients were identified as having a *PLPHP* variant and two (6%) had a variant in the *PRRT2* gene, detected through WES. Specific variants were not reported.

Age at seizure onset and developmental delay were reported separately for patients positive for *ALDH7A1* variant (n=4), those positive for a *PLPBP* variant (n=12), and those with neither of these variants (n=19). Age at seizure onset was lower for those with *ALDH7A1* variants (30 minutes to 1 hour) and delayed development reported in a higher proportion (75% (3/4)) compared to the other groups. For those with *PLPBP* variant, seizure onset was between one hour and ten days old and 17% had developmental delay; for those with neither variant, seizure onset was reported at four hours to 29 months and 37% (7/10) had developmental delay. It is not clear to what extent these differences are a result of Selection into the study, and furthermore, based on the small numbers reported, results cannot be considered representative of the whole population of patients with clinically-biochemically defined PDE.

##### Gene (ALDH7A1) positive PDE

The most tightly defined populations are those defined genetically. PDE-ALDH7A1 refers to PDE resulting specifically from variants in *ALDH7A1*. The primary focus for studies in this group is to characterise the genetic spectrum associated with *ALDH7A1* variants.

Two studies are reported. One reported an International study including 185 patients with clinical suspicion of PDE and at least one confirmed pathogenic variant in *ALDH7A1*.^31^ Participants were recruited from four clinical genetics laboratories that perform clinical testing of *ALDH7A1*, the International registry (which appears to cover North America, Europe, the Middle East and Australia) for PDE.^35^ and the pyridoxine dependent seizures patient registry.^36^ Specific details of the genetic tests used to identify genetic variants was not reported. The second study, based in China, included 33 participants from 31 families, 31 with PDE-ALDH7A1 and two with PLPB variants.^32^ Selection into the study appears to have been based on genetic testing results. A targeted PCR panel was used to sequence each exon (1 to 18) and exon-intron boundary of the *ALDH7A1* gene.

Biallelic variants in *ALDH7A1* were identified in 98% (182/185) of patients,^31^ resulting in 367 alleles with variants; three patients had only one variant allele identified (1.6%). The percentage of patients with compound heterozygous variants was 58.9% (109/185)^31^ and 84% (26/31)^32^ remaining patients were homozygous. Jiao et al. (2020) reported two (6.5%) patients as homozygous for *ALDH7A1*, however the supplementary table to the report clearly reports five patients as having the same genetic variant on both alleles indicating homozygosity (16% of total).^32^ The reason for this discrepancy is not clear.

Types of variants were reported slightly differently between the studies, however some similarities can be observed. In the registry-based study,^31^ 209 (57%) of 367 variantalleles were missense (compared to 65% (17/26) of variants identified in Jiao et al.^32^), 66 (18%) were splicing errors (compared to 12% (n=3) of variants^32^), 29 (8%) inDel (insertion and/or deletion of nucleotides), 29(8%) single nucleotide variant (SNV) terminations, 18 (5%) synonymous SNVs, and 15 (4%) CNVs. The remaining variants in Jiao et al. (2020) were nonsense (8%; 2/26) or deletions (15%; 4/26).^32^

Of the 26 different variants identified in Jiao et al. (2020), nine were recurrent within the study population (7/17 missense and 2/3 splicing site) and 17 (65%) occurred in only one patient each.^32^ The two most commonly identified variants were observed in 23% (7/31) of all PDE patients (a missense variant) and 19% (6/31) patients (a splicing site variant), respectively; remaining recurrent variants were observed in between two and four patients each.

The total number of recurrent variants identified in the study population was not reported by Coughlin et al. (2019), however four individual variants accounted for 38% (140/367) of all variant alleles, one of which was identified in a quarter of all alleles (94/367, 25.6%).^31^ Forty-nine missense variants accounted for the 209 alleles with missense variants, however the majority (65.3%; 32/49) of these were only identified in a single individual and 17 were recurrent (responsible for 177 of all variant alleles identified).

##### Summary of results

Using the broadest definition of a target population who might be suspected of having PDE, up to 6% could be confirmed as having *ALDH7A1* variants following genetic testing (based on a sample of 103 children with infantile onset (<12 months of age) pharmaco-resistant seizures.^29^) Restricting the population to those with a pyridoxine response, increased the percentage detected by WES to 11% (4/35), 75% (3/4) of whom also had elevated biochemical markers for PDE.^30^ Studies of patients with clinical and biochemical indicators of PDE have reported higher percentages with biallelic *ALDH7A1* variants of up to 86% (18/21).^37^

The International registry-based study of patients with clinical suspicion of PDE and at least one *ALDH7A1* variant allele^31^ provides the most comprehensive current picture of the genetic spectrum of patients with PDE, with data regarding Chinese patients with PDE reported in Jiao et al. (2020).^32^ The majority of patients in both studies had compound heterozygous *ALDH7A1* variants (58.9%, 109/185^31^ and 83.9%, 26/31^32^) compared to homozygous variants. Missense variants were the most commonly observed, accounting for 57% of variant alleles,^31^ and 65% of all variants.^32^ Although four individual (missense) variants accounted for 38% (140/367) of all variants identified in Coughlin et al. (2020),^31^ the majority of identified variants each occurred in a single individual. The occurrence of novel and often ‘private’ variants was a recurrent phenomenon across the included studies (17/26 variants occurred in a single individual in Jiao et al. (2020)^32^), with sequencing of one or both parents to confirm pathogenicity of identified variants commonly reported (e.g. in 2 of 6 children reported in Boonnsimma et al. (2023).^29^

##### Quality assessment

Of the four extracted studies, only one was considered to have met all quality assessment criteria.^29^ This was a single centre study with clearly defined population and period of recruitment, the sequencing methods were described in detail and cases and variants adequately ascertained. The study also broadly indicates the population of patients with unexplained infantile-onset seizures who may benefit from newborn WGS. We had several concerns about the other three studies extracted based on lack of information about the Selection of participants, level of detail provided concerning the participants,^32^ or testing methods used.^30, 31^ Two of the four studies were considered to report the genetic spectrum of PDE-*ALDH7A1* based only on sequencing techniques.^29, 32^ It is worth noting however that the majority of PDE presents in the first 4 weeks of life, such that the window of opportunity for WGS to benefit infants prior to developing seizures is small.

#### Heritable retinoblastoma

The three subgroups and studies for hRB were: any RB,^38^ sporadic RB^39^ and a combined category of familial, bilateral, or germline RB.^40^ Details per study are provided in Appendix 5, Table 21. All three studies were retrospective and conducted at a single-centre and all exclusively considered the RB1 gene. None of the selected studies exclusively used sequencing to identify variants.

##### Any RB

Retinoblastoma is a childhood onset cancer of the eye caused by biallelic variants in the *RB1* gene. RB can be either heritable (hRB) or somatic. hRB occurs where a variant on one of the alleles is present from conception (either inherited from a parent or occurring sporadically) and is therefore present in every cell of the body (germline) and the second variant occurs within the cells of the eye at some point after conception (this can occur prenatally or at any time point after birth). Around 40% of all RB is heritable. Somatic RB occurs where both variants occur within the cells of the eye, such that the variants can only be identified by genetic testing of tumour tissue as opposed to testing peripheral blood samples. Patients with bilateral RB are frequently assumed to have hRB, the majority of which occurs sporadically; only approximately 10% of all cases of RB have a known family history of the disease (familial RB). Studies that include ‘any RB’ provide the best estimate of the percentage of RB patients who would benefit from newborn WGS.

Salviat et al. (2020) included 1371 consecutive RB cases (including bilateral and unilateral, familial and non-familial) who successfully completed genetic counselling at a single centre.^38^ Multiple genetic screening methods were used dependent on the year of testing, but included various combinations of: denaturing high performance liquid chromatography (HPLC), quantitative multiplex PCR of short fluorescent fragments, methylation-restriction PCR, multiplex ligation-dependent probe amplification (MLPA), comparative genomic hybridization, Sanger sequencing and NGS. The promoter region and all exons with their flanking intronic sequences were screened. The identified pathogenic variants were classified as germline (identifiable in blood and therefore present from the point of conception) or somatic (occurring at any point following conception, and therefore only identifiable in tumour tissue) and then as associated with presence or absence (complete loss) of RB protein. Where tumour tissue was available, this was screened first to identify the *RB1* variants, with peripheral blood then tested for the identified variants to determine the germline (heritable) status (n=293). The remaining patients with no tumour tissue available underwent germline screening (of peripheral blood) only.

Of the original 1404 eligible participants, 118 (8.4%) had a known family history of RB (defined as families with at least two germline carriers of a *RB1* pathogenic variant). Of those who completed the study, 44.2% (606/1371) were found to have a germline *RB1* variant (hRB), including 497 with bilateral RB and 109 with unilateral RB. Germline mosaicism was identified in 28 of 606 (4.6%). Of the 765 patients with no germline variant identified, 248 (32.4%) were identified as having somatic RB (bi-allelic variants in tumour tissue only) and 517 (67.6%) were negative on germline screening (no tumour tissue available). Of those with germline variants identified, the majority (561/606, 92.6%) were identified by the germline screening strategy and 7.4% (n=45) were identified through the first-line tumour screening strategy. The most common variants (comprising 77.2% of identified germline variants) were nonsense (222, 36.6%), frameshift (140, 23.0%) or out-of-frame splice variants (110, 18.2%), all of which are associated with a loss of RB protein (total of 537/606 identified germline variants were associated with loss of RB protein).

Amongst those with hRB (germline variants) (n=606), the incidence of bilateral RB was higher in patients with variants associated with a loss of RB protein (84.2%; 452/537) compared to those with variants with no loss of RB protein (65.2%; 45/69) (P=0.01). Germline variants associated with a loss of RB protein were also associated with earlier mean age at diagnosis of RB (P<0.001), and later stage at diagnosis (P=0.047) compared to variants not associated with complete loss of RB protein.

##### Sporadic RB

Sporadic RB occurs where there is no family history of disease and can be either heritable (the variant is present in the germline and can therefore be passed on) or somatic (occurring only in the tumour). Children who develop sporadic germline RB (hRB) are the population with the greatest potential to benefit from newborn screening with WGS, as regular intensive surveillance can be initiated to allow earlier detection and treatment. This disease definition identifies the additional patients who might undergo surveillance for RB as a result of WGS. It is worth noting however that WGS is still of benefit to those with a known family history of RB because genetic testing will identify those who do not need to undergo such intensive surveillance because the *RB1* variant has not been passed on.

The study by Temming et al. (2013) included 195 patients who presented with unilateral sporadic RB and who underwent genetic testing at the request of patients or their legal guardians.^39^ Only those with ophthalmological follow-up until age 5 were eligible for inclusion. One or more of the following methods was used for genetic testing of blood or tumour tissue: analysis of allele loss in tumours, cytogenetic analysis, denaturing HPLC, exon-by-exon sequencing, MLPA, methylation-sensitive PCR, quantitative fluorescent multiplex PCR, quantitative real-time PCR, real-time PCR and single strand conformation polymorphism (SSCP). Forty (20.5%) patients were identified as having a germline *RB1* variant (hRB), 29 (72.5%) of whom had a heterozygous *RB1* variant and 11 (27.5%) had germline mosaicism (which can be passed on to offspring of the proband if it is found to be present in the particular germ cell that forms the embryo).^41^ Of those with heterozygous germline variants (hRB), 10 (34%) were classified as whole gene deletions, 13 (45%) as premature terminations and 6 (21%) as ‘mild’ variants.

Of the 195, nine (4.6%) developed bilateral RB during five-year follow-up, eight of whom had a heterozygous RB1 variant (3 with whole gene deletions and 5 with premature terminations) and one had germline mosaicism.

##### Heritable / Bilateral RB

Heritable RB includes cases who have a family history of RB (familial or inherited germline variant) and those whose germline variant in *RB1* occurs sporadically at the time of conception (sporadic germline variant). Most patients with bilateral RB have hRB such that studies frequently defined study eligibility as familial or bilateral RB as a proxy for identifying cases of germline hRB. Consideration of studies in this category provides an indication of the most commonly found types of variant, however in many studies multiple genetic tests are used to characterise the genetic spectrum and it is not always possible to identify those variants that would be most easily identified on WGS.

Hulsenbeck et al. (2021) included 287 cases with RB from a total population of 815 patients (342, 42% of whom were identified as having a germline variant).^40^ Patients with confirmed heterozygous pathogenic constitutional *RB1* variants (hRB) who had not previously undergone ophthalmological screening for familial RB were included in the study report. Those with mosaicism and non *RB1* variants were excluded. Multiple genetic screening methods were used, some of which included sequencing or sequential testing. A retinoblastoma variant effect class (REC) was developed to classify the identified pathogenic variants according to their effect on RB protein structure and quantity (i.e. extent of loss in RB protein), from REC-I (largest effect) to REC-V (smallest effect).

The most common variants (comprising 98.6% of identified variants) were nonsense or frameshift variants (REC-I) (199, 69.3%), whole *RB1* gene deletions (REC-II) (39, 13.6%) and missense or in-frame SNVs (REC-III) (45, 15.7%). Of those with whole *RB1* gene deletions (n=39), 27 (69%) were identified as also having deletion of the *MED4* gene (hypothesised as being associated with lower penetrance of RB.^42^

The percentage of bilateral RB was highest in those with nonsense or frameshift variants (REC-I) (186, 93.5%) compared to those with REC-II (30, 76.9%) and REC-III (36, 80.0%). Age at diagnosis was lowest in patients with REC-I variants (median: 7.3 months [range: 0.2–48.0]), followed by REC-II (10.3 months, [0.4–40.9]) and REC-III (11.6 months [0.9–45.1]) variants.

##### Summary of results across disease definitions

To summarise, the percentage of RB cases with an identified germline *RB1* variant(i.e. hRB) using various testing strategies varied from 20.5%^39^ for the most narrowly defined population (sporadic unilateral RB) to 44.2%^38^ for the most broadly defined population (any RB). Both of these studies included participants with germline mosaicism (4.6%^38^ and 27.5%^39^ of germline cases) which is not always detectable in peripheral blood. Mosaicism is where a percentage of cells in the body carry the variant allele, but others carry a normal copy. Sequencing is often done on DNA extracted from peripheral blood samples, which means patients with mosaicism (especially low-level mosaicism) may not be detected by WGS strategies if the DNA extracted from the blood does not carry the variant allele.^43^ Specific techniques, such as use of unique molecular identifiers and next-Generation sequencing are of interest for detecting mosaicism.^44^ Results from the largest, most inclusive study^38^ demonstrate that without genetic testing, as few as 10% of RB cases might have been identified for ophthalmologic surveillance from birth, based on known family history of the condition. Salviat et al. (2020) did however employ a stringent definition of familial RB.^38^

Salviat et al. (2020) further demonstrated that as much as 38% of the total population (n=517) had no RB variant identified (negative on germline screening and no tumour tissue available for testing).^38^ While it is likely that the majority of the 517 patients had somatic RB, a small proportion may have germline RB that was not detected by the genetic testing strategy. In Hulsenbeck et al. (2021), for example, three of the 821 patients at the centre were excluded as genetic data revealed no *RB1* variant and high *MYCN* (a different gene) amplification suggesting a different genetic pathway to development of RB which may not be identified using WGS.^40^

In two studies that reported the variant types, the most common were nonsense and frameshift variants.^38, 40^ The percentage of those with bilateral RB was reported as higher in those with nonsense variants in both of these studies.^38, 40^

##### Quality assessment

Quality assessment raised similar concerns about all three extracted studies for hRB.^38–40^ Insufficient detail about the genetic testing methods used led to concerns about ascertainment of variants by sequencing and about replication or application of results beyond the study. It was not possible to determine the applicability of study results to the review question, as techniques other than sequencing may have been used to determine the genetic status of the patients.

#### X-linked hypophosphataemic rickets

XLHR is one form of hereditary rickets (Supplement 1). Clinical symptoms of rickets are not specific to XLHR but are characteristic for a much broader range of conditions. Some conditions can be distinguished based on biochemical markers while others require information on the inheritance pattern or genetic testing. The proportion of rickets patients identified by sequencing the *PHEX* gene will therefore depend on whether disease is defined clinically, biochemically or genetically. The findings of studies on *PHEX* frequency in children with disease defined in four different ways are summarised in Appendix 5, Table 22.

Overall, six studies were selected across four different categories of hereditary rickets. One study was identified and described for the broad hereditary rickets category,^45^ and two studies were included for each of hypophosphatemic rickets^46, 47^ and hereditary hypophosphatemic rickets.^48, 49^ The largest study in these two categories only tested for the *PHEX* gene, while the second largest study tested for additional genes, thus providing information about the *PHEX*-negative cases which could either be undetected *PHEX* cases or caused by different genes. One study was selected for the XLHR category.^50^

Four studies were conducted at a single centre, ^45–48^ while one study included patients from all paediatric hospitals in Norway^49^ and one study was an International registry study.^50^ Three studies were prospective^45–47^ and three were retrospective.^48–50^ Sequencing techniques were employed to identify variants in the three prospective studies, while both sequencing and MLPA were used in two retrospective studies. The registry study of XLHR patients did not specify the genetic test(s) used.^50^

##### Hereditary rickets

This category is an umbrella term of any type of hereditary rickets (excluding nutritional rickets). It consists of two main types 1) vitamin-D-dependent rickets (low phosphate levels secondary to vitamin D deficiency) and 2) hypophosphatemic rickets (low phosphate is primary defect and rickets is therefore vitamin-D-resistant). Each type consists of several conditions caused by a series of genes with similar clinical symptoms. Some have distinct biochemical markers.

Jacob et al. (2023) conducted a comprehensive assessment to identify the genotypic spectrum of rickets in 10 Indian families with 10 patients suspected of having hereditary rickets.^45^ All 10 patients had symptom onset in childhood, however, two patients did not receive a diagnosis until early adulthood. Exome sequencing identified variants in six different genes including three patients (3/10, 33%) with a *PHEX* variant. The results revealed three known truncating variants, c1482+5G>C, c1586_1586+1del and c.58C>T. The c1586_1586+1del variant resulted in a more severe phenotype compared to the other two variants. Other implicated genes included *CYP27B1* (n = 3 patients), *CYP2R1*, *VDR*, *SLC34A3* and *SLC2A2* (all one patient each). No cases had an unidentified genetic cause.

##### Hypophosphatemic rickets

In hypophosphatemic rickets, low serum phosphate due to renal losses is the primary defect, which is either mediated by the hormone fibroblast growth factor 23 (FGF-23) (raised FGF-23 levels) or is independent of FGF-23 (normal FGF-23 levels). Measuring levels of FGF-23 can aid the distinction between the two types. There are 15 genetically distinct disorders which are grouped into FGF-23-dependent and FGF-23-independent hypophosphatemic rickets.^51^

Gaucher et al. (2009) analysed the *PHEX* gene in 118 families, including 56 familial and 62 sporadic cases using classical sequencing. Sequencing covered all 22 exons, intronic regions and the region at the 3 prime end which is not translated into a protein but serves regulatory processes.^47^ The study was conducted at a single centre but encompassed a multi-ethnic population, comprising individuals of European, North African, Caribbean, and Asian backgrounds. The inclusion criteria were based on low serum phosphate and TmP/GFR levels, bone deformities and radiological evidence of rickets. *PHEX* variants were found in 78% (93/118) of probands. The 93 variants comprised 78 different variants of which 60 (77%) were novel. Variant types included nonsense (28%), frameshift (30%), splice site variant (23%), and missense variant (19%).

Some uncertainty regarding the pathogenicity of novel *PHEX* variants was noted. One patient with a novel c.1206A>G variant (a single nucleotide replacement) also exhibited a second missense variant. In two other cases, both patients with the c.505G>A variant (another single nucleotide replacement) harboured additional variants, one involving an insertion leading to frameshift and the other a deletion leading to frameshift. The findings emphasise the significant role of *PHEX* in X-linked dominant hypophosphatemic rickets and suggest that family members should be screened when a *PHEX* variant is found in a sporadic case. Additionally, when a missense variant is detected, a search for another *PHEX* variant should be conducted.

The study’s approach to sequence only the *PHEX* gene meant that a large proportion of *PHEX* negative cases remained unexplained. For 3/25 negative cases a reason was proposed. Missing PCR samples for two exons in one case let to the conclusion that one patient had a large deletion. One patient was subsequently diagnosed with a different type of hypophosphatemic rickets, and one patient was diagnosed with tumour-induced osteomalacia with secondary hypophosphatemia.

Marik et al. (2022) screened 66 consecutive Indian patients with refractory hypophosphatemic rickets, using whole exome sequencing (WES).^46^ Patients were characterised by a lack of healing despite treatment with cholecalciferol and had lower-than-normal phosphate levels for their age. The mean age of onset of symptoms was 22.5±14.3 months. 24/66 (26.4%) patients had a confirmed *PHEX* variant and 40/66 (60.6%) patients had a confirmed variant in a different gene. The remaining two cases had a variant of unknown significance which were classified as negative and for whom genetic testing could not confirm the diagnosis. All 24 *PHEX* variants were different and 13/24 were novel.

In the context of expressivity, one patient carried two *PHEX* variations (c.2048T > A; p.(Leu683His) and c.2071-1G > C), whereas her mother, who was clinically mildly affected, had only one *PHEX* variation (c.2048T > A; p.(Leu683His)). This difference may explain the variability in disease severity between them.

##### Hereditary hypophosphatemic rickets

Hereditary hypophosphatemic rickets is marked by increased FGF23 activity leading to hypophosphatemia due to renal phosphate wasting. Genes associated with hereditary hypophosphatemic rickets include *PHEX, FGF23, ENPP1, DMP1*, and *FAM20C* which are clinically and biochemically similar but follow different inheritance patterns.

del Pino et al. (2022) included 96 patients diagnosed with hereditary hypophosphatemic rickets, of whom 42 underwent molecular testing of *PHEX* by Sanger sequencing and MLPA to detect gene deletions and duplications.^48^ The condition was characterised by the typical presence of combination of clinical, laboratory, and radiographic findings. Deleterious sequence alterations or large deletions in the *PHEX* gene were identified in 85.7% (36/42) of patients. The remaining six patients were not genetically confirmed.

Rafaelesen et al. (2016) investigated 28 Norwegian children with hereditary hypophosphatemic rickets from 19 families.^49^ Inclusion was based on serum phosphate levels below the age dependent reference range combined with tubular reabsorption rate of phosphate (not due to hyperparathyroidism). Sanger sequencing and MLPA analysis (to look for deletions and insertions in sequencing negative patients) of the *PHEX* gene was conducted. This was followed by Sanger sequencing of *FGF23, DMP1, ENPP1KL, and FAM20C* successively in *PHEX* negative patients. Overall, 13/19 (68.4%) probands were identified with *PHEX* variants. The 13 variants were all different and none were novel. There was one variant each in *FAM20C* and *SLC34A3*. 4/19 (21.1%) had no confirmed variant. Exploration of the effect of different types of variants (missense versus nonsense) revealed no difference in clinical outcomes.

##### X-linked hypophosphatemic rickets (XLHR)

X-linked hypophosphatemic rickets is the most common form of hereditary hypophosphatemic rickets. It is caused by variants in the *PHEX* gene which is inherited in an X-linked dominant fashion. Genetic testing or knowledge of a family history with a typical X-linked dominant inheritance pattern can support the diagnosis of XLHR.

Ariceta et al. (2023) is a registry-based study including multinational data of 579 participants with XLHR.^50^ XLHR diagnosis was based on clinical judgement of an XLH-treating expert physician (FH, clinical, radiological and biochemical findings), and /or by genetic testing. However, genetic testing was not required for patient registration. 282 of children underwent genetic testing (not further described). A total of 89.7% (253/282) children tested were identified with variants in the *PHEX* gene. There was a small number of patients with different genetic disease incorrectly registered including four with a variant in *FGF23*, one on *SLC34A3* and seven where the gene was not specified. 17/282 (6.0%) had no variant confirmed. The study only reported genetic findings at the gene level. While symptoms were not reported by *PHEX* variants, the study concluded overall that children with XLHR diagnosis despite early detection and treatment did not do too well.

##### Summary of results

The range of genes identified varied across the different categories of rickets. The frequency of *PHEX* varied from 33% in the most broadly defined population^45^ to 89.7% in the most narrowly defined population.^50^ ‘Pre-screening’ of the population using biochemical testing resulted in greater proportion of patients with *PHEX* in both the hypophosphatemic rickets category^47^ and the hereditary hypophosphatemic rickets category.^48^ The extent of test negatives (no confirmed pathogenic variant identified) varied from 3%^48^ to 21.1%^49^ which was largely due to the testing strategy used (number of genes considered, extent of sequencing, sequencing method (e.g. WGS versus Sanger sequencing), additional testing). One study sequenced the untranslated 3-prime region as well as exons and intronic regions in recognition that XLHR may be caused by variants in the regulatory region of the mRNA.^49^ MLPA was used in two studies to overcome limitations of sequencing to detect deletions and duplications.^48, 49^ Neither study reported the proportion of variants detected by MLPA in addition to those detected by sequencing. XLHR is characterised by many different variants precluding investigation of expressivity of specific variants, with one study reporting that there is no clear genotype-phenotype link.^49^ Furthermore, a great proportion of variants in each study was novel which presents challenges for the confirmation of variant pathogenicity.

#### Quality assessment

The quality assessment using the Murad tool raised concerns across the different dimensions assessed. The Selection of patients in both the Gaucher et al. (2009) and Jacob et al. (2022) is unclear whether the included participants were representative of all eligible patients.^45, 47^ The ascertainment of patients in Rafaelsen et al. (2016) is unclear.^49^ Except for Marik et al. (2022)all studies did not describe the cases in sufficient detail to enable other investigators to replicate the research.^45–50^

In three studies concerns regarding the applicability of study findings to WGS was low because the reported genetic spectrum was based on sequencing that more closely resembles WGS in the screening application,^45–47^ while concerns were high in the two studies that used MLPA in addition to sequencing^48, 49^ and unclear in one study where ‘genetic testing’ was not further specified.^50^

#### Familial hemophagocytic lymphohistiocytosis

Haemophagocytic lymphotistiocytosis is not a single disease but a syndrome that is associated with several heritable and non-heritable conditions. Symptoms and biochemical markers are, therefore, nonspecific and do not aid in the differential diagnosis, and a population of patients with fHLH is difficult to define based on clinical and biochemical characteristics. Historically, HLH was divided into primary (early onset, genetic condition) and secondary HLH (later onset, secondary to underlying medical condition such as cancers, infections or autoimmune disorders) using the main underlying trigger of symptomatic disease to define subgroups. More recently the boundary between primary and secondary HLH has blurred with a better understanding of the complexity of the syndrome (discovery of new genes involved, differences in severity, genetic involvement in secondary HLH, detection of digenic disease). It is more accepted now that primary HLH is an artificial and ill-defined category which was reflected in the published studies and could not be adopted here. fHLH is used to describe a subset of primary HLH disorders caused by bi-allelic variants in the four genes *PRF1, UNC13D, STX11 and STXBP2*. distinguishing between the four conditions is not feasible using clinical symptoms or biochemical markers and relies on genetic testing. The three categories and representative studies were, therefore, haemophagocytic lymphotistiocytosis (HLH) more broadly^52^, fHLH encompassing the four genes *PRF1, UNC13D, STX11* and *STXBP2*^53^ and any one of the four conditions, of which we identified studies for three of the four genes.^54–56^ The findings of the studies are summarised in Appendix 5, Table 23.TmP/GFR – ratio of tubular maximum reabsorption rate of phosphate to glomerular filtration rate; WES – whole exome sequencing; MLPA – multiplex ligation-dependent probe amplification; FH – family history, NR – not reported

Four of the studies were retrospective and multi-centre studies,^53^ ^52, 55, 56^ while Amirifar et al. (2021) was a systematic review.^54^ The test was well defined in one of the five studies which specified different sequencing methods (Sanger sequencing, NGS, WES) for patients with different indications based on biochemical assays.^53^

##### Haemophagocytic lymphotistiocytosis (HLH)

Haemophagocytic lymphotistiocytosis (HLH) is a hyperinflammatory syndrome generally defined by the diagnostic criteria recommended by the Histiocyte Society including symptoms of fever, splenomegaly, cytopenia, elevated cytokines and hemophagocytosis.^57^

Cetica et al. (2016) is an Italian registry-based study which analysed 500 HLH patients over 25 years.^52^ The multi-ethnic study included patients of southern European, Eastern European, African, Asian, and Hispanic origin. In 426 patients who underwent sequencing, a genetic diagnosis was possible in 171/426 (40.1%) of patients, while 43/426 (10.1%) had monoallelic disease and 197/426 (46.2%) were believed to have non-genetic disease. 15/426 (3.5%) cases with assumed genetic disease were missed. Of 171 patients with a genetic diagnosis, 141 (82.5%) had fHLH with *PRF1* and *UNC13D* variants accounting for 131/141 (92.9%) of fHLH cases. The 69 patients with *PRF1* and the 62 patients with *UNC13D* biallelic disease carried 34 and 37 different variants, respectively. A small number of variants were common occurring in up to 19 patients. The data on disease onset in sibling pairs revealed that there is no clear genotype-phenotype relationship. In 9/26 sibling pairs disease onset varied up to 17 years while in one pair, one sibling developed the disease at 6.7 years while the other remained unaffected at 25 years. The number of patients with monoallelic disease indicates that HLH likely results from both genetic predisposition and exogenous triggers.

##### fHLH

Familial HLH is an artificial category of four conditions caused by variations in four distinct genes. Patients with different fHLH subtypes are clinically and biochemically similar.

Shabrish et al. (2021) investigated 101 molecularly confirmed fHLH patients of which 98 were under the age of 18 years over 10 years from 20 referral centres in India.^53^ 86/98 (87.8%) had biallelic disease. 12/98 (12.2%) patients had monoallelic disease and would be considered test negative on sequencing. *PRF1* and *UNC13D* variants accounted for 70/86 (81.4%) of cases with biallelic disease. Molecular analysis revealed that missense variations were the most common type of variation in all four genes. The number of different variants was significant (25 different variants in 34 patients with *PRF1* and 28 different variants in 23 patients with *UNC13D* fHLH). Patients with homozygous variants across all four genes had an earlier disease onset (median 10 months) compared to those with compound heterozygous variants (median 3 years).

##### Single gene/variant

The narrowest category consists of patient populations that had confirmed fHLH caused by one specific gene while patients with confirmed variants in other genes were excluded. The studies’ aim was to characterise the variant spectrum in this tightly defined patient population.

Trizzino et al. (2008) included 124 patients with confirmed biallelic *PRF1* disease and a median age of disease onset of three months from six different International centres.^56^ They detected 63 different variants: 11 nonsense, 10 frameshift, 38 missense, and 4 in-frame deletions. 15/63 variants were novel. The most common single variant was a missense variant in 32 patients. Specific *PRF1* variants were strongly linked to Turkish, African American, and Japanese ethnic groups. Patients with two disruptive variants had a younger age at onset than patients with missense variants only.

The systematic review by Amirifar et al. (2021) analysed clinical features, immunologic data, and genetic findings from 57 articles covering 322 patients with *UNC13D* variant with a median age of onset of six months.^54^ 269/322 (83.5%) had biallelic disease, 50/322 (15.5%) had monoallelic disease and for three patients this information was not reported. Missense variations were the most common type of variation. Severe features appeared to be associated with a homozygous genotype and missense variants. Splice-site errors and compound heterozygosity were more prevalent in patients with mild features.

Pagel et al. (2012), a multinational study with patients included mainly from Germany and Turkey, included 37 patients with confirmed biallelic *STXBP2* variants.^55^ One of the 37 patients was 19 years of age at the time of diagnosis. Nine novel variants were reported. Variants included nine different missense variants, four different splice-site variants and several small deletions or insertions. Three variants were seen in more than five patients. 13/37 patients carried one of two splice-site variants affecting exon 15. The exon 15 splice-site variant was associated with mild disease and an atypical disease course. These patients often experienced chronic, recurrent episodes with long periods without HLH symptoms, and their reactivations typically responded to steroids-only treatment or underwent spontaneous remission.

##### Summary of results

In patients with HLH symptoms that met the HLH diagnostic criteria of the Histiocyte Society, a genetic diagnosis of fHLH was established in one third of the patients.^52^ This means, a third of clinically defined HLH patients could be detected by sequencing the four fHLH genes. 7% of patients would be missed because they carried variants in different genes, 3.5% would be missed because the genetic cause could not be identified, 10% would be missed because they had monoallelic disease and 46% would be missed because they had non-genetic disease. The number of patients with monoallelic disease identified in 3/5 studies led to the suggestion in one study that there is a gene-dosage effect which means that fHLH can no longer be regarded as a simple recessive disease.^52^ This needs to be considered in the interpretation of sequencing results and is further complicated by the occurrence of digenic disease (monoallelic variants in two of the four genes). However, the risk of disease in patients with monoallelic disease is currently unknown. Across all four conditions, there is some indication that homozygosity is associated with earlier onset and more severe disease^53^ and severity is linked to the type of variant^54^ but that there is no clear genotype-phenotype link as siblings with the same genotype displayed different phenotypes and different age of disease onset.^52^ Overall, the studies suggest that there are a few common variants that are linked to particular ethnicities highlighting the need to understand the genetic disease in a broad spectrum of patients that is applicable to the screening setting before considering the implementation of sequencing as a screening tool in a diverse population.

##### Quality assessment

The quality assessment of Amirifar et al. (2021)^54^ was conducted using the ROBIS-2 tool for systematic reviews,^18^ while other studies were evaluated using a modified Murad tool. We observed that four studies lacked clear or sufficient information on patient selection.^52, 53, 55, 56^ These studies provided limited details regarding the inclusion of patients from different centres and did not report the time frames of recruitment. Amirifar et al. (2021) presented an unclear risk of bias due to insufficient details on the review’s conduct, study selection, and data synthesis.^54^

#### Medium Chain Acyl-CoA Dehydrogenase Deficiency

MCADD is part of a group of conditions called inborn errors of metabolism. MCADD symptoms overlap with those of multiple inborn errors of metabolism. Differential diagnosis can be achieved by determining the disease specific biomarker profile, relevant enzyme activity levels and the underlying genetic variant. Because MCADD is on the current NBS screening panel, MCADD can be defined based on screening outcomes as positive on Initial MS/MS screening test and positive on confirmatory serum and urine tests.

The five disease definition categories for MCADD (from broadest to narrowest) and representative studies were: Inborn Errors of Immunity/Inherited Metabolic Disease (IEM/IMD),^58^ FAOD,^59^ Acyl CoA Dehydrogenase (ACAD) Deficiencies,^60^ clinical-biochemical MCADD^61, 62^ and gene (*ACADM*) positive MCADD.^63^ We included two studies for the clinical-biochemical MCADD category; one study that detected cases through NBS screening (genetic spectrum in MS/MS positives and in follow-up positives)^61^ and one that diagnosed patients clinically or biochemically following family screening.^62^

Details per study for each of the five disease definitions considered are provided in Appendix 5, Table 24. The genetic testing method was not reported in two studies,^58, 62^ three used a sequencing technique^59, 60, 63^ and one used sequential testing.^61^ All six studies were conducted retrospectively.

##### IEM/IMD

Inborn errors of metabolism (IEM), also known as inherited metabolic disorders (IMD), includes a group of approximately 600 conditions that are individually rare but collectively common. They are hard to diagnose given the nonspecific symptoms that many affected patients experience.^64^ IEM can be caused by variants in different genes that affect the same metabolic pathway at different stages resulting in different conditions with similar symptoms. Overall, IEM are very heterogeneous resulting in groups of disorders affecting different metabolic pathways with different epidemiology, presentation, and heritability.

Martin-Rivada et al. (2022)^58^ reported details for 224 Spanish newborns who underwent genetic testing following biochemical indication of an IEM as part of the national screening programme. The original cohort included 902 consecutive newborns with an Initial abnormal NBS test result. The molecular genetic testing method was not reported; however, 30 different genes were included. Of the 224 babies, 222 (99.1%) were diagnosed with a genetically confirmed IEM; two participants were considered to have biochemical hyperphenylalaninemia (no variants identified). In the wider group of Initial NBS positive babies, this equates to 24.6 % (222/902) with genetically confirmed IEM. Of those with a genetic variant, 19.3% (43/ 222) were identified with a variant in the *ACADM* gene. Amongst these 43 children, 14 different genotypes were identified. The 985A>G variant made up 70% of all alleles (60/86) in 43 newborns; 22 newborns were homozygous and 16 compound heterozygous for this variant. The remaining five cases were compound heterozygous for other variants. Only one patient (homozygous for 985A>G) showed symptoms of MCADD before newborn screening results were available.

##### FAOD

Fatty acid oxidation disorders (FAOD) are a particular group of IEM caused by variants of the genes associated with the metabolic pathway of fatty acids in the mitochondria. Symptoms overlap and different FAOD are identified by their specific acylcarnitine (fatty acid metabolites) profiles and can be confirmed by gene sequencing.

Maguolo et al. (2020) included 30 Italian patients with FAOD; 20 were infants diagnosed following NBS screening and 10 were clinically diagnosed with mean age at onset of 29 years and therefore excluded from review.^59^ Five of the 20 infants were biochemically positive for MCADD. Sequencing was conducted using a custom designed FAOD panel including 15 genes. Biallelic MCADD was identified in 15% of newborns (3/20) or 60% (3/5) infants with biochemical MCADD; while two infants carried monoallelic ACADM variants and would be classed as negative on genetic testing. Fifteen patients (75%) had different FAOD subtypes (5/15 with genetically confirmed disease, 1/15 with monoallelic disease and 10/15 without genetic information). Of six *ACADM* variants in those with biallelic disease, five were different. One infant was homozygous for the 985A>G variant and two infants were compound heterozygous carrying a total of four different variants. All three infants with biallelic *ACADM* variants had a residual MCAD enzyme activity of less than 5% associated with severe disease.

##### Acyl-CoA dehydrogenase deficiencies

The FAOD subgroup of ACAD deficiencies are caused by variants in 11 genes with *ACADM* being one of them.

Wang et al. (2019) reported genetic testing results for 20 newborns with ACAD confirmed on diagnostic biochemical testing from a cohort of 83 newborns with an Initial positive screening result for ACAD deficiency on NBS screening.^60^ Out of the 20 newborns, four had biochemically confirmed MCADD. High throughput sequencing and Sanger sequencing was used with a wider IEM panel of 306 genes. Three of the 20 (15%) newborns with confirmed ACAD were identified with biallelic compound heterozygous variants in the *ACADM* gene. In the wider group of babies with an Initial positive screening test, this would be 3.6% (3/83). One newborn was test negative on sequencing due to a monoallelic variant. Therefore, genetic testing identified 3/4 patients with biochemical MCADD. The three biallelic MCADD patients carried five different variants of which two were novel. None of the Chinese newborns carried the 985A>G variant common in European cohorts.

##### Clinical-biochemical MCADD

This disease definition includes those with a diagnosis of MCADD either confirmed by clinical characteristics or by biochemical testing. Ideally, we would have considered the two categories separately to investigate the impact of different disease definitions on the genetic spectrum of disease. However, studies tended to include a mix of patients or poorly defined the populations included.

We included two studies for this disease definition category, one of which included patients identified with MCADD through an NBS programme.^61^ The other study in this subgroup included children who were clinically diagnosed with MCADD or detected through family screening and presumably confirmed biochemically.^62^

Nichols et al. (2008) included 511 newborns with NBS octanoylcarnitine (C8) levels ≥0.3 µmol/L who were subsequently referred for molecular genetic testing using a sequential sequencing method.^61^

First tier testing specifically aimed to identify two of the most common variants prevalent in the US c.985A>G and c.199T>C, second tier testing used full *ACADM* sequencing in those with at least one variant or those without variant detected on first tier testing but repeat C8 levels of at least 0.4 µmol/L. Mesbah et al. (2020) included 17 children younger than 18 with clinically diagnosed MCADD, four were diagnosed via family screening and two postmortem.^62^ The genetic testing method was not reported. Both studies solely considered the *ACADM* gene.^61, 62^

The percentage of patients positive (biallelic *ACADM* variants) on first tier screening for two variants initially was 1.6% (8/511 with MS/MS positive screening test) in Nichols et al (2008).^61^ A further 157/511 (30.7%) of newborns were monoallelic and 83/511 (16.2%) had neither variant on first tier testing. In 20 newborns with positive clinical follow-up, sequencing of the full *ACADM* gene revealed 17 (85%) with biallelic *ACADM* variants and three with monoallelic variants classified as test negative. Mesbah et al. (2020) reported genetic test results of 14 of 17 included patients, 11 of whom had biallelic *ACADM* variants and three cases were missed by sequencing as only one variant was identified.^62^ The most common variant in these two studies from the US and Ireland was 985A>G (12/17 and 11/11^61^ either homozygote or compound heterozygote, respectively)^61, 62^. Nichols et al. (2008) reported 13 different variants of which five were novel.^61^ They reported that the c.199Y>C/c.134A>G genotype resulted in ‘mild’ MCADD.

##### Gene (ACADM) positive MCADD

This disease definition includes those with confirmed *ACADM* variants and provides useful information on variant frequency.

The study by Touw et al. (2012) included 68 children from the Dutch newborn screening programme with confirmed variants in the *ACADM* gene by sequencing all exons and adjacent intron regions.^63^ Seventeen different genotypes were reported of which seven were novel. Most of the children (42/68, 61.8%) were homozygous for the 985A>G variant; a further twenty were compound heterozygous including the 985A>G variant. The authors categorised genotypes into ‘classic’ (previously recognised in clinically confirmed cases, n=53) and ‘variant’ (genotypes not previously recognised in clinically confirmed cases, n=15), and reported median residual MCAD enzyme activity of 0% for the former and 25% for the latter group. This may support the theory that screening identifies disease with milder MCADD due to genotypes not recognised in clinical cases.

##### Summary of results across disease definitions

Five of the six studies described above reported the experience of using genetic confirmatory testing in national newborn screening programmes. The percentage of patients with a genetic MCADD diagnosis varied from 15%^58, 59^ to 19%^60^ in populations with a broad disease definition; and 85% in a population of patients with biochemically confirmed MCADD.^13^ As biochemical testing is readily available for MCADD, the latter category is the most relevant to consider. Genetically confirmed disease was less common in those with only an Initial positive NBS screening test for MCADD compared to those confirmed on second-tier or follow-up biochemical testing.^60^ Testing for only two common *ACADM* variants had a low yield in initially NBS test MCADD positive babies (8/511 (1.6%) biallelic and 157/511 (30.7%) monoallelic)^61^, however, this strategy could not be fully evaluated because results were not reported for those with biochemical MCADD confirmed on follow-up testing.

Results from the study which included patients with a clinical diagnosis of MCADD suggest that 18.0% (almost one in five) of MCADD cases cannot be detected through genetic screening.^65^ However, it is important to note that the sample size was small (with only 17 cases) and the genetic testing method used was not reported, so it is not clear how applicable these results are to WGS.

Of the studies that presented variant frequency, the 985A>G variant was found to be the most common.^58, 61–63^ In the study restricted to genetically confirmed cases of MCADD, 61.8% were homozygous with this variant.^63^ However, all four studies were from countries with populations of mainly Caucasian origin. The only study from Asia did not report this variant in any of their three genetic MCADD cases. *ACADM* variants present little heterogeneity in the studies (17 genotypes in 68 genetically confirmed MCADD cases), however, this may be misleading as ethnicity appears to affect the genotypes detected.^63^

Little information regarding expressivity was presented in the included studies. Generally, the evidence points towards some variants causing ‘milder’ disease based on residual enzyme activity studies and it appears that those were newly detected in the screening context.^63^

##### Quality assessment

Three of the studies met all the quality assessment criteria.^59–61^ We had concerns regarding the Selection criteria for sequencing in one study,^63^ and we felt that reporting was inadequate in the study by Mesbah et al. (2020).^62^ Applicability to the review question was unclear in two study, because the testing method was not reported.^58, 62^

##### Evidence on early vs late treatment

Question 4: What is the evidence on early (following screen detection or sibling detection (cascade testing)) versus late (following clinical presentation) treatment?

In the absence of RCTs investigating the outcomes of pre-symptomatic vs symptomatic treatment, we defined early vs late treatment for the five selected conditions based on the natural history of the disease (e.g. age of symptom onset, progressive vs relapsing / remitting conditions) and the type of available treatment/management (e.g. preventative vs symptom management). Table 6 summarises the natural history for the five conditions that informed our definitions of early vs late. Table 7 summarises the management strategies for the five conditions and Table 8 our definitions of early vs late based on the available treatment studies. The process illustrates that the different condition-treatment pairs require individual definitions of early vs late.

**Table 9.** Overview of the natural history of the 5 conditions.

| Condition | Natural history/characteristics of condition |
| --- | --- |
| PDE | Progressive<br>Deficiency of the enzyme alpha aminoadipic semialdehyde (α-AASA) dehydrogenase which is involved in the breakdown of lysine in the brain results in the accumulation of metabolites which inactivate pyridoxine. Pyridoxine depletion causes intractable neonatal seizures which become recurrent and prolonged if left untreated (with pyridoxine). Uncontrolled seizures can ultimately lead to death, but this appears to be less common. Classic PDE usually presents during the neonatal period (i.e. within 28 days of birth) with prolonged seizures that are difficult to control with anti-seizure medication; in 75% of cases seizures may occur within the first few hours of life. These seizures last several minutes and involve loss of consciousness, spasticity and |
|  | convulsions. If untreated, periods of encephalopathy are common (irritability, crying, fluctuating tone, poor feeding). In some cases (up to 30%), affected individuals do not experience seizures until they are 1 to 3 years old (late-onset PDE). Intellectual disability and developmental delay are often present (around 75% of cases), especially in those with classic PDE]<br>>75% of PDE patients have intellectual developmental delay (IDD) which is not thought to be correlated with lack of seizure control. IDD may be due to accumulation of neurotoxic metabolites associated with α-AASA dehydrogenase deficiency (LRT is targeted at reducing impact of this on outcome) |
| hRB | Progressive<br>Disrupted function of the tumour suppressor protein RB results in cancer of the eye which can be heritable (either familial or sporadic) or somatic and non-heritable. Bilateral disease usually considered heritable but unilateral can also be heritable. Heritable RB usually occurs at an average of 15 months of age and may be picked up by targeted ocular screening before any symptoms develop if there is family history of the disease. The most common first symptom of RB is leukocoria or visible whiteness of the pupil, which may be noticed in photographs taken using flash photography. Other common symptoms include strabismus (squint), proptosis (protruding eye(s)), glaucoma and hypopyon (presence of pus). If the tumour is large, the eye may become painful and inflamed. High-risk features on presentation (e.g. optic nerve invasion) are more common with increasing age and are associated with poorer outcome. If RB is left untreated, blindness can occur and metastases will most likely develop. Studies have reported detection of RB at birth in some infants. |
| XLHR | Progressive<br>Loss of function of the phosphate-regulating neutral endopeptidase (PHEX protein) leads to increased FGF23 which consequently decreases renal phosphate reabsorption, which increases urinary phosphate excretion and decreases calcitriol production leading to hypophosphatemia and other imbalances.<br>Hypophosphatemia manifests as rickets (i.e. lower leg deformities (from six months), waddling gait, progressive lower leg deformities, delayed gross motor development, widening of the distal metaphysis at the wrists and ankles (from age 1), dental abscesses/malpositions (3 years+), stunted growth, bone pain and hearing loss (older children)).<sup>66</sup> noted that while lower leg deformities may start at six months to one year, it's typically only recognised when toddlers start walking.<br>four criteria are used for clinical diagnosis: hypophosphataemia, increased levels of serum alkaline phosphatase, decreased tubular reabsorption of phosphate, and radiological evidence of rickets.<br>For all four markers to be positive can take up to six months and infants develop them inconsistently.<br>Generally, biochemical markers are detectable before radiological evidence.<br>Presentation with symptoms is often later when stunted growth and bowed legs become apparent. |
| fHLH | Condition requiring activation/reactivation to become symptomatic<br>Malfunction of the perforin/granzyme cytotoxic pathway leads to a proliferation of lymphocytes and overactive macrophages impeding normal downregulation of immune response macrophages and subsequently to an escalation of the immune |
|  | response, including abnormal targeting of red blood cells and cytokine storms leading to anaemia and organ damage.<br>Hyper-inflammatory syndrome with several genetic causes.<br>Usually activates in infancy (during the first year of life in 70%, though later onset is possible, and timing of onset may be dependent on fHLH subtype/variant) following infection. The most common symptoms include: fever that does not respond to antibiotics, rash, hepatomegaly, splenomegaly, enlarged lymph nodes and can lead to seizures, unconsciousness and coma. It is fatal if untreated and reactivates when immune system is triggered again. |
| MCADD | Condition requiring activation/reactivation to become symptomatic<br>Inactivity or deficiency of the medium chain acyl-CoA dehydrogenase (MCAD) protein prevents catalysing the beta-oxidation of fatty acids for formation of ketone bodies in the liver, leading to lack of an alternative energy source during periods of prolonged fasting or increased energy demands resulting in metabolic crises.<br>Individuals present as healthy at birth; first clinical presentation is typically between 3 and 24 months of life (a reduction in overnight feedings can trigger onset of symptoms or common infections in previously asymptomatic individuals), some may present with a metabolic crisis in the neonatal phase before screening results would be available. Others do not present with symptoms until childhood or even adulthood. Most symptomatic cases present from three months to three years of age. Under normal conditions, MCADD patients can use alternative glycogen stores elsewhere in the body. A person with MCADD who never experiences low blood sugar would never experience symptoms.<br>Up to 25% of MCADD-affected individuals will die during their first clinical manifestation.<br>Clinical presentation: unexplained lethargy, vomiting, altered consciousness, hypoglycemia, encephalopathy which may progress to seizures and coma.<br>"Not all individuals with MCADD develop such a clinical presentation, whether a severe metabolic crisis or milder symptoms. This causes some confusion as to whether the MCADD phenotype consists of specific clinical signs and symptoms or the biochemical evidence of an enzyme disorder".<sup>67</sup> Because signs and symptoms are highly variable, no consistent definition of what is regarded as symptomatic MCADD is used in the literature. |

**Table 10.** Overview of management strategies for the 5 conditions.

| Condition | Treatment category<br>(used to select conditions) | Treatment relevant to screening context | Treatments not considered |
| --- | --- | --- | --- |
| PDE | Low cost to both NHS and patient/family | 1st screening relevant Rx: Guideline recommendations are for pyridoxine initiation following PDE-ALDH7A1 diagnosis to prevent or reduce the severity of further seizures. Diagnosis usually occurs following a pyridoxine trial for intractable seizures or based on the combination of seizure and biochemical indicators of PDE, rather than following a genetic diagnosis. The guideline does not explicitly mention asymptomatic treatment initiation, however theoretically, asymptomatic treatment should prevent seizures or reduce severity if seizures occur. Expert advice suggests that treatment is not usually initiated asymptotically because of potential harm from high dose pyridoxine; those who are identified as gene positive are more likely to be treated on first seizure.<br>2nd screening relevant Rx: lysine reduction therapy [LRT] (lysine restricted diet, and arginine supplementation) aims to reduce the accumulation of potentially neurotoxic metabolites (e.g. pipecolic acid or α-AASA, amongst others) in order to prevent or reduce developmental delay.<sup>68</sup> LRT can be initiated soon after birth for infants who are not being breast fed as there are lysine restricted formula milk options.<br>Risk of Rx: The most well-known adverse effect of pyridoxine is sensory neuropathy, however this requires very high doses and seems to have been primarily reported in adults.<sup>69</sup> Clinical expert<br>advises that due to reports of toxicity associated with high dose Rx in infants/children (e.g. pyridoxine-induced seizures), asymptomatic treatment is unlikely to be initiated even where there is a known *ALDH7A1* variant.<br>LRT: Intolerance to a lysine-free amino acid formula, or severe adverse effects (nutritional, neurological or other) (diet should be terminated. In the former case, a natural protein-restricted diet may be considered" van Karnebeek et al. (2015)<sup>70</sup>) | Some children require a combination of pyridoxine and standard anticonvulsants for adequate seizure control. |
| hRB | Long-term surveillance with the associated anxiety and costs | 1st screening relevant Rx: surveillance to identify clinically presenting RB as early as possible (e.g. with fewer tumour foci). [Surveillance is usually based around red reflex testing, which begins after birth and may be repeated every few months until the child is 5 years old. Children with dim or absent red reflex are referred to a specialist ophthalmology service for eye examination under general anaesthetic. In some countries, children with a FH of RB are recommended to have more intensive ophthalmologist surveillance from birth].<br>2nd screening relevant Rx: for clinically presenting cases, 'early' treatment is where local therapies e.g. cryotherapy or laser treatment, can be used to firstly, save vision in the affected eye and ultimately, saving the eye (prevent enucleation). In some cases, these focal therapies may be used alongside chemotherapy. Focal treatments are considered curative but a risk of recurrence or development of RB in the other eye remains<sup>71</sup>, particularly for heritable forms, and children with RB will undergo continuous follow-up care. | Radical Rx: enucleation, radiotherapy or chemotherapy |
|  |  | Risk of Rx: No obvious clinical risks other than that associated with general anaesthetic; all surveillance programmes carry some risk of harm (e.g. psychological distress), particularly for those who do not go on to develop the condition. Risks associated with focal therapy include retinal thinning and hole formation, retinal detachment, vitreous condensation, and vitreous haemorrhage with tumour seeding and cataract.<sup>72</sup> |  |
| XLHR | High and long-term cost to both patient/family and NHS | 1st screening relevant Rx: Calcitriol and oral phosphate (Pi/D) for newborns/ asymptomatic patients = supplements to address low levels of phosphate and active vitamin D and prevent symptoms to develop / severity (traditional first line treatment, can be administered to newborns or before the development of clinical or radiological signs of rickets, started once diagnosed either by biochemical markers, radiographic images or onset of clinical symptoms, +/- genetic testing), feasibly at 2-3 months of age based on hypophosphataemia and increased levels of alkaline phosphatase<sup>73</sup> or following genetic confirmation,<sup>74</sup> treatment is preventative.<br>2nd screening relevant Rx (but not licenced for <1 year olds): Burosumab = monoclonal antibody to neutralise FGF23 to prevent low levels of phosphate and active vitamin D (could be given pre-symptomatically as first line treatment according to clinical advisor, so probably a future treatment), only curative treatment<br>Risk of Rx: | Pi/D regimen for symptomatic disease<br>Cinacalcet (suppresses parathyroid hormone secretion, which influences serum phosphate concentration, suggested as an adjunct treatment)<br>Vitamin D only (dated management strategy)<br>25-hydroxycholecalciferol (=circulating form of vitamin D which is converted to calcitriol the active form. Does not work in vitamin d resistant rickets because a defect in the conversion of vitamin D(3) to its active 25-hydroxy metabolite is not the cause of XLHR) |
|  |  | Toxicity of alfacalcidol (vomiting, diarrhoea, nephrocalcinosis, nephrolithiasis, and reduced kidney function) | Paricalcitol (a vitamin D analogue that prevents and treats secondary hyperparathyroidism)<br>Recombinant human growth hormone (never first line treatment)<br>Corrective surgery (late treatment option when nothing else has worked, not relevant to screening context but could be used as outcome measure for earlier treatments) |
| fHLH | Short-term high costs to NHS but long-term lower costs to NHS and patients | 1st screening relevant Rx: allogenic HSCT (+/- prophylactic treatment up until HSCT) with matched related (1st line) or unrelated (2nd line) donor. Once engrafted, new stem cells can differentiate into healthy blood cells and restore normal immune response. Includes a pre-transplant conditioning regimen: aims to immunosuppress host to that the graft takes. Conditioning regimens can be myeloablative (full dose, carries higher risks) or 'reduced intensity'. Different drug combinations are used in conditioning regimens, that is usually specified in the studies. Only curative treatment.<br>Risk of Rx:<br>conditioning: drug toxicity<br>HSCT itself: graft versus host disease, transplant related mortality (85-90% success rate (alive, cured)) | Chemoimmunotherapy of active disease: Induction therapy following the HLH-2004 protocol (HLH-1994) to suppress life-threatening inflammation by targeting the abnormally activated immune cells: 8 weeks of chemoimmunotherapy to control inflammation and then continued therapy until HSCT available |
|  |  | HSCT is the only presymptomatic treatment option. No official guidance about pre-symptomatic treatment: risks of HSCT need to be weighed against risk of waiting until activation.<sup>75</sup><br>2nd screening relevant Rx: Prophylactic treatment to reduce the chances of disease activation while waiting for HSCT |  |
| MCADD | Existing screening and treatment pathways so impact of WGS would be incremental | Screening relevant Rx: Dietary advice consisting of avoidance of fasting, strict feeding regimens and emergency regimen, with or without low fat diet or dietary supplementation of L-carnitines, preventative measure to avoid low blood sugar levels triggering decompensation (metabolic crisis)<br>Risk of management: None, but low compliance may be an issue | Carbohydrates given by mouth/IV administration of dextrose solution (treatments for symptoms following a trigger/ treatments to reverse catabolism and prevent metabolic crisis) |

PDE – pyridoxine-dependent epilepsy; hRB – heritable retinoblastoma; XLHR – X-linked hypophosphatmic rickets; fHLH – familial haemophagocytic lymphohistiocytosis; Rx – treatment; LRT – lysine reduction therapy; FH – family history; RB – retinoblastoma; Pi/D – calcitriol and oral phosphate; HSCT – hematopoietic stem cell transplantation; HLH – haemophagocytic lymphohistiocytosis

**Table 11.** Definition of early vs late treatment initiation for the 5 conditions.

| Condition | Aim of screening | Definition of “early” * | Definition of “late” |
| --- | --- | --- | --- |
| PDE | To identify PDE in patients before their first seizure. While pyridoxine may not be given pre-symptomatically, it can be given immediately at the time of first seizure without trials of ineffective anticonvulsants. | 1. Asymptomatic or pre-clinical (i.e. prior to first seizure)<br>2. After first seizure (if FH present or known to be *ALDH7A1*+ve)<br>3. After trial of single anticonvulsant<br>The asymptomatic stage ranges from a few hours to days (too short for screening to identify asymptomatic disease) or, less typically, months after birth. There are also reports of unusual fetal behaviour indicating likely antenatal seizures. Some family studies document antenatal treatment with pyridoxine or asymptomatic, prophylactic treatment of second born siblings, however treatment is more usually initiated soon after first seizure (if sibling affected or known to be *ALDH7A1* positive) or following failure to control seizures with anticonvulsant medication (i.e. within days/weeks of first seizure). Pyridoxine is unlikely to be a first line treatment for neonatal seizures per se. | Depends on definition of early<br>1. Any clinical presentation (a to c below)<br>2. After trial of one or more anticonvulsants (b or c below)<br>3. After trial of multiple anticonvulsants (c below)<br>Clinical presentations, include:<br>a. After first seizure<br>b. After trial of single anticonvulsant<br>c. After trial of multiple anticonvulsants |
| hRB | To identify the predisposition of RB in patients before cancer becomes symptomatic to allow surveillance for timely management of early signs of cancer to improve prognosis. | 1. Surveillance from birth, either because gene positive or known FH<br>2. Outcomes in patients with known FH (this option presumes at least some will be screen detected, or detected early because of family awareness of the risk of RB)<br>3. Detection of RB at stage A or B (the earlier the stage at which RB is detected, the greater the chance of local<br>therapeutic options being feasible, or at the very least avoidance of enucleation)<br>4. RB by age on presentation (e.g. <1 year) (age at diagnosis is a proxy for earlier detection, however children can present with more aggressive or advanced tumours even at very young ages) | 1. No surveillance, clinical presentation of RB<br>2. Outcomes in patients with no FH (presume no screening)<br>3. Detection of RB at stage C, D, E<br>4. RB by age on presentation (e.g. ≥6 months or 1 year) |
| XLHR | To identify XLHR in patients before rickets signs present and treat hypophosphatemia to prevent manifestation of rickets. | 1. Asymptomatic or pre-clinical (i.e. before signs of rickets like leg bowing, stunted growth, waddling gait become apparent). Early can mean detected based on a) genetically confirmed XLHR, b) abnormal biochemical markers and/or c) radiological signs of rickets through screening or knowledge of FH with relevant inheritance pattern<br>2. Age <1 year at treatment start (onset of main clinical symptoms from year 1 onwards) to increase the chance that the early group includes asymptomatic or "early symptomatic" patients (unless all <1 year olds are symptomatic indicated by treatment not first line, i.e. on treatment at enrolment, treatment switching reported or type of treatment not screening relevant) | 1. Symptomatic or with clinical signs of rickets (symptoms specified, treatment not first line, patients <1 year excluded)<br>2. Age >1 year at treatment start |
| fHLH | To identify the likelihood of HLH due to a genetic variation before disease activation by infection to consider curative Haematopoietic Stem Cell Transplantation (HSCT). Presymptomatic treatment aims to minimise the time the hyperinflammatory response can be triggered which requires chemoimmunotherapy with limited treatment success. | 1. Pre-symptomatic (=before 1st disease activation), identified through screening (*PRF1*, *STX11*, *STBPB2* or *UNC13D* positive / any biochemical screening (i.e. reduced/absent perforin expression; reduced natural killer cell activity) of either newborns or individuals identified through FH<br>2. Age <3 months at HSCT (diagnosis to HSCT in 6-8 weeks should be achievable) | 1. After 1st disease activation (regardless of whether patient still in active disease or in remission)<br>2. Age >3months at HSCT |
| MCADD | To identify MCAD deficiency in patients before the first metabolic crisis occurs. Pre-symptomatic detection allows management of the deficiency by avoiding fasting as compared to treating life-threatening metabolic decompensation events. | 1. Presymptomatic, i.e. before first trigger (low blood sugar levels that cannot be compensated) resulting in decompensation, identified either through biochemical screening, genetic screening or a combination of both (neonatally or because of FH)<br>2. Age <6 months at start of dietary advice | 1. Symptomatic, i.e. with clinical symptoms of active disease<br>2. Age >6 months at treatment start |

PDE – pyridoxine-dependent epilepsy; hRB – heritable retinoblastoma; XLHR – X-linked hypophosphatmic rickets; fHLH – familial haemophagocytic lymphohistiocytosis; MCADD – medium chain acyl-CoA dehydrogenase deficiency; FH – family history; RB – retinoblastoma; HSCT – hematopoietic stem cell transplantation; \* Option 1. is the preferred option. All subsequent options (2 onwards) are considered a step down and were only taken forward to data extraction if no studies for option 1 existed

Appendix 5 (Table 25 to Table 29) summarises the outcomes from studies investigating early vs late treatment as defined by the studies for the five conditions. The detailed quality assessment is included in Appendix 3Appendix 1.

#### Pyridoxine dependent epilepsy

There were five studies that compared relevant outcomes in children ’treated early’ for PDE and children ‘treated late’ for PDE (Appendix 5, Table 25).^68, 76–79^ Two studies included a series of families^76, 77^ and three^68, 78, 79^ were single-armed studies where patients were recruited retrospectively. Treatments included pyridoxine monotherapy,^76, 77^ lysine reduction therapy^78^ or a combination of the two.^68, 79^ The definition of ‘early’ and ‘late’ treatment varied between the studies. Two studies defined ‘early treated’ as the sibling with the shortest delay in receiving treatment after seizures began.^77, 79^ One study investigated the impact of treatment given antenatally (or asymptomatically) compared to symptomatically,^76^ and the remaining two studies used age at treatment initiation to differentiate early and late treatment.^68, 78^ The number of PDE cases in the included studies were low and ranged from four (from two families)^76^ to 60.^68^

None of the studies reported seizure control as an outcome but instead focused on developmental outcomes. Three studies reported intelligence quotient (IQ) scores,^76, 78, 79^ one reported a ‘standardised developmental assessment’ score (similar to IQ score),^68^ and one reported psychomotor development.^77^ Other reported outcomes included motor performance^76^ and a standardised neurological outcome.^78^ The timing of the outcome assessment was not reported in two studies^77, 78^ and ranged between 4 years (age of one child)^76^ and a mean of 15 years of age^79^ in the other studies.

There is some indication of improved developmental outcomes in ‘early treated’ children. In studies that reported IQ scores^76, 78, 79^ or a quantitative measure of developmental assessment,^68^ scores were higher in the ‘early treated’ groups, apart from the in the study by Tseng and colleagues.^79^ When treated with pyridoxine and lysine reduction therapy, the full-scale IQ score was slightly lower in the ‘early treated’ group (76.0 versus 77.4). Psychomotor development^77^ and motor performance^76^ were slightly better in the ‘early treated’ group; all three children in the ‘early treated’ group were assessed as normal in the study by Jiao et al. (2021)^77^ and compared to only one child assessed as normal in the ‘late treated’ group, and one ‘early treated’ child out showed a slightly better outcome than their ‘late treated’ sibling in the study by Bok et al. (2010) (walking four months earlier).^76^ There was insufficient evidence to determine whether there were any apparent differences as a result of type of treatment (pyridoxine monotherapy, lysine reduction therapy or a combination of the two).

##### Quality assessment

We had methodological concerns about all studies^76–79^ but one^68^ regarding the Selection of patients. The Selection process was not clearly reported in these studies and no recruitment dates were given. The follow-up period was not reported in one study,^78^ and in a second^77^ one child was only six months old at last follow-up. In addition, the description of the assessment processes was inadequate in the study by Jiao et al. (2021).^77^

##### Case studies or ‘early only’ studies

In addition, we identified six case studies that compared outcomes in early versus late treated siblings.^80–85^ Results from four of these case studies suggest that earlier treatment does not improve clinical outcomes.^80–82, 84^ Two studies reported better outcomes in the ‘early treated’ sibling.^83, 85^ No studies that looked at outcomes in ‘early treated’ patients alone were identified.

#### Heritable retinoblastoma

Five studies compared relevant outcomes in children who underwent pre-emptive surveillance to allow early detection of RB (‘treated early’) compared to those who presented clinically (‘treated late’) (Appendix 5, Table 26).^86–90^ All five studies were retrospective and single-armed.^86–90^ Two studies^87, 88^ were multi-centre and three^86, 89, 90^ were single-centre studies. The number of included RB cases ranged from 13^90^ to 264.^86^ Four^87–90^ of the five studies included children with a family history of RB, and the fifth study^86^ included all cases of RB, however the main comparisons of relevance that were reported were in those with a family history of RB (with or without surveillance).

Four studies^86, 88–90^ compared outcomes in ‘screened’ versus ‘not screened’ at the participant level, one^89^ of which further categorised the ‘screened’ group into intensively screened and screened. In the remaining study,^87^ the income status of the country was used as a proxy for ‘screened’ versus ‘not screened’, since it was assumed that more children in a high-income country will be detected by screening than their counterparts in low- and middle-income countries.

There was some variability in reported outcomes between studies. In three studies, mean or median age at diagnosis was lower in screen detected versus clinically detected RB suggesting that screening does allow earlier detection of RB than would happen otherwise (mean 4.9 months versus 17.2 months,^88^ mean 4.7 months versus 16.7 months^90^ and median 0 months for intensively screened, four months for screened and nine months for not screened.^89^ In terms of patient health outcomes, some measure of both ocular survival and survival was reported in three studies on a per participant level.^86, 89, 90^ The largest study^86^ using Kaplan-Meier analysis demonstrated considerably higher rates of ocular survival at one year for the screened family history group (n=86) compared to the not screened family history group (n=178) (83.2% compared to 47.5%), with a smaller difference at five years (67.7% compared to 58.2%). In contrast however, a marginal difference in survival at one year (100% versus 97.3%) had increased slightly by five years (93.2% versus 87.4%), potentially suggesting a benefit from earlier detection.^86^

Similar results were reported by Rothschild et al,^89^ with enucleation rates of 0% (0/16) for those intensively screened, 8.7% (2/23) for those ‘screened’, and 65% (13/20) for those not screened, and mortality rates of 0% for both screened groups, and of 5% (1/20) for the not screened group (follow-up time point not reported). The smallest study reported enucleation rates of 0% for those screened (0/5) and 75% for those not screened (6/8), with no deaths (median follow-up of 4.8 years).^90^

The final study reported considerably lower enucleation rates in the USA (25%; 8/32) compared to in developing countries (71.7%; 43/60), and a considerably higher probability of event free survival at five years (0.92 (SD 0.05) in the USA compared to 0.81 (SD 0.07) in developing countries).^87^ It is likely that other differences in the delivery of care have contributed to the observed differences in outcomes, and it is not possible to properly attribute these differences to the effect of screening alone.

*Quality assessment* We had concerns regarding whether the length of follow-up was long enough for outcomes to occur in one study.^88^ In four of the five studies, reporting was not sufficient to allow others to replicate the research.^86–88, 90^

In addition, we identified three studies only reporting on ‘early treatment’, i.e. Identification of RB via surveillance.^91–93^ Results from two of the three studies identified high proportions of infants with RB present in the first one to two weeks after birth (70% (12/17)^91^ and 50% (4/8)^92^). The third study reported screening of 23 asymptomatic siblings of probands with RB; 13% (3/23) were identified as having active RB on screening at a median of six months of age.^93^ It is not possible to determine whether the use of surveillance in these studies resulted in ‘better’ outcomes than would have occurred if they had presented clinically.

#### X-linked hypophosphataemic rickets

There were three studies that compared the outcomes in children with XHLR who underwent early treatment with those who received treatment later.^49, 94, 95^ All studies implemented standard treatments for rickets as opposed to the more recently licensed Burosomab. No studies using Burosumab as first line treatment were identified. Appendix 5, Table 27 provides the summary of their findings. All three studies were retrospective and conducted without control groups. Two of these studies involved multiple centres,^49^ ^95^ and one was conducted at a single centre.^94^ All three studies used the same definition of early (before one year of age) versus late treatment (at or after one year of age). In one study patients in the early treatment group were diagnosed prior to the onset of clinical signs of rickets, while those in the late treatment group were diagnosed after the appearance of clinical symptoms.^94^

Treatment involved administering oral phosphate, vitamin D, or an analogue of vitamin D (Alfacalcidol) (Pi/D) daily. The reported results in the studies included measurements of height and various biochemical parameters such as serum calcium, phosphate, ALP, creatinine, parathyroid hormone, and vitamin D3 levels. Makitie et al. (2003) and Quinlan et al. (2012) assessed the activity of rickets through radiographic examination.^94, 95^ Additionally, Makitie et al. (2003) made predictions regarding adult height. The evaluation of results occurred at different time intervals across the three studies.^94^ Makitie et al. (2003) measured outcomes at the end of the first year of treatment and before puberty.^94^ Rafaelsen et al. (2016) conducted measurements at each clinic visit and assessed the results at the last recorded consultation. The mean age at the last recorded consultation of the early group was documented as 11.1 years, while the late group had a mean age of 8.4 years.^49^ Quinlan et al. (2012) analysed outcomes at medium treatment durations of 8.5 years and 11.9 years for early and late treatment groups, respectively.^95^

The findings from these studies suggest that initiating treatment during the early stages of growth moderately enhances outcomes for patients with XLHR. Both Makitie et al. (2003) and Quinlan et al. (2012) reported median height closer to the expected average in the early treatment compared to the late treatment groups (SD scores of -0.7 (n=8) vs. -1.8 (n=11), p=0.009;^94^ and -0.7 (n=10) vs. -2.0 (n=13), p=0.009^95^). Additionally, Makitie et al. (2003) reported pre-pubertal height closer to the expected average in the early (-1.3 SDS; n=8) compared to the late treatment group (-2.0 SDS; n=11) (p= 0.054). Treatment effect in the early treated group (z-score -0.2) compared to the late treated group (z-score -1.2), was within that expected by chance (P=0.06) was observed by Makitie et al. (2003).^94^ In contrast, results from Rafaelsen et al. (2016) showed same trend as Makitie et al. (2003) but were within that expected by chance.^49, 94^

Regarding biochemical parameters, Makitie et al. (2003) found a similar degree of hypophosphatemia between groups, however serum alkaline phosphatase levels remained elevated in the late treatment group throughout childhood.^94^ Quinlan et al. (2012) reported no difference in median levels of serum phosphate or serum alkaline phosphatase between the early and late treatment groups.^95^

In terms of rickets activity, Makitie et al. (2003) observed more pronounced radiographic signs of rickets in the late treatment group, whereas patients receiving early treatment still displayed significant skeletal rickets changes.^94^ Quinlan et al. (2012) reported similar ricket severity scores in both groups.^95^

##### Quality assessment

The evaluation of study quality revealed that Quinlan et al. (2012) lacked sufficient information on patient selection, while it remained uncertain whether the follow-up duration in Rafaelsen et al. (2016) was adequate to assess outcomes.^49, 95^ The earlier study by Makitie et al. (2003) satisfactorily addressed all aspects of the quality assessment.^94^

In addition, we identified three studies only reporting on early treatment of which one of them is a case report. Exploration on early treatment indicated that early treatment could enhance metabolism, promote growth, and mitigate deformities. However, these findings are not entirely consistent with previous studies by Makitie et al. (2003) and Quinlan et al. (2012), which suggested that early intervention does not always result in complete normalisation of outcomes.^94, 95^ Conversely, Rafaelsen et al. (2016) reported no substantial differences in outcomes between early and late treatment groups.^49^

#### Familial hemophagocytic lymphohistiocytosis

There was one retrospective, multi-centre study that evaluated outcomes for children who received treatment for fHLH following asymptomatic detection before activation (treated ‘early’) and children who received treatment following clinical symptomatic detection or activation of fHLH (treated ‘late’) (Appendix 5, Table 28).^75^The study included 32 genetically confirmed sibling pairs/triplets with fHLH. The asymptomatic children were diagnosed following diagnosis of their sibling. Outcomes included mortality, cause of death and number of patients in complete remission at end of follow-up. The follow-up period for each patient was different.

We present results separately for this study, for per-protocol and intention to treat populations, using a slightly different definition for ‘early’ and ‘late’ treatment.

##### Intention to treat population

For the intention to treat population, we defined ‘early’ and ‘late’ treatment as cases that were asymptomatic and symptomatic at diagnosis, regardless of activation before or after treatment initiation. In other words, the ‘early’ treated group in the intention to treat population included patients who were asymptomatic at diagnosis but with some subsequently experiencing activation either before or shortly after commencing treatment, and this may confound the true effect of ‘early’ treatment on outcomes.

Mortality was lower amongst the ‘early’ treated group (15% versus 38%). Six patients (two (8%) in the ‘early’ treated group and four (15%) in the ‘late’ treated group) died due to transplant complications, which was the most common cause of death. The proportion in complete remission was higher in the ‘early’ treated group (81% versus 62%).

##### Per-protocol population

Ideally, ‘early’ treatment would begin in individuals before their first activation. However, individuals who present as asymptomatic may have experience disease activation before or shorty after initiation of treatment. Therefore, for the per-protocol population, we defined ‘early’ and ‘late’ treatment using four distinct groups, dependent on activation status. The ‘early’ treated group can be split into two categories: asymptomatic patients who were treated with HSCT +/- prophylactic treatment and did not activate (group one, n=15), and asymptomatic patients who were treated with prophylactic treatment and subsequently activated (group two, n=3). Similarly, for the ‘late’ treated patients, two groups can be defined: those who were symptomatic and subsequently treated with active disease protocol +/- HSCT (group three, n=26), and those who were asymptomatic at diagnosis but experienced disease activation before the start of treatment and thus were treated with active disease protocol +/- HSCT (group four, n=7). Defining the treatment groups in this way helps to reduce any potential confounding effects that activation may have had on outcomes and allows us to better isolate the impact of ‘early’ treatment of truly asymptomatic individuals on clinical outcomes. Mortality was lowest amongst group one (7%), and similar between the remaining three groups (group two 33%, group three 38%, group four 27%). Note that group one reflects those with less severe disease than group two, since these patients did not activate after treatment initiation. Also, group two only included three patients, one of whom died. The most common cause of death was disease progression. Three patients died following transplant complications (four in group three and two in group four). The number of patients in complete remission was highest in group one (93%). Proportions were considerably lower in the other three groups (group two 33%, group three 10%, group four 7%). However, group one had the shortest median follow-up time, so it is possible that patients in this group subsequently experienced symptoms or other events. One patient in group two was lost to follow-up.

Group three, who were symptomatic at diagnosis, and likely represent those with most severe disease, had the highest proportion of deaths and lowest proportion of patients in complete remission at the end of follow-up. Outcomes in the groups who had experienced activation following an asymptomatic diagnosis either before (group four) or after (group two) the start of treatment, were similar to group three. The opposite was true however for those in group one, which indicates that ‘early’ treatment before activation has been experienced may be effective. Further, only three asymptomatic patients with no previous activation subsequently activated after commencing ‘early’ treatment. As previously mentioned, this reduces our confidence in results from group two, but does however increase confidence in the conclusion that treatment in asymptomatic individuals with no previous activation may be effective.

##### Quality assessment

The primary quality concern in this study is the lack of detail regarding which International centres provided the data, and whether they sent in details for all eligible patients in their care. The patient characteristics, treatment and outcomes were reported adequately, and the follow-up time was sufficient to assess treatment outcomes.

We did not identify any studies looking at early (pre-symptomatic) treatment for fHLH only. We found one case report presenting a case of fHLH2 in one twin, with the other twin also harbouring the same homozygous variant but not presenting with either clinical or biochemical signs of the disease.^96^ Treatment was not presented for either twin.

#### Medium Chain Acyl-CoA Dehydrogenase Deficiency

There were nine studies that looked at relevant outcomes in children with MCADD who received early management following asymptomatic detection through screening (treated ‘early’) and those who received management following clinical symptomatic detection of MCADD (treated ‘late’) (Appendix 5, Table 29)^97–105^ All but one^97^ studies were single-arm and all were conducted retrospectively.

One study presented two patients, one received dietary management from five months following diagnosis by NBS screening, the other received management following symptom presentation. ^97^ Three Australian studies had overlapping patient cohorts.^101, 103, 104^ The sample sizes in the included studies ranged from two^97^ to 90.^99, 106^

The definitions of ‘early’ and ‘late’ treatment varied slightly between the studies. In eight of the nine studies,^97–104^ ‘early’ treatment was defined as management following detection of MCADD through NBS screening. This group also included those detected through family screening in one study.^98^ In six of the eight studies,^97, 99–101^ ^103, 104^ patients were all asymptomatic, but in two studies this group also included symptomatic patients.^98, 102^ In Wilson et al. (1999),^105^ ‘early’ treatment was management following asymptomatic screening due to an affected sibling. In all nine studies, ‘late’ treated was defined as management following clinical presentation of MCADD.^97–105^ One study also included those detected through family screening in the ‘late’ group, and it is unclear how many patients this group included and whether they were symptomatic or asymptomatic.^99^ It is important to bear these differences in the definitions of ‘early’ and ‘late’ treatment in mind when interpreting results. Where reported, management strategies included avoidance of fasting,^98, 99, 102, 103, 105^ carnitine supplementation,^98–100, 102, 105^ various diets^97, 99^ ^100, 102, 103^ and sick-day regimens.^103, 105^

The reported outcomes were heterogenous and included mortality/severe episodes, descriptions of patients’ clinical statuses, various measures of physical and psychological development, healthcare use and biomarker levels. Results for each of these outcomes are described separately below. Follow-up length was varied, but two studies reported outcomes within the first four years of life.^101, 103^

##### Mortality/severe episodes

Four studies reported mortality as an outcome.^100, 101, 103, 105^ This outcome was assessed at four years of age in two studies,^101, 103^ and follow-up was variable in the other two studies study.^101, 105^ In two studies, the proportion of patients who died by age four was lower in the ‘early’ treated groups (4% versus 17%^101^ and 4% versus 19%^103^ in the early and late treated groups, respectively). In the study by Wilson et al. (1999) 21% and 17% of children had died by age six in the early treated group and late treated groups, respectively.^105^ However, it is important to note that in this study, the definition of early treatment was not those detected by NBS screening and instead this group included those diagnosed due to an affected older sibling. No patients died in the study by Gong et al. (2021).^100^ One study reported the number of severe episodes by age two and four.^103^ At both ages, the percentage who had experienced a severe episode was lower in the ‘early’ treated groups.

##### Description of clinical status

Four studies reported the clinical status of patients after varying follow-up periods.^97, 98, 100, 107^ Where reported, age at assessment ranged between 24 months^97^ and 11 years.^107^ No studies assessed patients after a standardised follow-up period. In one study, all six children (four ‘early’ treated and two ‘late’ treated) were assessed as normal.^107^ In two studies, the ‘early’ treated groups were assessed as normal but some children in the ‘late’ treated groups showed clinical abnormalities including severe seizure disorder and cerebral palsy, nasogastric feeding in one child,^97^ one patient with intermittent fasting hypoglycaemia and one patient with hemiplegia due to disease episode.^100^ In the fourth study, nine (29%) were symptomatic in the ‘early’ treated group, and both children in the ‘late’ treated group were symptomatic.^98^

##### Physical and psychological outcomes

Various measures of physical and psychological outcomes were reported and overall, there appeared to be few differences between the ‘early’ and ‘late’ treated groups. There were no differences in terms of height, weight or neuropsychological function (within the first four years of life) in one study,^101^ or in terms of intellectual ability score (assessed at more than four years of age) in another study.^103^ In Wilcken et al. (2009), one child (4%) in the late treated group had a mild intellectual handicap, and two (8%) required extra assistance at school whereas the children in the ‘early’ treated group were all assessed as normal (at six years of age or last follow-up).^104^

##### Healthcare use

Four studies reported some measure of number of hospital/emergency rooms visits.^99, 101, 103, 105^ The mean (95% CI) number of hypoglycaemia related hospital days and ER visits per patient years was slightly lower in the ‘early’ treated group (0.09 (0.03-0.15) versus 0.11 (0.04-0.19)) in the study by Anderson et al. (2020).^99^ However, in this study it is important to note that the ‘late’ treated group may have included asymptomatic, screen detected patients. In two studies, the percentage of patients who had previously been admitted to hospital was lower in the ‘early’ treated groups (42% versus 85% (by age 4)^103^ and 25% versus 36% (variable follow-up)^105^). One study reported whether the hospital visits were inpatient, emergency, or outpatient within the first four years of life.^101^ The percentage of children with inpatient stays and emergency room visits was lower in the ‘early’ treated group, but the number of outpatient visits was higher in this group. Length of inpatient stay in those admitted was also similar between the ‘early’ and ‘late’ treated groups.

##### Biomarker levels

Biomarker levels at diagnosis were reported in one study.^100^ Mean levels of C6, C8 and C10 were lower in the ‘early’ treated children but there were no differences in the mean C8:C2 or C8:C10 ratios. However, it should be noted that these measures were reported inconsistently.

##### Quality assessment

In four of the studies Gong (2021), Haas (2007), Li (2019), and Wilcken (2007), we found that cases were not described in sufficient detail to allow for replication of the study or to make inferences, this was mostly due to the paucity of treatment definition and description.^100, 101, 103, 107^ In 4 studies, the method for Selection of patients was unclear.^97, 98, 100, 107^ The follow-up times were variable, and it was unknown in one study.^98^

In addition, we identified nine studies only reporting on early treatment in which MCADD patients were detected through NBS screening or family studies and no patients detected and treated following symptom onset.^59, 108–114^ One further study identified five cases admitted to hospital before their NBS test results became available but outcomes were not reported separately for the NBS screening group.^63^

#### Conclusions and learning from the review of five conditions

The five traditional reviews yielded insufficient evidence in populations relevant to a screening context to inform a UK NSC decision about WGS in newborns. The lack of evidence on penetrance and expressivity of the genetic variants for all five conditions does not appear to support the Identification of specific conditions or groups of individuals for which WGS may be beneficial in a screening context.

Available evidence about the genetic spectrum of patients with symptomatic or biochemical disease frequently pointed to small numbers of recurrent variants accounting for considerable proportions of cases, nevertheless large numbers of variants were novel or occurred only in a very small number of cases. The pathogenicity and penetrance of such variants is unclear. We also saw that genetic spectrum can be strongly affected by factors such as ethnicity and the prevalence of consanguinity. Studies often employed a suite of genetic testing methods rather than relying on sequencing alone, such that we could not determine the proportion of cases that could have been identified with WGS alone. As such the results observed are not necessarily transferable to the screening context of apparently healthy newborns.

In terms of identifying benefit from earlier treatment, our results generally reflect acknowledged difficulties in evaluating the effectiveness of interventions for rare conditions. Studies were generally small, with variable definitions of ‘early’ and ‘late’ both within and between conditions, and a frequent reliance on earlier intervention in siblings or where there was a known family history. Only a few examples of asymptomatic or very early initiation of treatment were identified. Outcome measures were often short-term, with limited follow-up to identify longer term patient-relevant outcomes such that any benefits from earlier diagnosis resulting from newborn screening will be difficult to quantify.

A few aspects were identified that would render certain conditions less likely candidates for a newborn screening programme using WGS:

- The genetic heterogeneity is large and novel potentially pathogenic variants are common causing a lot of uncertainty which has downstream implications in terms of time needed to determine pathogenicity
- The type of frequent variations (mosaicism, large deletions etc.) are not sufficiently captured by WGS, and require additional genetic testing
- The presymptomatic phase is likely to be shorter than the time until test results are available for diagnosis
- An early intervention phase cannot be defined
- The preferred curative treatment option is not licensed for newborns
- The curative treatment option carries a high risk of adverse events
- The available treatment option is for symptom management only

A single approach to reviewing five conditions was not feasible and a review of 200 conditions would require 200 individual reviews. The five reviews were undertaken by three full time and two part time reviewers and took seven months to complete without writing up the findings. A review team of similar size could take as much as 280 months (23 years) to undertake 200 consecutive reviews for 200 conditions. Some learning may be transferable between reviews of similar conditions shortening certain review processes. For instance, inborn errors of metabolism are a group of related conditions that are generally managed with a specific diet or dietary advice, therefore some thinking and decisions may be applicable more widely across several conditions. We could not explore this with the five conditions reviewed which we selected for a range of treatment scenarios. The five conditions were highly varied in their characteristics, treatment and aim of screening and the review process had to be tailored for each condition. Firstly, each search was developed individually, requiring an understanding of the condition in terms of:

- The presence of a non-genetic version in addition to a genetic version of the condition
- Alternative names and aliases
- Relevant umbrella terms.

Secondly, categorisation of sequencing studies by the studies’ disease definition required condition-specific categories depending on:

- The availability of biochemical tests (i.e. the definition of disease in biochemical terms is possible)
- The number of disease groups with overlapping symptoms
- Whether conditions were only defined genetically
- Whether the condition is already screened for.

Thirdly, the definition of early versus late was specific for each condition and depended on:

- Whether the relevant intervention is earlier detection or therapeutic, and whether the treatment is preventative, curative or management of symptoms
- Whether an early intervention phase could be defined
- Whether conditions are progressive or present following a trigger
- Whether early could be defined in other terms than presymptomatic (e.g. early stage of progressive disease).

### 2 Exploring ClinGen as an evidence source for the UK NSC

ClinGen is an evidence review resource that could be potentially used for the evaluation of WGS for 200 paediatric conditions. We, therefore, assessed the evidence base provided in ClinGen for the five conditions reviewed in our traditional review and compared ClinGen scoring dimensions and cores to the UK NSC criteria and our decisions. We also assessed the alignment of the dimensions and UK NSC criteria to the four principles used by Genomics England for decision on gene inclusion. High agreement between criteria from the three resources would mean that decisions from ClinGen and/or Genomics England could inform UK NSC recommendations in the future. The methods for this assessment are reported in section 2 of the methods chapter.

#### ClinGen as an evidence source for the five conditions in our review

We considered the gene-disease validity, variant classification (level of pathogenicity) and the actionability scores reported by ClinGen. The gene-disease validity has been confirmed in ClinGen for the eight genes of the five conditions considered in our review, and four of the five conditions reviewed (PDE, MCADD, hRB and XLHR) had a paediatric actionability report available on ClinGen in March 2024. However, no information on variant classification in terms of pathogenicity was available for any of the genes. Therefore, the resource has got limited informative value for decision making on the variant level. The evidence provided on the actionability of the four conditions with a paediatric actionability report is explored in the next chapter.

#### Comparison of the paediatric reports for PDE, MCADD, hRB and XLHR from ClinGen with our review findings

The paediatric actionability reports for PDE, MCADD, hRB and XLHR provide information and scores on four dimensions: severity of disease, penetrance, treatment effectiveness and burden of intervention. These are the most relevant aspects in determining the medical actionability for genetic conditions identified as incidental findings according to ClinGen. Table 12 summarises the ClinGen scores for the four conditions.

According to ClinGen, PDE, hRB and MCADD are conditions with high actionability (overall score 10 or 11 out of 12) and XLHR is pending an actionability assertion. XLHR has got an overall score of 9 suggesting moderate actionability based on the score alone, however, the Paediatric Actionability Working Group may override the score based on their clinical expertise. At the time of review, the evidence searches for the paediatric reports were 3.5 to 6 years old (Appendix 6, Table 30).

The severity of disease for all four conditions was scored 2 out of 3 (a reasonable possibility of death or major morbidity) and penetrance was scored 3 out of 3 (>40% chance of serious outcome). A summary of the penetrance findings is provided in Appendix 7. The score for penetrance was based on minimal evidence (level C evidence) for PDE, XLHR, and hRB (Table 22) and on a single published study identified non-systematically through expert review for MCADD. The evidence on penetrance for PDE and XLHR was provided by GeneReviews without further references to a primary data source (Appendix 6, Table 30).^115^ For hRB, the penetrance information came from a non-systematic review and a published guideline, in addition to GeneReviews. The penetrance information for MCADD was based on a published study of 81 NBS test positive newborns identified through the Danish NBS screening prgramme.^116^ Overall, 7/12 references that form the evidence base for penetrance and expressivity for these four conditions were websites, no evidence cited was based on sequencing data of an unselected newborn population, and no additional references to our four systematic reviews were identified (Appendix 6, Table 30).

The treatment available for PDE and MCADD was rated as highly effective (score 3 of 3) with minimal risk (score 3 of 3) (Table 12). Surveillance for hRB, while highly effective (score of 3), was judged to be associated with moderate risk (score of 2). The treatment for XLHR was classed as moderately effective with moderate risk (both score of 2). The treatments considered for the conditions are summarised based on the ClinGen information in Appendix 7, Table 31. The evidence base for the effectiveness of treatment and surveillance was classed as moderate for hRB, MCADD and PDR where at least one guideline or treatment recommendation was identified (5/8 as websites only, 3/8 published in peer reviewed journals), and minimal for XLHR in the absence of a clinical guideline or treatment recommendation. The evidence on the treatment effectiveness of oral phosphate and calcitriol (Pi/D) for XLHR was based on one non-systematic review, information from the OMIM website and GeneReviews. The main information that ClinGen included from GeneReviews was the findings from the retrospective study looking at Pi/D treatment in patients with XLHR <1 year of age versus >1 year of age by Makitie et al. (2003) included in our review above.^94^ One more reference to early versus late management was available for hRB, where the information provided was for screened (family members of probands with hRB) versus probands. This study concluded that surveillance resulted in earlier diagnosis and better outcomes measured by likelihood of enucleation, eye radiation and visual acuity, however less than half of the early group actually received surveillance.^117^ This study formed part of the treatment recommendation by Skalet et al. (2018),^118^ cited in the ClinGen paediatric actionability report for hRB. This study was missed by our searches but would not have been included in our review because the definition of early treatment did not meet our inclusion criteria.

Comparison of the ClinGen scores of the five conditions to our review findings using the UK NSC criteria and Genomics England’s decisions for gene inclusion is shown in Table 12. This was complicated by three aspects. Firstly, the ClinGen scores out of 3 (with being best for actionability) for the four dimensions do not provide a definitive yes/no assessment precluding an evaluation of the level of agreement for the individual criteria.

Secondly, the criteria for assessment do not map well across ClinGen, the UK NSC and GEL. While ClinGen assessed the severity of disease in the diagnostic context of an incidental finding in an individual, the UK NSC criterion 1 requires the assessor to consider the importance of disease from a public health perspective, considering whether screening for the condition is worth it, whether the natural history of the disease is understood and the link between the risk factor (here a genetic variant) and the disease is known. GEL’s principle A focuses more simplistically on the level of evidence that proves the gene-disease link.

The criterion around penetrance differs between ClinGen and GEL’s principle B versus UK NSC criterion 3 because the UK NSC criterion focuses on screen detected variants rather than variants detected in patients with confirmed disease. This means that the studies identified for question 2 in our traditional review only partially address this criterion and do not provide the evidence needed to understand the penetrance in the complete spectrum of genetic disease identified through screening. This explains at least partly the different outcomes of assessment for this criterion.

There are also some important differences in the evaluation of the evidence of treatment effectiveness. While ClinGen requires an available and effective treatment that can be used in presymptomatic individuals, the UK NSC and GEL also require evidence on the benefits of earlier treatment compared to treating symptomatic disease. And while ClinGen considers the burden of the treatment separately in their fourth dimension, GEL’s principle D is focused around equity in access to treatment and UK NSC criterion 10 focuses on available evidence of a treatment pathway for early treatment. While some criteria map reasonably well across GEL and the NSC, the evidence bar appears to be higher for the UK NSC which has led to different decisions for 5/5 conditions concerning the treatment effectiveness. In contrast to the UK NSC, who requires a systematic and comprehensive approach to evidence synthesis, GEL’s approach was similar to that used by ClinGen in that a single supporting reference was sufficient to meet Genomic England’s principles for gene inclusion and the evidence was often based on information provided by ClinGen or GeneReviews. This means that all five conditions met the Genomics England principles, while the current evidence base for none of the conditions meets all UK NSC criteria. Furthermore, ClinGen and GEL assessed gene-condition pairs against four criteria while the UK NSC requires evidence to a further 16 criteria which are not part of this comparison.

Thirdly, the subjective approach to reach a final decision is not transparent and decisions may not follow from the evidence. GEL and ClinGen heavily rely on the expertise and opinion of an expert clinical panel. The UK NSC on the other hand relies on a balanced interpretation of the amount and quality of evidence which is reviewed during stakeholder consultations.

Overall, it appears that different outcomes in the assessment of the five conditions are due to a combination of 1) different focus in the assessment criteria used (particularly the individual patient versus the public health perspective), 2) the expected limited evidence base for these rare conditions and a higher evidence bar used by the UK NSC and 3) the subjective interpretation of the evidence base to reach a final decision. This means that neither the ClinGen scores, nor the GEL decisions would be an appropriate proxy for a UK NSC recommendation for the five conditions. And using ClinGen as a short cut into the evidence base for a future review of genomic screening of 200 conditions may not be feasible nor appropriate.

**Table 12.** Summary of actionability for the five conditions based on ClinGen, our review and Genomics England.

| Condition | ClinGen dimensions (score* / level of evidence**) | NSC criteria*** (met / not met / rationale) | GEL Principles (met [y/n] / evidence) |
| --- | --- | --- | --- |
| THE CONDITION |  |  |  |
|  | Severity: What is the nature of the threat to health to an individual carrying a clearly deleterious allele in this gene? | 1.The condition should be an important health problem as judged by its frequency and/or severity. The epidemiology, incidence, prevalence and natural history of the condition should be understood, including development from latent to declared disease and/or there should be robust evidence about the association between the risk or disease marker and serious or treatable disease. | A) There is strong evidence that the genetic variant(s) causes the condition and can be reliably detected. |
| PDE | 2 / - | Met / intractable neonatal seizures associated with intellectual development delay in >75% cases and if uncontrolled leading to death, gene-phenotype link has been verified | Y / ClinGen<sup>23</sup> |
| fHLH | - / - | Met / abnormal immune response leading to fatal multi-organ failure in infancy, gene-phenotype link has been verified | Y / Trizzino 2008<sup>56</sup> – PRF1, Gadoury-Levesque 2020<sup>119</sup> – UNC13D, Al Ahmari 2021<sup>120</sup> – STX11, Gadoury-Levesque 2020<sup>119</sup> |
| MCADD | 2 / - | Met / metabolic disease with symptomatic presentation within first 2 years of life leading to death in 20% cases if undiagnosed, gene-phenotype link has been verified, already part of NBS program | Y / ClinGen<sup>23</sup> |
| hRB | 2 / - | Met / malignant neoplasm of the eye with symptoms onset at about 15 months of age leading to blindness and metastases, gene-phenotype link has been verified | Y / ClinGen<sup>23</sup> |
| XLHR | 2 / - | Met / hypophosphatemia leading within first 2 years of life to bone deformity, dental abscesses, stunted growth and bone and joint pain, gene-phenotype link has been verified | Y / Ruppe 2011<sup>121</sup> |
|  | Likelihood of disease: What is the chance that a serious outcome will materialize given a deleterious variant (akin to penetrance)? | 3.If the carriers of a variant are identified as a result of screening, the natural history of people with this status should be understood, including the psychological implications. | B) A high proportion of individuals who have the genetic variant(s) would be expected to have symptoms that would have a debilitating impact on quality of life if left undiagnosed. |
| PDE | 3 / C | Not met / penetrance of variants detected through screening/sequencing is not known, only information on genetic spectrum in children with disease is available | Y / GeneReviews<sup>115</sup> |
| fHLH | - / - | Not met / penetrance of variants detected through screening/sequencing is not known, only information on genetic spectrum in children with disease is available | Y / GeneReviews<sup>115</sup> |
| MCADD | 3 / N | Not met / penetrance of variants detected through sequencing is not known, only information on genetic spectrum in children with disease or with positive NBS test is available | Y / GeneReviews<sup>115</sup> |
| hRB | 3 / C | Not met / penetrance of variants detected through screening/sequencing is not known, only information on genetic spectrum in children with disease is available | Y / GeneReviews<sup>115</sup> |
| XLHR | 3 / C | Not met / penetrance of variants detected through screening/sequencing is not known, only information on genetic spectrum in children with disease is available | Y / GeneReviews<sup>115</sup> |
| THE INTERVENTION |  |  |  |
|  | Effectiveness: How effective is the selected, specific intervention for preventing or significantly diminishing the risk of harm when initiated during childhood (< 18 years)? | 9. There should be an effective intervention for patients identified through screening, with evidence that intervention at a pre-symptomatic phase leads to better outcomes for the screened individual compared with usual care. Evidence relating to wider benefits of screening, for example those relating to family members, should be taken into account where available. However, where there is no prospect of benefit for the individual screened then the screening programme should not be further considered. | C) Early or pre-symptomatic intervention for the condition has been shown to lead to substantially improved outcomes in children, compared to intervention after the onset of symptoms. |
| PDE | 3 / B | Not met / the evidence-based direction is that there might be some benefit in early treatment; however, the quality and volume of the evidence is low because of the definition of early vs late, the type of study, the number of participants, and the number of studies therefore, there is insufficient evidence to clearly judge the effect. | Y / GeneReviews<sup>115</sup> |
| fHLH | - / - | Not met / the evidence-based direction is that there might be some benefit in early treatment; however, the quality and volume of the evidence is low because of the definition of early vs late, the type of study, the number of participants, and the number of studies therefore, there is insufficient evidence to clearly judge the effect. | Y / GeneReviews<sup>115</sup> |
| MCADD | 3 / B | Not met / the evidence-based direction is that there might be some benefit in early treatment; however, the quality of the evidence is low because of the definition of early vs late, the type of study, and the number of participants, there is insufficient evidence to clearly judge the effect. While MCADD is on the current NBS screening panel, there is no indication that the decision was based on a strong evidence base. | Y / GeneReviews<sup>115</sup> |
| hRB | 3 / B | Not met / the evidence-based direction is that there might be some benefit in early treatment; however, the quality and volume of the evidence is low because of the definition of early vs late, the type of study, the number of participants, and the number of studies therefore, there is insufficient evidence to clearly judge the effect. | Y / GeneReviews<sup>115</sup> |
| XLHR | 2 / C | Not met / the evidence-based direction is that there might be some benefit in early treatment; however, the quality and volume of the evidence is low because of the definition of early vs late, the type of study, the number of participants, and the number of studies therefore, there is insufficient evidence to clearly judge the effect. | Y / NICE HST8 2018<sup>122</sup> |
|  | Nature of the intervention: How risky, medically burdensome or intensive is the given intervention? | 10. There should be agreed evidence-based policies covering which individuals should be offered interventions and the appropriate intervention to be offered. | D) Conditions screened for are only those for which the interventions are equitably accessible for all. |
| PDE | 3/ - | Not met / Published consensus guidelines state: "All newborns with PDE-*ALDH7A1* should be treated with 100 mg/day of pyridoxine supplementation." Not mentioning asymptomatic treatment specifically.<sup>123</sup> Clinical advice was that pyridoxine should not be given asymptotically due to reports of toxicity, it is unclear what common practice is. | Y / GeneReviews<sup>115</sup> |
| fHLH | - / - | Met / Clinical advice was: "I think most physicians would agree with pre-emptive HSCT for Perforin, Syntaxin, MUNC13-4 and MUCN18-2 deficiency (although even for these disorders there are some patients who follow a mild course). | Y / Clinical Commissioning Policy: Anakinra for Haemophagocytic Lymphohistiocytosis (HLH) for adults and children in all ages<sup>124</sup> |
| MCADD | 3 / - | Met / dietary advice is provided following treatment guidance such as... | Y / BIMDG guidelines<sup>125</sup> |
| hRB | 2 / - | Met / surveillance as offered currently for patients detected through FH | Y / NHS commissioning board NHS standard contract RB service<sup>124</sup> |
| XLHR | 2 / - | Met / Clinical advice was to start patients on Pi/D and published recommendations provide dosage specification for presymptomatic children<br>Not met / burosumab is not licensed for children <1 year of age currently | Y / NICE HST8 2018<sup>126</sup> |
| FINAL RATING |  |  |  |
|  | Total Score/final assertion |  | Category**** |
| PDE | 11 CB/strong actionability | 1/8 criteria considered in review were met, overall rating: not met | 1 |
| fHLH | -/- | 2/8 criteria considered in review were met, overall rating: not met | 1 |
| MCADD | 11 NB /strong actionability | 2/8 criteria considered in review were met, overall rating: not met | 1 |
| hRB | 10 CB / strong actionability | 2/8 criteria considered in review were met, overall rating: not met | 1 |
| XLHR | 9 CC/ assertion pending | 2/8 criteria considered in review were met, overall rating: not met | 1 |

\*Score between 0 and 3 (3 is best for actionability), sum of scores gives total score
\*\*Level of evidence: A substantial, B moderate, C minimal, D poor, N non-systematically identified or expert contributed evidence
\*\*\*The rating should be interpreted as a recommendation and can be changed following stakeholder consultation
\*\*\*\*Out of 4: Category 1: gene/condition appears to satisfy the four principles, Category 2: Unclear whether gene/condition satisfies the four principles - expert input required. Category 3: Gene/condition does not satisfy the four principles and is childhood onset, Category 4: Gene/condition does not meet the four principles and is adult onset.

#### Conclusions and learnings from ClinGen

ClinGen is an excellent and important resource of information on medical actionability for incidental findings of monogenic diseases. It produces reports separately for the paediatric context, uses a transparent and standardised approach of evidence collection and a semi-quantitative metric to score the actionability of diseases across four domains. An expert group reviews the decisions and the evidence and formulates an overall assertion on actionability. The process allows an early rule out for conditions that do not meet the evidence threshold. While this is a rich source of information and potentially a good starting point for discussion around reporting of genetic findings, the resource has got some limitations as an evidence source for the UK NSC:

1. The protocol that the paediatric actionability working groups follow is a generic one and there is no information on what each group did in their evidence review and how consistent the approach was across groups.
2. The focus is on individuals with incidental findings rather than the public health perspective for a screening programme decision
3. The reports tend to be quite dated and would require updating considering the fast-moving field of genetics.
4. The evidence bar for actionability is lower than what the UK NSC requires in terms of quantity and quality of evidence.
5. Knowledge of penetrance is not a requirement for actionability as early rule out is not triggered in the absence of information on penetrance.
6. Treatment effectiveness is judged by the availability of a treatment guideline with an existing intervention for an undiagnosed paediatric population without the requirement of evidence on benefits of early versus late treatment.
7. The number of conditions covered is limited as of March 2024 and no information on variant pathogenicity was available for any of the five conditions of interest.

Considering the relatively low evidence requirement, it may be feasible to explore using ClinGen as a tool to rule out conditions for reporting that did not meet the requirements for full review, but it is inappropriate for the UK NSC to base decisions on potential screening programs on the actionability reported in ClinGen without further assessment.

### 3 Using existing genomic studies of paediatric screening cohorts reporting penetrance as an evidence source

The purpose of this review was to explore the feasibility of identifying pathogenic variants of paediatric conditions that are known to have high penetrance and expressivity in an unselected paediatric population to be considered for an Initial newborn screening programme that maximises benefit and minimises harm. The review focused on studies using sequencing in newborns for childhood onset disease that reported penetrance or an approximation.

Out of 4970 articles identified, 105 were taken through to full text sifting. The majority were excluded because the test did not meet the inclusion criteria (indirect sequencing or sequencing was second- or third-line test), or the outcome was irrelevant (focus on carrier frequency). See Appendix 4, Figure 7 for the study flow diagram and Supplement 2 for the list of excluded studies and their reason for exclusion. There were 14 studies that reported experiences with gene sequencing in newborns which were reported in 16 references.^127–142^ Green et al. (2023),^127^ Green et al. (2022)^128^ and Ceyhan-Birsoy et al. (2019)^129^ all report on the BabySeq project cited as Green et al. (2023) from hereon.^127^ Full data extractions are reported in Supplement 4. Four studies sequenced genes for a single condition^139, 142–144^ and 10 studies sequenced genes for 74 or more conditions (Supplement 4).^127, 130–138^ Information on penetrance could be inferred from five of the 10 studies with clinical follow-up after a positive sequencing test.^127, 130, 132–134^ The remaining five studies compared sequencing data to follow-up test results,^131^ conventional newborn screening with clinical review,^135^ categorised sequencing outcomes into levels of penetrance based on existing literature,^137^ or categorised sequencing outcomes into pathogenic/likely pathogenic based on existing classification systems with^136^ or without clinical review.^133^ The five studies with clinical follow-up are summarised in Table 23 and Table 24. The studies originated from two countries, the US (n=2^127, 132^ and China (n=3^130, 133, 134^) While the US studies used WGS,^127, 132^ the Chinese studies used gene panel tests^130, 133, 134^ The number of included genes ranged from 134 to 954 and the number of newborns sequenced ranged from 127 to 29,989. Gene Selection and variant interpretation varied across studies. In 4/5 studies the country’s newborn screening programme acted as a starting point,^130, 132–134^ and 2/3 Chinese studies stated that they considered conditions on the recommended unified screening panel (RUSP) which details the mandatary conditions for newborn screening in the US in addition to their own.^133, 134^ However, the final scope of included conditions differed. Green et al. (2023) had the widest scope including a small number of adult onset conditions.^127^ The level of detail provided on variant Selection also varied. In general, as a minimum, studies reported Consideration of the ACMG guidelines and/or ClinGen/ClinVar for classification of variants from pathogenic to benign. Four of the five studies clearly stated that only pathogenic and likely pathogenic variants were reported.^127, 130,133, 134^

**Figure 7.**
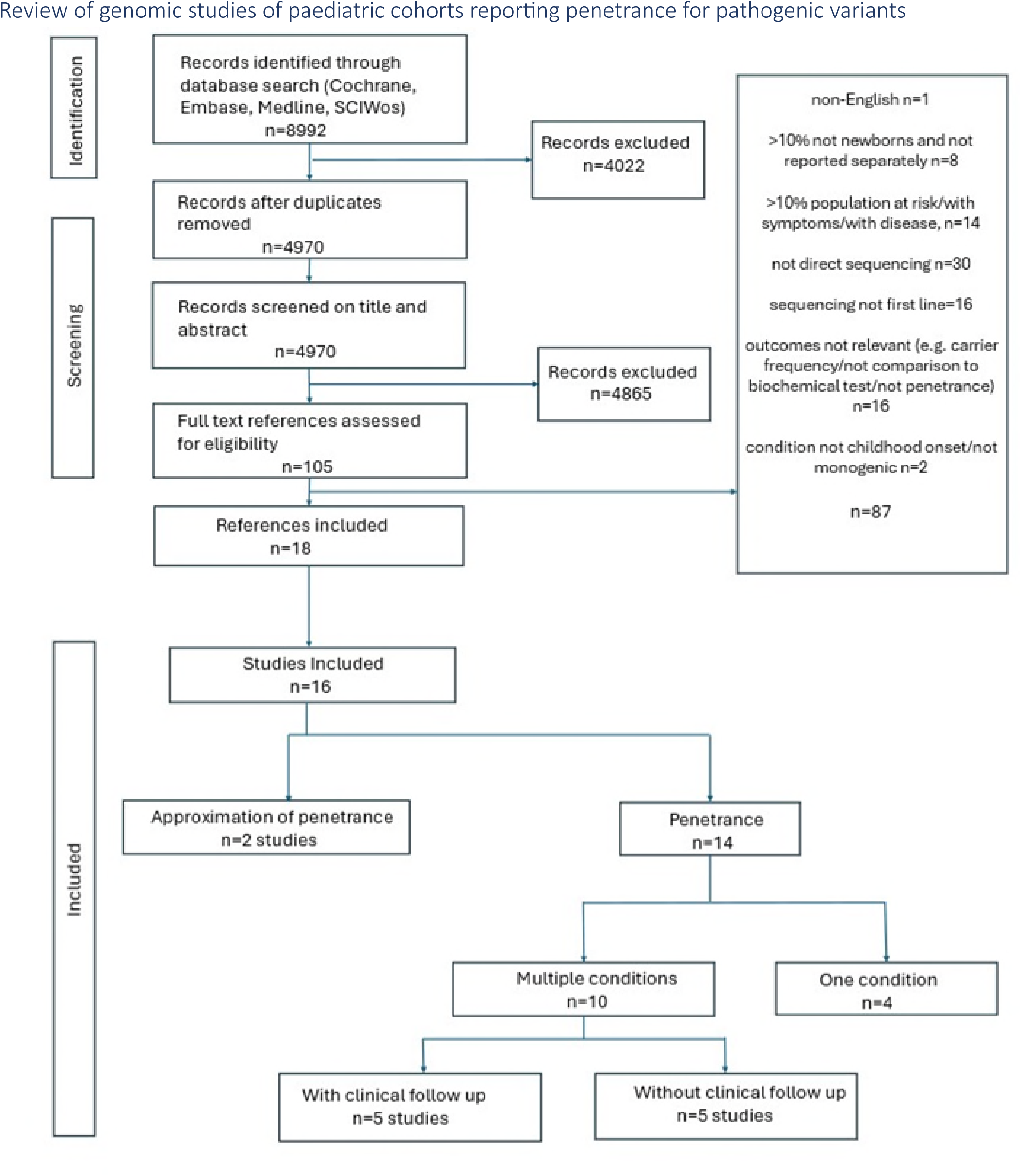
PRISMA flowchart for the review of genomic studies of paediatric cohorts reporting.

**Table 13.**
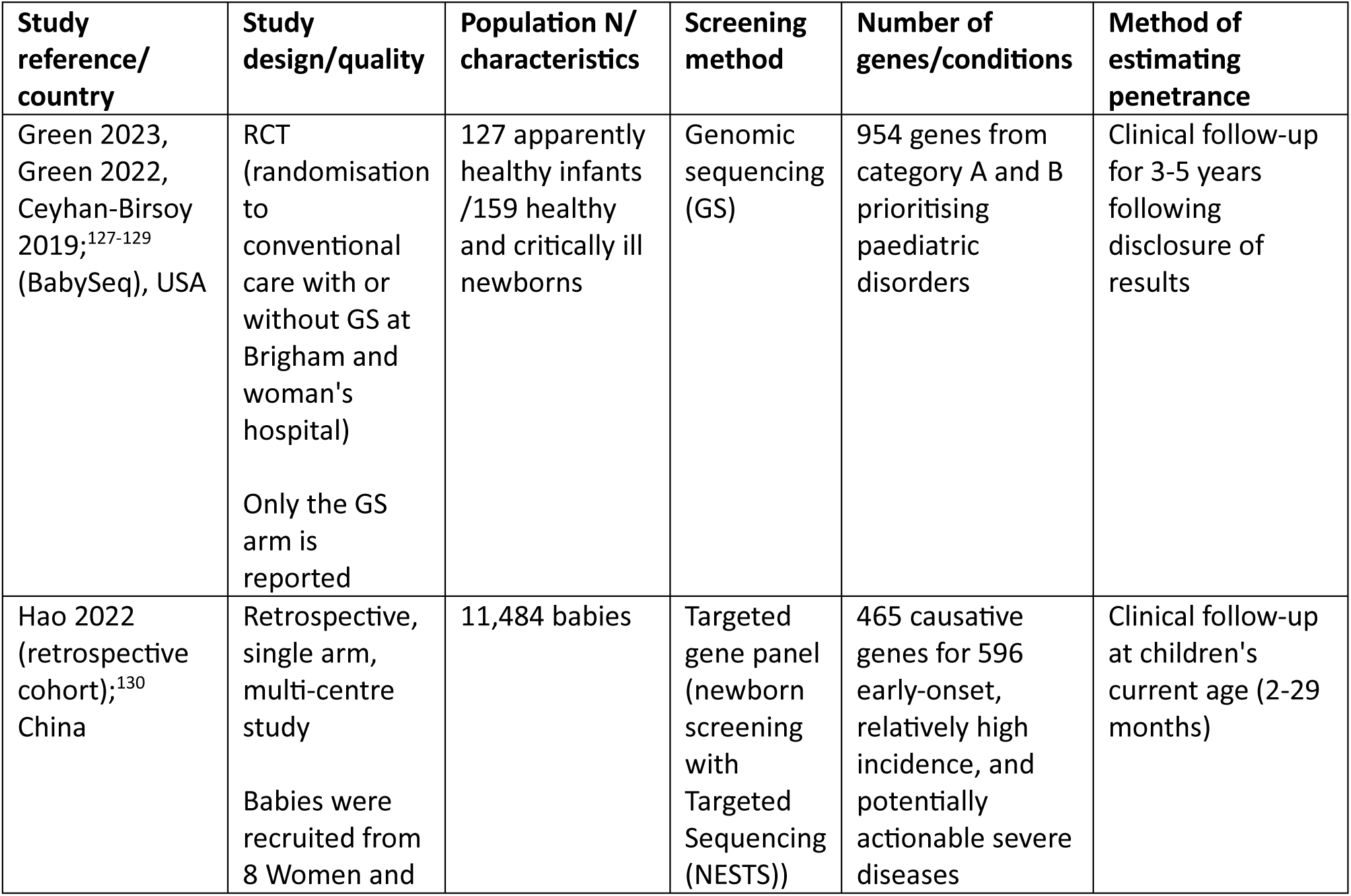
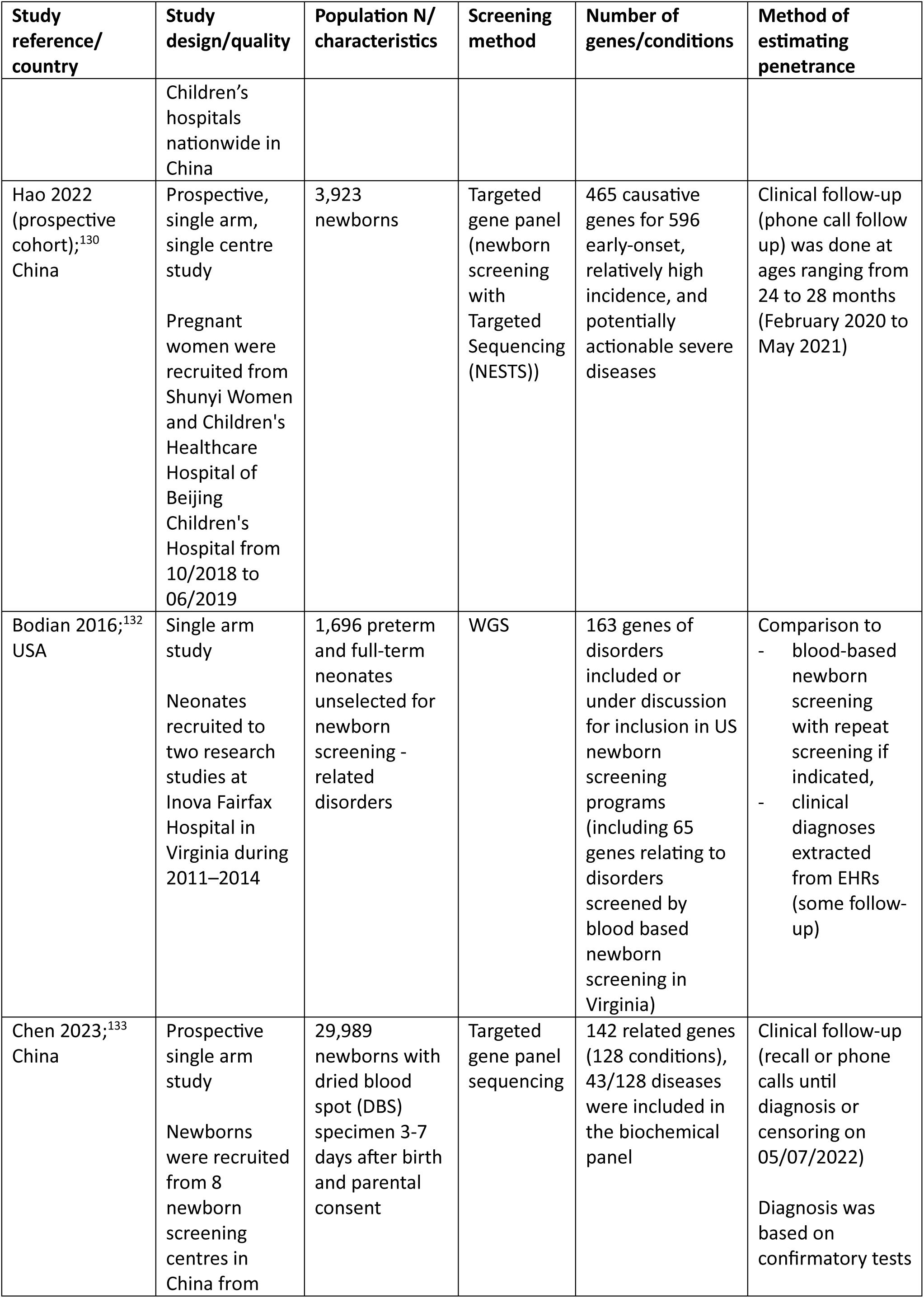
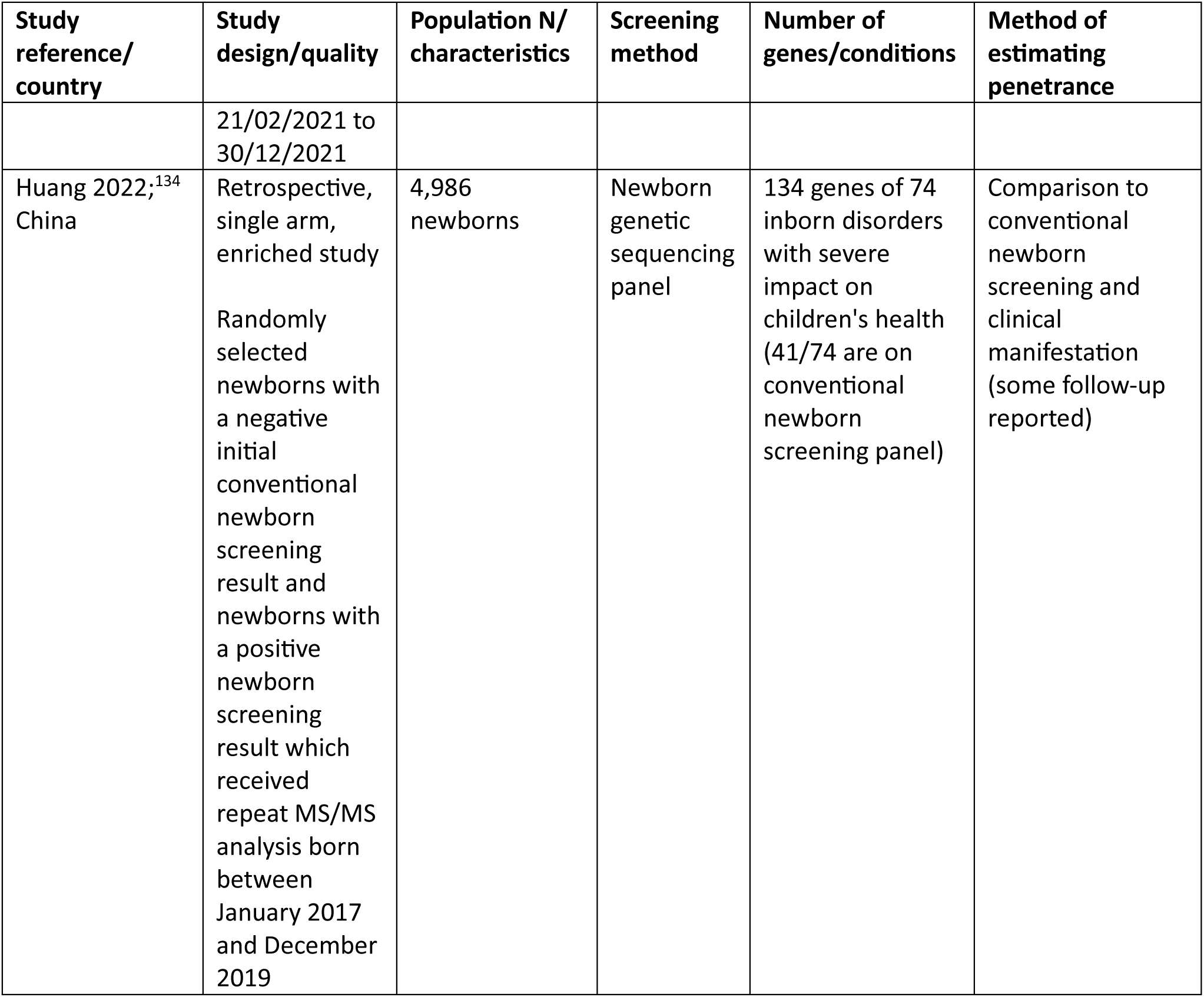
Characteristics of studies reporting results of genetic sequencing of newborns in the screening setting.

The findings of the five studies are reported in Table 14. The proportion of sequencing positive babies ranged from 1.7%.^132^ to 9.7%.^130^ Comparison to conventional newborn screening revealed that genetic screening and conventional screening are complementary each detecting and missing different cases. Genetic screening has the potential to identify a significant proportion of additional conditions that are not on the conventional screening panel in the US and/or China. Two of the three studies with a comparison to conventional screening reported that over half of the positive screens were made up of conditions not on the conventional screening panel.^132, 133^ However, 83.3% to 100% of these could not be confirmed clinically within the studies’ follow-up, so we do not know if detecting these was overdiagnosis of clinically insignificant disease or misdiagnosis of disease or early detection of later onset disease. In Chen et al. (2023), 359/395 (90.9%) of these unconfirmed additional cases were female babies with the X-linked inherited condition Glucose-6-Phosphate Dehydrogenase Deficiency.^133^ This represents an example where sequencing performs differently in males and females.

The great proportion of unconfirmed cases can be partially explained by the short follow-up which ranged within a retrospective study from 2 months to 29 months^130^ and from a median of 1.2years^133^ to >5 years^127^ in the prospective studies. Overall follow-up was insufficiently described, variable within studies and too short to capture disease onset for all included conditions. The short follow-up had an impact on the estimates of penetrance because cases developing clinical symptoms after follow-up will be unaccounted for. This impact would differ for early onset and later onset childhood conditions. Therefore, the Identification of variants in healthy newborns does not exclude a pathogenic role for these variants. Considering the short follow-up, penetrance, approximated by the number of confirmed cases after clinical follow-up for all genes considered, ranged from 1.6% after follow-up of 24-48 months^130^ to 50.4% after a median follow-up of 1.2 years.^133^ While some of the studies were large enough to report a significant number of sequencing positive cases, these large studies did not report findings on clinical outcome by gene variant for all cases to enable an estimate of penetrance on the variant level. One study reported 16 cases of Phenylketonuria, all of which were compound heterozygote with c.158G>A.^134^ All were weakly positive on conventional NBS testing due to slightly elevated PHE levels at regular intervals. However, none of the infants received any intervention, and the study authors concluded that the variant may be causing a mild phenotype. More such data are needed from sufficiently large studies to enable penetrance estimates to be derived for individual variants. However, the majority of variants reported in the studies appear to be single occurrences precluding the estimation of penetrance.

Clinical management was considered where appropriate in all five studies following a confirmed diagnosis including for ‘mild’ and subclinical cases on confirmatory testing. While clinical management in the studies was considered confirmatory of the positive sequencing result, early intervention also means that penetrance cannot be estimated for all cases from these studies. It is unknown whether symptoms would have developed even without treatment in confirmatory testing positive cases.

All five studies were single arm either retrospective or prospective cohorts. One study used an enriched study design, however, this was not reflected in the proportion of sequencing positive cases.^134^ Applicability may be a concern as conditions and variant frequencies vary across geographical regions (even within countries) and 3/5 studies were undertaken in Chinese populations. Sequencing negatives were not followed up apart from one study which reported that 82/445 newborns that received both genetic and conventional screening were missed by targeted gene panel sequencing.^133^ While this information is needed to evaluate test accuracy of WGS, it does not contribute to penetrance estimates (Box 1).

**Table 14.** Summary of findings from studies of genetic sequencing of newborns in the screening setting.

| Study ref | Number of cases detected by sequencing/ total sequenced (%) | Agreement with conventional newborn screening | Additional cases/conditions detected | Information on penetrance |
| --- | --- | --- | --- | --- |
| Green 2023, Green 2022, Ceyhan-Birsoy 2019; <sup>127-129</sup> (BabySeq), US | 10/127 (7.9%) | NA | NA | 2/10 (20%) confirmed on clinical follow-up at 55.5 and 60.1 months, on active management<br>8/10 (80%) unconfirmed on clinical follow-up between 37.6 and 60.3 months (no phenotype as variants predict risk for future disease, 7/8 with predicted moderate penetrance and 1/8 with predicted high penetrance) |
| Hao 2022 (retrospective cohort);<sup>130</sup> China | 902/11,484 (7.9%)<br>488/902 lost to follow-up | NR | NR | 50/414 (12.1%) confirmed on follow-up at age 2-29 months<br>364/414 (87.9%) unconfirmed at age 2-29 months (without obvious related phenotype) |
| Hao 2022 (prospective cohort);<sup>130</sup> China | 381/3,923 (9.7%) | NR | NR | 6/381 (1.6%) confirmed on clinical follow-up at age 24-28 months, 4/6 with active management<br>375/381 (98.4%) unconfirmed on clinical follow-up at age 24-28 months (follow-up may be too short for childhood onset disease) |
| Bodian 2016;<sup>132</sup> US | 33/1,969 (1.7%)<br>(19/33 conventional newborn screening conditions) | 1/19 WGS positive/conventional screening positive (agreement)<br>18/19 WGS positive/conventional screening negative (only detected by sequencing)<br>3 WGS negative/conventional screening positive (missed by sequencing – not included in total of 19) | 14/33 additional conditions not on the newborn screening panel (14/14 unconfirmed) | 2/33 (6.1%) confirmed by clinical diagnosis (1/2 newborn screening negative, affected by condition not medically actionable, healthy without follow-up time specified, 1/2 newborn screening positive, with positive follow-up test and active management, healthy without follow-up time specified)<br>17/33 (51.5%) unconfirmed by clinical follow-up (newborn screening negative, healthy at >1 year of age) |
|  |  |  |  | 14/33 (42.4%) unconfirmed by newborn screening (not on newborn screening panel, no clinical data presented) |
| Chen 2023;<sup>133</sup><br>China | 797/29,989<br>(2.7%) | 445 were positive for conditions on both panels and either positive on conventional newborn screening, genetic screening or both<br>343/445 conditions were positive on both conventional and genetic newborn screening (agreement)<br>20/445 conditions were conventional screening negative/genetic screening positive (only detected by sequencing)<br>82/445 conditions were conventional screening positive / genetic screening negative (missed by sequencing – not included in total of 797) | 434/797 were positive for disorders screened solely by genetic screening (395 unconfirmed) | 402/797 (50.4%) confirmed on follow-up testing and follow-up to median age of 1.2 years (39/402 only detected on WGS, with symptoms and on active management)<br>395/797 (49.6%) unconfirmed by follow-up testing and follow-up to median age of 1.2 years<br>Genetic reason for unconfirmed conditions:<br>359 (X-linked inheritance) females (unconfirmed genetic G6PD)<br>1 male with XXY karyotype (unconfirmed G6PD)<br>1 male with c.1386G>T located on exon 12 which was duplicated (unconfirmed genetic G6PD)<br>1 homozygous deletion of exon 7 in SMN1 but 3 copies of the SMN2 gene (acting as genetic modifier in unconfirmed genetic SMN)<br>33 cases may be attributed to childhood onset or mild phenotypes |
| Huang 2022<sup>134</sup><br>China | 113/ 4,986 (2.3%) | 56/113 conditions were conventional screening positive/ genetic screening positive (agreement, including 20 not confirmed by clinical manifestation)<br>45/113 conditions were conventional screening negative/genetic screening positive (only detected by sequencing) | 12/113 additional conditions not on conventional newborn screening panel (including 10 without clinical manifestation) | 36/113 (31.9%) confirmed by repeat MS/MS and clinical manifestation<br>30/113 (26.5%) confirmed by clinical manifestation (26/30 newborn screening positive /NGS positive; mild phenotype in female patients, 2/30 newborn screening negative/NGS positive, clinical diagnosis and symptom onset at 14 months and 3 years of age, 2/30 not on newborn screening panel, clinical diagnosis, disease onset at 14 months and 3 years of age))<br>47/113 (41.6%) unconfirmed (27/47 not on newborn screening panel, no clinical data or no clinical manifestation, 16/47 newborn screening positive/NGS positive, slightly elevated biochemical markers, no intervention, variant may be causing mild phenotype, 4/47 newborn screening negative /NGS positive, follow-up testing negative) |

#### Conclusions and learnings from existing newborn sequencing studies

In comparison to our traditional review of individual conditions, the focused review of studies reporting penetrance of any gene/condition pair in the newborn screening setting took around four weeks to develop and produced a robust search strategy which can be re-run in the future without Consideration of different conditions. The search picked up all publications from current genomics projects in newborns that were known to us.

Overall, the studies identified highlight that genomic sequencing for newborn screening is still in its infancy and cannot be implemented without further research. There was a lack of consensus on which genes to include, no indication of how to interpret discordant results from NBS programmes and genetic screening, uncertainty over which test is most appropriate for which condition, evidence of overdiagnosis and a large number of cases that could not be confirmed within the studies’ follow-up resulting in interventions, routine surveillance and regular follow-up of uncertain benefit. The studies do not lend themselves to determine a variant threshold for individual genes, i.e. the number and types of variants to include in a screening programme that focuses on detecting highly pathogenic and highly penetrant variants with low risk of producing harm. This is because:

- The number of infants with a specific condition displaying a range of variants is too low even in large studies so variant frequencies cannot be estimated.
- Infants with confirmed genetic disease received management which precludes estimation of penetrance and expressivity for cases without symptomatic confirmation of disease.
- Clinical follow-up was not sufficiently long to include all childhood onset cases.

The studies however give some insight into aspects of WGS not identified from our review of five conditions including challenges and promises of WGS of newborns:

- the potential complementary role of WGS to traditional newborn screening programmes
- levels of agreement with conventional newborn screening
- conditions for which conventional screening may be better (e.g. 4/4 NBS test positive congenital hypothyroidism infants missed by sequencing^134^)
- conditions where regional differences may exist (e.g. G6PD and PKU in China^131^)
- potential differences in PPV between sexes (e.g. G6PD^133^)
- proportion of infants positive on sequencing with two recessive variants where variants are in cis (i.e. in the same copy of the gene leaving the other gene copy intact) (7/55 (12.7%)^133^), therefore highlighting the potential role of parental testing in cases with two heterozygote variants to determine phase
- the impact of including analysis of copy number variations (in one study sequence analysis detected 33/47 (70.2%) at-risk genotypes, CNV analysis contributed additional 14/47 (29.8%)^137^ highlighting potential limitations of WGS)_
- Some indication of turnaround times for WGS (mean turnaround time 1.5 times longer for WGS versus exome gene panel testing (56 vs 37 days),^137^ sequencing to issuing formal report was within 11 days,^130^ turnaround time of newborn WGS 16 weeks to 24 weeks^135^)

In addition to the included studies described above, the review also identified two studies that reported findings that approximate penetrance.^145, 146^ In these studies, variants of a group of childhood onset conditions were identified and the number of healthy adults with disease-associated genotypes with these variants in a general population database were determined.^145, 146^ Gold et al. (2022) concluded that the false positive rate of genomic screening of newborns for diseases treatable with Hematopoietic stem cell transplantation was 0.04% based on 59/141,456 healthy adults with implicated genotypes using variants of 127 genes of severe childhood onset conditions treatable with HSCT.^145^ Breilyn et al (2023) found that clinically relevant variants according to ClinVar in *ACADS* (gene associated with short-chain acyl-CoA dehydrogenase deficiency) were not associated with evidence of metabolic disease in a large and ancestrally diverse adult population (2,035/30,000 healthy adults with implicated genotypes).^146^ These findings could imply that the variants of the 127 genes of severe childhood onset conditions treatable with HSCT may be suitable for newborn screening while variants in *ACADS* investigated may not be suitable. These studies may contribute useful insights on penetrance and expressivity in a future review if the studies meet the following inclusion criteria:

- population database should be of healthy adults rathe than newborns
- variants considered should be of genes associated with childhood onset disease
- outcomes should be number of adults with implicated genotype (not simply allele frequency)
- outcomes should be reported on variant level

### 4. Results for review of cost-effectiveness evaluations of WGS and WES

#### Search Results

Following the searches of databases and registries, the titles and abstracts of 2,325 records were screened, of which 226 records were identified as potentially meeting the eligibility criteria and were flagged for full text review (see PRISMA diagram in Appendix 4, Figure 8). Seventy-one of these studies were judged to meet the eligibility criteria.

**Figure 8.**
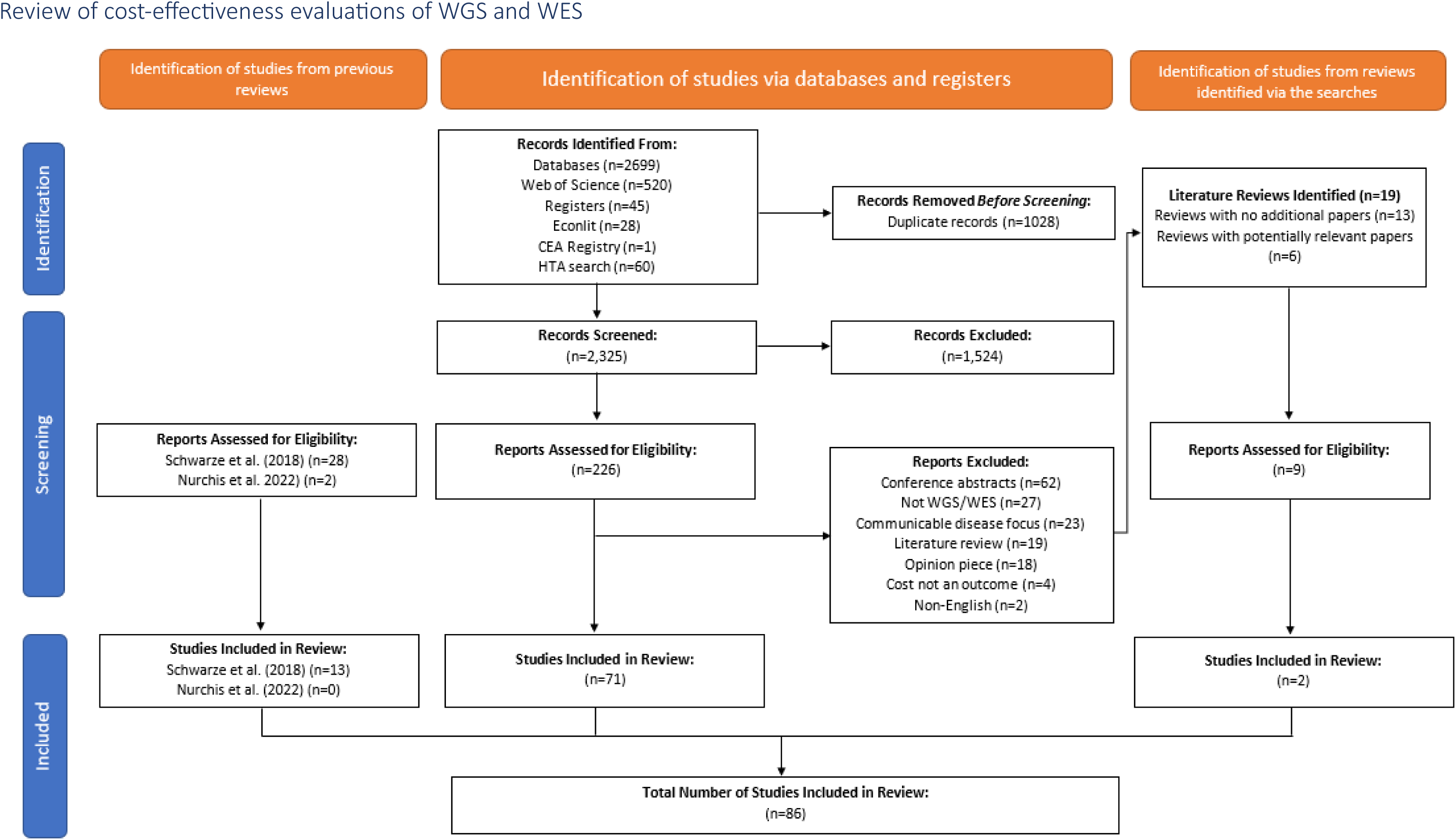
PRISMA flowchart for the review of cost-effectiveness evaluations of WGS and WES.

Nineteen of the records identified at the full text stage process were literature reviews. The reference lists of all nineteen reviews were checked to identify whether there were any potentially relevant records missed by our search or the original Schwarze et al.(2018) review.^147^ All included records in thirteen of the nineteen reviews had already been identified, but there were 6 reviews that included nine additional studies that had not been identified. Of these, two studies were eligible for inclusion.

Of the 28 costing studies included in the Schwarze et al. (2018) systematic review,^147^ 13 met our pre-defined eligibility criteria. Four studies were excluded because they were conference abstracts.^148–151^ One study focused on the use of whole genome sequencing for a communicable disease.^152^ Six studies were included in the Schwarze et al. (2018) review as Partial economic evaluations, but cost was not included as an outcome in the methods or results sections of the studies, cost was just mentioned briefly at some point in the paper.^153–158^ One study was categorised as a Partial economic evaluation, but we could not find any mention of costs in the paper.^159^ A further study was excluded because the focus of the analysis was on the cost associated with identifying incidental findings, and the main analysis excluded the costs associated with WES and WGS.^160^

Two literature reviews were included in the Schwarze et al. (2018) review; we didn’t find any additional papers to assess for eligibility in these reviews.^26, 161^

No additional papers were eligible from the Nurchis et al. (2022) scoping review.^27^

In total, there were 86 studies included in the review.

#### Study Characteristics

#Table 25 provides an overview of the characteristics of the studies included in this review. The full data extraction is included in Supplement 5. Most of the included studies were conducted in high income settings such as Europe (n=27/86, 31%), North America (n=28/86, 33%) or Australia (n=20/86, 23%). Of those conducted in Europe, around half (n=13/27) were from the Netherlands and four from the United Kingdom. The earliest study include in our review was published in 2014, and there has been an increase in the number of studies published on this topic since 2017 (see Figure 1).

#### Population

None of the included studies focused on the use of WGS or WES specifically in a screening context; all studies focused on symptomatic populations or the cost of WGS or WES more generally. Over three-quarters of the included studies included newborns or children in their target population (n=70/86, 81%), reflecting the early onset and presentation for most of the conditions targeted by WES and WGS.

The studies evaluated the use of WES and WGS for a wide range of conditions with a genetic component. The description of the target diseases was very broad for many of the papers, particularly those that focused on newborns or children. Terms including ‘genetic disorders’, ‘monogenic disorders’, ‘variety of conditions’, ‘rare diseases’, ‘mendelian disorders’, and ‘mitochondrial disorders’ were used, demonstrating the broad spectrum of possible genetic diseases potentially discoverable by WES and WGS in contexts where there is very little indication of a clear diagnosis for an individual. There were some common recurring themes such as neurological or neurodevelopmental disorders, intellectual disability or developmental delay (n=20/86, 23%), cancer (n=12/86, 14%), congenital anomalies (n=6/86, 7%), different types of epilepsy (n=4/86, 5%), and autism spectrum disorder (n=3/86, 3%).

#### Intervention

Over half of the studies focused on WES as the intervention (n=48, 56%), with just over a quarter focusing on WGS (n=22/86, 26%) and 16 studies (19%) focusing on both tests. Around a third of the studies explored the impact of putting WES or WGS testing earlier or later in the diagnostic pathway (n=29/86, 34%). In some of these studies, WES was already an established part of the diagnostic pathway under evaluation, and the question focused on whether it should be moved to later in the pathway (i.e. more targeted screening to reduce costs) or moved to earlier in the pathway to reduce the need for other diagnostic tests and potentially arrive at a diagnosis earlier.

There were also some studies (n=10/86, 12%) which focused specifically on rapid WES or WGS, which is different to standard sequencing in that it has a much shorter turnaround time, potentially increasing the clinical utility of the test by returning actionable results quicker. All these studies focused on children and newborns, and were looking for rare diseases, suspected genetic diseases or ‘diseases of an unknown cause’.

**Table 25.**
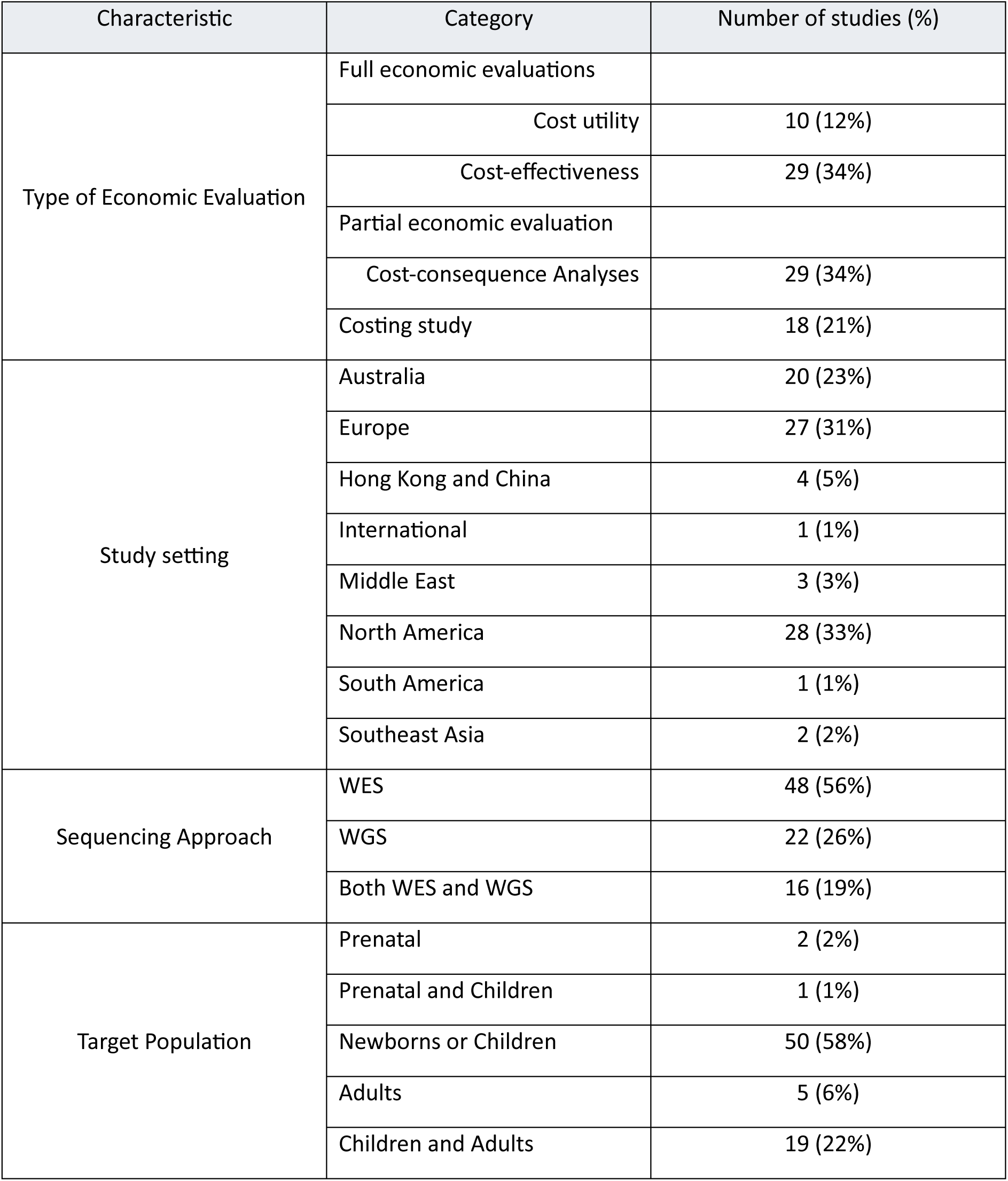
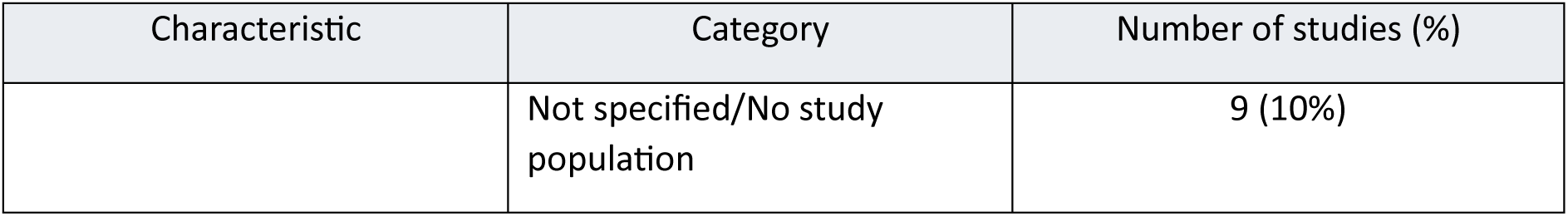
Characteristics of included studies.

| Characteristic | Category | Number of studies (%) |
| --- | --- | --- |
| Type of Economic Evaluation | Full economic evaluations |  |
|  | Cost utility | 10 (12%) |
|  | Cost-effectiveness | 29 (34%) |
|  | Partial economic evaluation |  |
|  | Cost-consequence Analyses | 29 (34%) |
|  | Costing study | 18 (21%) |
| Study setting | Australia | 20 (23%) |
|  | Europe | 27 (31%) |
|  | Hong Kong and China | 4 (5%) |
|  | International | 1 (1%) |
|  | Middle East | 3 (3%) |
|  | North America | 28 (33%) |
|  | South America | 1 (1%) |
|  | Southeast Asia | 2 (2%) |
| Sequencing Approach | WES | 48 (56%) |
|  | WGS | 22 (26%) |
|  | Both WES and WGS | 16 (19%) |
| Target Population | Prenatal | 2 (2%) |
|  | Prenatal and Children | 1 (1%) |
|  | Newborns or Children | 50 (58%) |
|  | Adults | 5 (6%) |
|  | Children and Adults | 19 (22%) |
|  | Not specified/No study population | 9 (10%) |

#### Comparator(s)

Of the studies that included a comparator (n=78/86, 91%), 44 (56%) explicitly stated that the comparator was current standard of care testing. This typically consisted of a broad range of tests, some focusing specifically on more targeted genetic testing, and others also encompassing a wide range of imaging, biochemical, biopsy tests. In some studies, these were very clearly broken down by individual test. Different assumptions were made in terms of which tests would no longer be needed following the incorporation of WES or WGS in the diagnostic pathway, typically dependent on the proposed timing of the different tests.

Many of these studies used cohort data to underpin their analyses (n=58/78, 74%), capturing the healthcare resource use and, where applicable, outcomes for the cohort. Of these, over two-thirds (n=39/58, 67%) used the same cohort of individuals in inform both their intervention and comparator(s). These studies relied on a single cohort who had received either the intervention or comparator, making assumptions about what would have happened to those individuals had they received the opposite. For example, the cohort may have received current standard of care and assumptions were made about what would have happened (usually which diagnostic tests could have been avoided) had the intervention been available. Some studies (n=18, 23%) had two distinct cohorts where one had received the intervention and one had received the comparator. Sixteen studies were modelling exercises, so the cohorts for the intervention and comparator(s) had been simulated, based on aggregate data obtained from the literature.

#### Outcome

Of the 86 studies included in this review, only 10 were cost utility studies and defined the outcome of their analysis as cost per quality-adjusted life year. Over a third of the included studies were cost-effectiveness evaluations (n=29/86, 34%) i.e. they reported the cost for a change in a specific health outcome. All of these studies focused on the number of diagnoses, additional diagnoses or diagnostic yield as the health outcome of interest. The costing studies included (n=18, 21%) tended to report either the total cost per patient or the cost per test.

#### Methodology

The costing perspective was not specified in 33 of the included studies (38%). Where it was reported, many adopted a broad health care system perspective (n=29/86, 36%) or a more specific health system perspective such as a hospital perspective, a clinical genetics service perspective or a laboratory perspective (n=15). Some studies considered costs to the patient or payer (n=8) or a broader, societal perspective (n=5). The costing perspective was largely driven by whether the cost of WES or WGS testing would fall to a public healthcare system or to the individual.

Over half of the included studies (n=50, 58%) didn’t state the time horizon of their analyses. Only 7 studies (8%) adopted a lifetime horizon, with 8 (9%) studies including costs over a 1-year time period or less, typically to only capture costs associated with the diagnostic testing pathway. Where time horizons of more than one year were explicitly implemented (n=24, 28%), seventeen studies (71%) stated that they applied a discount rate to costs beyond the first year.

Only two-thirds of the included studies (n=54/86, 63%) conducted some sort of uncertainty analyses, be it scenario or sensitivity analyses.

##### Cost utility studies

Cost utility studies, i.e. economic evaluations which produce estimates of the incremental cost per quality-adjusted life year (QALY), are typically the preferred economic evaluation method for health technology assessment in the UK. Of the 86 studies included in this review, only 10 were cost utility studies. All of these studies have been published since 2019. Three of these evaluations focused on individuals with non-small cell lung cancer.^162–164^ One study focused on children with prenatally diagnosed nonimmune hydrops fetalis and fetal effusions.^165^ One study focused on adults and children with suspected monogenic kidney disease.^166^

Crawford et al. (2021) evaluated the cost-effectiveness of early exome sequencing in Critically ill babies who are admitted to neonatal intensive care units and are suspected to have mitochondrial disorders compared to current typical care.^167^ They used a decision tree–Markov hybrid model to estimate costs and QALYs over a lifetime horizon. The model was populated using data from the published literature, expert opinion, and the Paediatric Health Information System database in the US. The model simplified the health outcome to two broad health states: within the NICU and post- NICU. Utilities from parents with a child in the ICU were used as a proxy for the utility of a newborn with severe mitochondrial disease in a level III/IV NICU.

Sanford Kobayashi et al. (2022) evaluated the cost-effectiveness of rapid WGS in Critically ill children admitted to paediatric (not neonatal) intensive care units.^168^ Their analysis was based on a cohort of 38 children ranging from 4 months to 17 years old, where rapid WGS had resulted in a molecular diagnosis in 17 of the 38 children. Eight of these 17 children had a change in clinical care, and QALY savings were estimated by Delphi Consensus for two of these children. One of these children had autoimmune polyendocrinopathy syndrome and vaccination for encapsulated organisms was assumed to reduce the risk of mortality. The other child was diagnosed with Factor XIIIA deficiency, initiation of prophylactic Factor XIII replacement decreased the risk of repeat central nervous system bleeds and associated mortality and neurologic complications. Cost savings were estimated for 4 of the 8 children based on the change in clinical management.

Stark et al. (2019) report a cost utility analysis based on the same cohort of individuals used to underpin a previously published cost-effectiveness analysis also included in our review.^169^ They collected data from a cohort of 80 infants aged 0-2 presenting with multiple congenital abnormalities and dysmorphic features, or other features strongly suggestive of monogenic disorders, from a single tertiary paediatric centre in Australia.^170^ In their cost utility analysis, they analyse cost-effectiveness of informative and uninformative results on continuing diagnostic investigation, changes in management, cascade testing in first-degree relatives, and parental reproductive planning and outcomes. The time horizon over which costs and outcomes are estimated is unclear, but the longest time frame mentioned is 18 months. Schofield et al. (2019) build on this analysis, adopting a 20-year time horizon, which the authors justify as being adequate for examining the long-term cost-effectiveness of WES without projecting beyond reasonable certainty in the outcomes of rare genetic diseases.^171^ Across both studies, health utility values were assigned based on parent-reported preferences for health states using utility values derived another study.

Lavelle et al. (2022) evaluated the use of WGS and WES in children with suspected genetic conditions.^172^ They modelled two separate patient populations, infants (<1 year old) who are Critically ill and children (aged <18 years) who were not Critically ill but with suspected genetic conditions. They conducted two analyses, one which produced estimates of cost per additional diagnosis and another which produced cost per quality-adjusted life year (QALY) over a lifetime horizon. To achieve the latter analysis, they made a range of assumptions about the proportion of children who receive a change in clinical management following a diagnosis and the proportion who improve following this change in management. Children were assigned costs and QALYs based on their assumed level of long-term disability (none, mild, moderate, and severe). The parameters for the model were taken from the literature.

##### Cost-effectiveness evaluations

Over a third of the included studies were cost-effectiveness evaluations (n=29/86, 34%) i.e. they reported the cost for a change in a specific health outcome. Twelve of these studies did not report the time horizon over which costs and outcomes were estimated. Seven studies estimated costs and outcomes over 3 years or less, or the time horizon was described as the diagnostic trajectory. Most studies focused only on costs associated with arriving at a diagnosis. Two studies had a lifetime horizon.^173, 174^ These two studies were led by the same researchers in Italy and both explored the cost-effectiveness of WGS versus WES and standard testing in paediatric patients with suspected genetic disorders. The key difference between the studies was that one used a decision tree modelling approach, and the later study used a Markov model approach. Both models were developed using the same data from a cohort of 870 paediatric patients who underwent testing in Rome. Clinical effectiveness was measured in terms of number of diagnoses. Costs included diagnostic tests as well as management and therapeutic procedures. The authors explain that, although a cost utility analysis is the preferred economic evaluation method in Italy, there was no follow-up data for the cohort and therefore QALY impacts from change in clinical management were not available. Both studies concluded that WGS sequencing would be cost-effective compared to WES and conventional testing.

##### Cost-consequence analyses

Around a third of the included studies (n=29/86, 34%) were cost-consequence analyses i.e. they reported cost and health outcomes separately and did not compare results to a cost-effectiveness decision threshold based on the acceptable amount of incremental cost per improvement in a health outcome. Most of these studies also focused on outcomes relating to the number of diagnoses, but some also focused on outcomes such as time to diagnosis and change in clinical management.

##### Costing studies

Eighteen were costing studies and did not report health outcomes. Nine of these studies were micro-costing studies where each individual resource use associated with conducting WGS or WES has been measured and reported. These types of studies are particularly useful in the absence of a standard tariff cost for a test in public healthcare settings. One of these studies was conducted as part of the Scottish Genomes Partnership (SGP) study, where each step of conducting a trio-based WGS is described from clinical assessment and recruitment to participant feedback of the WGS result. They identified which section of the standard genetic testing pathway WGS is intended to replace, a section of the pathway estimated to cost £1841 (2018 prices) on average. This cost varied considerably by and within disease, however, and the authors provide a useful breakdown of the costs by condition. The total cost of WGS was estimated to be £6625 per trio, of which the sequencing itself. There was one other UK-based micro-costing study, Schwarze et al. (2018), who estimated the cost of Illumina-based whole genome sequencing in a UK National Health Service laboratory for a cancer case and a rare disease trio case.^26^ Cost data were collected for all steps in the sequencing pathway and sensitivity analyses were conducted to identify key cost drivers. The estimated average cost for a rare disease WGS trio was £7050.11 (2016 values), with sequencing alone costing £4659 per case and bioinformatics and reporting costing £677. Interestingly, the cost for WGS for a cancer case and a rare disease case were quite similar, and the small difference was largely because the cancer case was based on two samples (tumour and germline), whereas the rare disease case was a trio (parents and proband).

#### Conclusions

In summary, none of the studies identified evaluated WGS or WES in a screening context and, even when looking at WGS or WES in other clinical contexts, only 10 of the 86 included studies in the review were cost utility analyses which is the preferred outcome for UK NSC cost-effectiveness evaluations. The majority of the included studies focused their time horizon or analyses solely on the diagnostic process, rather than incorporating any changes in patient management and the downstream cost and health consequences of these changes. Our review did identify some valuable micro-costing studies, which provide a detailed breakdown of the exact resource use required for WGS testing. These are likely to be useful for any future economic evaluations of WGS for newborn screening, however there is likely to be additional infrastructure requirements to facilitate WGS testing at this scale to consider, and possibly some economies of scale.

### 5. The public voice on evaluating whole genome sequencing

#### PPIE Group reflections on the topic-discussions

At the outset, the group largely supported WGS for newborn screening. There was some diversity of views underpinning this in terms of the way that participants viewed potential harms and benefits, but overall, the benefits were seen to outweigh any harms. As meetings progressed and the complexities were explored, views became more nuanced, for example, one participant mentioned that they now were “sitting on the fence a bit”. By the final meeting, there was still a sense of general positivity around the potential for WGS in newborns, but the group also articulated as many harms as benefits in the whiteboard exercise, illustrating their considered approach to balancing harms and benefits. Benefits identified included saving lives, improving outcomes and quality of life, a reduction in unnecessary and potentially invasive testing while searching for a diagnosis, information for future reproductive choices, and enabling vigilance for the onset of symptoms. Harms identified included parental anxiety and unnecessary worry, the possibility of uncertainty rather than clarity resulting from test results, concerns around education of what screening results mean (both among the public and health professionals), questions of who owns the data and who they are shared with, the strain on health and support services, and concerns around whether doctors will treat children as aggressively if the condition is thought to have a poor prognosis.

Reduction of harms focussed on provision of adequate support for parents and children (and children transitioning to adulthood) including emotional support, clear clinical pathways for diagnosis and treatment for babies identified as having a ‘condition suspected’ result, along with education on what screening results mean (including the possibility of uncertainty and the difference between a screening result and a diagnosis). Several group members reflected on poor experiences with receiving genetic diagnoses that led to harm, and in some cases trauma, and they expressed the view that these issues with the current system must be addressed before any expansion of screening. Under the headings of ‘potential harms’ and ‘what should happen when the evidence is not available?’) several group members mentioned the option of targeted screening being a preferable approach, where results are only disclosed if they meet a threshold of confidence in the result. In these circumstances, it was suggested that the other data gathered could be used for research, but that families should be notified if/when more became known about any uncertain findings.

#### Evaluation of the PPIE process

All participants engaged and participated in the process; they freely shared their views and experiences and openly listened to, and respected, the views of others. Participants responded that taking part was a positive experience. Some found it difficult to attend meetings due to other commitments (including caring commitments) and this led to one person withdrawing from the group. It also meant that at least some of the group were missing from every meeting.

Reflecting on the process, one area that will require careful consideration for future iterations is the planning of the timescale for PPIE. Recruitment of PPIE representatives took several weeks, which meant that the time available for meetings became condensed. With a largely established PPIE group, this would be less of a concern for future reviews involving PPIE, but if this process were to be initiated again more time should be allowed for recruitment and introductions to key concepts. The process would have benefited from having more time to develop and discuss ideas. A lot of complex concepts were discussed over a short timeframe and, despite the group having a relatively high level of familiarity with genetics,some participants commented they would have liked more time to continue discussing particularly challenging topic areas. When establishing a new PPIE group, it is important that participants are given time to share their stories and build rapport with other group members (and the team), which needs to be built into the process. The group built rapport quickly, helped by their sense of shared experiences, as most were parents to a child (or adult child) with a rare genetic condition – although their journeys through these experiences (and the conditions their children lived with) were very different. With PPIE groups with less shared experience, early rapport building activities and careful chairing of the group are likely to be required.

Producing the documentation for pre-reading also required time and careful consideration to pitch the information at the correct level whilst also presenting a broad and balanced overview of the topics. The importance of not over-burdening participants also had to be taken into account. Most of the documentation provided to the group was ‘bespoke’, and summarised information from the literature to ensure it was at the right level. This was also necessary to ensure that participants were not overwhelmed, and reading could be done within the hour allowed for meeting preparation. Again, for future iterations the importance of carefully curating the materials provided to the group should not be underestimated.

The evaluation questionnaires were a useful source of evaluation data on the process, but also proved helpful to allow group members to express views they may not have had the time for, or felt comfortable to express, in meetings. The comments on the forms (which included questions on whether there was anything that surprised them, or particularly stuck in their minds following a meeting) gave an insight into how their views were developing over time.

Comments from the group on the integration of PPIE into future reviews during the whiteboard exercise included the widening of perspectives to include other stakeholders, such as more charity representation, adults with later onset conditions and children and young adults living with rare conditions. Some also commented on the inclusion of medical professionals, ethicists and representatives from commercial entities (insurance companies, pharmaceutical companies) in debates about particular topics. It was felt that the inclusion of members of the general public, while important, could be challenging in terms of recruitment and finding people with interest in this area, and that they would need more background information and training in the subject area.

## Discussion

### Main findings

The five traditional reviews yielded little of the evidence necessary for decision making by the UK NSC. This is belied by the large scale of the five reviews which took seven months for Identification, Selection and review of published studies and comprised sifting nearly 20000 titles and abstracts and 1348 full texts. The 221 included studies addressed only two of the six review questions. No studies addressed the questions on penetrance or test accuracy. No studies addressed the questions on clinical effectiveness and harms of genetic screening. In fact, we did not identify any studies that reported on WGS in the newborn screening setting for any of the five conditions. Between one and nine studies across each of the five reviews reported observations on early versus late treatment where ‘early’ tended to only approximate screen detected and no study was designed to compare treatment in screen detected versus symptomatically detected children. There was some indication that earlier is better across all conditions, but studies were observational, small and we had concerns over their quality mainly around reporting and insufficiently clear definitions of disease.

Between 26 and 89 studies across the five reviews reported the genetic make-up of patients with suspected or confirmed disease for the five conditions. These studies were of interest for two reasons. Firstly, we wanted to compare the variant spectrum in children with clinical disease with that in asymptomatic children detected by genetic screening to gauge how applicable variant information from children with disease is to the screening context. The example of MCADD illustrated that the variant spectrum and consequently the variant frequency shifts when moving from sequencing symptomatic children to sequencing those with biochemical risk factors (positive NBS test) suggesting that studies in children with disease have limited applicability to screening.^175^ However, no studies were identified that used sequencing in unselected newborns. We were, therefore, unable to compare the variant spectrum in unselected and symptomatic newborns. This means that the review provided no information on variant frequency, variant interpretation or penetrance in screening populations or on how much this differs from populations with confirmed disease (additional to the shift from symptomatic to biochemical disease).

Secondly, we wanted to determine the proportion of children with disease that can be detected by sequencing to assess how many patients may be missed by newborn sequencing. This was complicated by the fact that the studies included different populations of patients illustrating that disease can be defined in different ways. Before we can determine the detection rate of WGS in screening (i.e. the proportion of patients with disease identified through sequencing), a clear definition of disease is needed. This is not straighfforward as disease can be defined symptomatically or biochemically using various thresholds. Many diseases have similar clinical and biochemical characteristics and can only be diagnosed genetically. For instance, determining the proportion of children with XLHR caused by variants in the *PHEX* gene is complicated by the fact that XLHR has no disease specific symptoms or biochemical markers. Defining XLHR symptomatically includes additional conditions cause by different genes which affects the proportion detectable by sequencing the *PHEX* gene. This illustrates that measuring the detection rate of WGS in this way is impractical.

Our review identified other challenges for WGS for the screening of newborns. Firstly, the type of test had an impact on genetic outcomes reported. This included for instance the scope of sequencing, i.e. the number of exons covered and whether intron/exon boundaries were considered, the use of additional tests (e.g. MLPA to detect deletions and insertions) as studies generally used exhaustive genetic testing to identify genetic causes in clinically affected patients, and year of testing (since there is rapid change in test development with addition of genes and variants over time). This highlights potential concerns over the applicability of some of the study findings to WGS in the screening setting and limitations of WGS as a standalone test. Secondly, ethnicity had an impact on the number and type of variants identified.^175^ This limits the generalisability of some study findings to the UK context and highlights the importance of considering an ethnically diverse population in the evaluation of WGS for newborn screening and an evaluation of the potential impact of WGS on inequity and inequality in screening. Thirdly, we excluded several studies that focused on specific subgroups of patients. While unrepresentative to the review question they identified particular challenges. For instance, patients with mosaicism are not identifiable by WGS, patients with digenic disease would be classified as negative and patients with atypical symptoms may not be identified if knowledge of their variants is limited. Finally, PDE and XLHR had a high number of novel and private variants for which pathogenicity can be difficult to ascertain. These and conditions with similarly high numbers of private variants may pose a challenge to WGS in screening because of potentially high numbers of variants of uncertain (or unknown) significance.

ClinGen is a useful resource for information on medical actionability of rare monogenetic diseases. Four of the five conditions reviewed have currently got an actionability report available on ClinGen. When compared to the UK NSC criteria, we identified two main limitations. Firstly, the availability of information on penetrance is not a requirement for inclusion of a condition on ClinGen. Secondly, the benefits of early versus late treatment are not a focus of the ClinGen review. In addition, the evidence bar for a positive inclusion decision is lower than for the UK NSC recommendations. As a result, it would be inappropriate for the UK NSC to base decisions for potential screening programs on the actionability reported in ClinGen without further assessment.

Our review of genomic studies of newborn screening populations provided information on International experiences with whole genome and panel testing for screening and illustrated that genetic screening is still in its infancy. Synthesis of the evidence revealed a lack of consensus on which genes to include, no indication of how to interpret discordant results from NBS programmes and genetic screening, uncertainty over which test is most appropriate for which condition, evidence of overdiagnosis and a large number of cases that could not be confirmed by the studie’s follow-up processes resulting in interventions, routine surveillance and regular follow-up of uncertain benefit. All of these would cause problems if WGS was implemented prematurely. The studies did not provide the information on variant penetrance and expressivity that would be needed to determine a variant threshold for individual genes (number and types of pathogenic variants) to select variants for evaluation for a screening programme that maximises benefits and minimises harm from overdiagnosis. One study reported 31 cases of Phenylketonuria of which 15 were caused by 13 different variants which were all confirmed by clinical presentation. This heterogeneity precluded penetrance estimates to be determined for individual variants even in this large study. The remaining 16 cases were clinically unconfirmed and involved a common variant which was believed to cause mild disease. This may represent one example where study information could be used to exclude a variant from Consideration for reporting.

The three main approaches explored do not offer an immediate solution to the evaluation of WGS for newborn screening in the future. Our reviews highlight many evidence gaps and challenges for implementation which may be a contributing factor to the wide variation between studies in decisions of which variants are reported to parents as potentially clinically significant. We conclude that information on penetrance of pathogenic variants selected for inclusion in genomic studies in the screening context should be one focus of future research efforts.

Our review, unsurprisingly, identified no economic evaluations of WGS in a screening context. All the evaluations focused on individuals with suspected genetic diseases and explored the cost, cost consequences, or cost-effectiveness of WES or WGS compared to standard of care diagnostic testing or to different types of sequencing (rapid vs. non-rapid) or positions in the diagnostic pathway. Many of the studies based their evaluations on data collected from reasonably small (all less than 1,500) cohorts of individuals, either receiving the intervention or current standard of care or both. Most evaluations did not adopt a lifetime horizon for capturing costs and health outcomes, as recommended, and most were limited to outcomes and costs for the diagnostic trajectory rather than including costs and outcomes associated with changes in clinical management and the consequences of this change in management. Very few studies conducted a cost utility analysis due to the difficulties in measuring the QALY impact of any diagnoses or downstream health benefits from treatment changes. Where cost utility studies were developed, broad classifications of health states and assumptions around outcomes were made to assign utility values.

Our update of the Schwarze et al. (2018) systematic review did identify a number of high-quality micro-costing studies where a granular account of all the resource use associated with WGS or WES had been recorded and costed.^26^ These types of studies are incredibly useful as they can be updated as unit costs change over time, and they also help to understand which aspects of the testing pathway is driving the cost. One of the studies identified in the review was published as part of the Scottish Genomes Partnership study and we have been made aware that similar activity is underway but not yet published by Genomics England. We would encourage future UK micro-costing studies to be undertaken, particularly when conducted in a routine clinical practice setting, rather than research setting, to help underpin future UK-based cost-effectiveness evaluations of WGS. It would be useful to have these available across a range of clinical contexts to understand more fully when and why costs may differ.

In this review, standard of care testing which frequently included some sort of genetic testing, was often listed as a comparator. Costs associated with standard of care varied widely between individuals, demonstrating the importance of developing large, heterogenous cohorts of individuals to truly capture the breadth of resource use. Standard of care will be very difficult to define when evaluating WGS in a screening context unless there is clear comparative data on which conditions would have been detected later, when and using which diagnostics. Standard of care in terms of clinical management was rarely included in the economic evaluations in this review, and is likely to add another layer of complexity, especially when trying to elucidate the benefits and harms associated with earlier diagnosis.

### Contribution to existing knowledge

There are no previous systematic reviews of the benefits and harms of WGS of newborns and so our review is novel. We have not identified any other group who is approaching the question from the same perspective. The International Consortium on Newborn Sequencing (ICoNS) is a Consortium of International genomic projects aiming to bring together, exchange and harmonise ideas and efforts to responsibly implement newborn sequencing.^176^ This is a useful source of information, but the focus of the Consortium is implementation rather than evidence synthesis for policy advisors. While the Consortium website portrays certainty of health benefits from newborn sequencing, our review highlights the uncertainties and evidence gaps that could result in harms if WGS is implemented prematurely.

There are currently at least 15 genomic projects underway Internationally aiming to sequence over 400,000 newborns.^176^ The study by Downie et al. (2024) compared gene lists across six of these genomics projects and reported substantial differences in genes lists due to different starting points with some prioritising the clinical validity of the gene disease association while others prioritise treatability.^177^ We have observed similar disagreement in gene lists due to different processes and priorities used across the genomic studies included in our review of penentrance in newborn screening populations. This lack of consensus may reflect uncertainty of what to report to parents as potentially significant which is based on current evidence. The gene Selection process is complex and multifactorial. It requires evidence to assess whether the condition is monogenic, whether the genotype/phenotype link is established by identifying pathogenic variants, and the penetrance and expressivity of these variants. Evidence on pathogenicity, penetrance and expressivity is limited for general population cohorts. Because there is less proof of pathogenicity for likely pathogenic variants there is no consensus on reporting likely pathogenic variants in screening, while VUS are generally not reported. It is important to understand and evaluate the underlying gene and variant Selection processes that underlies the decision what to report to parents and our work on this could be expanded in the future to a full systematic review to understand the different approaches to gene and variant curation and to synthesise commonalities and differences to inform a robust and unified process.

The ClinGen resource represents one open-access effort to develop and implement standardised, evidence-based methods to characterise the clinical actionability of genetic variations. However, our review found that the evidence bar for inclusion is low, the focus around treatment is clinical and therefore on treatability rather than benefits of early versus late treatment and while some aspects are relevant to the screening context (e.g. treatment for presymptomatic children), focus is on secondary findings in individuals in the diagnostic setting where the definition of an important condition may be different to the public health perspective of the screening setting.

### Strengths, challenges and limitations

#### Strengths

To the best of our knowledge this is the first attempt to identify an approach to assess the benefits and harms of WGS for newborn screening for policy advisors. This was a huge systematic review applying both traditional and novel approaches and has the potential to guide policymakers towards a new approach for both synthesising and producing the evidence required to make evidence based policy decisions in this complex area. We consulted widely with policy advisors and clinicians during protocol development to frame the right research questions for this review. We used a stratified random sample of conditions for our traditional review and published our protocol on PROSPERO. We consulted with clinicians specialising in the five disease areas, geneticists, clinical and laboratory advisors to the UK NBS programme and members of the UK NSC throughout the review process to ensure we deliver what is required by decision makers and undertook an extensive piece of work to consider the public and patient view on this ethically challenging topic. While all these aspects ensured a high-quality review, we learned from several challenges that can inform subsequent reviews.

#### Challenges

Reviewing five complex, very different conditions at the same time was challenging. It is impossible to gain the knowledge and insight for five conditions in a similar time frame as we would normally have to review a single condition. We attempted to work as one team for consistency in decision making, but had to focus on 2-3 conditions each. We attempted to use one review approach across all conditions, but quickly recognised that the conditions required individual Considerations. A review of 200 conditions would ideally require 200 review teams or an extensive time for a smaller number of teams to ensure the rigour and subject knowledge can be developed for each condition. Issues around consistency would need to be factored in and addressed if several review teams undertake the evidence syntheses.

The outcomes presented here represent a snapshot of the evidence in this fast-progressing field. Gene and variant lists are constantly changing and developing in light of new evidence. New variants are identified, variant classification changes based on new insights from clinical practice and research. Evaluations and policy decisions of 200 conditions once completed will not be up to date for long and will require a system that allows continuous evaluation.

The review highlights the challenge of defining disease because we were thinking across genetic, biochemical and clinical disease. It is important to be clear about what aspects of disease we would aim to detect and prevent with WGS. This is linked to the challenging concept of penetrance which depends on the definition of disease and at what time point we measure disease. This will determine the required follow-up time of screen-positive newborns for the estimation of penetrance and is likely to be different for different conditions.

A further challenge was the integration of expert clinical advice into the review and this will be a challenge in the future on how to incorporate clinical knowledge into the evidence base to inform a policy decision. Clinical knowledge was important and nuanced and was complementary to the review findings. The issue is that the information comes from individuals, however, in these rare diseases it would be challenging to identify a group of experts large enough to establish a consensus. We were advised that 1) targeted gene panels would be more economical than WGS (and others share that view^178^) but this is not currently a question that is being addressed, 2) meaningful time scales from testing to reporting are unlikely to be achievable in the overstretched NHS where current waiting times for WGS results for the diagnosis of symptomatic children often exceed one year,^179^ 3) storage of huge amounts of data is expensive and may not be accessible in the future, while re-sequencing may be more appropriate than accessing stored data in the future. These are important issues that our current review did not ask or identify from the published evidence. Furthermore, one clinical advisor said that they would never initiate asymptomatic vitamin B6 (pyridoxine) because of treatment induced seizures, yet this is the early intervention of interest here. This was an important finding but was not backed up by published studies. It was impossible for us to know how common this view is.

#### Limitations

Our approach of selecting the five conditions aimed to include a range of scenarios that would allow us to explore different challenges that the NHS would have to face with newborn sequencing. This meant that the conditions, the aim of screening for the conditions, the definitions of early versus late treatment, and their underlying genetics were very different. Consequently, we were unable to use one approach to reviewing as planned and concluded that it is unfeasible to use one review approach for 200 conditions. However, we do not know whether this applies to a group of similar conditions, e.g. metabolic conditions, where learnings from one review may be transferable to other conditions speeding up the review process. Repeating this approach with a group of similar conditions could shed light on this.

The number of studies reporting the genetic spectrum in children with confirmed disease was unexpectedly high for all five conditions. The studies were heterogenous in terms of population, test and outcomes and would have been difficult to synthesise. The most appropriate population with disease to determine the detection rate for WGS was difficult to define for XLHR and fHLH because of non-specific symptoms. We, therefore, decided to categorise the studies by definition of disease and extracted data from the largest study only (or more than one where results were complementary) to present the breadth of the evidence rather than aiming for completeness. The aim was to provide examples of outcomes depending on disease definition that could inform the inclusion and exclusion criteria of a future review for this question. However, this meant that we were unable to draw on all reported findings on variant frequency and expressivity or to highlight aspects where studies may have agreed or disagreed. We are uncertain how useful synthesising all the included studies would have been.

We were unable to undertake a number of explorations detailed in the protocol because the shorffall and type of data meant that they were unfeasible. We wanted to explore reporting of penetrance information from the genomic studies of paediatric screening cohorts 1) by subgroups of conditions predefined by the studies, 2) by the GEL category the conditions would fall into and 3) combined for the conditions with a top quintile ClinGen score. The aim was to explore the feasibility of determining the variant threshold that corresponds with the most severe phenotype for conditions. However, this was not feasible because 1) categories are at condition / gene level and penetrance is reported on variant level, 2) the penetrance information reported in studies was inadequate numerically and qualitatively, 3) the ClinGen score was not reported on variant level and 4) the number of conditions covered in ClinGen was limited. This means that we were unsuccessful in exploring whether a variant threshold for individual conditions could be used to develop a restricted but safe screening programme.

Studies comparing genetic screening with traditional NBS screening suggest that tests are complementary in terms of cases they detect and miss, but they do not tell us anything about the place of WGS in an existing screening program i.e. how both tests could be integrated, how to interpret contradictory outcomes, whether the sequence of tests should be different for different conditions or whether newborn sequencing should be a completely separate programme. In general, the combination of metabolomics alongside genomics, in whichever order, is likely to add to our understanding of genetic variants and their significance and considering all research in a future review will help to identify the best screening strategy for individual conditions.

Furthermore, there are several questions that we did not address or where there was insufficient evidence available from the studies we reviewed, but that are fundamental for the evaluation of newborn genetic screening.

1. The differences in variant frequency and penetrance in screened (unselected) and symptomatic cohorts is important to understand but data are lacking for most conditions. It is widely accepted that risk estimates for genetic disease from high-risk groups do not translate to the general population as has been shown for cancer predisposition genes in individuals with and without family history.^180^ And in MCADD, a condition that is already screened for by NBS screening with genetic confirmatory testing, the shift from symptomatic disease to biochemical disease definition resulted in a different variant spectrum.^116^ In this study, a lower proportion of screen detected newborns were homozygous for the c.985A>G variant which is very common in symptomatic children. A significant number of the newborns had genotypes with variants that had not been observed in patients detected clinically and some, like c.199T>C and c.127G>A, were associated with a milder biochemical phenotype. This knowledge is important for all conditions considered for genetic screening and needs to be extended to understanding the variant spectrum in infants designated screen positive based on genetic testing only.
2. There was insufficient evidence to explore differences in variant frequency and penetrance by sex, geographical region and ethnicity. Variants of uncertain (or unknown) significance are more common in non-European ethnic groups because of an underrepresentation in reference population databases used for variant frequency annotation.^8^ Cosenquently, there is a lack of genotype-phenotype correlations in populations such as the UK and US with significant ethnic diversity. This is a particular problem for whole population screening and means that it will be difficult to establish an equitable screening programme if ongoing genomic studies do not generate the evidence from an ethnically diverse population.
3. The review did not address the question of whether WGS is the best test for expanded newborn screening compared to WES, panel testing, sequencing combined with other tests or biochemical assays. Many conditions could be tested for using biochemical assays or bespoke genetic tests which could be superior to WGS.^179^ WGS has got limitations and is generally more expensive. It is unable to detect large deletions and needs to be adapted for instance to identify copy number diseases like SMA to ensure they are reliably detected. Furthermore, it can also produce inconclusive results as in cystic fibrosis screening.^181^ The gene by gene and exon by exon performance of WGS is improving all the time but this remains a factor at the time of writing.
4. Test failure of WGS was insufficiently reported by the included studies.
5. We did not explore the impact of sequencing results on clinician behaviour and clinical care.
6. We did not investigate workforce challenges which will include the number of genetic counsellors needed, and training of staff to correctly interpret WGS results.
7. We did not identify evidence-based pathways for those with rare asymptomatic genetic disease.
8. More work is needed to explore ethical issues around knowing but not reporting certain variants.
9. More work is needed to explore ethical issues of storing blood samples/genetic information.
10. Existing resource use data associated with WGS needs to be adapted/adjusted to account for the additional infrastructure and staff required to deliver WGS at scale i.e. in the context of a national screening programme.
11. Long-term follow-up on cohorts of, ideally, asymptomatic newborns undergoing WGS is needed to understand the implications of testing on patient management in terms of costs and patient outcomes.

#### Patient and Public Involvement and Engagement

The PPIE process itself was successful and viewed positively by participants, who all expressed a view that they would be interested in future PPIE work. The group started the process with a broadly positive view of the benefits of WGS for newborn screening, but the more they identified and developed their understanding of the potential harms the more cautious many of the group became. This process of increasingly Critical, or ambivalent, views being expressed as information and discussion increase has also been observed in other groups considering complex topics in the field of genomics and screening.^182^ In the final exercise, several expressed the view that a targeted approach to genomic screening may be preferrable to high throughput screens. This was not true of all of the group, however, and some remained largely supportive of WGS despite potential risks or harms, seeing all forms of knowledge as useful. This divergence in opinions has been highlighted in the literature^183^ where for some, non-actionable or uncertain results can be seen as empowering, whereas for others concerns about risk and unnecessary anxiety and stigma were identified. Some parents of children with health conditions demonstrate a greater tolerance for uncertainty from screening because they are already experienced in medical uncertainty.^183^

The way conflicting perspectives on complex topics in genomics and screening should be prioritised and weighted in research outputs and policy decisions is a debated topic within acceptability research.^184^ Further research with collaborative PPIE engagement that can map such diverse views to social characteristics, backgrounds and/or particular lived experiences may prove particularly useful in facilitating a nuanced understanding of the distributions of harms and benefits of genomic screening to inform policy recommendations and implementation. For this reason, it would be beneficial to increase the diversity of viewpoints if this PPIE process were to be repeated. A limitation of our work was that the recruitment strategy did not target participants based on cultural diversity (although there was some), nor did it aim to be reflective of the UK population. This was not achievable given the timescale. Future work should allocate more resources to recruitment and recruit participants from outside of the rare disease community to achieve more representation from members of the public without experience of living with rare conditions. This will be particularly important as most members of the public offered screening will have no, or limited, experience with rare genetic conditions. The group mentioned the potential for difficulty in recruiting members of the public where the relevance of the topic may not be immediately evident to them (especially if they are not new or expectant parents) and the added time that would be needed to educate public participants on the complexities around genetic screening and diagnoses. Familiarity and factual knowledge around genomics among the public is low^185^ and so time would need to be invested in introducing key terms and concepts around genomics, sequencing, and screening to enable meaningful Contribution s.^186^ When this is done, research has shown that previously ‘genetics agnostic’ members of the public can make substantial and rich Contribution s.^187^ Given the backgrounds of members of this group, and their existing familiarity with genetics, this step could be greatly condensed, but future PPIE work should accommodate additional time for information sharing and the incremental building of knowledge and debate, so that a common language for talking about genomic screening can be established.^186^ The group also mentioned the value of bringing in specialist contributors/viewpoints for particular topic areas, such as representatives from the insurance industry.

It is important to allow adequate time and Consideration to the composition and recruitment of future PPI groups, as well as time for new groups to build rapport and trust. It would also be valuable to allow time for the training of participants in PPIE in reviews and give them the resources to co-develop the aims, terms of reference, and timetable for their Contribution to the work (which was not possible in this rapid review setting). Finally, the importance of investing time and Consideration into the development of materials, resources and viewpoints to share with the group ahead of discussions, in an accessible format, should not be underestimated. This is an essential component of creating a deliberative space to allow meaningful Contribution and collaboration between the research team and PPIE contributors.

#### Equality, Diversity and Inclusion

Recruitment was targeted to people with experience with rare conditions (adults living with a rare condition, parents of children with rare genetic conditions), and advocates working in the area of rare genetic conditions. We were successful in recruiting people with a broad range of experience with genetic conditions. None of the group members with lived experience of rare genetic conditions (parents, adult living with a rare condition) had experience with the same genetic condition as anyone else in the group. Most of the group had not been involved with research or PPIE previously.

Due to the time constraints on recruitment and the focus on experience with rare conditions, we did not target recruitment by demographic variables (e.g. age, socioeconomic status, geographical location, ethnicity, and so on). Most group members were female, which may reflect the tendency for mothers to become the primary caregivers when they have a child with ‘medical complexities’.^188^ There was diversity in ages of the children they supported (under 5 years to over 20 years) and therefore the stage of caring for / supporting a child with a rare condition. As indicated in the discussion, for future work (and with more time available) we would wish to expand the viewpoints represented, by for example, including more public voices, and ensure diversity of background and experiences, including recruiting from underserved groups.

Much Consideration was given to the amount, format and content of documentation provided to participants to maximise accessibility and their understanding of concepts and complexities (e.g. terms such penetrance and expressivity). This also included producing a lay summary of the draft report for Meeting 4, where the PPIE group had the opportunity to discuss the findings and ask questions of a representative of the review team. Evaluation forms sent after each form checked with the participants that they were happy with the information that had been provided and asked if we could improve on this; feedback for this was entirely positive and all participants expressed an interest in being involved in future work in this area.

#### Impact and learning

This review will inform the UK NSC in their approach to evaluating WGS for newborn screening. The learning from this review is that a traditional review will unlikely be an effective approach for the evaluation of 200 conditions. We propose an alternative pragmatic approach focusing on first addressing the evidence for penetrance in the screening context for variants identified as actionable by current genomics projects like the Generation Study, a Critical question with implications for screening benefits and harms for which appropriate quality evidence is currently not available.

Penetrance may be defined differently for different conditions. A biochemical confirmatory test may be sufficient for some conditions to estimate penetrance if the link between biochemical and clinical disease is well understood and strong, for others a follow-up test may not be sufficiently predictive of clinical disease and for those conditions without an available confirmatory test, follow-up to clinical symptoms is essential. Where there is good quality evidence for high penetrance the evidence synthesis could be expanded to other questions relevant to benefits and harms of screening.

The review may also prompt the UK NSC to discuss evidence requirements for decisions on screening programs for rare and ultra rare genetic conditions. This will be useful as it will re-focus discussion on evidence in relation to WGS for newborn sequencing.

#### Implications for decision makers

Currently there is no evidence supporting large scale implementation of WGS of newborns with concomitant simultaneous detection of many conditions. The cost and the balance of benefit and harm is unknown, and implementation would prevent the research required to measure the benefits and harms (see research recommendations section). Our review of genomic studies of newborn screening cohorts desmonstrates unequivocally that introduction of WGS without substantial further research would cause many problems and uncertainties.

We found that the traditional approach to evidence synthesis cannot be applied to WGS of newborns, the quantity of work is very high, and the data provided is of insufficient quality and therefore value. Evidence from clinically detected cases is often not generalisable to screening, where the spectrum of disease differs, and less clinically significant disease types are more common. Whilst there are many unknowns about the benefits and harms of screening, we advocate addressing the accurate measurement of penetrance first, and filtering review efforts concerning other aspects such as benefit of earlier treatment only in those conditions with variants with sufficient penetrance. This proposal is a pragmatic stepwise approach based on the paucity of data and the need to engage with the broader scientific community to deliver the types of studies which will deliver the required penetrance data (see research recommendations section).

Another possible approach to evidence synthesis is to use existing gene and variant curation by genomics projects. Decisions on which conditions, genes and variants to report are generally based on severity of disease, disease onset, penetrance and expressivity of pathogenic variants, treatability and access to treatment. Gene curation processes vary greatly but are all based on a condition-by-condition approach. Our assessment of the Genomics England approach and ClinGen approach showed that they do not meet the requirements of the UK NSC and are not suitable for adoption. An assessment of apporaches more widely may be needed.

Therefore, what is needed is commissioning of carefully designed research to generate new evidence with a focus on penetrance data in a screening setting for genes previously selected by gene curation processes. Future review efforts can focus on the narrower questions of penetrance and expressivity of pathogenic variants in large screening cohorts first, with additional questions about earlier treatment benefits only in those with sufficient penetrance and expressivity. Onward data collection and monitoring of any future WGS screening programme will be essential to determine immediate and longer term costs, benefits and harms (managed access screening programmes similarly to NICE managed access schemes).

Economic evaluations to date of WGS have focused on symptomatic populations, and have focused predominantly on costs and outcomes associated with the diagnostic process itself. Studies that have attempted to estimate cost per quality-adjusted life years associated with WGS/WES testing, of which there are few, were either focused on very specific heath conditions where it is easier to define distinct health states, or have applied very crude quality of life and survival estimates to extremely broad health states. Creating a model which accounts for all the conditions is highly likely to be unfeasible. It may be possible to group some of the conditions by their biological pathways or clinical manifestations, providing a mechanism for identifying more homogenous health states to which you could apply more precise health-related quality of life, survival and cost estimates. There will inevitably be a high degree of uncertainty associated with any model developed for this evaluation question, driven by an inability to capture the whole decision problem, rather than parameter uncertainty or model structure limitations. Methods to interrogate this type of uncertainty and understand the risk of making the wrong decision (i.e. recommending/not recommending WGS for newborn screening) in this context would be valuable to support policy decision-making.

#### Research recommendations

Whole genome sequencing of newborn babies is a complex intervention with interdependent factors (such as gene selection, variant pathogenicity, penetrance, expressivity, benefits of earlier treatment) contributing to the balance of benefits and harm. There is a paucity of evidence around these factors for the screening context and very low levels of knowledge about the balance of benefit and harm.

We have highlighted many research gaps that span the complete evaluation process of WGS for newborn screening and propose possible research approaches to address these. It is imperative that this reserarch is undertaken as part of large joint-up and possibly International collaborations to produce the evidence that is needed to thoroughly assess the benefits and harms of WGS for newborn screening for rare and ultra rare conditions.

A systematic review of studies reporting gene and variant Selection approaches will help in the understanding of prioritisation and reporting decision of genomic projects. This may aid the formulation of a consensus, best practice gene curation approach for newborn genetic screening and inform the evaluation of Genomic England’s gene list and classification of pathogenicity of included variants as part of the future evaluation of WGS for newborn screening.

We recommend addressing the key question of penetrance in a screening population and produce the evidence needed for an evaluation. The evaluation of the subsequent interdependent factors can then focused on conditions only where there is good evidence of high penetrance for at least some pathogenic variants. Large research studies implementing newborn screening using WGS and reporting what they detect (i.e. diagnostic yield) and offering treatment do not provide the penetrance and expressivty data required for decisions about whether to implement. This is because if you report variants to parents as positive diagnosis in their newborns and offer them treatment, and they go on to have a good health outcome there is no way of knowing whether treatment was curative or whether they would never have had any symptoms or effects anyway and did not require treatment. So you cannot know if they benefitted or were harmed. For example, the Generation study is sequencing 100,000 newborns in England and will report results for variants in over 400 genes to parents, whose babies will receive clinician-guided management.^6^ This treatment provision precludes estimation of penetrance from the study. If studies were to choose to report fewer pathogenic variants, only those where there is existing good evidence that penetrance is high, that would provide the evidence that policymakers need on penetrance of other pathogenic variants (which can be accurately measured through follow-up to symptomatic disease in the absence of diagnosis and treatment). The Generation Study also demonstrates another key challenge in this area, which is that even with a very large sample of 100,000 newborns, power to analyse results by condition or variant is extremely low due to the rarity of most relevant conditions.

To understand penetrance the following combinations of research studies could be employed:

1. The ideal studies would be large cohort studies where either screening with WGS is given to newborn babies, without reporting results to parents or very few conditions of well evidenced penetrance and expressivity are reported to ensure more good than harm. Other potential conditions/genes should not be reported to participants unless they present symptomatically, to measure the penetrance and clinical significance of pathogenic variants and therefore benefits and overdiagnosis harms of revealing the test results. This would enable accumulation of evidence about penetrance for conditions/variants, which could be assessed on an ongoing basis for addition to the study/programme, if there is sufficient evidence of more good than harm. This may generate the evidence needed to allow a gradual or stepped implementation in the future based on levels of evidence on pathogenicity and penetrance/expressivity, burden and cost of available treatment, availability of confirmatory tests and disease onset. However, these studies would raise considerable ethical questions and would only be possible with parents’ complete understanding, agreement and consent. There would be concern that, once parents were asked whether they want to participate in a trial of WGS without telling them what you find, none of them will be in equipoise.
2. The existing large genomic screening cohort studies can produce evidence on penetrance for variants which are not included on their panels for reporting and treatment. This may provide some useful data because of the large variation in gene lists between studies. However, a note of caution should be applied because some of the rationale for different inclusion of variants may be differences between populations, which would mean we would be primarily measuring penetrance for variants that are of lesser importance in that population and penetrance measures may not be generalisable to the UK population. A further complication is that good quality follow-up to confirmed disease is necessary, such as using robust disease registries, which is not always available.
3. Genetic information from healthy adult cohorts such as the UK Biobank and worldwide datasets like GNOMAD could be used to identify low penetrance variants which are not suitable for inclusion on a screening panel. If large numbers of healthy adults have a variant believed to be associated with childhood onset disease, then it will not be a highly penetrant variant suitable for use in newborn screening. However, these studies are less useful for identifying which variants to include because they will exclude people with the disease who have died before reaching adulthood.
4. Whole genome or exome sequencing of stored dried blood spot samples could provide excellent data on penetrance if the samples are of sufficient quality for sequencing, and there is a robust system of follow-up to ascertain symptomatic disease status. An example of this approach has been successfully applied in California, where Adhikari et al. (2020) performed WES on dried blood spot samples and achieved exomes comparable to exomes from fresh blood in 1090/1416 samples.^189^ However, only samples from IEM-affected and MS/MS false positive samples were included and WES results were compared to follow-up testing precluding estimates of penetrance for the screening context. The challenges of doing these studies at scale are the cost and linking to good phenotype data.

For evidence on the clinical effectiveness of genetic screening in newborns, future research could explore the role of rare disease registries. A recent study from Germany evaluated the effectiveness of genetic newborn screening for spinal muscular atrophy (SMA) by comparing outcomes in screen detected and symptomatically detected patients (asymptomatic versus symptomatic treatment start) within the same healthcare system.^190^ This was feasible because two pilot projects for genetic SMA newborn screening were performed in Germany in two federal states before its nationwide implementation in 2021. This represents an excellent example how a study can be designed within a rare disease registry. However, registry data are not collected to address specific research questions and limitations will be common. Registry studies rely on high quality of data collection and reporting. The registry studies identified in our reviews were of low quality because none was comparative, no information on the type of test was provided and the information on the definitions of disease was insufficient.

Comparative (test accuracy) studies of gene panel tests, WES, WGS and expanded newborn blood spot testing are required to provide the evidence on which type of test is most promising for newborn screening. A technology centric approach (WGS for all conditions) has been identified as inappropriate for population screening^179^ and the best suited test needs to be identified for each condition under Consideration. For instance, WES has been shown to have insufficient sensitivity and specificity to replace MS/MS for IEMs in general but effectiveness varied among individual IEMs.^189^

UK micro-costing studies are needed to help underpin future cost-effectiveness evaluations of WGS, particularly when conducted in a routine clinical practice setting, rather than a research setting and across a range of clinical contexts to understand more fully when and why costs may differ.

Finally, there is a need for broader research about public perception and understanding of WGS and whether parents would still be in equipoise once they are fully informed. Further research should aim to map diverse views on complex topics in genomics and screening and associated views on risks and uncertainty to social characteristics, backgrounds and particular lived experiences to facilitate a nuanced understanding of the distributions of harms and benefits of genomic screening to inform policy recommendations and implementation. Future research also needs to investigate acceptability and accuracy of WGS in different populations (including island populations who are genetically diverse from mainland UK) and ethnic groups including those where consanguinity is common, and any resultant ethical and equity issues. Population genomic testing experience is skewed towards western Europeans and has resulted for instance in poorer prediction of polygenic risk scores in non-European populations.^179^ Concerns over deepening health disparities with WGS are warranted.

#### Conclusions

A traditional approach to systematic reviewing for WGS of newborns is unfeasible and the review does not reveal a new way to evaluate WGS for newborn screening in a single mechanism. There are two reasons for this: 1. there is insufficient evidence on each condition to allow a conventional UK NSC assessment; and 2. the variations in penetrance, natural history, test accuracy and effectiveness of treatments means that an aggregate is not informative. Our review highlights the main evidence gaps and informs the direction of future research efforts.

We propose a series of possible research approaches undertaken in large joint-up collaborations to produce the evidence that is needed for policy advisors before an evaluation of WGS is feasible. This may include a coordinated International approach to collecting penetrance data. This could be followed by a staged approach of evaluation considering only pathogenic variants with very high penetrance for screening.

## Data Availability

No new data have been created in the preparation of this article and therefore there is nothing available for access and further sharing. All queries should be submitted to the corresponding author.

## Additional information

### Declaration of competing interests

#### Full disclosure of interests

Completed ICMJE forms for all authors, including all related interests, are available in the toolkit on the NIHR Journals Library report publication page

#### Primary conflicts of interest

KF, JD, BS, CC, IK, KS, SC, AO, ND, RC, FB, FB, YT and STP received funding from the National Institute for Health and Care Research Evidence Synthesis Programme (ESG_HTA_NIHR1599280) to undertake this work. JD received funding from the Birmingham Biomedical Research Centre (BRC). FB received funding from Genomics England Limited for a process and impact evaluation. BS is a member of the UK NSC. CV is a Principal Evidence Review Manager of the UK NSC. JRB, DE and GS are clinical advisors to the UK NSC. AM is the Director of Programmes for the UK NSC. JRB is President of the International Society of Neonatal Screening. ZM is co-chair of the Scottish Clinical Genomics Forum and a committee member of the Clinical Genetics Society, the Scottish Strategic Network for genomic medicine, the NHSE rare disease test evaluation working group and board member of the Royal Colleges Genomics Advisory Board. FB is a member of the UK NSC Foetal, Maternal and Child Health Reference Group, the Bloodspot Task group and the Spinal Muscular Atrophy In Service Evaluation Partnership Board. STP is a member of the UK NSC AI task group, and chair of the UK NSC Research Methodology Group. SM has nothing to declare.

#### Contribution of authors and acknowledgements

**Karoline Freeman**: Conceptualisation, Investigation, Methodology, Project administration, Supervision, Writing – Original draft, Writing – reviewing and editing. **Jacqueline Dinnes**: Conceptualisation, Investigation, Methodology, Supervision, Writing – Original draft, Writing – reviewing and editing. **Bethany Shinkins**: Conceptualisation, Investigation, Writing – Original draft, Writing – reviewing and editing. **Corinna Clark**: Investigation, Writing – Original draft, Writing – reviewing and editing, lead of PPI work. **Inès Kander**: Investigation, Original draft. **Katie Scandrett**: Investigation, Original draft. **Shivashri Chockalingam**: Investigation, Original draft. **Aziza Osman**: Investigation. **Naila Dracup**: Investigation, Writing – Original draft. **Rachel Court**: Investigation, Supervision, Writing – Original draft. **Furqan Butt**: Investigation. **Cristina Visitin**: Conceptualisation, Methodology, Writing – reviewing and editing. **James R. Bonham**: Conceptualisation, Methodology, Writing – reviewing and editing. **David Elliman**: Conceptualisation, Methodology, Writing – reviewing and editing. **Graham Shortland:** Conceptualisation, Methodology, Writing – reviewing and editing. **Anne Mackie**: Conceptualisation, Methodology, Writing – reviewing and editing. **Zosia Miedzybrodzka**: Writing – reviewing and editing. **Sian Morgan**: Writing – reviewing and editing. **Felicity Boardman**: Methodology, Writing – reviewing and editing. **Yemisi Takwoingi**: Conceptualisation, Funding acquisition, Methodology, Writing – reviewing and editing. **Sian Taylor-Phillips**: Conceptualisation, Funding acquisition, Methodology, Supervision, Writing – Original draft.

We are grateful for the input of the seven members of the PPIE group, who included Kerri Pearce, Matt Howard-Murray, Frances Othen-Wales, Luis Canto E Castro and Helen Bates. The authors would also like to thank Dr Helen Jenkinson, Dr Austen Worth, Dr Emma Footit and Professor Raja Padidela for their specialist clinical advice on the rare conditions, and Professor Aileen Clarke for her Critical review of the manuscript.

#### Ethics statement

This report concerns secondary research, for which ethics approval is not required.

#### Information governance statement

This project did not involve the handling of personal information.

#### Department of Health and Social Care disclaimer

This publication presents independent research commissioned by the National Institute for Health and Care Research (NIHR). The views and opinions expressed in this publication are those of the authors and do not necessarily reflect those of the NHS, the NIHR, MRC, NIHR Coordinating Centre, the HTA programme or the Department of Health and Social Care.

## Appendices

### Appendix 1. Search development methods

*Overview of the search development methods for the review of five conditions, the review of genomic studies of paediatric cohorts reporting penetrance for pathogenic variants and the review of cost-effectiveness evaluations of WGS and WES*.

#### Review of five conditions

Search strategies were developed for each condition by an Information Specialist. The searches were developed in a test database (MEDLINE (Ovid)) and were informed and refined through a series of scoping searches, checks of a proportion of results from these searches and iterative discussions between the Information Specialist (ND), project lead (KF) and members of the reviewing team (IK, JD, SC).

Exploratory scoping searches were carried out for each condition to gain familiarity with the condition and the genetic cause. The scoping searches revealed that searching for the 5 conditions in one single search would not be a feasible approach, due to the complexity and differences across each condition and their genetic causalities. In our scoping searches, we tested searching for the specific genes associated with each condition and the condition, for example, rb1 and retinoblastoma. Combining the search terms for the condition and the gene with the Boolean operator ‘Or’ yielded an unmanageable volume of irrelevant results of the specific gene related to other conditions. For example, variants of the RB gene are associated with a large amount of other cancer types. Combining the terms for the gene and the condition using the Boolean operator ‘And’ would produce an overly specific search, which would have resulted in potentially relevant results being missed. Therefore, an iterative approach was adopted. A standardised search strand for terms related to hereditary/ inherited conditions or genetics was utilised for the conditions that can also present for reasons that are not due to the specific genetic variants. These included: familial hemophagocytic lymphohistiocytosis, retinoblastoma and X-linked hypophosphatemic rickets. The searches for Medium Chain Acyl-CoA Dehydrogenase Deficiency (MCADD) and pyridoxine-dependent epilepsy did not need to include the search terms for genetics, as the numbers retrieved were manageable without the addition of any other concepts and the draft search results for these conditions did not yield such a high proportion of irrelevant results. The process of developing and running the searches took a period of 6 weeks, which is longer than average for our Information Specialist team. This was largely due to the complexity of the topic.

Database-specific subject headings and free text words were identified for use in the search concepts by analysing the free text and indexing terms of the results from the scoping searches, text analysis software including medical subject heading (MeSH) on Demand, Anne O’Tate and PubMed ReMiner by the Information Specialist. Further terms were identified and tested from known relevant papers and resources including ClinGen and GeneReviews. The searches were peer-reviewed by a Senior Information Specialist and the project lead.

#### Review of genomic studies of paediatric cohorts reporting penetrance for pathogenic variants

Searches were developed iteratively in a single database (MEDLINE via Ovid) by an experienced information specialist (RC) and the project lead (KF), with input from members of the reviewing team. Developing and running the searches took approximately five weeks. The development process took longer than usual because there appears to be little evidence in the newborn screening setting. Despite running scoping searches, we did not have enough examples of published literature to confidently base an Initial search on. Therefore, we began with a narrow search combining the concepts of Whole Genome Sequencing (WGS) and newborn screening, checking samples of records for potentially relevant literature. Simultaneously, we undertook targeted searches to check for outputs from known large genomic studies, such as the BabySec project. It became evident that most of these ongoing studies had not published results at the time. We question whether a future review of just large genomic studies would be helpful to answer the question around penetrance because 1) conditions are so rare that there is limited scope for evidence on penetrance from one or two years’ worth of data and 2) newborns positive on WGS will be treated and not followed up to symptoms. Due to not finding much of relevance in this first iteration of the search and our other concerns, we broadened it by adding search terms for related concepts, such as genetic testing and sequencing, ran test searches with and without certain concepts and checked samples for potentially relevant literature. We also considered other approaches, such as searching for specific terms within the main body of an article using a database of full-text publications. The final search combines the concept of newborn screening with either WGS, WES, penetrance, actionability, sequencing or allele frequency. This search retrieved a large, but manageable number of records in MEDLINE and found all five known studies we had identified up to this point.^127, 131, 133, 134, 145^

##### Search strategy

The following databases were searched from inception to January 2024 (see Appendix 2 for exact dates and full search details): MEDLINE (via Ovid), Embase (via Ovid), Science Citation Index (via Clarivate) and the Cochrane Library (via Wiley). No date, language or study type filters were applied. Records were exported to EndNote and systematically de-duplicated using a process based on the University of Leeds method.^17^

#### Review of cost-effectiveness evaluations of WGS and WES

The searches for the review of cost-effectiveness studies were developed by an Information Specialist with input from the lead economist (BS) and peer reviewed by the Senior Information Specialist (RC). Comprehensive database searches were undertaken to identify evidence relating to cost effectiveness and whole genome sequencing. The search strategies are reported in Appendix 2.

We considered updating the search strategy carried out for a systematic review by Schwarze et al. (2018);^26^ however, as their review was carried out in July 2016, they were able to significantly make use of the NHS Economic Evaluation Database (NHS EED), produced by the Centre for Reviews and Dissemination (CRD). NHS EED ceased adding new records in March 2015 and the searches to identify them stopped at the end of 2014;^191^ therefore it was agreed that we would need to carry out broader searches on MEDLINE, Embase and the Science Citation Index, utilising an economic search filter. Supplementary searches was also carried out on the CEA registry and the review by Scwartz et al (2018)^26^ was used as a source, by cross-checking their included studies.

The search terms were derived from analysing the free text and indexing terms of relevant known studies and analysing the results from Initial scoping searches. The terms for the cost effectiveness search strand was developed from an economic search filter.^192^ The Medline (Ovid), Embase (Ovid), Science Citation Index (Web of Science – Clarivate), EconLit (EBSCO), the CEA Registry, the HTA International database and Google were searched from inception on the 13^th^ February 2023 for economic evaluations, health technology assessment reports and economic models. The search results were limited to English language studies. The search results were stored and de-duplicated in EndNote 20 using the University of Leeds method.^17^

### Appendix 2. Search strategy

*Search details including databases, date of search, terms and breakdown of number of results for the traditional review of 5 conditions, the review of penetrance or actionability of gene variants of rare genetic childhood-onset diseases identified in newborn screening populations using WGS (alternative review) and the review of cost-effectiveness evaluations of WGS and WES*.

#### Review of five conditions

##### PDE

###### Search summary

| Database (Platform) | Date searched | Concepts | Hits | Notes |
| --- | --- | --- | --- | --- |
| MEDLINE (Ovid) | 29/09/2023 | Pyridoxamine dependent epilepsy | 545 | Removed animal studies<br>English language limit |
| Embase (Ovid) | 29/09/2023 | Pyridoxamine dependent epilepsy | 706 | Removed animal studies<br>No Emtree terms focussed<br>English language limit |
| Science Citation Index (Web of Science) | 29/09/2023 | Pyridoxamine dependent epilepsy | 584 |  |
| Cochrane Library | 29/09/2023 | Pyridoxamine dependent epilepsy | 7 |  |

Total from database searches: 1835
Total after systematically removing duplicates in EndNote (using University of Leeds method)<sup>17</sup>: 994

###### Ovid MEDLINE(R) ALL 1946 to September 18, 2023

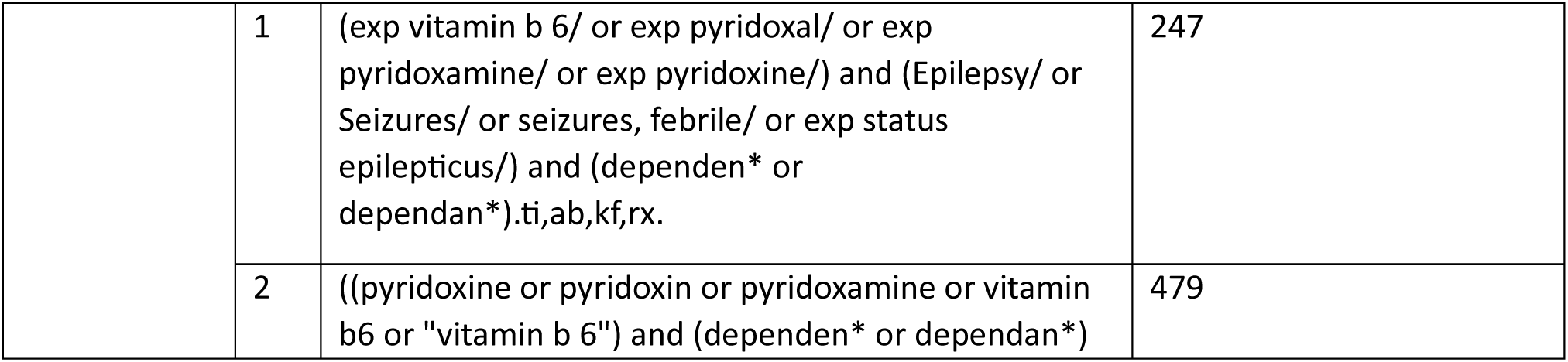
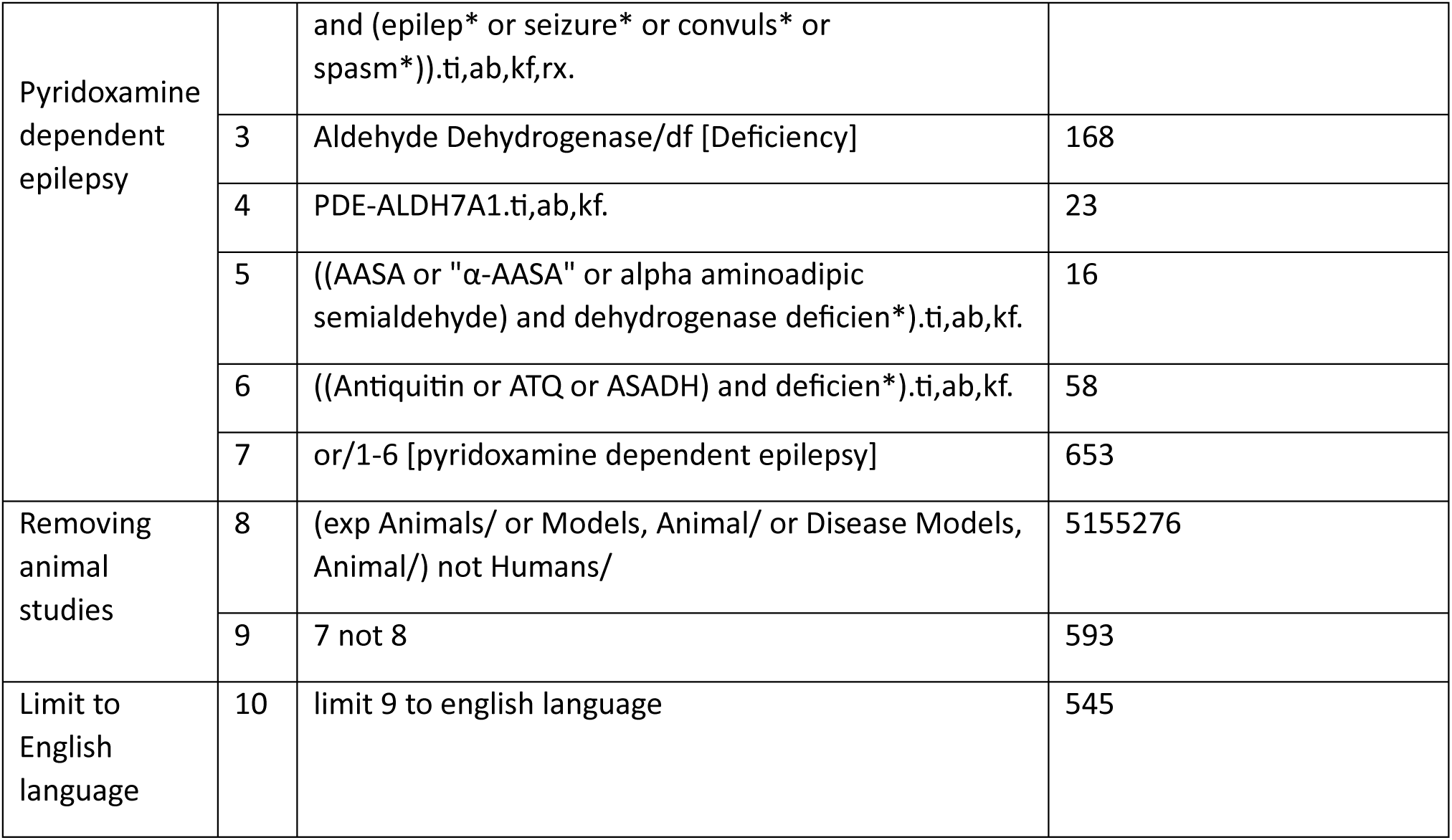

|  |  |  |
| --- | --- | --- |
| 1 | (exp vitamin b 6/ or exp pyridoxal/ or exp pyridoxamine/ or exp pyridoxine/) and (Epilepsy/ or Seizures/ or seizures, febrile/ or exp status epilepticus/) and (dependen* or dependan*).ti,ab,kf,rx. | 247 |
| 2 | ((pyridoxine or pyridoxin or pyridoxamine or vitamin b6 or "vitamin b 6") and (dependen* or dependan*)) | 479 |

|  |  |  |  |
| --- | --- | --- | --- |
| Pyridoxamine dependent epilepsy |  | and (epilep* or seizure* or convuls* or spasm*).ti,ab,kf,rx. |  |
|  | 3 | Aldehyde Dehydrogenase/df [Deficiency] | 168 |
|  | 4 | PDE-ALDH7A1.ti,ab,kf. | 23 |
|  | 5 | ((AASA or "α-AASA" or alpha aminoadipic semialdehyde) and dehydrogenase deficien*).ti,ab,kf. | 16 |
|  | 6 | ((Antiquitin or ATQ or ASADH) and deficien*).ti,ab,kf. | 58 |
|  | 7 | or/1-6 [pyridoxamine dependent epilepsy] | 653 |
| Removing animal studies | 8 | (exp Animals/ or Models, Animal/ or Disease Models, Animal/) not Humans/ | 5155276 |
|  | 9 | 7 not 8 | 593 |
| Limit to English language | 10 | limit 9 to english language | 545 |

###### Embase Classic+Embase 1947 to 2023 September 19

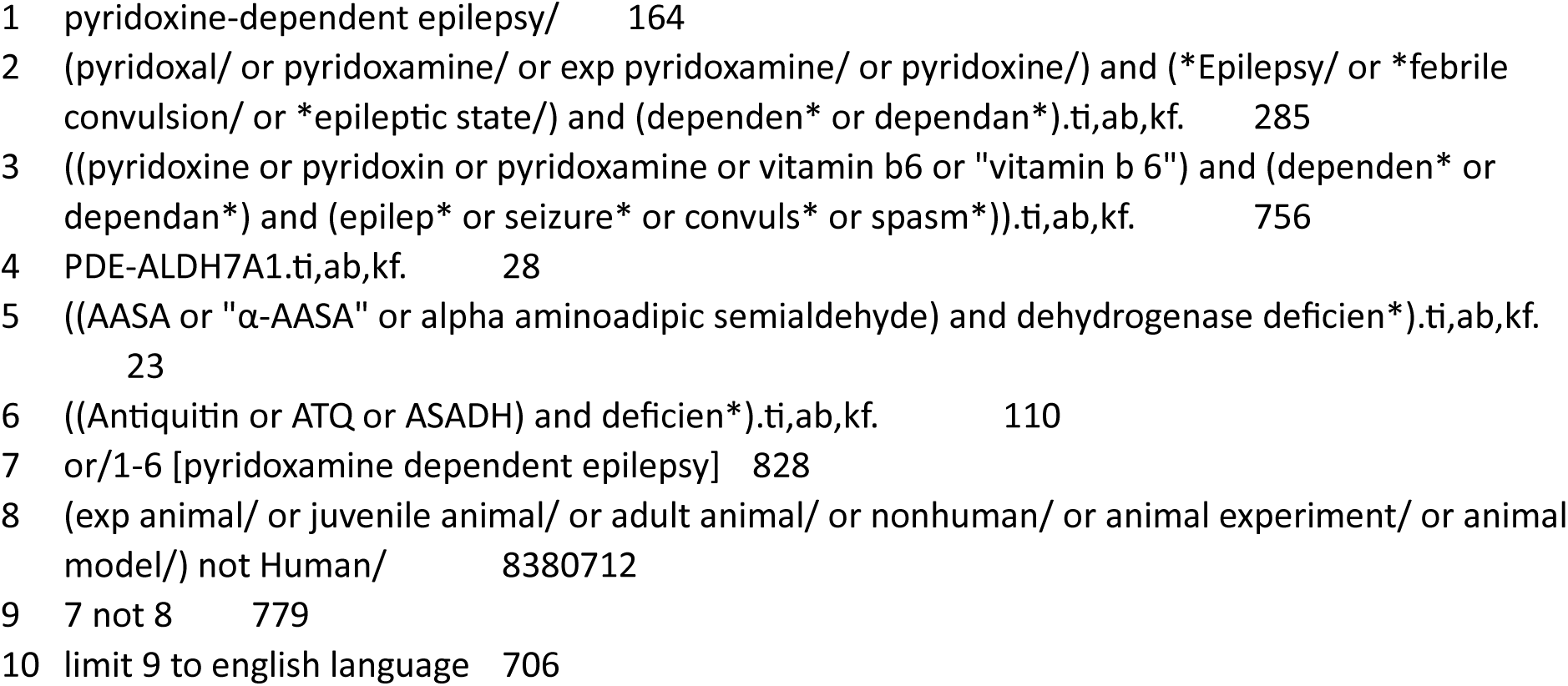

###### Web of Science - Science Citation Index Expanded, (SCI-EXPANDED) 1970-present, Social Sciences Citation Index (SSCI) 1900-present

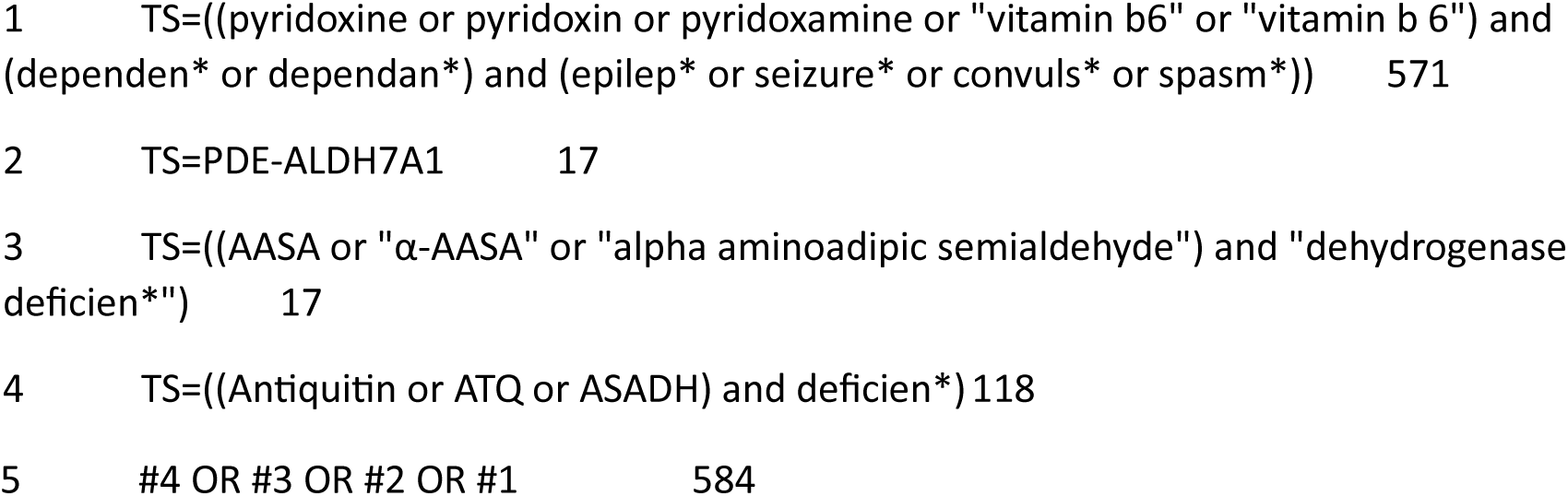

###### Cochrane library

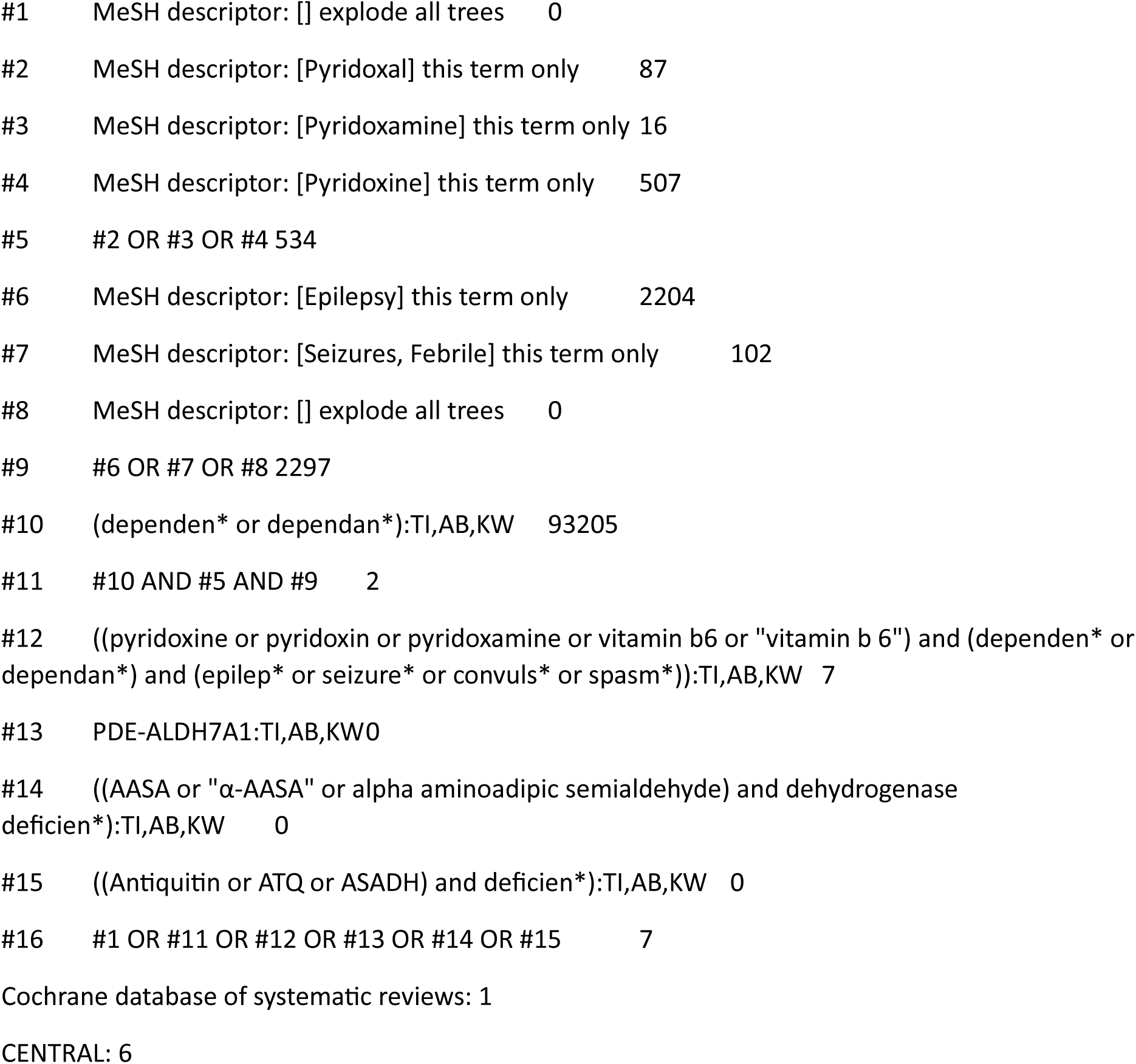

##### hRB

###### Search summary

| Database (Platform) | Date searched | Concepts | Hits | Notes |
| --- | --- | --- | --- | --- |
| MEDLINE (Ovid) | 04/10/2023 | Retinoblastoma<br>AND<br>Genetics/ hereditary | 4521 | Removed animal studies<br>English language limit |
| Embase (Ovid) | 04/10/2023 | Retinoblastoma<br>AND<br>Genetics/ hereditary | 3345 | Removed animal studies<br>English language limit |
| Science Citation Index (Web of Science) | 04/10/2023 | Retinoblastoma<br>AND<br>Genetics/ hereditary | 471 | Removed animal studies<br>English language limit |
| Cochrane Library | 04/10/2023 | Retinoblastoma<br>AND<br>Genetics/ hereditary | 18 |  |

Total from database searches: 8355
Total after systematically removing duplicates in EndNote (using University of Leeds method)<sup>17</sup>: 5797

###### Ovid MEDLINE(R) ALL 1946 to October 02, 2023

|  |  |  |  |
| --- | --- | --- | --- |
| Retinoblastoma | 1 | Retinoblastoma/ | 8207 |
|  | 2 | Retinal Neoplasms/ge | 919 |
|  | 3 | Genes, Retinoblastoma/ | 1772 |
|  | 4 | (Retina* adj3 (cancer* or tumor* or tumour* or neoplasm* or glioblastoma* or glioma* or neuroblastoma*)).ti,ab,kf. | 1582 |
|  | 5 | or/1-4 [retinoblastoma] | 10873 |
|  | 6 | Genetics/ | 12921 |
|  | 7 | Genetics.fs. | 4015866 |
|  | 8 | Genetic disorder/ | 14547 |
| Hereditary/<br>genetics | 9 | exp genetic predisposition to disease/ | 157705 |
|  | 10 | Genetic Diseases, Inborn/ | 14547 |
|  | 11 | (Genetic* or gene or genes or family or families or familial or DNA or hereditary or heritable or heredodegenerative* or inherit* or congenital* or germline or germinal*).ti,ab,kf. | 5102985 |
|  | 12 | or/6-11 [Hereditary/ genetics] | 6533487 |
| Retinoblastoma<br>and genetics/<br>hereditary | 13 | 5 and 12 [Retinoblastoma and genetics/<br>hereditary] | 5401 |
| Removed<br>animal studies | 14 | (exp Animals/ or exp Models, Animal/ or<br>Disease Models, Animal/) not Humans/ | 5161657 |
|  | 15 | 13 not 14 | 5025 |
| Limit to English<br>language | 16 | limit 15 to english language | 4521 |

###### Embase Classic+Embase 1947 to 2023 Week 39

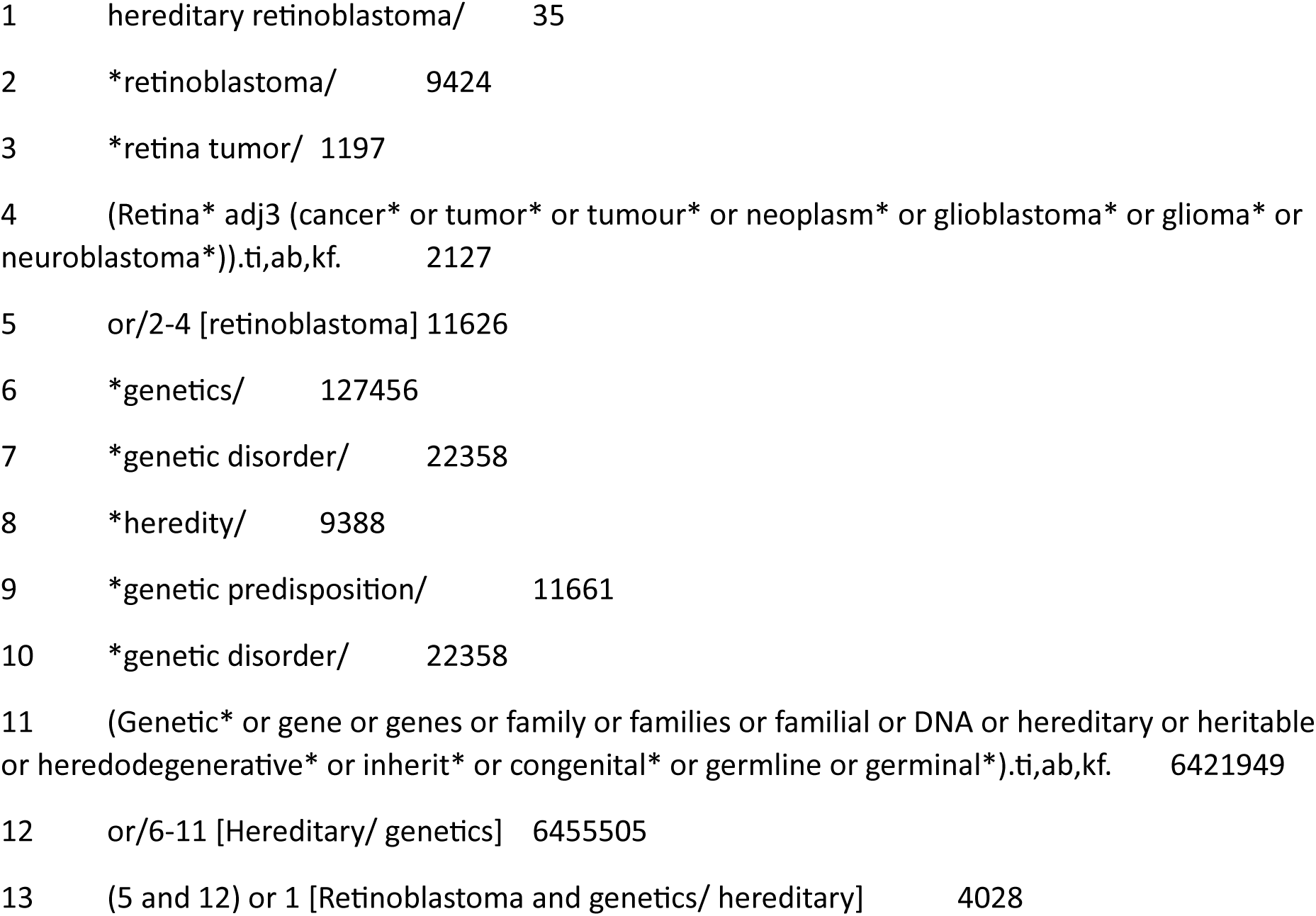
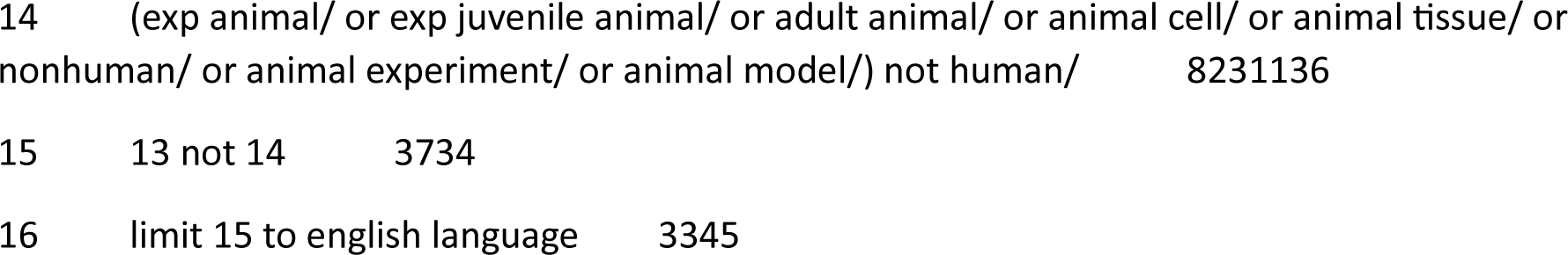

###### Web of Science

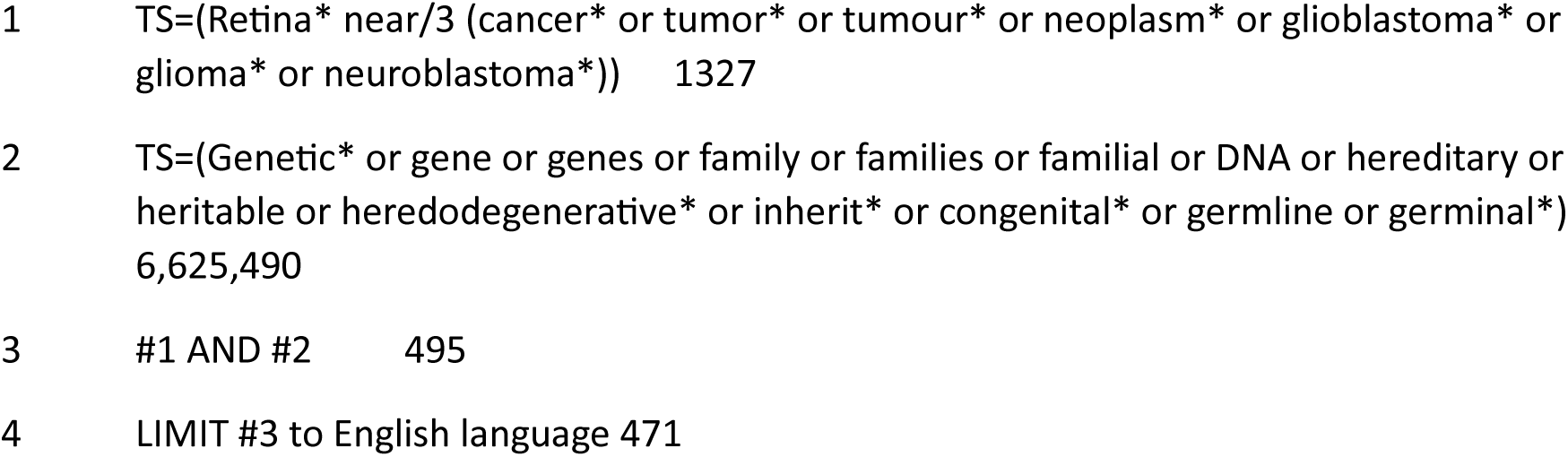

###### Cochrane library

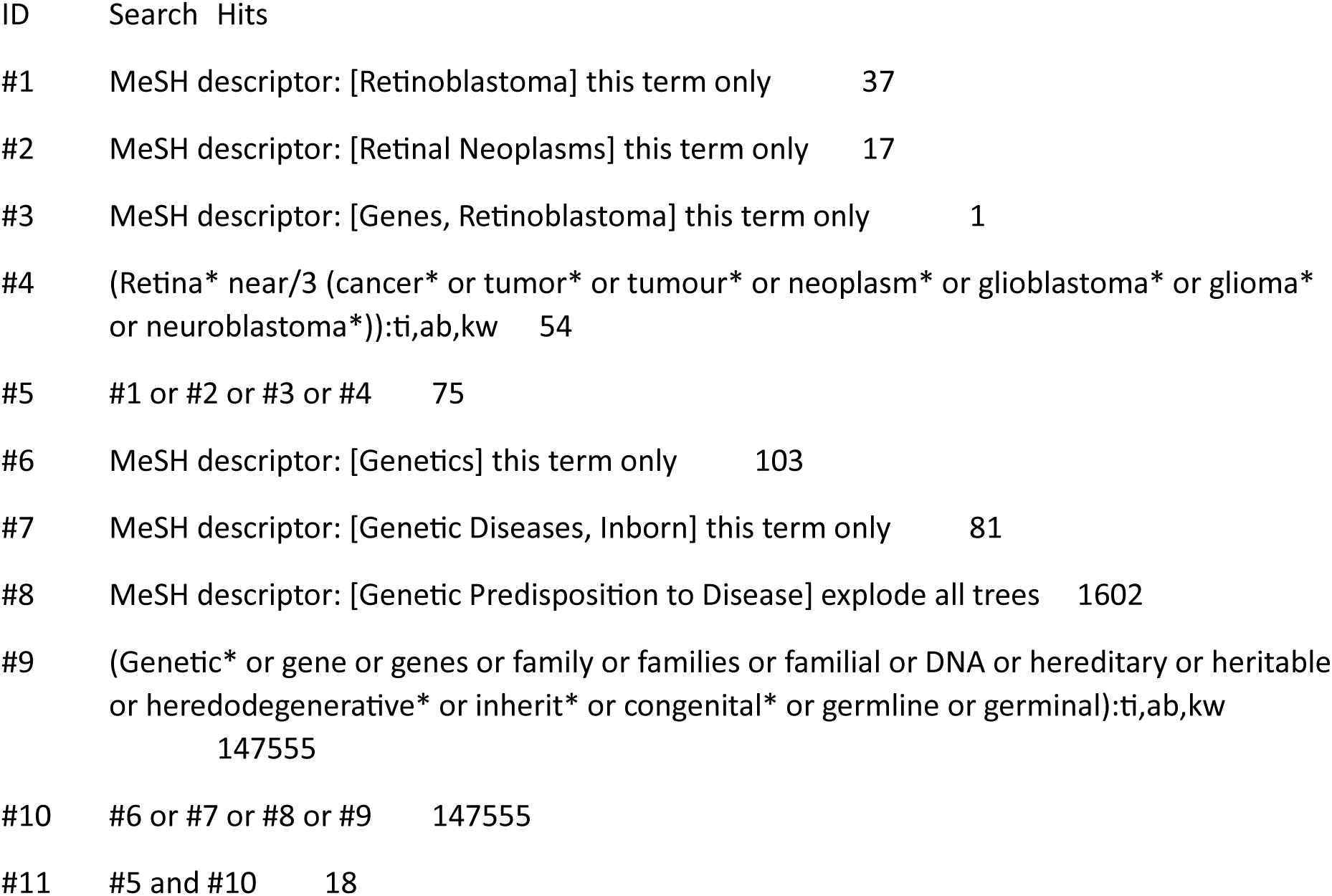

##### XLHR

###### Search summary

| Database (Platform) | Date searched | Concepts | Hits | Notes |
| --- | --- | --- | --- | --- |
| MEDLINE (Ovid) | 05/10/2023 | X-linked Hypophosphatemic Rickets<br>AND<br>Genetics/ hereditary | 2078 | Removed animal studies<br>English language limit |
| Embase (Ovid) | 05/10/2023 | X-linked Hypophosphatemic Rickets<br>AND<br>Genetics/ hereditary | 3474 | Removed animal studies<br>English language limit |
| Science Citation Index (Web of Science) | 05/10/2023 | X-linked Hypophosphatemic Rickets<br>AND<br>Genetics/ hereditary | 2088 | English language limit |
| Cochrane Library | 05/10/2023 | X-linked Hypophosphatemic Rickets<br>AND<br>Genetics/ hereditary | 278 |  |

Total from database searches: 7918
Total after systematically removing duplicates in EndNote (using University of Leeds method)<sup>17</sup>: 4787

###### Ovid MEDLINE(R) ALL 1946 to September 18, 2023

|  |  |  |  |
| --- | --- | --- | --- |
| Hypophosphatemic rickets | 1 | exp Rickets, Hypophosphatemic/ | 952 |
|  | 2 | Hypophosphat?emic.ti,ab,kf,rx. | 2433 |
|  | 3 | rickets/ or (rickets or rachitides or rachitis).ti,ab,kf,rx. | 10233 |
|  | 4 | 2 and 3 | 1644 |
|  | 5 | 1 or 4 [Hypophosphatemic rickets] | 2080 |
| Hypophosphatemia | 6 | Hypophosphatemia/ | 1908 |
|  | 7 | Hypophosphat?emia*.ti,ab,kf,rx. | 5397 |
|  | 8 | or/6-7 [Hypophosphatemia] | 5864 |
| Vitamin d resistant rickets | 9 | (Vitamin d resistant and (rickets or rickets or rachitides or rachitis)).ti,ab,kf,rx. | 608 |
|  | 10 | VDRR.ti,ab,kf. | 30 |
|  | 11 | or/9-10 [Vitamin d resistant rickets] | 612 |
| Hypophosphatemia or Hypophosphatemic rickets or vitamin d resistant rickets | 12 | 5 or 8 or 11 [Hypophosphatemia or Hypophosphatemic rickets or vitamin d resistant rickets] | 7318 |
| Genetics/ hereditary | 13 | Genetics/ | 12922 |
|  | 14 | Genetics.fs. | 4016108 |
|  | 15 | Genetic disorder/ | 14548 |
|  | 16 | exp genetic predisposition to disease/ | 157711 |
|  | 17 | Genetic Diseases, Inborn/ | 14548 |
|  | 18 | (Genetic* or gene or genes or family or families or familial or DNA or hereditary or heritable or heredodegenerative* or inherit* or congenital* or germline or germinal*).ti,ab,kf. | 5103147 |
|  | 19 | or/13-18 [Genetics/ hereditary] | 6533601 |
| Hypophosphatemia or Hypophosphatemic rickets and genetics/ hereditary | 20 | 12 and 19 [Hypophosphatemia or Hypophosphatemic rickets and genetics/ hereditary] | 2327 |
| Phrase searches for X-linked hypophosphatemic hypophosphataemic rickets | 21 | ((XLHR or XLH or X Linked) adj3 (hypophosphatemia* or hypophosphataemia*)).ti,ab,kf,rx. | 659 |
|  | 22 | ((((XLHR or XLH or X Linked) adj3 (Hypophosphatemic or hypophosphataemic)) and (rickets or rachitides or rachitis)).ti,ab,kf,rx. | 580 |
|  | 23 | ((((XLHR or XLH or X Linked) adj3 vitamin d resistant) and (rickets or rachitides or rachitis)).ti,ab,kf,rx. | 41 |
|  | 24 | or/20-23 | 2656 |
| Removal of animal studies | 25 | (exp Animals/ or exp Models, Animal/ or Disease Models, Animal/) not Humans/ | 5162107 |
|  | 26 | 24 not 25 | 2286 |
| Limit to English language | 27 | limit 26 to english language | 2078 |

###### Embase Classic+Embase 1947 to 2023 October 04

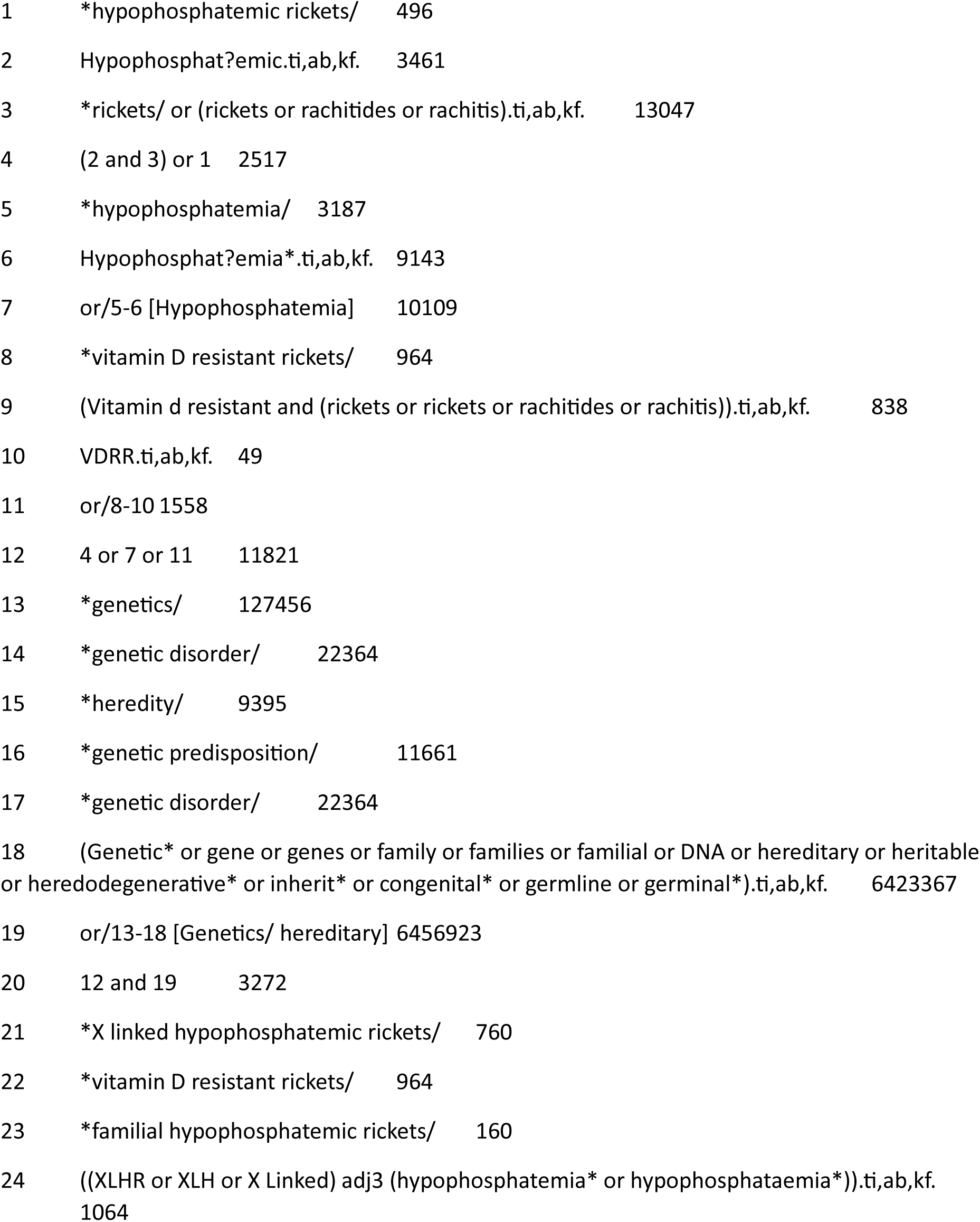
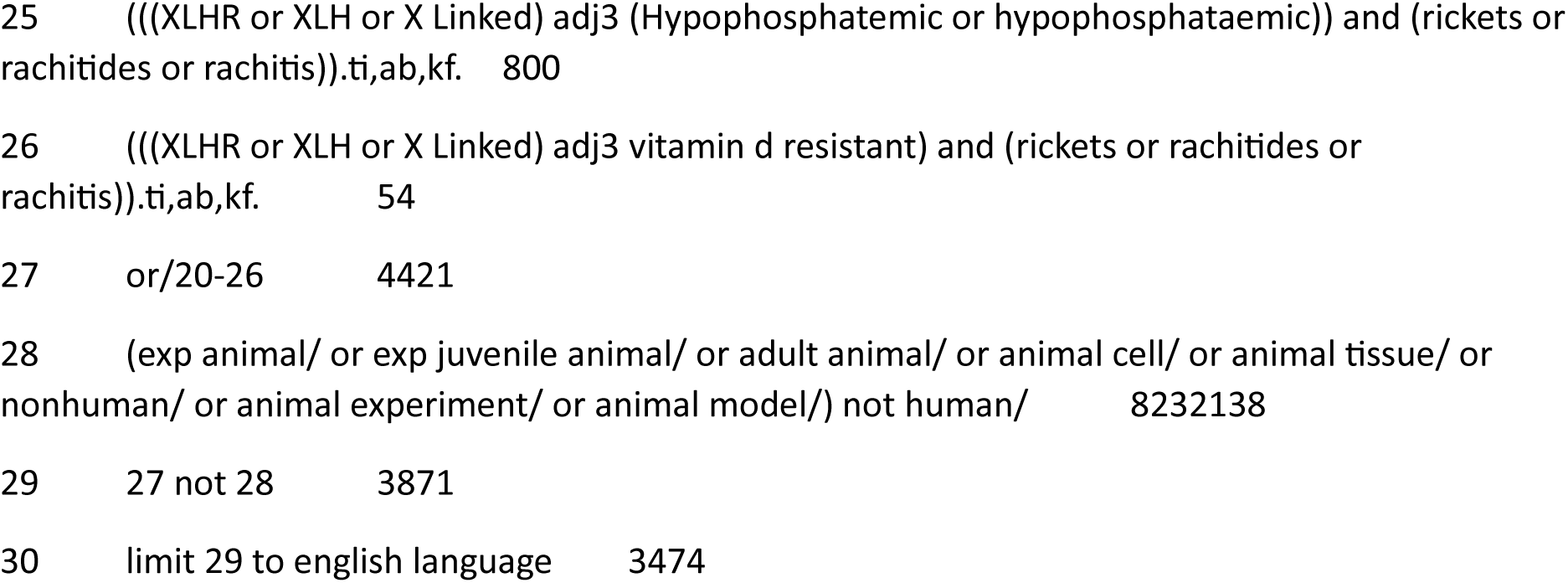

###### Web of Science - Science Citation Index Expanded (SCI-EXPANDED) -1970-present, Social Sciences Citation Index (SSCI)-1900-present

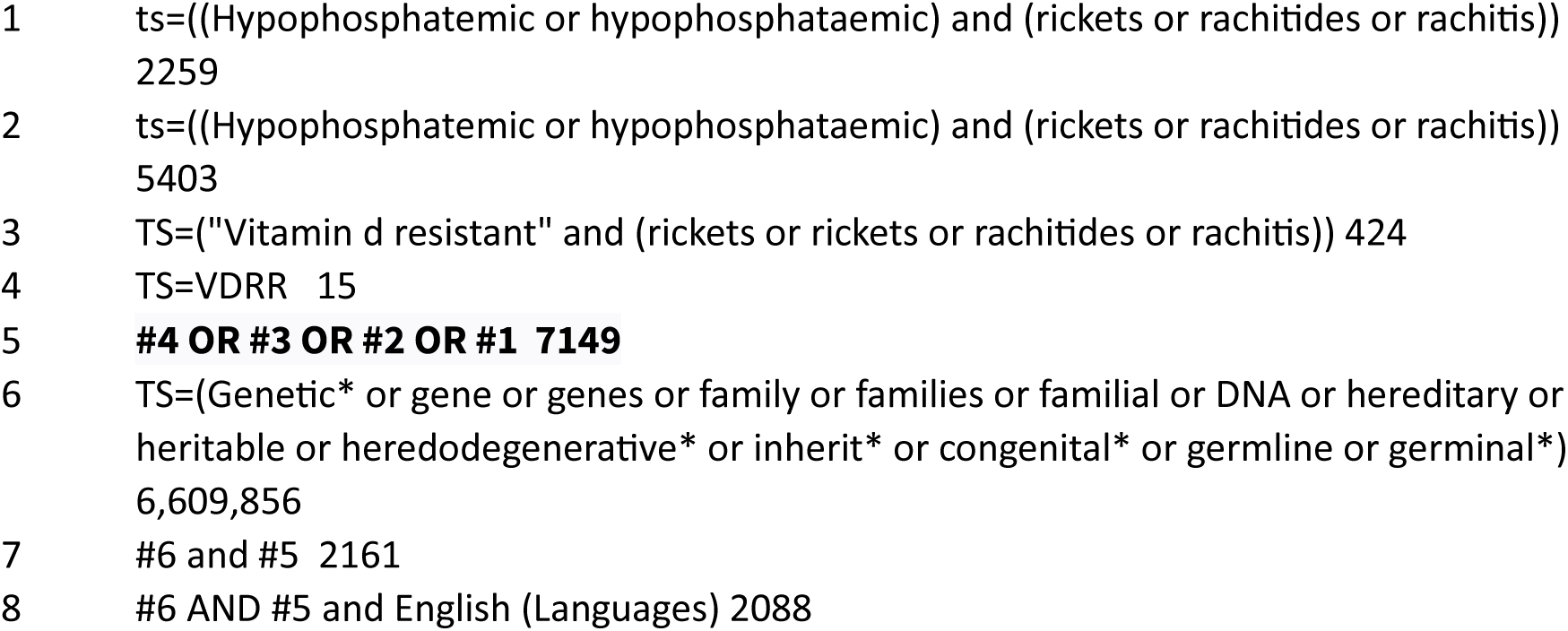

###### Cochrane library

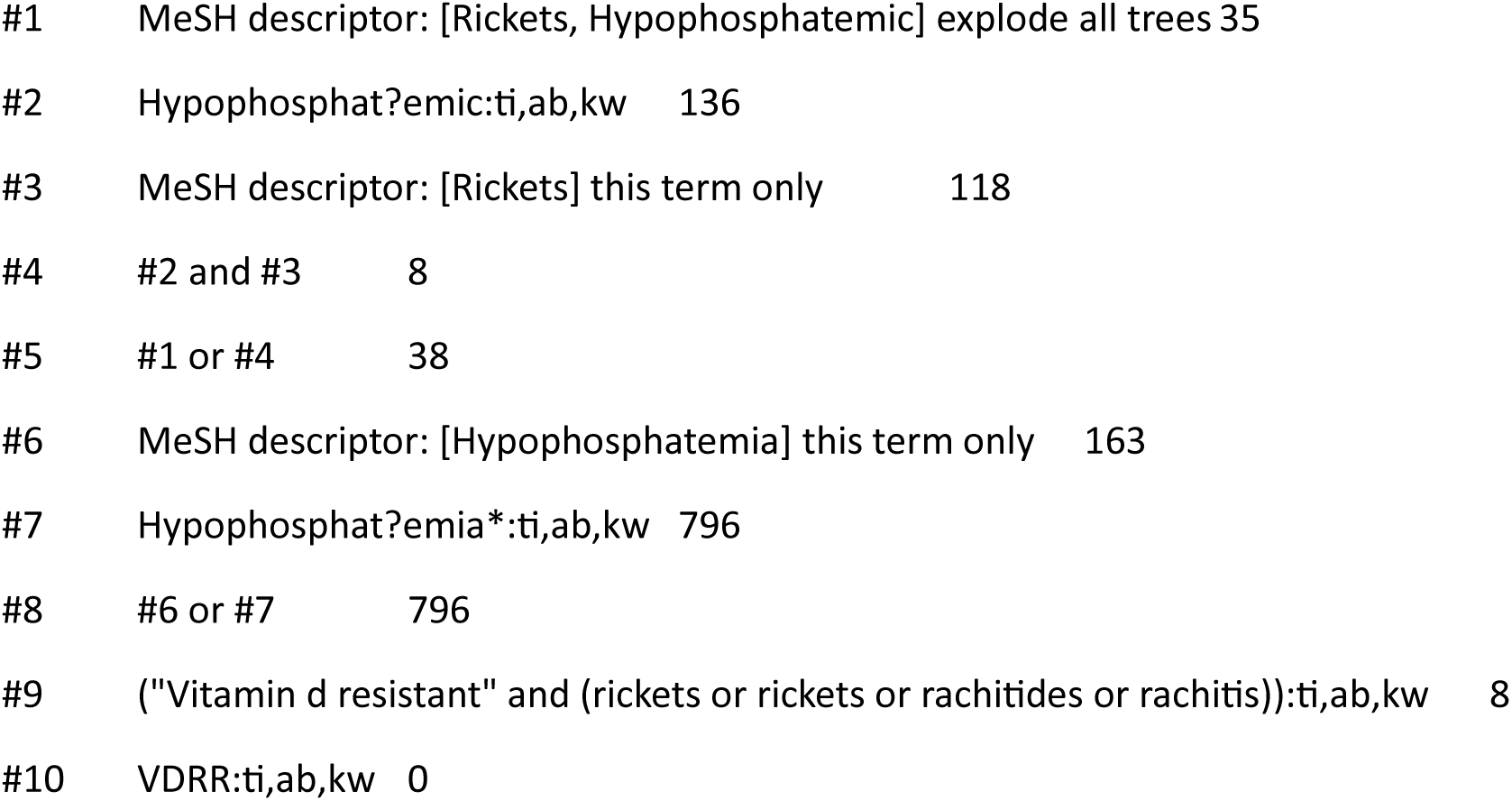
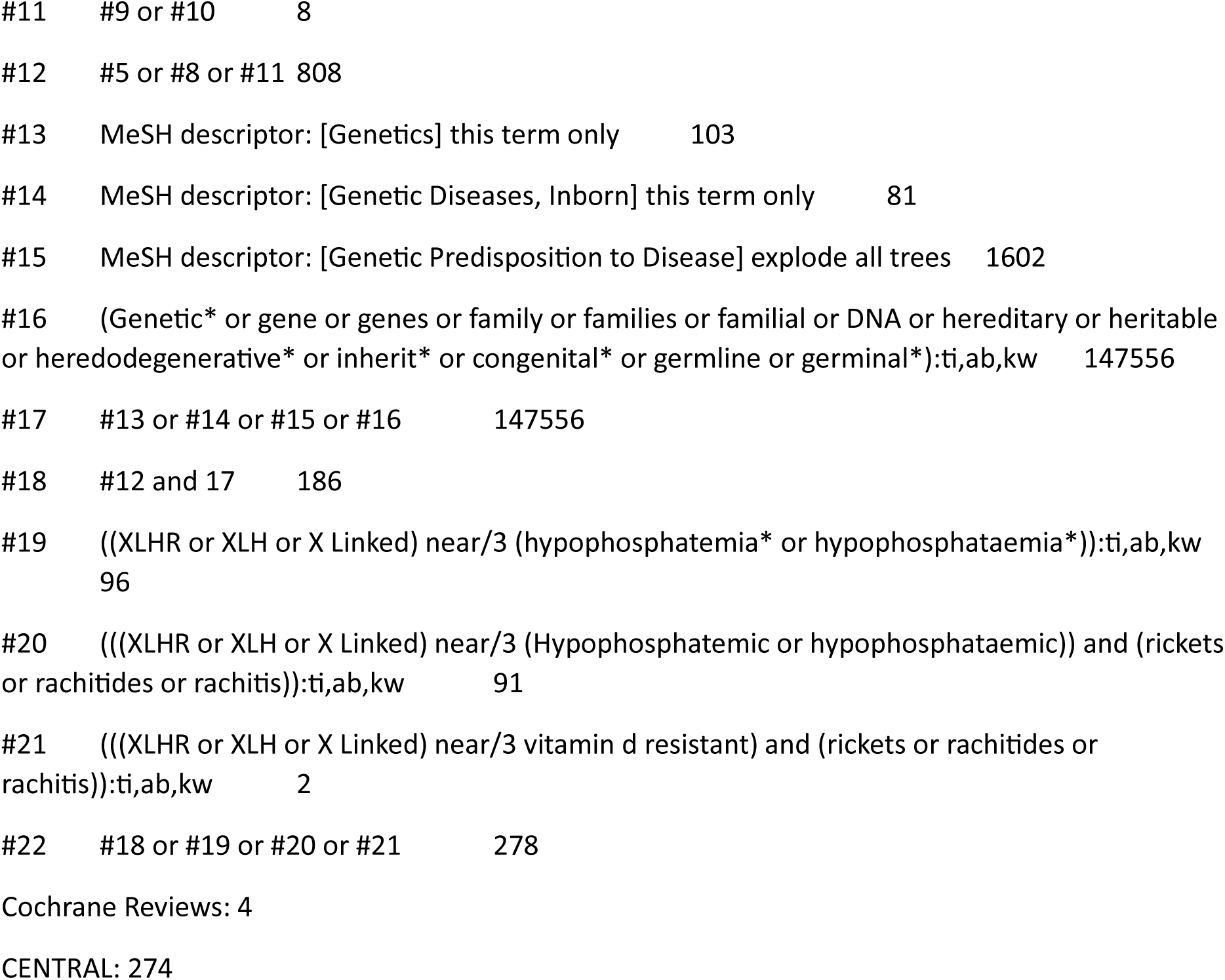

##### fHLH

###### Search summary

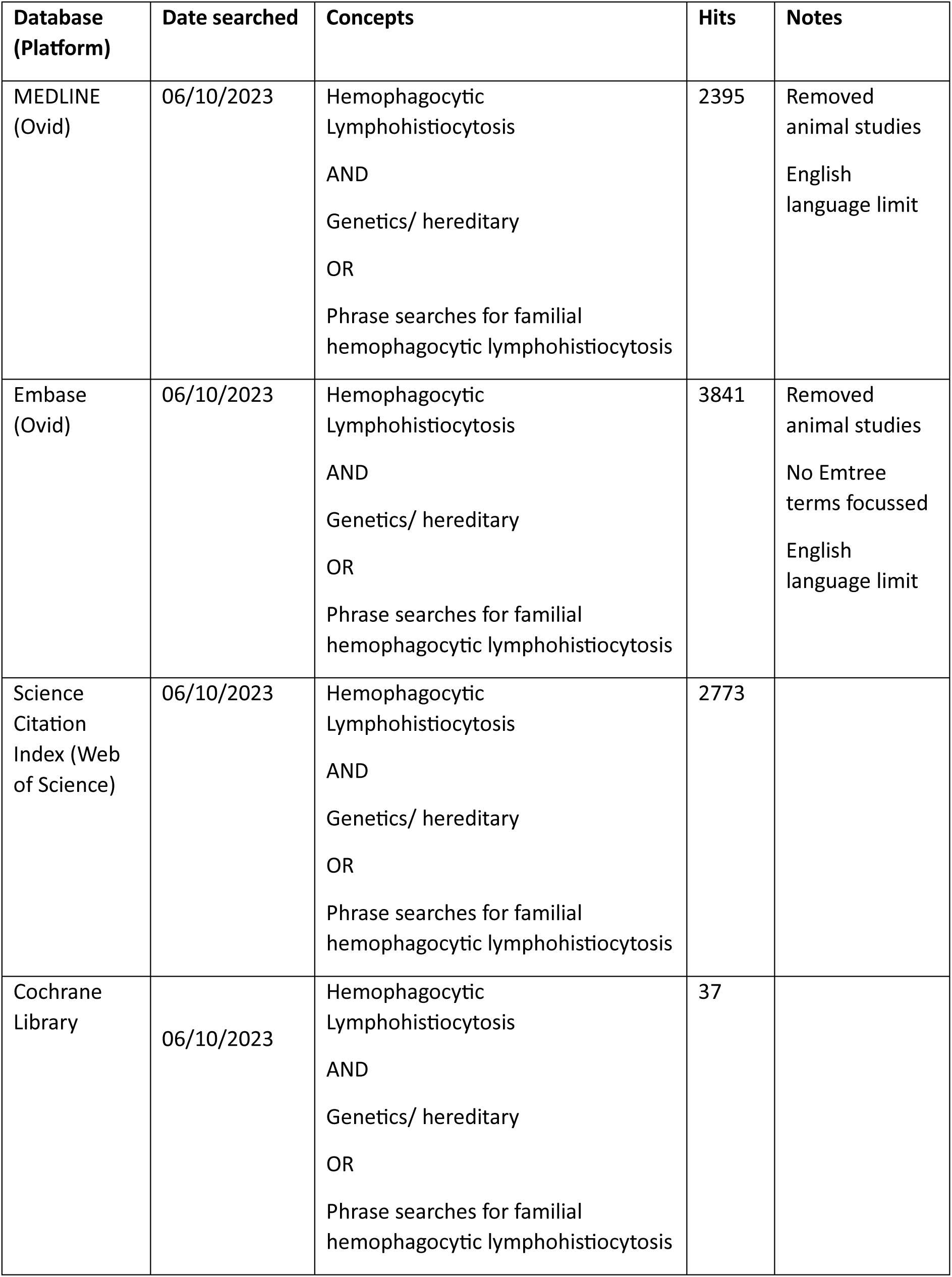
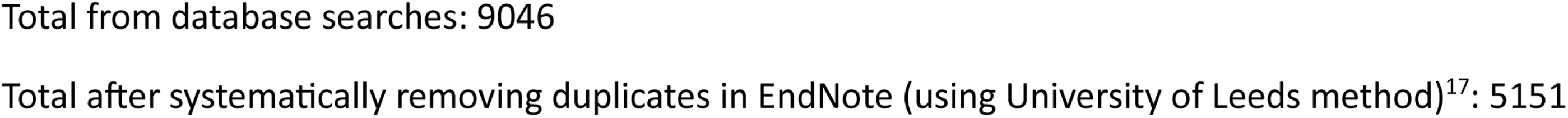

###### Ovid MEDLINE(R) ALL 1946 to October 05, 2023

|  |  |  |  |
| --- | --- | --- | --- |
|  | 1 | Lymphohistiocytosis, Hemophagocytic/ | 3949 |
| Hemophagocytic<br>Lymphohistiocytosis | 2 | ((Hemophagocytic or haemophagocytic or erythrophagocytic) adj3 (lymphohistiocytos* or lymphocytos* or histiocytos* or reticulos* or lymphohistiocytos* or syndrome*)).ti,ab,kf,rx. | 7098 |
|  | 3 | or/1-2 [Hemophagocytic Lymphohistiocytosis] | 7556 |
| Genetics/ hereditary | 4 | Genetics/ | 12922 |
|  | 5 | Genetics.fs. | 4016776 |
|  | 6 | Genetic disorder/ | 14549 |
|  | 7 | exp genetic predisposition to disease/ | 157723 |
|  | 8 | Genetic Diseases, Inborn/ | 14549 |
|  | 9 | (Genetic* or gene or genes or familial or family or families or DNA or hereditary or heritable or heredodegenerative* or inherit* or congenital* or germline or germinal*).ti,ab,kf. | 5104100 |
|  | 10 | or/4-9 [Genetics/ hereditary] | 6534784 |
| Hemophagocytic<br>Lymphohistiocytosis<br>and genetics/<br>hereditary | 11 | 3 and 10 | 2618 |
| Familial<br>Hemophagocytic<br>Lymphohistiocytosis | 12 | ((Primary adj4 (hemophagocytic or haemophagocytic or erythrophagocytic)) and (lymphohistiocytosis or lymphocytos* or histiocytos* or reticulos* or lymphohistiocytos* or syndrome*)).ti,ab,kf,rx. | 210 |
|  | 13 | (FHLH or PHLH).ti,ab,kf. | 99 |
|  | 14 | or/11-13 | 2709 |
| Excluding animal<br>studies | 15 | (exp Animals/ or exp Models, Animal/ or Disease Models, Animal/) not Humans/ | 5162502 |
|  | 16 | 14 not 15 | 2657 |
| English language | 17 | limit 16 to english language | 2395 |

###### Embase Classic+Embase 1947 to 2023 October 05

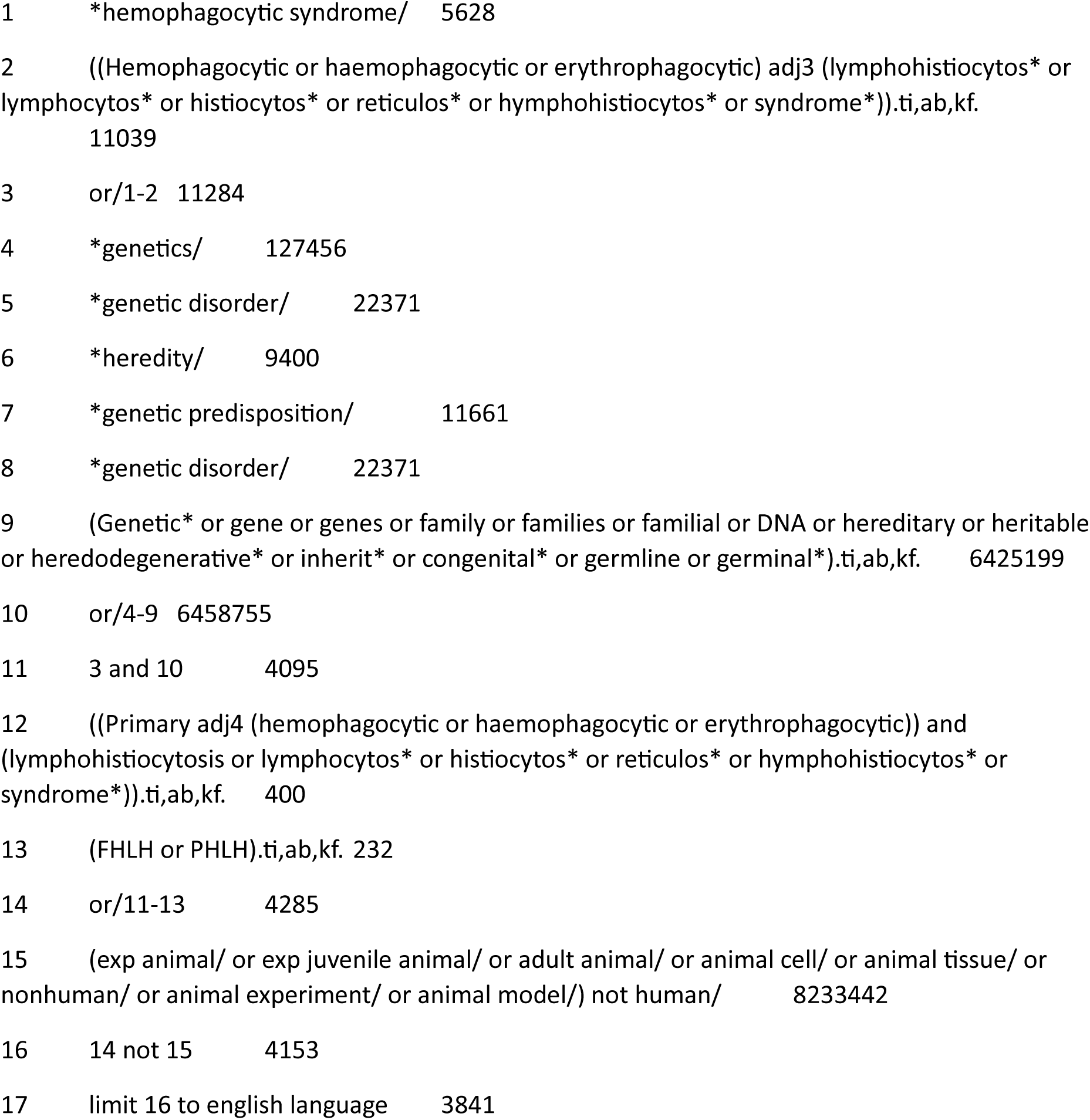

###### Web of Science Science Citation Index Expanded (SCI-EXPANDED) 1970-present, Social Sciences Citation Index (SSCI) 1900-present

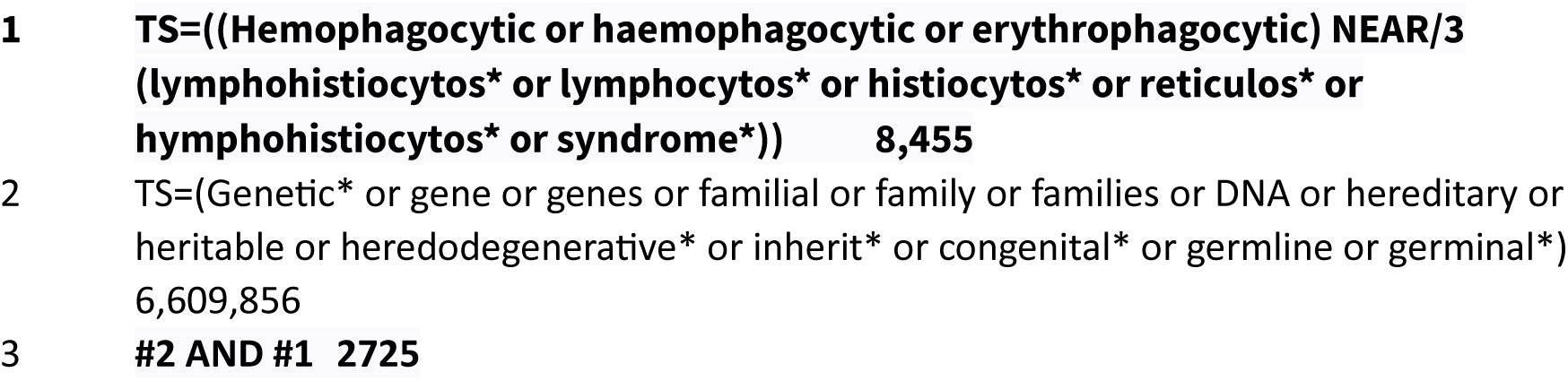
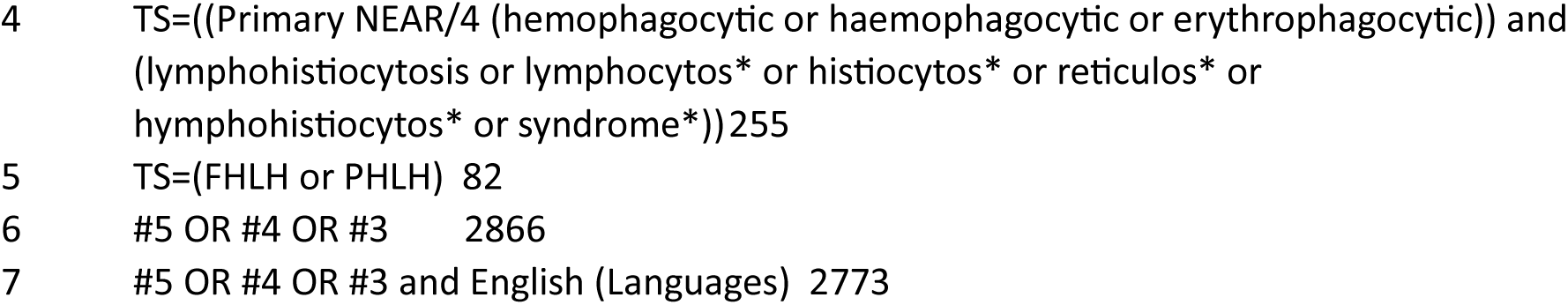

###### Cochrane library

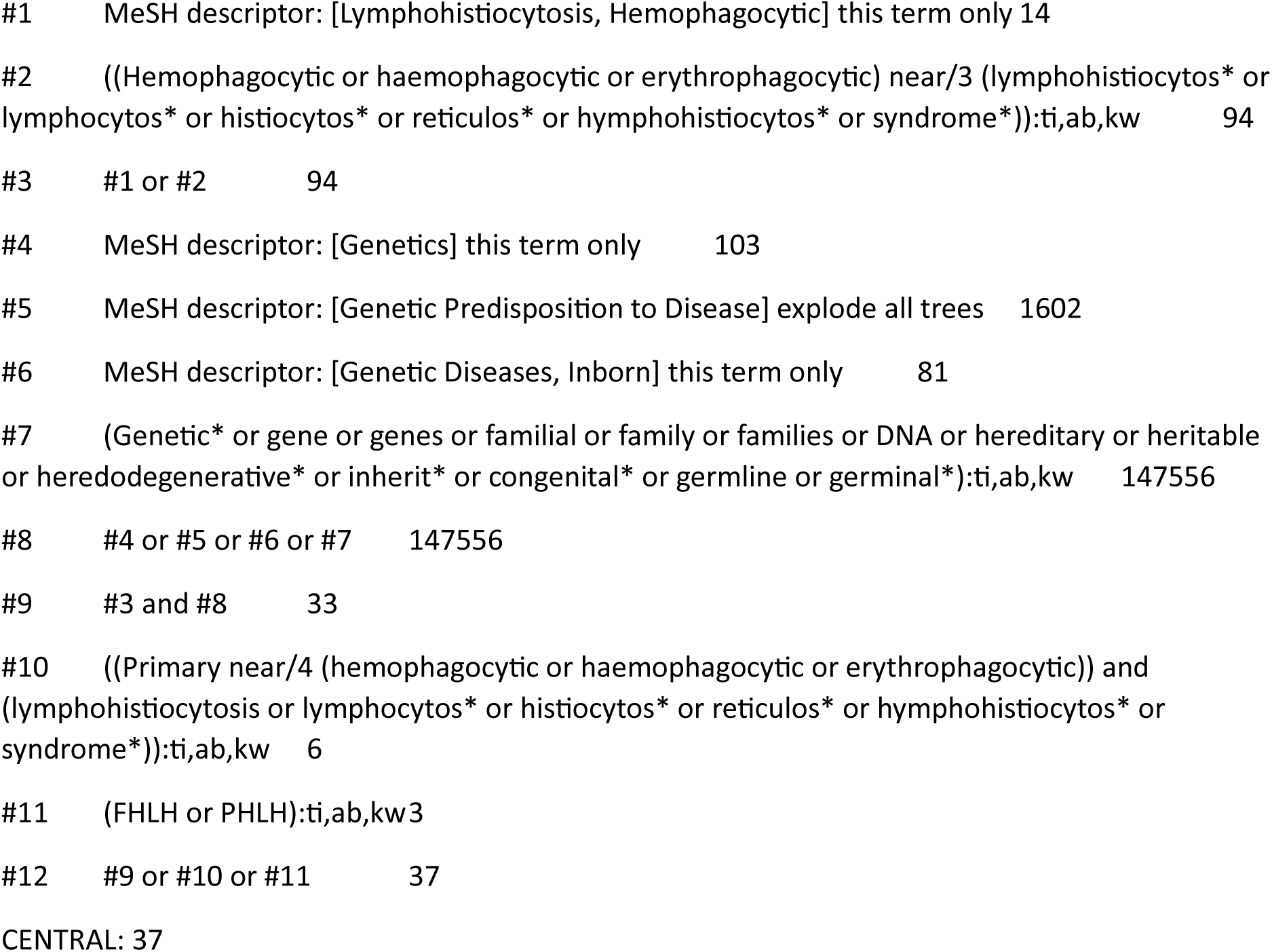

##### MCADD

###### Search summary

| Database (Platform) | Date searched | Concepts | Hits | Notes |
| --- | --- | --- | --- | --- |
| MEDLINE (Ovid) | 20/09/2023 | MCADD | 931 | Removed animal studies<br>English language limit |
| Embase (Ovid) | 20/09/2023 | MCADD | 1750 | Removed animal studies<br>No Emtree terms focussed<br>English language limit |
| Science Citation Index (Web of Science) | 20/09/2023 | MCADD | 1813 | No date or language limits. |
| Cochrane Library | 20/09/2023 | MCADD | 17 | No date or language limits. |

Total pre-duplication: 4511
Total after systematically removing duplicates in EndNote (using University of Leeds method)<sup>17</sup>: 2962

###### Ovid MEDLINE(R) ALL 1946 to September 18, 2023

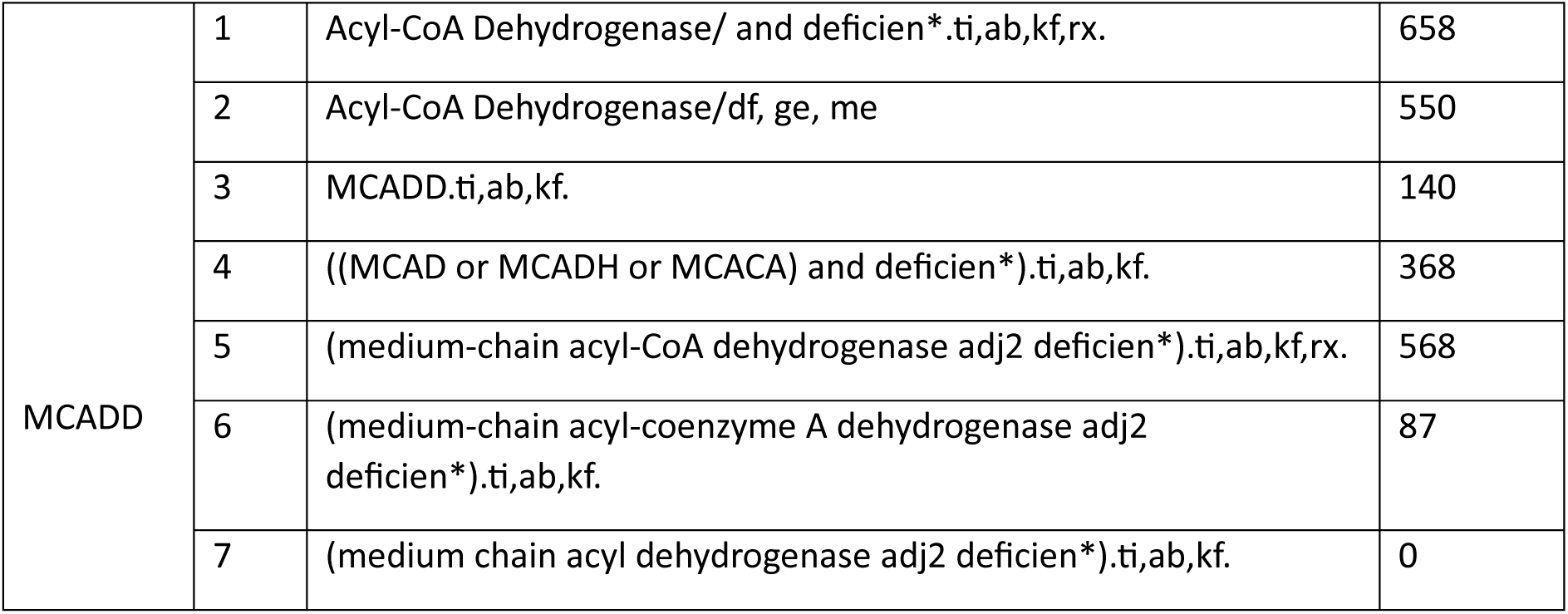
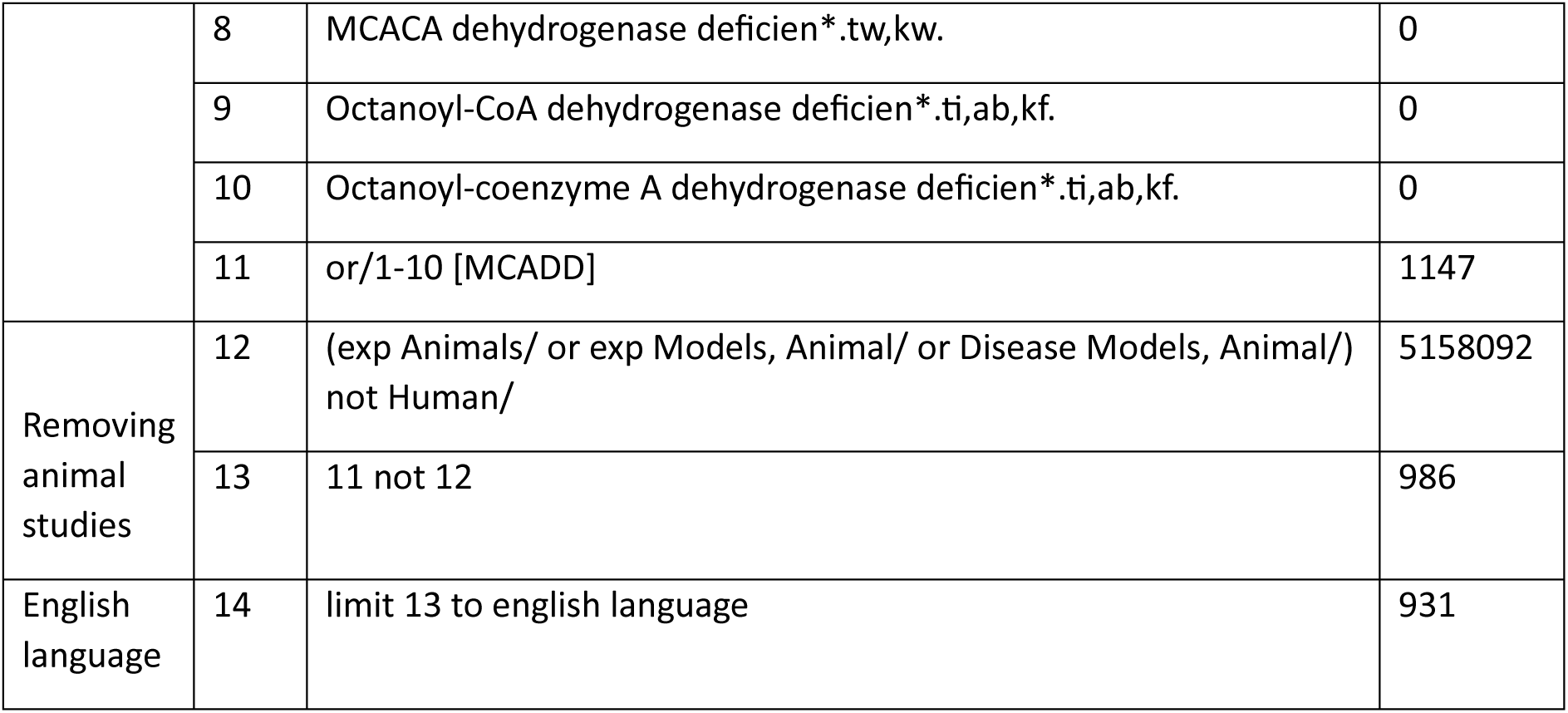

###### Embase Classic+Embase 1947 to 2023 September 19

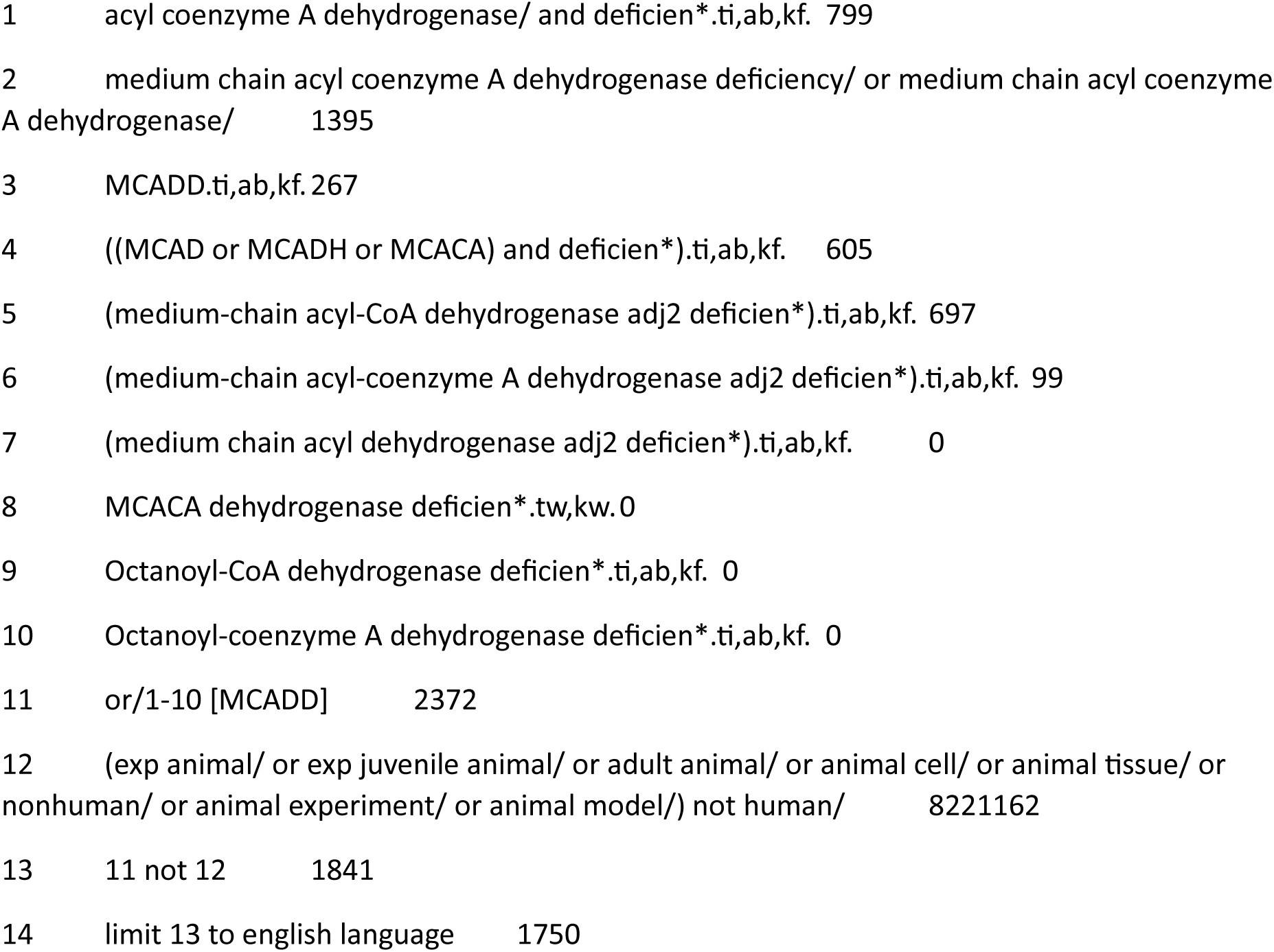

###### Web of Science - Science Citation Index Expanded (SCI-EXPANDED) 1970-present, Social Sciences Citation Index (SSCI) 1900-present

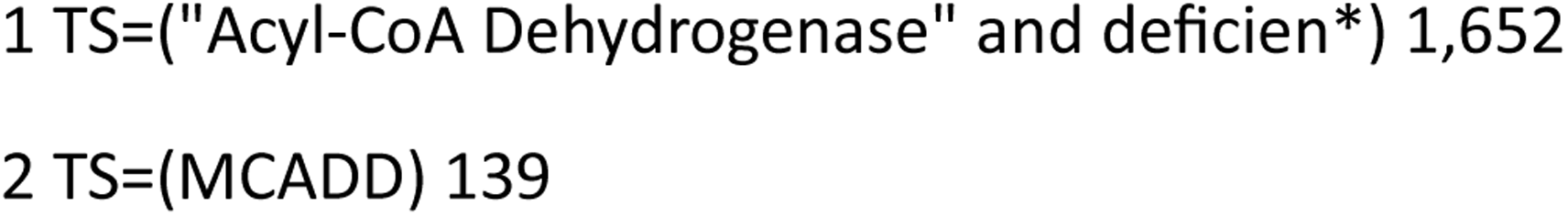
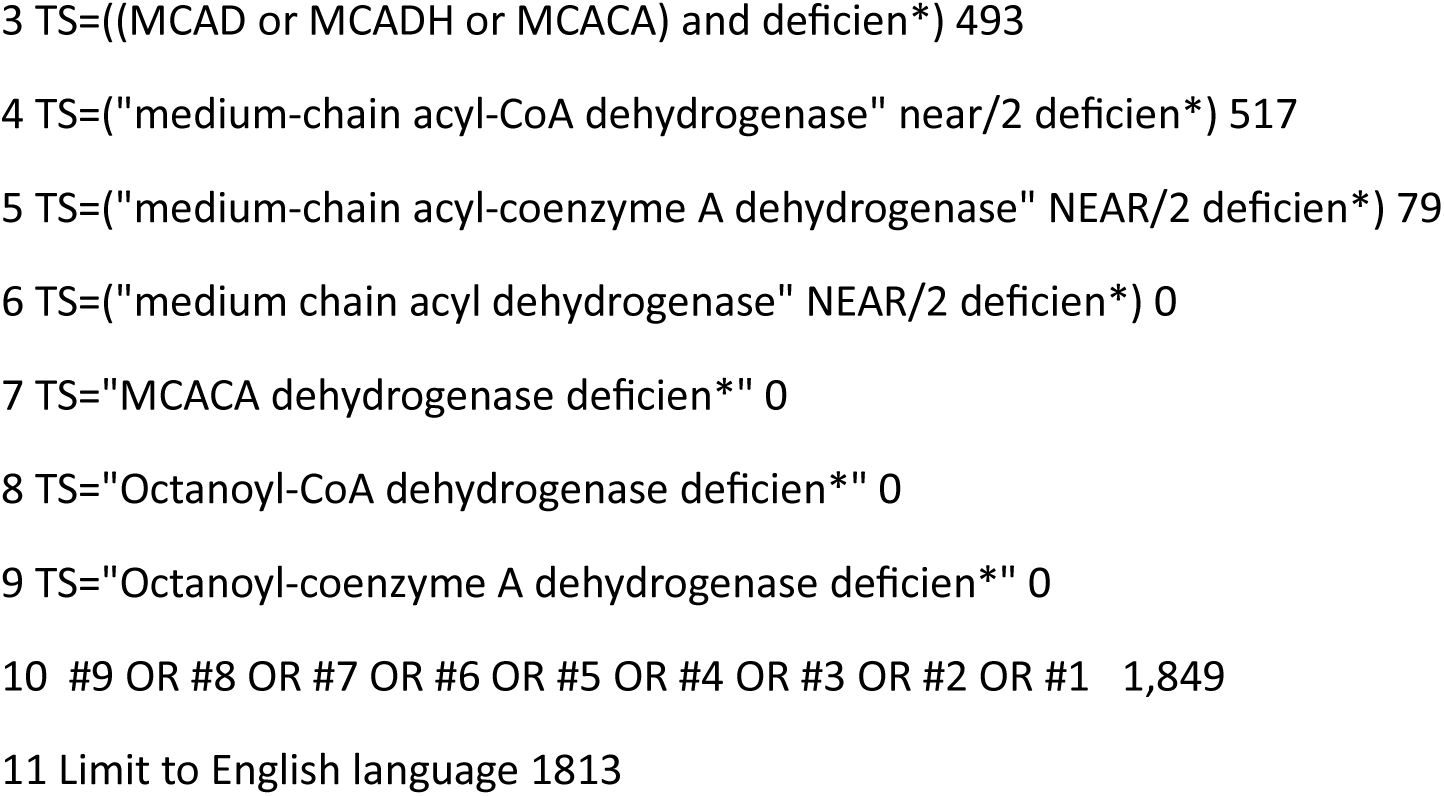

###### Cochrane library

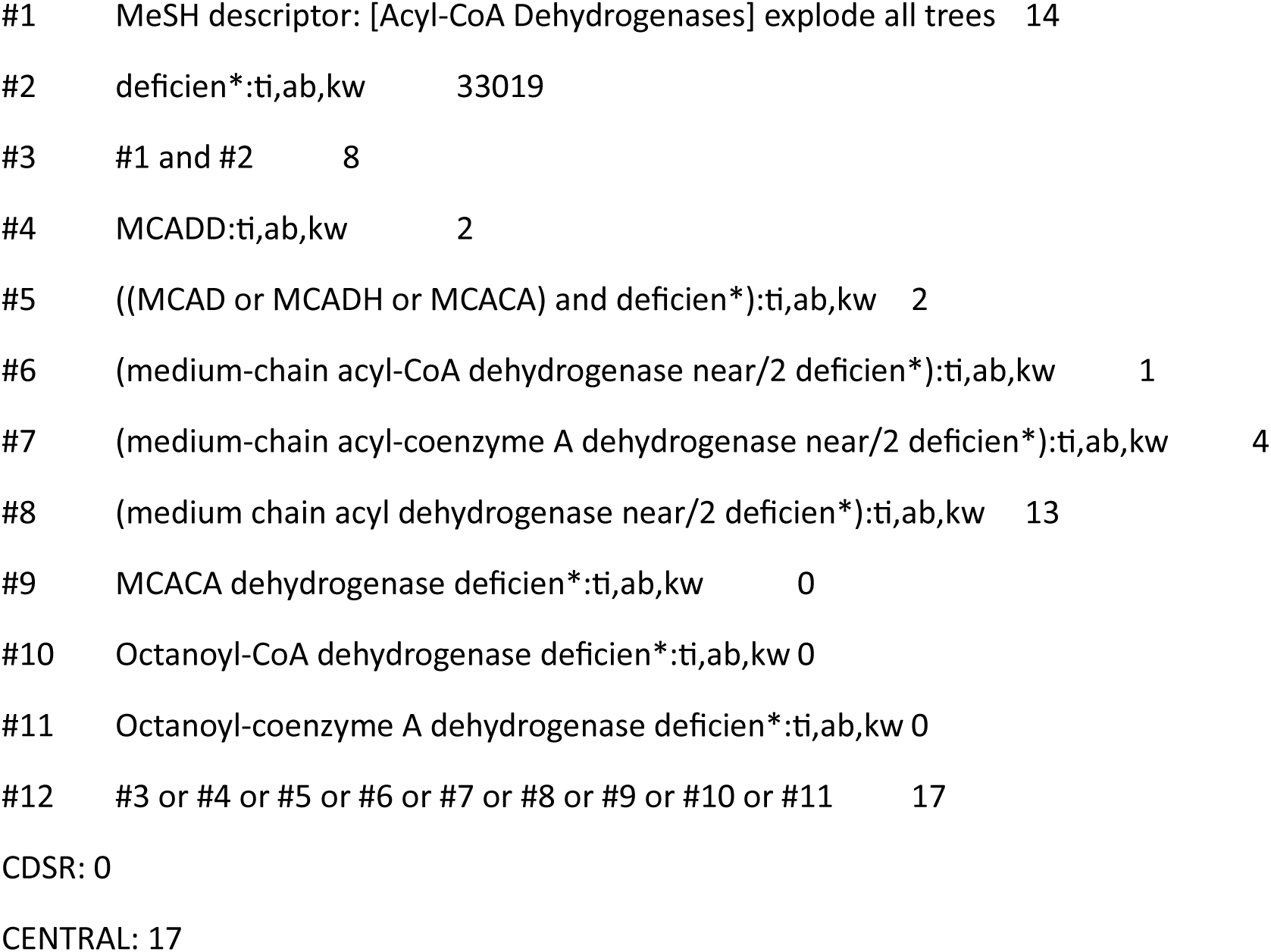

##### Review of genomic studies of paediatric cohorts reporting penetrance for pathogenic variants

###### Search summary

| Database (Platform) | Date searched | Concepts | Hits | Notes |
| --- | --- | --- | --- | --- |
| MEDLINE (Ovid) | 18/01/2024 | WGS/WES/Penetrance/Actionability/<br>Gene frequency/Sequencing/Allele frequency<br>AND<br>Newborn screening | 3019 | No date or language limits. Found 5 known studies |
| Embase (Ovid) | 19/01/2024 | WGS/WES/Penetrance/Actionability/<br>Gene frequency/Sequencing/Allele frequency<br>AND<br>Newborn screening | 3764 | Removed conference abstracts. No date or language limits. No Emtree headings focussed. |
| Science Citation Index (Web of Science) | 19/01/2024 | WGS/WES/Penetrance/Actionability/<br>Gene frequency/Sequencing/Allele frequency<br>AND<br>Newborn screening | 2076 | No date or language limits. |
| Other? Cochrane Library | 19/01/2024 | WGS/WES/Penetrance/Actionability/<br>Gene frequency/Sequencing/Allele frequency<br>AND<br>Newborn screening | 133 | No date or language limits. |

Total from database searches: 8992
Total after systematically removing duplicates in EndNote (using University of Leeds method)<sup>17</sup>: 4970

###### MEDLINE

Date searched: 18/01/2024

Database segment: Ovid MEDLINE(R) ALL <1946 to January 17, 2024>

https://ovidsp.ovid.com/ovidweb.cgi?T=JS&NEWS=N&PAGE=main&SHAREDSEARCHID=62stKMwQjirbNXTZKB33Ey3QPoKWoPeo9lIbQ7b1QNoyW2UjVAZu6ArPkcdDGqva7

|  |  |  |  |
| --- | --- | --- | --- |
| Whole Genome Sequencing (WGS) or Whole Exome Sequencing (WES) | 1 | Whole Genome Sequencing/ | 9616 |
|  | 2 | Exome Sequencing/ | 8205 |
|  | 3 | (whole genome sequenc* or complete genome sequenc*).ti,ab,kf. | 45870 |
|  | 4 | WGS.ti,ab,kf. | 8462 |
|  | 5 | (whole exome sequenc* or complete exome sequenc*).ti,ab,kf. | 19981 |
|  | 6 | WES.ti,ab,kf. | 6937 |
|  | 7 | genomic sequenc*.ti,ab,kf. | 20077 |
|  | 8 | Genomics/ | 68324 |
|  | 9 | 1 or 2 or 3 or 4 or 5 or 6 or 7 or 8 [Whole Genome Sequencing (WGS) or Whole Exome Sequencing (WES)] | 153084 |
| Newborn screening | 10 | Neonatal Screening/ | 12112 |
|  | 11 | ((newborn* or neonat* or infan* or baby or babies) adj5 (screen* or test*)).ti,ab,kf. | 36554 |
|  | 12 | Mass Screening/ | 117096 |
|  | 13 | exp infant/ or infant disease*.ti,ab,kf. or (babies or baby or infan* or neonat* or neo-nat* or newborn* or new-born*).ti,ab,kf. or infan*.jn,jw. or infan*.in. | 1663069 |
|  | 14 | 12 and 13 | 10501 |
|  | 15 | 10 or 11 or 14 [Newborn screening] | 48363 |
| Penetrance/Actionability | 16 | 9 and 15 [WGS/WES AND Newborn screening] | 528 |
|  | 17 | Penetrance/ | 2542 |
|  | 18 | penetrance.ti,ab,kf. | 14501 |
|  | 19 | ((("proportion of carriers" or "proportion of individuals") adj10 phenotyp*).ti,ab,kf. | 22 |
|  | 20 | actionability.ti,ab,kf. | 894 |
|  | 21 | 17 or 18 or 19 or 20 [Penetrance/Actionability] | 16140 |
|  | 22 | 15 and 21 [Penetrance/actionability AND Newborn screening] | 73 |
| Sequencing | 23 | Sequence Analysis/ or exp High-Throughput Nucleotide Sequencing/ or exp Molecular Sequence | 499723 |
|  |  | Annotation/ or exp Nanopore Sequencing/ or exp Oligonucleotide Array Sequence Analysis/ or exp Position-Specific Scoring Matrices/ or exp Sequence Analysis, DNA/ |  |
|  | 24 | sequenc*.ti,ab,kf. | 1482656 |
|  | 25 | 23 or 24 [Sequencing] | 1706387 |
|  | 26 | 15 and 25 [Sequencing AND Newborn screening] | 2656 |
| Gene frequency | 27 | exp Gene Frequency/ | 84475 |
|  | 28 | ((allele* or gene*) adj3 frequenc*).ti,ab,kf. | 62822 |
|  | 29 | 27 or 28 | 123910 |
|  | 30 | 15 and 29 [Gene frequency and Newborn screening] | 396 |
|  | 31 | 16 or 22 or 26 | 2765 |
|  | 32 | 16 or 22 or 26 or 30 | 3019 |

###### Embase

Date searched: 19/01/2024

Database segment: Embase Classic+Embase <1947 to 2024 Week 02>

https://ovidsp.ovid.com/ovidweb.cgi?T=JS&NEWS=N&PAGE=main&SHAREDSEARCHID=4zKpZuDwhGs6myjL7S8Z46JqaFTuk80EWrwhr8tsaG6QSMVKpxP5critdRCUddTRd

Embase Classic+Embase <1947 to 2024 Week 02>

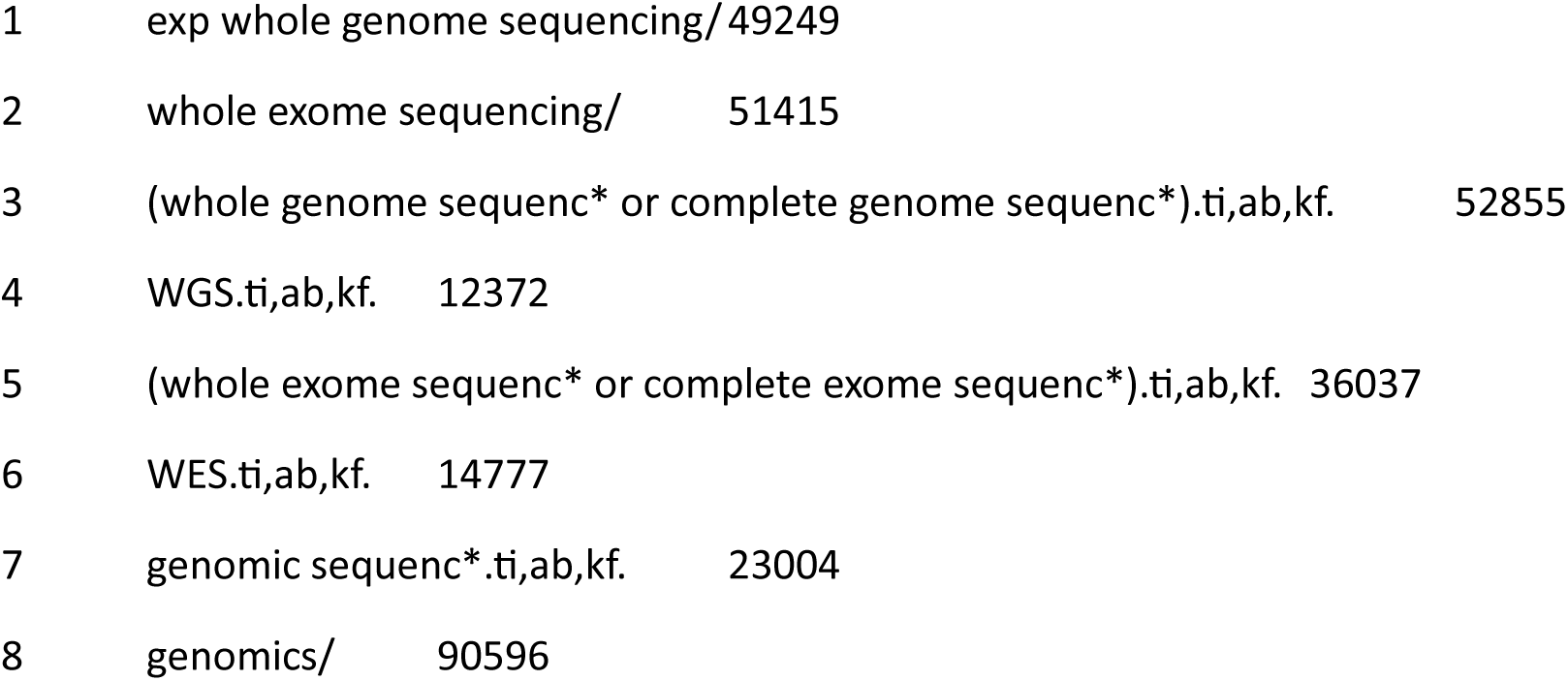
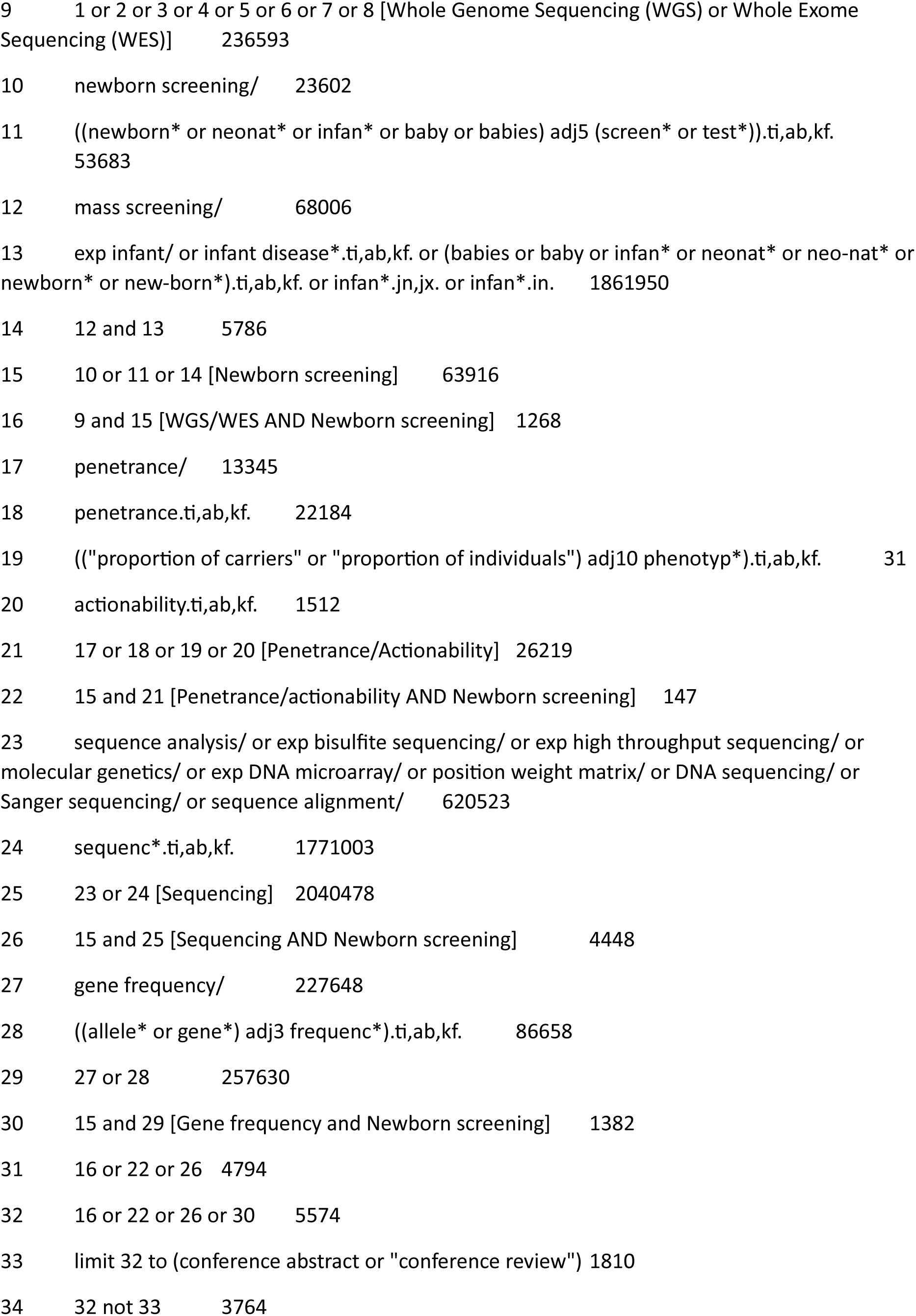

###### Science Citation Index (Web of Science)

Date searched: 19/01/2024

Note: search reads from bottom to top.

|  |  |  |
| --- | --- | --- |
| 18 | #8 OR #13 OR #15 OR #17 | 2,076 |
| 17 | #7 AND #16 | 194 |
| 16 | TS= ((allele* OR gene*) NEAR/2 frequenc*) | 82,970 |
| 15 | #7 AND #14 | 1,901 |
| 14 | TS=sequenc* | 2,047,808 |
| 13 | #12 AND #7 | 62 |
| 12 | #9 OR #10 OR #11 | 15,124 |
| 11 | TS=actionability | 881 |
| 10 | TS= (("proportion of carriers" OR "proportion of individuals") NEAR/10 phenotyp*) | 22 |
| 9 | TS=penetrance | 14,255 |
| 8 | #6 AND #7 | 318 |
| 7 | TS= ((newborn* OR neonat* OR infan* OR baby OR babies) NEAR/5 (screen* OR test*)) | 32,897 |
| 6 | #1 OR #2 OR #3 OR #4 OR #5 | 90,839 |
| 5 | TS="genomic sequenc*" | 20,189 |
| 4 | TS=WES | 6,697 |
| 3 | TS= ("whole exome sequenc*" OR "complete exome sequenc*") | 19,578 |
| 2 | TS=WGS | 10,344 |
| 1 | TS= (("whole genome sequenc*" OR "complete genome sequenc*")) | 47,997 |

#### Cochrane Library (Wiley)

Date searched: 19/01/2024

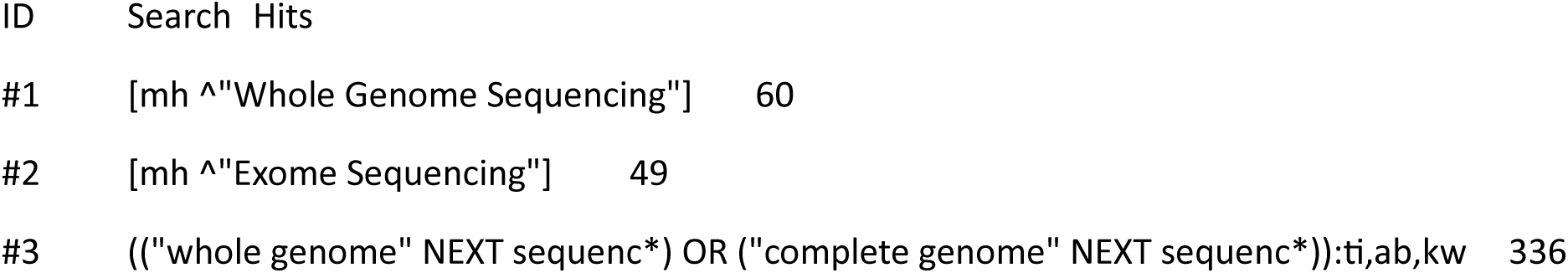
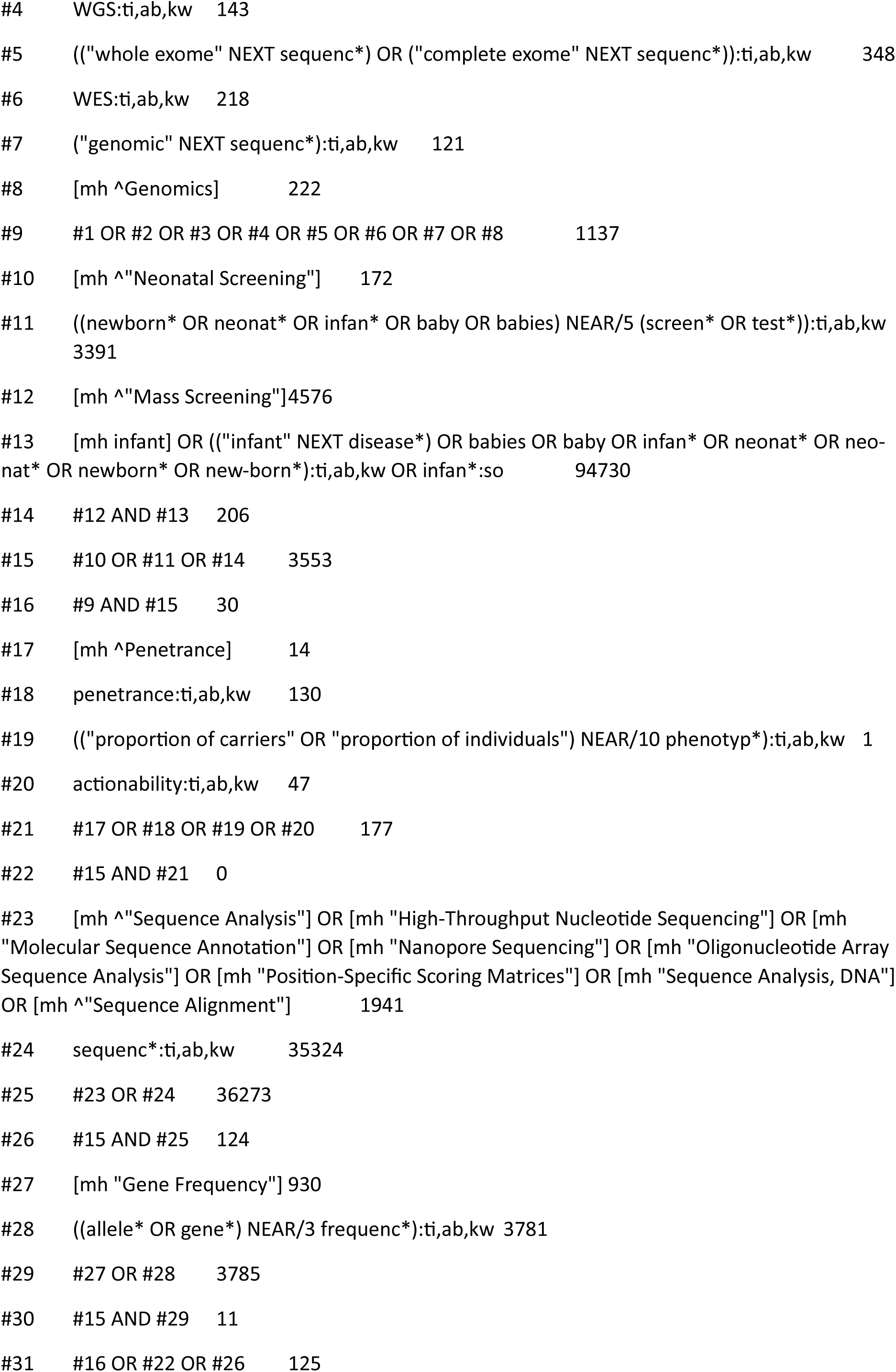
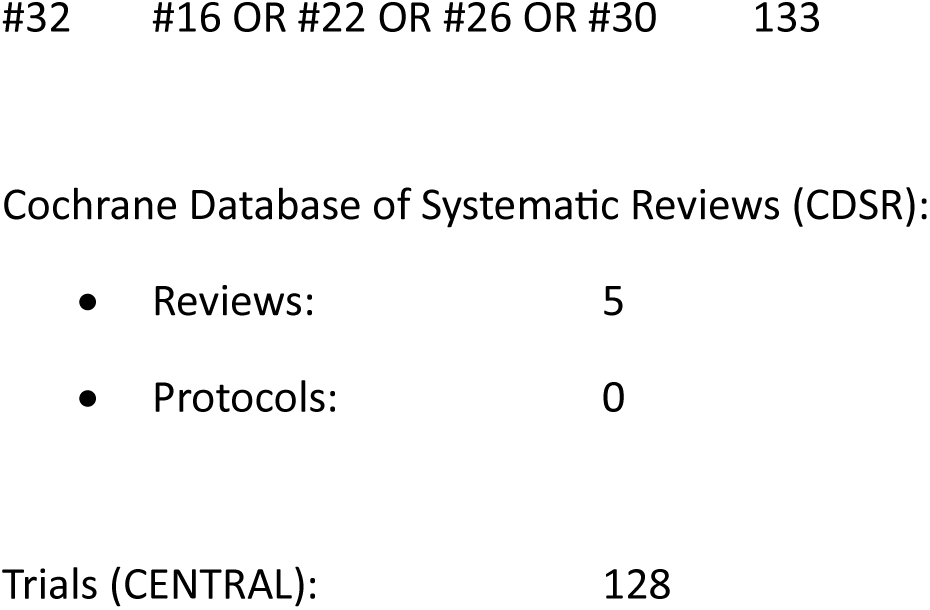

##### Review of cost-effectiveness evaluations of WGS and WES

###### Search summary

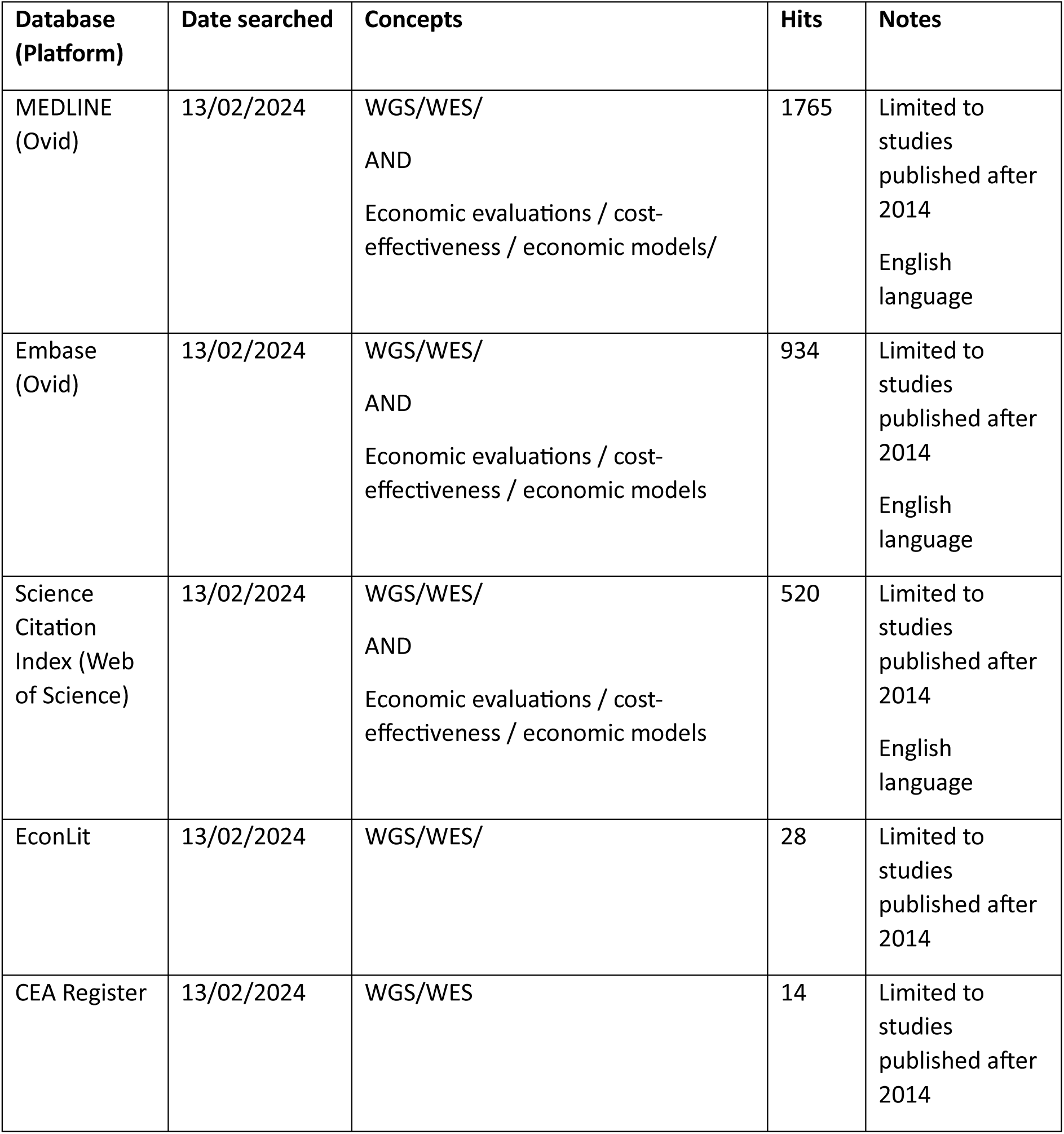

###### Search summary

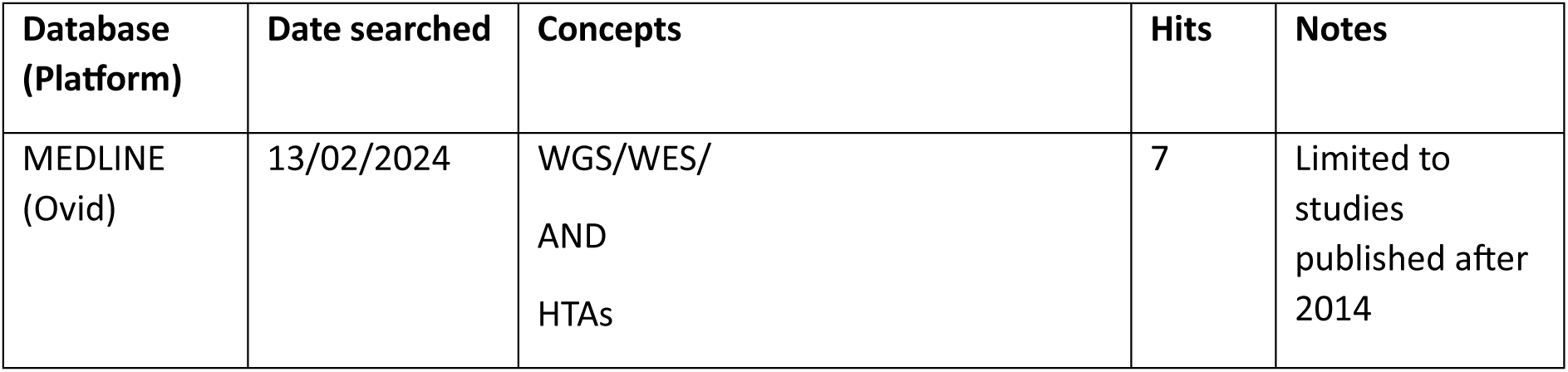
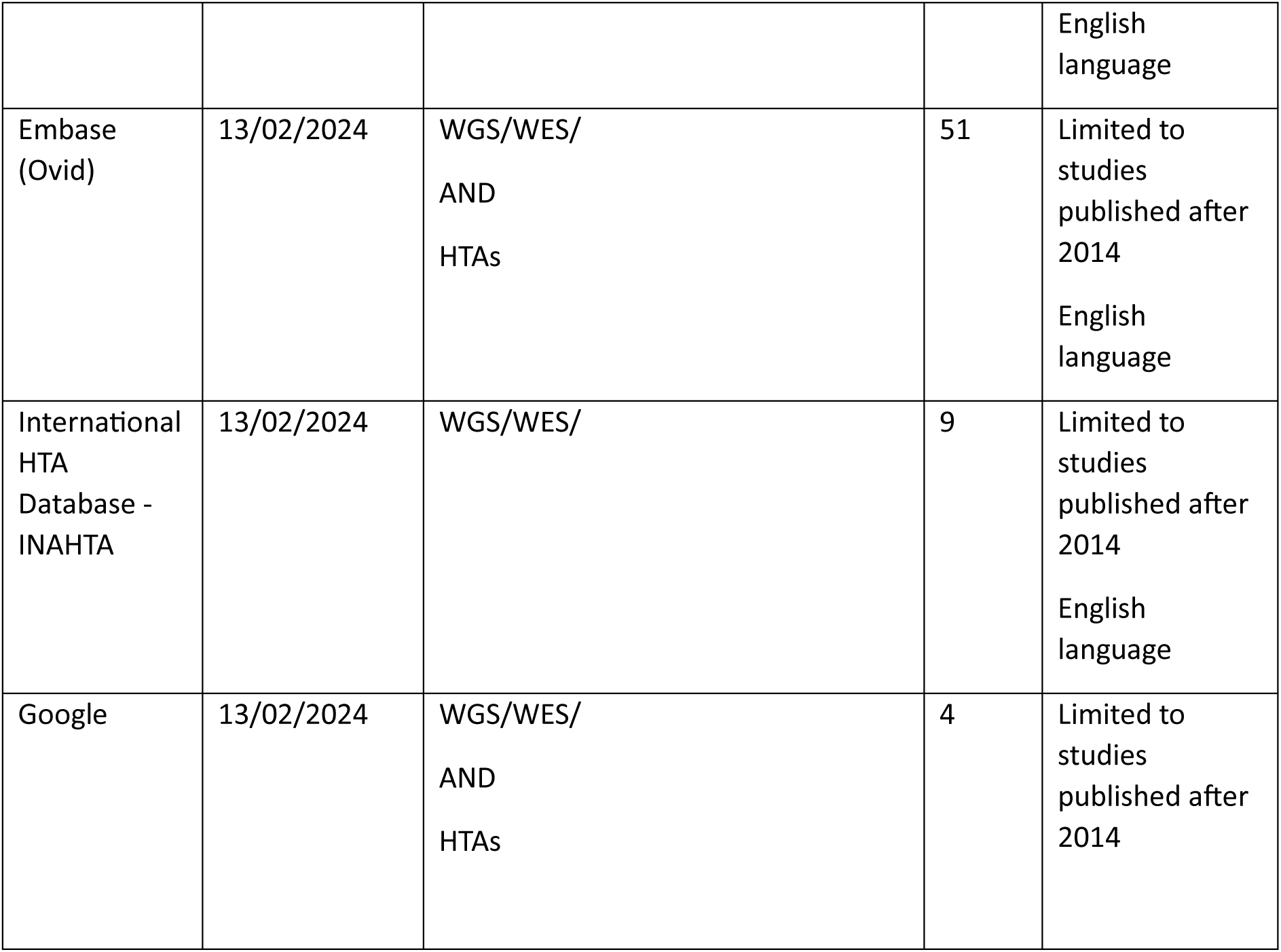

###### Ovid MEDLINE(R) ALL 1946 to February 12, 2024

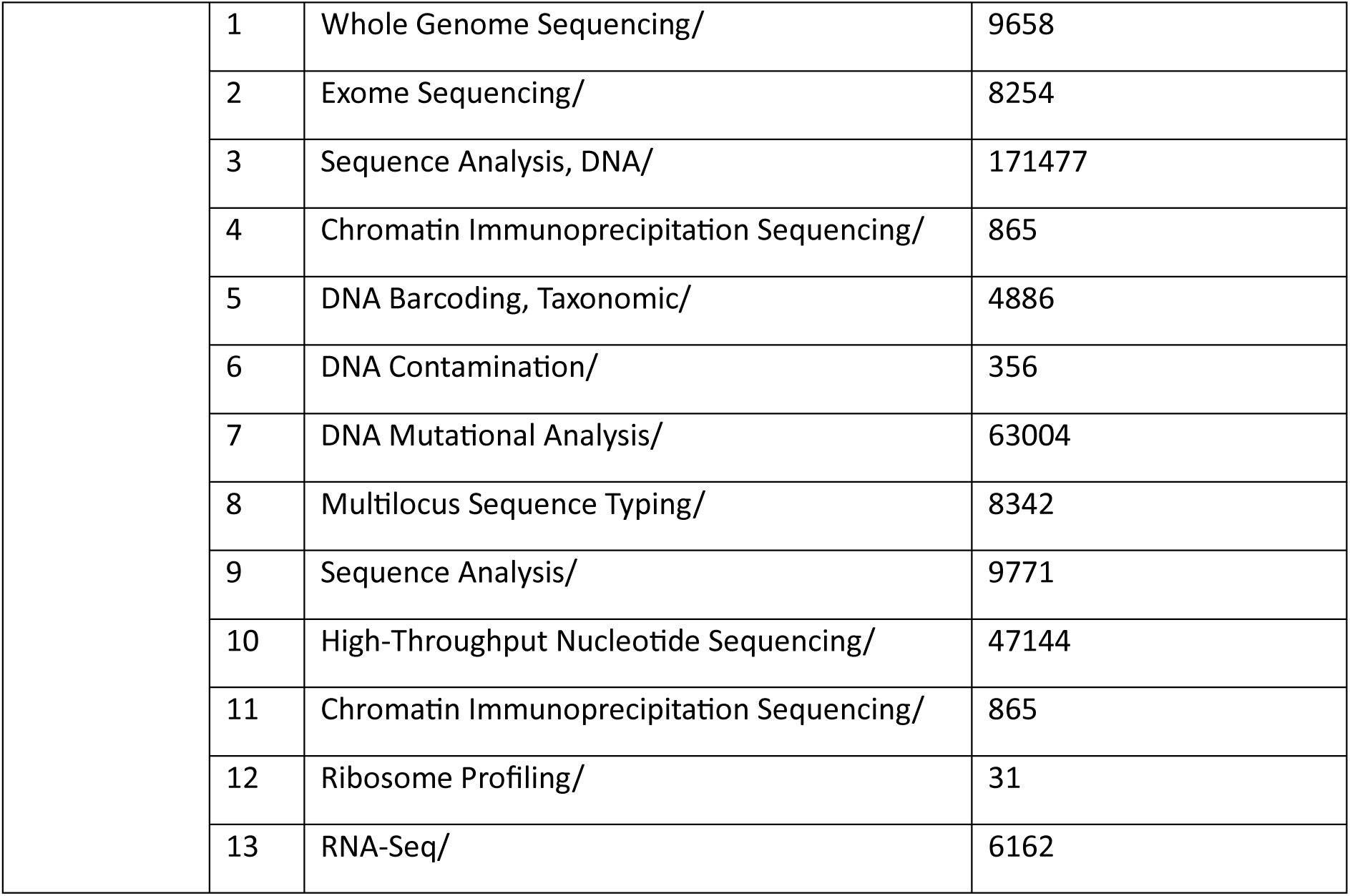
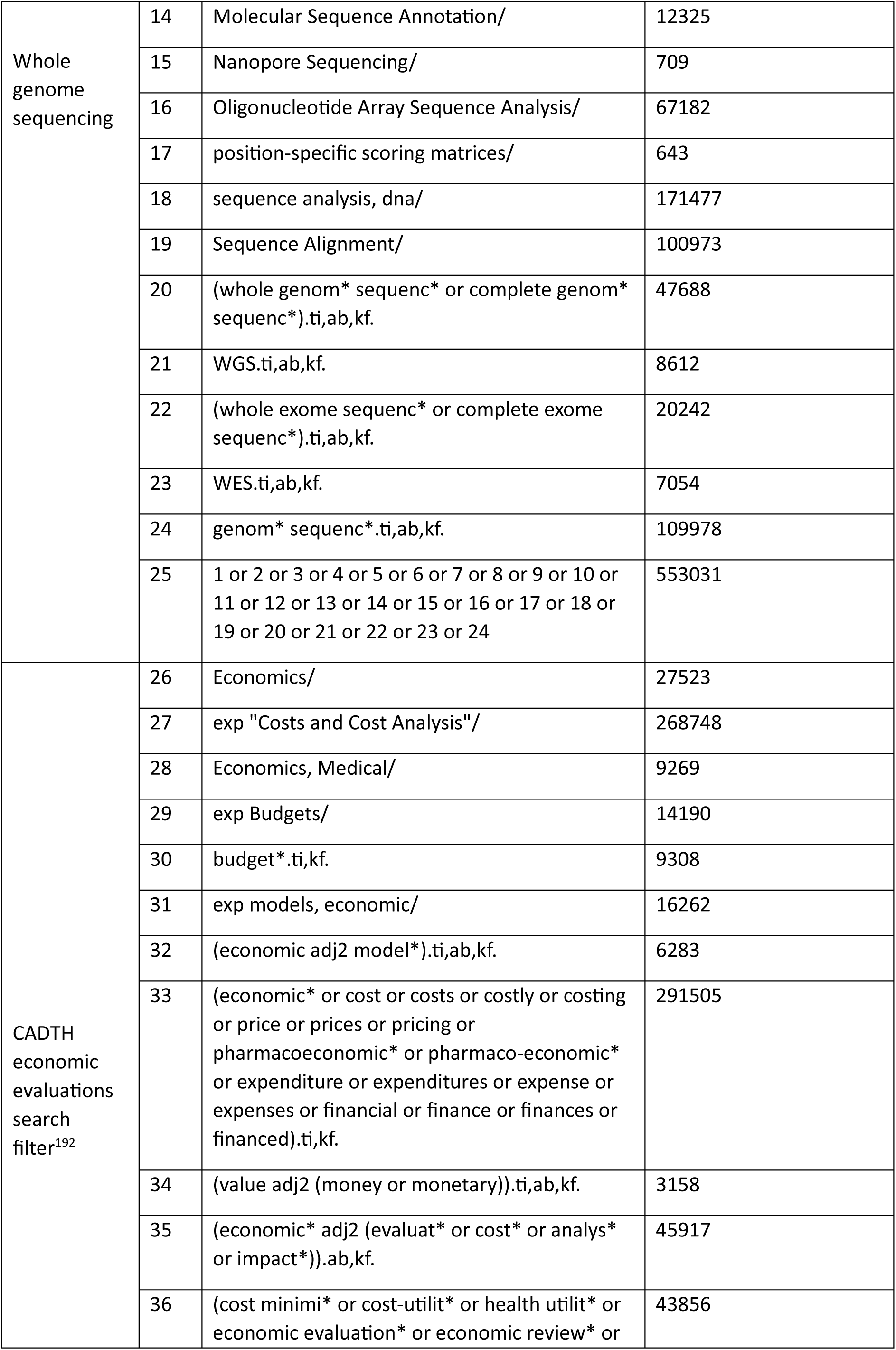
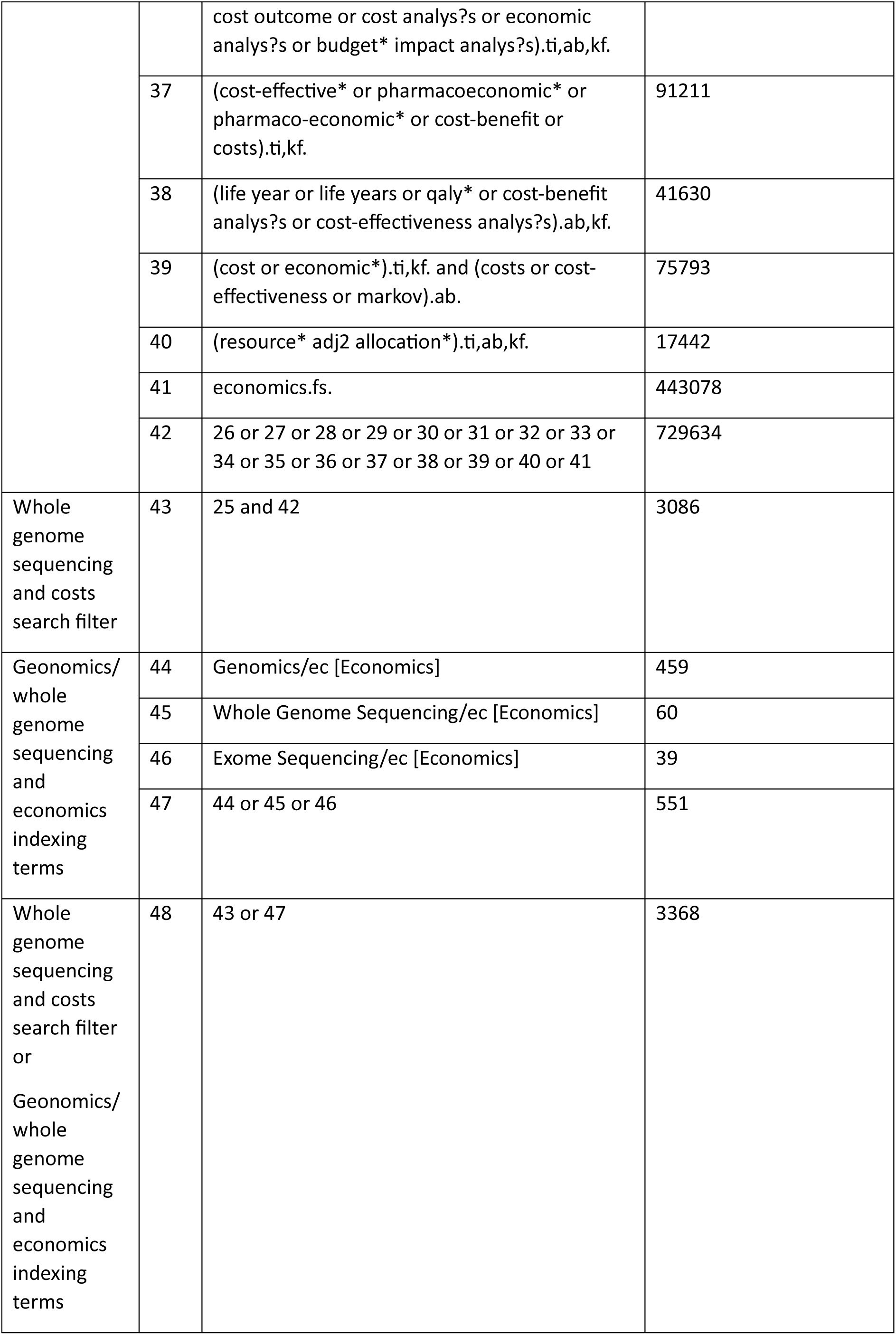
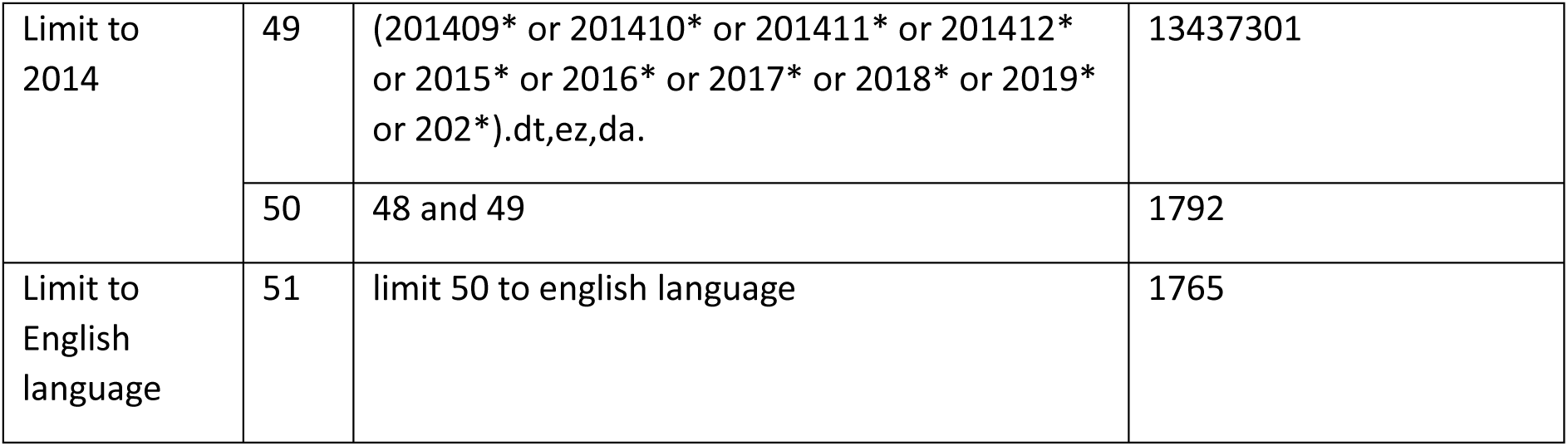

###### Embase Classic+Embase 1947 to 2024 February 13

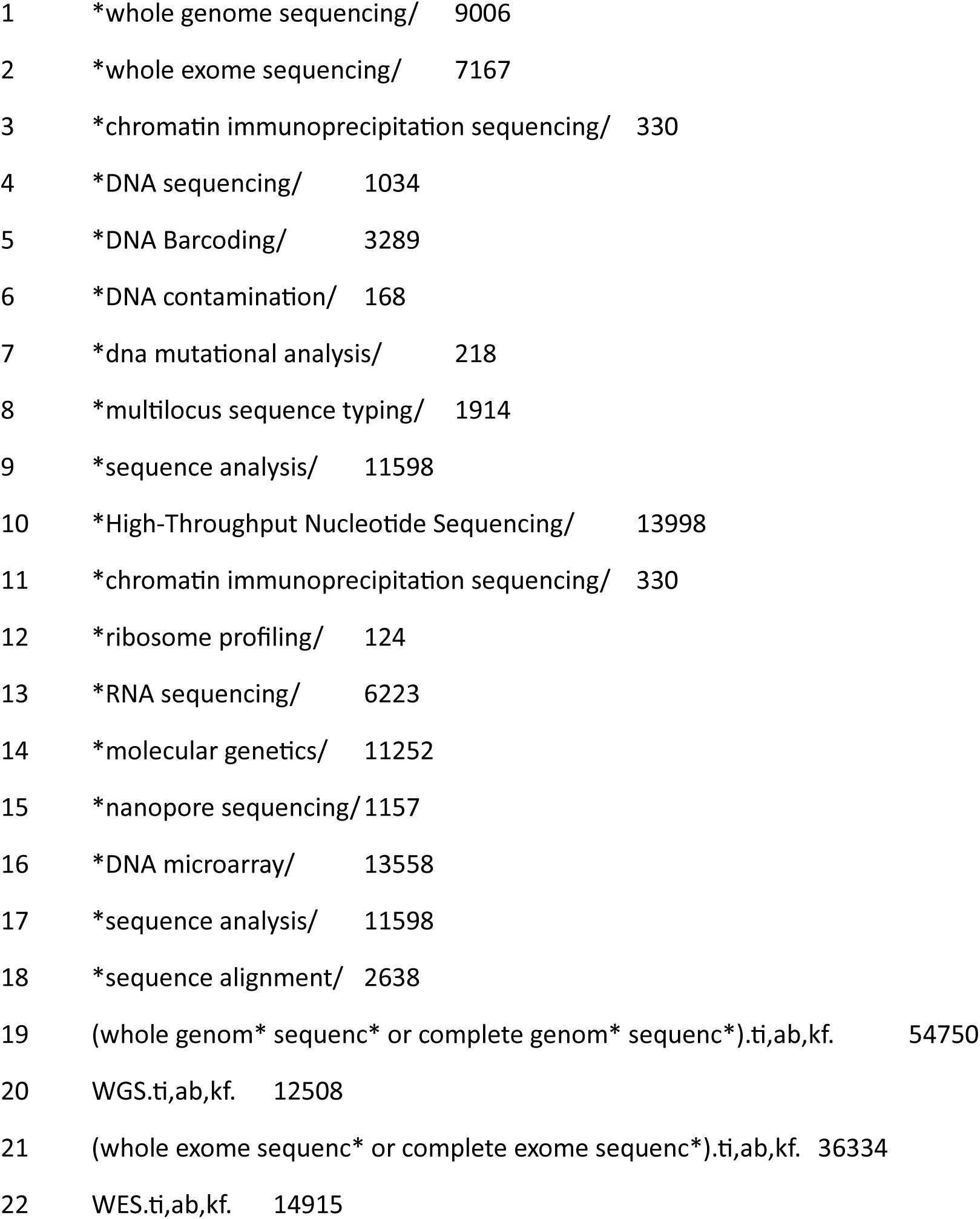
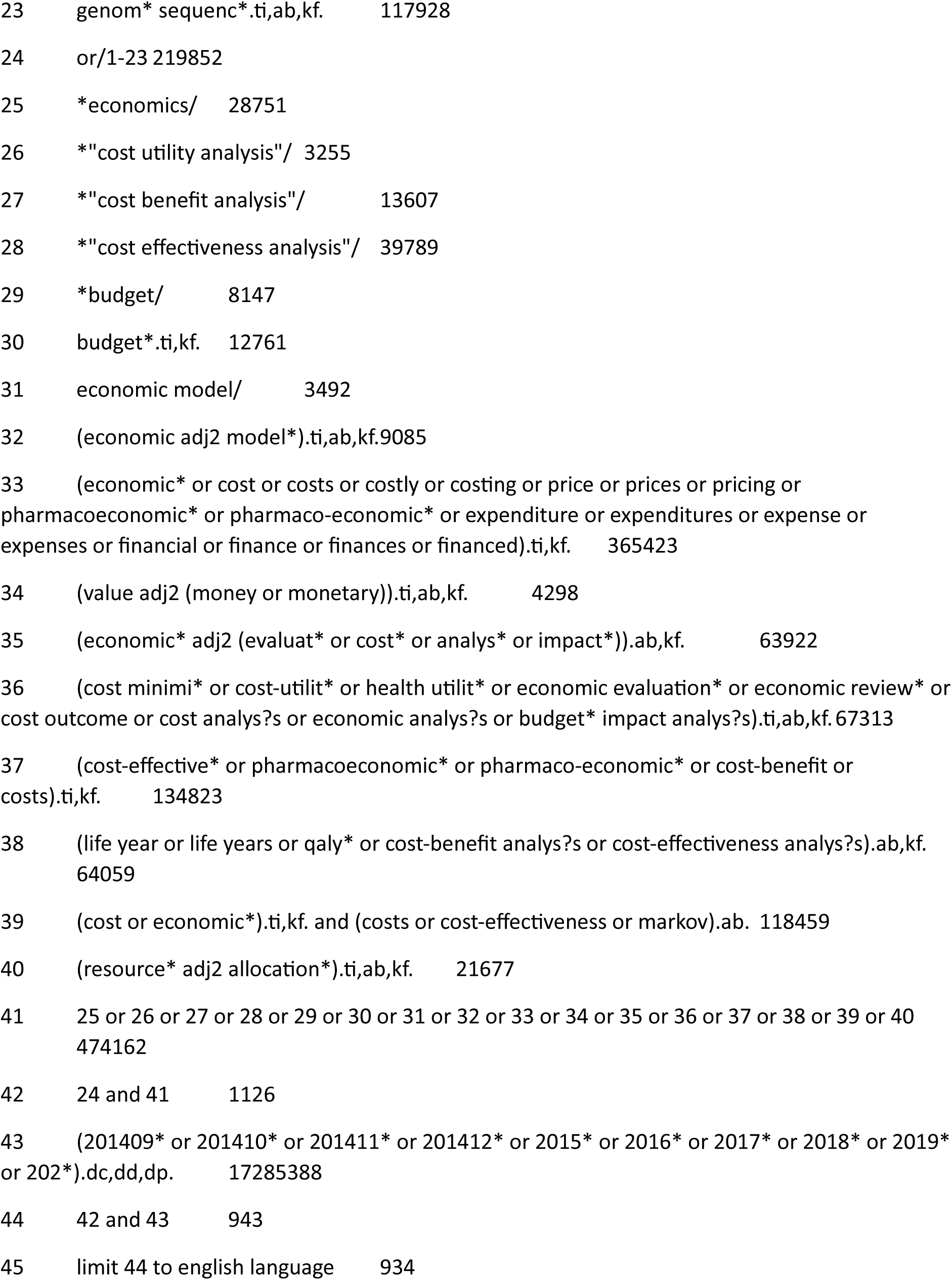

###### Web of Science Search Strategy

Run 14^th^ Feb 2024

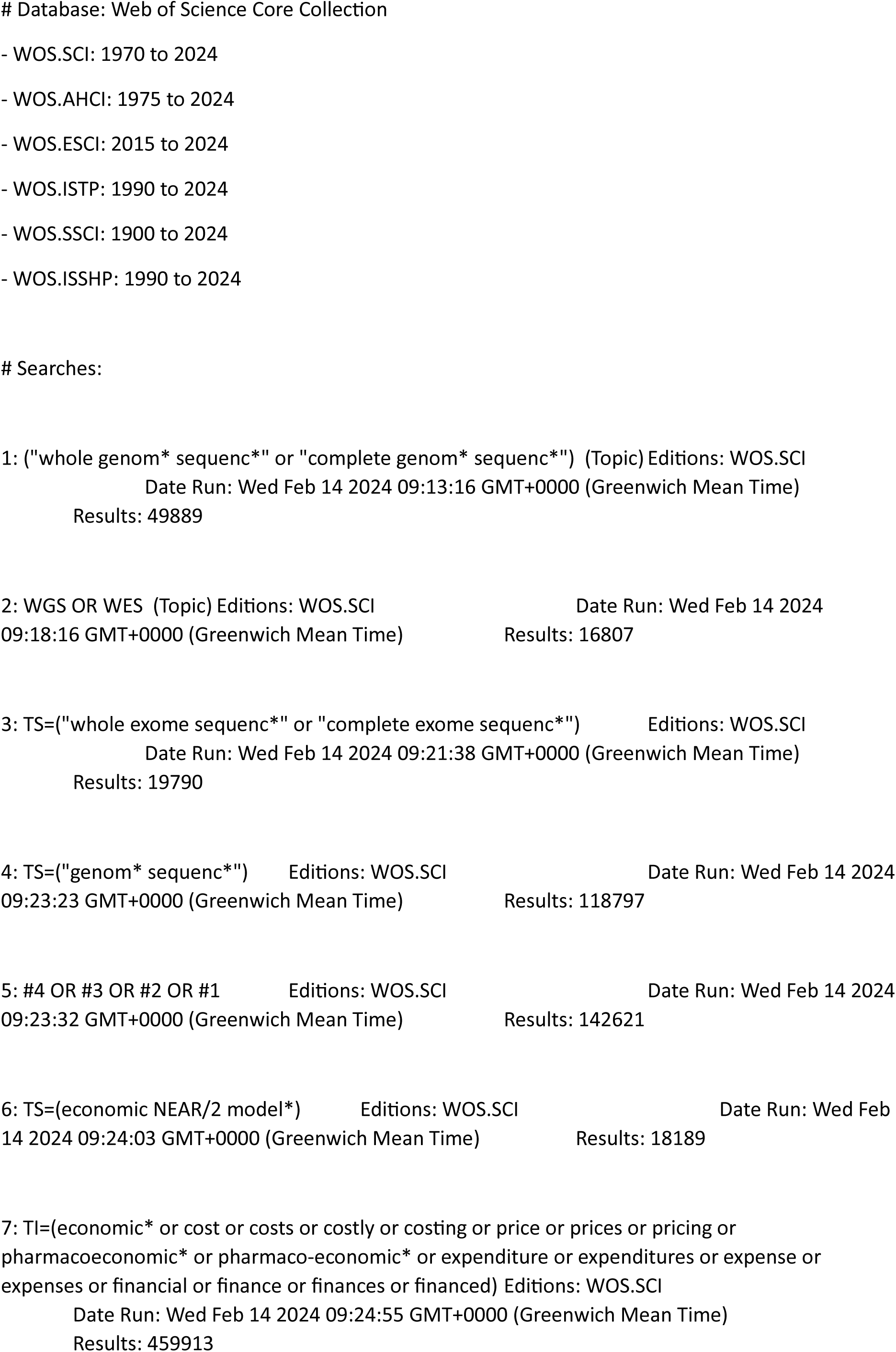
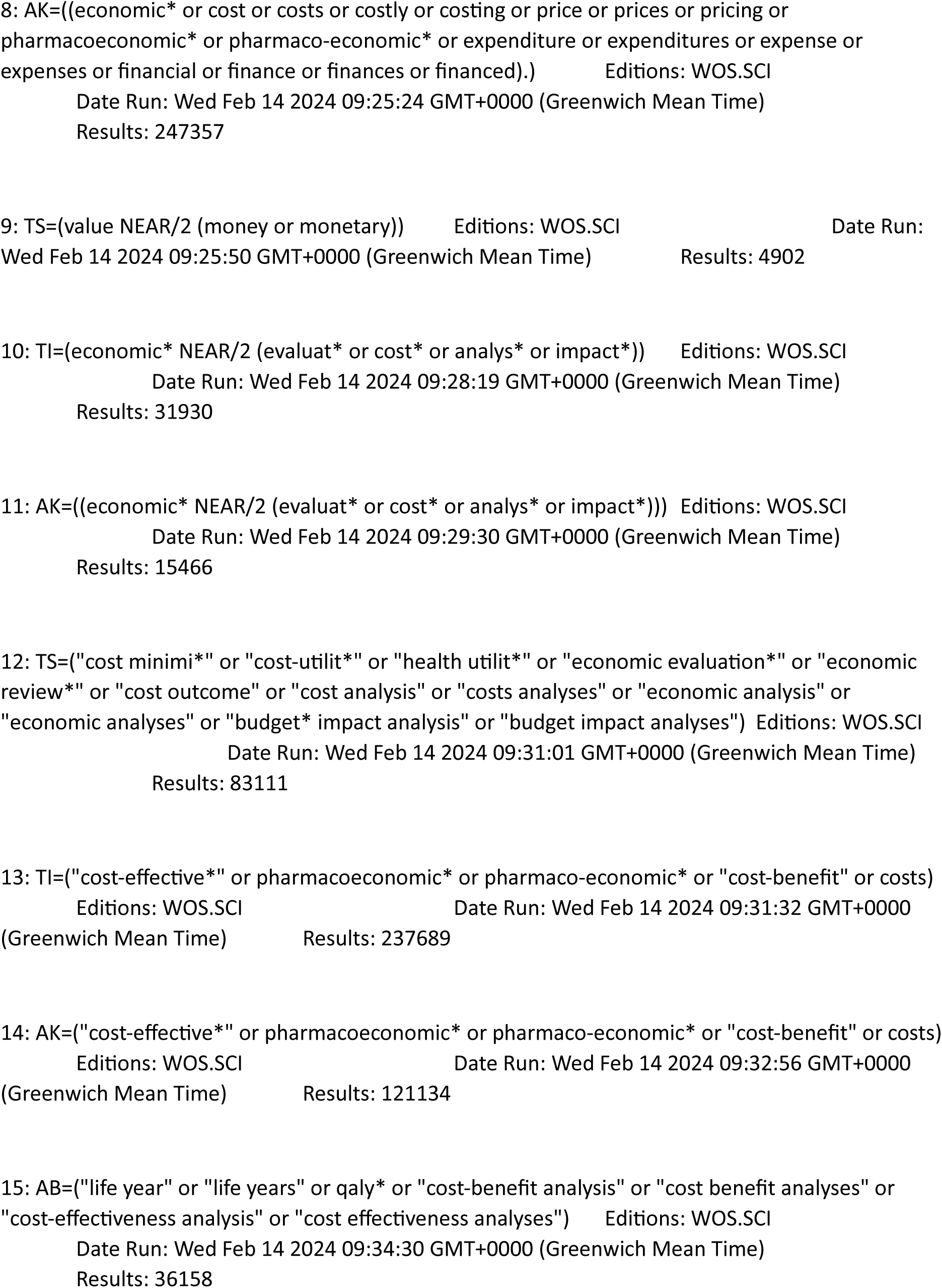
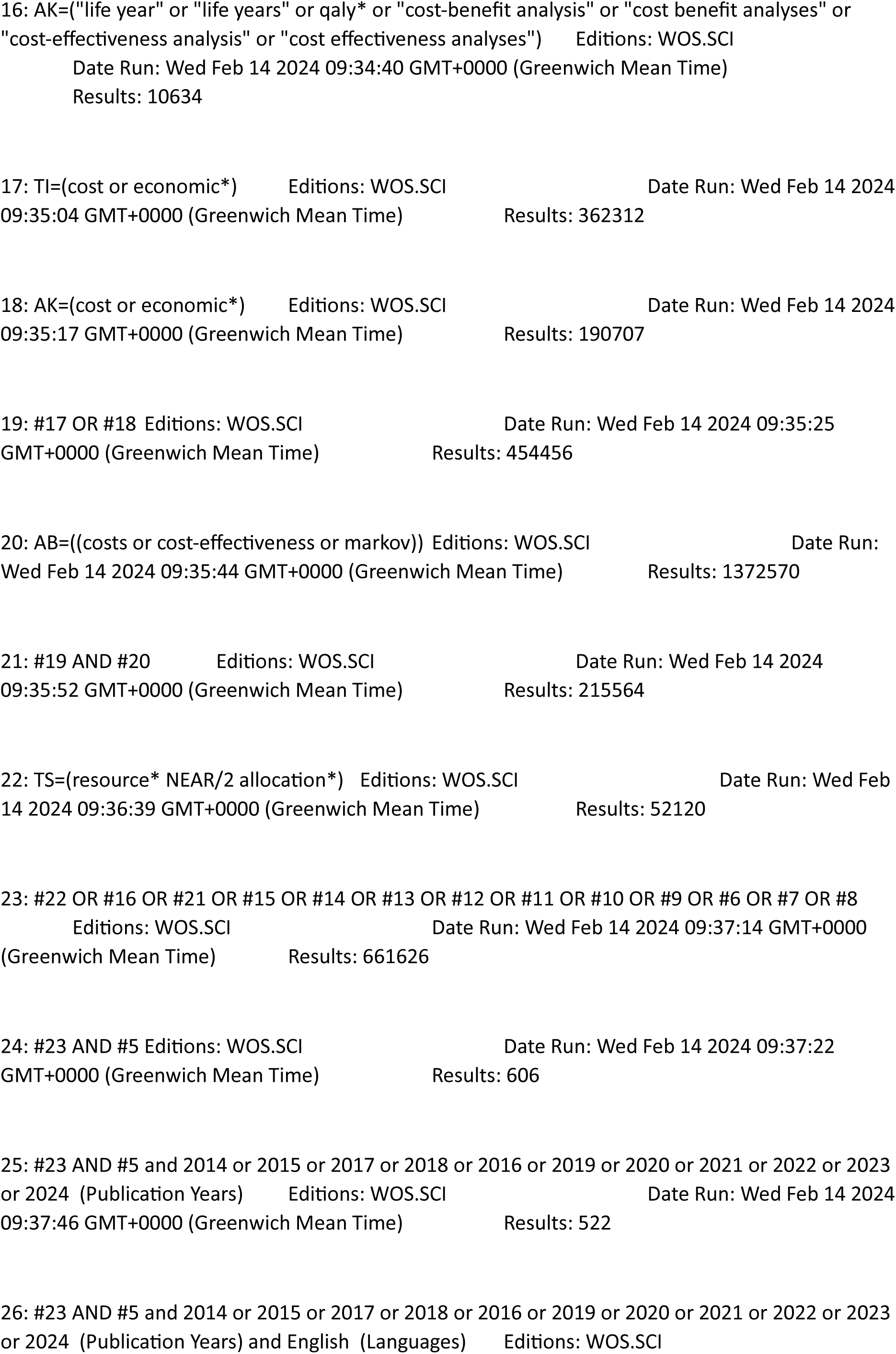
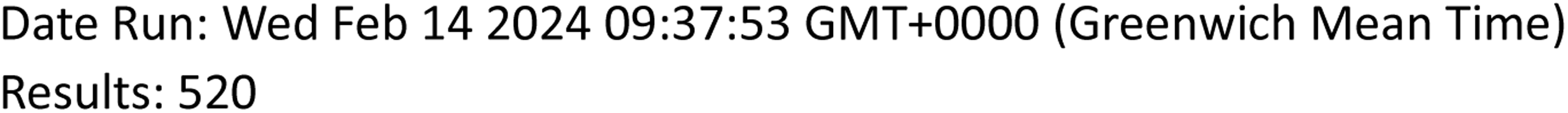

###### EconLit

|  |  |  |
| --- | --- | --- |
| S1 | TI ( ("whole genom* sequenc*" or "complete genom* sequenc*") ) OR AB ( ("whole genom* sequenc*" or "complete genom* sequenc*") ) OR SU ( ("whole genom* sequenc*" or "complete genom* sequenc*") ) | 10 |
| S2 | TI ( (WGS OR WES) ) OR AB ( (WGS OR WES) ) OR SU ( (WGS OR WES) ) | 58 |
| S3 | TI ( ("whole exome sequenc*" or "complete exome sequenc*") ) OR AB ( ("whole exome sequenc*" or "complete exome sequenc*") ) OR SU ( ("whole exome sequenc*" or "complete exome sequenc*") ) | 2 |
| S4 | TI "genom* sequenc*" OR AB "genom* sequenc*" OR SU "genom* sequenc" | 17 |
| S5 | S1 OR S2 OR S3 OR S4 | 71 |
|  | S1 OR S2 OR S3 OR S4 |  |
|  | Limiters - Publication Date: 20140101-20231231 |  |
| S6 |  | 28 |

###### CEA registry

Title, Abstract or Keyword genomic sequencing 3 results

Title, Abstract or Keyword genome 7 results

Title, Abstract or Keyword exome sequencing 3 results

Title, Abstract or Keyword WGS 0 results

Title, Abstract or Keyword WES 1 result

###### Ovid MEDLINE(R) ALL 1946 to February 13, 2024

|  |  |  |
| --- | --- | --- |
| 1 | Whole Genome Sequencing/ | 9662 |
| 2 | Exome Sequencing/ | 8258 |
| 3 | Sequence Analysis, DNA/ | 171483 |
| 4 | Chromatin Immunoprecipitation Sequencing/ | 866 |
| 5 | DNA Barcoding, Taxonomic/ | 4888 |
| 6 | DNA Contamination/ | 356 |
| 7 | DNA Mutational Analysis/ | 63005 |
| 8 | Multilocus Sequence Typing/ | 8343 |
| 9 | Sequence Analysis/ | 9772 |
| 10 | High-Throughput Nucleotide Sequencing/ | 47158 |
| 11 | Chromatin Immunoprecipitation Sequencing/ | 866 |
| 12 | Ribosome Profiling/ | 31 |
| 13 | RNA-Seq/ | 6162 |
| 14 | Molecular Sequence Annotation/ | 12326 |
| 15 | Nanopore Sequencing/ | 709 |
| 16 | Oligonucleotide Array Sequence Analysis/ | 67183 |
| 17 | position-specific scoring matrices/ | 643 |
| 18 | sequence analysis, dna/ | 171483 |
| 19 | Sequence Alignment/ | 100974 |
| 20 | (whole genom* sequenc* or complete genom* sequenc*).ti,ab,kf. | 47719 |
| 21 | WGS.ti,ab,kf. | 8618 |
| 22 | (whole exome sequenc* or complete exome sequenc*).ti,ab,kf. | 20252 |
| 23 | WES.ti,ab,kf. | 7063 |
| 24 | genom* sequenc*.ti,ab,kf. | 110021 |
| 25 | 1 or 2 or 3 or 4 or 5 or 6 or 7 or 8 or 9 or 10 or 11 or 12 or 13 or 14 or 15 or 16 or 17 or 18 or 19 or 20 or 21 or 22 or 23 or 24 | 553104 |
| 26 | exp Technology Assessment, Biomedical/ | 12286 |
| 27 | (technology assessment* or HTA or HTAs or technology overview* or technology appraisal*).ti,ab,kf. | 11825 |
| 28 | (biomedical technology assessment* or bio-medical technology assessment*).mp,hw. | 27 |
| 29 | (health adj2 technology assessment).jw. | 4924 |
| 30 | (cochrane or (health adj2 technology assessment) or evidence report).jw. | 21864 |
| 31 | 26 or 27 or 28 or 29 or 30 | 38796 |
| 32 | 25 and 31 | 115 |
| 33 | (2022* or 2023* or 2024*).dt,ez,da. | 3788244 |
| 34 | 32 and 33 | 7 |

###### Embase Classic+Embase 1947 to 2024 February 13

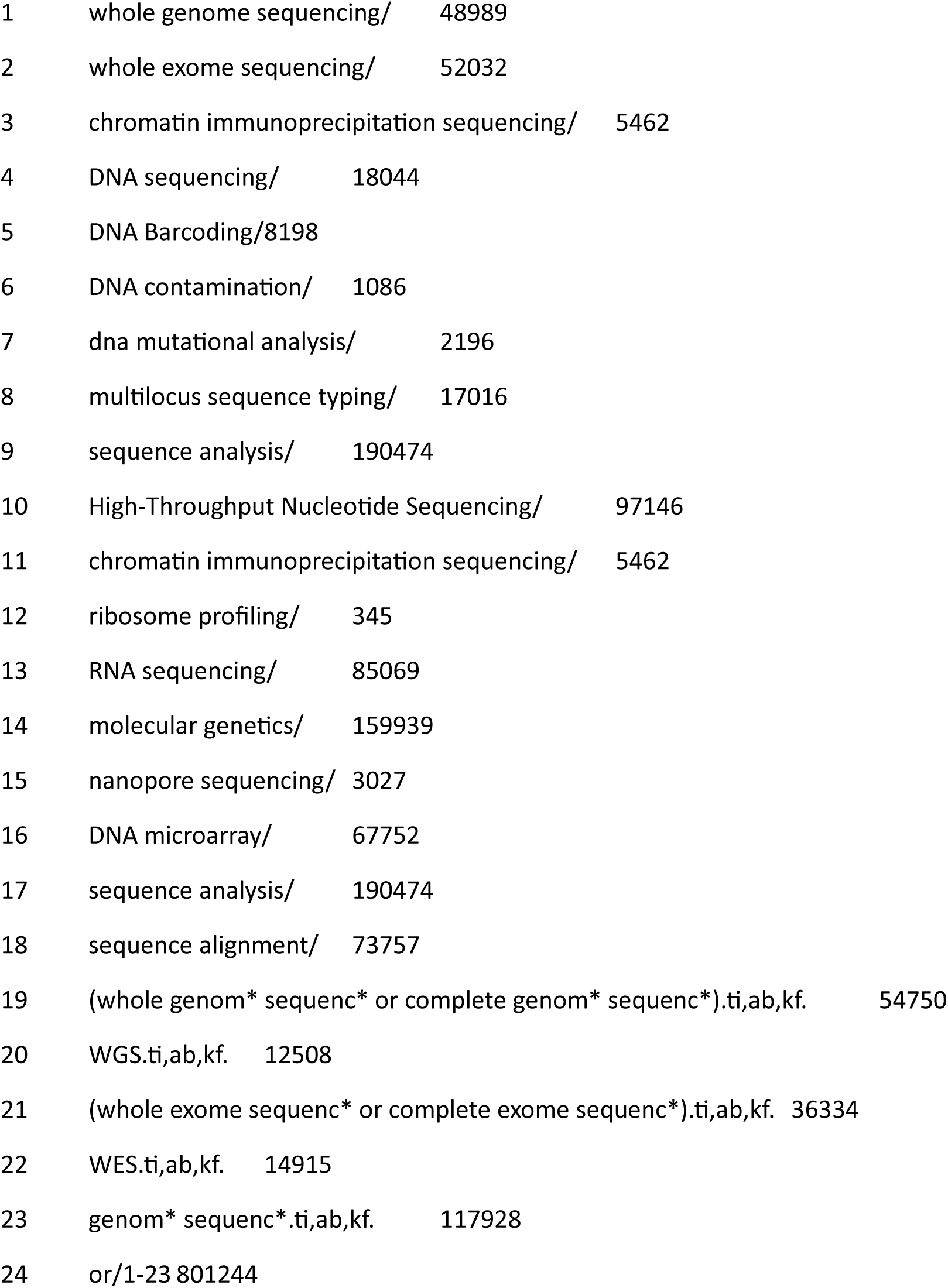
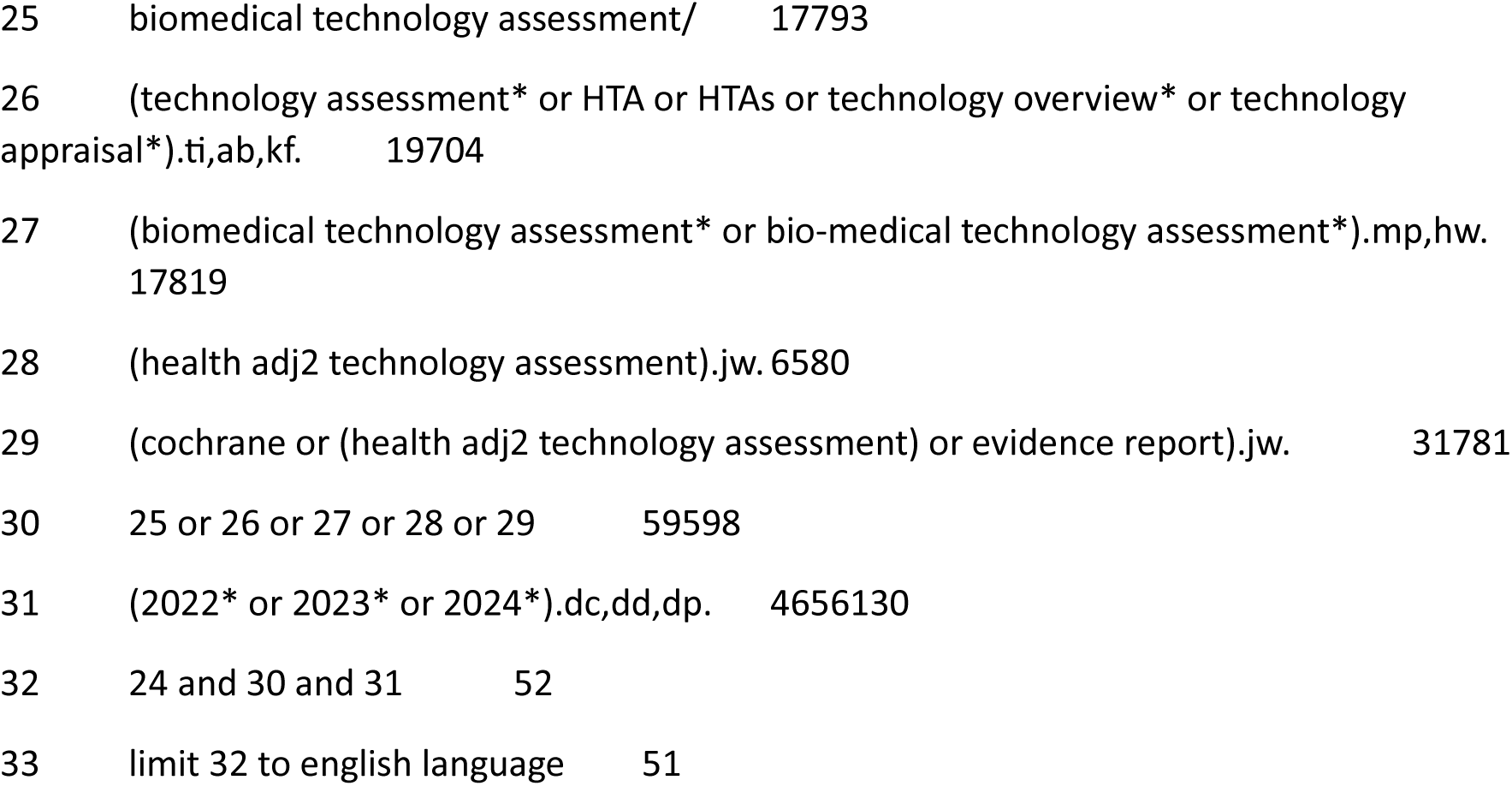

###### International HTA Database - INAHTA

(“whole genom* sequenc*” or “complete genom* sequenc*” OR WGS OR WES OR “whole exome sequenc*” or “complete exome sequenc*” or “genom* sequenc*”) 9 results – limited to 2022-2024 0 results

###### Google

“Health technology assessment” ”whole genome sequencing” 2 results

### Appendix 3. Quality appraisal

*Quality appraisal for the review of five conditions, including details of tailoring of Murad et al.’s (2018)*^21^ *quality appraisal tool for the assessment of case series and case reports for Q2 (studies exploring the prevalence of different genetic variants) and Q4 (studies exploring the impact of earlier versus later treatment), the ROBIS-2*^18^ *tool used to appraise one systematic review exploring the prevalence of different genetic variants in fHLH and assessment of bias and applicability of included studies according to these tools*.

#### Gene/variant frequency in patients with the condition(s) of interest

##### Subset of items from Murad et al.’s (2018) ^21^ quality appraisal tool for the assessment of case series and case reports used for Q2 (studies exploring the prevalence of different genetic variants)

**Table 15.** Tailoring of Murad’s quality appraisal tool for Q2 (studies exploring the prevalence of genetic variants in children with disease)

| Domain | Questions | Rating |
| --- | --- | --- |
| Selection | **1a. Does the patient(s) represent(s) the whole experience of the investigator (centre) or is the selection method unclear to the extent that other patients with similar presentation may not have been reported?** | yes - all subjects/consecutive subjects/random subjects over a certain time period who meet inclusion criteria are included (registry studies should include all patients in the respective country/countries, for 'yes' information on adequate completeness should be provided)<br>no - no information/unclear selection of subjects included in the study/only proportion of eligible patients had sequence data |
|  | **1a (comment). Comment if needed (e.g justification for 'No' response)** | NA |
|  | **1b. Were the criteria for selection for sequencing clear and appropriate (e.g not based on disease severity)?** | yes - all eligible patients (refer to Q1a) were sequenced or selection of eligible patients who were sequenced was random / not selective<br>no - not all eligible patients were sequenced and patients who were sequenced were selected inappropriately<br>unclear - not all eligible patients were sequenced and it is not reported how patients who were sequenced were selected (appropriateness unclear) |
|  | **1b (comment). Comment if needed (e.g justification for 'No' response)** | NA |
| Ascertainment | **2a. Were cases (i.e. presence of clinical features and /or biochemical features of the relevant condition) adequately ascertained?** | yes - clinical or biochemical diagnosis of condition ascertained from clinical records/ clinical assessment<br>no - self reporting/parent reporting<br>unclear - not reported |
|  | **2b. Were cases (i.e. presence of clinical features and /or biochemical features of the relevant condition) adequately defined?** | yes - unambiguous label of condition / group of conditions with clear clinical and / or biochemical definitions<br>no - some ambiguity in label of condition / group of conditions and /or no clear clinical and / or biochemical definitions (Ines to think about whether inconsistency in using terms in fHLH, inability to clearly categorise conditions based on symptoms/biochemical test is actually a risk of bias or just reporting) |
|  | **3a. Was the presence of variants (genetic make-up) adequately ascertained?** | yes - genetic make-up ascertained from clinical records (retrospective) or genetic testing (prospective)<br>no - outcomes are self reported/parent reported<br>unclear - not reported |
| **Reporting** | **4. Is the case(s) described with sufficient details to allow other investigators to replicate the research or to allow practitioners make inferences related to their own practice?** | yes - definition of disease (clinical and/or biochemical) / eligibility criteria / extent of sequencing (genes/variants/number of exons...) specified / definition of genetic disease (annotation / calling or prioritisation rules) (all present) |
| **Applicability** | **5. Is the reported genetic spectrum based only on sequencing techniques or were additional tests needed to fully identify variants (i.e are the results applicable to the the review question (WGS))?** | yes - genetic make-up reported only based on sequencing based techniques<br>no - sequencing was followed by additional methods to identify for instance deletions / insertions<br>unclear - type of genetic test not report (e.g. registry studies) or multiple types of genetic test were used and unclear which variants were identified by sequencing |

#### Quality appraisal tables

The majority of studies were appraised using the tailored Murad et al. (2018) tool, one (^54^) was a systematic review and was assessed with the ROBIS-2 tool^193^ and is presented below in a separate table.

**Table 16.**
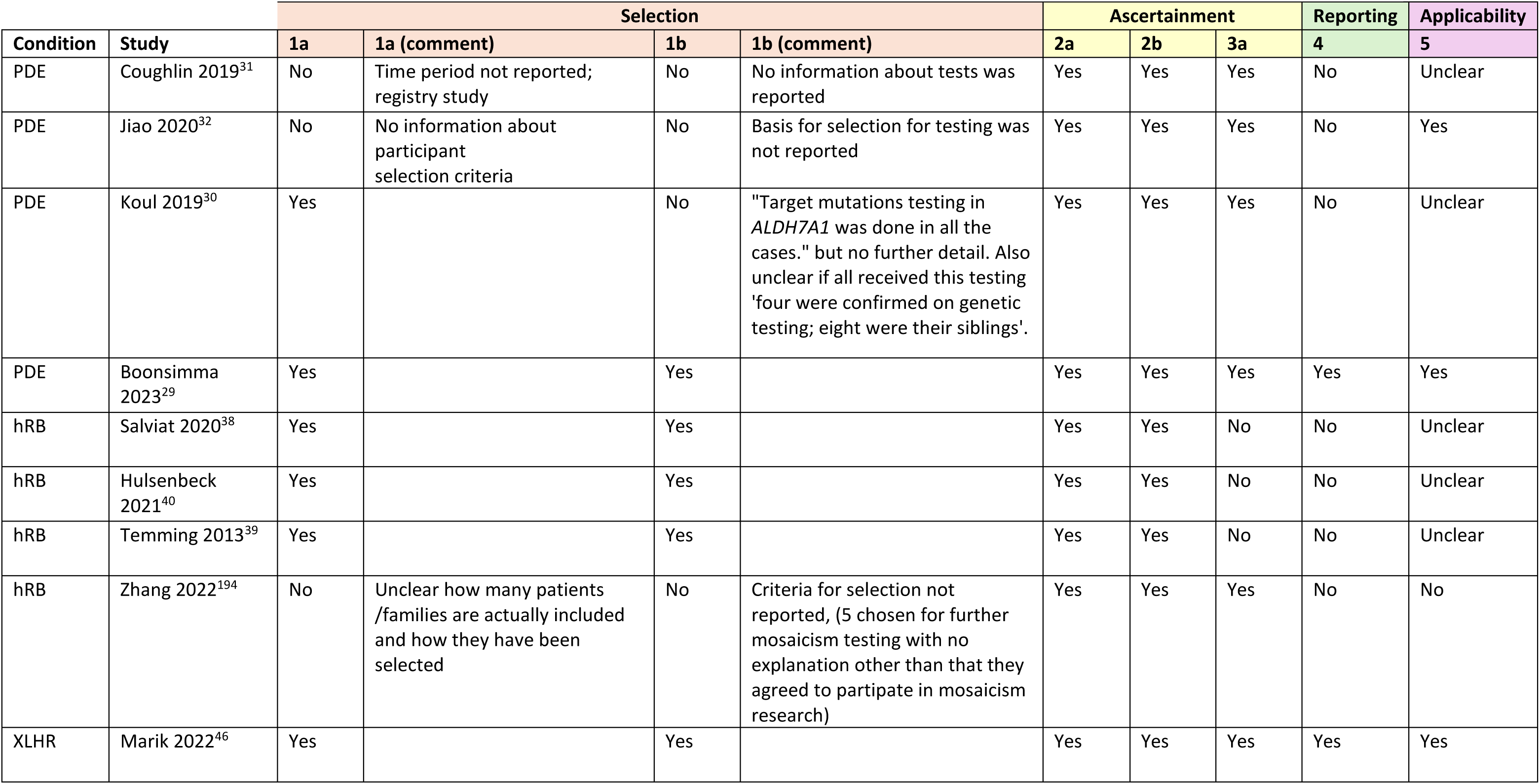
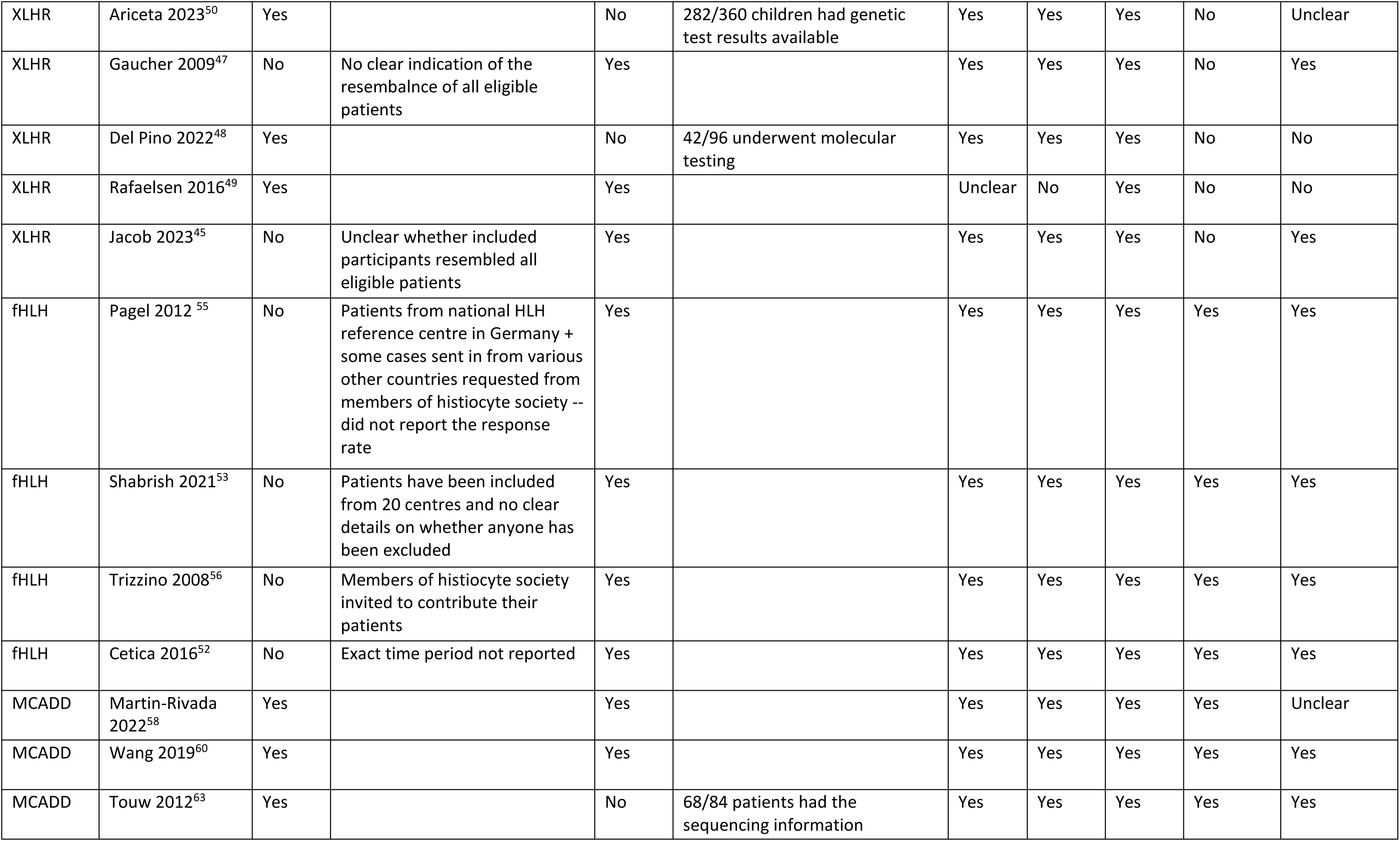
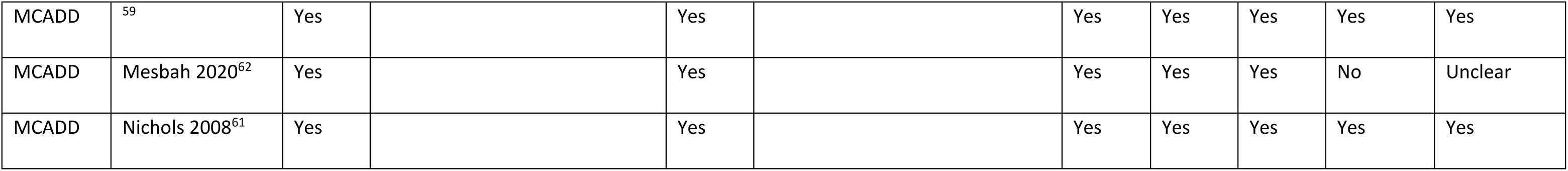
Quality appraisal of studies exploring the gene/variant frequency in patients with the condition(s) of interest using the tailored Murad’s tool.

| Condition | Study | Selection |  |  |  | Ascertainment |  |  | Reporting | Applicability |
| --- | --- | --- | --- | --- | --- | --- | --- | --- | --- | --- |
|  |  | 1a | 1a (comment) | 1b | 1b (comment) | 2a | 2b | 3a | 4 | 5 |
| PDE | Coughlin 2019<sup>31</sup> | No | Time period not reported; registry study | No | No information about tests was reported | Yes | Yes | Yes | No | Unclear |
| PDE | Jiao 2020<sup>32</sup> | No | No information about participant selection criteria | No | Basis for selection for testing was not reported | Yes | Yes | Yes | No | Yes |
| PDE | Koul 2019<sup>30</sup> | Yes |  | No | "Target mutations testing in *ALDH7A1* was done in all the cases." but no further detail. Also unclear if all received this testing 'four were confirmed on genetic testing; eight were their siblings'. | Yes | Yes | Yes | No | Unclear |
| PDE | Boonsimma 2023<sup>29</sup> | Yes |  | Yes |  | Yes | Yes | Yes | Yes | Yes |
| hRB | Salviat 2020<sup>38</sup> | Yes |  | Yes |  | Yes | Yes | No | No | Unclear |
| hRB | Hulsenbeck 2021<sup>40</sup> | Yes |  | Yes |  | Yes | Yes | No | No | Unclear |
| hRB | Temming 2013<sup>39</sup> | Yes |  | Yes |  | Yes | Yes | No | No | Unclear |
| hRB | Zhang 2022<sup>194</sup> | No | Unclear how many patients /families are actually included and how they have been selected | No | Criteria for selection not reported, (5 chosen for further mosaicism testing with no explanation other than that they agreed to participate in mosaicism research) | Yes | Yes | Yes | No | No |
| XLHR | Marik 2022<sup>46</sup> | Yes |  | Yes |  | Yes | Yes | Yes | Yes | Yes |
| XLHR | Ariceta 2023<sup>50</sup> | Yes |  | No | 282/360 children had genetic test results available | Yes | Yes | Yes | No | Unclear |
| XLHR | Gaucher 2009<sup>47</sup> | No | No clear indication of the resemblance of all eligible patients | Yes |  | Yes | Yes | Yes | No | Yes |
| XLHR | Del Pino 2022<sup>48</sup> | Yes |  | No | 42/96 underwent molecular testing | Yes | Yes | Yes | No | No |
| XLHR | Rafaelsen 2016<sup>49</sup> | Yes |  | Yes |  | Unclear | No | Yes | No | No |
| XLHR | Jacob 2023<sup>45</sup> | No | Unclear whether included participants resembled all eligible patients | Yes |  | Yes | Yes | Yes | No | Yes |
| fHLH | Pagel 2012<sup>55</sup> | No | Patients from national HLH reference centre in Germany + some cases sent in from various other countries requested from members of histiocyte society -- did not report the response rate | Yes |  | Yes | Yes | Yes | Yes | Yes |
| fHLH | Shabrish 2021<sup>53</sup> | No | Patients have been included from 20 centres and no clear details on whether anyone has been excluded | Yes |  | Yes | Yes | Yes | Yes | Yes |
| fHLH | Trizzino 2008<sup>56</sup> | No | Members of histiocyte society invited to contribute their patients | Yes |  | Yes | Yes | Yes | Yes | Yes |
| fHLH | Cetica 2016<sup>52</sup> | No | Exact time period not reported | Yes |  | Yes | Yes | Yes | Yes | Yes |
| MCADD | Martin-Rivada 2022<sup>58</sup> | Yes |  | Yes |  | Yes | Yes | Yes | Yes | Unclear |
| MCADD | Wang 2019<sup>60</sup> | Yes |  | Yes |  | Yes | Yes | Yes | Yes | Yes |
| MCADD | Touw 2012<sup>63</sup> | Yes |  | No | 68/84 patients had the sequencing information | Yes | Yes | Yes | Yes | Yes |
| MCADD | <sup>59</sup> | Yes |  | Yes |  | Yes | Yes | Yes | Yes | Yes |
| MCADD | Mesbah 2020<sup>62</sup> | Yes |  | Yes |  | Yes | Yes | Yes | No | Unclear |
| MCADD | Nichols 2008<sup>61</sup> | Yes |  | Yes |  | Yes | Yes | Yes | Yes | Yes |

**Table 17.** Quality appraisal of one systematic review exploring the gene/variant frequency in patients with the condition(s) of interest using the ROBIS-2 tool.

|  |  |  |
| --- | --- | --- |
| **Condition:** fHLH |  |  |
| **Study:** (<sup>54</sup>) |  |  |
| **PHASE 1: ASSESSING RELEVANCE (optional)**<br>Not applicable in this review |  |  |
| **PHASE 2: IDENTIFYING CONCERNS WITH THE REVIEW PROCESS** |  |  |
| **DOMAIN 1: STUDY ELIGIBILITY CRITERIA** |  |  |
| 1.1 Did the review adhere to pre-defined objectives and eligibility criteria? | Probably yes | No protocol to check |
| 1.2 Were the eligibility criteria appropriate for the review question? | Yes |  |
| 1.3 Were eligibility criteria unambiguous? | Yes |  |
| 1.4 Were any restrictions in eligibility criteria based on study characteristics appropriate (e.g. date, sample size, study quality, outcomes measured)? | Yes | Selection criterion on study design is very broad so no risk of bias by excluding certain study types |
| 1.5 Were any restrictions in eligibility criteria based on sources of information appropriate (e.g. publication status or format, language, availability of data)? | Yes |  |
| **Concerns regarding specification of study eligibility criteria** | **Low** |  |
| **DOMAIN 2: IDENTIFICATION AND SELECTION OF STUDIES** |  |  |
| 2.1 Did the search include an appropriate range of databases/electronic sources for published and unpublished reports? | Yes |  |
| 2.2 Were methods additional to database searching used to identify relevant reports? | Yes |  |
| 2.3 Were the terms and structure of the search strategy likely to retrieve as many eligible studies as possible? | Probably yes |  |
| 2.4 Were restrictions based on date, publication format, or language appropriate? | Probably yes |  |
| 2.5 Were efforts made to minimise error in selection of studies? | No information | No details on how many people sifted and how discrepancies were discussed |
| **Concerns regarding methods used to identify and/or select studies** | **Low** |  |
| **DOMAIN 3: DATA COLLECTION AND STUDY APPRAISAL** |  |  |
| 3.1 Were efforts made to minimise error in data collection? | Probably yes | No information on how data extraction form was developed and whether it was piloted |
| 3.2 Were sufficient study characteristics available for both review authors and readers to be able to interpret the results? | Probably yes |  |
| 3.3 Were all relevant study results collected for use in the synthesis? | Yes |  |
| 3.4 Was risk of bias (or methodological quality) formally assessed using appropriate criteria? | No information |  |
| 3.5 Were efforts made to minimise error in risk of bias assessment? | No information |  |
| **Concerns regarding methods used to collect data and appraise studies** | **Unclear** | **Rationale: No information on the risk of bias assessment** |
| **DOMAIN 4: SYNTHESIS AND FINDINGS** |  |  |
| 4.1 Did the synthesis include all studies that it should? | Probably yes |  |
| 4.2 Were all pre-defined analyses reported or departures explained? | No information | No protocol to check |
| 4.3 Was the synthesis appropriate given the nature and similarity in the research questions, study designs and outcomes across included studies? | No information | No information on individual studies and no justification on methods of synthesis provided: "Data were combined without being weighted, and the analysis was done as if the data were derived from a single sample" but probably ok for the simple analysis of frequencies of types of variants |
| 4.4 Was between-study variation (heterogeneity) minimal or addressed in the synthesis? | No information | No information on individual included studies so impossible to judge the level of heterogeneity and the type of adjustment that would have been appropriate. |
| 4.5 Were the findings robust, e.g. as demonstrated through funnel plot or sensitivity analyses? | No information | Probably not appropriate for this study |
| 4.6 Were biases in primary studies minimal or addressed in the synthesis? | No information |  |
| **Concerns regarding the synthesis and findings** | **Unclear** | **Rationale: No information on heterogeneity of studies and the synthesis method is not justified, no information provided on included study characteristics, list of included studies or a protocol** |
| **PHASE 3: JUDGING RISK OF BIAS** |  |  |
| 1. Concerns regarding specification of study eligibility criteria | Low | Rationale: not applicable |
| 2. Concerns regarding methods used to identify and/or select studies | Low | Rationale: not applicable |
| 3. Concerns regarding methods used to collect data and appraise studies | Unclear | Rationale: No information about the risk of bias assessment |
| 4. Concerns regarding the synthesis and findings | Unclear | Rationale: No information on heterogeneity of studies and the synthesis method is not justified, no information provided on included study characteristics, list of included studies or a protocol |

| RISK OF BIAS IN THE REVIEW |  |  |
| --- | --- | --- |
| A. Did the interpretation of findings address all of the concerns identified in Domains 1 to 4? | No |  |
| B. Was the relevance of identified studies to the review's research question appropriately considered? | Probably yes |  |
| C. Did the reviewers avoid emphasizing results on the basis of their statistical significance? | Probably no |  |
| **Risk of bias in the review** | **Unclear** | **Rationale: The study did not provide sufficient detail on the conduct of the review, the include studies and the synthesis of the studies to make an informed assessment. No protocol is appended or referenced.** |

#### Evidence on early vs late treatment

##### Subset of items from Murad et al.’s (2018)^21^ quality appraisal tool for the assessment of case series and case reports tailored for *Q4 (studies exploring the impact of earlier versus later treatment)*

**Table 18.**
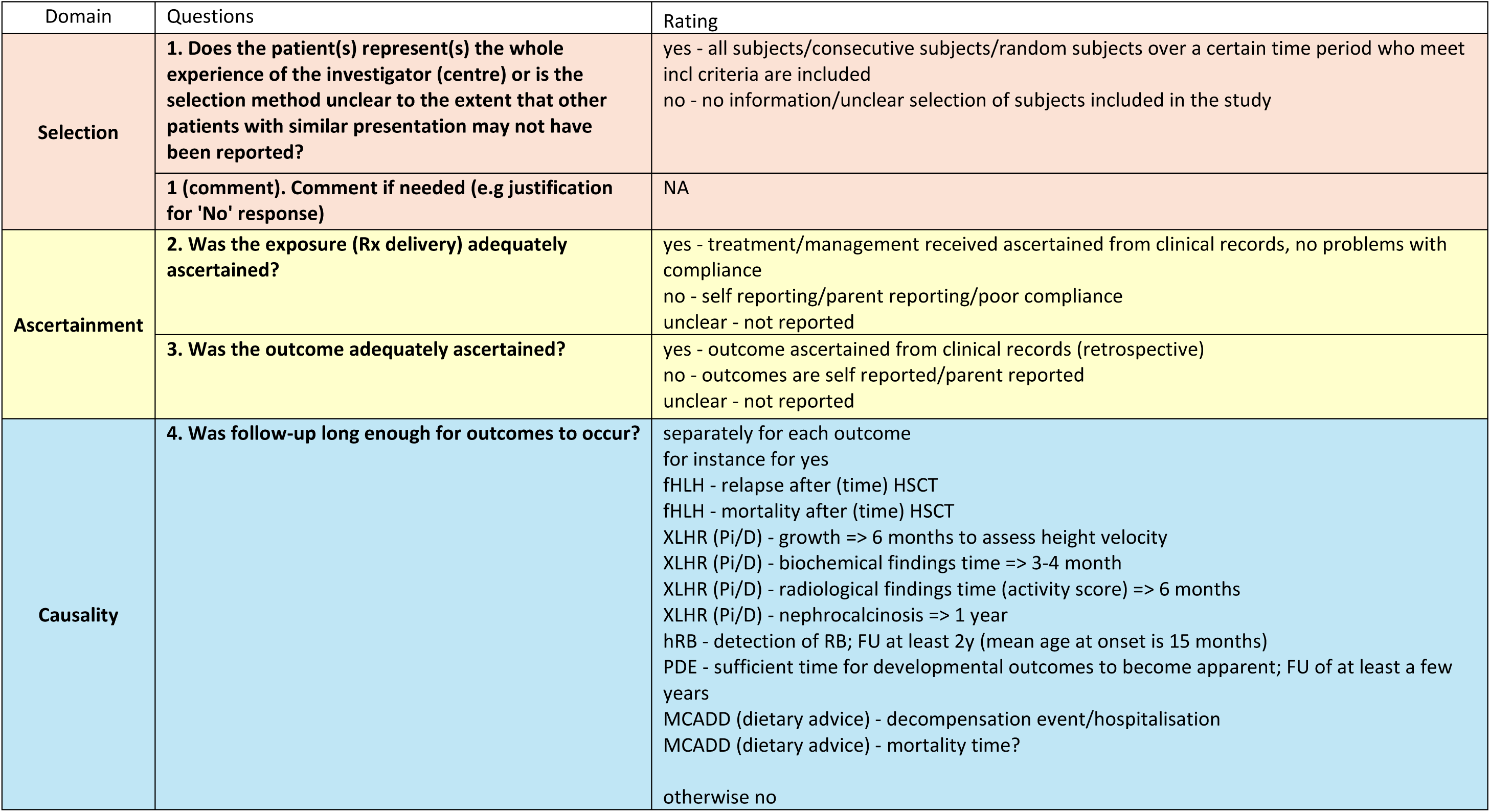
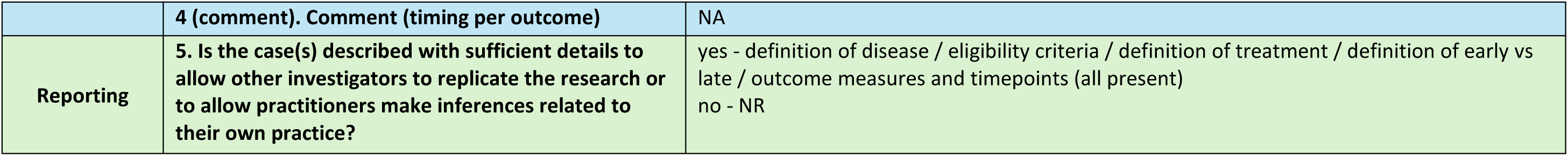
Tailoring of Murad’s quality appraisal tool for Q4 (studies exploring the benefit of earlier vs later treatment)

| Domain | Questions | Rating |
| --- | --- | --- |
| Selection | **1. Does the patient(s) represent(s) the whole experience of the investigator (centre) or is the selection method unclear to the extent that other patients with similar presentation may not have been reported?** | yes - all subjects/consecutive subjects/random subjects over a certain time period who meet incl criteria are included<br>no - no information/unclear selection of subjects included in the study |
|  | **1 (comment). Comment if needed (e.g justification for 'No' response)** | NA |
| Ascertainment | **2. Was the exposure (Rx delivery) adequately ascertained?** | yes - treatment/management received ascertained from clinical records, no problems with compliance<br>no - self reporting/parent reporting/poor compliance<br>unclear - not reported |
|  | **3. Was the outcome adequately ascertained?** | yes - outcome ascertained from clinical records (retrospective)<br>no - outcomes are self reported/parent reported<br>unclear - not reported |
| Causality | **4. Was follow-up long enough for outcomes to occur?** | separately for each outcome<br>for instance for yes<br>fHLH - relapse after (time) HSCT<br>fHLH - mortality after (time) HSCT<br>XLHR (Pi/D) - growth => 6 months to assess height velocity<br>XLHR (Pi/D) - biochemical findings time => 3-4 month<br>XLHR (Pi/D) - radiological findings time (activity score) => 6 months<br>XLHR (Pi/D) - nephrocalcinosis => 1 year<br>hRB - detection of RB; FU at least 2y (mean age at onset is 15 months)<br>PDE - sufficient time for developmental outcomes to become apparent; FU of at least a few years<br>MCADD (dietary advice) - decompensation event/hospitalisation<br>MCADD (dietary advice) - mortality time?<br>otherwise no |
|  | **4 (comment). Comment (timing per outcome)** | NA |
| **Reporting** | **5. Is the case(s) described with sufficient details to allow other investigators to replicate the research or to allow practitioners make inferences related to their own practice?** | yes - definition of disease / eligibility criteria / definition of treatment / definition of early vs late / outcome measures and timepoints (all present)<br>no - NR |

##### Quality appraisal table

**Table 19.**
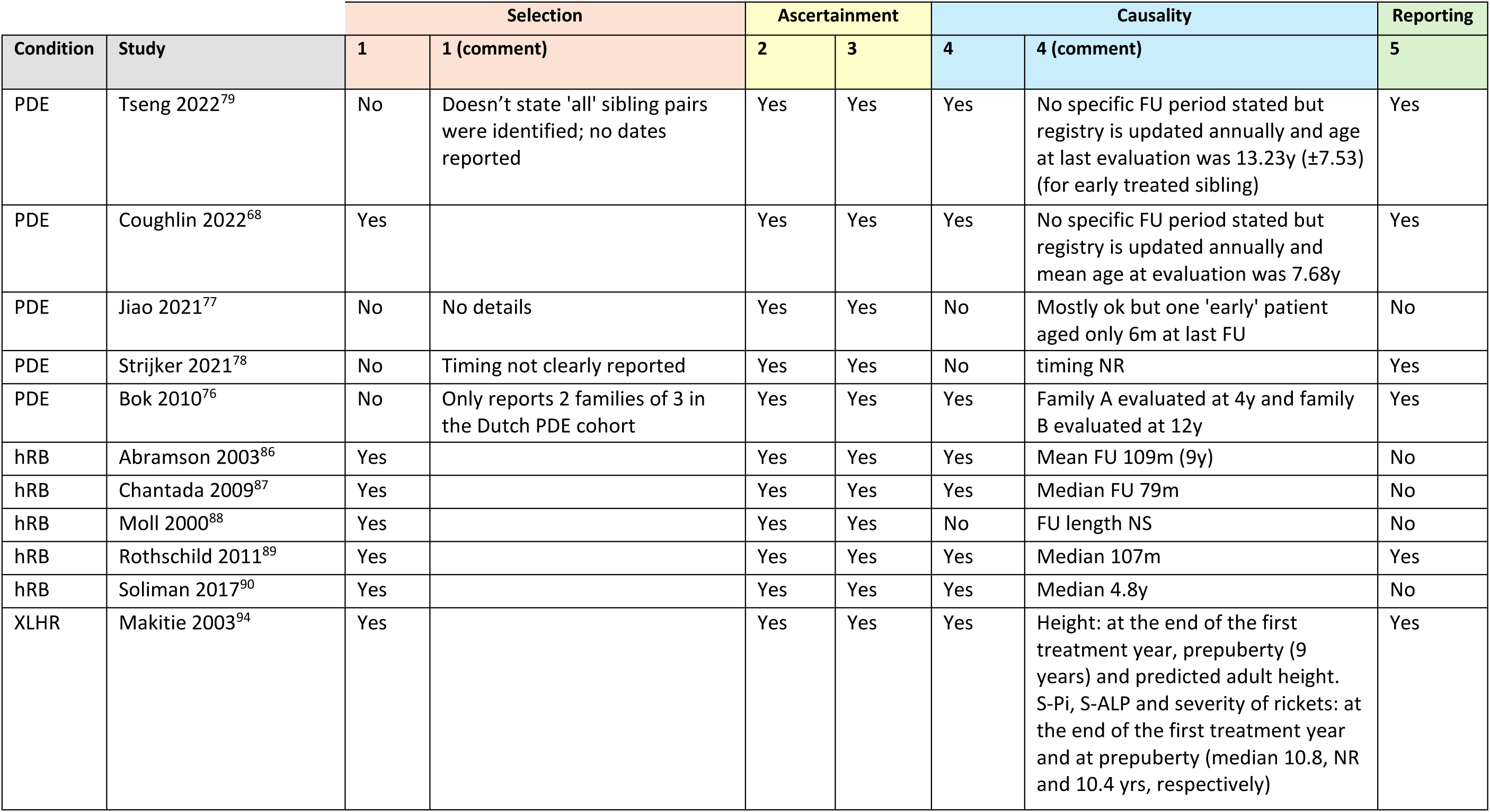
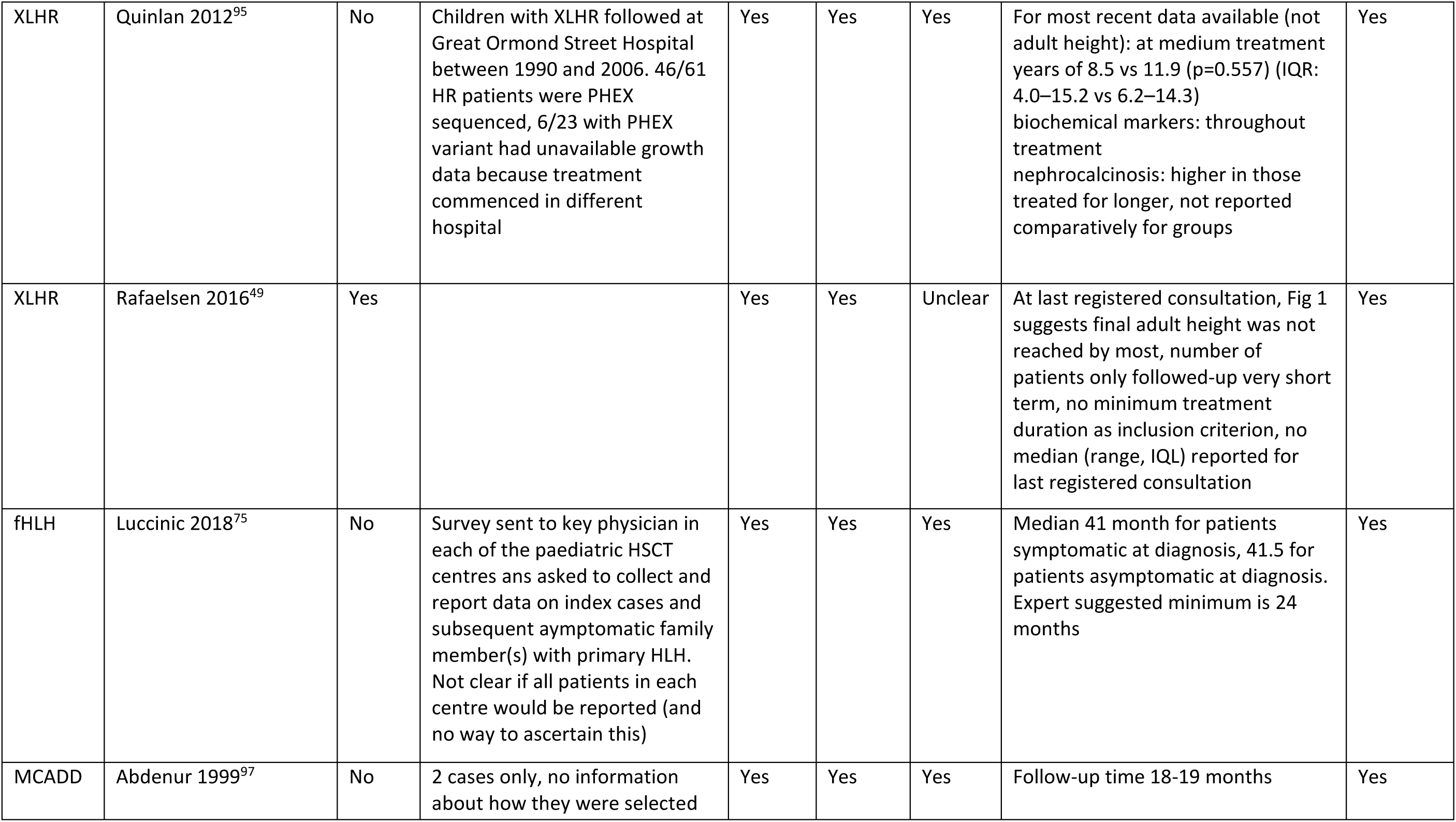
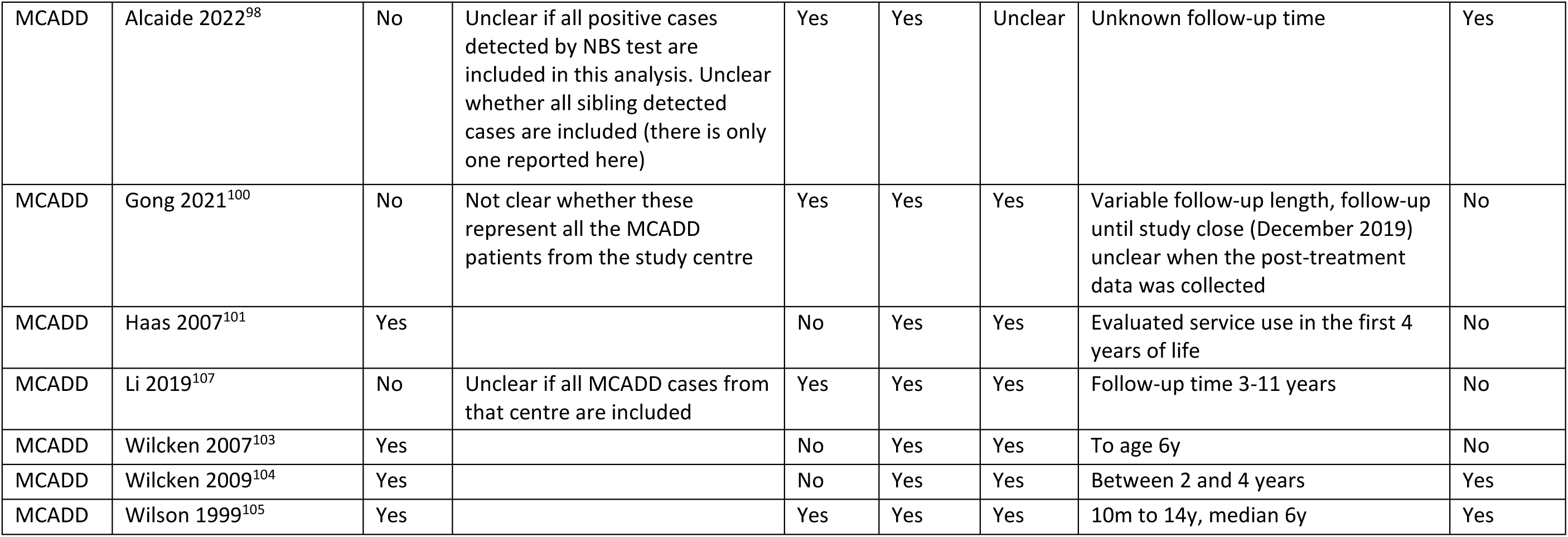
Quality appraisal of studies exploring early vs late treatment in patients with the condition(s) of interest using the tailored Murad et al. tool.

### Appendix 4. PRISMA flowcharts

*PRISMA flow charts illustrating the Selection process of studies for the review of five conditions, the review of genomic studies of paediatric cohorts reporting penetrance for pathogenic variants and the review of cost-effectiveness evaluations of WGS and WES*.

#### Review of five conditions

### Appendix 5. Summary tables for the review of the five conditions

*Data extraction tables for the traditional review including studies exploring gene/variant frequency in patients with the condition(s) of interest (including study design and recruitment dates, number and definition of cases, test description, genes/variants considered, gene frequency in cases and number of negative tests, variant frequency and expressivity), and studies presenting evidence on early vs late treatment including study design, number and definition of cases, definitions of early and late treatment, outcome measure and timepoint, and results)*.

#### Gene/variant frequency in patients with condition(s) of interest

##### PDE

**Table 20.**
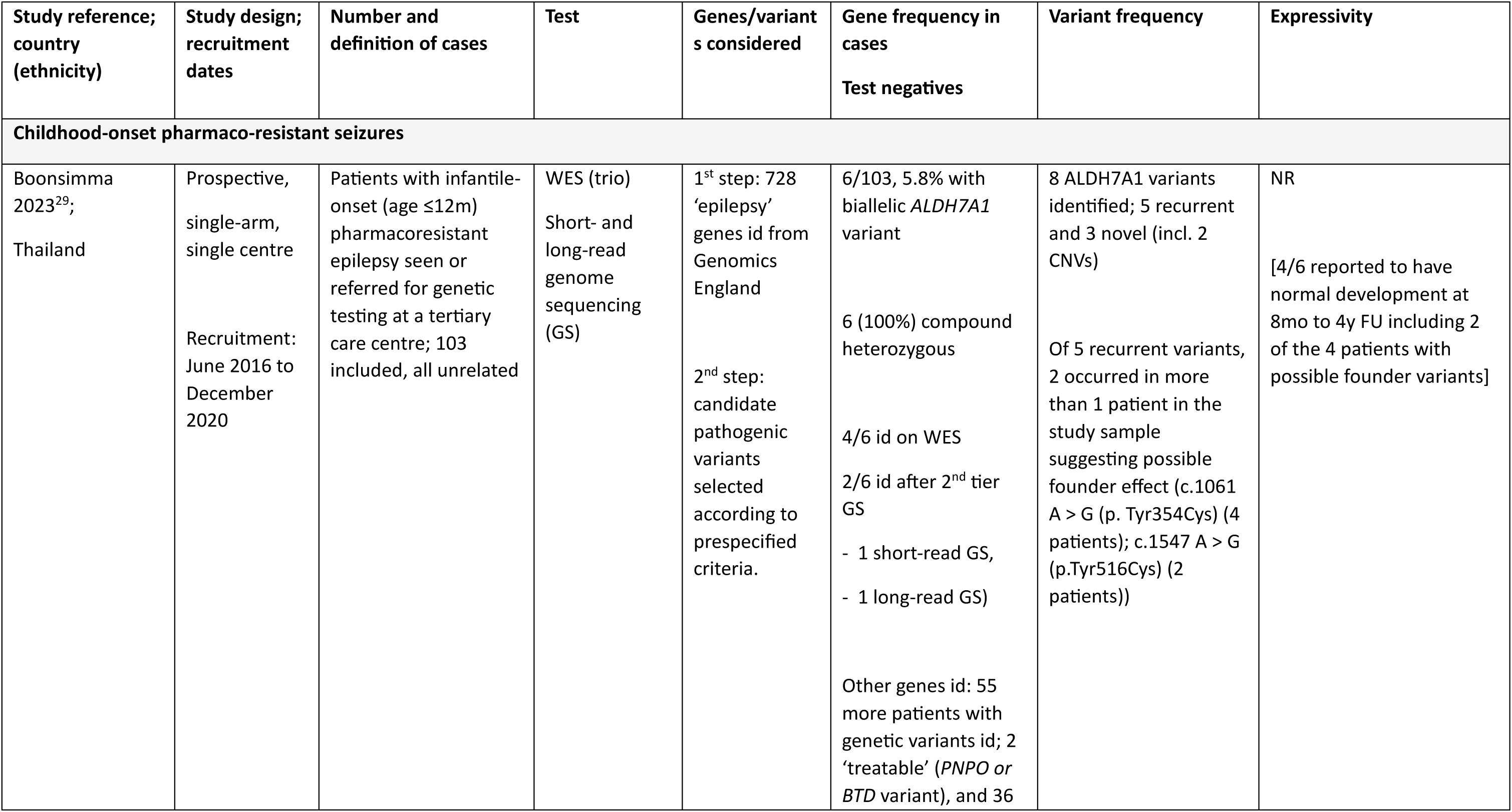
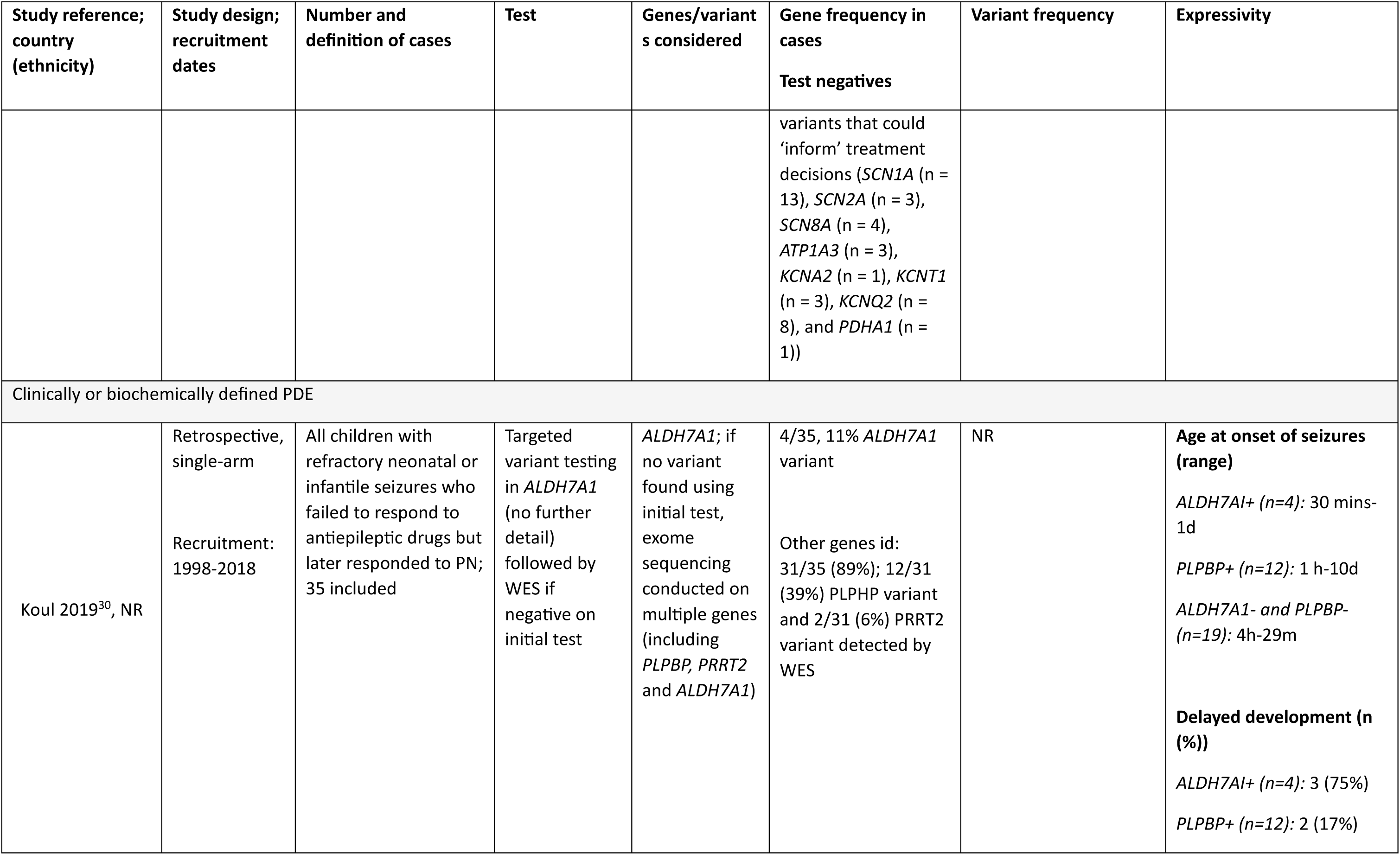
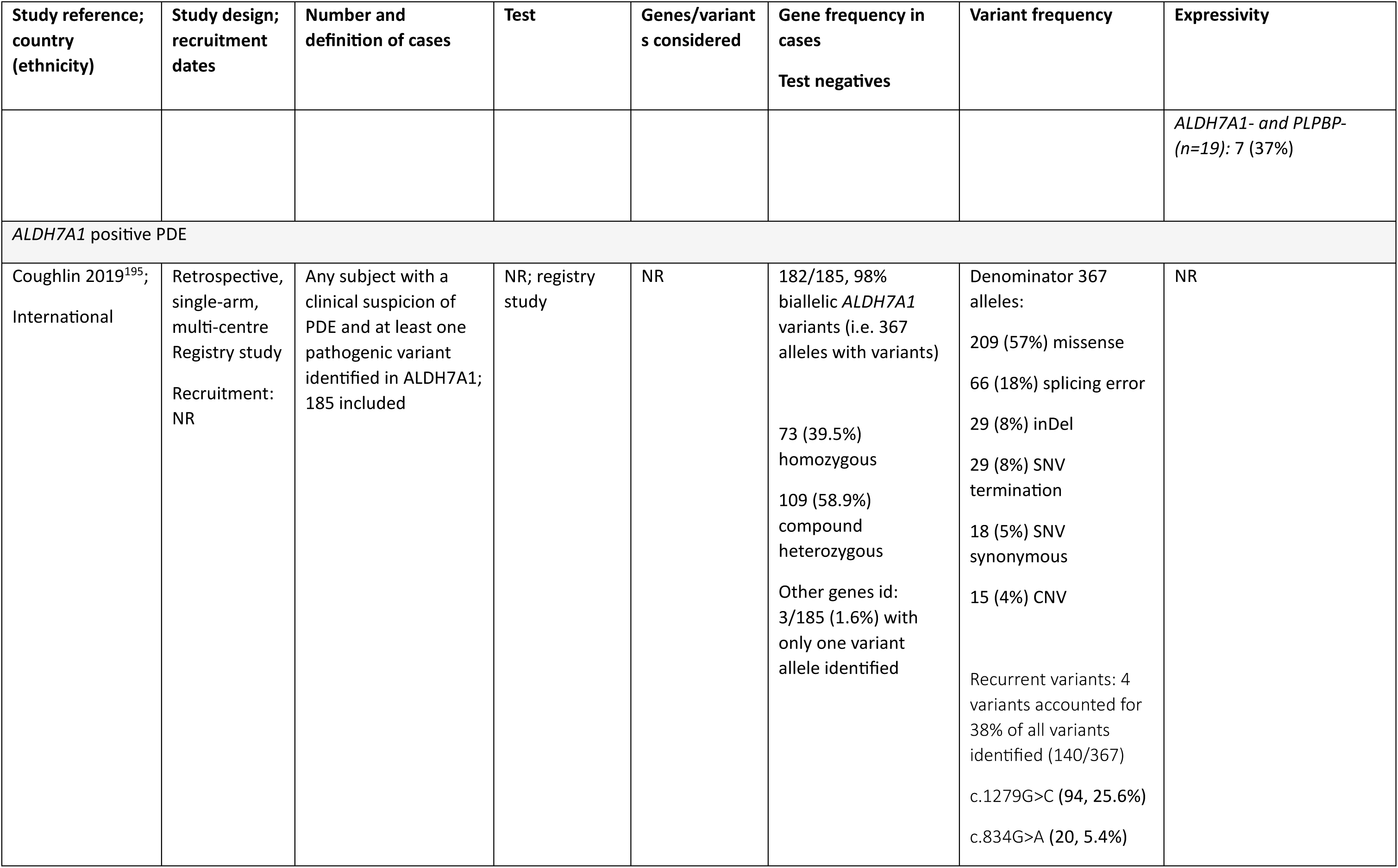
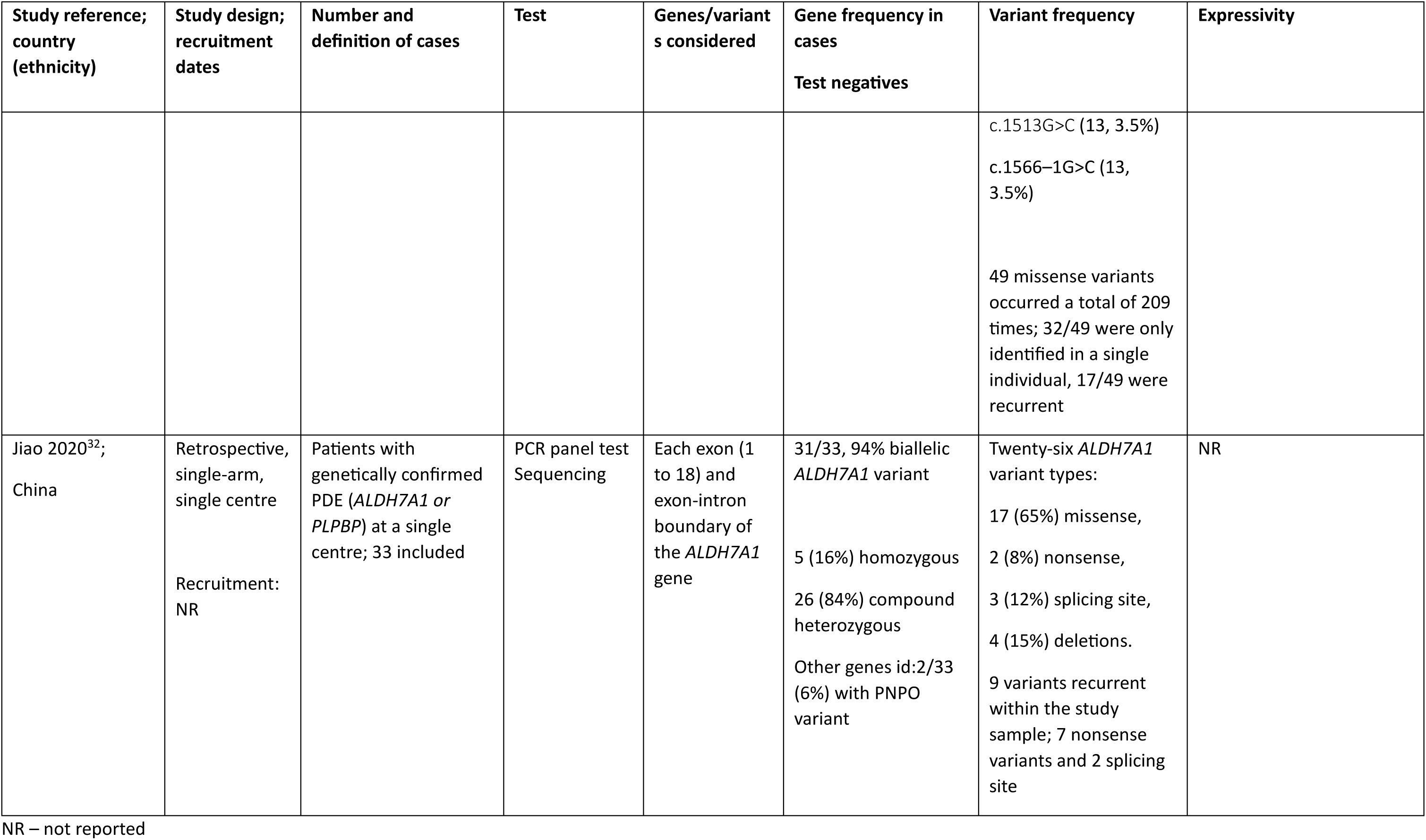
Summary tables for the studies exploring gene/variant frequency in patients with PDE.

##### hRB

**Table 21.** Summary tables for the studies exploring gene/variant frequency in patients with hRB.

| Study reference;<br>country | Study design;<br>recruitment | Number and<br>definition of cases | Test | Genes/variants<br>considered | Gene frequency in<br>cases<br>Test negatives | Variant frequency | Expressivity |
| --- | --- | --- | --- | --- | --- | --- | --- |
| *Any RB* |  |  |  |  |  |  |  |
| Salviat 2020<sup>38</sup>,<br>France | Retrospective,<br>single-arm,<br>single-centre;<br>January 1<br>2000-September 30<br>2017 | Unrelated RB index<br>cases consecutively<br>referred for genetic<br>consultation; 1371<br>patients out of 1404<br>completed genetic<br>counselling | Multiple<br>methods<br>dependent on<br>year of testing<br>(including<br>sequencing<br>and<br>sequential<br>testing); first-line screening<br>done on<br>tumour when<br>available,<br>followed by<br>germline<br>screening | *RB1*; whole<br>spectrum of<br>pathogenic<br>variants | 606/1371 (44.2%)<br>with germline *RB1*<br>variant (including<br>28/606 (4.6%) with<br>germline<br>mosaicism); 45/606<br>(7.4%) identified<br>through first-line<br>tumour screening;<br>561/606 (92.6%)<br>identified through<br>germline screening<br>765/1371 (54.5%)<br>with no germline<br>*RB1* variant;<br>517/765 (67.6%)<br>without tumour<br>screening; 248/765<br>(32.4%) with<br>somatic pathogenic<br>variant | Nonsense*: 222 (36.6%)<br>Frameshift*: 140<br>(23.0%)<br>Out-of-frame splice<br>variants*: 110 (18.2%)<br>Large rearrangements*:<br>65 (10.8%)<br>In-frame splice variants:<br>35 (5.8%)<br>Missense: 25 (4.1%)<br>Promoter sequence<br>pathogenic variants: 5<br>(0.8%)<br>In-frame deletion: 4<br>(0.7%)<br>*Associated with<br>absence of *RB* protein | **Percentage of bilateral RB**<br>*Variants with loss of RB*<br>*protein (n=537): 84.2%*<br>*Variants with no loss of RB*<br>*protein (n=69):*<br>65.2%<br>*Nonsense variants*<br>*(n=222): 89.2%*<br>**Mean (SD) age at**<br>**diagnosis**<br>*Variants with loss of RB*<br>*protein (n=537): 12.3m*<br>*(11.3)*<br>*Variants with no loss of RB*<br>*protein (n=69):*<br>16.3m (13.2) |
|  |  |  |  |  |  | Novel low-penetrance<br>region identified in exon<br>24 | **Percentage of stage D/E**<br>**RB**<br>*Variants with loss of RB*<br>*protein (n=537): 63.5%*<br>*Variants with no loss of RB*<br>*protein (n=69): 49.3%* |
| *Sporadic RB* |  |  |  |  |  |  |  |
| Temming 2013<sup>39</sup>;<br>Germany | Retrospective,<br>single-arm,<br>single-centre;<br>1961-2006 | Patients treated and<br>undergoing<br>ophthalmological<br>follow-up (to age 5y)<br>for unilateral RB with<br>genetic data available<br>and no family history<br>of RB; 195 patients<br>selected out of 868<br>diagnosed at the<br>centre | Multiple<br>methods<br>dependent on<br>year of<br>testing;<br>including<br>sequencing<br>and<br>sequential<br>testing | *RB1*; no further<br>detail | 40/195 (20.5%) with<br>germline *RB1*<br>variant (including<br>11/40 (27.5%) with<br>germline mosaicism)<br>155/195 (79.5%)<br>with no germline<br>*RB1* variant | *For the 29 patients with*<br>*a heterozygous RB1*<br>*variant*<br>Whole gene deletion:<br>10 (34%)<br>Mild variant: 6 (21%)<br>Premature termination:<br>13 (45%) | 9/195 (4.6%) developed<br>metachronous bilateral<br>RB; 8/9 had a<br>heterozygous variant and<br>1/9 had germline<br>mosaicism |
| *Germline/hereditary RB* |  |  |  |  |  |  |  |
| Hulsenbeck<br>2021<sup>40</sup>; Germany<br>(includes national<br>and international<br>patients) | Retrospective,<br>single-arm,<br>single-centre;<br>1992-2011 | Patients with a<br>heterozygous<br>pathogenic<br>constitutional *RB1*<br>variant (somatic<br>mosaicism excluded)<br>with complete | Multiple<br>methods;<br>including<br>sequencing<br>and | *RB1*; no further<br>detail | 287/287 (100.0%)<br>with germline *RB1*<br>variant | Nonsense or frameshift<br>variant (REC-I): 199<br>(69.3%) | **Percentage of bilateral RB**<br>*REC-I (n=199): 93.5%* *REC-*<br>*II (n=39): 76.9%*<br>*REC-III (n=45): 80.0%* |
|  |  | phenotype data who did not undergo screening for RB; 287 patients selected out of 821 diagnosed at the centre | sequential testing |  | None with no germline variant<br>*3/821 without RB1 variants characterized by high MYCN amplification* | Whole RB1 gene deletions (REC-II): 39 (13.6%)<br>Missense or in-frame SNV (REC-III): 45 (15.7%)<br>SNV or indels 3' end (REC-IV): 1 (0.3%)<br>SNV in promotor region (REC-V): 3 (1.0%) | **Median (range) age at diagnosis**<br>*REC-I (n=199): 7.3m (0.2-48.0)*<br>*REC-II (n=39): 10.3m (0.4-40.9)*<br>*REC-III (n=45): 11.6m (0.9-45.1)* |

RB – retinoblastoma; m – months

##### XLHR

**Table 22.** Summary tables for the studies exploring gene/variant frequency in patients with XLHR.

| Study reference; country (ethnicity) | Study design; recruitment | Number and definition of cases | Test | Genes/variants considered | Gene frequency in cases<br>Test negatives | Variant frequency | Expressivity |
| --- | --- | --- | --- | --- | --- | --- | --- |
| Hereditary rickets |  |  |  |  |  |  |  |
| Jacob 2023;<sup>45</sup><br>India | Prospective,<br>single-arm<br>single centre<br>Recruitment:<br>2018 | Unrelated individuals with suspected hereditary rickets<br>10 probands with childhood onset disease, 2/10 with diagnosis in early adulthood | Exome sequencing | Not specified | 3/10 (33%) PHEX<br>Test 'negatives':<br>CYP27B1 (n=3)<br>CYP2R1 (n=1)<br>VDR (n=1)<br>SLC34A3 (n=1)<br>SLC2A2 (n=1) | 3 recurrent (previously reported) truncating PHEX variants identified<br>c1482+5G>C, c1586_1586+1del and c.58C>T. The c1586_1586+1del | Phenotype of c1586_1586+1del comparatively more severe than c1482+5G>C and c.58C>T. |
| Hypophosphatemic rickets |  |  |  |  |  |  |  |
| Gaucher 2009;<sup>47</sup><br>France | Prospective,<br>single-arm,<br>single centre | Hypophosphatemic rickets associated with tubular phosphate wasting (low serum phosphate and TmP/GFR), along with | Sequencing | PHEX<br>22 exons plus adjacent intronic | 93/118 (78%) PHEX (44 sporadic and 49 familial cases) | 78 different variants identified; 60/78 (77%) were novel | Some uncertainty about pathogenicity of novel variants, e.g. |
| (European, North African, Caribbean, Asian) | Recruitment: NR | bone deformities and radiological evidence of rickets<br>118 probands (62 sporadic; 56 familial) /209 patients of 118 families with hypophosphatemic rickets 'pre-screened' to exclude patients with tubulopathy, hypercalciuria, or hyperparathyroidism, all indicative of other HR types |  | sequences and the untranslated region at the 3-prime end | PHEX negative patients:<br>25/118<br>Further analysis revealed:<br>1 case large deletion,<br>1 case different variant (FGF23),<br>1 case tumour-induced osteomalacia,<br>Rest remains unexplained (possible reasons: large deletions, mosaicism, other causal genes) | 22/78 (28%) nonsense,<br>23/78 (30%) frameshift,<br>18/78 (23%) splice sites,<br>15/78 (19%) missense | c.1206A>G (single nucleotide replacement): unclear relationship between variant and disease (patient also displays a missense variant)<br>2x c.505G>A variant (single nucleotide replacement): both patients carry additional variants (1x insertion leading to frameshift and 1x deletion leading to a frameshift)<br>c.849 + 6insT maybe responsible for late onset disease (10 to 12 years) |
| Marik 2022;<sup>46</sup> India | Prospective, single-arm | Refractory **hypophosphatemic** rickets (lack of radiological healing despite treatment); | WES | **PHEX**, FGF23, DMP1, ENPP1, CLCN5, CTNS, SLC2A2, | 24/66 (36.4%) PHEX | 24 different PHEX variants,<br>13/24 novel variants, | One patient was carrying two PHEX variations c.2048T > A, c.2071-1G > C while her mother who was clinically mildly affected had only one |
| | single centre (consecutive patients)<br>Recruitment: May 2015 - July 2019 | blood levels of phosphate below normal values for age<br>66 unrelated patients (42 familial cases); mean age at onset of symptoms $22.5 \pm 14.3$ months. | | GATM, SLC34A1, EHHADH, SLC4A1, ATP6V1B1 | 40/66 (60.6%) other affected genes: ENPP1 (n = 2), CLCN5 (n=3), CTNS (n = 3), SLC2A2 (n = 6), SLC4A1 (n = 7), ATP6V1B1 (n = 4), CYP27B1 (n = 5), VDR (n = 4); n=1 each for FGF23, DMP1, SLC34A1, EHHADH, GATM, ATP6V0A4, FGFR1<br>2/66 (3.0%) VUS; 1 PHEX and 1 HRAS variant considered negative | 11/24 known variants<br>19 pathogenic, 4 likely pathogenic and 1 VUS | PHEX variation c.2048T > A; p.(Leu683His) which could be the reason behind the variability in disease severity<br>Patients carrying PHEX deletions/insertions/nonsense for a premature stop codon and truncated PHEX protein showed a severe phenotype.<br>Patients with: PHEX insertion c.985dup showed an additional feature of dolicocephaly, PHEX deletion c.1202del had multiple pseudofractures, PHEX deletion c.1965del showed loss of permanent teeth, sensorineural hearing loss in left ear and severe enthesopathy |
| Hereditary hypophosphatemic rickets |  |  |  |  |  |  |  |
| del Pino 2022;<sup>48</sup> Argentina | Retrospective, single-arm<br>single centre<br>Recruitment: 1992 - 2019 | Patients diagnosed with hereditary hypophosphatemic rickets (following clinical and biochemical selection of patients, diagnosis was based on clinical examination, laboratory tests and X-ray)<br>42/96 included children underwent molecular testing | Sequencing + MLPA (to detect gene deletions and duplications) | *PHEX*<br>Entire coding region (exons 1–22) and splice sites in flanking intronic regions | 36/42 (85.7%) *PHEX*<br>No confirmed variant<br>6/42 (14.3%) | 36/36 deleterious alterations or large deletions | NR |
| Rafaelsen 2016;<sup>49</sup> Norway | Retrospective, single-arm<br>multi centre<br>Recruitment: 2009 – 2014 | Patients with hereditary hypophosphatemia; defined by serum phosphate below age-dependent reference range in repeated samples combined with TmP/GFR not due to primary or secondary hyperparathyroidism, | Sanger sequencing + MLPA (if a disease-causing variant was not found, the inheritance pattern suggested a sporadic case or X-linked | *PHEX* and *FGF23*, *DMP1*, *ENPP1*, *KL*, *FAM20C* in successive order if *PHEX* negative<br>All exons and intron–exon | 13/19 (68.4%) *PHEX*<br>Other genes:<br>1/19 (5.3%) *FAM20C*<br>1/19 (5.3%) *SLC34A3*<br>4/19 (21.1%) no confirmed variant | 13 different *PHEX* variants,<br>9/13 novel *PHEX* variants<br>9 novel variants:<br>1 large duplication,<br>2 frameshift and | There were no differences in growth, dental involvement, persistent bowing, or development of nephrocalcinosis in a comparison of non-sense *PHEX* variants with missense *PHEX* variants |
|  |  | Fanconi syndrome or other tubulopathy, vitamin D dependent rickets, vitamin D deficiency or hypophosphatemia secondary to acute metabolic derangements. +/- FH or genetic diagnosis<br>19 probands / 28 patients (22 familial and 6 sporadic cases) from 19 families<br>Median age at diagnosis was 2.1 years (range 0.1–15.5 years) | dominant disease to look for mid-size deletions and insertions) | boundaries of selected genes | (possible reason: variants in other genes) | premature stop codons,<br>2 triplet deletions,<br>2 missense,<br>1 nonsense,<br>1 splice site |  |
| x-linked hypophosphatemic rickets |  |  |  |  |  |  |  |
| Ariceta 2023;<sup>50</sup><br>Multinational | Retrospective, single-arm registry study | People of all ages diagnosed with XLHR based on clinical judgement of an XLH-treating expert | Genetic testing | NR | 253/282 (89.7%)<br>*PHEX* | NR | NR |
|  | Recruitment:<br>Aug 2017 - Nov 2020 | physician (FH, clinical, radiological and biochemical findings), or by genetic testing (positive for *PHEX* variant), small number of non-*PHEX* variants (n = 15; 2.6%)<br>282/579 children with genetic test results included |  |  | Other genes:<br>4/282 (1.4%) *FGF23*,<br>1/282 (0.4%) *SLC34A3*,<br>7/282 (2.5%) other<br>17/282(6.0%) no confirmed variant |  |  |

TmP/GFR – ratio of tubular maximum reabsorption rate of phosphate to glomerular filtration rate; WES – whole exome sequencing; MLPA – multiplex ligation-dependent probe amplification; FH – family history, NR – not reported

##### fHLH

**Table 23.** Summary tables for the studies exploring gene/variant frequency in patients with fHLH.

| Study reference; country (ethnicity) | Study design; recruitment | Number and definition of cases | Test | Genes/variants considered | Gene frequency in cases<br>Test negatives | Variant frequency | Expressivity |
| --- | --- | --- | --- | --- | --- | --- | --- |
| **HLH** |  |  |  |  |  |  |  |
| Cetica 2016;<sup>52</sup><br>Italy<br>(Southern European; Eastern European; African; Asian; Hispanic; others) | Retrospective, single-arm multi-centre<br>Recruitment: time reported to the Italian registry: 1989 - 2014 | Patients with a clinical diagnosis of HLH as defined by the diagnostic criteria recommended by the Histiocyte Society that was subsequently confirmed.<br>426 sequenced/500 (33/500 no material, 41/500 normal on immunological testing no suspicion of genetic disease, i.e. no consanguinity, familial recurrence, pigment deficiency, | NR; registry study | Testing strategy based on results from immunologic assays:<br>- *PRF1* in patients with perforin expression deficit<br>- *UNC13D* then *STX11* then *STXBP2* in patients with degranulation defect<br>- *RAB27a* then *LYST* in patients | 171/426 (40.1%) with biallelic pathogenic variants<br>69/426 (16.2%) *PRF1*<br>62/426 (14.6%) *UNC13D*<br>1/426 (0.2%) *STX11*<br>9/426 (2.1%) *STXBP2*<br>Other HLH genes:<br>*RAB27a* (n=10)<br>*LYST* (n=3)<br>XIAP (n=4)<br>SD2D1A (n=13) | *PRF1*:<br>28/69 homozygous<br>34 different variants<br>- missense (n=20)<br>- nonsense (n=6)<br>- deletions/insertions (n=8)<br>Most frequent variants: c.1122G>A (n=19 patients); c.272C>T (n=11); c.657C>A (n=10); c.695G>A (n=9)<br>*UNC13D*:<br>22/62 homozygous | 26 families with 2 siblings: 16 pairs with similar age<br>at which the disease manifested, 9 pairs with age difference of between 46 and 207 months, and 1 pair sibling 1 with disease onset at 6.7 years, and sibling 2 unaffected at the age of 25 years<br>50.8% of the 171 patients with biallelic variants died. |
|  |  | or disease reactivation)<br>For 500 patients:<br>mostly children,<br>44/500 (8.8%)<br>adults /<br>median age (range)<br>at diagnosis: 2.2y (0 - 60y) |  | with pigment deficiency<br>- *SH2D1A* in male patients with defective *SAP* expression and/or inhibitory instead of activating 2B4 receptor function<br>- *XIAP* in male patients<br>Coding exons and exon-intron boundaries were sequenced | Test negative:<br>255/426 (59.9%)<br>Monoallelic variants:<br>43/426 (10.1%) (in 1 (n=41) or 2 (n=2) HLH-related genes*)<br>25/426 *PRF1*,<br>10/426 *UNC13D*,<br>2/426 *STX11*,<br>6/426 *STXBP2*<br>Other HLH genes<br>*RAB27A* (n=2)<br>No variants found (secondary HLH):<br>197/426 (46.2%) | 37 different variants<br>- missense (n=11)<br>- nonsense (n=8)<br>- deletions/insertions (n=12)<br>- splicing (n=6)<br>Most frequent variants:<br>c.75311G>T (n=19);<br>c.2346_2349delGGAG (n=11); c.1847A>G (n=8)<br>*STX11*: 1 variant (deletion/insertion)<br>*STXBP2*: 7 different variants<br>- missense (n=4)<br>- deletions/insertions (n=3) |  |
|  |  |  |  |  | No variants found (assumed missed genetic cases):<br>15/426 (3.5%) (potentially underlying genes not yet discovered at time of testing) |  |  |
| fHLH (4 genes only) |  |  |  |  |  |  |  |
| Shabrish 2021,<sup>53</sup><br>India | Retrospective single-arm multi-centre<br>Recruitment: 2010 - 2020 | Patients fitting into HLH criteria (including perforin expression and degranulation assay on NK cells) of the Histiocyte Society referred to ICMR-National Institute of Immunohematology or collaborating FPID centres or other tertiary care centres in India | Sanger sequencing (for perforin-deficient patients)<br>NGS or WES (for patients with degranulation abnormalities) | *PRF1*,<br>*UNC13D*,<br>*STX11*,<br>*STXBP2* | 86/98 (87.8%) with biallelic disease<br>40/98 (40.8%) *PRF1*<br>30/98 (30.6%) *UNC13D*<br>7/98 (7.1%) *STX11*<br>9/98 (9.2%) *STXBP2*<br>Test negatives:<br>12/98 (12.2%) | Details of variant spectrum available for n=72/98<br>*PRF1* (n=34)<br>- 25 different variants<br>- missense (19/25)<br>- nonsense (4/25)<br>- frameshift (2/25)<br>*UNC13D* (n=23) | Across all 4 genes, patients with homozygous variants had earlier disease onset (median 10m) than patients with compound heterozygous variants (median 3y)<br>all but 2 patients (1 homozygous, 1 compound heterozygous) with *PRF1* had <10% perforin expression.<br>patients with compound heterozygous variants in |
|  |  | Diagnosis confirmed by molecular analysis<br>98/101 under 18 years age |  |  | Individuals with monoallelic disease:<br>8/98 (8.2%) *PRF1*<br>2/98 (2.0%) *UNC13D* | - 28 different variants<br>- missense (10/28)<br>- nonsense (1/28)<br>- frameshift (11/28)<br>- splice-site and intronic (6/28)<br>*STX11* (n=6)<br>- 5 different variants<br>- 2/5 novel<br>- missense (3/5)<br>- nonsense (1/5)<br>- frameshift (1/5)<br>*STXBP2* (n=9)<br>- 8 different variants<br>- 2/8 novel<br>- missense (5/8) | *UNC13D* (n=7), *STXBP2* (n=1) had a natural killer cell degranulation <10% apart from 2 (*UNC13D*)<br>3 patients with atypical presentations (2 with *PRF1* variants, 1 with *STXBP2* variant) |
|  |  |  |  |  |  | - splice-site and intronic (3/8) |  |
| Single gene/variant |  |  |  |  |  |  |  |
| Trizzino 2008;<sup>56</sup><br>Multinational<br>(USA, Italy, Germany, Japan, Sweden, Czech Republic)<br>(Caucasian, Turkish, African American, Japanese, Hispanic, Arabic, Asian, African, Moroccan) | Retrospective<br>single-arm<br>multi-centre<br>Recruitment:<br>1983 - 2006 | Patients with FHL2 with documented biallelic PRF1 variants<br>124 patients / median (range) age at presentation: 3m (15d -- 26.3y) | Direct sequencing | PRF1 (exons 2 and 3) and exon-intron boundaries | 124/124 with biallelic PRF1 variants | 63 different variants (inc. 15 novel):<br>- 11 nonsense<br>- 10 frameshift<br>- 38 missense<br>- 4 in-frame deletion<br>missense variants only (n=34)<br>1 missense + 1 disruptive (nonsense/frameshift) variant (n=28)<br>disruptive variants only (n=62)<br>Most common variants: 1122G>A in 32 patients | Younger age of onset for patients with 2 disruptive variants compared to patients with missense variants only (p<0.001)<br>Nonsense variants significantly associated with absent natural killer cell activity (p=0.008) |
|  |  |  |  |  |  | (22 homozygous, 10 compound heterozygous), 17 Turkish, 5 Italian<br>50delT variant in 21 patients (16 homozygous, 5 compound heterozygous), mainly African or African American<br>1090-91delCT variant in 7 patients (2 homozygous, 5 compound heterozygous), all Japanese) |  |
| Amirifar 2021;<sup>54</sup><br>Multinational | Systematic review | Patients with *UNC13D* variant meeting $\geq 5$ (severe) or $\leq 4$ (mild) HLH 2004 criteria (confirmed | NR | NR | 269/322 (83.5%) with biallelic disease<br>Test negatives: | 117/269 homozygous<br>152/269 compound heterozygous | 60% of severe feature and 30% of mild feature patients carried homozygous *UNC13D* variants ($p = .001$) |
|  |  | immunologic and genetic evaluation)<br>322 patients with *UNC13D* variant/<br>median age of onset 6 months |  |  | Monoallelic disease<br>50/322 (15.5%)<br>Not reported:<br>3/322 (0.9%) | For complete sample (n=322):<br>- Missense 131 (20.47%)<br>- Nonsense 69 (10.87%)<br>- Deletion 4 (0.63%)<br>- Frameshift 87 (13.59%)<br>- Splice error 224 (35.0%)<br>- Inversion 38 (5.94%)<br>- Unknown 85 (13.28%)<br>- 53.4% of variants in exons<br>- 46.6% of variants in introns | Missense variants (20.5%) were the most prevalent variation in severe feature patients<br>Splice errors (35%) are the most prevalent alteration in mild form of the disease<br>Frequency of frameshift and nonsense variants was 14% in severe feature patients and 11% in mild feature patients<br>Severe features patients had exonic variants 1.6-fold higher than intronic variants. The frequency of exonic and intronic variants was equal in mild feature patients |
| Pagel 2012;<sup>55</sup><br>Multinational (Germany, Turkey)<br>(White, Turkish, Arab, Asian) | Retrospective single-arm multi-centre<br>Recruitment: NR | Patients with confirmed biallelic *STXBP2* variants<br>37 (28 families) patients / median (range) age at diagnosis 2 months (3 days to 19 years); 1/37 (2.7%) >18 years | Direct sequencing | *STXBP2* (exons 1 to 19) and adjacent intronic sequences | 37/37 with biallelic *STXBP2* variants | 25/37 homozygous<br>12/37 compound heterozygous<br>9 novel variants<br>9 different missense variants<br>4 different splice-site variants<br>small deletions or insertions<br>frequent variants: c.1430C>T (detected in 5 patients of Arab origin), c.1621G>A (detected in 7 patients of mainly white origin), c.247-1 G>C splice-site variant in | 24 patients with biallelic missense variants, in-frame or frameshift deletions and splice-site variants other than exon 15: classic course of disease with early onset and rapidly fatal course if HSCT could not be performed, variable ethnic backgrounds.<br>13 patients with either homozygous or compound heterozygous variants with 1 allele carrying an exon 15 splice-site variant: mild and atypical course. Most developed chronic recurrent course with long episodes of absent HLH symptoms and recurrent reactivations with spontaneous remission or response to steroids-only treatment. Mainly central European ethnic background |

| Study reference;<br>country<br>(ethnicity) | Study design;<br>recruitment | Number and<br>definition of cases | Test | Genes/varia<br>nts<br>considered | Gene frequency in<br>cases<br>Test negatives | Variant frequency | Expressivity |
| --- | --- | --- | --- | --- | --- | --- | --- |
|  |  |  |  |  |  | exon 15 (detected in 12<br>mainly German and<br>Turkish patients) |  |

\*i.e. 43 patients have a total of 45 monoallelic variants HLH – haemophagocytic lymphohistiocytosis; NK – natural killer; NGS: next generation sequencing; WES – whole exome sequencing; y – years; m – months; FPID – foundation of primary immunodeficiency; FHL/fHLH – familial haemophagocytic lymphohistiocytosis

##### MCADD

**Table 24.** Summary tables for the studies exploring gene/variant frequency in patients with MCADD.

| Study reference; country | Study design; recruitment | Number and definition of cases | Test | Genes/variants considered | Gene frequency in cases<br>Test negatives | Variant frequency | Expressivity |
| --- | --- | --- | --- | --- | --- | --- | --- |
| **IEM/IMD** |  |  |  |  |  |  |  |
| Martin-Rivada 2022<sup>58</sup><br>Spain | Retrospective<br>Single-arm<br>newborn screening program experience (14 IEM conditions)<br>Recruitment : 2011 - 2019 | All patients who received an initial abnormal IEM screening result on NBS test and subsequently referred for biochemical and molecular genetic diagnosis. Genetic testing conducted in all biochemically confirmed cases.<br>224/902 with positive initial screening test had biochemically confirmed disease and underwent genetic testing | NR | **ACADM**,<br>*PAH*,<br>*DNAJC12*,<br>*PCBD1*,<br>*GCDH*,<br>*MCCC1*,<br>*MCCC2*,<br>*PCCB*,<br>*MAT1A*, *FAH*,<br>*HPD*,<br>*BCKDHB*,<br>*BCKHBA*,<br>*DBT*, *BCAT2*,<br>*CBS*, *SLC3A1*,<br>*OTC*, *ASS1*,<br>*HMGCL*,<br>*ACADVL*,<br>*HADHA*,<br>*SLC22A5*,<br>*CPT2*, *ETFB*,<br>*CPT1A*,<br>*MMACHC*,<br>*MMADHC*, | 222/224 (99.1%) with biochemically confirmed disease also had a genetically confirmed **diagnosis**<br>**43/224 (19.2%)**<br>**ACADM**<br>*Other IEM genes:*<br>*PAH* (n=83)<br>*DNAJC12* (n=4)<br>*PCBD1* (n=2)<br>*GCDH* (n=12)<br>*MCCC1* (n=2)<br>*MCCC2* (n=5)<br>*PCCB* (n=5)<br>*MAT1A* (n=6)<br>*FAH* (n=3)<br>*HPD* (n=1)<br>*BCKDHB* (n=2) | 14 different genotypes for *ACADM*, 4 of which recurred within the study sample and 10 occurred only in 1 patient each<br>Most common variant 985A>G (59/86 alleles, 66%); 22 cases homozygous 985A>G and 16 cases compound heterozygous including 985A>G.<br>Remaining 5 cases compound heterozygous for other variants | 1 MCADD case with clinical symptoms before newborn screening results were available (and very high C8 levels), homozygous for 985A>G |
|  |  |  |  | MMUT,<br>MMAB | *BCKHBA (n=1)*<br>*DBT (n=1)*<br>*BCAT2 (n=2)*<br>*CBS (n=2)*<br>*SLC3A1 (n=1)*<br>*OTC (n=2)*<br>*ASS1 (n=2)*<br>*HMGCL (n=1)*<br>*ACADVL (n=13)*<br>*HADHA (n=2)*<br>*SLC22A5 (n=11)*<br>*CPT2 (n=2)*<br>*ETFB (n=1)*<br>*CPT1A (n=1)*<br>*MMACHC (n=6)*<br>*MMADHC (n=1)*<br>*MMUT (n=2)*<br>*MMAB (n=2)*<br>**Test negative:**<br>n=2 (patients with biochemical HPA) |  |  |
| **FAOD** |  |  |  |  |  |  |  |
| Maguolo 2020;<sup>59</sup> Italy | Retrospective<br>Single-arm newborn screening program experience<br>Recruitment : Feb 2014 – April 2019 | Patients diagnosed with FAODs via newborn screening or clinical diagnosis<br>20/23 newborns with initial positive screening test had biochemically confirmed disease<br>5/20 biochemical MCADD positive (diagnosed via NBS test)<br>Further 10/11 with clinical dx reported (mean age at onset 29.3y (SD 26.5y)) excluded from review | Sequencing (NGS) with custom designed panel; variants confirmed by Sanger sequencing | **ACADM**,<br>*ACADVL*,<br>*CPT1A*, *CPT2*,<br>*ETFA*, *ETFB*,<br>*ETFDH*,<br>*FLAD1*,<br>*HADHA*,<br>*HADHB*,<br>*SLC23A2*-,<br>*SLC25A32*,<br>*SLC52A1*,<br>*SLC52A2*,<br>*SLC52A3* | 3/20 (15%) *ACADM*<br>Test negative:<br>2/20 (10%) *ACADM* monoallelic<br>Other FAOD genes:<br>5/20 (25%)<br>*ACADVL* (*VLCADD*) n=3<br>*ETFDH* (*MADD*) n=2<br>Other monoallelic genes:<br>*ACADVL* (*VLCADD*) n=1<br>No genetic information<br>10/20<br>*SLC22A5* (*CUD*) (n=3) | 3 biallelic variants: 1 homozygous 985A>G, 2 compound heterozygous:<br>c.817_829del/c.388-14A>G (n=1)<br>c.244insT/c.978G>A (n=1)<br>2 missense<br>1 nonsense<br>1splicing<br>1 frameshift<br>5/5 previously reported | 3/3 enzymatic activity <5%<br>(activity < 10% correlates with certainly symptomatic, 10-20% insufficient evidence, >20% might never manifest severe symptoms, i.e. mild biochemical phenotype) |
|  |  |  |  |  | ACADS (SCADD) (n=6)<br>VLCADD) (n=1) |  |  |
| **ACAD** |  |  |  |  |  |  |  |
| Wang 2019;<sup>60</sup><br>China | Retrospective<br>Single-arm<br>newborn screening program experience<br>Recruitment : 2014 to 2018 | Newborns recalled for further testing following initial positive result for ACAD deficiency on NBS test (HPLC-MS/MS assay). Cases with subsequent positive result on 2<sup>nd</sup> specimen referred for diagnostic testing and genetic analysis.<br>20/83 newborns with positive screening result had confirmed ACAD on diagnostic testing | NSG + Sanger sequencing | 306 genes expanded IEM panel reporting **ACADM**, ACADS, ACADVL | 3/20 (15%) biallelic *ACADM*<br>Test negative:<br>1/20 (5%) monoallelic *ACADM*<br>Other genes:<br>15/20 (75%)<br>*ACADS* (SCAD) (n=10)<br>*ACADVL* (VLCAD) (n=5)<br>No genetic information<br>*SCAD* n=1 | 3/3 cases compound heterozygous<br>5 different variants<br>1 occurred twice: c.449_452DelCTGA(Het)<br>two novel variants: c.589A > G and c.1248T > G,<br>3 previously reported variants: c.449_452delCTGA, c.970G > A, c.1238G > A | NR |
|  |  | 4/20 with confirmed MCADD |  |  |  |  |  |
| **Clinical-biochemical MCADD** |  |  |  |  |  |  |  |
| Mesbah 2020;<sup>62</sup><br>Ireland | Retrospective<br>Single-arm<br>Recruitment :<br>01/01/1998 –<br>30/08/2016 | Children <18 yrs with clinical dx of MCADD identified via the national centre for inherited metabolic disorders and metabolic laboratory at Temple Street Children's University Hospital<br>17 children; 4 dx by family screening and 2 postmortem<br>Average age at clinical presentation: 1.48y [0.005 to 2.86] | Genetic testing (not further specified) | ACADM | 11/17 (64.7%) *ACADM*<br>Missed cases (one variant identified only), 3/17 (17.6%)<br>3/17 clinical MCADD cases had no reported genetic results; likely no genetic testing conducted | 8/11 biallelic *ACADM* variant homozygous (985A>G)<br>3/11 compound heterozygous (985A>G + a second different variant) | NR |
| Nichols 2008;<sup>61</sup><br>United States | Retrospective<br>Single-arm<br>newborn screening program experience<br>Recruitment : first 18 months from MCADD inclusion in NYS NBS programme | Newborns with C8 levels $\geq 0.3 \mu\text{mol/L}$ on NBS test referred for molecular testing<br>511<br>newborns with molecular test results | Sequential:<br>1. PCR/FRET c.985A>G & c.199T>C variant detection<br>If 1 or 2 variants detected OR C8$\geq 0.4 \mu\text{mol/L}$<br>2. complete *ACADM* sequence | 12 exons of *ACADM* | MS/MS positives (n=511):<br>PCR/FRET (2 *ACADM* variants):<br>8/511 (1.6%) biallelic *ACADM*<br>Test negative:<br>157/511 (30.7%) monoallelic<br>Neither variant<br>83/511 (16.2%)<br>Follow-up testing positive (n=20):<br>PCR/FRET + *ACADM* sequencing: | 17/20 with biallelic MCADD<br>13 different variants<br>5 novel variants<br>6/17 homozygote<br>11/17 compound heterozygote<br>Homozygote:<br>6/17 985A>G/985A>G<br>Compound heterozygote:<br>5/17 985A>G compound heterozygote<br>1/17 c.985A>G/c.199T>C | "mild" MCADD<br>c.199Y>C/c.134A>G |
|  |  |  |  |  | 17/20 (85%) biallelic *ACADM*<br>Test negative:<br>3/20 (15%) monoallelic | 2/17 c.199T>C compound heterozygote<br>3/17 compound heterozygote different variants |  |
| Gene (*ACADM*) positive MCADD |  |  |  |  |  |  |  |
| Touw 2012;<sup>63</sup> Netherlands | Retrospective<br>Single-arm newborn screening program experience<br>Recruitment : 2007 - 2010 | Children from Dutch birth cohort with a clinical follow-up in a metabolic centre after positive result for MCADD on population NBS programme<br>Diagnosis confirmed by *ACADM* gene analysis<br>68 children with *ACADM* genotypes / 84 MCADD confirmed | Sequencing of all exons and adjacent intron regions | *ACADM* | 68/68 *ACADM* | 10 known (previously reported) genotypes<br>7 novel genotypes<br>Classic genotypes: 53/68, 77.9%:<br>985A>G/985A>G (n=42)<br>985A>G compound heterozygous (n=8)<br>233T>C/233T>C (n=2)<br>233T>C compound heterozygous (n=1) | Median residual MCAD enzyme activity:<br>In patients with classic *ACADM* genotypes: 0% (range 0-8% and 0-5% in leukocytes and lymphocytes, respectively)<br>In patients with clinically unrecognised genotypes: 25% (range 0-63%) |
|  |  | patients; 108 with initial positive NBS test |  |  |  | Genotypes not previously recognised in clinically confirmed cases: 15/68, 22.1%<br>985A>G compound heterozygous (n=12)<br>c.233 T>C/c.1066A>T (n=1)<br>c.250 C>T/c.199 T>C<br>c.799 G>A/c.865 G>A | 6/17 genotypes with enzyme activity 0%:<br>985A>G/985A>G<br>c.233 T>C/c.789A>G<br>c.985A>G /c.216 + 1 G>T<br>c.985A>G/c.470 C>T<br>c.233 T>C/c.1066A>T<br>c.985A>G/c.928 G>A |

IEM – inborn error of metabolism; IMD – inherited metabolic disorders; FAOD – fatty acid oxidation disorder; NGS – next generation sequencing; NBS – newborn blood spot, MCADD - Medium-chain acyl-CoA dehydrogenase deficiency

#### Evidence on early vs late treatment

##### PDE

**Table 25.** Summary tables for the studies exploring the impact of early vs late treatment in patients with PDE.

| Study reference | Study design | Number and definition of cases | Definition early vs late | Outcome measure and time point | Results |
| --- | --- | --- | --- | --- | --- |
| Bok 2010;<sup>76</sup><br>Netherlands | Family study | 4 (2 families); families in which the mother used PN daily during the second pregnancy from the first trimester onwards (all genetically confirmed) | Antenatal **PN** (following birth, asymptotically or following first seizure); no antenatal or asymptomatic **PN** | Total IQ<br>Motor performance<br>Outcomes measured at ages 5 and 4 (Family 1) and ages 14 and 12 (Family 2) | **Total IQ**<br>*Family 1*<br>Early treated sibling: 98<br>Late treated sibling: 73<br>*Family 2*<br>Early treated sibling: 106<br>Late treated sibling: 80<br>**Motor performance**<br>*Family 1*<br>Early treated sibling: Hypotonic, walking at 27m<br>Late treated sibling: Hypotonic, walking at 31m<br>*Family 2*<br>Early treated sibling: Normal<br>Late treated sibling: Normal |
| Jiao 2021;<sup>77</sup><br>China | Family study | 6 (3 families); patients with PDE (genetically confirmed) | Second born siblings treated with **PN** immediately after first seizure; first born siblings treated with **PN** at 10m (onset 8d), 4y (onset 3d), and 7y (onset 5m) | Psychomotor development evaluated through clinical judgement and parental questionnaires; timing not specified | **Psychomotor development**<br>*Family 1*<br>Early treated sibling: Normal<br>Late treated sibling: Mild delay<br>*Family 2*<br>Early treated sibling: Normal<br>Late treated sibling: Severe delay<br>*Family 3*<br>Early treated sibling: Normal<br>Late treated sibling: Normal |
| Tseng 2022;<sup>79</sup><br>international | Retrospective, single-arm, multi-centre registry study | 37 (18 families); families with PDE (confirmed by elevation of α-AASA or genetically confirmed) where at least one sibling has confirmed ALDH7A1 and there is a difference in age at initiation of treatment of at least seven days | Author defined: Sibling with the shortest delay in treatment initiation (**PN and/or LRT**); sibling with the longest delay in treatment initiation | Full-scale IQ (two scales used); mean age 12.5y in early treated group and 15.3y in late treated group<br>Clinical assessments (if standardised assessment not available); timing not specified | <u>PN monotherapy</u><br>**Mean full-scale IQ**<br>*Available for one sibling pair*<br>Early treated sibling: 106.0<br>Late treated sibling: 80.0<br>**Clinical assessments**<br>*Available for 9 sibling pairs* |
|  |  |  |  |  | Early treated sibling showed a better outcome in four pairs (44%), similar outcome in four pairs (44%) and a worse outcome in one pair (11%).<br><u>PN & LRT</u><br>**Mean (SD) full-scale IQ**<br>*Early treated (n=3): 76.0 (21.63)*<br>*Late treated (n=5): 77.40 (27.93)*<br>**Clinical assessments**<br>*Available for 8 sibling pairs*<br>Early treated sibling showed a better outcome in 5 pairs (62.5%), similar outcome in 2 pairs (25%), and a worse outcome in one pair (12.5%). |
| Coughlin 2022;<sup>68</sup><br>International | Single-arm, multi-centre; patients recruited using registry and ambispective clinical data collected | 60; individuals (any age) with a confirmed diagnosis of PDE-ALDH7A1 (either elevated α-AASA or genetically confirmed) and at least one developmental assessment | **PN+LRT** in the first 6m of life; **PN** monotherapy or **PN+LRT** at >6m of age | Standardised developmental assessment; mean (SD) age at developmental assessment 7.68y (5.49y) | **Mean standardised developmental assessment**<br>*Early treated (n=14 assessments for 8 patients): 87.3 (95% CI: 79.5-95.0)*<br>*Late treated (n=98 assessments from 46 patients): 73.8 (95% CI: 73.8-77.5)* |
|  |  |  |  |  | **Difference in mean standardised developmental assessment**<br>21.9 (95% CI: 1.7-42.0)<br>*After adjustment for confounders* |
| Strijker 2021;<sup>78</sup><br>Netherlands | Cross-sectional, single-arm; retrospective data obtained from international registry | 12; patients with PDE (genetically confirmed) | **LRT** started at <3y; **LRT** started at ≥3y, **no LRT** given | IQ (several scales used in different patients); timing not specified<br>Neurological outcome (two standardised, age-specific neurological assessments); timing not specified | **IQ**<br>*Early treated (n=3):* All had IQ>70<br>*Late treated (n=4):* 1 had IQ>70 and 3 had IQ≤70<br>*No LRT treatment (n=5):* 2 had IQ>70 and 3 had IQ≤70<br>**Neurological outcome**<br>*Early treated (n=3):* 2 neurologically normal, 1 with complex minor neurological dysfunction (MND)<br>*Late treated (n=4):* 3 neurologically normal, 1 with complex MND<br>*No LRT treatment (n=5):* 2 neurologically normal, 3 with complex MND |

PDE – pyridoxine dependent epilepsy; PN – pyridoxine; LRT – lysine reduction therapy; m – months; y – years; d – days; IQ – intelligence quotient

##### hRB

**Table 26.** Summary tables for the studies exploring the impact of early vs late treatment in patients with hRB.

| Study reference | Study design | Number and definition of cases | Definition early vs late | Outcome measure and time point | Results |
| --- | --- | --- | --- | --- | --- |
| Abramson 2003;<br><sup>86</sup> USA | Retrospective,<br>single-arm,<br>single-centre | 264 from sample of 1831;<br>all RB patients who were<br>seen at a single centre<br>between 1914 and 2000 | RB detected by routine<br>screening due to a family<br>history of RB (SFH); no<br>screening, clinical<br>presentation of RB with<br>(FH+) or without FH (FH-) | Age at diagnosis<br>Ocular survival; 1 and<br>5 years<br>Survival; 1 and 5<br>years | **Mean age at Dx, months (SD)**<br>SFH (n=86): 7.7 (1.5)<br>all FH+ (n=264): 10.8 (0.8); NR separately for<br>FH+ with no screening<br>**Ocular survival rate (Kaplan-Meier)**<br>1 year<br>SFH (n=86): 83.2%<br>FH+ (n=178): 47.5%<br>5 year<br>SFH (n=86): 67.7%<br>FH+ (n=178): 58.2%<br>**Survival rate (Kaplan-Meier)**<br>1 year<br>SFH (n=86): 100.0%<br>FH+ (n=178): 97.3%<br>5 year |
|  |  |  |  |  | *SFH (n=86): 93.2%*<br>*FH+ (n=178): 87.4%* |
| Moll 2000;<sup>88</sup> Netherlands | Retrospective, single-arm, multi-centre registry study | 75; patients with a positive family history for RB identified on the Dutch retinoblastoma register | RB detected by screening from birth; no screening (hereditary aspect of RB was not known to proband's parents), clinical presentation of RB | Age at diagnosis | **Mean (median [range]) age at diagnosis**<br>*Screened (n=60): 4.9m (1.9m [1d-48m])*<br>*Not screened (n=25): 17.2m (10.0m [1.5m-63.0m])* |
| Rothschild 2011;<sup>89</sup> France | Retrospective, single-arm, single-centre | 59; all RB patients at a national referral centre with familial RB between 1995 and 2004 | RB detected by screening due to a family history of RB, either 'intensively screened' (IS) or 'screened' (S) if the screening did not meet recommendations; fundus examination due to clinical presentation of RB (NS) | Age at diagnosis;<br>Enucleation; timing not specified<br>Mortality; timing not specified | **Age at diagnosis; p<0.001**<br>*IS (n=16): median 0m (range 0-7 )*<br>*S (n=23): median 4m (range 0-35)*<br>*NS (n=20): median 9m (range 2-57)*<br>**Enucleation of at least one eye (n (%))**<br>*IS (n=16): 0 (0%)*<br>*S (n=23): 2 (8.7%)*<br>*NS (n=20): 13 (65.0%)*<br>**Mortality (n (%))**<br>*IS (n=16): 0 (0%)* |
|  |  |  |  |  | *S (n=23): 0 (0%)*<br>*NS (n=20): 1 (5.0%)* |
| Soliman 2017;<br><sup>90</sup> Egypt | Retrospective, single-arm, single-centre | 13 (10 families); RB cases with a family history at presentation at hospital | RB detected by screening; no screening, clinical presentation of RB | Age at diagnosis<br>Enucleation; evaluated at last follow-up (median (range) follow-up: 4.8y (1.2-9.1))<br>Mortality; timing not specified | **Age at diagnosis (mean)**<br>*Screened 4.7m*<br>*Not screened 16.7m*<br>**Enucleation of at least one eye (n (%))**<br>*Screened (n=5): 0 (0%)*<br>*Not screened (n=8): 6 (75%)*<br>**Mortality**<br>No deaths |
| Chandata 2009;<sup>87</sup> USA, Argentina, Brazil, Turkey, Jordan, Venezuela | Retrospective, single-arm, multi-centre | 92; patients with familial RB (fRB), diagnosed at one of five referral centres | Developed country (USA) with more screening for fRB; developing countries (DC) with limited screening for fRB | Enucleation; timing not specified<br>Probability of event free survival, where an event is defined as extraocular relapse, second malignancies, and death; 5 years | **Enucleation of at least one eye (n (%))**<br>*USA (n=32): 8 (25.0%)*<br>*DC (n=60): 43 (71.7%)*<br>**Probability (SE) of event free survival**<br>*USA (n=32): 0.92 (0.05)*<br>*DC (n=60): 0.81 (0.07)* |

RB – retinoblastoma; y – years; m – months; d – days; SE – standard error

##### XLHR

**Table 27.** Summary tables for the studies exploring the impact of early vs late treatment in patients with XLHR.

| Study reference | Study design | Number and definition of cases | Definition early vs late | Outcome measure and time point | Results |
| --- | --- | --- | --- | --- | --- |
| Makitie 2003;<sup>94</sup> Canada | Retrospective, single arm, single centre | 19; all patients with XLHR (confirmed by medical history, physical examination, radiography, biochemistry tests and FH) | Before development of clinically manifest rickets based on positive family history and biochemistry; Treatment onset age <1.0 year<br>vs<br>After development of clinical signs of rickets; Onset age ≥1.0 year | Height, predicted adult height,<br>Rickets activity.<br>End of first treatment year. | Group 1 (early): n=8, Group 2 (late): n=11<br>**Height:** median z-score:<br>Group 1: -0.7 SDS vs. Group 2: -1.8 SDS; p=0.009, at pre-puberty (9.0 yr): -1.3 SDS vs -2.0 SDS; p= 0.054<br>**Predicted adult height:** z-score Group 1: -0.2 SDS vs Group 2: -1.2 SDS; p=0.06.<br>**S-Pi** (mean change from diagnosis to end of first treatment year): z-score Group 1: +2.2 SDS vs Group 2 +0.8 SDS; p=0.005. Median at end of the first treatment year: Group 1: -1.5 SDS vs Group 2: -3.0 SDS, median at pre-puberty (median age 10.8 y): Group 1: -2.6 SDS vs Group2: -2.5 SDS<br>**S-ALP** improved in both groups during the first treatment year. Median at end of the first treatment year: Group 1: -0.8 SDS vs Group 2: +2.6 SDS, median at pre-puberty (median age 10.8 y): Group 1: -0.7 SDS vs Group2: +2.0 SDS<br>**Rickets activity** (median score at the end of the first treatment): no change +2.0 Group 1 vs improved to 4.0±0.4 Group 2; p=0.052; (median score at prepuberty, median age 10.4 yr): 4.0±0.5 Group 1 vs 5.0 ± 0.7 Group 2; p=0.27. |
| Rafaelsen 2016,<sup>49</sup> Norway | Retrospective, single arm, multi-centre | 19; children who had Serum phosphate below the age dependent reference range in repeated samples combined with tubular maximum reabsorption rate of phosphate per glomerular filtration rate. | Treatment start age < 1.0 y<br>vs<br>Treatment start age > 1.0 y | Height, biochemical parameters, Nephrocalcinosis<br>Skeletal X-ray examinations<br>At each clinic visit from time of diagnosis to the time of study inclusion. | Group 1 (early): n=10, Group 2 (late): n=9<br>At last registered consultation:<br>**Height:** z-score: Group 1: -1.4 (-2.6 to 0.8); Group 2: -2 (-6.3 to 0.3)<br>Delta z-score: Group 1: -0.4 (-3.1 to 2.0); Group 2: 0 (-1.1 to 1.3)<br>**Persistent bowing:** Group 1: 5/10; Group 2: 7/9<br>**Dental involvement:** Group 1 2/10; Group 2: 7/9 |
| Quinlan 2012,<sup>95</sup><br>UK | Retrospective,<br>single arm, single<br>centre | 23; Children with PHEX<br>variant, Treatment<br>duration >2 years, >2 clinic<br>visits per year | Treatment start age < 1.0 y<br>vs<br>Treatment start age > 1.0 y | Height, biochemical<br>parameters,<br>Rickets activity,<br>Renal ultrasound | Group 1 (early): n=10,<br>Group 2 (late): n=13<br>**Height** (score (SD): G1: -0.7 (-1.5, 0.3) vs.G2:<br>-2.0 (-2.3 to -1.0); p=0.009 at medium<br>treatment years of 8.5 vs 11.9 (p=0.557)<br>**Rickets severity score** (available for 20<br>patients, 11 in G1 and 9 in G2): median=1 in<br>both groups<br>**s-Pi**: median standard deviation score: G1: -<br>0.54 (-0.45 to -0.60) vs G2: -0.52 (-0.49 to<br>-0.59); p=0.92<br>**s-ALP** median: G1: 0.9 (0.8–1.2) vs G2: 0.8 (0.7–<br>0.9); p= 0.13<br>throughout treatment for both biochemical<br>markers |

y – year

##### fHLH

**Table 28.** Summary tables for the studies exploring the impact of early vs late treatment in patients with fHLH.

| Study reference<br>(extraction approach) | Study design | Number and definition of cases | Definition early vs late | Outcome measure and time point | Results |
| --- | --- | --- | --- | --- | --- |
| Lucchini 2018;<sup>75</sup> International<br>(per protocol analysis) | Retrospective, multi-centre | 51/66; fHLH (sibling pairs/triplets) with variant on either PRF1, UNC13D, STX11 or STXBP2, either symptomatic or asymptomatic at diagnosis (and diagnosed following sibling diagnosis)<br>One asymptomatic case who did not receive any treatment excluded | **Early**<br>Cases that were asymptomatic at diagnosis and did not activate before treatment initiation<br>*Group 1:* Asymptomatic cases without activation prior to HSCT +/- prophylactic treatment<br>*Group 2:* Asymptomatic cases with activation after the start of prophylactic treatment<br>**Late**<br>Cases that were symptomatic at diagnosis (index cases) or asymptomatic cases that activated before treatment initiation<br>*Group 3:* Index cases who were symptomatic at diagnosis and treated<br>with active disease protocol +/- HSCT<br>*Group 4:* Asymptomatic cases with activation before treatment initiation and treated with active disease protocol + HSCT | Mortality, cause of death, those in complete remission at end of follow-up; variable follow-up | **Mortality (n (%)); cause of death**<br>*Group 1 (n=15):* 1 (7%); sudden death while in complete remission<br>*Group 2 (n=3):* 1 (33%); disease progression<br>*Group 3 (n= 26):* 10 (38%); 4 transplant related mortality, 5 disease progression, 1 died as a result of infection<br>*Group 4 (n=7):* 2 (29%); 2 transplant related mortality<br>**Complete remission (n (%)); median [range] length of follow-up (months)**<br>*Group 1 (n=15):* 14 (93%); 37 [17-89]<br>*Group 2 (n=3):* 1 (33%); 48<br>*Group 3 (n=26):* 10 (38%); 41.5 [12-144]<br>*Group 4 (n=7):* 5 (7%); 48 [12-72]<br>*Note that one patient in group 2 was lost to follow-up* |
| Lucchini 2018;<sup>75</sup> International (Intention to treat) | Retrospective, multi-centre | 52/66; fHLH (sibling pairs/triplets) with variant on either *PRF1*, *UNC13D*, *STX11* or *STXBP2*, either symptomatic or asymptomatic at diagnosis (and diagnosed following sibling diagnosis) | **Early**<br>Cases that were asymptomatic at diagnosis<br>**Late**<br>Cases that were symptomatic at diagnosis | Mortality, cause of death, those in complete remission at end of follow-up; variable follow-up | **Mortality (n (%)); cause of death**<br>*Early (n=26):* 4 (15%); 2 transplant related mortality, 1 HLH progression, 1 sudden death in complete remission<br>*Late (n=26):* 10 (38%); 4 transplant related mortality, 5 HLH progression, 1 "died as a result of infection"<br>**Complete remission (n (%)); median [range] length of follow-up (months)**<br>*Early (n=26):* 21 (81%); 42 [12 - 144]<br>*Late (n=26):* 16 (62%); 41.5 [12-144]<br>*Note that one patient in the early treated group was lost to follow-up* |

fHLH – familial haemophagocytic lymphohistiocytosis; HLH – haemophagocytic lymphohistiocytosis; HSCT – Haematopoietic Stem Cell Transplant; y – year; m – months; d – days

##### MCADD

**Table 29.** Summary tables for the studies exploring the impact of early vs late treatment in patients with MCADD.

| Study reference | Study design | Number and definition of cases | Definition early vs late | Outcome measure and time point | Results |
| --- | --- | --- | --- | --- | --- |
| Abdenur 1999;<sup>97</sup> Argentina | Retrospective, 2 patient case series | 2; MCADD confirmed by biochemical testing and variant analysis | **Early**<br>Management with high carbohydrate diet from 5m age following positive NBS test or sibling detected (asymptomatic)<br>**Late**<br>Management with high carbohydrate diet following symptomatic clinical presentation (Reye-like syndrome) | Description of the clinical status of the patients; assessed at age 34m for early treated case and 24m for late treated case | **Clinical status description**<br>*Early (n=1):* no decompensation, height, weight and head circumference at 25<sup>th</sup> to 50<sup>th</sup> centile, physically normal<br>*Late (n=1):* severe seizure disorder and cerebral palsy, nasogastric feeding |
| Alcaide 2022;<sup>98</sup> Spain | Retrospective, single-arm, multi-centre | 33; neonates with genetically confirmed or strong clinical and biochemical features of MCADD | **Early**<br>Management with avoidance of fasting and carnitine supplementation following positive NBS test (includes symptomatic patients), or detected through family screening (asymptomatic)<br>**Late**<br>Management with avoidance of fasting and/or carnitine supplementation following clinical presentation | Description of the clinical status of the symptomatic patients; no standardised follow-up time | **Clinical status description of symptomatic patients**<br>*Early (n=31):* 9/31 patients symptomatic<br>Conduct disorder (age 10y) n=1<br>Autistic behaviour, language retardation and macrosomy n=1<br>Hospital admissions for vomiting and food intolerance n=1<br>Fasting hypoglycaemia n=1<br>Autistic behaviour and hospitalisation for decompensation (age 2y) n=1<br>Hypotonia and food intolerance n=1 |
|  |  |  | *Note that one patient in the early treated group had no management and management was poorly followed in another patient* |  | Creatine kinase elevation n=1<br>Jaundice n=1<br>Liver crisis and decompensation (age 2y) n=1<br>*Late (n=2): both patients symptomatic*<br>Convulsive status epilepticus, fasting hypoglycaemias and conduct disorder (age 16m) n=1<br>Cyclical vomiting, abdominal pain, temperature episode (age 10y) n=1 |
| Anderson 2020;<sup>99</sup> USA | Retrospective, single-arm, single-centre | 90; diagnosed with MCADD following NBS screening, family history or clinical presentation | **Early**<br>Management with avoidance of fasting, carnitine supplementation and diet following positive NBS test (asymptomatic)<br>**Late**<br>Management with avoidance of fasting, carnitine supplementation and diet following symptomatic clinical presentation, or detected through family screening | Number of hypoglycaemia related hospital admissions and emergency room visits; timing not reported | **Mean (95% CI) number of hypoglycaemia related hospital days and ER visits per patient**<br>*Early (n=76): 0.62 (0.21-1.04)*<br>*Late (n=16): 2.15 (0.67-3.58)*<br>**Mean (95% CI) number of hypoglycaemia related hospital days and ER visits per patient years**<br>*Early (n=76): 0.09 (0.03-0.15)*<br>*Late (n=16): 0.11 (0.04-0.19)* |
| Gong 2021;<sup>100</sup> China | Retrospective, single-arm, single-centre | 24; genetically confirmed MCADD patients recruited between 2009 and 2019 | **Early**<br>Management with carnitine supplementation and diet following positive NBS test (asymptomatic)<br>**Late**<br>Management with carnitine supplementation and diet following symptomatic clinical presentation | Biomarkers; measured at presentation (aged between 0.5m and 36m)<br>Mortality, description of the clinical status of the patients; at study close (range of follow-up times depending on age at start of study) | **Mean C6**<br>*Early (n=18): 0.52*<br>*Late (n=4): 0.16*<br>**Mean C8**<br>*Early (n=18): 2.09*<br>*Late (n=6): 0.60*<br>**Mean C10**<br>*Early (n=16): 0.27*<br>*Late (n=5): 0.10*<br>**Mean C8:C2 ratio**<br>*Early (n=15): 0.25*<br>*Late (n=4): 0.07*<br>**Mean C8:C10 ratio**<br>*Early (n=18): 9.23* |
|  |  |  |  |  | *Late (n=5): 7.94*<br>**Mortality (n (%))**<br>*Early (n=15): 0 (%)*<br>*Late (n=5): 0 (0%)*<br>**Clinical status description**<br>*Early (n=15): all healthy and asymptomatic*<br>*Late (n=5): one patient with intermittent fasting hypoglycaemia, one patient with hemiplegia due to disease episode and 3 healthy and asymptomatic* |
| Haas 2007;<br><sup>101</sup> Australia* | Retrospective, single-arm, multi-centre | 59; children with MCADD diagnosed clinically or by NBS screening between April 1994 and March 2002 | **Early**<br>Management following positive NBS test (asymptomatic)<br>**Late**<br>Management following symptomatic clinical presentation | Mortality, healthcare use (inpatient stays, emergency department visits, outpatient services); within the first four years of life | **Mortality (n (%))**<br>*Early (n=24): 1 (4%)*<br>*Late (n=35): 6 (17%)*<br><u>Inpatient stays</u><br>**Patients with inpatient stays (n (%))** |
|  |  |  |  |  | *Early (n=24): 10 (42%)*<br>*Late (n=35): 25 (71%)*<br>**Mean (SD) number of admissions per year in children admitted at least once**<br>*Early (n=10): 1.1 (1.4)*<br>*Late (n=25): 0.9 (0.9)*<br>**Mean (SD) length of stay**<br>*Early (n=10): 2.5d (2.6)*<br>*Late (n=25): 2.6d (3.1)*<br><u>Emergency department visits</u><br>**Patients with emergency department visits (n (%))**<br>*Early (n=24): 5 (21%)*<br>*Late (n=35): 12 (34%)*<br>**Mean (SD) number of visits per year in children who visited the ED at least once** |
|  |  |  |  |  | *Early (n=5): 0.5 (0.2)*<br>*Late (n=12): 1.3 (1.3)*<br><u>Outpatient services</u><br>**Patients who used outpatient services (n (%))**<br>*Early (n=24): 15 (63%)*<br>*Late (n=35): 21 (60%)*<br>**Mean (SD) number of uses per annum in children who used outpatient services at least once**<br>*Early (n=15): 1.5 (1.0)*<br>*Late (n=21): 1.6 (1.4)* |
| Li 2019;<sup>102</sup> China | Retrospective, single-arm | 6; children with MCADD diagnosed clinically or by NBS screening between January 2007 and June 2017 | **Early**<br>Management with diet following positive NBS test (asymptomatic) or management with carnitine supplementation, avoidance of fasting and diet following positive NBS test (symptomatic)<br>**Late** | Description of the clinical status of the patients; variable follow-up (from 3y to 11y) | **Clinical status description**<br>*Early (n=4): all normal*<br>*Late (n=2): all normal* |
|  |  |  | Management with diet±carnitine supplementation following symptomatic clinical presentation |  |  |
| Wilcken, 2007;<sup>103</sup> Australia* | Retrospective, single-arm, multi-centre | 81; children with MCADD diagnosed clinically or by NBS screening born between 01/04/1994 and 31/03/2004<br>**Cohort 1** includes those diagnosed between **1994-2002** (n=59); **cohort 2** also includes patients diagnosed between **2002-2004** (n=81) | **Early**<br>Management with avoidance of fasting, sick-day regimen following positive screening<br>**Late**<br>Management with avoidance of fasting, sick-day regimen following symptomatic clinical presentation | Diagnosed by age 2 (cohort 2)<br>Diagnosed by age 4 (cohort 1)<br>Mean age at diagnosis (cohort 1)<br>Mortality, severe episodes; assessed at age 2 (cohort 2) and age 4 (cohort 1)<br>Number admitted to hospital, total length of stay; assessed at age 4 (cohort 1) | **Diagnosed by age 2 (n (%))**<br>*Early (n=41):* 41 (100%)<br>*Late (n=40):* 28 (70%)<br>**Diagnosed by age 4 (n (%))**<br>*Early (n=24):* 24 (100%)<br>*Late (n=35):* 26 (74%)<br>**Mean (range) age at diagnosis (cohort 1)**<br>*Early (n=24):* 0.5m (0.1-3)<br>*Late (n=26):* 16m (0.1-93)<br>**Mortality (n (%))**<br>At age 2<br>*Early (n=41):* 1 (2%)<br>*Late (n=28):* 4 (14%) |
|  |  |  |  | Neuropsychological outcome (intellectual ability score WJ III); assessed at age >4 (cohort)<br>*Note that outcomes assessed at age 4 are for those in cohort 1 and outcomes assessed at age 2 are for those in cohort 2* | At age 4<br>*Early (n=24): 1 (4%)*<br>*Late (n=26): 5 (19%)*<br>**Severe episodes (n (%))**<br>At age 2<br>*Early (n=41): 1 (2%)*<br>*Late (n=28): 18 (64%)*<br>At age 4<br>*Early (n=24): 2 (8%)*<br>*Late (n=26): 18 (69%)*<br>**Admission to hospital (n (%)) (cohort 1)**<br>*Early (n=24): 10 (42%)*<br>*Late (n=26): 22 (85%)*<br>**Mean (SD) length of stay per admission days (cohort 1)**<br>*Early (n=10): 2.35 (2.6)* |
|  |  |  |  |  | *Late (n=22): 2.95 (3.4)*<br>**Mean (SD) WJ III score (cohort 1)**<br>*Early (n=25): 103.6 (11.7)*<br>*Late (n=13): 104.9 (14.8)* |
| Wilcken 2009;<sup>104</sup> Australia* | Retrospective, single-arm, multi-centre | 59; children with MCADD diagnosed clinically or by NBS screening between 01/04/1994-31/03/2002 (of 116 diagnosed with IEM) | **Early**<br>Management following positive screening<br>**Late**<br>Management following symptomatic clinical presentation | Mortality, intellectual handicap, school placement (extra help/special class/special school), physical handicap; outcomes measured at 6y or latest clinical review for those lost to follow-up | **Mortality (n (%))**<br>*Early (n=24): 5 (21%)*<br>*Late (n=35): 6 (17%)*<br>**Intellectual handicap**<br>*Early (n=22): 0 (0%)*<br>*Late (n=28): 1 (4%) (mild handicap)*<br>**School placement (extra help/special class/special school) (n (%))**<br>*Early (n=15): 0 (0%)*<br>*Late (n=26): 2 (8%) (extra help)*<br>**Physical handicap** |
|  |  |  |  |  | *Early (n=22): 0 (0%)*<br>*Late (n=29): 0 (0%)* |
| Wilson 1999;<sup>105</sup><br>United Kingdom | Retrospective, single-arm, single-centre | 41; all children with MCADD seen acutely or as outpatients between September 1993 and September 1997 | **Early**<br>Management with avoidance of fasting, sick-day regimen and carnitine supplementation following asymptomatic sibling-detection<br>**Late**<br>Management with avoidance of fasting, sick-day regimen and carnitine supplementation following symptomatic clinical presentation | Number (and severity) of episodes requiring hospital admission before diagnosis<br>Mortality, number (and severity) of episodes requiring hospital admission after diagnosis;<br>at latest follow-up<br>variable follow up median 6y (10m – 14y) | **Admissions before diagnosis (n (%))**<br>*Early (n=8): 0 (0%)*<br>*Late (n=33): 17 (52%)* (8 admitted for a coma, 9 admitted for lethargy/hypoglycaemia)<br>**Mortality (n (%))**<br>*Early (n=8): 0 (0%)*<br>*Late (n=33): 0 (0%)*<br>**Admissions after diagnosis (n (%))**<br>*Early (n=8): 2 (25%)* (admitted for lethargy/hypoglycaemia)<br>*Late (n=33): 12 (36%)* (2 admitted for coma, 10 admitted for lethargy/hypoglycaemia) |

\*cohort overlap; MCADD – Medium Chain Acyl CoA Dehydrogenase Deficiency; NBS – Newborn Blood Spot; y – years, m – months, d – days

### Appendix 6. Reference lists from the ClinGen paediatric reports for XLHR, MCADD, PDE and hRB

*List of references cited in the ClinGen paediatric reports for XLHR, MCADD, PDE, and hRB and assessment against our inclusion and exclusion criteria for the review of five conditions*.

**Table 30.** List of references from the ClinGen paediatric reports for XLHR, MCADD, PDE and hRB.

| Condition | Date of search | References underlying ClinGen scores for penetrance and treatment effectiveness and final assertion | Reference in WGS review |
| --- | --- | --- | --- |
| XLHR | 05/11/2018 | MD Ruppe. X-Linked Hypophosphatemia. 2012 Feb 09 [Updated 2017 Apr 13]. In: MP Adam, HH Ardinger, RA Pagon, et al., editors. GeneReviews® [Internet]. Seattle (WA): University of Washington, Seattle; 1993-2024. Available from: http://www.ncbi.nlm.nih.gov/books/NBK83985<br>Online Medelian Inheritance in Man, OMIM®. Johns Hopkins University, Baltimore, MD. HYPOPHOSPHATEMIC RICKETS, X-LINKED DOMINANT; XLHR. MIM: 307800: 2017 Mar 03. World Wide Web URL: http://omim.org.<br>Carpenter TO, Imel EA, Holm IA, Jan de Beur SM, Insogna KL. A clinician's guide to X-linked hypophosphatemia. J Bone Miner Res. (2011) 26(7):1381-8.<br>Carpenter TO, Whyte MP, Imel EA, Boot AM, Hogler W, Linglart A, Padidela R, Van't Hoff W, Mao M, Chen CY, Skrinar A, Kakkis E, San Martin J, Portale AA. Burosumab Therapy in Children with X-Linked Hypophosphatemia. N Engl J Med. (2018) 378(21):1987-1998.<br>Whyte MP, Carpenter TO, Gottesman GS, Mao M, Skrinar A, San Martin J, Imel EA. Efficacy and safety of burosumab in children aged 1-4 years with X-linked hypophosphataemia: a multicentre, open-label, phase 2 trial. Lancet Diabetes Endocrinol. (2019) | NA<br>NA<br>Excluded on T/A (non-systematic review)<br>Excluded on FT (children with active rickets – not early vs late)<br>Excluded on T/A (children with conventional therapy – not early vs late) |
| PDE | 29/05/2019 | SM Gospe. Pyridoxine-Dependent Epilepsy. 2001 Dec 07 [Updated 2017 Apr 13]. In: MP Adam, HH Arding, RA Pagon, et al., editors. GeneReviews® [Internet]. Seattle (WA): University of Washington, Seattle; 1993-2024. Available from: http://www.ncbi.nlm.nih.gov/books/NBK1486<br>Stockler S, Plecko B, Gospe SM Jr, Coulter-Mackie M, Connolly M, van Karnebeek C, Mercimek-Mahmutoglu S, Hartmann H, Scharer G, Struijs E, Tein I, Jakobs C, Clayton P, Van Hove JL. Pyridoxine dependent epilepsy and antiquitin deficiency: clinical and molecular characteristics and recommendations for diagnosis, treatment and follow-up. Mol Genet Metab. (2011) 104(1-2):48-60.<br>van Karnebeek CD, Stockler-Ipsiroglu S, Jaggumantri S, Assmann B, Baxter P, Buhas D, Bok LA, Cheng B, Coughlin CR 2nd, Das AM, Giezen A, Al-Hertani W, Ho G, Meyer U, Mills P, Plecko B, Struys E, Ueda K, Albersen M, Verhoeven N, Gospe SM Jr, Gallagher RC, Van Hove JK, Hartmann H. Lysine-Restricted Diet as Adjunct Therapy for Pyridoxine-Dependent Epilepsy: The PDE Consortium Consensus Recommendations. JIMD Rep. (2014) 15:1-11. | NA<br>excluded on T/A (non-systematic review)<br>excluded on T/A (consortium recommendation) |
| MCADD | 03/08/2020 | Merritt JL, Chang JJ. Medium-Chain Acyl-Coenzyme A Dehydrogenase Deficiency. GeneReviews® (1993)<br>New England Consortium of Metabolic Programs. Medium Chain Acyl-CoA Dehydrogenase Deficiency (MCADD). (2013) Website: https://newenglandconsortium.org/for-professionals/acute-illness-materials/fatty-acid-oxidation-disorders/medium-chain-acyl-coa-dehydrogenase-deficiency-mcadd/<br>Frazier, D.M. Medium chain acyl CoA dehydrogenase deficiency (MCADD). Genetic Metabolic Dietitians International: Nutrition Guidelines. (2008) Website: http://gmdi.org/Resources/Nutrition-Guidelines/MCAD<br>British Inherited Metabolic Diseases Group. Medium Chain Acyl-CoA Dehydrogenase Deficiency (MCADD) - Acute Illness/Decomposition (standard version). (2020) Website: http://www.bimdg.org.uk/store/guidelines/ER-MCADD-v5_232766_05042017.pdf | NA<br>NA<br>NA<br>NA |
|  |  | British Inherited Metabolic Diseases Group. Adult emergency management; Medium chain fat oxidation disorders. (2018) Website: http://www.bimdg.org.uk/store/guidelines/ADULT_MCAD-rev_2015_566641_09012016.pdf<br>Dixon, M. Champion, M. British Inherited Metabolic Diseases Group - Dietitians' Group. Medium Chain Acyl-CoA Dehydrogenase Deficiency (MCADD) - Dietary management guidelines for dietitians. (2007) Website: http://www.bimdg.org.uk/store/guidelines/ER-MCADD-v5_232766_05042017.pdf<br>Andresen BS, Lund AM, Hougaard DM, Christensen E, Gahrn B, Christensen M, Bross P, Vested A, Simonsen H, Skogstrand K, Olpin S, Brandt NJ, Skovby F, Nørgaard-Pedersen B, Gregersen N. MCAD deficiency in Denmark. Mol Genet Metab. (2012) 106(1096-7206):175-88. | NA<br>NA<br>Included in Q2 |
| hRB | 30/04/2018 | National Retinoblastoma Strategy Canadian Guidelines for Care: Strategie therapeutique du retinoblastome guide clinique canadien. Can J Ophthalmol. Canadian journal of ophthalmology. Journal canadien d'ophtalmologie. (2009) 44 Suppl 2:S1-88.<br>Lohmann D, Gallie B, Dommering C, Gauthier-Villars M. Clinical utility gene card for: retinoblastoma. Eur J Hum Genet. (2011) 19(3).<br>Skalet AH, Gombos DS, Gallie BL, Kim JW, Shields CL, Marr BP, Plon SE, Chevez-Barrios P. Screening Children at Risk for Retinoblastoma: Consensus Report from the American Association of Ophthalmic Oncologists and Pathologists. Ophthalmology. (2018) 125(3):453-458. | Excluded on T/A (guideline)<br>Excluded on T/A (Gene card)<br>Excluded on T/A (Consensus report) |
|  |  | DR Lohmann, BL Gallie. Retinoblastoma. 2000 Jul 18 [Updated 2015 Nov 19]. In: MP Adam, HH Ardinger, RA Pagon, et al., editors. GeneReviews® [Internet]. Seattle (WA): University of Washington, Seattle; 1993-2024. Available from: http://www.ncbi.nlm.nih.gov/books/NBK1452<br>Aerts I, Lumbroso-Le Rouic L, Gauthier-Villars M, Brisse H, Doz F, Desjardins L. Retinoblastoma. Orphanet J Rare Dis. (2006) 1:31.<br>Online Medelian Inheritance in Man, OMIM®. Johns Hopkins University, Baltimore, MD. RETINOBLASTOMA; RB1. MIM: 180200: 2016 Aug 04. World Wide Web URL: http://omim.org. | NA<br>Excluded on T/A<br>(non-systematic<br>review)<br>NA |

### Appendix 7. ClinGen information on penetrance and treatment effectiveness for XLHR, MCADD, PDE and hRB

#### Information on penetrance and treatment effectiveness from the paediatric actionability reports available on ClinGen for XLHR, MCADD, PDE and hRB

**Table 31.** ClinGen information on penetrance and treatment effectiveness for XLHR, MCADD, PDE and hRB.

| Condition | Gene | Penetrance | Expressivity | Patient Management |
| --- | --- | --- | --- | --- |
| XLHR | *PHEX* | Penetrance is believed to be 100% by age one year (GeneReviews, OMIM, **Tier 3**) | Individuals with XLHR have variable expressivity (OMIM, **Tier 3**)<br>The severity can differ among members of the same family. (GeneReviews, **Tier 4**) | Treatment for XLHR focuses on improving pain and correcting bone deformities. Treatment from time of diagnosis until growth is complete.<br>Treatment: oral phosphate administered three to five times daily and high-dose calcitriol. Treatment effectiveness: partially corrects leg deformities, decreases the number of necessary surgeries, improves adult height. Many patients can still have suboptimal growth and bone healing with treatment. Early treatment (<1 year of age) had higher median height z-score lower serum alkaline phosphatase, less marked radiographic signs of rickets early treatment does not completely normalize skeletal development. (GeneReviews, OMIM, Carpenter 2011, **Tier 3**)<br>Burosumab has not been incorporated into practice guidelines to date. An open-label study showed improved renal tubular phosphate reabsorption, serum phosphorus levels, standing height, and physical function, reduced pain and severity of rickets at week 64 of treatment (Carpenter 2018, Whyte 2019, **Tier 5**) |
| MCADD | *ACADM* | In effected individuals without established diagnosis of MCADD, at least 18% and up to 25% individuals die | MCADD may remain asymptomatic. A “milder” biochemical phenotype can still develop into life-threatening symptoms. | The standard treatment is lifelong supplementation of pyridoxine. One registry study showed that 75% of patients were seizure-free on monotherapy, 13% were controlled on pyridoxine plus additional |
|  |  | during their first metabolic crisis. (GeneReviews, **Tier 3**)<br>This MCADD estimate is four times higher in screened newborns that in clinically presenting cases during the 10-year period prior to initiation of newborn screening (potentially due to a) a different in variant spectrum - screened population with high numbers of variants associated with a milder biochemical phenotype, and b) a reduced penetrance of 50% of the common c.985A>G variant). (Andresen 2012, **Tier 5**) | (GeneReviews, **Tier 3**) | anticonvulsants, 13% continued to have seizures (Stockler 2011, **Tier 2**) |
| PDE | ALDH7A1 | No penetrance data from unselected populations<br>All individuals with PDE have intractable seizures that respond both clinically and electrographically to large daily supplements of pyridoxine. (GeneReviews, **Tier 4**) | Substantial inter-patient differences in the required effective dose related to the underlying *ALDH7A1* pathogenic variant<br>Seizure types are variable even in the individual patient. Patients treated as early as in the neonatal period may have developmental delay and intellectual disability, whereas other patients with prolonged status epilepticus and later diagnosis may have a normal IQ. (Stockler 2011, **Tier 3**)<br>No clear genotype-phenotype correlations are known. Pathogenic missense variants that result in residual enzyme activity may be associated with | Management by avoidance of prolonged fasting to prevent primary manifestations in asymptomatic patients with MCADD. Infants and children require frequent feedings, with maximum “safe fasting times” of 4-12 hours depending on age. Times are based on the few single patient reports of controlled fasting studies and feeding practices in screened populations (Frazier 2008, Dixon 2007, **Tier 2**)<br>Immediate management during an acute crisis is key to preventing sudden death. Treatment is aimed at reversal of catabolism and prevention of hypoglycemia by giving simple carbohydrates by mouth or intravenous fluids. (New England Consortium of Metabolic Programs 2013, Frazier 2008, British Inherited Metabolic Disease Group 2018, Inherited Metabolic Disease Group 2020, Dixon 2007, **Tier 2**) |
|  |  |  | a more favourable developmental phenotype. (GeneReviews, van Karnebeek, **Tier 3**) | Low-dose L-carnitine supplementation is recommended when carnitine levels are below the normal range (Frazier 2008, **Tier 2**) |
| hRB | *RB1* | > 90% patients with bilateral RB and 15% of patients with unilateral RB have germinal RB1 variants. (National Retinoblastoma Strategy Canadian Guidelines for Care, Lohmann 2011, GeneReviews, Aerts 2006, **Tier 3**)<br><10% of families show a “low penetrance” phenotype with reduced expressivity and incomplete penetrance (i.e., ≤25%). (GeneReviews, **Tier 4**) | RB1 gene has 3 expression patterns: unilateral or bilateral retinoblastoma, retinoma, or no visible retinal pathology except for “normal degeneration” with age. (OMIM, **Tier 3**) | Clinical screening, including examination by an ophthalmologist with experience in RB, from birth. If abnormal frequent examinations under anaesthesia by a paediatric anaesthetist.<br>Early diagnosis, when tumours are small, maximizes survival and vision outcomes and reduces the need for chemotherapy, enucleation, and radiotherapy (one retrospective study) (National Retinoblastoma Strategy Canadian Guidelines of care 2009, Skalet 2018, **Tier 2**) |

## List of supplements

Supplement 1 Characteristics of the five conditions under review

*Overview of the natural history, genetics and epidemiology, screening and diagnosis and treatment of PDE, hRB, XLHR, fHLH and MCADD*

Supplement 2 Excluded studies

*Studies excluded at full text sifting stage and reasons for exclusion for the review of five conditions, the review of genomic studies of paediatric cohorts reporting penetrance for pathogenic variants and the review of cost-effectiveness evaluations of WGS and WES*.

Supplement 3 Included studies for Q2 (prevalence of genetic variants in those with biochemical or biochemical and clinical features of each condition)

*Overview and characteristics of included studies for Q2 (prevalence of genetic variants in those with biochemical or biochemical and clinical features of each condition) of the review of five conditions including study aim, population, total population number in study, eligible population number for the review, study outcome and disease category of included population*.

Supplement 4 Data extraction of 14 studies included in the review of genomic studies of paediatric cohorts reporting penetrance for pathogenic variants

*Data extraction sheet for studies included in the review of genomic studies of paediatric cohorts reporting penetrance for pathogenic variants*

Supplement 5 Data extraction of 86 studies included in the review of cost-effectiveness evaluations of WGS and WES

*Data extraction sheet for studies included in the review of cost-effectiveness evaluations of WGS and WES*.

## Glossary

Biallelic: Referring to both alleles of a gene
ClinGen: Resource funded by the National Institutes of Health that centralises information about the clinical relevance of genes and variants
Clinical actionability: Level of evidence about pathogenicity, penetrance and expressivity of a genetic variant and the extent to which interventions can be used to mitigate the effect of the disease
Digenic: Referring to two genes
Dosage sensitivity: Refers to the impact of changes in gene dosage (the number of copies of a gene) on an organism’s phenotype
Exon: Coding sequence of DNA within a gene that is retained during the RNA splicing process before translation into protein
Expressivity: Degree to which a trait/condition is expressed in individuals with a particular genetic variant
Founder effect: Phenomenon where a small group of individuals establishes a new population, leading to reduced genetic diversity compared to the original population. The presence of an allele at an unusually high frequency in an isolated population.
Gene-disease validity: Level of evidence supporting the association between a specific gene and a disease/phenotype
Gene dosage: Number of copies of a particular gene present in a cell/organism and hence related to the amount of gene product
Generation Study: UK study run by Genomics England in partnership with the NHS to sequence the genome of newborn babies in the UK and screen for 200 rare genetic conditions
Genomics England: Company set up by the UK Department of Health and Social care to run the 100,000 Genomes Project. It is now overseeing the Generation Study
Germline variant: A variant within germ cells that can be passed on to offspring
Intron: Non-coding sequence of DNA within a gene that is removed during the RNA splicing process before translation into protein
Monoallelic: Referring to a single allele of a gene
Mosaicism: Refers to the presence of multiple populations of cells with different genotypes in a single individual due to variants occurring in some cells during development
Penetrance: Proportion of individuals with a given genetic variant who display the traits/condition
Private variants: Gene variants that is specific to an individual or family and not commonly found in the wider population
Proband: Individual in a family who is first identified as having a particular genetic condition
Sporadic variants: Gene variants that arise spontaneously in an individual without being inherited from a parent
Vairant annotation: Variant annotation is the process of assigning functional information to an DNA alteration such as the effect on protein structure.
Variant: Change/alteration in the DNA sequence that may affect how a gene functions. Variants can be benign, pathogenic or of unknown significance
Variant interpretation: Variant interpretation is the process of drawing direct links between individual variants and a disease phenotype for decisions on reporting. This requires expert interpretation and literature review and Consideration of context (other genetic factors, environmental factors, ancestry, sex, type of phenotype (symptomatic disease) under Consideration). Guidelines for variant interpretation for instance by ACMG are available to standardise interpretation of variant pathogenicity and categorisation into pathogenic, likely pathogenic, uncertain, likely benign and benign.
Variant of unknown significance: Variant for which there is insufficient information to determine its impact on disease risk/health.
Variant prioritisation: Variant prioritisation is the process of filtering variants identified through sequencing using bioinformatic tools to identify variants most likely linked to a disease phenotype. More advanced tools include information on predictions about effect on protein structure, conservation (conserved sequences across all vertebrates), constraint (gene regions intolerant to loss of function variants), variant frequency, mode of inheritance and gene-disease associations.
Pathogenicity (variant pathogenicity): A variant is classified as pathogenic if evidence confirms that it causes disease based on variant interpretation. Information for variant interpretation and variant prioritisation is generally based on family, clinical and case control studies which are enriched for etiological co-factors, which means penetrance in these cohorts is overestimated. Pathogenicity annotations used in the screening context have got implications for specificity as ‘known’ pathogenic variants have lower penetrance in population-based studies. Degree to which a variant impacts the function of a gene. >99% probability of pathogenicity defined as pathogenic variant and 90%–99% probability of pathogenicity defined as likely pathogenic variant.
Whole exome sequencing: Technique for sequencing all the protein-coding regions of genes (exons) in a genome
Whole genome sequencing: Technique for sequencing the entire DNA (genome) of an individual
AABR: Automated Auditory Brainstem Response
ACMG: American College of Medical Genetics and Genomics
CEA: Cost-Effectiveness Analysis
ClinGen: Clinical Genome Resource
ClinVar: Clinical Variation
CNV: Copy Number Variant
CRD: Centre For Reviews And Dissemination
DBS: Dried Blood Spot
DEIB: Diversity, Equity, Inclusion, And Belonging
DNA: Deoxyribonucleic Acid
FAOD: Fatty Acid Oxidation Disorders
FH: Family History
fHLH: Familial Hemophagocytic Lymphohistiocytosis
G6PDD: Glucose-6-phosphate dehydrogenase deficiency
GEL: Genomics England Limited
GS: Genomic Sequencing
HLH: Haemophagocytic Lymphohistiocytosis
HPLC: High Performance Liquid Chromatography
hRB: Heritable Retinoblastoma
HSCT: Hematopoietic Stem-Cell Transplantation
HTA: Health Technology Assessment
ICoNS: International Consortium On Newborn Sequencing
ICU: Intensive Care Unit
IDD: Intellectual Developmental Delay
IEI: Inborn Errors of Immunity
IEM: Inborn Errors of Metabolism
IMD: Inherited Metabolic Disease
IQ: Intelligence Quotient
LRT: Lysine Reduction Therapy
MCADD: Medium Chain Acyl-Coa Dehydrogenase Deficiency
MeSH: Medical Subject Headings
MLPA: Multiplex Ligation Dependent Probe Amplification
mRNA: Messenger Ribonucleic Acid
NBS: Newborn Bloodspot
NGS: Next Generation Sequencing
NESTS: NEwborn Screening with Targeted Sequencing
NHS EED: NHS Economic Evaluation Database
NICU: Neonatal Intensive Care Unit
NSC: National Screening Committee
PCR: Polymerase Chain Reaction
PDE: Pyridoxine Dependent Epilepsy
Pi/D: Oral phosphate and calcitriol (active vitamin D)
PKU: Phenylketonuria
PPIE: Patient And Public Involvement Engagement
QALY: Quality Adjusted Life Year
QUIPS: Quality In Prognostic Studies
RCT: Randomized Controlled Trial
REC: Retinoblastoma Variant Effect Class
ROBIS: Risk Of Bias In Systematic Reviews
SD: Standard Deviation
SGP: Scottish Genomes Partnership
SMA: Spinal Muscular Atrophy
SNV: Single Nucleotide Variant
SSCP: Single Strand Conformation Polymorphism
VUS: Variant of Unknown Significance
WES: Whole Exome Sequencing
WGS: Whole Genome Sequencing
XLHR: X-Linked Hypophosphataemic Rickets

## Notes

### Clinical Protocols

https://www.crd.york.ac.uk/PROSPERO/display_record.php?RecordID=475529

