## Supplement 1 for "Evaluating whole genome sequencing for rare diseases in newborn screening: evidence synthesis from a series of systematic reviews"

### Characteristics of the 5 conditions under review

#### Pyridoxine-dependent epilepsy

##### *Background*

Pyridoxine-dependent epilepsy (PDE or PDE-ALDH7A1, henceforth PDE, OMIM 266100) is a rare inherited form of epilepsy, mostly caused by mutation in the ALDH7AI gene. Other forms not considered here include PDE-PNPO and PDE-PLPHP, which are caused by mutations in the PNPO and PLPHP genes, respectively.[1] The ALDH7A1 gene is responsible for production of the enzyme alpha aminoadipic semialdehyde (α-AASA) dehydrogenase, which is involved in the breakdown of lysine in the brain. Deficiency of α-AASA dehydrogenase results in the accumulation of metabolites including piperideine-6-carboxylate (P6C), which in turn inactivates pyridoxal 5'-phosphate (PLP), the active form of pyridoxine. PLP depletion is thought to contribute to the epileptic features observed in PDE and treatment with large daily doses of pyridoxine (vitamin B6) leads to adequate seizure control for most patients. Neurotoxic metabolite accumulation associated with α-AASA dehydrogenase deficiency has also been purported to contribute to intellectual disability and developmental delay, which occurs in as much as 75% of cases of PDE.

##### *Natural history*

Classic PDE usually presents during the neonatal period with prolonged seizures that are difficult to control with anti-seizure medication; in 75% of cases seizures may occur within the first few hours of life.[2] These seizures last several minutes and involve loss of consciousness, spasticity and convulsions. If untreated, periods of encephalopathy are common (irritability, crying, fluctuating tone, poor feeding). In some cases, affected individuals do not experience seizures until they are one to three years old (late-onset PDE). Intellectual disability and developmental delay are often present (around 75% of cases), especially in those with classic PDE.[1, 3, 4] Due to the symptoms of PDE being similar to other, more common neonatal disorders, the condition can be missed, and this can led to death.[5]

##### *Genetics and epidemiology*

PDE is an autosomal recessive condition usually caused by a homozygous or compound heterozygous mutation in the ALDH7A1 gene on chromosome 5q23.[6] Over 165 pathogenic variants of ALDH7A1 have been identified and the vast majority are biallelic.[7] A recent review reported prevalences of PDE based on clinical diagnosis,  to, as low as 1:396,000 in the Netherlands, and 1:783,000 in the United Kingdom.[8] However, these early studies only included patients who responded to a pyridoxine trial and are therefore likely to have under-estimated prevalence; more recent studies estimate the prevalence of PDE as 1:64,352. Children born to couples who are both carriers of the mutation have a 25% risk of developing PDE.[9]

##### *Screening and Diagnosis*

PDE should be considered when investigating intractable seizures in patients aged three and under. Historically, diagnosis was ascertained by a positive clinical response to pyridoxine treatment,[4, 10] however, techniques to diagnose PDE now include measurement of biomarkers in the urine, blood, or cerebral spinal fluid (i.e. $\Delta^1$-piperideine-6-carboxylate (or Δ1-P6C) α-AASA)[11] and genetic testing for pathogenic (or likely pathogenic) variants in ALDH7A1.[7] A 2020 consensus guideline recommended the use of α-AASA or Δ1-P6C as diagnostic biomarkers of PDE, either alone or in combination with "other biomarkers" but did not make specific recommendations about levels that should be considered diagnostic.[7] A 2007 series of 11 patients with definite, probable or possible PDE reported the control values used for both α-AASA and Δ1-P6C; α-AASA was above control values in 10 of 11 patients, while PA in plasma was elevated in all patients with elevated α-AASA levels.[11] Although MRI abnormalities have been reported,[11, 12] there are currently no imaging or electroencephalogram features that can confirm a diagnosis of PDE.[4]

PDE is not currently screened for in newborn programmes in the UK.

##### *Treatment*

There is no cure for PDE. The mainstay of treatment has traditionally been daily, high doses of pyridoxine for seizure control. However, outcomes for patients are often still poor, even with early diagnosis. Adjunct lysine reduction therapies (LRT) (e.g., lysine restricted diet and arginine supplementation) aim to reduce the accumulation of metabolites thought to contribute to intellectual disability and developmental delay.[13, 14] The combination of vitamin B6 and LRT is known as 'Triple Therapy'.[15] Small observational studies have suggested possible improvements in clinical outcomes from triple therapy. However results are likely to be confounded by earlier age on initiation of treatment.[13],[16, 17]

### Hereditary retinoblastoma

#### *Background*

Hereditary retinoblastoma (RB, OMIM 180200) is a rare embryonic malignant neoplasm of the eye. RB is caused by biallelic mutations to the human retinoblastoma susceptibility (RB1) gene on chromosome 13q14 that codes for the RB protein.[18] Mutations to the RB1 gene prevent production of functional protein leading to uncontrollable growth of cells in the retina, resulting in tumours.[19] It is estimated that 40 to 45% of all RBs are hereditary (resulting from a germline mutation and a second mutation occurring in retinal cell precursors); the remaining cases are somatic (caused by two allele mutations at the cellular level).[20] Hereditary RB is usually bilateral (80% of cases), 5% are trilateral (including a pineal/midline neuroectodermal tumour) and 15% are unilateral.[21] However, it is estimated only 10% of children with RB have a family history of the disease.[22]The remaining cases, where there is no family history of the disease, are known as sporadic. Sporadic cases can either be hereditary or somatic.[23]

#### *Natural history*

Hereditary RB usually occurs at an average of 15 months of age and may be picked up by targeted ocular screening before any symptoms develop if there is family history of the disease.[24, 25] The most common first symptom of RB is leukocoria or visible whiteness of the pupil, which may be noticed in photographs taken using flash photography. Leukocoria was cited as primary reason for treatment referral in 62.8% of cases in a global cohort of 4351 patients, followed by strabismus (squint) in 10.2% and proptosis (protruding eye(s)) in 7.4%.[24] Other common symptoms include glaucoma and hypopyon (presence of pus), and if the tumour is large, the eye may become painful and inflamed.[26] High-risk features on presentation (e.g. optic nerve invasion) are more common with increasing age and are associated with poorer outcome.[27] If RB is left untreated, blindness can occur and metastases will most likely develop.[28, 29] In 5% of cases, heritable RB is associated with a midline brain tumour.[30]

RB is considered to be largely curable, with 10-year survival rates from non-neoplastic causes no lower than the general population.[31] Survival rates have been shown to vary globally according to national income levels, with 3-year survival ranging from 99·5% (95% CI 98·8–100·0) for children from high income countries to 57·3% (52·1–63·0) for children from low-income countries.[24] Following curative treatment for hereditary RB, survivors have an increased risk of subsequent malignancy (standardised incidence ratio of 11.9, 95% CI 10.4, 13.5), with considerably higher risks for sarcoma of the bone or soft tissue.[32]

#### *Genetics and epidemiology*

Hereditary RB is caused by a heterozygous germline mutation on one allele and a second, somatic mutation on the other allele of the RB1 gene on chromosome 13q14.[33] Over 900 mutations in RB1 have been reported; research is ongoing to investigate whether the type of mutation (e.g., nonsense, deletion, frameshift, or splicing mutations) is associated with the clinical features of RB.[34, 35]Most mutations are of very high penetrance and expressivity.[32] In 10-20% of cases, the mutation is inherited from a parent who also has hereditary RB.[36] Children with one parent who has heritable RB have a 25% risk of developing RB (50% risk of inheritance and 90% penetrance).[34, 37] There are two known RB1 allele mutations which show a parent-of-origin effect; c.607+1G→T substitution and c.1981C→T (p.Arg661Trp) missense mutation.[38] Otherwise, the mutation in the affected child is new.[36]

Around 44 cases of RB are diagnosed every year in the UK, and 40% of these are of the hereditary form.[39],[40] There is a slightly higher incidence of bilateral RB amongst males.[39] In around 5% of people with RB, the part of chromosome 13q that contains the RB1 gene is missing. This rare form of RB is classed as hereditary and is known as chromosome 13q deletion.[21]

##### *Screening and Diagnosis*

Many countries offer targeted ophthalmological screening for RB in children born into families where there is a history of RB.[25] Family members may have already undergone genetic counselling and testing, particularly where a germline RB1 mutation was identified in the original proband, and genetic testing is likely to be offered to offspring of those identified as having a familial RB1 mutation.[41] Subsequent ophthalmological screening of those identified as at risk of developing RB is usually based around red reflex testing, begins after birth and, may be repeated every few months until the child is 5 years old.[42] Children with dim or absent red reflex are referred to a specialist ophthalmology service for eye examination under general anaesthetic.[21] Unlike other cancers, RBs can be diagnosed by their appearance so a biopsy is usually not necessary. After RB is diagnosed, other tests are conducted to stage the tumour. These can include an ultrasound or MRI scans, a lumbar puncture, a bone marrow sample or a bone scan.[43] For those with bilateral or multifocal RB (hereditary), alterations in the RB1 gene can usually be detected in blood samples. For children with unilateral RB, genetic testing can clarify whether the disease is hereditary or somatic.[21]

##### *Treatment*

Management of RB is complex and treatment regimens must be tailored dependent on the circumstances, including factors such as tumour stage, number of foci, localization and size of the tumour(s).[44, 45] Treatment options include enucleation, cryotherapy, laser treatment, chemotherapy or radiotherapy.[46] Small, localised tumours can be successfully treated with laser treatment or cryotherapy, however chemotherapy is often needed for more advanced cases or when RB is present in both eyes, as is often the case for the hereditary form of the disease. Chemotherapy has been shown to lead to tumour control and avoidance of enucleation (eye removal) or external beam radiotherapy in over 90% of patients with no evidence of seeding (tumour invasion) into the subretinal space or vitreous cavity prior to commencing treatment.[45, 47, 48] In a series of 869 eyes (540 patients) undergoing chemotherapy for RB, a total of 161 (19%) underwent enucleation at a mean of 15 months (range 1 to 191 months).[45]

Sometimes, enucleation must be performed.[46] According to the NHS, there is a high chance the child will lose some or all vision in the affected eye therefore successful treatment is highly dependent on identifying RB early. A UK retrospective case study of patients with bilateral retinoblastoma identified visual impairment in 38% (14/44) of children (i.e., Snellen acuity between 20/40 and 20/200 in the better eye) and legal blindness in 19% of children (vision of 20/200 or worse in the better eye) following chemotherapy.[49]

#### Medium Chain Acyl-CoA Dehydrogenase Deficiency (MCADD)

##### *Background*

Medium chain Acyl-CoA Dehydrogenase Deficiency (MCADD, OMIM 201450) is an inherited metabolic disease in which medium-chain fatty acids cannot be oxidised. This leads to an accumulation of fatty acids in the body and to a disruption in energy production mechanisms, particularly that of ketone synthesis. People with MCADD thus cannot mobilise energy stores in periods of increased metabolic demand (i.e., fasting, intense exercise, illness, etc.) which sends them into a state of metabolic crisis.

A deficiency of MCAD can result in a build-up of acylcarnitines (esters that bind to fat molecules to transport them into mitochondria) in blood, which may be observed in patients with biochemical testing.[50]

##### *Natural history*

MCADD typically presents in the first 2 years of life with hypoglycaemic episodes concurrent with illness or increased periods of fasting (i.e., with the reduction of night-time feeds). Severe hypoglycaemic episodes may lead to seizure, and metabolic decompensation characterised by vomiting, coma and even death. MCADD accounts for around 1% of Sudden Infant Death Syndrome / Sudden Unexpected Death in Infants though the inclusion of MCADD in screening programmes has greatly reduced this.[51, 52] Mortality following a metabolic crisis episode in undiagnosed people with MCADD is around 20%.[53-55]

##### *Genetics and epidemiology*

MCADD is an autosomal recessive condition affecting the ACADM gene. In the majority of cases (>80%), the condition is caused by a homozygous 985A→G mutation.[56-59] It is more prevalent in Caucasian populations and has a prevalence of 1 in 10,000 in the UK.[60] Other mutations are more common in other ethnic groups (e.g., Japan).[61] In the UK, the prevalence of homozygous 985A→G carriers is estimated at 6.2 per 100,000.[62]

##### *Screening and Diagnosis*

In the UK, MCADD has been part of the standard blood spot screening battery since 2009.[ref] The current testing and diagnostic pathways are presented in Figure 1. The current approach to screening for MCADD consists of measuring the concentration of acylcarnitines (primarily C8 and C10) in the blood. A raised level of these markers (C8 > 0.5 µmol/L and C8:C10 ≥ 1 [62, 63]) is suggestive of an incomplete breakdown of medium-chain fatty acids due to MCADD. In the current screening and diagnosis pathway, positive metabolite findings trigger genetic testing looking for the common 985A→G mutation in the first instance and followed by an extended mutation screening for patients who do not have a homozygous 985A→G mutation. The sensitivity of the screening programme for MCADD in England is estimated to be 94%.[62]

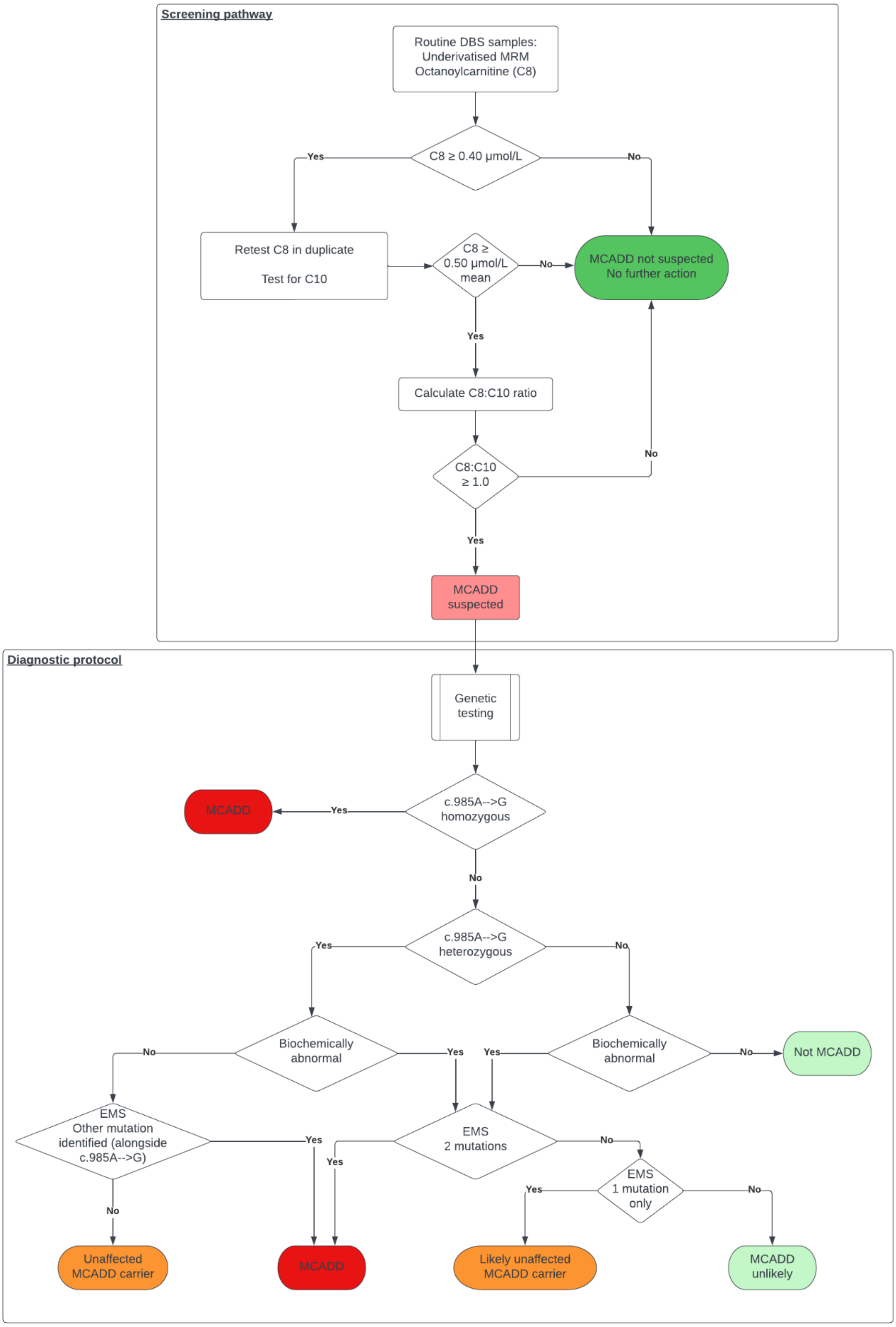


**Figure 1.1 Screening and Diagnostic pathways for MCADD (adapted from [64])**

Key: C8: Octanoylcarnitine; C10: Decanoylcarnitine; DBS: Dried Blood Spot; EMS: Extended Mutation Screening; MCADD: Medium Chain Acyl-CoA Dehydrogenase Deficiency; MRM: Multiple Reaction Monitoring

*Treatment*

There is no cure for MCADD, but it can be effectively managed through diet. MCADD management typically consists of preventing hypoglycaemic episodes through limiting fasting periods.[65] Normal diet composition is normally acceptable, with the exception of coconut and coconut-derived products.[66] Diets should include sufficient complex carbohydrate intake, especially before fasting periods (i.e., night-time).

Acute illness increases risk of metabolic crisis. Emergency plans include prevention or treatment of hypoglycaemia through intake of fast carbohydrate by mouth or intravenous glucose infusion.[67]

#### X-linked hypophosphataemic rickets

##### *Background*

X-linked hypophosphataemic rickets (XLHR, OMIM 307800), also known as X-linked hypophosphataemia, X-linked rickets, or vitamin-D resistant rickets, is a hereditary disorder of phosphate processing that causes a form of rickets. This is primarily characterized by osteomalacia (soft bones) and associated complications (bone deformity, bone and joint pain, dental problems). XLHR was first reported in 1957 and mutation to the PHEX gene identified as a cause in 1995.[68, 69]

The pathophysiology of XLHR is not fully understood but is known to primarily involve an increase in the FGF23 hormone. This triggers changes in both the kidneys and the parathyroid glands and ultimately results in increased renal phosphate wasting.[70]

##### *Natural history*

Features of XLHR can be broadly divided into acute and chronic signs of hypophosphataemia. Signs of acute hypophosphataemia include muscle weakness, respiratory and cardiac insufficiency, neurological dysfunction, and blood disorder. Chronic signs include bone deformity, dental abscesses, stunted growth and bone and joint pain.[71] Clinical symptoms usually appear in the first two years of life, becoming more obvious with delayed walking or slowing down of growth/ exacerbation of leg bowing once toddlers become weight-bearing.[72]

##### *Genetics and epidemiology*

XLHR is caused by a mutation in the PHEX gene. Although most patients with XLHR have inherited a pathogenic variant from a parent, around 20% present with de novo mutations, meaning that pathogenic changes to the gene have occurred spontaneously.[73] XLHR is the most common type of hereditary rickets, and penetrance is generally assumed to be 100% with no sex differences in penetrance.[71, 74] Prevalence estimates range from 1.7:100,000 children to 4.8:100,000 children and adults.[75]

##### *Screening and diagnosis*

X-linked hypophosphataemia is not currently screened for in newborn programmes in the UK. It is generally diagnosed in early childhood (usually before the 2nd birthday) through a combination of clinical features, biochemical characteristics, and radiological signs. Table **Error! No text of specified style in document.**.1 summarises some key features of X-linked hypophosphataemic rickets.

**Table** Error! No text of specified style in document.**.1. Key clinical, biochemical and radiological features of X-linked hypophosphataemic rickets**

| Clinical features | Biochemical characteristics | Radiological signs |
|---|---|---|
| - Short stature<br>- Leg bowing/knock-knees<br>- Delayed walking/abnormal gait<br>- Dental abscesses | - Low serum phosphate*<br>- High urine phosphate | - Widening/cupping of metaphyses<br>- Rachitic rosary of ribs<br>- Sometimes “green stick” fractures |

*0-15 days: <5.6 mg/dL; 15-365 days: 4.8 mg/dL; 1-4 years: <4.3 mg/dL; 5-12 years: <4.1 mg/dL; 13-15 years: <3.2(female) / 3.5(male) mg/dL; 16-18 years: <2.9 mg/dL [76, 77]

##### *Treatment*

There's no cure for X-linked hypophosphataemia, but it can be managed, with a goal of normalising serum phosphate concentration. This consists of oral supplementation of phosphorus (20 to 40 mg/kg/day) and calcitriol (active vitamin D, 20 to 30 ng/kg/day) multiple times a day.[70] This management approach is not always effective, but evidence suggests that earlier intervention is beneficial.[78] This strategy should include regular follow up to limit the risks of complications associated with treatment, which are commonplace. These include hypercalcaemia, hypercalciuria, kidney stones, nephrocalcinosis, impaired renal function and can lead to chronic kidney disease.[79]

An alternative approach using burosumab injections was approved in the UK in 2018.[80] Burosumab is an antibody against FGF23, which leads to an increase in renal phosphate reuptake/reduced wasting, increase in serum calcitriol, and increased gastrointestinal absorption of phosphate.[70]

### Familial haemophagocytic lymphohistiocytosis

#### *Background*

Familial haemophagocytic lymphohistiocytosis (fHLH) is an immunological disorder characterized by abnormal immune activation in which overactive macrophages target red blood cells. In terms of pathogenesis, fHLH involves a dysfunction of the cytotoxic perforin/granzyme pathway used by lymphocytes to target infected cells and downregulate the immune response as needed. The inability to neutralise overactive macrophages leads to an escalation of the immune response, including abnormal targeting of red blood cells and cytokine storms leading to organ damage.

#### *Natural history*

In fHLH, key downregulation mechanisms of the immune system are defective. Specifically, T-cells and Natural Killer (NK) cells have defective perforin/granzyme pathway (which is used by lymphocytes to trigger lysis of targeted cells, including overactive macrophages). This leads to a proliferation of lymphocytes and overactive macrophages which attack red blood cells causing anaemia. The cytokine storm associated with the unbridled immune response can lead to fatal multi-organ failure. fHLH usually manifests in infancy, with minor infections triggering an abnormal immune response. The prognosis for fHLH is poor but new treatments are promising.[81]

#### *Genetics and epidemiology*

Different genes are associated with different types of fHLH.[82] fHLH is inherited in an autosomal recessive pattern.[83] Information pertaining to genes and pathophysiology of different types of fHLH are provided in Table **Error! No text of specified style in document.**.2. Here, the focus is on types of haemophagocytic lymphohistiocytosis that specifically involve the malfunction of the perforin/granzyme cytotoxic pathway.

**Table** Error! No text of specified style in document.**.2. Types of haemophagocytic lymphohistiocytosis affecting the perforin/granzyme pathway**

| HLH subtype (OMIM) | Gene involved | Pathophysiology |
|---|---|---|
| fHLH type 2 (OMIM 603553) | PRF1 | Affects perforin (a protein on the lytic granule that lets the granzymes into the target cells) |
| fHLH type 3 (OMIM 608898) | UNC13D | Affects munc13-4 (a protein that is involved in the fusion of the lytic granule and the target cell membrane) |
| fHLH type 4 (OMIM 603552) | STX11 | Affects syntaxin-11 (a protein involved in the docking of the lytic granule to the target cell) |
| fHLH type 5 (OMIM 613101) | STXBP2 | Affects munc18-2 (a protein involved in the fusion of the lytic granule and the cell membrane) |

There is little information about the prevalence of these disorders though they are rare. [84] The prevalence in Sweden is estimated to be 1.8:100k and 1:100k in Texas.[85, 86]

#### *Screening and diagnosis*

Familial HLH usually presents in infancy with symptoms including fever, enlarged spleen and liver, lymphadenopathy, and an array of neurological symptoms. Complications of fHLH can include anaemia, haemorrhage and secondary infection linked to decreased red blood cell, platelet, and neutrophil counts. These non-specific signs and symptoms associated with the rarity of fHLH can render diagnosis difficult. Classical fHLH laboratory findings can help with diagnosis, including high ferritin, abnormal cell counts, disturbed liver function markers.[83] Table Error**! No text of specified style in document.**.3 summarises the diagnosis criteria for fHLH.

**Table** Error! No text of specified style in document.**.3 Diagnostic criteria for familial haemophagocytic lymphohistiocytosis (adapted from [87])**

| Diagnostic can be established if either A or B is fulfilled: | |
|---|---|
| A. Genetic variation consistent with fHLH | B. Any 5 of the following:<br>- Fever >38.5°C<br>- Splenomegaly<br>- Abnormal cell counts<br>o Haemoglobin <9g/dL (<100g/dL for infants 4 weeks and under)<br>o Platelets <100x10$^9$/L<br>o Neutrophils <1.0x10$^9$/L<br>- High fasting triglycerides >3.0 mmol/L (>265mg/dL) and/or low fibrinogen (≤1.5g/L)<br>- Haemophagocytosis in bone marrow, spleen, liver, lymph nodes, or other tissue<br>- Decreased NK cell activity<br>- Ferritin ≥ 500µL<br>- High soluble IL-2 receptor ≥2,400U/mL |

##### *Treatment*

Treatment of acute disease consists of supressing the immune response, through immunotherapy (i.e., corticosteroids) and chemotherapy but an important proportion of patients do not respond to these approaches. The only curative treatment currently available is allogeneic haematopoietic stem cell transplantation.[88]

##### *References*

1. Coughlin CR, II, Gospe SM, Jr. Pyridoxine-dependent epilepsy: Current perspectives and questions for future research. *Annals of the Child Neurology Society* 2023;**1**(1):24-37. http://dx.doi.org/10.1002/cns3.20016

2. de Rooy RLP, Halbertsma FJ, Struijs EA, van Spronsen FJ, Lunsing RJ, Schippers HM*, et al.* Pyridoxine dependent epilepsy: Is late onset a predictor for favorable outcome? *Eur J Paediatr Neurol* 2018;**22**(4):662-6. http://dx.doi.org/10.1016/j.ejpn.2018.03.009

3. Medline Plus. *Pyridoxine-dependent epilepsy*. Bethesda, MD National Library of Medicine; 2013. URL: https://medlineplus.gov/genetics/condition/pyridoxine-dependent-epilepsy/ (Accessed 17 June 2023).

4. Gospe SM. Gene Reviews: Pyridoxine-Dependent Epilepsy – ALDH7A1. *Gene Reviews: Pyridoxine-Dependent Epilepsy - ALDH7A1* 2022.

22. Trent RJ. 4 - COMPLEX GENETIC TRAITS. In: Trent RJ, editor. *Molecular Medicine (Third Edition)*Burlington: Academic Press; 2005:77-118. http://dx.doi.org/https://doi.org/10.1016/B978-012699057-7/50004-7
23. Yun J, Li Y, Xu CT, Pan BR. Epidemiology and Rb1 gene of retinoblastoma. *Int J Ophthalmol* 2011;**4**(1):103-9. http://dx.doi.org/10.3980/j.issn.2222-3959.2011.01.24
24. Global Retinoblastoma Study G, Fabian ID, Abdallah E, Abdullahi SU, Abdulqader RA, Adamou Boubacar S*, et al.* Global retinoblastoma presentation and analysis by national income level. *JAMA Oncol* 2020;**6**(5):685. http://dx.doi.org/10.1001/jamaoncol.2019.6716
25. Vempuluru VS, Kaliki S. Screening for retinoblastoma: A systematic review of current strategies. *Asia Pac J Ophthalmol (Phila)* 2021;**10**(2):192-9. http://dx.doi.org/10.1097/apo.0000000000000378
26. GARD. *Retinoblastoma*. Gaithersburg, MD Genetic and Rare Diseases

Information Center; 2024. URL: https://rarediseases.info.nih.gov/diseases/7563/retinoblastoma (Accessed 17 November 2023).
27. Kaliki S, Maniar A, Patel A, Palkonda VAR, Mohamed A. Clinical presentation and outcome of retinoblastoma based on age at presentation: a review of 1450 children. *Int Ophthalmol* 2020;**40**(1):99-107. http://dx.doi.org/10.1007/s10792-019-01155-z
28. Institute NE. *Treatment for retinoblastoma*. Bethesda, MD National Eye Institute; 2022. URL: https://www.nei.nih.gov/learn-about-eye-health/eye-conditions-and-diseases/retinoblastoma/treatment-retinoblastoma (Accessed 16 December 2023).
29. Fabian ID, Reddy A, Sagoo MS. Classification and staging of retinoblastoma. *Community Eye Health* 2018;**31**(101):11-3. https://www.ncbi.nlm.nih.gov/pubmed/29915461

https://www.ncbi.nlm.nih.gov/pmc/articles/PMC5998397
30. Soliman SE, ElManhaly M, Dimaras H. Knowledge of genetics in familial retinoblastoma. *Ophthalmic genetics* 2017;**38**(3):226-32. http://dx.doi.org/https://dx.doi.org/10.1080/13816810.2016.1195846
31. Yu CL, Tucker MA, Abramson DH, Furukawa K, Seddon JM, Stovall M*, et al.* Cause-specific mortality in long-term survivors of retinoblastoma. *J Natl Cancer Inst* 2009;**101**(8):581-91. http://dx.doi.org/10.1093/jnci/djp046
32. Kleinerman RA, Schonfeld SJ, Tucker MA. Sarcomas in hereditary retinoblastoma. *Clin Sarcoma Res* 2012;**2**(1):15. http://dx.doi.org/10.1186/2045-3329-2-15
33. Kelly J. *# 180200 RETINOBLASTOMA; RB1*. Baltimore, MD: OMIM; 2019. URL: https://www.omim.org/entry/180200

(Accessed 20th January 2024).
34. Valverde JR, Alonso J, Palacios I, Pestaña Á. RB1 gene mutation up-date, a meta-analysis based on 932 reported mutations available in a searchable database. *BMC Genet* 2005;**6**(1):53. http://dx.doi.org/10.1186/1471-2156-6-53
35. Mehyar M, Mosallam M, Tbakhi A, Saab A, Sultan I, Deebajah R*, et al.* Impact of RB1 gene mutation type in retinoblastoma patients on clinical presentation and management outcome. *Hematol Oncol Stem Cell Ther* 2020;**13**(3):152-9. http://dx.doi.org/10.1016/j.hemonc.2020.02.006
36. St. Jude Children's Research Hospital. *Hereditary retinoblastoma*. Memphis, TN St. Jude Children's Research Hospital; 2020. URL: https://www.stjude.org/disease/hereditary-retinoblastoma.html (Accessed 03 November 2023).
37. AlAli A, Kletke S, Gallie B, Lam W-C. Retinoblastoma for Pediatric Ophthalmologists. *The Asia-Pacific Journal of Ophthalmology* 2018;**7**(3):160-8. http://dx.doi.org/10.22608/apo.201870
38. Soliman SE, Dimaras H, Khetan V, Gardiner JA, Chan HSL, Heon E*, et al.* Prenatal versus Postnatal Screening for Familial Retinoblastoma. *Ophthalmology* 2016;**123**(12):2610-7. http://dx.doi.org/https://dx.doi.org/10.1016/j.ophtha.2016.08.027

39. Children with Cancer UK. *Retinoblastoma*. London: Children with Cancer UK; 2024. URL: https://www.childrenwithcancer.org.uk/childhood-cancer-info/cancer-types/retinoblastoma/ (Accessed 16 February 2024).
40. MacCarthy A, Birch JM, Draper GJ, Hungerford JL, Kingston JE, Kroll ME*, et al.* Retinoblastoma in great Britain 1963-2002. *Br J Ophthalmol* 2009;**93**(1):33-7. http://dx.doi.org/10.1136/bjo.2008.139618
41. Canadian Retinoblastoma Society. Canadian Retinoblastoma Society. National Retinoblastoma Strategy Canadian Guidelines for Care: Stratégie thérapeutique du rétinoblastome guide clinique canadien. *Can J Ophthalmol* 2009;**44**:S9-S47. http://dx.doi.org/http://dx.doi.org/10.3129/i09-194
42. Shafiq A. Seeing red in young children: the importance of the red reflex. *Br J Gen Pract* 2015;**65**(633):209-10. http://dx.doi.org/10.3399/bjgp15x684625
43. Inform N. *Retinoblastoma: Children*. Edinburgh: NHS Scotland; 2023. URL: https://www.nhsinform.scot/illnesses-and-conditions/cancer/cancer-types-in-children/retinoblastoma-children (Accessed 06 November 2023).
44. Lohmann DR, Gallie BL. Gene Reviews: Retinoblastoma. *Gene Reviews: Retinoblastoma University* 2018.
45. Shields C, Ancona-Lezama D, Dalvin L. Modern treatment of retinoblastoma: A 2020 review. *Indian J Ophthalmol* 2020;**68**(11):2356. http://dx.doi.org/10.4103/ijo.ijo_721_20
46. NHS. *Treatment for retinoblastoma*. London: National Health Service; 2023. URL: https://www.nhs.uk/conditions/retinoblastoma/treatment/ (Accessed 18 February 2024).
47. Shields CL, Mashayekhi A, Au AK, Czyz C, Leahey A, Meadows AT*, et al.* The international classification of retinoblastoma predicts chemoreduction success. *Ophthalmology* 2006;**113**(12):2276-80. http://dx.doi.org/10.1016/j.ophtha.2006.06.018
48. Bartuma K, Pal N, Kosek S, Holm S, All-Ericsson C. A 10-year experience of outcome in chemotherapy-treated hereditary retinoblastoma. *Acta Ophthalmol* 2014;**92**(5):404-11. http://dx.doi.org/10.1111/aos.12282
49. Stacey AW, Clarke B, Moraitis C, Fabian ID, Smith V, Sagoo MS*, et al.* The incidence of binocular visual impairment and blindness in children with bilateral retinoblastoma. *Ocul Oncol Pathol* 2019;**5**(1):1-7. http://dx.doi.org/10.1159/000489313
50. Van Hove JL, Zhang W, Kahler SG, Roe CR, Chen YT, Terada N*, et al.* Medium-chain acyl-CoA dehydrogenase (MCAD) deficiency: diagnosis by acylcarnitine analysis in blood. *Am J Hum Genet* 1993;**52**(5):958-66. https://www.ncbi.nlm.nih.gov/pubmed/8488845

https://www.ncbi.nlm.nih.gov/pmc/articles/PMC1682046
51. Marsden D, Bedrosian CL, Vockley J. Impact of newborn screening on the reported incidence and clinical outcomes associated with medium- and long-chain fatty acid oxidation disorders. *Genet Med* 2021;**23**(5):816-29. http://dx.doi.org/10.1038/s41436-020-01070-0
52. McConkie-Rosell A, Iafolla AK. Medium-chain acyl CoA dehydrogenase deficiency: Its relationship to SIDS and the impact on genetic counseling. *J Genet Couns* 1993;**2**(1):17-27. http://dx.doi.org/10.1007/bf00962557
53. Iafolla AK, Thompson RJ, Jr., Roe CR. Medium–chain acyl-coenzyme A dehydrogenase deficiency: Clinical course in 120 affected children. *J Pediatr* 1994;**124**(3):409-15. http://dx.doi.org/10.1016/s0022-3476(94)70363-9
54. Touma EH, Charpentier C. Medium chain acyl-CoA dehydrogenase deficiency. *Arch Dis Child* 1992;**67**(1):142-5. http://dx.doi.org/10.1136/adc.67.1.142
55. Wilcken B, Carpenter KH, Hammond J. Neonatal symptoms in medium chain acyl coenzyme A dehydrogenase deficiency. *Archives of disease in childhood* 1993;**69**(3 Spec No):292-4. http://ovidsp.ovid.com/ovidweb.cgi?T=JS&PAGE=reference&D=med3&NEWS=N&AN=8215568
56. Gregersen N, Blakemore AI, Winter V, Andresen B, Kolvraa S, Bolund L*, et al.* Specific diagnosis of medium-chain acyl-CoA dehydrogenase (MCAD) deficiency in dried blood spots by a polymerase chain reaction (PCR) assay detecting a point-mutation (G985) in the MCAD gene. *Clinica*

*chimica acta; international journal of clinical chemistry* 1991;**203**(1):23-34. http://ovidsp.ovid.com/ovidweb.cgi?T=JS&PAGE=reference&D=med3&NEWS=N&AN=1769118
57. Yokota I, Coates PM, Hale DE, Rinaldo P, Tanaka K. Molecular survey of a prevalent mutation, 985A-to-G transition, and identification of five infrequent mutations in the medium-chain Acyl-CoA dehydrogenase (MCAD) gene in 55 patients with MCAD deficiency. *Am J Hum Genet* 1991;**49**(6):1280-91. https://www.ncbi.nlm.nih.gov/pubmed/1684086

https://www.ncbi.nlm.nih.gov/pmc/articles/PMC1686456
58. Pollitt RJ, Leonard JV. Prospective surveillance study of medium chain acyl-CoA dehydrogenase deficiency in the UK. *Arch Dis Child* 1998;**79**(2):116-9. http://dx.doi.org/10.1136/adc.79.2.116
59. Derks TGJ, Reijngoud D-J, Waterham HR, Gerver W-JM, van den Berg MP, Sauer PJJ*, et al.* The natural history of medium-chain acyl CoA dehydrogenase deficiency in the Netherlands: Clinical presentation and outcome. *J Pediatr* 2006;**148**(5):665-70.e3. http://dx.doi.org/10.1016/j.jpeds.2005.12.028
60. Oerton J, Khalid JM, Besley G, Dalton RN, Downing M, Green A*, et al.* Newborn screening for medium chain acyl-CoA dehydrogenase deficiency in England: Prevalence, predictive value and test validity based on 1.5 million screened babies. *J Med Screen* 2011;**18**(4):173-81. http://dx.doi.org/10.1258/jms.2011.011086
61. Tajima G, Hara K, Tsumura M, Kagawa R, Okada S, Sakura N*, et al.* Screening of MCAD deficiency in Japan: 16years' experience of enzymatic and genetic evaluation. *Mol Genet Metab* 2016;**119**(4):322-8. http://dx.doi.org/10.1016/j.ymgme.2016.10.007
62. Leal J, Wordsworth S, Oerton J, Khalid JM, Dezateux C. Synthesis framework estimating prevalence of MCADD and sensitivity of newborn screening programme in the absence of direct evidence. *J Clin Epidemiol* 2014;**67**(10):1131-8. http://dx.doi.org/10.1016/j.jclinepi.2014.05.011
63. Downing M, Pollitt R. Newborn bloodspot screening in the UK – past, present and future. *Ann Clin Biochem* 2008;**45**(1):11-7. http://dx.doi.org/10.1258/acb.2007.007127
64. Public Health England. *NHS Newborn Blood Spot Screening Programme A laboratory guide to newborn blood spot screening for inherited metabolic diseases*. London: Public Health England; 2017. URL: https://assets.publishing.service.gov.uk/media/5a82314fed915d74e623648e/IMD_laboratory_handbook_2017.pdf (Accessed 15 March 2023).

72. Linglart A, Biosse-Duplan M, Briot K, Chaussain C, Esterle L, Guillaume-Czitrom S*, et al.* Therapeutic management of hypophosphatemic rickets from infancy to adulthood. *Endocr Connect* 2014;**3**(1):R13-R30. http://dx.doi.org/10.1530/ec-13-0103
73. Dixon PH, Christie PT, Wooding C, Trump D, Grieff M, Holm I*, et al.* Mutational analysis of PHEX gene in X-linked Hypophosphatemia1. *J Clin Endocrinol Metab* 1998;**83**(10):3615-23. http://dx.doi.org/10.1210/jcem.83.10.5180
74. Ruppe MD. *Gene Reviews: X-Linked Hypophosphatemia*. Seattle (WA: GeneReviews; 2017.
75. Haffner D, Emma F, Eastwood DM, Duplan MB, Bacchetta J, Schnabel D*, et al.* Clinical practice recommendations for the diagnosis and management of X-linked hypophosphataemia. *Nat Rev Nephrol* 2019;**15**(7):435-55. http://dx.doi.org/10.1038/s41581-019-0152-5
76. Adeli K, Higgins V, Trajcevski K, White-Al Habeeb N. The Canadian laboratory initiative on pediatric reference intervals: A CALIPER white paper. *Crit Rev Clin Lab Sci* 2017;**54**(6):358-413. http://dx.doi.org/10.1080/10408363.2017.1379945
77. Colantonio DA, Kyriakopoulou L, Chan MK, Daly CH, Brinc D, Venner AA*, et al.* Closing the gaps in pediatric laboratory reference intervals: A CALIPER database of 40 biochemical markers in a healthy and multiethnic population of children. *Clin Chem* 2012;**58**(5):854-68. http://dx.doi.org/10.1373/clinchem.2011.177741
78. Makitie O, Doria A, Kooh SW, Cole WG, Daneman A, Sochett E. Early treatment improves growth and biochemical and radiographic outcome in X-linked hypophosphatemic rickets. *The Journal of clinical endocrinology and metabolism* 2003;**88**(8):3591-7. http://ovidsp.ovid.com/ovidweb.cgi?T=JS&PAGE=reference&D=med5&NEWS=N&AN=12915641
79. Carpenter TO, Imel EA, Holm IA, Jan de Beur SM, Insogna KL. A clinician's guide to X-linked hypophosphatemia. *J Bone Miner Res* 2011;**26**(7):1381-8. http://dx.doi.org/10.1002/jbmr.340
80. NICE. *Burosumab for treating X-linked hypophosphataemia in children and young people*. London: National Institute for Health and Care Excellence; 2018. URL: https://www.nice.org.uk/guidance/hst8 (Accessed 18 March 2024).
81. Bayram C, Tahtakesen TN, Arslantaş E, Yilmaz E, Özdemir GN, Pasli Uysalol E*, et al.* Prognostic factors and long-term outcomes in 41 children with primary hemophagocytic lymphohistiocytosis: Report of a single-center experience and review of the literature. *J Pediatr Hematol Oncol* 2023;**45**(5):262-6. http://dx.doi.org/10.1097/MPH.0000000000002653
82. Brisse E, Wouters CH, Matthys P. Hemophagocytic lymphohistiocytosis (HLH): A heterogeneous spectrum of cytokine-driven immune disorders. *Cytokine Growth Factor Rev* 2015;**26**(3):263-80. http://dx.doi.org/10.1016/j.cytogfr.2014.10.001
83. Zhang K, Astigarraga I, Bryceson Y, Lehmberg K, Machowicz R, Marsh R. Gene Reviews: Familial Hemophagocytic Lymphohistiocytosis. *Gene Reviews: Familial Hemophagocytic Lymphohistiocytosis University of Washington* 2021.
84. George M. Hemophagocytic lymphohistiocytosis: review of etiologies and management. *J Blood Med* 2014;**5**:69. http://dx.doi.org/10.2147/jbm.s46255
85. Meeths M, Horne A, Sabel M, Bryceson YT, Henter J-I. Incidence and clinical presentation of primary hemophagocytic lymphohistiocytosis in Sweden. *Pediatr Blood Cancer* 2015;**62**(2):346-52. http://dx.doi.org/10.1002/pbc.25308
86. Niece JA, Rogers ZR, Ahmad N, Langevin A-M, McClain KL. Hemophagocytic lymphohistiocytosis in Texas: Observations on ethnicity and race. *Pediatr Blood Cancer* 2010;**54**(3):424-8. http://dx.doi.org/10.1002/pbc.22359
87. Henter J-I, Horne A, Aricó M, Egeler RM, Filipovich AH, Imashuku S*, et al.* HLH-2004: Diagnostic and therapeutic guidelines for hemophagocytic lymphohistiocytosis. *Pediatr Blood Cancer* 2007;**48**(2):124-31. http://dx.doi.org/10.1002/pbc.21039
88. Canna SW, Marsh RA. Pediatric hemophagocytic lymphohistiocytosis. *Blood* 2020;**135**(16):1332-43. http://dx.doi.org/10.1182/blood.2019000936
