## Supplement 2 for "Evaluating whole genome sequencing for rare diseases in newborn screening: evidence synthesis from a series of systematic reviews"

### Excluded studies

Studies excluded at full text sifting stage and reasons for exclusion for the review of five conditions, the review of genomic studies of paediatric cohorts reporting penetrance for pathogenic variants and the review of cost-effectiveness evaluations of WGS and WES.

#### Review of five conditions

##### Studies eligible for Q1 (What is the penetrance and expressivity of different gene variants associated with each condition?), Q2 (What is the prevalence of genetic variants in those with biochemical or biochemical and clinical features of each condition?) and Q3 (What is the diagnostic accuracy of gene sequencing for each condition?)

###### PDE

| **Reference** | **Reason for exclusion (Q2)** |
|---|---|
| Celik H, Ozdemir FMA, Aksoy E, Oztoprak U, Kilic M, Yuksel D. Evaluation of clinical and electroencephalographic findings in patients with early childhood epilepsy and inborn errors of metabolism. Acta Neurol Belg. 2022 Dec;122(6):1575-1581. | Wrong population |
| Coughlin CR, Tseng LA, Bok LA, Hartmann H, Footitt E, Striano P, Tabarki BM, Lunsing RJ, Stockler-Ipsiroglu S, Gordon S, Van Hove JLK, Abdenur JE, Boyer M, Longo N, Andrews A, Janssen MCH, van Wegberg A, Prasad C, Prasad AN, Lamb MM, Wijburg FA, Gospe SM Jr, van Karnebeek C; International PDE Consortium. Association Between Lysine Reduction Therapies and Cognitive Outcomes in Patients With Pyridoxine-Dependent Epilepsy. Neurology. 2022 Dec 5;99(23):e2627-e2636. | Irrelevant outcomes |
| Perry MS. Genetic Testing in Epileptic Encephalopathy: Rosetta Stone or Just an Expensive Rock? Epilepsy Curr. 2016 Jan-Feb;16(1):12-3. doi: 10.5698/1535-7597-16.1.12. | Publication type |
| Toldo I, Bonardi CM, Bettella E, Polli R, Talenti G, Burlina A, Sartori S, Murgia A. Brain malformations associated to Aldh7a1 gene | Case study |

| | |
|---|---|
| mutations: Report of a novel homozygous mutation and literature review. Eur J Paediatr Neurol. 2018 Nov;22(6):1042-1053 | |
| Weckhuysen S, Pearl PL. Gene sleuthing in pyridoxine-dependent epilepsy. Neurology. 2015 Sep 1;85(9):748-9. | Publication type |
| Xue J, Wang J, Gong P, Wu M, Yang W, Jiang S, Wu Y, Jiang Y, Zhang Y, Yuzyuk T, Li H, Yang Z. Simultaneous quantification of alpha-aminoadipic semialdehyde, piperideine-6-carboxylate, pipecolic acid and alpha-aminoadipic acid in pyridoxine-dependent epilepsy. Sci Rep. 2019 Aug 6;9(1):11371. | Wrong population |

#### hRB

| **Reference** | **Reason for exclusion (Q2)** |
|---|---|
| Blanquet V, Gross MS, Turleau C, Sénamaud-Beaufort C, Doz F, Besmond C. Three novel germline mutations in exons 8 and 18 of the retinoblastoma gene. Hum Mol Genet. 1994 Jul;3(7):1185-6 | Test for specific variants or types of mutation |
| Abouzeid H, Schorderet DF, Balmer A, Munier FL. Germline mutations in retinoma patients: relevance to low-penetrance and low-expressivity molecular basis. Mol Vis. 2009;15:771-7 | Wrong population |
| Ahmad NN, Dixon P, Shields JA, Dua HS, Donoso LA. Identification and rapid screening of a DraI RFLP by PCR in the retinoblastoma gene. Br J Ophthalmol. 1994 Feb;78(2):159. doi: 10.1136/bjo.78.2.159-a | Wrong population |
| Berry JL, Xu L, Polski A, Jubran R, Kuhn P, Kim JW, Hicks J. Aqueous Humor Is Superior to Blood as a Liquid Biopsy for Retinoblastoma. Ophthalmology. 2020 Apr;127(4):552-554 | Wrong population |
| Blanquet V, Turleau C, de Grouchy J, Creau-Goldberg N. Physical map around the retinoblastoma gene: possible genomic imprinting suggested by NruI digestion. Genomics. 1991 Jun;10(2):350-5 | Wrong population |
| Bonaïti-Pellié C, Clerget-Darpoux F, Babron MC. Hereditary retinoblastoma: can balanced insertion entirely explain the differences of expressivity among families? Hum Genet. 1990 Dec;86(2):203-8 | Wrong population |
| Chaussade A, Millot G, Wells C, Brisse H, Laé M, Savignoni A, Desjardins L, Dendale R, Doz F, Aerts I, Jimenez I, Cassoux N, Stoppa Lyonnet D, Gauthier Villars M, Houdayer C. Correlation between RB1germline mutations and second primary malignancies in hereditary retinoblastoma patients treated with external beam radiotherapy. Eur J Med Genet. 2019 Mar;62(3):217-223. | Wrong population |
| Cohen JG, Dryja TP, Davis KB, Diller LR, Li FP. RB1 genetic testing as a clinical service: a follow-up study. Med Pediatr Oncol. 2001 Oct;37(4):372-8 | Wrong population |

| | |
|---|---|
| Connolly MJ, Payne RH, Johnson G, Gallie BL, Allderdice PW, Marshall WH, Lawton RD. Familial, EsD-linked, retinoblastoma with reduced penetrance and variable expressivity. Hum Genet. 1983;65(2):122-4 | Wrong population |
| Cowell JK. Genetics of paediatric solid tumours. Br Med Bull. 1994 Jul;50(3):600-23 | Wrong population |
| Dommering CJ, van der Hout AH, Meijers-Heijboer H, Marees T, Moll AC. IVF and retinoblastoma revisited. Fertil Steril. 2012 Jan;97(1):79-81 | Wrong population |
| Gregersen PA, Funding M, Alsner J, Olsen MH, Overgaard J, Staffieri SE, Lou S, Urbak SF. Genetic testing in adult survivors of retinoblastoma in Denmark: A study of the experience and impact of genetic testing many years after initial diagnosis. Eur J Med Genet. 2022 Sep;65(9):104569 | Wrong population |
| Harini R, Ata-ur-Rasheed M, Shanmugam MP, Amali J, Das D, Kumaramanickavel G. Genetic profile of 81 retinoblastoma patients from a referral hospital in southern India. Indian J Ophthalmol. 2001 Mar;49(1):37-42 | Wrong population |
| Hogg A, Onadim Z, Baird PN, Cowell JK. Detection of heterozygous mutations in the RB1 gene in retinoblastoma patients using single-strand conformation polymorphism analysis and polymerase chain reaction sequencing. Oncogene. 1992 Jul;7(7):1445-51 | Wrong population |
| Houdayer C, Gauthier-Villars M, Laugé A, Pagès-Berhouet S, Dehainault C, Caux-Moncoutier V, Karczynski P, Tosi M, Doz F, Desjardins L, Couturier J, Stoppa-Lyonnet D. Comprehensive screening for constitutional RB1 mutations by DHPLC and QMPSF. Hum Mutat. 2004 Feb;23(2):193-202 | Wrong population |
| Mamatha. G, Joseph. B, Shanmugam. M and Kumaramanickavel. G<br><br>CGA codons multiplex PCR in rapid diagnosis of retinoblastoma.<br><br>Indian Journal of Human Genetics 2006 Vol. 12 Issue 1 Pages 34-38 | Wrong population |
| Seminara SB, Dryja TP. Unbiased transmission of mutant alleles at the human retinoblastoma locus. Hum Genet. 1994 Jun;93(6):629-34 | Wrong population |
| Zhang K, Nowak I, Rushlow D, Gallie BL, Lohmann DR. Patterns of missplicing caused by RB1 gene mutations in patients with retinoblastoma and association with phenotypic expression. Hum Mutat. 2008,, 29(4), 475-84 | Wrong population |
| Quiñonez-Silva, G., Dávalos-Salas, M., Recillas-Targa, F. *et al.* “Monoallelic germline methylation and sequence variant in the promoter of the *RB1* gene: a possible constitutive epimutation in hereditary retinoblastoma”. *Clin Epigenet* **8**, 1 (2016). | Test for specific variants or types of mutation |
| Yazici H, Wu HC, Tigli H, Yilmaz EZ, Kebudi R, Santella RM. High levels of global genome methylation in patients with retinoblastoma. Oncol Lett. 2020 Jul;20(1):715-723 | Test for specific variants or types of mutation |

| | |
|---|---|
| Zhang XL, Fu WL, Wang JH, Zhou LX, Lei MP, Hua C. New suspicious gene loci correlated with the generation and development of children's retinoblastoma. *Chin J of Clin Rehab*, 9, 11 (2005), 197-1999 | Test for specific variants or types of mutation |
| Ewens KG, Bhatti TR, Moran KA, Richards-Yutz J, Shields CL, Eagle RC, Ganguly A. Phosphorylation of pRb: mechanism for RB pathway inactivation in MYCN-amplified retinoblastoma. Cancer medicine 2016, 6 (3), 619-630 | Not blood sample test |
| Aerts L, Sastre-Garau X, Savignoni A, Lumbroso-Le Rouic L, Thebaud-Leculee E, Frappaz D, et al. Results of a multicenter prospective study on the postoperative treatment of unilateral retinoblastoma after primary enucleation. J Clin Onco: official journal of the American Society of Clinical Oncology 2013, 31, 11, 1458-63 | No direct sequencing |
| Castela, G., Providência, J., Monteiro, M. *et al.* Characterization of the Portuguese population diagnosed with retinoblastoma. Sci Rep 12, 4378 (2022) | No direct sequencing |
| Chaudhry, Shabana, Onadim, Zerrin, Sagoo, Mandeep, Reddy, Ashwin. The recognition of cavitary Retinoblastoma tumours: implications for management and genetic analysis 2018. Retina 38(4): 782-787 | No direct sequencing |
| Joseph B, Paul PG, Elamparithi A, Roy J, Vidhya A, Shanmugam MP, Kumaramanickavel G. Mutational scanning of RB1 gene by multiplex PCR. Ind J of Biotechnol. 2005, 4, 2,194-200 | No direct sequencing |
| Joseph B, Raman R, Uthra S, Jagadeesan M, Ganesh A, Paul PG. Genotype-phenotype correlation analysis in retinoblastoma patients from India. Asian Pacific journal of cancer prevention: APJCP 2006, 7 (4), 619-22 | No direct sequencing |
| Kato MV, Ishizaki K, Toguchida J, Kaneko A, Takayama J, Tanooka H. Mutations in the retinoblastoma gene and their expression in somatic and tumor cells of patients with hereditary retinoblastoma. Human mutation 1994, 3(1), 44-51 | No direct sequencing |
| Kato MV, Ishizaki K, Shimizu T, Ejima Y, Tanooka H, Takayama J. Parental origin of germ-line and somatic mutations in the retinoblastoma gene. Human genetics 1994, 94(1), 31-8 | No direct sequencing |
| Ketteler, Hulsenbeck, Frank, Schmidt, Jockel, Lohmann. The influence of the type of predisposing RB1 variants on the incidence of malignancies. European Journal of Human Genetics 2020, 28, 1, 550 | No direct sequencing |
| Klutz M, Brockmann D, Lohmann DR. A parent-of-origin effect in two families with retinoblastoma is associated with a distinct splice mutation in the RB1 gene. American journal of human genetics 2002, 71(1), 174-9 | No direct sequencing |
| Liu Z, Song Y, Bia B, Cowell JK. Germline mutations in the RB1 gene in patients with hereditary retinoblastoma. Genes, chromosomes & cancer 1995, 14(4), 277-84. | No direct sequencing |
| Ohtani-Fujita N, Dryja TP, Rapaport JM, Fujita T, Matsumura S, Ozasa K, Watanabe Y, Hayashi K, Maeda K, Kinoshita S, Matsumura T, Ohnishi Y, Hotta Y, Takahashi R, Kato MV, Ishizaki K, Sasaki MS, Horsthemke B, Minoda K, Sakai T. | No direct sequencing |

| | |
|---|---|
| Hypermethylation in the retinoblastoma gene is associated with unilateral, sporadic retinoblastoma. Cancer Genet Cytogenet. 1997 Oct 1;98(1):43-9 | |
| Onadim Z, Cowell JK. Application of PCR amplification of DNA from paraffin embedded tissue sections to linkage analysis in familial retinoblastoma. J Med Genet. 1991, 28(5),312-6 | No direct sequencing |
| Onadim Z, Hogg A, Baird PN, Cowell JK. Oncogenic point mutations in exon 20 of the RB1 gene in families showing incomplete penetrance and mild expression of the retinoblastoma phenotype. Proc Natl Acad Sci U S A. 1992 ;89(13):6177-81 | No direct sequencing |
| Plowman PN, Pizer B, Kingston JE. Pineal Parenchymal Tumours: II: On the Aggressive Behaviour of Pineoblastoma in Patients with an Inherited Mutation of the RB1 Gene, Clinical Oncology, 16(4), 2004, 244-247 | No direct sequencing |
| Sana Qureshi, Jasmine H. Francis, Sofia S. Haque, Ira J. Dunkel, Mark M. Souweidane, Danielle N. Friedman, David H. Abramson, Magnetic Resonance Imaging Screening for Trilateral Retinoblastoma: The Memorial Sloan Kettering Cancer Center Experience 2006–2016, Ophthalmology Retina, 4, 3, 2020, 327-335 | No direct sequencing |
| Ramprasad VL, Madhavan J, Murugan S, Sujatha J, Suresh S, Sharma T, Kumaramanickavel G. Retinoblastoma in India : microsatellite analysis and its application in genetic counseling. Mol Diagn Ther. 2007;11(1):63-70 | No direct sequencing |
| Ruiz del Río N, Abelairas Gómez JM, Alonso García de la Rosa FJ, Peralta Calvo JM, de las Heras Martín A. Genetic analysis in retinoblastoma and peripheral blood correlation. Arch Soc Esp Oftalmol. 2015 Dec;90(12):562-5 | No direct sequencing |
| Salamanca-Gómez F, Luengas F, Antillón F. Genetic and cytogenetic studies in children with retinoblastoma. Cancer Genet Cytogenet. 1984 Oct;13(2):129-38. | No direct sequencing |
| Scheffer H, te Meerman GJ, Kruize YC, van den Berg AH, Penninga DP, Tan KE, der Kinderen DJ, Buys CH. Linkage analysis of families with hereditary retinoblastoma: nonpenetrance of mutation, revealed by combined use of markers within and flanking the RB1 gene. Am J Hum Genet. 1989 Aug;45(2):252-60 | No direct sequencing |
| Shields CL, Dockery P, Ruben M, Yaghy A, Sunday MA, Duffner ER, Levin HJ, Taylor OS, Calotti M, Lally SE, Shields JA. Likelihood of Germline Mutation With Solitary Unilateral Retinoblastoma Based on Patient Age at Presentation: Analysis of 482 Consecutive Patients. J Pediatr Ophthalmol Strabismus. 2021 Nov-Dec;58(6):355-364 | No direct sequencing |
| Shimizu T, Toguchida J, Kato MV, Kaneko A, Ishizaki K, Sasaki MS. Detection of mutations of the RB1 gene in retinoblastoma patients by using exon-by-exon PCR-SSCP analysis. Am J Hum Genet. 1994 May;54(5):793-800 | No direct sequencing |

| | |
|---|---|
| Toncheva D, Genkova P, Tzoneva M, Konstantinov I, Markova V. High-resolution chromosome analysis in retinoblastoma. Neoplasma. 1987;34(1):23-6 | No direct sequencing |
| Tran HV, Schorderet DF, Gaillard MC, Balmer A, Munier FL. Risk assessment of recurrence in sporadic retinoblastoma using a molecular-based algorithm. Ophthalmic Genet. 2012 Mar;33(1):6-11 | No direct sequencing |
| Yazici H, Kir N, Taş S, Ayan I, Kebidi R, Peksayar G, Dalay N. Investigation of the loss of heterozygosity and familial segregation by PCR in retinoblastoma. Clin Biochem. 1996 Dec;29(6):595-8 | No direct sequencing |
| Yu YS, Kim IJ, Ku JL, Park JG. Identification of four novel RB1 germline mutations in Korean retinoblastoma patients. Hum Mutat. 2001 Sep;18(3):252 | No direct sequencing |
| Zhang Q, Minoda K. Detection of RB germline mutations using exon-by-exon heteroduplex analysis compared with SSCP. Yan Ke Xue Bao. 1996 Sep;12(3):151-7 | No direct sequencing |
| Macías M, Dean M, Atkinson A, Jiménez-Morales S, García-Vazquez FJ, Saldaña-Alvarez Y, Ramírez-Bello J, Chávez M, Orozco L. Spectrum of RB1 gene mutations and loss of heterozygosity in Mexican patients with retinoblastoma: identification of six novel mutations. Cancer Biomark. 2008;4(2):93-9 | FT not retrievable |
| Mallipatna A, Marino M, Singh AD. Genetics of Retinoblastoma. Asia Pac J Ophthalmol (Phila). 2016 Jul-Aug;5(4):260-4 | FT not retrievable |
| Mathew L, Miale TD, Rao S, Lobel SA, Fishman GA, Goldberg MF. Retrospective analysis of 58 children with retinoblastoma. Ophthalmic Paediatr Genet. 1984 Aug;4(2):67-74 | FT not retrievable |
| Temming P, Lohmann D, Bornfeld N, Sauerwein W, Goericke SL, Eggert A. Current concepts for diagnosis and treatment of retinoblastoma in Germany: aiming for safe tumor control and vision preservation. Klin Padiatr. 2012 Oct;224(6):339-47 | FT not retrievable |
| Zhang Q, Minoda K, Zeng R, Wu Z, Xiao X, Li S, Zhang F. Exon-by-exon screening for RB germline mutations using Heteroduplex-SSCP analysis. Yan Ke Xue Bao. 1997 Mar;13(1):5-11 | FT not retrievable |
| Tanwar, M., Balaji, S., Vanniarajan, A. *et al.* Parental age and retinoblastoma—a retrospective study of demographic data and genetic analysis. *Eye* **36**, 57–63 (2022) | Not related to genetics |
| Torbidoni AV, Sampor C, Laurent VE, Aschero R, Iyer S, Rossi J, Alderete D, Alonso DF, Szijan I, Chantada GL. Minimal disseminated disease evaluation and outcome in trilateral retinoblastoma. Br J Ophthalmol. 2018 Nov;102(11):1597-1601 | Not related to genetics |

| | |
|---|---|
| Leone, P.E., Vega, M.E., Jervis, P. *et al.* Two new mutations and three novel polymorphisms in the *RB1* gene in Ecuadorian patients. *J Hum Genet* **48**, 639–641 (2003). | Test for specific variants or types of mutation |
| Alonso J, García-Miguel P, Abelairas J, Mendiola M, Sarret E, Vendrell MT, Navajas A, Pestaña A. Spectrum of germline RB1 gene mutations in Spanish retinoblastoma patients: Phenotypic and molecular epidemiological implications. Hum Mutat. 2001 May;17(5):412-22 | Publication type |
| Vanniarajan Ayyasamy, Thirumalairaj Kannan, Aloysius Abraham, Bharanidharan Devarajan, Namrata Gaikwad, Veerappan Muthukkaruppan, Usha Kim; A New Sequential Screening Strategy for Rapid Diagnosis of Retinoblastoma. *Invest. Ophthalmol. Vis. Sci.* 2015;56(7 ):1242 | Publication type |
| Bookstein R, Lee EY, To H, Young LJ, Sery TW, Hayes RC, Friedmann T, Lee WH. Human retinoblastoma susceptibility gene: genomic organization and analysis of heterozygous intragenic deletion mutants. Proc Natl Acad Sci U S A. 1988 Apr;85(7):2210-4 | Publication type |
| A. Fernandez-Teijeiro, F. J. Alonso Garcia De La Rosa, A. Varo Rodriguez, M. Benitez Carabante, D. Garcia Aldana, N. Conde Cuevas and F. Espejo Arjona. Germline RB1 gene mutations analysis in 60 patients from a spanish national reference unit for retinoblastoma. Pediatric Blood and Cancer 2013 Vol. 60 Issue SUPPL. 3 Pages 128 | Publication type |
| S. Frenkel and J. Pe'er. The clinical presentation of retinoblastoma patients with mosaics. Investigative Ophthalmology and Visual Science 2016 Vol. 57, 12, 3672 | Publication type |
| N. Guha, S. A. Deepak, S. Lateef, A. Padmanabhan, S. Gundimeda, A. Ghosh, et al. A multiomics approach to identify biomarkers of clinically advanced retinoblastoma for diagnostics and therapeutic applications. FASEB Journal 2015 Vol. 29 | Publication type |
| Herzog H, Darby K, Hort YJ, Shine J. Intron 17 of the human retinoblastoma susceptibility gene encodes an actively transcribed G protein-coupled receptor gene. Genome Res. 1996 Sep;6(9):858-61 | Publication type |
| Horsthemke B, Greger V, Barnert HJ, Höpping W, Passarge E. Detection of submicroscopic deletions and a DNA polymorphism at the retinoblastoma locus. Hum Genet. 1987 Jul;76(3):257-61 | Publication type |
| Hung CC, Lin SY, Lee CN, Chen CP, Lin SP, Chao MC, Chiou SS, Su YN. Low penetrance of retinoblastoma for p.V654L mutation of the RB1 gene. BMC Med Genet. 2011 May 26;12:76 | **Publication type** |
| Munier FL, Wang MX, Spence MA, Thonney F, Balmer A, Pescia G, Donoso LA, Murphree AL. Pseudo low penetrance in retinoblastoma. Fortuitous familial aggregation of sporadic cases caused by independently derived mutations in two large pedigrees. Arch Ophthalmol. 1993 Nov;111(11):1507-11 | Publication type |

| | |
|---|---|
| Munier F, Spence MA, Pescia G, Balmer A, Gailloud C, Thonney F, van Melle G, Rutz HP. Paternal selection favoring mutant alleles of the retinoblastoma susceptibility gene. Hum Genet. 1992 Jul;89(5):508-12 | Publication type |
| Sasaki O, Meguro K, Tohmiya Y, Funato T, Shibahara S, Sasaki T. Altered expression of retinoblastoma protein-interacting zinc finger gene, RIZ, in human leukaemia. Br J Haematol. 2002 Dec;119(4):940-8 | Publication type |
| Seo SH, Ahn HS, Yu YS, Kang HJ, Park KD, Cho SI, Park JS, Hyun YJ, Kim JY, Seong MW, Park SS. Mutation spectrum of RB1 gene in Korean bilateral retinoblastoma patients using direct sequencing and gene dosage analysis. Clin Genet. 2013 May;83(5):494-6 | Publication type |
| Seo SH, Ahn HS, Yu YS, Kang HJ, Park KD, Cho SI, Park JS, Hyun YJ, Kim JY, Seong MW, Park SS. Mutation spectrum of RB1 gene in Korean bilateral retinoblastoma patients using direct sequencing and gene dosage analysis. Clin Genet. 2013 May;83(5):494-6 | Publication type |
| Tomar S, Sethi R, Sundar G, Quah TC, Quah BL, Lai PS. AB109. Novel constitutional and somatic *RB1* mutations underlying retinal cancers in addition to *TNFα*, *KIF13A* and *MGMT* alterations. Ann Transl Med. 2017 Sep;5(Suppl 2):AB109 | Publication type |
| Valverde JR, Alonso J, Palacios I, Pestaña A. RB1 gene mutation up-date, a meta-analysis based on 932 reported mutations available in a searchable database. BMC Genet. 2005 Nov 4;6:53 | Publication type |
| Michael Francis Walsh et al., The heritability of retinoblastoma: An institutional review.. JCO 34, 1591-1591(2016) | Publication type |
| Xu Y, Fu Z, Gao X, Wang R, Li Q. Long non-coding RNA XIST promotes retinoblastoma cell proliferation, migration, and invasion by modulating microRNA-191-5p/brain derived neurotrophic factor. Bioengineered. 2021 Dec;12(1):1587-1598 | Publication type |
| Yilmaz S, Horsthemke B, Lohmann DR. Twelve novel RB1 gene mutations in patients with hereditary retinoblastoma. Mutations in brief no. 206. Online. Hum Mutat. 1998;12(6):434 | Publication type |
| Zhu XP, Dunn JM, Phillips RA, Goddard AD, Paton KE, Becker A, Gallie BL. Preferential germline mutation of the paternal allele in retinoblastoma. Nature. 1989 Jul 27;340(6231):312-3 | Publication type |
| Mitter D, Ullmann R, Muradyan A, Klein-Hitpass L, Kanber D, Ounap K, Kaulisch M, Lohmann D. Genotype-phenotype correlations in patients with retinoblastoma and interstitial 13q deletions. Eur J Hum Genet. 2011 Sep;19(9):947-58 | Test for specific variants or types of mutation |
| Zhang, XL, Fu, WL, Wang, JH, Zhou, LX, Lei, MP, Hua, C. New suspicious gene loci correlated with the generation and development of children's retinoblastoma. Chinese Journal of Clinical Rehabilitation. 9(11), 197-199 | Test for specific variants or types of mutation |

| | |
|---|---|
| Yusof, H.A.,Ramlee, N.,Ishak, S.R.,Ab Rajab, N.S.,Ghani, S.A.,Bachok, N.S.,Aziz, S.H.S.A.,Tajudin, L.-S.A.,Alagaratnam, J.V.,San, L.P.,Nishio, H.,Alwi, Z.B. (2010-06). Identification of single nucleotide polymorphism (SNP) 153104 (A to G) of RB1 gene in Malaysian retinoblastoma children and its association with laterality and staging of the disease. International Medical Journal 17 (2) : 129-133 | Test for specific variants or types of mutation |
| Dryja TP, Morrow JF, Rapaport JM. Quantification of the paternal allele bias for new germline mutations in the retinoblastoma gene. Hum Genet. 1997 Sep;100(3-4):446-9 | Test for specific variants or types of mutation |
| Ayari-Jeridi H, Moran K, Chebbi A, Bouguila H, Abbes I, Charradi K, Benammar-Elgaaïed A, Ganguly A. Mutation spectrum of RB1 gene in unilateral retinoblastoma cases from Tunisia and correlations with clinical features. PLoS One. 2015 Jan 20;10(1):e0116615 | Test for specific variants or types of mutation |
| Raizis A, Clemett R, Corbett R, McGaughran J, Evans J, George P. Improved clinical management of retinoblastoma through gene testing. N Z Med J. 2002 May 24;115(1154):231-4 | Not blood sample test |
| Abramson DH, Mandelker D, Francis JH, Dunkel IJ, Brannon AR, Benayed R, Berger MF, Arcila ME, Ladanyi M, Friedman DN, Jayakumaran G, Diosdado MS, Robbins MA, Haggag-Lindgren D, Shukla N, Walsh M, Kothari P, Tsui DWY. Retrospective Evaluation of Somatic Alterations in Cell-Free DNA from Blood in Retinoblastoma. Ophthalmol Sci. 2021 Mar 16;1(1):100015 | Not blood sample test |
| Dunn JM, Phillips RA, Zhu X, Becker A, Gallie BL. Mutations in the RB1 gene and their effects on transcription. Mol Cell Biol. 1989 Nov;9(11):4596-604 | Not blood sample test |
| Ganguly A, Shields CL. Differential gene expression profile of retinoblastoma compared to normal retina. Mol Vis. 2010 Jul 13;16:1292-303 | Not blood sample test |
| McEvoy J, Nagahawatte P, Finkelstein D, Richards-Yutz J, Valentine M, Ma J, Mullighan C, Song G, Chen X, Wilson M, Brennan R, Pounds S, Becksfort J, Huether R, Lu C, Fulton RS, Fulton LL, Hong X, Dooling DJ, Ochoa K, Mardis ER, Wilson RK, Easton J, Zhang J, Downing JR, Ganguly A, Dyer MA. RB1 gene inactivation by chromothripsis in human retinoblastoma. Oncotarget. 2014 Jan 30;5(2):438-50. | Not blood sample test |
| Rushlow, Diane E, Mol, Berber M, Kennett, Jennifer Y, Yee, Stephanie<br>Pajovic, Sanja, Theriault, Brigitte L, Prigoda-Lee, Nadia L, Spencer, Clarellen, Dimaras, Helen, Corson, Timothy W, Pang, Renee, Massey, Christine, Godbout, Roseline et al. Characterisation of retinoblastomas without RB1 mutations: genomic, gene expression, and clinical studies. 2013, The Lancet. Oncology, 14, 4, 327-34. | Not blood sample test |
| Schmidt MJ, Prabakar RK, Pike S, Yellapantula V, Peng CC, Kuhn P, Hicks J, Xu L, Berry JL. Simultaneous Copy Number Alteration and Single-Nucleotide Variation Analysis in Matched Aqueous Humor and Tumor Samples in Children with Retinoblastoma. Int J Mol Sci. 2023 May 11;24(10):8606 | Not blood sample test |

| | |
|---|---|
| Su G, Senft S, Cowell J. Oncogenic mutations in the rb1 gene in retinoblastoma tumors from patients from the kingdom of Saudi-Arabia. Int J Oncol. 1995 Mar;6(3):687-91 | Not blood sample test |
| Xu L, Shen L, Polski A, Prabakar RK, Shah R, Jubran R, Kim JW, Biegel J, Kuhn P, Cobrinik D, Hicks J, Gai X, Berry JL. Simultaneous identification of clinically relevant *RB1* mutations and copy number alterations in aqueous humor of retinoblastoma eyes. Ophthalmic Genet. 2020 Dec;41(6):526-532 | Not blood sample test |
| Yandell DW, Dryja TP. Detection of DNA sequence polymorphisms by enzymatic amplification and direct genomic sequencing. Am J Hum Genet. 1989 Oct;45(4):547-55 | Not blood sample test |
| Kato MV, Shimizu T, Nagayoshi M, Kaneko A, Sasaki MS, Ikawa Y. Genomic imprinting of the human serotonin-receptor (HTR2) gene involved in development of retinoblastoma. Am J Hum Genet. 1996 Nov;59(5):1084-90 | Not blood sample test |
| Kothari P, Marass F, Yang JL, Stewart CM, Stephens D, Patel J, Hasan M, Jing X, Meng F, Enriquez J, Huberman K, Viale A, Francis JH, Berger MF, Shukla N, Abramson DH, Dunkel IJ, Tsui DWY. Cell-free DNA profiling in retinoblastoma patients with advanced intraocular disease: An MSKCC experience. Cancer Med. 2020 Sep;9(17):6093-6101 | Validation of new test |
| Jessica Le Gall, Catherine Dehainault, Camille Benoist, Alexandre Matet, Livia Lumbroso-Le Rouic, Isabelle Aerts, Irene Jiménez, Gudrun Schleiermacher, Claude Houdayer, François Radvanyi, Eleonore Frouin, Victor Renault, François Doz, Dominique Stoppa-Lyonnet, Marion Gauthier-Villars, Nathalie Cassoux, Lisa Golmard, Highly Sensitive Detection Method of Retinoblastoma Genetic Predisposition and Biomarkers, The Journal of Molecular Diagnostics, 23, 12, 2021, 1714-1721 | Validation of new test |
| Li M, Chen XM, Wang DM, Gan L, Qiao Y. Effects of miR-26a on the expression of Beclin 1 in retinoblastoma cells. Genet Mol Res. 2016 Jul 14;15(2) | Validation of new test |
| Gerrish A, Bowns B, Mashayamombe-Wolfgarten C, Young E, Court S, Bott J, McCalla M, Ramsden S, Parks M, Goudie D, Carless S, Clokie S, Cole T, Allen S. Non-Invasive Prenatal Diagnosis of Retinoblastoma Inheritance by Combined Targeted Sequencing Strategies. J Clin Med. 2020 Oct 30;9(11):3517 | Validation of new test |
| Kiet NC, Khuong LT, Minh DD; Nguyen The Vinh; Quan NHM, Xinh PT, Trang NNC, Luan NT, Khai NM, Vu HA. Spectrum of mutations in the RB1 gene in Vietnamese patients with retinoblastoma. Mol Vis. 2019;25:215-221 | Subpopulation |
| Ahani A, Akbari MT, Saliminejad K, Behnam B, Akhondi MM, Vosoogh P, Ghassemi F, Naseripour M, Bahoush G, Khorshid HR. Screening for large rearrangements of the RB1 gene in Iranian patients with retinoblastoma using multiplex ligation-dependent probe amplification. Mol Vis. 2013;19:454-62 | Subpopulation |
| Lan X, Xu W, Tang X, Ye H, Song X, Lin L, Ren X, Yu G, Zhang H, Wu S. Spectrum of RB1 Germline Mutations and Clinical Features in Unrelated Chinese Patients With Retinoblastoma. Front Genet. 2020;11:142 | Subpopulation |

| | |
|---|---|
| Mendonça V, Evangelista AC, P Matta B, M Moreira MÂ, Faria P, Lucena E, Seuánez HN. Molecular alterations in retinoblastoma beyond RB1. Exp Eye Res. 2021;211:108753 | Subpopulation |
| Davies HR, Broad KD, Onadim Z, Price EA, Zou X, Sheriff I, Karaa EK, Scheimberg I, Reddy MA, Sagoo MS, Ohnuma SI, Nik-Zainal S. Whole-Genome Sequencing of Retinoblastoma Reveals the Diversity of Rearrangements Disrupting RB1 and Uncovers a Treatment-Related Mutational Signature. Cancers (Basel). 2021;13(4):754 | Subpopulation |
| Tsai T, Fulton L, Smith BJ, Mueller RL, Gonzalez GA, Uusitalo MS, O'Brien JM. Rapid identification of germline mutations in retinoblastoma by protein truncation testing. Arch Ophthalmol. 2004;122(2):239-48 | Subpopulation |
| Sippel KC, Fraioli RE, Smith GD, Schalkoff ME, Sutherland J, Gallie BL, Dryja TP. Frequency of somatic and germ-line mosaicism in retinoblastoma: implications for genetic counseling. Am J Hum Genet. 1998;62(3):610-9 | Subpopulation |
| Zhang H, Qiu X, Song Z, Lan L, Ren X, Ye B. CircCUL2 suppresses retinoblastoma cells by regulating miR-214-5p/E2F2 Axis. Anticancer Drugs. 2022;33(1):e218-e227 | Subpopulation |
| Rushlow D, Piovesan B, Zhang K, et al. Detection of mosaic RB1 mutations in families with retinoblastoma. Human Mutation. 2009;30(5):842-851 | Subpopulation |
| Albrecht P, Ansperger-Rescher B, Schüler A, Zeschnigk M, Gallie B, Lohmann DR. Spectrum of gross deletions and insertions in the RB1 gene in patients with retinoblastoma and association with phenotypic expression. Hum Mutat. 2005;26(5):437-45 | Subpopulation |
| Reddy MA, Butt M, Hinds AM, Duncan C, Price EA, Sagoo MS, Onadim Z. Prognostic Information for Known Genetic Carriers of RB1 Pathogenic Variants (Germline and Mosaic). Ophthalmol Retina. 2021;5(4):381-387 | Subpopulation |
| Grotta S, D'Elia G, Scavelli R, Genovese S, Surace C, Sirleto P, Cozza R, Romanzo A, De Ioris MA, Valente P, Tomaiuolo AC, Lepri FR, Franchin T, Ciocca L, Russo S, Locatelli F, Angioni A. Advantages of a next generation sequencing targeted approach for the molecular diagnosis of retinoblastoma. BMC Cancer. 2015;15:841 | Subpopulation |
| Barbosa RH, Vargas FR, Aguiar FC, Ferman S, Lucena E, Bonvicino CR, Seuánez HN. Hereditary retinoblastoma transmitted by maternal germline mosaicism. Pediatr Blood Cancer. 2008;51(5):598-60 | Single family |
| Genuardi M, Klutz M, Devriendt K, Caruso D, Stirpe M, Lohmann DR. Multiple lipomas linked to an RB1 gene mutation in a large pedigree with low penetrance retinoblastoma. Eur J Hum Genet. 2001;9(9):690-4 | Single family |
| Hung, CC., Lin, SY., Lee, CN. et al. Low penetrance of retinoblastoma for p.V654L mutation of the RB1 gene. BMC Med Genet. 2011;12;76 | Single family |

| | |
|---|---|
| Sánchez-Sánchez F, Ramírez-Castillejo C, Weekes DB, Beneyto M, Prieto F, Nájera C, Mittnacht S. Attenuation of disease phenotype through alternative translation initiation in low-penetrance retinoblastoma. Hum Mutat. 2007;28(2):159-67 | Single family |

#### XLHR

| Reference | Reason for exclusion (Q2) |
|---|---|
| Acar S, Al-Rijjal RA, Meyer B, Shi Y, Demir K, Bober E, et al. Clinical and genetic features of our patients with hypophosphatemic rickets. JCRPE Journal of Clinical Research in Pediatric Endocrinology. 2017;9(Supplement 1):28. | publication type (inc. abstract only) |
| Acar S, BinEssa HA, Demir K, Al-Rijjal RA, Zou M, Catli G, et al. Clinical and genetic characteristics of 15 families with hereditary hypophosphatemia: Novel Mutations in PHEX and SLC34A3. PloS one. 2018;13(3):e0193388. | duplicate |
| Al Kaissi A, Farr S, Ganger R, Klaushofer K, Grill F. Windswept lower limb deformities in patients with hypophosphataemic rickets. Swiss medical weekly. 2013;143:w13904. | no sequencing |
| Alikasifoglu A, Unsal Y, Gonc N, Ozon A, Kandemir N, Alikasifoglu M. Is conventional treatment still the first choice in pediatric patients with PHEX mutations in an era of monoclonal FGF-23 antibody? Hormone Research in Paediatrics. 2021;94(SUPPL 1):210-1. | publication type (inc. abstract only) |
| Al-Juraibah F, Al-Dubayee M, Babiker A. Experience of burosumab therapy in four children with X-linked hypophosphataemia in Saudi Arabia. Hormone Research in Paediatrics. 2019;91(Supplement 1):373. | publication type (inc. abstract only) |
| Alonso G, Plantalech L, Guelman R, Gonzalez S, Cassinelli H, Redal M, et al. Familial and sporadic hypophosphatemic rickets: Clinical and molecular findings. Actualizaciones en Osteologia. 2014;10(2):e97830. | language |
| Alonso G, Plantalech L, Guelman R, Gonzalez S, Redal MA, Cassinelli H, et al. Familiar hypophosphatemic rickets: Molecular findings in thirteen Argentinean families. Bone. 2011;48(6):S288-S9. | publication type (inc. abstract only) |
| Alonso G, Plantalech L, Guelman R, Gonzalez S, Redal MA, Cassinelli H, et al. Familiar hypophosphatemic rickets: Molecular findings in thirteen Argentinean families. Bone. 2011;48(6):S288-S9. | publication type (inc. abstract only) |

| | |
|---|---|
| Ambrosetti I, Olivucci G, Graziano C, Lanzoni G, Severi G, Ambrosini E, et al. Genotype-phenotype correlation in patients with PHEX-related hypophosphatemia: identification of novel variants and a case of mosaicism. European Journal of Human Genetics. 2023;31(Supplement 1):384. | publication type (inc. abstract only) |
| Arenas MA, Jaimovich S, Perez Garrido N, Del Pino M, Viterbo G, Marino R, et al. Hereditary hypophosphatemic rickets and craniosynostosis. Journal of pediatric endocrinology & metabolism : JPEM. 2021;34(9):1105-13. | subpopulation |
| Baroncelli GI, Zampollo E, Manca M, Toschi B, Bertelloni S, Michelucci A, et al. Pulp chamber features, prevalence of abscesses, disease severity, and PHEX mutation in X-linked hypophosphatemic rickets. Journal of bone and mineral metabolism. 2021;39(2):212-23. | no sequencing |
| Beck-Nielsen SS, Brixen K, Gram J, Brock-Jacobsen B, Brusgaard K. Mutational analysis of the PHEX, FGF23, DMP1, SCL34A3 and CLCN5 genes in patients with Hypophosphatemic Rickets. Hormone Research in Paediatrics. 2011;76(SUPPL. 2):59. | publication type (inc. abstract only) |
| Beck-Nielsen SS, Brixen K, Gram J, Brusgaard K. Mutational analysis of PHEX, FGF23, DMP1, SLC34A3 and CLCN5 in patients with hypophosphatemic rickets. Journal of human genetics. 2012;57(7):453-8. | unclear age |
| Burnett CH, Dent CE, Harper G, Warland BJ. Vitamin d-resietant rickets. analysis of twenty-four pedigrees with hereditary and sporadic cases. American Journal of Medicine. 1964;36(2):222-32. | no sequencing |
| Christie PT, Harding B, Nesbit MA, Whyte MP, Thakker RV. X-linked hypophosphatemia attributable to pseudoexons of the PHEX gene. The Journal of clinical endocrinology and metabolism. 2001;86(8):3840-4. | subpopulation |
| Clausmeyer S, Hesse V, Clemens PC, Engelbach M, Kreuzer M, Becker-Rose P, et al. Mutational analysis of the PHEX gene: novel point mutations and detection of large deletions by MLPA in patients with X-linked hypophosphatemic rickets. Calcified tissue international. 2009;85(3):211-20. | subpopulation |
| Colares G, Kaupert L, Bussmann L, Silveira P, Matsunaga R. Identification of three novel PHEX mutations in brazilian pediatric patients with X-linked hypophosphatemic rickets. Hormone Research in Paediatrics. 2011;76(SUPPL. 4):29. | publication type (inc. abstract only) |
| Dahir K, Rush E, Beltran D, Eisenbeis S, Johnson B, Ramesan P, et al. Hypophosphatemia Gene Panel Sponsored Program: A High Yield of Molecular Diagnoses from Clinically Confirmed XLH and | publication type (inc. abstract only) |

| | |
|---|---|
| Suspected Genetic Hypophosphatemia. Journal of Bone and Mineral Research. 2020;35(SUPPL 1):304. | |
| De Paula Colares Neto G, Ferreira De Assis Funari M, Ferraz De Souza B, Matsunaga Martin R. Molecular basis of 47 patients with FGF23-mediated hypophosphatemic rickets in a single-center study. Hormone Research in Paediatrics. 2016;86(Supplement 2):21. | publication type (inc. abstract only) |
| De Paula Colares Neto G, Silveira Corre PH, Martin RM. Evaluation of nephrocalcinosis and nephrolithiasis in eleven children with X-linked hypophosphataemic rickets confirmed with mutations in PHEX gene. Hormone Research in Paediatrics. 2013;80(SUPPL. 1):219. | publication type (inc. abstract only) |
| Dgraham JB. Genetic errors of metabolism and environmental interaction. familial hypophosphatemia: an inherited demand for increased vitamin. Ann NY Acad Sd. 1961;91(3):667-73. | no sequencing |
| Dixon PH, Christie PT, Wooding C, Trump D, Grieff M, Holm I, et al. Mutational analysis of PHEX gene in X-linked Hypophosphatemia1. J Clin Endocrinol Metab. 1998;83(10):3615-23. | unclear age |
| Durmaz E, Zou M, Al-Rijjal RA, Baitei EY, Hammami S, Bircan I, et al. Novel and de novo PHEX mutations in patients with hypophosphatemic rickets. Bone. 2013;52(1):286-91. | duplicate |
| Econs MJ, Friedman NE, Rowe PS, Speer MC, Francis F, Strom TM, et al. A PHEX gene mutation is responsible for adult-onset vitamin D-resistant hypophosphatemic osteomalacia: evidence that the disorder is not a distinct entity from X-linked hypophosphatemic rickets. The Journal of clinical endocrinology and metabolism. 1998;83(10):3459-62. | subpopulation |
| Econs MJ, Friedman NE, Rowe PS, Speer MC, Francis F, Strom TM, et al. A PHEX gene mutation is responsible for adult-onset vitamin D-resistant hypophosphatemic osteomalacia: evidence that the disorder is not a distinct entity from X-linked hypophosphatemic rickets. The Journal of clinical endocrinology and metabolism. 1998;83(10):3459-62. | publication type (inc. abstract only) |
| Fahiminiya S, Almuriekhi M, Nawaz Z, Staffa A, Lepage P, Ali R, et al. Whole exome sequencing unravels disease-causing genes in consanguineous families in Qatar. Clinical genetics. 2014;86(2):134-41. | publication type (inc. abstract only) |
| Fahiminiya S, Almuriekhi M, Nawaz Z, Staffa A, Lepage P, Ali R, et al. Whole exome sequencing unravels disease-causing genes in consanguineous families in Qatar. Clinical genetics. 2014;86(2):134-41. | unclear age? |

| | |
|---|---|
| Fortuno J, Oliva M, Royo I, Flores M, Torrents A, Martinez F, et al. Mosaicism in the PHEX gene. Is it more common than we thought? European Journal of Human Genetics. 2020;28(SUPPL 1):832. | publication type (inc. abstract only) |
| Fortuno J, Oliva M, Royo I, Flores M, Torrents A, Martinez F, et al. Mosaicism in the PHEX gene. Is it more common than we thought? European Journal of Human Genetics. 2020;28(SUPPL 1):832. | unclear age |
| Gottesman G, Wollberg V, Mumm S, Whyte M. Mild X-linked hypophosphatemia (PHEX c. 231A>G): Normalization of Serum Phosphorus Levels with Low-Dose Burosumab Therapy. Journal of Bone and Mineral Research. 2020;35(SUPPL 1):286. | publication type (inc. abstract only) |
| Gu J, Wang C, Zhang H, Yue H, Hu W, He J, et al. Targeted resequencing of phosphorus metabolism-related genes in 86 patients with hypophosphatemic rickets/osteomalacia. International journal of molecular medicine. 2018;42(3):1603-14. | unclear age |
| Huang Y, Mei L, Pan Q, Tan H, Quan Y, Gui B, et al. Novel de novo nonsense mutation of the PHEX gene (p.Lys50Ter) in a Chinese patient with hypophosphatemic rickets. Gene. 2015;565(1):150-4. | duplicate |
| Huskey M, Duan S, Wollberg V, Mack KE, Gu CC, Gottesman G. Next generation sequencing for hypophosphatasia and X-linked hypophosphatemia. Journal of Bone and Mineral Research. 2015;30(Supplement 1). | publication type (inc. abstract only) |
| Ichikawa S, Traxler EA, Estwick SA, Curry LR, Johnson ML, Sorenson AH, et al. Mutational survey of the PHEX gene in patients with X-linked hypophosphatemic rickets. Bone. 2008;43(4):663-6. | unclear age |
| Ishihara Y, Ohata Y, Takeyari S, Kitaoka T, Fujiwara M, Nakano Y, et al. Genotype-phenotype analysis, and assessment of the importance of the zinc-binding site in PHEX in Japanese patients with X-linked hypophosphatemic rickets using 3D structure modeling. Bone. 2021;153:116135. | unclear age |
| Kang Q-l, Xu J, Zhang Z, He J-w, Lu L-s, Fu W-z, et al. Three novel PHEX gene mutations in four Chinese families with X-linked dominant hypophosphatemic rickets. Biochemical and biophysical research communications. 2012;423(4):793-8. | >10% adults and not reported separately |
| Kienitz T, Ventz M, Kaminsky E, Quinkler M. Novel PHEX nonsense mutation in a patient with X-linked hypophosphatemic rickets and review of current therapeutic regimens. Experimental and clinical endocrinology & diabetes : official journal, German Society of Endocrinology [and] German Diabetes Association. 2011;119(7):431-5. | publication type (inc. abstract only) |

| | |
|---|---|
| Kulikova K, Kolodkina A, Vasilyev E, Petrov V, Gofman F, Horkin A, et al. The spectrum of molecular defects in 64 patients with hypophosphatemic rickets identified by targeted next-generation sequencing. Hormone Research in Paediatrics. 2015;84(SUPPL. 1):161. | publication type (inc. abstract only) |
| Lee S-H, Agashe MV, Suh S-W, Yoon Y-C, Song S-H, Yang J-H, et al. Paravertebral ligament ossification in vitamin D-resistant rickets: incidence, clinical significance, and genetic evaluation. Spine. 2012;37(13):E792-6. | >10% adults and not reported separately |
| Lin X, Li S, Zhang Z, Yue H. Clinical and Genetic Characteristics of 153 Chinese Patients With X-Linked Hypophosphatemia. Frontiers in cell and developmental biology. 2021;9:617738. | >10% adults and not reported separately |
| Makitie O, Liu J, Williams A, Wood S. First Interim Analysis of the International X-Linked Hypophosphataemia (XLH) Registry: Family history and genetic findings. European Journal of Human Genetics. 2023;31(Supplement 1):123. | publication type (inc. abstract only) |
| Marik B, Bagga A, Sinha A, Hari P, Sharma A. A study on the genetic and molecular basis of hypophosphatemic rickets (HR). Revista Argentina de Endocrinologia y Metabolismo. 2021;58(SUPPL 1):31. | publication type (inc. abstract only) |
| Marik B, Bagga A, Sinha A, Khandelwal P, Hari P, Sharma A. Genetic study on the patients with refractory rickets. European Journal of Human Genetics. 2020;28(SUPPL 1):1014-5. | publication type (inc. abstract only) |
| Miller N, Daugherty S, Sarafrazi S, Boada P, Carpenter T, Chunn L, et al. A comprehensive locus specific database of PHEX gene variants associated with X-linked hypophosphatemia vastly increases the number of known variants. Molecular Genetics and Metabolism. 2021;132(Supplement 1):S133. | publication type (inc. abstract only) |
| Morey M, Castro-Feijoo L, Barreiro J, Cabanas P, Gil M, Bernabeu I, et al. Genetic analysis of X-linked dominant hypophosphatemic rickets. Hormone Research in Paediatrics. 2010;74(SUPPL. 3):174. | publication type (inc. abstract only) |
| Mumm S, Huskey M, Cajic A, Wollberg V, Zhang F, Madson KL, et al. PHEX 3'-UTR c.*231A>G near the polyadenylation signal is a relatively common, mild, American mutation that masquerades as sporadic or X-linked recessive hypophosphatemic rickets. Journal of bone and mineral research : the official journal of the American Society for Bone and Mineral Research. 2015;30(1):137-43. | subpopulation |
| Mumm S, Huskey M, Cajic A, Wollberg V, Zhang F, Madson KL, et al. PHEX 3'-UTR c.*231A>G near the polyadenylation signal is a relatively common, mild, American mutation that masquerades as sporadic or X-linked recessive hypophosphatemic rickets. | publication type (inc. abstract only) |

| | |
|---|---|
| Journal of bone and mineral research : the official journal of the American Society for Bone and Mineral Research. 2015;30(1):137-43. | |
| Mumm S, Huskey M, Wollberg V, Madson K, Wenkert D, Gottesman GS, et al. Identification of the polya mutation (C.*231a>G) in the PHEX 3'UTR in five boys with X-linked hypophosphatemia (XLH). Journal of Bone and Mineral Research. 2012;27(SUPPL. 1). | publication type (inc. abstract only) |
| Nielsen LH, Rahbek ET, Beck-Nielsen SS, Christesen HT. Treatment of hypophosphataemic rickets in children remains a challenge. Danish medical journal. 2014;61(7):A4874. | no sequencing |
| Ohata Y, Kubota T, Takeyari S, Kitaoka T, Nakano Y, Miyata K, et al. Mutational analysis of the PHEX gene and genotype-phenotype correlation in 37 Japanese patients with X-linked hypophosphatemic rickets. Journal of Bone and Mineral Research. 2019;34(Supplement 1):265. | publication type (inc. abstract only) |
| Park PG, Lim SH, Ahn YH, Kang HG, Ha IS. Genotype and phenotype analysis in patients with X-linked hypophosphatemia. Journal of the American Society of Nephrology. 2021;32:432. | publication type (inc. abstract only) |
| Park PG, Lim SH, Lee H, Ahn YH, Cheong HI, Kang HG. Genotype and Phenotype Analysis in X-Linked Hypophosphatemia. Frontiers in pediatrics. 2021;9:699767. | publication type (inc. abstract only) |
| Popowska E, Pronicka E, Sulek A, Jurkiewicz D, Rowe P, Rowinska E, et al. X-linked hypophosphatemia in Polish patients. 1. Mutations in the PHEX gene. Journal of applied genetics. 2000;41(4):293-302. | unclear age |
| Prentice P, Owens M, Brain C, Allgrove J, Gevers E. Mosaic PHEX variants are important causes of X-linked hypophosphataemic rickets. Hormone Research in Paediatrics. 2021;94(SUPPL 1):98. | publication type (inc. abstract only) |
| Pronicka E, Popowska E, Rowinska E, Piekutowska D, Oglecka M, Krajewskawalasek M. BIOCHEMICAL AND DNA MARKERS OF X-LINKED HYPOPHOSPHATEMIC RICKETS - A STUDY OF SPORADIC CASES. Journal of Inherited Metabolic Disease. 1992;15(3):335-8. | no sequencing |
| Pronicka E, Popowska E, Rowinska E, Piekutowska D, Oglecka M, Krajewska-Walasek M. Biochemical and DNA markers of X-linked hypophosphataemic rickets: a study of sporadic cases. Journal of inherited metabolic disease. 1992;15(3):335-8. | duplicate |
| Puente-Ruiz N, Docio P, Unzueta MTG, Lavín BA, Maiztegi A, Vega AI, et al. Uncovering genetic causes of hypophosphatemia. J Intern Med. 2023;293(6):753-62. | >10% adults and not reported separately |

| | |
|---|---|
| Rafaelsen S, Raeder H, Johansson S, Bjerknes R. Hypophosphatemic rickets in norwegian children: Genotypes, phenotypes, and complications. Hormone Research in Paediatrics. 2014;82(SUPPL. 1):185. | publication type (inc. abstract only) |
| Rodriguez-Rubio E, Gil-Pena H, Chocron S, Madariaga L, de la Cerda-Ojeda F, Fernandez-Fernandez M, et al. Phenotypic characterization of X-linked hypophosphatemia in pediatric Spanish population. Orphanet journal of rare diseases. 2021;16(1):104. | no sequencing |
| Rojek A, Obara-Moszynska M, Niedziela M. Growth hormone treatment of 2 patients with x-linked hypophosphatemic rickets caused by PHEX mutation: Effects on linear growth. Hormone Research in Paediatrics. 2018;90(Supplement 1):186. | publication type (inc. abstract only) |
| Ruppe M. Clinical regulators of FGF23 in X-linked hypophosphatemic rickets: A cohort level analysis. Journal of Bone and Mineral Research. 2010;25(SUPPL. 1):S230. | publication type (inc. abstract only) |
| Ruppe MD, Brosnan PG, Au KS, Tran PX, Dominguez BW, Northrup H. Mutational analysis of PHEX, FGF23 and DMP1 in a cohort of patients with hypophosphatemic rickets. Clinical endocrinology. 2011;74(3):312-8. | publication type (inc. abstract only) |
| Ruppe MD, Brosnan PG, Au KS, Tran PX, Dominguez BW, Northrup H. Mutational analysis of PHEX, FGF23 and DMP1 in a cohort of patients with hypophosphatemic rickets. Clinical endocrinology. 2011;74(3):312-8. | unclear age |
| Rush ET, Johnson B, Aradhya S, Beltran D, Bristow SL, Eisenbeis S, et al. Molecular Diagnoses of X-Linked and Other Genetic Hypophosphatemias: Results From a Sponsored Genetic Testing Program. Journal of bone and mineral research : the official journal of the American Society for Bone and Mineral Research. 2022;37(2):202-14. | unclear age? |
| Samartino A, Azevedo E, Peixinho J, Marcatto F, Silva I, Nunes J, et al. Mutational analysis and genotype-phenotype correlation of the PHEX gene in Brazilian patients with X-linked hypophosphatemic rickets. Hormone Research in Paediatrics. 2021;94(SUPPL 1):212. | publication type (inc. abstract only) |
| Sato K, Tajima T, Nakae J, Adachi M, Asakura Y, Tachibana K, et al. Three novel PHEX gene mutations in Japanese patients with X-linked hypophosphatemic rickets. Pediatric research. 2000;48(4):536-40. | unclear age |
| Schutt SM, Schumacher M, Holterhus PM, Felgenhauer S, Hiort O. Effect of GH replacement therapy in two male siblings with | publication type (inc. abstract only) |

| | |
|---|---|
| combined X-linked hypophosphatemia and partial GH deficiency. European journal of endocrinology. 2003;149(4):317-21. | |
| Silva JM, Alexandre AR, Matsuura CO, Rodrigues TB, Pacheco NSS, Oliveira RS, et al. Four novel PHEX gene mutations in patients with X-linked hypophosphatemia. Hormone Research in Paediatrics. 2019;92(Supplement 1):43. | publication type (inc. abstract only) |
| Smith PS, Gottesman GS, Zhang F, Cook F, Ramirez B, Wenkert D, et al. X-Linked Hypophosphatemia: Uniquely Mild Disease Associated With PHEX 3'-UTR Mutation c.*231A>G (A Retrospective Case-Control Study). Journal of bone and mineral research : the official journal of the American Society for Bone and Mineral Research. 2020;35(5):920-31. | >10% adults and not reported separately |
| Smith PS, Gottesman GS, Zhang F, McAlister WH, Cook F, Wollberg V, et al. X-Linked Hypophosphatemia: PHEX 3'UTR c. *231A>G causes a uniquely mild phenotype including three large american kindreds (a retrospective, case-control study). Journal of Bone and Mineral Research. 2018;33(Supplement 1):425-6. | publication type (inc. abstract only) |
| Sun Y, Xia W, Li M, Jiang Y, Wang O, Nie M, et al. Genetic analysis of 128 Chinese patients with hypophosphatemic rickets. Bone. 2010;47(SUPPL. 3):S449. | publication type (inc. abstract only) |
| Tavana N, Ting TH, Lai K, Kennerson ML, Thilakavathy K. Whole exome sequencing identifies two novel variants in PHEX and DMP1 in Malaysian children with hypophosphatemic rickets. Italian journal of pediatrics. 2022;48(1):193. | duplicate |
| Thiele S, Stubbe A, Werner R, Hiort O, Hoeppner W. Establishing of a novel NGS tool for the diagnosis of X-linked hypophosphatemia (XLH). Hormone Research in Paediatrics. 2019;91(Supplement 1):120. | publication type (inc. abstract only) |
| Thiele S, Werner R, Stubbe A, Hiort O, Hoeppner W. Validation of a next-generation sequencing (NGS) panel to improve the diagnosis of X-linked hypophosphataemia (XLH) and other genetic disorders of renal phosphate wasting. European journal of endocrinology. 2020;183(5):497-504. | publication type (inc. abstract only) |
| Whyte MP, Schranck FW, Armamento-Villareal R. X-linked hypophosphatemia: a search for gender, race, anticipation, or parent of origin effects on disease expression in children. The Journal of clinical endocrinology and metabolism. 1996;81(11):4075-80. | no sequencing |
| Winters RW, Graham JB. Multiple genetic mechanisms in vitamin D-resistant rickets. Pediatrics. 1960;25:932-4. | publication type (inc. abstract only) |
| Winters RW, Graham JB, Williams TF, McFalls VW, Burnett CH. A genetic study of familial hypophosphatemia and vitamin D | no sequencing |

| | |
|---|---|
| resistant rickets. Transactions of the Association of American Physicians. 1957;70:234-42. | |
| Winters RW, Graham JB, Williams TF, McFalls VW, Burnett CH. A genetic study of familial hypophosphatemia and vitamin D resistant rickets with a review of the literature. Medicine. 1958;37(2):97-142. | no sequencing |
| Winters RW, Graham JB, Williams TF, McFalls VW, Burnett CH. A genetic study of familial hypophosphatemia and vitamin D resistant rickets with a review of the literature. 1958. Medicine. 1991;70(3):215-7. | no sequencing |
| Winters RW, McFalls VF, Graham JB. Genetic studies of vitamin D resistant rickets and familial hypophosphatemia. Helvetica paediatrica acta. 1959;14:533-8. | no sequencing |
| Xia WB, Sun Y, He XD, Jiang Y, Nie M, Li M, et al. Clinical features and gene mutation analysis of patients with hypophosphatemic rickets. Bone. 2010;47(SUPPL. 3):S377. | publication type (inc. abstract only) |
| Xiong F, Ran Q. Two novel PHEX gene mutations in chinese patients with X-linked hypophosphatemic rickets. Hormone Research in Paediatrics. 2017;88(Supplement 1):264. | publication type (inc. abstract only) |
| Yue H, Yu J-b, He J-w, Zhang Z, Fu W-z, Zhang H, et al. Identification of two novel mutations in the PHEX gene in Chinese patients with hypophosphatemic rickets/osteomalacia. PloS one. 2014;9(5):e97830. | duplicate |
| Zhang A, Zheng B, Wang C, Zhang Y, Ding G, Huang S, et al. Genetic and functional characterization of PHEX gene variants in 42 children with X-linked hypophosphatemic rickets. Journal of the American Society of Nephrology. 2018;29:309. | publication type (inc. abstract only) |
| Zhang C, Zhao Z, Sun Y, Xu L, JiaJue R, Cui L, et al. Clinical and genetic analysis in a large Chinese cohort of patients with X-linked hypophosphatemia. Bone. 2019;121:212-20. | unclear age |

#### fHLH

| **Reference** | **Reason for exclusion (Q2)** |
|---|---|
| Abbas AAH, Tapp HE, Rice MS, Mangos HM. Familial haemophagocytic lymphohistocytosis: A review of six cases. HAEMA. 2004;7(2):233-9. | FT not retrievable |
| Abdel-Mannan O, Absoud M, Benetou C, Hickson H, Chong WK, Hemingway C, et al. TEN YEAR FOLLOW-UP SURVEILLANCE OF PAEDIATRIC ACQUIRED DEMYELINATING SYNDROMES (ADS) IN | publication type (inc. abstract only) |

| | |
|---|---|
| THE UK. Journal of Neurology, Neurosurgery and Psychiatry. 2022;93(6):127. | |
| Abdel-Mannan O, Absoud M, Benetou C, Hickson H, Hemingway C, Lim M, et al. Ten year follow-up surveillance of paediatric acquired demyelinating syndromes (ADS) in the United Kingdom. Multiple Sclerosis Journal. 2021;27(2 SUPPL):44-5. | wrong condition |
| Abdel-Mannan O, Absoud M, Benetou C, Hickson H, Hemingway C, Lim M, et al. Incidence of paediatric multiple sclerosis and other acquired demyelinating syndromes: 10-year follow-up surveillance study. Developmental medicine and child neurology. 2022;64(4):502-8. | publication type (inc. abstract only) |
| Abdel-Mannan O, Absoud M, Benetou C, Hickson H, Hemingway C, Lim M, et al. Incidence of paediatric multiple sclerosis and other relapsing demyelination conditions: 10-year follow-up UK surveillance of paediatric acquired demyelinating syndromes (ADS). Developmental Medicine and Child Neurology. 2022;64(SUPPL 1):8. | publication type (inc. abstract only) |
| Abdel-Mannan O, Parida A, Foster K, Ramdas S, Ram D, Eleftheriou D, et al. Isolated CNS familial hemophagocytic lymphohistiocytosis (HLH) in children presenting as a mimic of demyelination in children. Multiple Sclerosis Journal. 2021;27(2 SUPPL):136. | publication type (inc. abstract only) |
| Abolhassani H, Kiaee F, Tavakol M, Chavoshzadeh Z, Mahdaviani SA, Momen T, et al. Fourth Update on the Iranian National Registry of Primary Immunodeficiencies: Integration of Molecular Diagnosis. Journal of Clinical Immunology. 2018;38(7):816-32. | wrong condition |
| Agarwal S, Radhakrishnan N, Dinand V, Chinnabhandar V, Sachdeva A. Hemophagocytic lymphohistiocytosis in children: A tertiary care center experience from India. Indian Journal of Hematology and Blood Transfusion. 2014;30(2 SUPPL. 1):479. | publication type (inc. abstract only) |
| Agarwal S, Radhakrishnan N, Thakkar D, Dinand V, Gupta A, Chinnabhandar V, et al. Hemophagocytic lymphohistiocytosis in children: A tertiary care center experience from india. Pediatric Blood and Cancer. 2014;61(SUPPL. 2):S226. | publication type (inc. abstract only) |
| Agarwal S, Rani S, Ahmed H, Rauthan A, Modumudi P. Hemophagocytic lymphohistiocytosis syndrome : A tertiary care centre experience from South India. Pediatric Blood and Cancer. 2016;63(Supplement 3):S160. | publication type (inc. abstract only) |
| Ahuja A, Chatterjee T, Kapoor R, Sharma A, Singh K. Variable spectrum of HLH presentation with two different outcomes. | publication type (inc. abstract only) |

| | |
|---|---|
| Indian Journal of Hematology and Blood Transfusion. 2019;35(1 Supplement):S85-S6. | |
| Akcay N, Kihtir HS, Kocoglu Barlas U, Sevketoglu E. The effect of plasmapheresis on mortality in patients monitored in the pediatric intensive care unit due to hemophagocytic lymphohistiocytosis: Single center experience. Critical Care. 2020;24(SUPPL 2). | publication type (inc. abstract only) |
| Aksu Uzunhan T, Caliskan M, Karaman S, Aydin K, Devecioglu O. A rare cause of acute cerebellar ataxia: familial hemophagocytic lymphohistiocytosis. Pediatric neurology. 2014;51(3):465-6. | publication type (inc. abstract only) |
| Akyol S, Ozcan A, Sekine T, Chiang SC, Yilmaz E, Karakurkcu M, et al. Different clinical presentation of 3 patients with familial hemophagocytic lymphohistiocytosis with two novel mutations. HemaSphere. 2018;2(Supplement 2):901. | publication type (inc. abstract only) |
| Al-Ahmari A, Sheereen A, Elamin T, Jabr A, Al-Awwami M, Alsmadi O, et al. Molecular changes in Saudi patients with familial hemophagocytic lymphohistiocytosis. Pediatric Blood and Cancer. 2014;61(SUPPL. 2):S108. | publication type (inc. abstract only) |
| Algeri M, Slatter M, Qasim W, Bertaina V, Pagliara D, Galaverna F, et al. Outcomes of children with primary immunodeficiencies receiving alpha/beta T-cell depleted HLA-haplo-HSCT followed by infusion of lymphocytes transduced with the inducible caspase 9 (iC9) suicide gene. Bone Marrow Transplantation. 2019;53:87-8. | publication type (inc. abstract only) |
| Alhumaidan W, Al-Otaibi A, Elyamany G, Alabbas F, Ali TB. Familial hemophagocytic lymphohistiocytosis induced by SARS-CoV-2. Pediatric hematology and oncology. 2021;38(4):406-9. | publication type (inc. abstract only) |
| Allen C, Chandrakasan S, Jordan M, Leiding J, Oladapo A, Pednekar P, et al. EMAPALUMAB TREATMENT PATTERNS/OUTCOMES IN PRIMARY HEMOPHAGOCYTIC LYMPHOHISTIOCYTOSIS: REAL-HLH STUDY. Pediatric Blood and Cancer. 2023;70(Supplement 3). | publication type (inc. abstract only) |
| Allen M, De Fusco C, Legrand F, Clementi R, Conter V, Danesino C, et al. Familial hemophagocytic lymphohistiocytosis: how late can the onset be? Haematologica. 2001;86(5):499-503. | No direct sequencing |
| Almarzooqi F, Souid A-K, Vijayan R, Al-Hammadi S. Novel genetic variants of inborn errors of immunity. PloS one. 2021;16(1):e0245888. | wrong condition |
| Alsaedi H, Lazarchik J, Jaroscak J. Hematopoietic stem cell transplant for familial hemophagocytic lymphohistiocytosis | publication type (inc. abstract only) |

| | |
|---|---|
| (HLH) in pediatric patients at the medical university of South Carolina. Pediatric Blood and Cancer. 2014;61(SUPPL. 1):S101. | |
| Alzahrani A, Elyamany G, Elfaraidi H, Othman N, Shaker A, Al-Batniji F, et al. Hemophagocytic syndromes in saudi children: Single center experience. Haematologica. 2015;100(SUPPL. 1):559-60. | publication type (inc. abstract only) |
| Amirifar P, Ranjouri MR, Abolhassani H, Moeini Shad T, Almasi-Hashiani A, Azizi G, et al. Clinical, Immunological and Genetic findings in Patients with UNC13D Deficiency (FHL3): a Systematic Review. Pediatric allergy and immunology : official publication of the European Society of Pediatric Allergy and Immunology. 2020. | duplicate |
| Arico M, Janka G, Fischer A, Henter JI, Blanche S, Elinder G, et al. Hemophagocytic lymphohistiocytosis. Report of 122 children from the International Registry. FHL Study Group of the Histiocyte Society. Leukemia. 1996;10(2):197-203. | not genes of interest |
| Ashok V, Kent P, Tamulonis K, Vavra K. To Transplant or Not to Transplant? Late-Onset Primary HLH in a Patient: A Case Report and Review of Literature. Journal of pediatric hematology/oncology. 2019;41(6):482-8. | publication type (inc. abstract only) |
| Astigarraga I, Garcia-Obregon S, Dapena JL, Mateos E, Echebarria A, Navajas A, et al. Genetic studies and familial predisposition to lymphohistiocytosis hemophagocytic in Spain. Pediatric Blood and Cancer. 2010;55(5):880. | publication type (inc. abstract only) |
| Aytac S, Balta G, Kuskonmaz B, Aksu T, Okur FV, Unal S, et al. Isolated cns hemophagocytic lymphohistiocytosis in children:what we know, what we don't know ? Blood. 2021;138(SUPPL 1):4190. | publication type (inc. abstract only) |
| Beken B, Aytac S, Kuskonmaz B, Balta G, Cetin M, Gurgey A, et al. Clinical and laboratory evaluation of patients with familial hemophagocytic lymphohistiocytosis in a single center; Striking the importance of central nervous system and hepatic involvement. Blood. 2014;124(21). | publication type (inc. abstract only) |
| Blincoe A, Heeg M, Campbell P, Khojah A, Klein-Gitelman M, Talano JA, et al. Isolated central nervous system disease in familial hemophagocytic lymphohistiocytosis a multicenter case series. Journal of Clinical Immunology. 2019;39(Supplement 1):S112-S3. | publication type (inc. abstract only) |
| Bloch C, Jais JP, Gil M, Boubaya M, Suarez F, Bader-Meunier B, et al. Genetic of Sporadic Hemophagocytic Lymphohistiocytosis. Blood. 2019;134(Supplement 1):82. | publication type (inc. abstract only) |

| | |
|---|---|
| Bock AM, Phillippi L, LeVeque M, Casper J, Thakar M, Margolis D, et al. The successful treatment of recurrent CNS disease post-HCT in patients with hemophagocytic lymphohistiocytosis. Pediatric Blood and Cancer. 2015;62(Supplement 2):S118. | publication type (inc. abstract only) |
| Borte S, Meeths M, Liebscher I, Krist K, Nordenskjold M, Hammarstrom L, et al. Combined newborn screening for familial hemophagocytic lymphohistiocytosis and severe T- and B-cell immunodeficiencies. The Journal of allergy and clinical immunology. 2014;134(1):226-8. | publication type (inc. abstract only) |
| Borte S, Meeths M, Nordenskjold M, Hammarstrom L, Henter JI, Von Dobeln U, et al. Combined newborn screening for familial hemophagocytic lymphohistiocytosis and severe T and B cell immunodeficiencies. Journal of Clinical Immunology. 2014;34(2 SUPPL. 1):S185. | publication type (inc. abstract only) |
| Botto LD, Meeths M, Campos-Xavier B, Bergamaschi R, Mazzanti L, Scarano E, et al. Chondrodysplasia and growth failure in children after early hematopoietic stem cell transplantation for non-oncologic disorders. American journal of medical genetics Part A. 2021;185(2):517-27. | wrong condition |
| Bujan W, Schandene L, Ferster A, De Valck C, Goldman M, Sariban E. Abnormal T-cell phenotype in familial erythrophagocytic lymphohistiocytosis. Lancet (London, England). 1993;342(8882):1296. | publication type (inc. abstract only) |
| Busiello R, Fimiani G, Miano MG, Arico M, Santoro A, Ursini MV, et al. A91V perforin variation in healthy subjects and FHLH patients. International journal of immunogenetics. 2006;33(2):123-5. | no information about age |
| Busiello R, Galgani M, De Fusco C, Poggi V, Adriani M, Racioppi L, et al. Role of A91V mutation in perforin gene in hemophagocytic lymphohistiocytosis. Blood. 2004;104(6):1910-. | publication type (inc. abstract only) |
| Caselli D, Sieni E, Gambineri E, Cetica V, Brugnolo F, Aric o M. Role of serotherapy for SCT in familial haemophagocytic lymphohistiocytosis. Bone Marrow Transplantation. 2012;47(SUPPL. 1):S386. | publication type (inc. abstract only) |
| Cetica V, Pende D, Griffiths GM, Arico M. Molecular basis of familial hemophagocytic lymphohistiocytosis. Haematologica. 2010;95(4):538-41. | publication type (inc. abstract only) |
| Chandra S, Filipovich A, Jordan M. Lymphopenia in patients with hemophagocytic lymphohistiocytosis: Are B cells suppressed in these patients? Biology of Blood and Marrow Transplantation. 2014;20(2 SUPPL. 1):S167-S8. | publication type (inc. abstract only) |

| | |
|---|---|
| Chang TY, Jaffray J, Woda B, Newburger PE, Usmani GN. Hemophagocytic lymphohistiocytosis with MUNC13-4 gene mutation or reduced natural killer cell function prior to onset of childhood leukemia. Pediatric blood & cancer. 2011;56(5):856-8. | wrong condition |
| Chinn IK, Eckstein OS, Peckham-Gregory EC, Goldberg BR, Forbes LR, Nicholas SK, et al. Genetic and mechanistic diversity in hemophagocytic lymphohistiocytosis. Journal of Allergy and Clinical Immunology. 2018;141(2 Supplement 1):AB19. | publication type (inc. abstract only) |
| Chinn IK, Goldberg BR, Forbes LR, Nicholas SK, Mace EM, Abhyankar HA, et al. Heterogeneity of molecular diagnoses and outcomes in a diverse cohort of patients with hemophagocytic lymphohistiocytosis. Pediatric Blood and Cancer. 2016;63(Supplement 2):S39-S40. | publication type (inc. abstract only) |
| Clementi R, Zur Stadt U, Savoldi G, Varotto S, Conter V, De Fusco C, et al. Six novel mutations in the PRF1 gene in children with haemophagocytic lymphohistiocytosis [10]. Journal of Medical Genetics. 2001;38(9):643-6. | duplicate |
| Coniglio ML, Da Ros M, Balasco D, Brizzi V, Dell'Acqua F, De Fusco C, et al. Flow cytometric perforin and cd107a screening for the diagnosis of familial hemophagocytic lymphohistiocytosis: Report from the Italian registry. Pediatric Blood and Cancer. 2020;67(SUPPL 1). | publication type (inc. abstract only) |
| Cui TT, Cui T, Wang Y, Wang J, Zhang J, Gao Z, et al. The clinical features and outcome of familial hemophagocytic lyphohistocytosis-2 : An experience from a single center. Pediatric Blood and Cancer. 2020;67(SUPPL 1). | publication type (inc. abstract only) |
| Delaney MM, Shafford EA, Al-Attar A, Pritchard J. Familial erythrophagocytic reticulosis. Complete response to combination chemotherapy. Archives of disease in childhood. 1984;59(2):173-5. | No direct sequencing |
| Delaney MM, Shafford EA, Alattar A, Pritchard J. FAMILIAL ERYTHROPHAGOCYTIC RETICULOSIS - COMPLETE RESPONSE TO COMBINATION CHEMOTHERAPY. Archives of Disease in Childhood. 1984;59(2):173-5. | duplicate |
| Drabko K, Salomonowicz M, Pieczonka A, Gozdzik J, Malinowska I, Kalwak K, et al. Hematopoietic stem cell transplantation in children with hemophagocytic lymfohistiocytosis (HLH)-report of polish pediatric stem cell transplantation group. Bone Marrow Transplantation. 2016;51(SUPPL. 1):S452. | publication type (inc. abstract only) |
| Du Y, Li L, Wang XY, Chen L, Jia SH, Mi R. Familial Hemophagocytic Lymphohistiocytosis Type 3. Indian journal of pediatrics. 2020;87(10):861. | publication type (inc. abstract only) |

| | |
|---|---|
| Dufourcq-Lagelouse R, Jabado N, Le Deist F, Stephan JL, Souillet G, Bruin M, et al. Linkage of familial hemophagocytic lymphohistiocytosis to 10q21-22 and evidence for heterogeneity. American journal of human genetics. 1999;64(1):172-9. | off topic |
| Duvall AS, Frame DG, Briones M, Ebens CL, Grimley MS, Joshi SA, et al. Neonatal hemophagocytic lymphohistiocytosis: A multi-center case series highlighting the challenges in diagnosis and management including the utility of hematopoietic stem cell transplant. Pediatric Blood and Cancer. 2017;64(Supplement 1):S97. | publication type (inc. abstract only) |
| DuVall AS, Palka C, Frame DG, Briones M, Ebens CL, Grimley MS, et al. Neonatal hemophagocytic lymphohistiocytosis: A multi-center case series highlighting the unique phenotypic presentation and challenges in diagnosis and management. Journal of Clinical Immunology. 2016;36(3):310. | publication type (inc. abstract only) |
| Ebens CL, DuVall AS, DeFor TE, Dimitrov MT, Frame D, Briones MA, et al. A Multi-Center Case Series, Systematic Review and Meta-Analysis of Neonatal Hemophagocytic Lymphohistiocytosis. Blood. 2020;136(Supplement 1):19-20. | publication type (inc. abstract only) |
| Eckstein OS, Chinn IK, Peckham-Gregory EC, Goldberg BR, Forbes LR, Nicholas SK, et al. Genetic and mechanistic diversity in pediatric hemophagocytic lymphohisitocytosis. Pediatric Blood and Cancer. 2019;66(Supplement 1). | publication type (inc. abstract only) |
| El Chazli Y, El-Maksoud MA, Elsharkawy A, Shoman W, Mikhael N, El-beheiry A. Hemophagocytic lymphohistiocytosis: A missed diagnosis in children with neurological manifestations. Pediatric Blood and Cancer. 2020;67(SUPPL 1). | duplicate |
| El Chazli Y, Elsharkawy A, El-Maksoud MA, Shoman W, Mikhael N. Hemophagocytic lymphohistiocytosis in egyptian children: Single center experience. Pediatric Blood and Cancer. 2020;67(SUPPL 1). | publication type (inc. abstract only) |
| Elsayed S, Elsobky E, Ragab E, Grandin V, Lambert N, De Saint Basile G. Molecular characterization and prenatal diagnosis of familial hemophagocytic lymophohistocytosis in Egyptians. Haematologica. 2013;98(SUPPL. 1):678. | publication type (inc. abstract only) |
| Elshinawy M, Al Rawas A, Wali Y. Familial hemophagocytic lymphohistiocytosis in Oman: An update on unique clinical and molecular features. Haematologica. 2016;101(Supplement 1):583. | publication type (inc. abstract only) |
| Elshinawy M, El-Beshlawi I, Al Nuaimi M, Al Dhuhli A, Alrawas A, Wali Y. Familial hemophagocytic lymphohistiocytosis in Oman: | publication type (inc. abstract only) |

| | |
|---|---|
| From non-existence to molecular characterization. Pediatric Blood and Cancer. 2012;58(7):1045. | |
| Elshinawy M, Wali Y. Familial hemophagocytic lymphohistiocytosis in Oman: An update on unique clinical and molecular features. European Journal of Pediatrics. 2016;175(11):1543-4. | publication type (inc. abstract only) |
| Entesarian M, Chiang SCC, Schlums H, Meeths M, Chan M-Y, Mya S-N, et al. Novel deep intronic and missense UNC13D mutations in familial haemophagocytic lymphohistiocytosis type 3. British journal of haematology. 2013;162(3):415-8. | publication type (inc. abstract only) |
| Entesarian M, Meeths M, Rudd E, Nordenskjold M, Henter JI. Frequency of PRF1, STX11 and UNC13D mutations in patients with a genetic diagnosis of familial hemophagocytic lymphohistiocytosis. Pediatric Blood and Cancer. 2011;56(4):693. | publication type (inc. abstract only) |
| Essa MF, Abujoub R, Elbashir E, Alsudairy R, Alomari A, Alsultan A. HLA-matched HSCT using targeted busulfan-based conditioning in children with primary hemophagocytic lymphohistiocytosis. Bone marrow transplantation. 2021;56(12):3097-9. | publication type (inc. abstract only) |
| Fatima Z, Khan A, Tariq U, Sohail MS. Hemophagocytic Lymphohistiocytosis: A Case Series. Cureus. 2018;10(4):e2545. | >=10% adults + not reported separately |
| Felber M, Steward C, Hauri Hohl M, Kentouche K, Fasth A, Pachlopnik Schmid J, et al. Excellent results after RIC-conditioning and T-cell replete HLA-identical transplants including cord blood in hemophagocytic lymphohistiosis (HLH). Bone Marrow Transplantation. 2016;51(SUPPL. 1):S47. | publication type (inc. abstract only) |
| Feng W-X, Yang X-Y, Li J-W, Gong S, Wu Y, Zhang W-H, et al. Neurologic Manifestations as Initial Clinical Presentation of Familial Hemophagocytic Lymphohistiocytosis Type2 Due to PRF1 Mutation in Chinese Pediatric Patients. Frontiers in genetics. 2020;11:126. | duplicate |
| Filipovich AH. Life-threatening hemophagocytic syndromes: current outcomes with hematopoietic stem cell transplantation. Pediatric transplantation. 2005;9 Suppl 7:87-91. | publication type (inc. abstract only) |
| Fiorillo A, Catera P, Guarino A, Grimaldi M, Menna G, Migliorati R, et al. Familial erythrophagocytic lymphohistiocytosis: adverse prognostic significance of delayed diagnosis. Haematologica. 1993;78(4):242-4. | No direct sequencing |
| Fiorillo A, Catera P, Guarino A, Grimaldi M, Menna G, Migliorati R, et al. FAMILIAL ERYTHROPHAGOCYTIC | duplicate |

| | |
|---|---|
| LYMPHOHISTIOCYTOSIS - ADVERSE PROGNOSTIC-SIGNIFICANCE OF DELAYED DIAGNOSIS. Haematologica. 1993;78(4):242-4. | |
| Fischer A, Cerf-Bensussan N, Blanche S, Le Deist F, Bremard-Oury C, Leverger G, et al. Allogeneic bone marrow transplantation for erythrophagocytic lymphohistiocytosis. The Journal of pediatrics. 1986;108(2):267-70. | publication type (inc. abstract only) |
| Gadoury-Levesque V, Dong L, Su R, Chen J, Risma K, Marsh R, et al. Frequency and variant spectrum in 227 patients with molecular diagnosis of hemophagocytic lymphohistiocytosis. Allergy, Asthma and Clinical Immunology. 2020;16(SUPPL 1). | publication type (inc. abstract only) |
| Gadoury-Levesque V, Dong L, Su R, Chen J, Risma K, Marsh R, et al. Frequency and variant spectrum in 227 patients with molecular diagnosis of hemophagocytic lymphohistiocytosis. Allergy, Asthma and Clinical Immunology. 2020;16(SUPPL 1). | >=10% adults + not reported separately |
| Gepte MB, Del Rosario ML, Rigor E. HEMOPHAGOCYTIC LYMPHOHISTIOCYTOSIS: CLINICAL REVIEW AT THE PHILIPPINE CHILDREN'S MEDICAL CENTER. Pediatric Blood and Cancer. 2023;70(Supplement 1). | publication type (inc. abstract only) |
| Giardino S, Faraci M, Morreale G, Bagnasco F, Micalizzi C, Marcenaro S, et al. Allogeneic hematopoietic stem cell transplantation in congenital hemophagocytic lymphohistiocytosis: 17-years single pediatric centre experience. Bone Marrow Transplantation. 2014;49(SUPPL. 1):S371. | publication type (inc. abstract only) |
| Gilbert EF, ZuRhein GM, Wester SM, Herrmann J, Hong R, Opitz JM. Familial hemophagocytic lymphohistiocytosis: report of four cases in two families and review of the literature. Pediatric pathology. 1985;3(1):59-92. | No direct sequencing |
| Graham GE, Graham LM, Bridge PJ, Maclaren LD, Wolff JEA, Coppes MJ, et al. Further evidence for genetic heterogeneity in familial hemophagocytic lymphohistiocytosis (FHLH). Pediatric research. 2000;48(2):227-32. | off topic |
| Greenmyer JR, Thompson WS, Mavis S, Hassan S, Weckwerth J, Hobbs C, et al. Neonatal familial hemophagocytic lymphohistiocytosis diagnosed with ultrarapid whole-genome sequencing. Pediatric blood & cancer. 2023;70(1):e29871. | publication type (inc. abstract only) |
| Gulati N, Eckstein O, Forbes L, Peckham-Gregory E, Ozuah N, Kamdar K, et al. Genomic characterization of a pediatric cohort with non-malignant lymphoproliferative disorders. Pediatric Blood and Cancer. 2019;66(Supplement 2):S70-S1. | publication type (inc. abstract only) |

| | |
|---|---|
| Gutovskaya E, Shekhovtsova Z, Shelikhova L, Shipitsina I, Balashov D, Shasheleva D, et al. Graft depletion of TCR as/CD19 in matched unrelated and haploidentical transplantation after treosulfan-based conditioning regimen for inherited hemophagocytic lymphohistiocytosis. Bone Marrow Transplantation. 2016;51(SUPPL. 1):S263-S4. | publication type (inc. abstract only) |
| Hadzic N, Molnar E, Height S, Kovacs G, Dhawan A, Andrikovics H, et al. High Prevalence of Hemophagocytic Lymphohistiocytosis in Acute Liver Failure of Infancy. The Journal of pediatrics. 2022;250:67-74.e1. | publication type (inc. abstract only) |
| Hazzazi K, Al-Saud B, Mohammed R. Two Unrelated Cases of Familial Hemophagocytic Lymphohistocytosis Presented as Severe Combined Immunodeficiency. Journal of Clinical Immunology. 2021;41(SUPPL 1):S122-S3. | publication type (inc. abstract only) |
| Henry MM, Julien D, Williams JA, Jacobsen J, Ngwube A. Isolated central nervous system hemophagocytic lymphohistiocytosis: A case series. Pediatric Blood and Cancer. 2019;66(Supplement 1). | publication type (inc. abstract only) |
| Henter JI, Elinder G. Familial hemophagocytic lymphohistiocytosis. Clinical review based on the findings in seven children. Acta paediatrica Scandinavica. 1991;80(3):269-77. | No direct sequencing |
| Henter JI, Elinder G, Finkel Y, Soder O. Successful induction with chemotherapy including teniposide in familial erythrophagocytic lymphohistiocytosis. Lancet (London, England). 1986;2(8520):1402. | publication type (inc. abstract only) |
| Henter J-I, von Bahr Greenwood T, Bergsten E. Emapalumab in Primary Hemophagocytic Lymphohistiocytosis. The New England journal of medicine. 2020;383(6):596-8. | publication type (inc. abstract only) |
| Herman TE, Siegel MJ. Familial hemophagocytic lymphohistiocytosis. Journal of perinatology : official journal of the California Perinatal Association. 2010;30(5):363-5. | publication type (inc. abstract only) |
| Hernandez-Sanchez A, Cerezo-Martin JM, Garcia-Bacelar A, Caballero-Alvarez D, Azibeiro R, Pablos A, et al. ABCL-297 Hemophagocytic Lymphohistiocytosis Singularity: A Multicenter Retrospective Study. Clinical Lymphoma, Myeloma and Leukemia. 2022;22(Supplement 2):S369-S70. | publication type (inc. abstract only) |
| Hofer-Popow I, Dworzak M, Hutter C, Mann G, Holter W, Matthes-Leodolter S, et al. Allogeneic stem cell transplantation with reduced intensity regimen as successful therapy for primary haemophagocytic lymphohistiocytosis: A single center | publication type (inc. abstract only) |

| | |
|---|---|
| experience. Oncology Research and Treatment. 2018;41(Supplement 4):62. | |
| Houshmand M, Pourpak Z, Safaei S, Badalzadeh B, Alizadeh Z, Fazlollahi MR, et al. Genetic diagnosis of primary immunodefeciency in iran. Iranian Journal of Allergy, Asthma and Immunology. 2013;12(1):S26-S7. | publication type (inc. abstract only) |
| Janka GE. Familial hemophagocytic lymphohistiocytosis: therapy in the German experience. Pediatric hematology and oncology. 1989;6(3):227-31. | No direct sequencing |
| Johnson J, Huizenga K, Kissell D, Jordan M, ZurStadt U, Filipovich A, et al. STXBP2 (MUNC18-2) mutations in North American patients with familial hemophagocytic lymphohistiocytosis. Pediatric Blood and Cancer. 2010;54(6):817. | publication type (inc. abstract only) |
| Jordan M, Locatelli F, Allen C, De Benedetti F, Grom AA, Ballabio M, et al. A novel targeted approach to the treatment of hemophagocytic lymphohistiocytosis (HLH) with an anti-interferon gamma (IFNgamma) monoclonal antibody (MAB), NI-0501: First results from a pilot phase 2 study in children with primary HLH. Blood. 2015;126(23):LBA3. | publication type (inc. abstract only) |
| Kamalanathan N, Kumar AA, Korulla A, Fouzia NA, Abraham A, George B, et al. Molecular diagnosis of primary immunodeficiency diseases. Indian Journal of Hematology and Blood Transfusion. 2018;34(1 Supplement):159. | publication type (inc. abstract only) |
| Kang SH, Koh YK, Choi ES, Kim H, Koh KN, Im HJ. EXPERIENCES WITH TREOSULFAN-BASED CONDITIONING FOR HEMOPHAGOCYTIC LYMPHOHISTIOCYTOSIS. Pediatric Blood and Cancer. 2022;69(Supplement 5):S282. | publication type (inc. abstract only) |
| Khanna R, Height S, Gilmour K, Hadzic N. Primary immunodeficiencies in cryptogenic acute liver failure in children: A single centre experience. Hepatology. 2015;62(SUPPL. 1):1048A. | publication type (inc. abstract only) |
| Kini P, Wasekar N, PreethamTauro RS, Asangi S, Khude S, Jadhav S, et al. Familial Hemophagocytic Lymphohistiocytosis (Fhl) - A Case Series. Pediatric Hematology Oncology Journal. 2018;3(3 Supplement):S16. | publication type (inc. abstract only) |
| Klaudel-Dreszler M, Jankowska I, Kaminska D, Socha P. Hemophagocytic lymphohistiocytosis should be considered in differential diagnostics of acute liver failure before making the decision concerning liver transplantation. Transplantation. 2015;99(7 SUPPL. 1):126. | publication type (inc. abstract only) |

| | |
|---|---|
| Klaudel-Dreszler M, Rutynowska-Pronicka O, Socha P, Kowalewska-Majewska E, Baku A, Perek D, et al. Haemophagocytic lymphohistiocytosis - A single centre experience. European Journal of Immunology. 2009;39(SUPPL. 1):S465-S6. | publication type (inc. abstract only) |
| Klaudel-Dreszler M, Rutynowska-Pronicka O, Socha P, Piatosa B, Bakua A, Dadalski M, et al. Acute liver insufficiency caused by haemophagocytic lymphohistiocytosis-experience of children's memorial health institute in Warsaw. Journal of Pediatric Gastroenterology and Nutrition. 2010;50(SUPPL. 2):E148. | publication type (inc. abstract only) |
| Koh KN, Kang SH, Kim H, Im HJ, Suh JK. Favorable Outcome with Durable Chimerism after Allogeneic Hematopoietic Cell Transplantation Using Busulfan and Fludarabine-Based Reduced-Intensity Conditioning for Children with Hemophagocytic Lymphohistiocytosis. Biology of Blood and Marrow Transplantation. 2020;26(3 Supplement):S290. | publication type (inc. abstract only) |
| Koh KN, Suh JK, Chung NG, Cho B, Kang HJ, Shin HY, et al. Characteristics and outcome of pediatric patients with hemophagocytic lymphohistiocytosis in korea. Vox Sanguinis. 2014;107(SUPPL. 1):236. | publication type (inc. abstract only) |
| Kram DE, Santarelli MD, Russell TB, Pearsall KB, Saldana BD. STX11-deficient familial hemophagocytic lymphohistiocytosis type 4 is associated with self-resolving flares and a milder clinical course. Pediatric blood & cancer. 2019;66(9):e27890. | publication type (inc. abstract only) |
| Krivokapic-Dokmanovic L, Krstovski N, Jankovic S, Lazic J, Radlovic N, Janic D. Clinical characteristics and disease course in children with haemophagocytic lymphohistiocytosis treated at the university children's hospital in Belgrade. Srpski Arhiv za Celokupno Lekarstvo. 2012;140(3-4):191-7. | publication type (inc. abstract only) |
| Langer S, Sachdeva D, Kundoo A, Kotwal J, Sachdeva A. Utility of perforin assay in the era of next generation sequencing. Indian Journal of Hematology and Blood Transfusion. 2020;36(1 SUPPL):S187. | publication type (inc. abstract only) |
| Lee SME, Sumegi J, Villanueva J, Tabata Y, Zhang KJ, Chakraborty E, et al. Patients of African ancestry with hemophagocytic lymphohistiocytosis share a common haplotype of <i>PRF1</i> with a 50delt mutation. Journal of Pediatrics. 2006;149(1):134-7. | duplicate |
| Lee SW, Koh KN, Im HJ, Chung NG, Cho B, Kang HJ, et al. Clinical features, genetics and outcome of pediatric patients with hemophagocytic lymphohistiocytosis in Korea: Report of a | publication type (inc. abstract only) |

| | |
|---|---|
| nationwide survey from Korea histiocytosis working party. Haematologica. 2014;99(SUPPL. 1):464. | |
| Lehmann LE, Nagy A. The contribution of cardiac hypertrophy on transplant outcome in patients with HLH undergoing BMT. Biology of Blood and Marrow Transplantation. 2014;20(2 SUPPL. 1):S177-S8. | publication type (inc. abstract only) |
| Lehmberg K, Albert M, Beutel K, Gruhn B, Meisel R, Schulz A, et al. Fludarabin, treosulfan, thiotepa, and alemtuzumab as conditioning regimen for children with haemophagocytic lymphohistiocytosis: Low toxicity and effective disease control. Bone Marrow Transplantation. 2013;48(SUPPL. 2):S23. | publication type (inc. abstract only) |
| Lehmberg K, Ledig S, Wustrau K, Kontny U, Westphal S, Hundsdorfer P, et al. ETOPOSIDE FOR PRIMARY HLH - BETTER THAN ITS REPUTATION. Pediatric Blood and Cancer. 2023;70(Supplement 1). | publication type (inc. abstract only) |
| Li Z, Wang Y, Wang J, Zhang J, Wang Z. Hematopoietic stem cell transplantation for haemophagocytic lymphohistiocytosis: A single-center report of 61 patients. Bone Marrow Transplantation. 2017;52(Supplement 1):449. | publication type (inc. abstract only) |
| Liao C-H, Lee N-C, Jou S-T, Chiang B-L, Yu H-H. UNC13D mutation presenting as fulminant familial hemophagocytic lymphohistiocytosis. Journal of microbiology, immunology, and infection = Wei mian yu gan ran za zhi. 2020;53(6):1039-41. | publication type (inc. abstract only) |
| Locatelli F, Jordan M, Allen C, Cesaro S, Fagioli F, Henry M, et al. Interferon gamma (IFN-gamma) neutralization as a valuable therapeutic target in primary HLH. Haematologica. 2016;101(Supplement 1):114-5. | publication type (inc. abstract only) |
| Lovsin E, Jenko Bizjan B, Zajc Avramovic M, Kovac J, Debeljak M, Avcin T. Whole exome sequencing of MIS-C patients: Analysis of genes involved in inborn errors of type I IFN immunity, hemophagocytic lymphohistiocytosis (HLH), Kawasaki disease (KD) and TLR7 gene. Pediatric Rheumatology. 2021;19(SUPPL 1). | publication type (inc. abstract only) |
| Lu G, Xie Z-d, Shen K-l, Ye L-j, Wu R-h, Liu C-y, et al. Mutations in the perforin gene in children with hemophagocytic lymphohistiocytosis. Chinese medical journal. 2009;122(23):2851-5. | publication type (inc. abstract only) |
| Lucchini G, Marsh RA, Gilmour K, Worth A, Rao A, Booth C, et al. Treatment dilemmas in asymptomatic children with primary haemophagocytic lymphohistiocytosis. Blood. 2017;130(Supplement 1). | publication type (inc. abstract only) |

| | |
|---|---|
| Lucchini G, Marsh R, Gilmour K, Worth A, Nademi Z, Rao A, et al. Treatment dilemmas in asymptomatic children with primary haemophagocytic lymphohistiocytosis. Bone Marrow Transplantation. 2019;53:730. | publication type (inc. abstract only) |
| Ma H, Zhang R, Zhang L, Wei A, Zhao X, Yang Y, et al. Treatment of pediatric primary hemophagocytic lymphohistiocytosis with the HLH-94/2004 regiments and hematopoetic stem cell transplantation in china. Pediatric Blood and Cancer. 2021;68(SUPPL 1). | publication type (inc. abstract only) |
| Madkaikar M, Mhatre S, Desai M, Ghosh K. Spectrum of perforin gene mutations in familial hemophagocytic lymphohistiocytosis 2 (FHL2) patients from India. Journal of Clinical Immunology. 2014;34(2 SUPPL. 1):S217-S8. | publication type (inc. abstract only) |
| Mai H, Li C, Wang X, Liu S, Fang X, Yuan X. Clinical characteristics of childhood hemophagocytic syndrome and analysis of underlying genetic deficiency. Pediatric Blood and Cancer. 2019;66(Supplement 1). | publication type (inc. abstract only) |
| Mai H, Yuan X, Fang X, Wang X, Liu S, Wang Y, et al. Clinical features and outcomes of 140 childhood hemophagocytic lymphohisticocytosis. Pediatric Blood and Cancer. 2016;63(Supplement 3):S161. | publication type (inc. abstract only) |
| Mancebo Sierra E, Aquilino C, Lopez Herradon A, Allende Martinez L, Ruiz Contreras J, Gil J, et al. Clinical and genetic characteristics in a series of haemophagocytic lymphohistiocytosis patients without perforin defects. Inmunologia. | publication type (inc. abstract only) |
| Mancebo Sierra E, Aquilino C, Lopez Herradon A, Allende Martinez L, Ruiz Contreras J, Gil J, et al. Clinical and genetic characteristics in a series of haemophagocytic lymphohistiocytosis patients without perforin defects. Inmunologia. | publication type (inc. abstract only) |
| Marois L, Touzot F, Haddad E, Fernandez I, Morin M-P, De Bruycker JJ, et al. Successful management of familial hemophagocytic lymphohistiocytosis by the JAK 1/2 inhibitor ruxolitinib. Pediatric blood & cancer. 2021;68(6):e28954. | publication type (inc. abstract only) |
| Marsh RA, Satake N, Biroschak J, Jacobs T, Johnson J, Jordan MB, et al. <i>STX11</i> Mutations and Clinical Phenotypes of Familial Hemophagocytic Lymphohistiocytosis in North America. Pediatric Blood & Cancer. 2010;55(1):134-40. | no information about age |
| Maschan M, Gutovskaya E, Balashov D, Raikina E, Zaharova V, Zhogov V, et al. TCR alphabeta-depleted hematopoietic stem cell transplantation in patients with primary hemophagocytic | publication type (inc. abstract only) |

| | |
|---|---|
| lymphohistiocytosis. Pediatric Blood and Cancer. 2016;63(Supplement 2):S49-S50. | |
| McCreary D, Omoyinmi EO, Hong Y, Mulhern C, Papadopoulou C, Al Obaidi M, et al. Development and clinical application of a targeted next generation sequencing gene panel for monogenic autoinflammatory diseases of the CNS. Pediatric Rheumatology. 2019;17(Supplement 1). | publication type (inc. abstract only) |
| McCreary D, Omoyinmi E, Hong Y, Jensen B, Burleigh A, Price-Kuehne F, et al. A rapid turnaround gene panel for severe autoinflammation: Genetic results within 48 hours. Frontiers in immunology. 2022;13:998967. | wrong population |
| Messina C, Zecca M, Fagioli F, Rovelli A, Lanino E, Bertaina A, et al. Hematopoietic stem cell transplantation for hemophagocytic lymphohistiocitosis : A national retrospective analysis of data from the italian association of pediatric hematology oncology (AIEOP). Blood. 2015;126(23):621. | publication type (inc. abstract only) |
| Nagai K, Yamamoto K, Fujiwara H, Jhun A, Ochi T, Suemori K, et al. Real incidence and variation of familial hemophagocytic lymphohistiocytosis in Japan based on genetic and functional analyses of cytotoxic T lymphocytes. Pediatric Blood and Cancer. 2011;56(4):696. | publication type (inc. abstract only) |
| Naik S, Eckstein OS, Sasa G, Krance RA, Allen CE, Heslop HE, et al. Incorporation of Thiotepa in a Reduced Intensity Conditioning Regimen Leads to Improved Engraftment after Stem Cell Transplant for Patients with Hemophagocytic Lymphohistiocytosis. Blood. 2019;134(Supplement 1):3273. | publication type (inc. abstract only) |
| Nazir HF, Ba Alawi FS, Al Hosni S, Al Rawas A, Dennison D. T Cell Depleted Haploidentical Hematopoietic Stem Cell Transplantation for Patients with Familial Hemophagocytic Lymphohistiocytosis Who Do Not Have Matched Family Donors: Experience in Oman. Biology of blood and marrow transplantation : journal of the American Society for Blood and Marrow Transplantation. 2020;26(6):1119-23. | No direct sequencing |
| Nazir HF, Hassanein N, Wali Y, Al Yazidi LS. Outcome of Viral-associated Hemophagocytic Lymphohistiocytosis at a Tertiary Hospital. The Pediatric infectious disease journal. 2022;41(4):330-4. | wrong condition |
| Nespoli L, Locatelli F, Bonetti F, Porta F, Zecca M, Gibardi A, et al. Familial haemophagocytic lymphohistiocytosis treated with allogeneic bone marrow transplantation. Bone marrow transplantation. 1991;7 Suppl 3:139-42. | No direct sequencing |

| | |
|---|---|
| Omoyinmi E, Standing A, Rowczenio D, Keylock A, Gomes SM, Price-Kuehne F, et al. A targeted next-generation sequencing gene panel for autoinflammation. Pediatric Rheumatology. 2017;15(Supplement 1). | publication type (inc. abstract only) |
| Oymak Y, Toret E, Ay Y, Karapinar T, Demirag B, Yaman Y, et al. Hemophagocytic lymphohistiocytosis: Single center experience. Haematologica. 2015;100(SUPPL. 1):299. | publication type (inc. abstract only) |
| Pagel J, Lehmberg K, Koch F, Janka G, Hennies HC, Ehl S, et al. Munc18-2 missense mutations are associated with onset of disease during infancy in FHL-5 patients. Pediatric Blood and Cancer. 2011;56(4):702-3. | publication type (inc. abstract only) |
| Poltavets NV, Maschan MA, Polyakov AV, Novichkova GA, Maschan AA. Mutations in UNC13D gene are the most frequent cause of FHL in a group of Russian patients. Pediatric Blood and Cancer. 2009;53(4):687. | publication type (inc. abstract only) |
| Qian Y, Johnson JA, Connor JA, Valencia CA, Barasa N, Schubert J, et al. The 253-kb inversion and deep intronic mutations in UNC13D are present in North American patients with familial hemophagocytic lymphohistiocytosis 3. Pediatric blood & cancer. 2014;61(6):1034-40. | no information about age |
| Qian YP, Johnson JA, Connor JA, Valencia CA, Barasa N, Schubert J, et al. The 253-kb Inversion and Deep Intronic Mutations in <i>UNC13D</i> are Present in North American Patients With Familial Hemophagocytic Lymphohistiocytosis 3. Pediatric Blood & Cancer. 2014;61(6):1034-40. | duplicate |
| Qian Y, Johnson J, Connor J, Husami A, Bryceson Y, Meeths M, et al. The 253 KB inversion and 118 (-308) C>T intronic mutations in UNC13D are common in patients with familial hemophagocytic lymphohistiocytosis type 3 (FHL3) in North America. Journal of Clinical Immunology. 2012;32(2):406-7. | publication type (inc. abstract only) |
| Qian Y, Johnson J, Connor J, Husami A, Bryceson Y, Meeths M, et al. The 253 KB inversion and 118 (-308) C>T intronic mutations in UNC13D are common in patients with familial hemophagocytic lymphohistiocytosis type 3 (FHL3) in North America. Journal of Clinical Immunology. 2012;32(2):406-7. | publication type (inc. abstract only) |
| Ramzan M, Yadav SP, Chinnabhandar V, Kaur D, Enteserian M, Sachdeva A. Hemophagocytic lymphohistiocytosis (HLH) in infants: A single centre experience. Pediatric Blood and Cancer. 2012;59(6):1045. | publication type (inc. abstract only) |
| Ramzan M, Yadav SP, Chinnabhandar V, Rastogi N, Enteserian M, Sachdeva A. Hemophagocytic lymphohistiocytosis (HLH) in | publication type (inc. abstract only) |

| | |
|---|---|
| infants: A single centre experience. Indian Journal of Hematology and Blood Transfusion. 2011;27(4):251. | |
| Rodrigues J, Cypriano M. Hemophagocytic lymphohistiocytosis: Clinical, laboratory characteristics and outcomes of twenty-one patients in a single institution. Pediatric Blood and Cancer. 2021;68(SUPPL 1). | publication type (inc. abstract only) |
| Rohr J, Beutel K, Maul-Pavicic A, Vraetz T, Thiel J, Warnatz K, et al. Atypical familial hemophagocytic lymphohistiocytosis due to mutations in UNC13D and STXBP2 overlaps with primary immunodeficiency diseases. Haematologica. 2010;95(12):2080-7. | publication type (inc. abstract only) |
| Rudd E, Ericson KG, Zheng C, Uysal Z, Özkan A, Gürgey A, et al. Spectrum and clinical implications of syntaxin 11 gene mutations in familial haemophagocytic lymphohistiocytosis:: association with disease-free remissions and haematopoietic malignancies -: art. no. e14. Journal of Medical Genetics. 2006;43(4):6. | duplicate |
| Rudd E, Meeths M, Uysal Z, Nordenskjold M, Henter J-I, Fadeel B. Multiple inherited sequence variations in two disease-causing genes in familial haemophagocytic lymphohistiocytosis. British journal of haematology. 2009;146(2):218-20. | duplicate |
| Rudd E, Meeths M, Uysal Z, Nordenskjold M, Henter JI, Fadeel B. Multiple inherited sequence variations in two disease-causing genes in familial haemophagocytic lymphohistiocytosis: Correspondence. British Journal of Haematology. 2009;146(2):218-20. | publication type (inc. abstract only) |
| Sadeghi P, Esslami GG, Rokni-Zadeh H, Changi-Ashtiani M, Mohsenipour R. Familial Hemophagocytic Lymphohistiocytosis secondary to UNC13D mutation: a report of two cases. BMC pediatrics. 2022;22(1):667. | publication type (inc. abstract only) |
| Salcioglu Z, Sayilan Sen H, Tugcu D, Akcay A, Aydogan G, Gokce M, et al. Familial hemophagocytic lymphohistiocytosis in children: Single centre results. Pediatric Blood and Cancer. 2015;62(Supplement 4):S301. | publication type (inc. abstract only) |
| Sanchez IP, Leal-Esteban LC, Orrego JC, Franco JL, Coll Y, Serna L, et al. The first two cases of familial haemophagocytic lymphohistiocytosis type II due to PRF1 gene mutations in Colombia. Journal of Clinical Immunology. 2011;31(SUPPL. 1):S23. | publication type (inc. abstract only) |
| Sanchez IP, Leal-Esteban LC, Orrego JC, Franco JL, Coll Y, Serna L, et al. The first two cases of familial haemophagocytic lymphohistiocytosis type II due to PRF1 gene mutations in | no information about age |

| | |
|---|---|
| Colombia. Journal of Clinical Immunology. 2011;31(SUPPL. 1):S23. | |
| Seo JJ. Genotype-phenotype studies on familial hemophagocytic lymphohistiocytosis (FHL) in Korea. Pediatric Blood and Cancer. 2017;64(Supplement 4):S7. | publication type (inc. abstract only) |
| Seo JY, Song J-S, Lee K-O, Won H-H, Kim J-W, Kim S-H, et al. Founder effects in two predominant intronic mutations of UNC13D, c.118-308C>T and c.754-1G>C underlie the unusual predominance of type 3 familial hemophagocytic lymphohistiocytosis (FHL3) in Korea. Annals of hematology. 2013;92(3):357-64. | duplicate |
| Shah N, Wolff JA. Long term management of familial erythrophagocytic lymphohistiocytosis. Pediatric Research. 1973;7(4):360. | publication type (inc. abstract only) |
| Shamsian BS, Rezaei N, Alavi S, Hedayat M, Amin Asnafi A, Pourpak Z, et al. Primary hemophagocytic lymphohistiocytosis in Iran: report from a single referral center. Pediatric hematology and oncology. 2012;29(3):215-9. | not genes of interest |
| Shing MMK, Lee V, Chiang GPK, Leung AWK, Cheng FWT, Li CK. The first reported primary haemophagocytic lymphohistiocytosis (HLH) of chinese children in Hong Kong. Pediatric Blood and Cancer. 2011;57(5):873. | publication type (inc. abstract only) |
| Sieni E, Cetica V, Beutel K, Santoro A, Mastrodicasa E, Meeths M, et al. Genotype-phenotype study of familial haemophagocytic lymphohistiocytosis due to UNC13D mutations. Haematologica. 2010;95(SUPPL. 2):89. | duplicate |
| Sieni E, Cetica V, Pende D, Griffiths G, Santoro A, Ciambotti B, et al. Heterogeneity of familial hemophagocytic lymphohistiocytosis. Haematologica. 2010;95(SUPPL. 2):85. | publication type (inc. abstract only) |
| Sikora KA, Fall N, Barnes MG, Sumegi J, Filipovich AH, Grom AA. Specific subgroup of new-onset systemic juvenile idiopathic arthritis without overt macrophage activation syndrome shares similar gene expression signature with a distinct subset of familial hemophagocytic lymphohistiocytosis. Arthritis and Rheumatism. 2010;62(SUPPL. 10):653. | publication type (inc. abstract only) |
| Stadt UZ, Beutel K, Kolberg S, Schneppenheim R, Kabisch H, Janka G, et al. Mutation spectrum in children with primary hemophagocytic lymphohistiocytosis:: Molecular and functional analyses of <i>PRF1</i>, <i>UNC13D</i>, <i>STX11</i>, and <i>RAB27A</i>. Human Mutation. 2006;27(1):62-8. | duplicate |
| Stark B, Hershko C, Rosen N, Cividalli G, Karsai H, Soffer D. Familial hemophagocytic lymphohistiocytosis (FHLH) in Israel. I. | No direct sequencing |

| | |
|---|---|
| Description of 11 patients of Iranian-Iraqi origin and review of the literature. Cancer. 1984;54(10):2109-21. | |
| Stepp SE, Dufourcq-Lagelouse R, Le Deist F, Bhawan S, Certain S, Mathew PA, et al. Perforin gene defects in familial hemophagocytic lymphohistiocytosis. Science (New York, NY). 1999;286(5446):1957-9. | no information about age |
| Summerlin J, Wells DA, Anderson MK, Halford Z. A Review of Current and Emerging Therapeutic Options for Hemophagocytic Lymphohistiocytosis. The Annals of pharmacotherapy. 2023;57(7):867-79. | publication type (inc. abstract only) |
| Sun Y, Xiao J, Fan S, Liu Z, Jiang F, Chen J. CLINICAL STUDY OF HAPLOIDENTICAL STEM CELL TRANSPLANTATION FOR PRIMARY HEMOPHAGOCYTIC LYMPHOHISTIOCYTOSIS IN CHILDREN. Pediatric Blood and Cancer. 2023;70(Supplement 1). | publication type (inc. abstract only) |
| Sun Y, Xiao J, Fan S, Liu Z, Jiang F, Chen J. CLINICAL STUDY OF HAPLOIDENTICAL STEM CELL TRANSPLANTATION FOR PRIMARY HEMOPHAGOCYTIC LYMPHOHISTIOCYTOSIS IN CHILDREN. Pediatric Blood and Cancer. 2023;70(Supplement 1). | No direct sequencing |
| Tang Y, Chen Y, Wang Z, Feng J. Mutations of PRF1 gene in chinese patients with hemophagocytic lymphohistiocytosis and malignant lymphomas. Pediatric Blood and Cancer. 2013;60(SUPPL. 3):152-3. | publication type (inc. abstract only) |
| Tesi B, Lagerstedt-Robinson K, Chiang SCC, Ben Bdira E, Abboud M, Belen B, et al. Targeted high-throughput sequencing for genetic diagnostics of hemophagocytic lymphohistiocytosis. Genome medicine. 2015;7:130. | >=10% adults + not reported separately |
| Thakkar D, Radhakrishnan N, Kalra M, Gupta A, Dinand V, Sachdeva A. Stem cell transplantation in primary hemophagocytic lymphohistiocytosis: Experience from a tertiary care centre in India. Pediatric Blood and Cancer. 2014;61(SUPPL. 2):S298-S9. | publication type (inc. abstract only) |
| Tong C, Liu H, Xie J, Wang H, Zhang X, Wang F, et al. High frequency of familial hemophagocytic lymphocytosis associated immune gene defects in refractory viral infection of unknown origin. Blood. 2010;116(21). | publication type (inc. abstract only) |
| Ueda I, Kurokawa Y, Koike K, Ito S, Sakata A, Matsumora T, et al. Late-onset cases of familial hemophagocytic lymphohistiocytosis with missense perforin gene mutations. American journal of hematology. 2007;82(6):427-32. | No direct sequencing |

| | |
|---|---|
| Ulm E, Tolusso L, Marsh R, Dyer L. Disease-causing copy number variants in hemophagocytic lymphohistiocytosis disease genes. Pediatric Blood and Cancer. 2018;65(Supplement 1):S240. | publication type (inc. abstract only) |
| Uribe RMNR, Del Pilar Salazar Rosales H, Vargas AO, Hernandez GL, De Martinez M. Hematopoietic stem cell transplantation for primary immune deficiencies: Experience from a tertiary medical center-Instituto Nacional de Pediatria. Bone Marrow Transplantation. 2019;54:393-4. | publication type (inc. abstract only) |
| Verhelst H, Verloo P, Mondelaers V, Haerynck F, Van Coster R, Bordon V. Neurological presentation in children with familial and acquired hemophagocytic lymphohistiocytosis. European Journal of Paediatric Neurology. 2011;15(SUPPL. 1):S17. | publication type (inc. abstract only) |
| Vinas L, Coniglio ML, Balasco D, Fotzi I, Favre C, Arico M, et al. Monoallelic variants in genes related to familial hemophagocytic lymphohistiocytosis: Report from the Italian registry. Haematologica. 2017;102(Supplement 2):287. | publication type (inc. abstract only) |
| Ware R, Friedman HS, Kinney TR, Kurtzberg J, Chaffee S, Falletta JM. Familial erythrophagocytic lymphohistiocytosis: late relapse despite continuous high-dose VP-16 chemotherapy. Medical and pediatric oncology. 1990;18(1):27-9. | No direct sequencing |
| Welsh F, Hague R. Characteristics of haemophagocytic lymphohistiocytosis at a teritiary paediatric centre. Archives of Disease in Childhood. 2015;100(SUPPL. 3):A192-A3. | publication type (inc. abstract only) |
| Woo S, Champion L, McCellars D. Treatment of familial erythrophagocytic lymphohistiocytosis with vinblastine loaded platelets. Proceedings of the American Association for Cancer Research. 1979;Vol. 20:280. | publication type (inc. abstract only) |
| Xinh PT, Chuong HQ, Diem TPH, Nguyen TM, Van ND, Mai Anh NH, et al. Spectrum mutations of PRF1, UNC13D, STX11, and STXBP2 genes in Vietnamese patients with hemophagocytic lymphohistiocytosis. International journal of laboratory hematology. 2021;43(6):1524-30. | no information about age |
| Xu X, Tang Y, Luo Z, Song H, Xu W. Role of serum cytokines in the diagnosis and treatment of pediatric hemophagocytic lymphohistiocytosis. Pediatric Blood and Cancer. 2022;69(SUPPL 1). | publication type (inc. abstract only) |
| Xu X-J, Wang H-S, Ju X-L, Xiao P-F, Xiao Y, Xue H-M, et al. Clinical presentation and outcome of pediatric patients with hemophagocytic lymphohistiocytosis in China: A retrospective multicenter study. Pediatric blood & cancer. 2017;64(4). | duplicate |

| | |
|---|---|
| Yacobovich J, Dgany O, Krasnov T, Stepansky P, Huri C, Attias D, et al. Molecular analysis of familial hemophagocytic lymphohistiocytosis in Israel. Pediatric Blood and Cancer. 2011;56(4):702. | publication type (inc. abstract only) |
| Yamamoto K, Ishii E, Horiuchi H, Ueda I, Ohga S, Nishi M, et al. Mutations of syntaxin 11 and SNAP23 genes as causes of familial hemophagocytic lymphohistiocytosis were not found in Japanese people. Journal of human genetics. 2005;50(11):600-3. | no information about age |
| Yamamoto K, Ishii E, Sako M, Ohga S, Furuno K, Suzuki N, et al. Identification of novel MUNC13-4 mutations in familial haemophagocytic lymphohistiocytosis and functional analysis of MUNC13-4-deficient cytotoxic T lymphocytes. Journal of medical genetics. 2004;41(10):763-7. | no information about age |
| Yang L, Tang Y, Xiao FX, Xiong J, Shen KF, Liu YN, et al. Hemophagocytic Lymphohistocytosis in the Chinese Han Population May Be Associated with an STXBP2 Gene Polymorphism. PloS one. 2016;11(8):e0159454. | no information about age |
| Zhang J, Sun Y, Shi X, Zhang R, Wang Y, Xiao J, et al. Genotype characteristics and immunological indicator evaluation of 311 hemophagocytic lymphohistiocytosis cases in China. Orphanet journal of rare diseases. 2020;15(1):112. | no information about age |
| Zhang K, Han J, Denton J, Nortman S, Dyer L, Valencia CA, et al. Comprehensive strategy to establish the clinical diagnosis for patients with hemophagocytic lymphohistiocytosis. Pediatric Blood and Cancer. 2017;64(Supplement 2):S43. | publication type (inc. abstract only) |
| Zhang K, Qian Y, Johnson J, Connor J, Meeths M, Bryceson YT, et al. The 253kb inversion and the C.118-308 C>T intronic mutations in UNC13D (MUNC13-4) are found in north american patients with familial hemophagocytic lymphohistiocytosis type 3. Journal of Clinical Immunology. 2012;32(SUPPL. 1):S186. | publication type (inc. abstract only) |
| Zhang K, Wei C, Ma M, Valencia CA, Corrigan D, Nicoletti P, et al. ARTIFICIAL INTELLIGENCE PROVIDES DEEPER GENETIC INSIGHT: PROFILE OF PRIMARY AND SECONDARY HLH USING LARGE SCALE ELECTRONIC HEALTH RECORDS. Pediatric Blood and Cancer. 2023;70(Supplement 1). | publication type (inc. abstract only) |
| Zhang L, Li Z, Wei A, Wang T, Zhang R. The genetic characterization, clinical evaluation of primary hemophagocytic lymphohistiocytosis and the importance of bilirubin abnormality: A single center experience. Pediatric Blood and Cancer. 2020;67(SUPPL 1). | publication type (inc. abstract only) |

| | |
|---|---|
| Zhang L, Ma HH, Wei A, Yang Y, Li Z, Zhang R, et al. Genetic characterization of pediatric primary hemophagocytic lymphohistiocytosis in China: A single center study. Pediatric Blood and Cancer. 2019;66(Supplement 1). | publication type (inc. abstract only) |
| Zhao Y, Li Z, Cheng H, Ding C, Zhang Q, Zhang L, et al. Neurological abnormality could be the first and only symptom of familial hehophagocytic lymphohistiocytosis: Report of two families. Pediatric Blood and Cancer. 2019;66(Supplement 1). | publication type (inc. abstract only) |
| Zhong Y, Villegas M, Yeap F, Ooi P, Ang E, Tan PL. Haemophagocytic lymphohistiocytosis: A decade of experience in a paediatric centre in South-East Asia. Pediatric Blood and Cancer. 2017;64(Supplement 2):S44-S5. | publication type (inc. abstract only) |
| Zieger B, Sandrock K, Nakamura L, Jurk K, Kehrel B, Schulz A, et al. Biochemical and molecular genetic characterization of platelet disorders in patients with immunodeficiency syndromes. European Journal of Pediatrics. 2011;170(2):269. | publication type (inc. abstract only) |
| Zur Stadt U, Beutel K, Weber B, Kabisch H, Schneppenheim R, Janka G. A91V is a polymorphism in the perforin gene not causative of an FHLH phenotype. Blood. 2004;104(6):1909-10. | publication type (inc. abstract only) |
| zur Stadt U, Kabisch H, Janka G, Schneider EM. Rapid LightCycler assay for identification of the perforin codon 374 Trp --> stop mutation in patients and families with hemophagocytic lymphohistiocytosis (HLH). Medical and pediatric oncology. 2003;41(1):26-9. | off topic |
| zur Stadt U, Schmidt S, Kasper B, Beutel K, Diler AS, Henter J-I, et al. Linkage of familial hemophagocytic lymphohistiocytosis (FHL) type-4 to chromosome 6q24 and identification of mutations in syntaxin 11. Human molecular genetics. 2005;14(6):827-34. | publication type (inc. abstract only) |
| Akyol S, Ozcan A, Sekine T, Chiang SC, Yilmaz E, Karakurkcu M, et al. Different clinical presentation of 3 patients with familial hemophagocytic lymphohistiocytosis with two novel mutations. HemaSphere. 2018;2(Supplement 2):901. | subpopulation |
| Benson LA, Li H, Henderson LA, Solomon IH, Soldatos A, Murphy J, et al. Pediatric CNS-isolated hemophagocytic lymphohistiocytosis. Neurology(R) neuroimmunology & neuroinflammation. 2019;6(3):e560. | subpopulation |
| Bergsten E, Horne A, Hed Myrberg I, Arico M, Astigarraga I, Ishii E, et al. Stem cell transplantation for children with hemophagocytic lymphohistiocytosis: results from the HLH-2004 study. Blood advances. 2020;4(15):3754-66. | subpopulation |

| | |
|---|---|
| Blincoe A, Heeg M, Campbell PK, Hines M, Khojah A, Klein-Gitelman M, et al. Neuroinflammatory Disease as an Isolated Manifestation of Hemophagocytic Lymphohistiocytosis. Journal of clinical immunology. 2020;40(6):901-16. | subpopulation |
| Cesaro S, Locatelli F, Lanino E, Porta F, Di Maio L, Messina C, et al. Hematopoietic stem cell transplantation for hemophagocytic lymphohistiocytosis: a retrospective analysis of data from the Italian Association of Pediatric Hematology Oncology (AIEOP). Haematologica. 2008;93(11):1694-701. | subpopulation |
| Cetica V, Santoro A, Gilmour KC, Sieni E, Beutel K, Pende D, et al. STXBP2 mutations in children with familial haemophagocytic lymphohistiocytosis type 5. Journal of medical genetics. 2010;47(9):595-600. | subpopulation |
| Danielian S, Basile N, Rocco C, Prieto E, Rossi J, Barsotti D, et al. Novel syntaxin 11 gene (STX11) mutation in three Argentinean patients with hemophagocytic lymphohistiocytosis. Journal of clinical immunology. 2010;30(2):330-7. | subpopulation |
| Feldmann J, Menasche G, Callebaut I, Minard-Colin V, Bader-Meunier B, Le Clainche L, et al. Severe and progressive encephalitis as a presenting manifestation of a novel missense perforin mutation and impaired cytolytic activity. Blood. 2005;105(7):2658-63. | subpopulation |
| Feng W-X, Yang X-Y, Li J-W, Gong S, Wu Y, Zhang W-H, et al. Neurologic Manifestations as Initial Clinical Presentation of Familial Hemophagocytic Lymphohistiocytosis Type2 Due to PRF1 Mutation in Chinese Pediatric Patients. Frontiers in genetics. 2020;11:126. | subpopulation |
| Gao L, Yang L, Huang L, Xiao Y, Deng J, Zheng M, et al. Clinical and genetic features of Epstein-Barr virus-triggered late-onset primary hemophagocytic lymphohistiocytosis: Ten pedigrees study. Clinical and translational medicine. 2021;11(6):e393. | subpopulation |
| Hadzic N, Molnar E, Height S, Kovacs G, Dhawan A, Andrikovics H, et al. High Prevalence of Hemophagocytic Lymphohistiocytosis in Acute Liver Failure of Infancy. The Journal of pediatrics. 2022;250:67-74.e1. | subpopulation |
| Li H, Benson LA, Henderson LA, Solomon IH, Kennedy AL, Soldatos A, et al. Central nervous system-restricted familial hemophagocytic lymphohistiocytosis responds to hematopoietic cell transplantation. Blood advances. 2019;3(4):503-7. | subpopulation |
| Merli P, Zecca M, Fagioli F, Rovelli A, Lanino E, Bertaina A, et al. Outcomes of children with Hemophagocytic | subpopulation |

| | |
|---|---|
| Lymphohistiocytosis given allogeneic Hematopoietic Stem Cell Transplantation in Italy. Bone Marrow Transplantation. 2017;52(Supplement 1):157-8. | |
| Rudd E, Ericson KG, Zheng C, Uysal Z, Özkan A, Gürgey A, et al. Spectrum and clinical implications of syntaxin 11 gene mutations in familial haemophagocytic lymphohistiocytosis:: association with disease-free remissions and haematopoietic malignancies -: art. no. e14. Journal of Medical Genetics. 2006;43(4):6. | subpopulation |
| Sadeghi P, Esslami GG, Rokni-Zadeh H, Changi-Ashtiani M, Mohsenipour R. Familial Hemophagocytic Lymphohistiocytosis secondary to UNC13D mutation: a report of two cases. BMC pediatrics. 2022;22(1):667. | subpopulation |
| Seo JY, Lee KO, Yoo KH, Sung KW, Koo HH, Kim SH, et al. Prevalence of type 5 familial hemophagocytic lymphohistiocytosis in Korea and novel mutations in STXBP2. Clinical genetics. 2016;89(2):222-7. | subpopulation |
| Seo JY, Song J-S, Lee K-O, Won H-H, Kim J-W, Kim S-H, et al. Founder effects in two predominant intronic mutations of UNC13D, c.118-308C>T and c.754-1G>C underlie the unusual predominance of type 3 familial hemophagocytic lymphohistiocytosis (FHL3) in Korea. Annals of hematology. 2013;92(3):357-64. | subpopulation |
| Swaminathan VV, Uppuluri R, Meena SK, Varla H, Chandar R, Ramakrishnan B, et al. Treosulfan-Based Conditioning in Matched Family, Unrelated and Haploidentical Hematopoietic Stem Cell Transplantation for Genetic Hemophagocytic Lymphohistiocytosis: Experience and Outcomes over 10 Years from India. Indian journal of hematology & blood transfusion : an official journal of Indian Society of Hematology and Blood Transfusion. 2022;38(1):84-91. | subpopulation |
| Tesi B, Chiang SCC, El-Ghoneimy D, Hussein AA, Langenskiold C, Wali R, et al. Spectrum of Atypical Clinical Presentations in Patients with Biallelic PRF1 Missense Mutations. Pediatric blood & cancer. 2015;62(12):2094-100. | subpopulation |
| Vahidi M, Badalzadeh M, Jannesar M, Mazinani M, Fazlollahi MR, Khodayari Namini N, et al. Clinical and Genetic Analysis of Nine Suspected Familial Haemophagocytic Lymphohistiocytosis Patients for MUNC13-4 Deficiency and Introducing Four Novel Mutations in UNC13D. Iranian journal of allergy, asthma, and immunology. 2019;18(5):487-92. | subpopulation |
| Wang Y, Wang Z, Zhang J, Wei Q, Tang R, Qi J, et al. Genetic features of late onset primary hemophagocytic | subpopulation |

| | |
|---|---|
| lymphohistiocytosis in adolescence or adulthood. PloS one. 2014;9(9):e107386. | |
| Zhang K, Chandrakasan S, Chapman H, Valencia CA, Husami A, Kissell D, et al. Synergistic defects of different molecules in the cytotoxic pathway lead to clinical familial hemophagocytic lymphohistiocytosis. Blood. 2014;124(8):1331-4. | subpopulation |

#### MCADD

| Reference | Reason for exclusion (Q1) | Reason for exclusion (Q2) |
|---|---|---|
| Adhikari AN, Gallagher RC, Wang Y, Currier RJ, Amatuni G, Bassaganyas L, et al. The role of exome sequencing in newborn screening for inborn errors of metabolism. Nat Med. 2020;26(9):1392-7. | N/A | >=10% other conditions + not reported separately |
| Alcaide P, Ferrer-Lopez I, Gutierrez L, Leal F, Martin-Hernandez E, Quijada-Fraile P, et al. Lymphocyte Medium-Chain Acyl-CoA Dehydrogenase Activity and Its Potential as a Diagnostic Confirmation Tool in Newborn Screening Cases. Journal of clinical medicine. 2022;11(10). | N/A | No direct sequencing |
| Andresen BS, Bross P, Jensen TG, Knudsen I, Winter V, Kolvraa S, et al. Molecular diagnosis and characterization of medium-chain acyl-CoA dehydrogenase deficiency. Scandinavian journal of clinical and laboratory investigation Supplementum. 1995;220:9-25. | N/A | publication type (inc. abstract only) |
| Andresen BS, Bross P, Jensen TG, Winter V, Knudsen I, Kolvraa S, et al. A rare disease-associated mutation in the medium-chain acyl-CoA dehydrogenase (MCAD) gene changes a conserved arginine, previously shown to be functionally essential in short-chain acyl-CoA dehydrogenase (SCAD). American journal of human genetics. 1993;53(3):730-9. | N/A | publication type (inc. abstract only) |
| Anonymous. Mutations causing medium-chain acyl-CoA dehydrogenase deficiency: a collaborative compilation of the data from 172 patients. Workshop on Molecular Aspects of MCAD Deficiency. Progress in clinical and biological research. 1992;375:499-506. | N/A | no information about age |
| Blois B, Riddell C, Dooley K, Dyack S. Newborns with C8-acylcarnitine level over the 90th centile have an increased frequency of the common MCAD 985A>G mutation. Journal of inherited metabolic disease. 2005;28(4):551-6. | N/A | No direct sequencing |
| Cantu-Reyna C, Zepeda LM, Montemayor R, Benavides S, Gonzalez HJ, Vazquez-Cantu M, et al. Incidence of inborn | N/A | >=10% other conditions + |

| | | |
|---|---|---|
| errors of metabolism by expanded newborn screening in a Mexican hospital. Journal of Inborn Errors of Metabolism and Screening. 2016;4(no pagination). | | not reported separately |
| Curtis D, Blakemore AI, Engel PC, Macgregor D, Besley G, Kolvraa S, et al. Heterogeneity for mutations in medium chain acyl-CoA dehydrogenase deficiency in the UK population. Clinical genetics. 1991;40(4):283-6. | N/A | No direct sequencing |
| de Vries HG, Niezen-Koning K, Kliphuis JW, Smit GP, Scheffer H, ten Kate LP. Prevalence of carriers of the most common medium-chain acyl-CoA dehydrogenase (MCAD) deficiency mutation (G985A) in The Netherlands. Human genetics. 1996;98(1):1-2. | irrelevant outcomes only | No direct sequencing |
| Derks TGJ, Duran M, Waterham HR, Reijngoud D-J, Ten Kate LP, Smit GPA. The difference between observed and expected prevalence of MCAD deficiency in The Netherlands: a genetic epidemiological study. European journal of human genetics : EJHG. 2005;13(8):947-52. | N/A | No direct sequencing |
| Ding JH, Bross P, Yang BZ, Iafolla AK, Millington DS, Roe CR, et al. Genetic heterogeneity in MCAD deficiency: frequency of K329E allele and identification of three additional mutant alleles. Progress in clinical and biological research. 1992;375:479-88. | N/A | no information about age |
| Ding JH, Roe CR, Iafolla AK, Chen YT. Medium-chain acyl-coenzyme A dehydrogenase deficiency and sudden infant death. The New England journal of medicine. 1991;325(1):61-2. | N/A | publication type (inc. abstract only) |
| Dong L, Ji C, Xu J, Cui Y. Screening and follow-up results of neonate medium-chain acyl-CoA dehydrogenase deficiency in Zibo, Shandong province. Zhejiang da xue xue bao Yi xue ban = Journal of Zhejiang University Medical sciences. 2022;51(3):284-9. | N/A | language |
| Dundar M, Tahiri S, Saatci C, Ozkul Y, Caglayan AO. Frequency of the common G985A mutation in the medium-chain acyl-coa dehydrogenase gene in Turkish population. Erciyes Tip Dergisi. 2007;29(4):263-7. | irrelevant outcomes only | No direct sequencing |
| Ensenauer R, Winters JL, Parton PA, Kronn DF, Kim J-W, Matern D, et al. Genotypic differences of MCAD deficiency in the Asian population: novel genotype and clinical symptoms preceding newborn screening notification. Genet Med. 2005;7(5):339-43 | N/A | publication type (inc. abstract only) |
| Ferreira ACS, Orlandi MPA, Oliveira VC, Malta FSV, Caxito FA, Gomes KB, et al. A985G mutation incidence in the medium- | N/A | No direct sequencing |

| | | |
|---|---|---|
| chain acyl-CoA dehydrogenase (MCAD) gene in Brazil. Genetics and molecular research : GMR. 2009;8(2):487-93. | | |
| Ged C, el Sebai H, de Verneuil H, Parrot-Rouleau F. Is genotyping useful for the screening of medium-chain acyl-CoA dehydrogenase deficiency in France? Journal of inherited metabolic disease. 1995;18(2):253-6. | N/A | duplicate |
| Ged C, Elsebai H, Deverneuil H, Parrotrouleau F. IS GENOTYPING USEFUL FOR THE SCREENING OF MEDIUM-CHAIN ACYL-COA DEHYDROGENASE-DEFICIENCY IN FRANCE. Journal of Inherited Metabolic Disease. 1995;18(2):253-6. | N/A | No direct sequencing |
| Giroux S, Dube-Linteau A, Cardinal G, Labelle Y, Laflamme N, Giguere Y, et al. Assessment of the prevalence of the 985A>G MCAD mutation in the French-Canadian population using allele-specific PCR. Clinical genetics. 2007;71(6):569-75. | irrelevant outcomes only | wrong population |
| Goncalves MM, Marcao A, Sousa C, Nogueira C, Ferreira F, Fonseca H, et al. Portuguese Neonatal Screening Programme: A Retrospective Cohort Study of 18 Years of MS/MS. Endocrine, metabolic & immune disorders drug targets. 2023. | N/A | publication type (inc. abstract only) |
| Gregersen N, Andresen BS, Bross P. Prevalent mutations in fatty acid oxidation disorders: diagnostic considerations. European journal of pediatrics. 2000;159 Suppl 3:S213-8. | N/A | publication type (inc. abstract only) |
| Gregersen N, Andresen BS, Bross P, Winter V, Rudiger N, Engst S, et al. Characterization of a disease-causing Lys329 to Glu mutation in 16 patients with medium-chain acyl-CoA dehydrogenase deficiency. Journal of inherited metabolic disease. 1991;14(3):314-6. | N/A | No direct sequencing |
| Gregersen N, Blakemore AIF, Winter V, Andresen B, Kolvraa S, Bolund L, et al. SPECIFIC DIAGNOSIS OF MEDIUM-CHAIN ACYL-COA DEHYDROGENASE (MCAD) DEFICIENCY IN DRIED BLOOD SPOTS BY A POLYMERASE CHAIN-REACTION (PCR) ASSAY DETECTING A POINT-MUTATION (G985) IN THE MCAD GENE. Clinica Chimica Acta. 1991;203(1):23-34. | N/A | duplicate |
| Gregersen N, Blakemore AI, Winter V, Andresen B, Kolvraa S, Bolund L, et al. Specific diagnosis of medium-chain acyl-CoA dehydrogenase (MCAD) deficiency in dried blood spots by a polymerase chain reaction (PCR) assay detecting a point-mutation (G985) in the MCAD gene. Clinica chimica acta; international journal of clinical chemistry. 1991;203(1):23-34. | N/A | No direct sequencing |
| Gregersen N, Winter V, Curtis D, Deufel T, Mack M, Hendrickx J, et al. MEDIUM-CHAIN ACYL-COA DEHYDROGENASE (MCAD) DEFICIENCY - THE PREVALENT MUTATION G985 (K304E) IS SUBJECT TO A STRONG FOUNDER EFFECT FROM | N/A | duplicate |

| | | |
|---|---|---|
| NORTHWESTERN EUROPE. Human Heredity. 1993;43(6):342-50. | | |
| Gregersen N, Winter V, Curtis D, Deufel T, Mack M, Hendrickx J, et al. Medium chain Acyl-CoA dehydrogenase (MCAD) deficiency: The prevalent mutation G985 (K304E) is subject to a strong founder effect from northwestern Europe. Human Heredity. 1993;43(6):342-50. | N/A | duplicate |
| Gregersen N, Winter V, Kolvraa S, Andresen BS, Bross P, Blakemore A, et al. Molecular analysis of medium-chain acyl-CoA dehydrogenase deficiency: a diagnostic approach. Progress in clinical and biological research. 1992;375:441-52. | N/A | No direct sequencing |
| Gregersen N, Winter V, Lyonnet S, Saudubray JM, Wendel U, Jensen TG, et al. MOLECULAR-GENETIC CHARACTERIZATION AND URINARY-EXCRETION PATTERN OF METABOLITES IN 2 FAMILIES WITH MCAD DEFICIENCY DUE TO COMPOUND HETEROZYGOSITY WITH A 13 BASE-PAIR INSERTION IN ONE ALLELE. Journal of Inherited Metabolic Disease. 1994;17(2):169-84. | N/A | duplicate |
| Huidekoper HH, Schneider J, Westphal T, Vaz FM, Duran M, Wijburg FA. Prolonged moderate-intensity exercise without and with L-carnitine supplementation in patients with MCAD deficiency. Journal of inherited metabolic disease. 2006;29(5):631-6. | N/A | off topic |
| Inagaki T, Ohishi N, Bachmann C, Ghisla S, Tsukagoshi N, Udaka S, et al. IMMUNOCHEMICAL AND MOLECULAR ANALYSIS OF MEDIUM-CHAIN ACYL COA DEHYDROGENASE-DEFICIENCY. J Clin Biochem Nutr. 1990;8(1):1-8. | N/A | No direct sequencing |
| Johansson A, Guthenberg C, Ahlman H, Von Dobeln U, Hagenfeldt L. Prevalence of the 985A > G mutation in the medium-chain acyl-CoA dehydrogenase (MCAD) gene in Sweden. Scandinavian Journal of Clinical and Laboratory Investigation. 1999;59(4):289-91. | irrelevant outcomes only | wrong population |
| Joy P, Black C, Rocca A, Haas M, Wilcken B. Neuropsychological functioning in children with medium chain acyl coenzyme a dehydrogenase deficiency (MCADD): the impact of early diagnosis and screening on outcome. Child neuropsychology : a journal on normal and abnormal development in childhood and adolescence. 2009;15(1):8-20. | N/A | No direct sequencing |
| Khalid JM, Oerton J, Cortina-Borja M, Andresen BS, Besley G, Dalton RN, et al. Ethnicity of children with homozygous c.985A>G medium-chain acyl-CoA dehydrogenase deficiency: | N/A | irrelevant outcomes only |

| | | |
|---|---|---|
| findings from screening approximately 1.1 million newborn infants. Journal of medical screening. 2008;15(3):112-7. | | |
| Khalid JM, Oerton J, Cortina-Borja M, Andresen BS, Besley G, Dalton RN, et al. Ethnicity of children with homozygous c.985A>G medium-chain acyl-CoA dehydrogenase deficiency: findings from screening approximately 1.1 million newborn infants. Journal of medical screening. 2008;15(3):112-7. | N/A | publication type (inc. abstract only) |
| Kozak L, Hrabincova E, Rudolfoa J, Vrabelova S, Freiberger T. Screening of the most common medium-chain acyl CoA dehydrogenase (MCAD) deficiency mutation (K329E) in the Czech newborn population. The Southeast Asian journal of tropical medicine and public health. 1999;30 Suppl 2:49-50. | irrelevant outcomes only | wrong population |
| Kozak L, Hrabincova E, Rudolfoa J, Vrabelova S, Freiberger T. Screening of the most common medium-chain acyl CoA dehydrogenase (MCAD) deficiency mutation (K329E) in the Czech newborn population. The Southeast Asian journal of tropical medicine and public health. 1999;30 Suppl 2:49-50. | N/A | No direct sequencing |
| Lam C, Mardach RR, Broome DL, Cederbaum S, Qu Y. Moderate increases of medium chain acylcarnitines: A real MCADD patient or a medication origin? Molecular Genetics and Metabolism. 2011;102(3):310. | N/A | publication type (inc. abstract only) |
| Leal J, Ades AE, Wordsworth S, Dezateux C. Regional differences in the frequency of the c.985A>G ACADM mutation: findings from a meta-regression of genotyping and screening studies. Clinical genetics. 2014;85(3):253-9. | N/A | study design |
| Lecoq I, Mallet E, Bonte JB, Travert G. The A985 to G mutation of the medium-chain acyl-CoA dehydrogenase gene and sudden infant death syndrome in Normandy. Acta paediatrica (Oslo, Norway : 1992). 1996;85(2):145-7. | irrelevant outcomes only | No direct sequencing |
| Lehotay DC, LePage J, Thompson JR, Rockman-Greenberg C. Blood acylcarnitine levels in normal newborns and heterozygotes for medium-chain acyl-CoA dehydrogenase deficiency: a relationship between genotype and biochemical phenotype? Journal of inherited metabolic disease. 2004;27(1):81-8. | N/A | wrong population |
| Liebl B, Nennstiel-Ratzel U, Roscher A, von Kries R. Data required for the evaluation of newborn screening programmes. European journal of pediatrics. 2003;162 Suppl 1:S57-61. | N/A | publication type (inc. abstract only) |
| Lillevali H, Margus K, Ounap K, Metspalu A. Mutation 985A>G in the MCAD gene shows low incidence in Estonian population. Human mutation. 2000;15(3):293-4. | N/A | No direct sequencing |

| | | |
|---|---|---|
| Longo N, Anderson DR, Viau K, Pasquali M. Clinical and biochemical outcome of patients with medium-chain acyl-CoA dehydrogenase deficiency. Journal of Inherited Metabolic Disease. 2016;39(Supplement 1):S152-S3. | N/A | publication type (inc. abstract only) |
| Maier EM, Liebl B, Roschinger W, Nennstiel-Ratzel U, Fingerhut R, Olgemoller B, et al. Population spectrum of ACADM genotypes correlated to biochemical phenotypes in newborn screening for medium-chain aryl-CoA dehydrogenase deficiency. Human Mutation. 2005;25(5):443-52. | N/A | duplicate |
| Maier EM, Pongratz J, Muntau AC, Liebl B, Nennstiel-Ratzel U, Busch U, et al. Dissection of biochemical borderline phenotypes in carriers and genetic variants of medium-chain acyl-CoA dehyrogenase deficiency: implications for newborn screening [corrected]. Clinical genetics. 2009;76(2):179-87. | N/A | wrong population |
| Matsubara Y, Narisawa K, Tada K, Ikeda H, Yao YQ, Danks DM, et al. Prevalence of K329E mutation in medium-chain acyl-CoA dehydrogenase gene determined from Guthrie cards. Lancet (London, England). 1991;338(8766):552-3. | irrelevant outcomes only | No direct sequencing |
| McKinney JT, Longo N, Hahn SH, Matern D, Rinaldo P, Strauss AW, et al. Rapid, comprehensive screening of the human medium chain acyl-CoA dehydrogenase gene. Molecular genetics and metabolism. 2004;82(2):112-20. | N/A | FT not retrievable |
| Narravula A, Garber KB, Askree SH, Hegde M, Hall PL. Variants of uncertain significance in newborn screening disorders: implications for large-scale genomic sequencing. Genet Med. 2017;19(1):77-82. | N/A | no information about age |
| Opdal SH, Vege A, Saugstad OD, Rognum TO. Is the medium-chain acyl-CoA dehydrogenase G985 mutation involved in sudden infant death in Norway? [6]. European Journal of Pediatrics. 1995;154(2):166-7. | N/A | publication type (inc. abstract only) |
| Romppanen EL, Mononen T, Mononen I. Molecular diagnosis of medium-chain acyl-CoA dehydrogenase deficiency by oligonucleotide ligation assay. Clinical chemistry. 1998;44(1):68-71. | N/A | FT not retrievable |
| Sander S, Janzen N, Janetzky B, Scholl S, Steuerwald U, Schafer J, et al. Neonatal screening for medium chain acyl-CoA deficiency: high incidence in Lower Saxony (northern Germany). European journal of pediatrics. 2001;160(5):318-9. | N/A | No direct sequencing |
| Seddon HR, Gray G, Pollitt RJ, Iitia A, Green A. Population screening for the common G985 mutation causing medium-chain acyl-CoA dehydrogenase deficiency with Eu-labeled | irrelevant outcomes only | No direct sequencing |

| | | |
|---|---|---|
| oligonucleotides and the DELFIA system. Clinical chemistry. 1997;43(3):436-42. | | |
| Seddon HR, Green A, Gray RG, Leonard JV, Pollitt RJ. Regional variations in medium-chain acyl-CoA dehydrogenase deficiency. Lancet (London, England). 1995;345(8942):135-6. | N/A | No direct sequencing |
| Smith EH, Thomas C, McHugh D, Gavrilov D, Raymond K, Rinaldo P, et al. Allelic diversity in MCAD deficiency: the biochemical classification of 54 variants identified during 5 years of ACADM sequencing. Molecular genetics and metabolism. 2010;100(3):241-50. | N/A | no information about age |
| Strauss AW, Duran M, Zhang ZF, Alpers R, Kelly DP. Molecular analysis of medium chain acyl-CoA dehydrogenase deficiency. Progress in clinical and biological research. 1990;321:609-23. | N/A | publication type (inc. abstract only) |
| Szalai C, Czinner A, Revai K. The frequency of medium-chain acyl-CoA dehydrogenase G985 mutation in the Hungarian population [4]. European Journal of Pediatrics. 1996;155(3):256. | N/A | publication type (inc. abstract only) |
| Tan J, Wagner M, Stenton SL, Strom TM, Wortmann SB, Prokisch H, et al. Lifetime risk of autosomal recessive mitochondrial disorders calculated from genetic databases. EBioMedicine. 2020;54:102730. | N/A | off topic |
| Tanaka K, Gregersen N, Ribes A, Kim J, Kolvraa S, Winter V, et al. A survey of the newborn populations in Belgium, Germany, Poland, Czech Republic, Hungary, Bulgaria, Spain, Turkey, and Japan for the G985 variant allele with haplotype analysis at the medium chain Acyl-CoA dehydrogenase gene locus: clinical and evolutionary consideration. Pediatric research. 1997;41(2):201-9. | N/A | FT not retrievable |
| Tanaka K, Yokota I, Coates PM, Strauss AW, Kelly DP, Zhang Z, et al. Mutations in the medium chain acyl-CoA dehydrogenase (MCAD) gene. Human mutation. 1992;1(4):271-9. | N/A | publication type (inc. abstract only) |
| Thompson JR, Manchur D, Gregory C, Dilling L, Lacson A, Seargeant L, et al. DNA analysis for detection of medium-chain acyl-CoA dehydrogenase deficiency in a Manitoba newborn population. Screening. 1995;4(1):9-15. | N/A | FT not retrievable |
| Udvari S, Bross P, Andresen BS, Gregersen N, Engel PC. Biochemical characterisation of mutations of human medium-chain acyl-CoA dehydrogenase. Advances in Experimental Medicine and Biology. 2000;466:387-93. | N/A | publication type (inc. abstract only) |

| | | |
|---|---|---|
| Wilcken B, Carpenter KH, Hammond J. Neonatal symptoms in medium chain acyl coenzyme A dehydrogenase deficiency. Archives of disease in childhood. 1993;69(3 Spec No):292-4. | N/A | No direct sequencing |
| Yang C, Shi C, Zhou C, Wan Q, Zhou Y, Chen X, et al. Screening and follow-up results of fatty acid oxidative metabolism disorders in 608 818 newborns in Jining, Shandong province. Zhejiang da xue xue bao Yi xue ban = Journal of Zhejiang University Medical sciences. 2021;50(4):472-80. | N/A | language |
| Yang N, Gong L-F, Zhao J-Q, Yang H-H, Ma Z-J, Liu W, et al. Inborn errors of metabolism detectable by tandem mass spectrometry in Beijing. Journal of pediatric endocrinology & metabolism : JPEM. 2020;33(5):639-45. | N/A | >=10% other conditions + not reported separately |
| Yokota I, Coates PM, Hale DE, Rinaldo P, Tanaka K. The molecular basis of medium chain acyl-CoA dehydrogenase deficiency: survey and evolution of 985A----G transition, and identification of five rare types of mutation within the medium chain acyl-CoA dehydrogenase gene. Progress in clinical and biological research. 1992;375:425-40. | N/A | No direct sequencing |
| Yokota I, Indo Y, Coates PM, Tanaka K. Molecular basis of medium chain acyl-coenzyme A dehydrogenase deficiency. An A to G transition at position 985 that causes a lysine-304 to glutamate substitution in the mature protein is the single prevalent mutation. The Journal of clinical investigation. 1990;86(3):1000-3. | N/A | off topic |
| Ziadeh R, Hoffman EP, Finegold DN, Hoop RC, Brackett JC, Strauss AW, et al. Medium chain acyl-CoA dehydrogenase deficiency in Pennsylvania: neonatal screening shows high incidence and unexpected mutation frequencies. Pediatric research. 1995;37(5):675-8. | N/A | No direct sequencing |
| Andresen BS, Bross P, Knudsen I, Winter V, Kolvraa S, Bolund L, et al. Medium-chain acyl-CoA dehydrogenase (MCAD) deficiency due to heterozygosity for the common mutation and an allele resulting in low levels of MCAD mRNA. Journal of inherited metabolic disease. 1994;17(3):275-8. | N/A | subpopulation |
| Andresen BS, Bross P, Udvari S, Kirk J, Gray G, Kmoch S, et al. The molecular basis of medium-chain acyl-CoA dehydrogenase (MCAD) deficiency in compound heterozygous patients: is there correlation between genotype and phenotype? Human molecular genetics. 1997;6(5):695-707. | N/A | subpopulation |
| Brackett JC, Sims HF, Steiner RD, Nunge M, Zimmerman EM, deMartinville B, et al. A novel mutation in medium chain acyl- | N/A | subpopulation |

| | | |
|---|---|---|
| CoA dehydrogenase causes sudden neonatal death. The Journal of clinical investigation. 1994;94(4):1477-83. | | |
| McCandless SE, Chandrasekar R, Linard S, Kikano S, Rice L. Sequencing from dried blood spots in infants with "false positive" newborn screen for MCAD deficiency. Molecular genetics and metabolism. 2013;108(1):51-5. | N/A | subpopulation |
| Conne B, Zufferey R, Belin D. The A985G mutation in the medium-chain acyl-CoA dehydrogenase gene: high prevalence in the Swiss population resident in Geneva. Journal of inherited metabolic disease. 1995;18(5):577-83. | subpopulation | subpopulation |
| Yusupov R, Finegold DN, Naylor EW, Sahai I, Waisbren S, Levy HL. Sudden death in medium chain acyl-coenzyme a dehydrogenase deficiency (MCADD) despite newborn screening. Molecular genetics and metabolism. 2010;101(1):33-9. | N/A | subpopulation |
| Zschocke J, Schulze A, Lindner M, Fiesel S, Olgemoller K, Hoffmann GF, et al. Molecular and functional characterisation of mild MCAD deficiency. Human genetics. 2001;108(5):404-8. | N/A | subpopulation |

### Studies eligible for Q4 (What is the effectiveness of earlier vs later intervention for each condition?)

#### PDE

| Reference | Reason for exclusion (Q4) |
|---|---|
| Akiyama T, Akiyama M, Hayashi Y, Shibata T, Hanaoka Y, Toda S, Imai K, Hamano SI, Okanishi T, Yoshinaga H, Kobayashi K. Measurement of pyridoxal 5'-phosphate, pyridoxal, and 4-pyridoxic acid in the cerebrospinal fluid of children. Clin Chim Acta. 2017;466:1-5 | Own cutoff needed |
| Jamali A, Kristensen E, Tangeraas T, Arntsen V, Sikiric A, Kupliauskiene G, Myren-Svelstad S, Berland S, Sejersted Y, Gerstner T, Hassel B, Bindoff LA, Brodtkorb E. The spectrum of pyridoxine dependent epilepsy across the age span: A nationwide retrospective observational study. Epilepsy Res. 2023;190:107099 | Own cutoff needed |
| Coughlin CR, 2nd, Tseng LA, Bok LA, Hartmann H, Footitt E, Striano P, et al. Association Between Lysine Reduction Therapies and Cognitive Outcomes in Patients With Pyridoxine-Dependent Epilepsy. Neurology. 2022;99(23):e2627-e36 | Own cutoff needed |
| Jiao X, Gong P, Niu Y, Zhang Y, Yang Z. A Rare Presentation Characterized by Epileptic Spasms in ALDH7A1, Pyridox(am)ine- | Own cutoff needed |

| | |
|---|---|
| 5'-Phosphate Oxidase, and PLPBP Deficiency. Frontiers in genetics. 2022;13:804461. | |
| Kanno J, Kure S, Narisawa A, Kamada F, Takayanagi M, Yamamoto K, et al. Allelic and non-allelic heterogeneities in pyridoxine dependent seizures revealed by ALDH7A1 mutational analysis. Molecular genetics and metabolism. 2007;91(4):384-9. | Own cutoff needed |
| Plecko B, Hikel C, Korenke GC, Schmitt B, Baumgartner M, Baumeister F, et al. Pipecolic acid as a diagnostic marker of pyridoxine-dependent epilepsy.Neuropediatrics 2005, 36;3; 200-5 | Own cutoff needed |
| Falsaperla, R., Vari, M.S., Toldo, I. et al. Pyridoxine-dependent epilepsies: an observational study on clinical, diagnostic, therapeutic and prognostic features in a pediatric cohort. Metab Brain Dis 2018, 33, 261–269 | Own cutoff needed |
| Schmitt B, Baumgartner M, Mills PB, Clayton PT, Jakobs C, Keller E, et al. Seizures and paroxysmal events: symptoms pointing to the diagnosis of pyridoxine-dependent epilepsy and pyridoxine phosphate oxidase deficiency. Developmental medicine and child neurology. 2010;52(7):e133-42. | Own cutoff needed |
| Jiao X, Xue J, Gong P, et al. Clinical and genetic features in pyridoxine-dependent epilepsy: a Chinese cohort study. Developmental Medicine and Child Neurology. 2020 Mar;62(3):315-321. DOI: 10.1111/dmcn.14385. PMID: 31737911. | Own cutoff needed |

#### hRB

| **Reference** | **Reason for exclusion (Q4)** |
|---|---|
| de Jong MC, Kors WA, de Graaf P, Castelijns JA, Kivela T, Moll AC. Trilateral retinoblastoma: a systematic review and meta-analysis. The Lancet Oncology. 2014;15(10):1157-67. | FT not retrievable |
| Jain IS, Mohan K, Jain S. Bilateral retinoblastoma in northern India--modes of presentation and heredity. Indian journal of ophthalmology. 1986;34:311-4. | FT not retrievable |
| Barbosa RH, Aguiar FCC, Silva MFL, Costa RA, Vargas FR, Lucena E, et al. Screening of RB1 alterations in Brazilian patients with retinoblastoma and relatives with retinoma: phenotypic and genotypic associations. Investigative ophthalmology & visual science. 2013;54(5):3184-94. | genetics only |

| | |
|---|---|
| Cohen JG, Dryja TP, Davis KB, Diller LR, Li FP. RB1 genetic testing as a clinical service: a follow-up study. Medical and pediatric oncology. 2001;37(4):372-8. | genetics only |
| Dhar SU, Chintagumpala M, Noll C, Chevez-Barrios P, Paysse EA, Plon SE. Outcomes of integrating genetics in management of patients with retinoblastoma. Archives of ophthalmology (Chicago, Ill : 1960). 2011;129(11):1428-34. | genetics only |
| Frenkel S, Pe'er J. The clinical presentation of retinoblastoma patients with mosaics. Investigative Ophthalmology and Visual Science. 2016;57(12):3672. | genetics only |
| Gargallo P, Oltra JS, Yanez Y, Segura V, Balaguer J, Canete A. Retinoblastoma: towards an earlier diagnosis. Retinoblastoma: hacia un diagnostico mas precoz. 2018;93(9):439-43. | genetics only |
| Gregersen PA, Funding M, Alsner J, Olsen MH, Overgaard J, Staffieri SE, et al. Genetic testing in adult survivors of retinoblastoma in Denmark: A study of the experience and impact of genetic testing many years after initial diagnosis. European journal of medical genetics. 2022;65(9):104569 | genetics only |
| Harbour JW. Overview of RB gene mutations in patients with retinoblastoma. Implications for clinical genetic screening. Ophthalmology. 1998;105(8):1442-7 | genetics only |
| Onadim Z, Hogg A, Baird PN, Cowell JK. Oncogenic point mutations in exon 20 of the RB1 gene in families showing incomplete penetrance and mild expression of the retinoblastoma phenotype. Proceedings of the National Academy of Sciences of the United States of America. 1992;89(13):6177-81 | genetics only |
| Parrilla-Vallejo M, Perea-Perez R, Relimpio-Lopez I, Montero-de-Espinosa I, Rodriguez-de-la-Rua E, Terron-Leon JA, et al. Retinoblastoma: The importance of early diagnosis. Retinoblastoma: la importancia de su diagnostico precoz. 2018;93(9):423-30. | genetics only |
| Qureshi S, Francis JH, Haque SS, Dunkel IJ, Souweidane MM, Friedman DN, et al. Magnetic Resonance Imaging Screening for Trilateral Retinoblastoma: The Memorial Sloan Kettering Cancer Center Experience 2006-2016. Ophthalmology Retina. 2020;4(3):327-35. | genetics only |
| Rushlow DE, Mol BM, Kennett JY, Yee S, Pajovic S, Theriault BL, et al. Characterisation of retinoblastomas without RB1 mutations: genomic, gene expression, and clinical studies. The Lancet Oncology. 2013;14(4):327-34 | genetics only |

| | |
|---|---|
| Torbidoni AV, Sampor C, Laurent VE, Aschero R, Iyer S, Rossi J, et al. Minimal disseminated disease evaluation and outcome in trilateral retinoblastoma. The British journal of ophthalmology. 2018;102(11):1597-601 | genetics only |
| Valverde JR, Alonso J, Palacios I, Pestana A. RB1 gene mutation up-date, a meta-analysis based on 932 reported mutations available in a searchable database. BMC genetics. 2005;6:53 | genetics only |
| Zhang R, Song Y-N, Duo X, Guo Z, Sun Y, Zhang Z, et al. Retinoblastoma cell-derived Twist protein promotes regulatory T cell development. Cancer immunology, immunotherapy : CII. 2021;70(4):1037-48 | genetics only |
| Fabian ID, Abdallah E, Abdullahi SU, Abdulqader RA, Abdulrahaman AA, Abouelnaga S, et al. The Global Retinoblastoma Outcome Study: a prospective, cluster-based analysis of 4064 patients from 149 countries. Lancet Glob Health. 2022;10(8):e1128-e40. | irrelevant outcomes only |
| Holladay DA, Holladay A, Montebello JF, Redmond KP. Clinical presentation, treatment, and outcome of trilateral retinoblastoma. Cancer. 1991;67(3):710-5 | irrelevant outcomes only |
| Kao L-Y, Su W-W, Lin Y-W. Retinoblastoma in Taiwan: survival and clinical characteristics 1978-2000. Japanese journal of ophthalmology. 2002;46(5):577-80 | irrelevant outcomes only |
| Wallach M, Balmer A, Munier F, Houghton S, Pampallona S, von der Weid N, et al. Shorter time to diagnosis and improved stage at presentation in Swiss patients with retinoblastoma treated from 1963 to 2004. Pediatrics. 2006;118(5):e1493-8. | irrelevant outcomes only |
| Wilson MW, Haik BG, Billups CA, Rodriguez-Galindo C. Incidence of new tumor formation in patients with hereditary retinoblastoma treated with primary systemic chemotherapy: is there a preventive effect? Ophthalmology. 2007;114(11):2077-82 | irrelevant outcomes only |
| Fabian ID, Abdallah E, Abdullahi SU, Abdulqader RA, Abdulrahaman AA, Abouelnaga S, et al. The Global Retinoblastoma Outcome Study: a prospective, cluster-based analysis of 4064 patients from 149 countries. Lancet Glob Health. 2022;10(8):e1128-e40. | irrelevant outcomes only |
| DerKinderen DJ, Koten JW, Van Romunde LK, Nagelkerke NJ, Tan KE, Beemer FA, et al. Early diagnosis of bilateral retinoblastoma reduces death and blindness. International journal of cancer. 1989;44(1):35-9 | irrelevant treatment (not early vs late, not early ToI) |

| | |
|---|---|
| Moll AC, Imhof SM, Bouter LM, Tan KE. Second primary tumors in patients with retinoblastoma. A review of the literature. Ophthalmic genetics. 1997;18(1):27-34 | irrelevant treatment (not early vs late, not early ToI) |
| Temming P, Viehmann A, Arendt M, Eisele L, Spix C, Bornfeld N, et al. Pediatric second primary malignancies after retinoblastoma treatment. Pediatric blood & cancer. 2015;62(10):1799-804 | irrelevant treatment (not early vs late, not early ToI) |
| Chang C-Y, Chiou T-J, Hwang B, Bai L-Y, Hsu W-M, Hsieh Y-L. Retinoblastoma in Taiwan: survival rate and prognostic factors. Japanese journal of ophthalmology. 2006;50(3):242-9 | irrelevant treatment (not early vs late, not early ToI) |
| Bouchoucha Y, Matet A, Berger A, Carcaboso AM, Gerrish A, Moll A, et al. Retinoblastoma: From genes to patient care. European journal of medical genetics. 2023;66(1):104674 | outcomes of treatment not reported comparatively |
| Brennan RC, Qaddoumi I, Billups CA, Kaluzny T, Furman WL, Wilson MW. Patients with retinoblastoma and chromosome 13q deletions have increased chemotherapy-related toxicities. Pediatric blood & cancer. 2016;63(11):1954-8. | outcomes of treatment not reported comparatively |
| Joseph B, Mamatha G, Raman G, Shanmugam MP, Kumaramanickavel G. Methylation status of RB1 promoter in Indian retinoblastoma patients. Cancer Biol Ther. 2004;3(2):184-7 | outcomes of treatment not reported comparatively |
| Kim JK, Kan G, Mao Y, Wu Z, Tan X, He H, et al. UHRF1 downmodulation enhances antitumor effects of histone deacetylase inhibitors in retinoblastoma by augmenting oxidative stress-mediated apoptosis. Molecular oncology. 2020;14(2):329-46 | outcomes of treatment not reported comparatively |
| Kivela T. Trilateral retinoblastoma: a meta-analysis of hereditary retinoblastoma associated with primary ectopic intracranial retinoblastoma. Journal of clinical oncology : official journal of the American Society of Clinical Oncology. 1999;17(6):1829-37 | outcomes of treatment not reported comparatively |
| Lim FPM, Soh SY, Iyer JV, Tan AM, Swati H, Quah BL. Clinical profile, management, and outcome of retinoblastoma in singapore. Journal of pediatric ophthalmology and strabismus. 2013;50(2):106-12 | outcomes of treatment not reported comparatively |
| Meel R, Kashyap S, Bakhshi S, Singh Bajaj M, Wadhwani M. Retinoblastoma in Children Older than 6 Years of Age. Ocular Oncology and Pathology. 2020;6(6):395-404 | outcomes of treatment not reported comparatively |

| | |
|---|---|
| Naseripour M, Nazari H, Bakhtiari P, Modarres-zadeh M, Vosough P, Ausari M. Retinoblastoma in Iran: outcomes in terms of patients' survival and globe survival. The British journal of ophthalmology. 2009;93(1):28-32 | outcomes of treatment not reported comparatively |
| Hall LS, Ceisler E, Abramson DH. Visual outcomes in children with bilateral retinoblastoma. Journal of AAPOS : the official publication of the American Association for Pediatric Ophthalmology and Strabismus. 1999;3(3):138-42 | outcomes of treatment not reported comparatively |
| Chan HSL, Heon E, Schechter T, Dimaras H, Gallie BL, Doyle JJ. Improving survival of extraocular, metastatic and trilateral retinoblastoma patients with combined intensive therapy. Cancer Research. 2013;73(8 SUPPL. 1) | publication type (inc. abstract only) |
| Frenkel S, Pe'er J. The clinical presentation of retinoblastoma patients with mosaics. Investigative Ophthalmology and Visual Science. 2016;57(12):3672 | publication type (inc. abstract only) |
| Lim FPM, Swati H, Chan MY, Tan AM, Quah BL, Soh SY. Clinical profile of retinoblastoma in a children's cancer centre in singapore. Pediatric Blood and Cancer. 2011;57(5):786. | publication type (inc. abstract only) |
| Mastrangelo D, Di Leonardo A, Lentini L, De Francesco S, Hadjistilianou T. Missing evidences in cancer genetics: the retinoblastoma paradigm. Cellular oncology : the official journal of the International Society for Cellular Oncology. 2008;30(6):509-10 | publication type (inc. abstract only) |
| Providencia J, Castela G, Silva S, Machado E, Murta JN. First 22-months results of retinoblastoma management in a national reference center. European Journal of Ophthalmology. 2017;27(5):e161 | publication type (inc. abstract only) |
| Rodriguez A, Kelley C, Patel A, Ramasubramanian A. Prenatal Diagnosis of Retinoblastomas: A Scoping Review. International Journal of General Medicine. 2023;16:1101-10 | publication type (inc. abstract only) |
| Shields CL, Shields JA. Diagnosis and management of retinoblastoma. Cancer control : journal of the Moffitt Cancer Center. 2004;11(5):317-27 | publication type (inc. abstract only) |
| Staffieri S, Kearns L, Elder J, McKenzie J, Curnow L, Amor D, et al. Familial retinoblastoma and genetic testing: A paradigm shift in clinical care. Twin Research and Human Genetics. 2016;19(5):563-4 | publication type (inc. abstract only) |
| Temming P, Viehmann A, Arendt M, Eisele L, Spix C, Bornfeld N, et al. Pediatric second primary malignancies after retinoblastoma treatment. Pediatric blood & cancer. 2015;62(10):1799-804 | publication type (inc. abstract only) |

| | |
|---|---|
| Waddell K, Kagame K, Picton S, Simmons I, Newton R. Survival from retinoblastoma in rural uganda. Pediatric Blood and Cancer. 2012;59(6):1064-5 | publication type (inc. abstract only) |
| Wijsard MH, Serne SH, Otten RH, Bosscha MI, Dommering CJ, Fabius AW, et al. At what age could screening for familial retinoblastoma be discontinued? A systematic review. Cancers. 2021;13(8):1942 | publication type (inc. abstract only) |
| de Jong MC, Kors WA, de Graaf P, Castelijns JA, Moll AC, Kivela T. The Incidence of Trilateral Retinoblastoma: A Systematic Review and Meta-Analysis. American journal of ophthalmology. 2015;160(6):1116-26.e5 | publication type (inc. abstract only) |
| Kivela T, Tuppurainen K, Riikonen P, Vapalahti M. Retinoblastoma associated with chromosomal 13q14 deletion mosaicism. Ophthalmology. 2003;110(10):1983-8 | study design |
| Pesin SR, Shields JA. Seven cases of trilateral retinoblastoma. American journal of ophthalmology. 1989;107(2):121-6 | study design |
| Raval V, DeBenedictis M, Bowen R, Soto H, Davanzo J, Singh A. Retinoblastoma in twins: Risk assessment of genotypic variants. Indian J Ophthalmol. 2021;69(5):1230-1233 | study design |
| Richter S, Vandezande K, Chen N, Zhang K, Sutherland J, Anderson J, et al. Sensitive and efficient detection of RB1 gene mutations enhances care for families with retinoblastoma. American journal of human genetics. 2003;72(2):253-69 | outcomes of treatment not reported comparatively |
| Taktikos A. Investigation of retinoblastoma with special reference to histology and prognosis. British journal of ophthalmology. 1996(5):225-34 | study design |

#### XLHR

| **Reference** | **Reason for exclusion (Q4)** |
|---|---|
| Alon U, Chan CM. Effects of 1,25-(OH)2-D3 on tubular phosphate (P) handling in X-linked dominant renal hypophosphatemic rickets (RHR). Federation Proceedings. 1982;41(3):No.-59. | publication type (inc. abstract only) |
| Anonymous. X-linked hypophosphatemia. FASEB Journal. 1994;8(11):801. | publication type (inc. abstract only) |
| Anonymous. Burosumab for X-linked hypophosphataemia. Australian prescriber. 2022;45(1):25-6. | publication type (inc. abstract only) |
| Brener A, Lebenthal Y, Cleper R, Kapusta L, Zeitlin L. Body composition and cardiometabolic health of pediatric patients with X-linked hypophosphatemia (XLH) under burosumab | irrelevant treatment (not early vs late, not early ToI) |

| | |
|---|---|
| therapy. Therapeutic advances in endocrinology and metabolism. 2021;12:20420188211001150. | |
| Cagnoli M, Richter R, Bohm P, Knye K, Empting S, Mohnike K. Spontaneous Growth and Effect of Early Therapy with Calcitriol and Phosphate in X-linked Hypophosphatemic Rickets. Pediatric endocrinology reviews : PER. 2017;15(Suppl 1):119-22. | outcomes of treatment not reported comparatively |
| Carpenter T. The authors are full-time employees of Amgen (AG, RBW) and UCB (CG) and own stock and stock options. JBMR Plus. 2019;3(Supplement 2):S9. | publication type (inc. abstract only) |
| Carpenter T, Imel E, G SG, Skrinar A, Mao M, Martin JS, et al. KRN23 effects on phosphate and vitamin d metabolism in children <5 years old with x-linked hypophosphatemia (XLH). Hormone Research in Paediatrics. 2017;88(Supplement 1):10-1. | publication type (inc. abstract only) |
| Cau ACA, Araya N, Carles M, Perez G, Barra P. Burosumab therapy: Case report of a 5-year-old girl with x-linked hypophosphatemic rickets. Hormone Research in Paediatrics. 2020;93(SUPPL 2):43. | publication type (inc. abstract only) |
| Chaussain-Miller C, Sinding C, Wolikow M, Lasfargues J-J, Godeau G, Garabedian M. Dental abnormalities in patients with familial hypophosphatemic vitamin D-resistant rickets: prevention by early treatment with 1-hydroxyvitamin D. The Journal of pediatrics. 2003;142(3):324-31. | >=10% other conditions + not reported separately |
| Connor P, Vondeling G, Marshall JD, Lloyd A. PRO26 THE NICE APPRAISAL OF BUROSUMAB FOR THE TREATMENT OF XLH: EVALUATING THE LIFELONG OUTCOMES AND COST-EFFECTIVENESS OF CORRECTING DEFORMITY WHEN TREATING CHILDREN AND ADOLESCENTS WITH A GROWING SKELETON. Value in Health. 2019;22(Supplement 3):S845. | publication type (inc. abstract only) |
| Costa T, Reade TM, Cole DEC. Renal handling of phosphate (Pi) and bone mineralization in X-linked hypophosphatemia (XLH) during treatment with Pi and 1,25-(OH)2D3. Pediatric Research. 1980;14(4 II):No.-571. | publication type (inc. abstract only) |
| Drezner MK, Harrelson JM. 1,25-Dihydroxyvitamin D3 and phosphate (Pi) therapy can completely heal the bone disease in X-linked hypophosphatemic rickets/osteomalacia (XLH). Clinical Research. 1982;30(2):523A. | publication type (inc. abstract only) |
| Fujimoto M, Senoo S, Kurosawa K, Yamaguchi Y, Hanaki K, Namba N. Effects of burosumab on two siblings with mild X-linked hypophosphatemia. Hormone Research in Paediatrics. 2023;96(Supplement 2):71. | publication type (inc. abstract only) |

| | |
|---|---|
| Glorieux F, Scriver C, Holick M, DeLuca H. The response to 1,25 dihydroxycholecalciferol (1,25 DHCC) in X linked hypophosphatemia. Pediatric Research. 1973;7(4):387. | publication type (inc. abstract only) |
| Gordon RJ, Levine MA. Burosumab treatment of children with X-linked hypophosphataemic rickets. Lancet (London, England). 2019;393(10189):2364-6. | publication type (inc. abstract only) |
| Gottesman G, Wollberg V, Mumm S, Whyte M. Mild X-linked hypophosphatemia (PHEX c. 231A>G): Normalization of Serum Phosphorus Levels with Low-Dose Burosumab Therapy. Journal of Bone and Mineral Research. 2020;35(SUPPL 1):286. | publication type (inc. abstract only) |
| Hogler W, Imel EA, Whyte MP, Munns C, Portale AA, Ward L, et al. Burosumab resulted in better clinical outcomes than continuation with conventional therapy in both younger (1-4 years-old) and older (5-12 years-old) children with X-linked hypophosphatemia. Hormone Research in Paediatrics. 2019;91(Supplement 1):71-2. | publication type (inc. abstract only) |
| Howard CP, Huse DM, Hayles AB, Stickler GB. Growth in patients with familial hypophosphatemic rickets treated with large doses of phosphate. Pediatric Research. 1979;13(4 II):No.-326. | FT not retrievable |
| Hsia DYY, Kraus M, Samuels J. Genetic studies on vitamin D resistant rickets (familial hypophosphatemia). American journal of human genetics. 1959;11(2 Part 1):156-65. | irrelevant outcomes only |
| Imel E, Whyte MP, Munns C, Portale AA, Ward L, Nilsson O, et al. Burosumab resulted in better clinical outcomes than continuation with conventional therapy in younger and older children with X-linked hypophosphatemia. Journal of Bone and Mineral Research. 2019;34(Supplement 1):12. | publication type (inc. abstract only) |
| Kulikova K, Kolodkina A, Vasiliev E, Petrov V, Kenis V, Petrov M, et al. Clinical and genetic characteristics of 168 Russian patients with hypophosphatemic rickets. Hormone Research in Paediatrics. 2019;91(Supplement 1):375. | publication type (inc. abstract only) |
| Liu ES, Carpenter TO, Gundberg CM, Simpson CA, Insogna KL. Calcitonin administration in X-linked hypophosphatemia. The New England journal of medicine. 2011;364(17):1678-80. | publication type (inc. abstract only) |
| Martos Moreno GA, Aparicio C, de Lucas C, Gil Pena H, Argente J. X-linked hypophosphatemic rickets due to mutations in PHEX: Clinical and evolutionary variability. Anales de Pediatria. 2016;85(1):41-3. | language |
| McNair SL, Stickler GB. Growth in familial hypophosphatemic vitamin D-resistant rickets. The Journal of pediatrics. 1969;74(5):828-9. | duplicate |

| | |
|---|---|
| Mehls O, Manz F, Kettenmann K, Bonjour JP, Trechsel U. Effect of calcitriol on serum 1,25 (OH) 2D3 levels and on renal phosphorous threshold in X-linked hypophosphatemic rickets. Advances in experimental medicine and biology. 1984;178:411-3. | publication type (inc. abstract only) |
| Nabeshima Y, Sato T, Zukeran H, Komatsu R, Nakano S, Ichihashi Y, et al. Fibroblast growth factor 23 levels in cord and peripheral blood during early neonatal period as possible predictors of affected offspring of X-linked hypophosphatemic rickets: report of three female cases from two pedigrees. Journal of pediatric endocrinology & metabolism : JPEM. 2023;36(8):786-90. | irrelevant outcomes only |
| Nilsson O, Imel E, Whyte M, Munns C, Portale A, Ward L, et al. Burosumab showed greater improvement in phosphate metabolism, rickets, and bowing than continuation with conventional therapy in children with X-linked hypophosphatemia (XLH). Calcified Tissue International. 2019;104(Supplement 1):S4. | publication type (inc. abstract only) |
| Padidela R, Nilsson O, Makitie O, Beck-Nielsen S, Ariceta G, Schnabel D, et al. The international X-linked hypophosphataemia (XLH) registry (NCT03193476): rationale for and description of an international, observational study. Orphanet journal of rare diseases. 2020;15(1):172. | publication type (inc. abstract only) |
| Padidela R, Nilsson O, Linglart A, Makitie O, Beck-Nielsen S, Ariceta G, et al. X-linked hypophosphatemia registry-an international prospective patient registry. Hormone Research in Paediatrics. 2018;90(Supplement 1):177. | publication type (inc. abstract only) |
| Paloian NJ, Nemeth BA, Modaff P, Steiner R. Improvement in biomarkers in pediatric x-linked hypophosphatemic rickets after 1 year of treatment with burosumab. Journal of the American Society of Nephrology. 2019;30:475-6. | publication type (inc. abstract only) |
| Pericas N, Ibars Z, Rufach A, Vives I, Vique C, Madrid A, et al. X linked Hypophosphatemic rickets. Pediatric Nephrology. 2010;25(3):577-8. | publication type (inc. abstract only) |
| Perwad F, Portale AA. Burosumab Therapy for X-Linked Hypophosphatemia and Therapeutic Implications for CKD. Clinical journal of the American Society of Nephrology : CJASN. 2019;14(7):1097-9. | publication type (inc. abstract only) |
| Poon KS, Sng AA, Ho CW, Koay ES-C, Loke KY. Genetic Testing Confirmed the Early Diagnosis of X-Linked Hypophosphatemic Rickets in a 7-Month-Old Infant. Journal of investigative medicine high impact case reports. 2015;3(3):2324709615598167. | irrelevant treatment (not early vs late, not early ToI) |
| Portale AA, Martin JS, Carpenter T. KRN23, a Fully Human Monoclonal Antibody to FGF23, Reverses Renal Phosphate Wasting and Improves Rickets in Children with X-Linked | publication type (inc. abstract only) |

| | |
|---|---|
| Hypophosphatemia. Journal of the American Society of Nephrology. 2016;27:218A. | |
| Portale AA, Imel E, De Beur SJ, Munns C, Pitukcheewanont P, Martin JS, et al. Postprandial serum phosphorus and calcium concentrations in adults and children with X-linked hypophosphatemia (XLH) are within normal range during burosumab treatment. Journal of the American Society of Nephrology. 2019;30:476. | publication type (inc. abstract only) |
| Portale AA, Imel E, Munns C, Ward LM, Whyte M, Simmons J, et al. Burosumab improved rickets and clinical outcomes compared to conventional therapy in children with XLH. Journal of the American Society of Nephrology. 2018;29:857. | publication type (inc. abstract only) |
| Rafaelsen S, Raeder H, Johansson S, Bjerknes R. Hypophosphatemic rickets in norwegian children: Genotypes, phenotypes, and complications. Hormone Research in Paediatrics. 2014;82(SUPPL. 1):185. | publication type (inc. abstract only) |
| Rothenbuhler A, Esterle L, Gueorguieva I, Salles J-P, Mignot B, Colle M, et al. Two-year recombinant human growth hormone (rhGH) treatment is more effective in pre-pubertal compared to pubertal short children with X-linked hypophosphatemic rickets (XLHR). Growth hormone & IGF research : official journal of the Growth Hormone Research Society and the International IGF Research Society. 2017;36:11-5. | irrelevant treatment (not early vs late, not early ToI) |
| Rothenbuhler A, Esterle L, Gueorguieva I, Salles JP, Mignot B, Bougneres P, et al. Two-year growth hormone treatment is only effective in pre-pubertal compared to pubertal patients in short children with X-linked hypophosphatemic rickets. Hormone Research in Paediatrics. 2012;78(SUPPL. 1):186. | publication type (inc. abstract only) |
| Russell RGG, Smith R, Preston C, Walton RJ, Woods CG, Henderson RG, et al. The effect of 1,25 dihydroxycholecalciferol on renal tubular reabsorption of phosphate, intestinal absorption of calcium and bone histology in hypophosphataemic renal tubular rickets. Clinical Science and Molecular Medicine. 1975;48(3):177-86. | >=10% adults + not reported separately |
| Schumacher M, Thieme M, Nissen S, Werner R, Jueppner H, Herting E, et al. Genetic characterization and genotype-phenotype correlation of a large cohort of patients with hypophosphatemic rickets. European Journal of Pediatrics. 2009;168(3):383. | publication type (inc. abstract only) |
| Siklar Z, Turan S, Bereket A, Abaci A, Bas F, Demir K, et al. Nationwide hypophosphatemic rickets study. Hormone Research in Paediatrics. 2018;90(Supplement 1):109-10. | publication type (inc. abstract only) |

| | |
|---|---|
| Skalova S, Kopecka M, Filipsky T. TREATMENT OPTIONS FOR X-LINKED HYPOPHOSPATAEMIA. Pediatric Nephrology. 2022;37(11):2905. | publication type (inc. abstract only) |
| Verge CF, Cowell CT, Howard NJ, Donaghue KC, Silink M. Growth in children with X-linked hypophosphataemic rickets. Acta paediatrica (Oslo, Norway : 1992) Supplement. 1993;388:70-6. | irrelevant treatment (not early vs late, not early ToI) |
| Verge CF, Lam A, Simpson JM, Cowell CT, Howard NJ, Silink M. Effects of therapy in X-linked hypophosphatemic rickets. The New England journal of medicine. 1991;325(26):1843-8. | irrelevant treatment (not early vs late, not early ToI) |
| Ward LM, Glorieux FH, Whyte MP, Munns CF, Portale AA, Hogler W, et al. Effect of Burosumab Compared With Conventional Therapy on Younger vs Older Children With X-linked Hypophosphatemia. The Journal of clinical endocrinology and metabolism. 2022;107(8):e3241-e53. | irrelevant treatment (not early vs late, not early ToI) |
| Ward LM, Imel EA, Skrinar A, San Martin J. Burosumab resulted in greater improvement in clinical outcomes than continuation with conventional therapy in younger (1-4 years-old) and older (5-12 years-old) children with X-linked hypophosphatemia (XLH). Hormone Research in Paediatrics. 2019;92(Supplement 1):4. | publication type (inc. abstract only) |
| Zivicnjak M, Schnabel D, Billing H, Staude H, Filler G, Querfeld U, et al. Age-related stature and linear body segments in children with X-linked hypophosphatemic rickets. Pediatric nephrology (Berlin, Germany). 2011;26(2):223-31. | outcomes of treatment not reported comparatively |

#### fHLH

| **Reference** | **Reason for exclusion (Q4)** |
|---|---|
| Abbas AAH, Tapp HE, Rice MS, Mangos HM. Familial haemophagocytic lymphohistocytosis: A review of six cases. HAEMA. 2004;7(2):233-9. | FT not retrievable |
| Aksionau A, Wei EX. Accuracy of the criteria for hemophagocytic lymphohistiocytosis. International journal of clinical and experimental pathology. 2020;13(12):3139-48. | irrelevant outcomes only |
| Aksu Uzunhan T, Caliskan M, Karaman S, Aydin K, Devecioglu O. A rare cause of acute cerebellar ataxia: familial hemophagocytic lymphohistiocytosis. Pediatric neurology. 2014;51(3):465-6. | publication type (inc. abstract only) |
| Alhumaidan W, Al-Otaibi A, Elyamany G, Alabbas F, Ali TB. Familial hemophagocytic lymphohistiocytosis induced by SARS-CoV-2. Pediatric hematology and oncology. 2021;38(4):406-9. | publication type (inc. abstract only) |

| | |
|---|---|
| Al-Lamki Z, Wali YA, Pathare A, Ericson KG, Henter J-I. Clinical and genetic studies of familial hemophagocytic lymphohistiocytosis in Oman: need for early treatment. Pediatric hematology and oncology. 2003;20(8):603-9. | irrelevant treatment (not early vs late, not early ToI) |
| Bujan W, Schandene L, Ferster A, De Valck C, Goldman M, Sariban E. Abnormal T-cell phenotype in familial erythrophagocytic lymphohistiocytosis. Lancet (London, England). 1993;342(8882):1296. | publication type (inc. abstract only) |
| Du Y, Li L, Wang XY, Chen L, Jia SH, Mi R. Familial Hemophagocytic Lymphohistiocytosis Type 3. Indian journal of pediatrics. 2020;87(10):861. | publication type (inc. abstract only) |
| Fatima Z, Khan A, Tariq U, Sohail MS. Hemophagocytic Lymphohistiocytosis: A Case Series. Cureus. 2018;10(4):e2545. | >=10% adults + not reported separately |
| Ferreira M, Martins J, Silvestre C, Abadesso C, Matias E, Loureiro H, et al. Familial haemophagocytic lymphohistiocytosis: two case reports. BMJ case reports. 2010;2010. | irrelevant treatment (not early vs late, not early ToI) |
| Filipovich AH. Life-threatening hemophagocytic syndromes: current outcomes with hematopoietic stem cell transplantation. Pediatric transplantation. 2005;9 Suppl 7:87-91. | publication type (inc. abstract only) |
| Fiorillo A, Catera P, Guarino A, Grimaldi M, Menna G, Migliorati R, et al. FAMILIAL ERYTHROPHAGOCYTIC LYMPHOHISTIOCYTOSIS - ADVERSE PROGNOSTIC-SIGNIFICANCE OF DELAYED DIAGNOSIS. Haematologica. 1993;78(4):242-4. | irrelevant treatment (not early vs late, not early ToI) |
| Fiorillo A, Catera P, Guarino A, Grimaldi M, Menna G, Migliorati R, et al. Familial erythrophagocytic lymphohistiocytosis: adverse prognostic significance of delayed diagnosis. Haematologica. 1993;78(4):242-4. | publication type (inc. abstract only) |
| Fischer A, Cerf-Bensussan N, Blanche S, Le Deist F, Bremard-Oury C, Leverger G, et al. Allogeneic bone marrow transplantation for erythrophagocytic lymphohistiocytosis. The Journal of pediatrics. 1986;108(2):267-70. | publication type (inc. abstract only) |
| Greenmyer JR, Thompson WS, Mavis S, Hassan S, Weckwerth J, Hobbs C, et al. Neonatal familial hemophagocytic lymphohistiocytosis diagnosed with ultrarapid whole-genome sequencing. Pediatric blood & cancer. 2023;70(1):e29871. | publication type (inc. abstract only) |
| Herman TE, Siegel MJ. Familial hemophagocytic lymphohistiocytosis. Journal of perinatology : official journal of the California Perinatal Association. 2010;30(5):363-5. | publication type (inc. abstract only) |
| Hofer-Popow I, Dworzak M, Hutter C, Mann G, Holter W, Matthes-Leodolter S, et al. Allogeneic stem cell transplantation with reduced intensity regimen as successful therapy for primary | publication type (inc. abstract only) |

| | |
|---|---|
| haemophagocytic lymphohistiocytosis: A single center experience. Oncology Research and Treatment. 2018;41(Supplement 4):62. | |
| Hoseynpour-Feyzi A, Tavakoly HR, Fayyazi A, Dezhakam A, Behbahan AG, Shhverdi M. Hemophagocytic lymphohistiocytosis: A four-year experience. Research Journal of Medical Sciences. 2008;2(3):105-8. | irrelevant treatment (not early vs late, not early ToI) |
| Janka GE. Familial hemophagocytic lymphohistiocytosis: therapy in the German experience. Pediatric hematology and oncology. 1989;6(3):227-31. | irrelevant treatment (not early vs late, not early ToI) |
| Janka GE. Familial hemophagocytic lymphohistiocytosis: therapy in the German experience. Pediatric hematology and oncology. 1989;6(3):227-31. | publication type (inc. abstract only) |
| Kumar S, Tahlan A, Cruz SD, Gupta M, Gupta V. Hemophagocytic Lymphohistiocytosis (HLH): Case Series. Indian Journal of Hematology and Blood Transfusion. 2022;38(Supplement 1):S83-S4. | publication type (inc. abstract only) |
| Lehmberg K, Ledig S, Wustrau K, Kontny U, Westphal S, Hundsdorfer P, et al. ETOPOSIDE FOR PRIMARY HLH - BETTER THAN ITS REPUTATION. Pediatric Blood and Cancer. 2023;70(Supplement 1). | publication type (inc. abstract only) |
| Liao C-H, Lee N-C, Jou S-T, Chiang B-L, Yu H-H. UNC13D mutation presenting as fulminant familial hemophagocytic lymphohistiocytosis. Journal of microbiology, immunology, and infection = Wei mian yu gan ran za zhi. 2020;53(6):1039-41. | publication type (inc. abstract only) |
| Lucchini G, Marsh RA, Gilmour K, Worth A, Rao A, Booth C, et al. Treatment dilemmas in asymptomatic children with primary haemophagocytic lymphohistiocytosis. Blood. 2017;130(Supplement 1). | publication type (inc. abstract only) |
| Lucchini G, Marsh R, Gilmour K, Worth A, Nademi Z, Rao A, et al. Treatment dilemmas in asymptomatic children with primary haemophagocytic lymphohistiocytosis. Bone Marrow Transplantation. 2019;53:730. | publication type (inc. abstract only) |
| Rego I, Severino M, Micalizzi C, Faraci M, Pende D, Dufour C, et al. Neuroradiologic findings and follow-up with magnetic resonance imaging of the genetic forms of haemophagocytic lymphohistiocytosis with CNS involvement. Pediatr Blood Cancer. 2012;58(5):810-4. | irrelevant treatment (not early vs late, not early ToI) |
| Rodrigues JB, Nasr BP, Cypriano MDS. Hemophagocytic lymphohistiocytosis: presentation and outcome of twenty-one | irrelevant treatment (not early vs late, not early ToI) |

| | |
|---|---|
| patients at a single institution. Hematology, transfusion and cell therapy. 2022;44(4):485-90. | |
| Shah N, Wolff JA. Long term management of familial erythrophagocytic lymphohistiocytosis. Pediatric Research. 1973;7(4):360. | publication type (inc. abstract only) |
| Shamsian BS, Rezaei N, Alavi S, Hedayat M, Amin Asnafi A, Pourpak Z, et al. Primary hemophagocytic lymphohistiocytosis in Iran: report from a single referral center. Pediatric hematology and oncology. 2012;29(3):215-9. | >=10% other conditions + not reported separately |

#### MCADD

| **Reference** | **Reason for exclusion (Q4)** |
|---|---|
| Abdenur JE, Chamoles NA, Specola N, Schenone AB, Jorge L, Guinle A, et al. MCAD deficiency - Acylcarnitines (AC) by tandem mass spectrometry (MS-MS) are useful to monitor dietary treatment. In: Quant PA, Eaton S, editors. Current Views of Fatty Acid Oxidation and Ketogenesis: From Organelles to Point Mutations. Advances in Experimental Medicine and Biology. 466. New York: Kluwer Academic/Plenum Publ; 1999. p. 353-63. | duplicate |
| Andresen BS, Bross P, Knudsen I, Winter V, Kolvraa S, Bolund L, et al. Medium-chain acyl-CoA dehydrogenase (MCAD) deficiency due to heterozygosity for the common mutation and an allele resulting in low levels of MCAD mRNA. Journal of inherited metabolic disease. 1994;17(3):275-8. | irrelevant outcomes only |
| Balci MC, Karaca M, Ergul Y, Omeroglu RE, Demirkol M, Gokcay GF. Cardiologic evaluation of Turkish mitochondrial fatty acid oxidation disorders. Pediatrics international : official journal of the Japan Pediatric Society. 2022;64(1):e15317. | irrelevant treatment (not early vs late, not early ToI) |
| Carroll JC, Gibbons CA, Blaine SM, Cremin C, Dorman H, Honeywell C, et al. Genetics: newborn screening for MCAD deficiency. Canadian family physician Medecin de famille canadien. 2009;55(5):487. | publication type (inc. abstract only) |
| Casey JL. MCAD deficiency in the Holderman Mennonite population in central Kansas. Kansas medicine : the journal of the Kansas Medical Society. 1992;93(11):306-8. | irrelevant outcomes only |
| Clayton PT, Doig M, Ghafari S, Meaney C, Taylor C, Leonard JV, et al. Screening for medium chain acyl-CoA dehydrogenase deficiency using electrospray ionisation tandem mass spectrometry. Archives of disease in childhood. 1998;79(2):109-15. | irrelevant treatment (not early vs late, not early ToI) |

| | |
|---|---|
| Curtis D, Blakemore AI, Engel PC, Macgregor D, Besley G, Kolvraa S, et al. Heterogeneity for mutations in medium chain acyl-CoA dehydrogenase deficiency in the UK population. Clinical genetics. 1991;40(4):283-6. | irrelevant outcomes only |
| Ding JH, Bross P, Yang BZ, Iafolla AK, Millington DS, Roe CR, et al. Genetic heterogeneity in MCAD deficiency: frequency of K329E allele and identification of three additional mutant alleles. Progress in clinical and biological research. 1992;375:479-88. | irrelevant outcomes only |
| Ding JH, Roe CR, Iafolla AK, Chen YT. Medium-chain acyl-coenzyme A dehydrogenase deficiency and sudden infant death. The New England journal of medicine. 1991;325(1):61-2. | publication type (inc. abstract only) |
| Dong L, Ji C, Xu J, Cui Y. Screening and follow-up results of neonate medium-chain acyl-CoA dehydrogenase deficiency in Zibo, Shandong province. Zhejiang da xue xue bao Yi xue ban = Journal of Zhejiang University Medical sciences. 2022;51(3):284-9. | language |
| Fowler DJ, Picker J, Waisbren SE, Levy HL. Neonatal screening for medium--chain acyl-CoA dehydrogenase deficiency. Lancet (London, England). 2002;359(9306):628. | publication type (inc. abstract only) |
| Frazier DM, Millington DS, McCandless SE, Koeberl DD, Weavil SD, Chaing SH, et al. The tandem mass spectrometry newborn screening experience in North Carolina: 1997-2005. Journal of inherited metabolic disease. 2006;29(1):76-85. | irrelevant outcomes only |
| Ged C, el Sebai H, de Verneuil H, Parrot-Rouleau F. Is genotyping useful for the screening of medium-chain acyl-CoA dehydrogenase deficiency in France? Journal of inherited metabolic disease. 1995;18(2):253-6. | irrelevant outcomes only |
| Goncalves MM, Marcao A, Sousa C, Nogueira C, Ferreira F, Fonseca H, et al. Portuguese Neonatal Screening Programme: A Retrospective Cohort Study of 18 Years of MS/MS. Endocrine, metabolic & immune disorders drug targets. 2023. | publication type (inc. abstract only) |
| Gregersen N, Winter V, Kolvraa S, Andresen BS, Bross P, Blakemore A, et al. Molecular analysis of medium-chain acyl-CoA dehydrogenase deficiency: a diagnostic approach. Progress in clinical and biological research. 1992;375:441-52. | irrelevant outcomes only |
| Huidekoper HH, Schneider J, Westphal T, Vaz FM, Duran M, Wijburg FA. Prolonged moderate-intensity exercise without and with L-carnitine supplementation in patients with MCAD deficiency. Journal of inherited metabolic disease. 2006;29(5):631-6. | irrelevant treatment (not early vs late, not early ToI) |

| | |
|---|---|
| Iafolla AK, Thompson RJ, Jr., Roe CR. Medium-chain acyl-coenzyme A dehydrogenase deficiency: clinical course in 120 affected children. The Journal of pediatrics. 1994;124(3):409-15. | outcomes of treatment not reported comparatively |
| Inagaki T, Ohishi N, Bachmann C, Ghisla S, Tsukagoshi N, Udaka S, et al. IMMUNOCHEMICAL AND MOLECULAR ANALYSIS OF MEDIUM-CHAIN ACYL COA DEHYDROGENASE-DEFICIENCY. J Clin Biochem Nutr. 1990;8(1):1-8. | irrelevant outcomes only |
| Jager EA, Kuijpers MM, Bosch AM, Mulder MF, Gozalbo ER, Visser G, et al. A nationwide retrospective observational study of population newborn screening for medium-chain acyl-CoA dehydrogenase (MCAD) deficiency in the Netherlands. Journal of inherited metabolic disease. 2019;42(5):890-7. | irrelevant outcomes only |
| Janeiro P, Jotta R, Ramos R, Florindo C, Ventura FV, Vilarinho L, et al. Follow-up of fatty acid -oxidation disorders in expanded newborn screening era. European Journal of Pediatrics. 2019;178(3):387-94. | duplicate |
| Janeiro P, Jotta R, Ramos R, Florindo C, Ventura FV, Vilarinho L, et al. Follow-up of fatty acid beta-oxidation disorders in expanded newborn screening era. European journal of pediatrics. 2019;178(3):387-94. | irrelevant outcomes only |
| Janzen N, Hofmann AD, Schmidt G, Das AM, Illsinger S. Non-invasive test using palmitate in patients with suspected fatty acid oxidation defects: disease-specific acylcarnitine patterns can help to establish the diagnosis. Orphanet journal of rare diseases. 2017;12(1):187. | irrelevant outcomes only |
| Kelly DP, Whelan AJ, Hale DE, Rinaldo P, Rutledge SL, Zhang Z, et al. Molecular characterization of medium-chain acyl-CoA dehydrogenase deficiency causing sudden death. Progress in clinical and biological research. 1992;375:463-72. | irrelevant outcomes only |
| Kennedy S, Potter BK, Wilson K, Fisher L, Geraghty M, Milburn J, et al. The first three years of screening for medium chain acyl-CoA dehydrogenase deficiency (MCADD) by newborn screening ontario. BMC pediatrics. 2010;10:82. | irrelevant outcomes only |
| Kim SZ, Jeon YM, Song WJ, Lee SH, Yamaguchi S, Strauss A. Clinical and molecular evaluation of 15 Korean mcad patients detected by newborn screening. Journal of Inborn Errors of Metabolism and Screening. 2017;5:221. | publication type (inc. abstract only) |
| Kirk JM, Laing IA, Smith N, Uttley WS. Neonatal presentation of medium-chain acyl-CoA dehydrogenase deficiency in two families. Journal of inherited metabolic disease. 1996;19(3):370-1. | publication type (inc. abstract only) |

| | |
|---|---|
| Landau YE, Lichter-Konecki U, Levy HL. Genomics in newborn screening. J Pediatr. 2014;164(1):14-9. | study design |
| Landau YE, Waisbren SE, Chan LMA, Levy HL. Long-term outcome of expanded newborn screening at Boston children's hospital: benefits and challenges in defining true disease. Journal of inherited metabolic disease. 2017;40(2):209-18. | study design |
| Lobo AM, Gaspar A, Cabral A, Silva MFB, Ventura F, Almeida IT, et al. Medium chain acyl-CoA dehydrogenase deficiency: 19 cases. Journal of Inherited Metabolic Disease. 2007;30:49-. | publication type (inc. abstract only) |
| Longo N, Anderson DR, Viau K, Pasquali M. Clinical and biochemical outcome of patients with medium-chain acyl-CoA dehydrogenase deficiency. Journal of Inherited Metabolic Disease. 2016;39(Supplement 1):S152-S3. | publication type (inc. abstract only) |
| Marsden D, Bedrosian CL, Vockley J. Impact of newborn screening on the reported incidence and clinical outcomes associated with medium- and long-chain fatty acid oxidation disorders. Genet Med. 2021;23(5):816-29. | >=10% other conditions + not reported separately |
| Marsden D. Expanded newborn screening by tandem mass spectrometry: the Massachusetts and New England experience. The Southeast Asian journal of tropical medicine and public health. 2003;34 Suppl 3:111-4. | publication type (inc. abstract only) |
| Mesbah Z, Sing Ho K, Fitzsimons P, Monavari AA, Crushell E, Mayne PD. Medium Chain Acyl-CoA Dehydrogenase Deficiency (MCADD) in the Irish Paediatric Population. Irish medical journal. 2020;112(10):1016. | irrelevant outcomes only |
| Morris AA, Taylor RW, Lightowlers RN, Aynsley-Green A, Bartlett K, Turnbull DM. Medium chain acyl-CoA dehydrogenase deficiency caused by a deletion of exons 11 and 12. Human molecular genetics. 1995;4(4):747-9. | duplicate |
| Morris AAM, Taylor RW, Lightowlers RN, Aynsleygreen A, Bartlett K, Turnbull DM. MEDIUM-CHAIN ACYL-COA DEHYDROGENASE DEFICIENCY CAUSED BY A DELETION OF EXON-11 AND EXON-12. Human Molecular Genetics. 1995;4(4):747-9. | publication type (inc. abstract only) |
| Nennstiel-Ratzel U, Arenz S, Maier EM, Knerr I, Baumkotter J, Roschinger W, et al. Reduced incidence of severe metabolic crisis or death in children with medium chain acyl-CoA dehydrogenase deficiency homozygous for c.985A>G identified by neonatal screening. Molecular genetics and metabolism. 2005;85(2):157-9. | irrelevant outcomes only |
| Opdal SH, Vege A, Saugstad OD, Rognum TO. Is the medium-chain acyl-CoA dehydrogenase G985 mutation involved in sudden | publication type (inc. abstract only) |

| | |
|---|---|
| infant death in Norway? [6]. European Journal of Pediatrics. 1995;154(2):166-7. | |
| Pollitt RJ, Manning NJ, Olpin SE, Young ID. Prenatal diagnosis of a defect in medium-chain fatty acid oxidation. Journal of Inherited Metabolic Disease. 1994;17(3):279-82. | publication type (inc. abstract only) |
| Prasad C, Speechley KN, Dyack S, Rupar CA, Chakraborty P, Kronick JB. Incidence of medium-chain acyl-CoA dehydrogenase deficiency in Canada using the Canadian Paediatric Surveillance Program: Role of newborn screening. Paediatrics & child health. 2012;17(4):185-9. | irrelevant outcomes only |
| Purevsuren J, Hasegawa Y, Fukuda S, Kobayashi H, Mushimoto Y, Yamada K, et al. Clinical and molecular aspects of Japanese children with medium chain acyl-CoA dehydrogenase deficiency. Molecular genetics and metabolism. 2012;107(1-2):237-40. | irrelevant outcomes only |
| Real LM, Gayoso AJ, Olivera M, Caruz A, Ruiz A, Gayoso F. Detection of nucleotide c985 A->G mutation of medium-chain acyl-CoA dehydrogenase gene by real-time PCR. Clinical Chemistry. 2001;47(5):958-9. | FT not retrievable |
| Sander S, Janzen N, Janetzky B, Scholl S, Steuerwald U, Schafer J, et al. Neonatal screening for medium chain acyl-CoA deficiency: high incidence in Lower Saxony (northern Germany). European journal of pediatrics. 2001;160(5):318-9. | irrelevant outcomes only |
| Seddon HR, Green A, Gray RG, Leonard JV, Pollitt RJ. Regional variations in medium-chain acyl-CoA dehydrogenase deficiency. Lancet (London, England). 1995;345(8942):135-6. | irrelevant outcomes only |
| Stanley CA, Hale DE, Coates PM. Medium-chain acyl-CoA dehydrogenase deficiency. Progress in clinical and biological research. 1990;321:291-302. | study design |
| Strauss AW, Duran M, Zhang ZF, Alpers R, Kelly DP. Molecular analysis of medium chain acyl-CoA dehydrogenase deficiency. Progress in clinical and biological research. 1990;321:609-23. | irrelevant outcomes only |
| Szalai C, Czinner A, Revai K. The frequency of medium-chain acyl-CoA dehydrogenase G985 mutation in the Hungarian population [4]. European Journal of Pediatrics. 1996;155(3):256. | publication type (inc. abstract only) |
| Tanner S, Sharrard M, Cleary M, Walter J, Wraith E, Lee P, et al. Screening for medium chain acyl-CoA dehydrogenase deficiency has still not been evaluated. BMJ (Clinical research ed). 2001;322(7278):112. | publication type (inc. abstract only) |
| Thompson SM, Dennison B, Wiley V, Carpenter K, Wilcken B, Bhattacharya K, et al. Dietetic issues in the management of medium chain acyl COA dehydrogenase deficiency diagnosed by | publication type (inc. abstract only) |

| | |
|---|---|
| newborn screening. Molecular Genetics and Metabolism. 2009;98(1-2):117. | |
| Udvari S, Bross P, Andresen BS, Gregersen N, Engel PC. Biochemical characterisation of mutations of human medium-chain acyl-CoA dehydrogenase. Advances in Experimental Medicine and Biology. 2000;466:387-93. | irrelevant outcomes only |
| Udvari S, Bross P, Andresen BS, Gregersen N, Engel PC. Biochemical characterisation of mutations of human medium-chain acyl-CoA dehydrogenase. In: Quant PA, Eaton S, editors. Current Views of Fatty Acid Oxidation and Ketogenesis: From Organelles to Point Mutations. Advances in Experimental Medicine and Biology. 466. New York: Kluwer Academic/Plenum Publ; 1999. p. 387-93. | duplicate |
| Van Gennip AH, Bakker HD, Duran M, Van Oudheusden LJ. The diagnosis and treatment of a patient with medium-chain acyl-CoA dehydrogenase deficiency: Overnight fasting does not result in the expected urinary metabolite profile. Journal of Inherited Metabolic Disease. 1986;9(SUPPL. 2):293-6. | publication type (inc. abstract only) |
| Van Hove JLK, Myers S, Kerckhove KV, Freehauf C, Bernstein L. Acute nutrition management in the prevention of metabolic illness: a practical approach with glucose polymers. Molecular genetics and metabolism. 2009;97(1):1-3. | publication type (inc. abstract only) |
| Wilcken B, Carpenter KH, Hammond J. Neonatal symptoms in medium chain acyl coenzyme A dehydrogenase deficiency. Archives of disease in childhood. 1993;69(3 Spec No):292-4. | irrelevant treatment (not early vs late, not early ToI) |
| Wilcken B, Hammond J, Silink M. Morbidity and mortality in medium chain acyl coenzyme A dehydrogenase deficiency. Arch Dis Child. 1994;70(5):410-2. | outcomes of treatment not reported comparatively |
| Yang C, Shi C, Zhou C, Wan Q, Zhou Y, Chen X, et al. Screening and follow-up results of fatty acid oxidative metabolism disorders in 608 818 newborns in Jining, Shandong province. Zhejiang da xue xue bao Yi xue ban = Journal of Zhejiang University Medical sciences. 2021;50(4):472-80. | language |
| Yang N, Gong L-F, Zhao J-Q, Yang H-H, Ma Z-J, Liu W, et al. Inborn errors of metabolism detectable by tandem mass spectrometry in Beijing. Journal of pediatric endocrinology & metabolism : JPEM. 2020;33(5):639-45. | irrelevant outcomes only |
| Yokota I, Coates PM, Hale DE, Rinaldo P, Tanaka K. The molecular basis of medium chain acyl-CoA dehydrogenase deficiency: survey and evolution of 985A----G transition, and identification of five rare types of mutation within the medium chain acyl-CoA | irrelevant outcomes only |

| |
|---|
| dehydrogenase gene. Progress in clinical and biological research. 1992;375:425-40. |

### Review of genomic studies of paediatric cohorts reporting penetrance for pathogenic variants

| **Reference** | **Reason for exclusion** |
|---|---|
| Howes RE, Chan ER, Rakotomanga TA, Schulte S, Gibson J, Zikursh M, et al. Prevalence and genetic variants of G6PD deficiency among two Malagasy populations living in Plasmodium vivax-endemic areas. Malar J. 2017;16(1):139. | >10% not newborns and not reported separately |
| Nuinoon M, Krithong R, Pramtong S, Sasuk P, Ngeaiad C, Chaimusik S, et al. Prevalence of G6PD deficiency and G6PD variants amongst the southern Thai population. Peerj. 2022;10:e14208. | >10% not newborns and not reported separately |
| Nivoloni Kde A, da Silva-Costa SM, Pomilio MC, Pereira T, Lopes Kde C, de Moraes VC, et al. Newborn hearing screening and genetic testing in 8974 Brazilian neonates. Int J Pediatr Otorhinolaryngol. 2010;74(8):926-9. | sequencing not first line |
| Kraan CM, Bui QM, Field M, Archibald AD, Metcalfe SA, Christie LM, et al. FMR1 allele size distribution in 35,000 males and females: a comparison of developmental delay and general population cohorts. Genet Med. 2018;20(12):1627-34. | not direct sequencing |
| Norrgard KJ, Pomponio RJ, Swango KL, Hymes J, Reynolds T, Buck GA, et al. Double mutation (A171T and D444H) is a common cause of profound biotinidase deficiency in children ascertained by newborn screening the the United States. Mutations in brief no. 128. Online. Hum Mutat. 1998;11(5):410. | >10% population at risk/symptoms/disease |
| Grueso Ceron AL, Arturo-Terranova D, Satizabal Soto JM. Characterization of genomic variants associated with congenital heart disease in patients from southwestern Colombian. Heliyon. 2024;10(1):e23678. | outcomes not relevant |
| Ellis G, Wilcock AR, Goldberg DM. Experience of routine live-birth screening for galactosaemia in a British hospital, with emphasis on heterozygote detection. Arch Dis Child. 1972;47(251):34-40. | not direct sequencing |

| | |
|---|---|
| Park KS. Carrier frequency and predicted genetic prevalence of Pompe disease based on a general population database. Mol Genet Metab Rep. 2021;27:100734. | outcomes not relevant |
| Kaler SG, Parad RB, Bhattacharjee A. Newborn Screening for Menkes Disease Based on Targeted-Next Generation Sequencing. Mol Ther. 2020;28(4):321-. | outcomes not relevant |
| Milko LV, O'Daniel JM, DeCristo DM, Crowley SB, Foreman AKM, Wallace KE, et al. An Age-Based Framework for Evaluating Genome-Scale Sequencing Results in Newborn Screening. J Pediatr. 2019;209:68-76. | not direct sequencing |
| Morris-Rosendahl DJ, Edwards M, McDonnell MJ, John S, Alton EWFW, Davies JC, et al. Whole-Gene Sequencing of CFTR Reveals a High Prevalence of the Intronic Variant c.3874-4522AG in Cystic Fibrosis. American Journal of Respiratory and Critical Care Medicine. 2020;201(11):1438-41. | >10% population at risk/symptoms/disease |
| He S, Li J, Li DM, Yi S, Lu X, Luo Y, et al. Molecular characterization of alpha- and beta-thalassemia in the Yulin region of Southern China. Gene. 2018;655:61-4. | >10% population at risk/symptoms/disease |
| Mantravadi V, Bednarski JJ, Ritter MA, Gu H, Kolicheski AL, Horner C, et al. Immunological Findings and Clinical Outcomes of Infants With Positive Newborn Screening for Severe Combined Immunodeficiency From a Tertiary Care Center in the U.S. Front. 2021;12(no pagination). | not direct sequencing |
| He X, Li X, Guo Y, Zhao Y, Dong H, Dong J, et al. Newborn Screening of Genetic Mutations in Common Deafness Genes With Bloodspot-Based Gene Chip Array. Am J Audiol. 2018;27(1):57-66. | >10% not newborns and not reported separately |
| Liu Y, Chen M, Liu J, Mao A, Teng Y, Yan H, et al. Comprehensive Analysis of Congenital Adrenal Hyperplasia Using Long-Read Sequencing. Clin Chem. 2022;68(7):927-39. | >10% population at risk/symptoms/disease |
| Kemper AR, Knapp AA, Green NS, Comeau AM, Metterville DR, Perrin JM. Weighing the evidence for newborn screening for early-infantile Krabbe disease. Genet Med. 2010;12(9):539-43. | sequencing not first line |
| Liu Z, Zhang P, He X, Liu S, Tang S, Zhang R, et al. New multiplex real-time PCR approach to detect gene mutations for spinal muscular atrophy. BMC Neurol. 2016;16(1):141. | >10% population at risk/symptoms/disease |

| | |
|---|---|
| Hao Z, Fu D, Ming Y, Yang J, Huang Q, Lin W, et al. Large scale newborn deafness genetic screening of 142,417 neonates in Wuhan, China. PLoS ONE. 2018;13(4):e0195740. | sequencing not first line |
| Klaassen T, Teder M, Viikmaa M, Metspalu A. Neonatal screening for the cystic fibrosis main mutation DELTAF508 in Estonia. J Med Screen. 1998;5(1):16-9. | not direct sequencing |
| Lacerda L, Amaral O, Pinto R, Oliveira P, Aerts J, Miranda MCS. GAUCHER DISEASE - N370S GLUCOCEREBROSIDASE GENE-FREQUENCY IN THE PORTUGUESE POPULATION. Clin Genet. 1994;45(6):298-300. | not direct sequencing |
| Larsen TB, Lassen JF, Brandslund I, Byriel L, Petersen GB, Norgaard-Pedersen B. The Arg506Gln mutation (FV Leiden) among a cohort of 4188 unselected Danish newborns. Thromb Res. 1998;89(5):211-5. | not direct sequencing |
| Mercimek-Mahmutoglu S, Pop A, Kanhai W, Fernandez Ojeda M, Holwerda U, Smith D, et al. A pilot study to estimate incidence of guanidinoacetate methyltransferase deficiency in newborns by direct sequencing of the GAMT gene. Gene. 2016;575(1):127-31. | outcomes not relevant |
| Mercimek-Mahmutoglu S, Sinclair G, van Dooren SJ, Kanhai W, Ashcraft P, Michel OJ, et al. Guanidinoacetate methyltransferase deficiency: first steps to newborn screening for a treatable neurometabolic disease. Mol Genet Metab. 2012;107(3):433-7. | sequencing not first line |
| Park JE, Noh SJ, Oh M, Cho DY, Kim SY, Ki CS. Frequency of hereditary neuropathy with liability to pressure palsies (HNPP) due to 17p11.2 deletion in a Korean newborn population. Orphanet J Rare Dis. 2018;13(1):40. | condition not childhood onset/not monogenic |
| Lev A, Sharir I, Simon AJ, Levy S, Lee YN, Frizinsky S, et al. Lessons Learned From Five Years of Newborn Screening for Severe Combined Immunodeficiency in Israel. Journal of Allergy and Clinical Immunology: In Practice. 2022;10(10):2722-31.e9. | sequencing not first line |
| Lin Y, Lin C, Lin B, Zheng Z, Lin W, Chen Y, et al. Newborn screening for fatty acid oxidation disorders in a southern Chinese population. Heliyon. 2024;10(1):e23671. | >10% population at risk/symptoms/disease |
| Fitzgerald T, Duva S, Ostrer H, Pass K, Oddoux C, Ruben R, et al. The frequency of GJB2 and GJB6 mutations in the New York State newborn population: feasibility of genetic screening for hearing defects. Clin Genet. 2004;65(4):338-42. | sequencing not first line |
| Fortin CA, Girard L, Bonenfant C, Leblanc J, Cruz-Marino T, Blackburn ME, et al. Benefits of Newborn Screening for Vitamin D- | >10% population at risk/symptoms/disease |

| | |
|---|---|
| Dependant Rickets Type 1A in a Founder Population. Front Endocrinol (Lausanne). 2022;13(no pagination). | |
| Hansen PS, Norgaard-Petersen B, Meinertz H, Jensen HK, Hansen AB, Klausen IC, et al. Incidence of the apolipoprotein B-3500 mutation in Denmark. Clin Chim Acta. 1994;230(1):101-4. | not direct sequencing |
| Koizumi A, Matsuura N, Inoue S, Utsunomiya M, Nozaki J, Inoue K, et al. Evaluation of a mass screening program for lysinuric protein intolerance in the northern part of Japan. Genet Test. 2003;7(1):29-35. | not direct sequencing |
| Gasparini P, Arbustini E, Restagno G, Zelante L, Stanziale P, Gatta L, et al. Analysis of 31 CFTR mutations by polymerase chain reaction/oligonucleotide ligation assay in a pilot screening of 4476 newborns for cystic fibrosis. J Med Screen. 1999;6(2):67-9. | sequencing not first line |
| Kham SK, Quah TC, Loong AM, Tan PL, Fraser A, Chong SS, et al. A molecular epidemiologic study of thalassemia using newborns' cord blood in a multiracial Asian population in Singapore: results and recommendations for a population screening program. J Pediatr Hematol Oncol. 2004;26(12):817-9. | not direct sequencing |
| Li SX, Chen DL, Zhao SB, Guo LL, Feng HQ, Zhang XF, et al. Cordblood-Based High-Throughput Screening for Deafness Gene of 646 Newborns in Jinan Area of China. Clin. 2015;8(3):211-7. | sequencing not first line |
| Lin M, Yang LY, Xie DD, Chen JT, Nguba SM, Ehapo CS, et al. G6PD Deficiency and Hemoglobinopathies: Molecular Epidemiological Characteristics and Healthy Effects on Malaria Endemic Bioko Island, Equatorial Guinea. PLoS ONE. 2015;10(4):e0123991. | >10% not newborns and not reported separately |
| Kun L, Jiexiang H, Hua L, Junlin H, Yijun R, Lixian Z, et al. Genetic screening of 15 hearing loss variants in 77,647 neonates with clinical follow-up. Mol Genet Genomic Med. 2023:e2324. | sequencing not first line |
| Hohenfellner K, Bergmann C, Fleige T, Janzen N, Burggraf S, Olgemoller B, et al. Molecular based newborn screening in Germany: Follow-up for cystinosis. Mol Genet Metab Rep. 2019;21:100514. | sequencing not first line |
| Li YY, Xu J, Sun XC, Li HY, Mu K. Newborn screening and genetic variation of medium chain acyl-CoA dehydrogenase deficiency in the Chinese population. J Pediatr Endocrinol Metab. 2022;35(10):1264-71. | sequencing not first line |

| | |
|---|---|
| Lei J, Han L, Deng X, Long M, Xiao Y, Lin X, et al. [Analysis of results of concurrent hearing and deafness genetic screening and follow up of 33 911 newborns]. Chung Hua I Hsueh I Chuan Hsueh Tsa Chih. 2021;38(1):32-6. | Non-English |
| Johansson A, Guthenberg C, Ahlman H, Von Dobeln U, Hagenfeldt L. Prevalence of the 985A>G mutation in the medium-chain acyl-CoA dehydrogenase (MCAD) gene in Sweden. Scand J Clin Lab Invest. 1999;59(4):289-91. | not direct sequencing |
| Li YY, Xu J, Sun XC, Li HY, Mu K. Characteristics, differential diagnosis, individualized treatment, and prevention of hyperhomocysteinemia in newborns. Eur J Med Genet. 2023;66(10):104836. | sequencing not first line |
| Nami AH, Kabiri M, Motlagh FZ, Shirzadeh T, Fakhari N, Karimi A, et al. Genetic attributes of Iranian cystic fibrosis patients: the diagnostic efficiency of CFTR mutations in over a decade. Front. 2023;14:8. | >10% population at risk/symptoms/disease |
| Joseph J, Joseph A. Carrier frequency of connexin26 W24X mutation in the population of Kerala, India. Indian Journal of Otology. 2021;27(4):222-4. | not direct sequencing |
| Du Y, Huang L, Cheng X, Zhao L, Ruan Y, Ni T. Analysis of p.V37I compound heterozygous mutations in the GJB2 gene in Chinese infants and young children. Biosci. 2016;10(3):220-6. | >10% population at risk/symptoms/disease |
| Pang S, Spence DA, New MI. Newborn screening for congenital adrenal hyperplasia with special reference to screening in Alaska. Ann N Y Acad Sci. 1985;458:90-102. | not direct sequencing |
| Kingsmore SF. Newborn testing and screening by whole-genome sequencing. Genet Med. 2016;18(3):214-6. | outcomes not relevant |
| Guomei C, Luyan Z, Lingling D, Chunhong H, Shan C. Concurrent Hearing and Genetic Screening among Newborns in Ningbo, China. Comput. 2022;2022:1713337. | sequencing not first line |
| Zhang J, Wang H, Yan C, Guan J, Yin L, Lan L, et al. The Frequency of Common Deafness-Associated Variants Among 3,555,336 Newborns in China and 141,456 Individuals Across Seven Populations Worldwide. Ear Hear. 2023;44(1):232-41. | outcomes not relevant |
| Pavlova Z, Sarafov S, Todorov T, Kirov A, Chamova T, Gospodinova M, et al. Characterization of population genetic structure of | >10% population at risk/symptoms/disease |

| | |
|---|---|
| hereditary transthyretin amyloidosis in Bulgaria. Amyloid. 2021;28(4):219-25. | |
| Solomon BD, Pineda-Alvarez DE, Bear KA, Mullikin JC, Evans JP. Applying Genomic Analysis to Newborn Screening. Mol Syndromol. 2012;3(2):59-67. | outcomes not relevant |
| Suzuki M, West C, Beutler E. Large-scale molecular screening for galactosemia alleles in a pan-ethnic population. Hum Genet. 2001;109(2):210-5. | not direct sequencing |
| Tabor HK, Auer PL, Jamal SM, Chong JX, Yu JH, Gordon AS, et al. Pathogenic variants for Mendelian and complex traits in exomes of 6,517 European and African Americans: implications for the return of incidental results. Am J Hum Genet. 2014;95(2):183-93. | >10% not newborns and not reported separately |
| Vicario-Feliciano R, Hernandez-Hernandez CI, Camacho-Pastor IC, Martinez-Cruzado JC. A Custom-Made Newborn Screening Test for Wilson's Disease in Puerto Rico. Cureus. 2022;14(4):e24446. | not direct sequencing |
| Wen C, Yang X, Cheng X, Zhang W, Li Y, Wang J, et al. Optimized concurrent hearing and genetic screening in Beijing, China: A cross-sectional study. Biosci. 2023;17(2):148-59. | not direct sequencing |
| Wojcik MH, Zhang T, Ceyhan-Birsoy O, Genetti CA, Lebo MS, Yu TW, et al. Discordant results between conventional newborn screening and genomic sequencing in the BabySeq Project. Genet Med. 2021;23(7):1372-5. | >10% population at risk/symptoms/disease |
| Xiao F, Yan K, Wang H, Wu B, Hu L, Yang L, et al. Protocol of the China Neonatal Genomes Project: An observational study about genetic testing on 100,000 neonates. Pediatric Medicine. 2021;4(no pagination). | not direct sequencing |
| Xue S, Zhu J, Zhang H, Han L, Yang R, Dai P, et al. Association Analysis of Gene Sequencing by NeoSeq Combined with Tandem Mass Spectrum and Four Neonatal Diseases. Clin Lab. 2023;69(9):01. | not direct sequencing |
| Zeng X, Liu Z, Wang J, Zeng X. Combined hearing screening and genetic screening of deafness among Hakka newborns in China. Int J Pediatr Otorhinolaryngol. 2020;136:110120. | not direct sequencing |
| Zhang T, Shen Y, Xu Y, Wu D, Chen C, Yang R. Clinical, biochemical characteristics and genotype-phenotype analysis of congenital | >10% population at risk/symptoms/disease |

| | |
|---|---|
| hypothyroidism diagnosed by newborn screening in China. Clin Chim Acta. 2023;547:117459. | |
| Putcha GV, Bejjani BA, Bleoo S, Booker JK, Carey JC, Carson N, et al. A multicenter study of the frequency and distribution of GJB2 and GJB6 mutations in a large North American cohort. Genet Med. 2007;9(7):413-26. | >10% not newborns and not reported separately |
| Ramadevi R, Savithri HS, Devi ARR, Bittles AH, Rao NA. AN UNUSUAL DISTRIBUTION OF GLUCOSE-6-PHOSPHATE-DEHYDROGENASE DEFICIENCY OF SOUTH INDIAN NEWBORN POPULATION. Indian J Biochem Biophys. 1994;31(4):358-60. | not direct sequencing |
| Shinwari ZMA, Almesned A, Alakhfash A, Al-Rashdan AM, Faqeih E, Al-Humaidi Z, et al. The Phenotype and Outcome of Infantile Cardiomyopathy Caused by a Homozygous <i>ELAC2</i> Mutation. Cardiology. 2017;137(3):188-92. | not direct sequencing |
| Torres-Serrant M, Ramirez SI, Cadilla CL, Ramos-Valencia G, Santiago-Borrero PJ. Newborn screening for hermansky-pudlak syndrome type 3 in Puerto Rico. J Pediatr Hematol Oncol. 2010;32(6):448-53. | not direct sequencing |
| Upadhye DS, Jain D, Nair SB, Nadkarni AH, Ghosh K, Colah RB. First case of Hb Fontainebleau with sickle haemoglobin and other non-deletional alpha gene variants identified in neonates during newborn screening for sickle cell disorders. J Clin Pathol. 2012;65(7):654-9. | not direct sequencing |
| Waddell L, Wiley V, Carpenter K, Bennetts B, Angel L, Andresen BS, et al. Medium-chain acyl-CoA dehydrogenase deficiency: genotype-biochemical phenotype correlations. Mol Genet Metab. 2006;87(1):32-9. | sequencing not first line |
| Yamaguchi-Kabata Y, Yasuda J, Uruno A, Shimokawa K, Koshiba S, Suzuki Y, et al. Estimating carrier frequencies of newborn screening disorders using a whole-genome reference panel of 3552 Japanese individuals. Hum Genet. 2019;138(4):389-409. | outcomes not relevant |
| Yorifuji T, Kawai M, Muroi J, Mamada M, Kurokawa K, Shigematsu Y, et al. Unexpectedly high prevalence of the mild form of propionic acidemia in Japan: presence of a common mutation and possible clinical implications. Hum Genet. 2002;111(2):161-5. | >10% not newborns and not reported separately |
| Yue L, Lin M, Chen JT, Zhan XF, Zhong DS, Monte-Nguba SM, et al. Rapid screening for sickle cell disease by polymerase chain reaction-high resolution melting analysis. Mol Med Report. 2014;9(6):2479-84. | >10% not newborns and not reported separately |

| | |
|---|---|
| Zhao JB, Zhao L, Gu YC, Huisman TH. Types of alpha-globin gene deficiencies in Chinese newborn babies in the Guangxi region, P. R. China. Hemoglobin. 1992;16(4):325-8. | not direct sequencing |
| Zhao P, Lin L, Lan L. Analysis of mutation spectrum of common deafness-causing genes in Hakka newborns in southern China by semiconductor sequencing. Medicine (Baltimore). 2018;97(38):e12285. | outcomes not relevant |
| Costa TEJ, Gerber VKQ, Ibañez HC, Melanda VS, Parise IZS, Watanabe FM, et al. Penetrance of the <i>TP53</i> R337H Mutation and Pediatric Adrenocortical Carcinoma Incidence Associated with Environmental Influences in a 12-Year Observational Cohort in Southern Brazil. Cancers (Basel). 2019;11(11):16. | sequencing not first line |
| Custodio G, Parise GA, Kiesel Filho N, Komechen H, Sabbaga CC, Rosati R, et al. Impact of neonatal screening and surveillance for the TP53 R337H mutation on early detection of childhood adrenocortical tumors. J Clin Oncol. 2013;31(20):2619-26. | sequencing not first line |
| Bliznetz EA, Martsul DN, Khorov OG, Markova TG, Polyakov AV. Spectrum of the GJB2 mutations in Belarussian patients with hearing loss. Findings of pilot genetic screening of hearing impairment in newborns. Russian Journal of Genetics. 2014;50(2):191-7. | >10% population at risk/symptoms/disease |
| Boonsimma P, Ittiwut C, Kamolvisit W, Ittiwut R, Chetruengchai W, Phokaew C, et al. Exome sequencing as first-tier genetic testing in infantile-onset pharmacoresistant epilepsy: diagnostic yield and treatment impact. Eur J Hum Genet. 2023;31(2):179-87. | >10% population at risk/symptoms/disease |
| Ceyhan-Birsoy O, Machini K, Lebo MS, Yu TW, Agrawal PB, Parad RB, et al. A curated gene list for reporting results of newborn genomic sequencing. Genet Med. 2017;19(7):809-18. | outcomes not relevant |
| Chen G, Wang X, Fu S. Prevalence of A1555G mitochondrial mutation in Chinese newborns and the correlation with neonatal hearing screening. Int J Pediatr Otorhinolaryngol. 2011;75(4):532-4. | sequencing not first line |
| Dai P, Huang LH, Wang GJ, Gao X, Qu CY, Chen XW, et al. Concurrent Hearing and Genetic Screening of 180,469 Neonates with Follow-up in Beijing, China. Am J Hum Genet. 2019;105(4):803-12. | not direct sequencing |
| Alfadhel M, Umair M, Almuzzaini B, Alsaif S, AlMohaimeed SA, Almashary MA, et al. Targeted SLC19A3 gene sequencing of 3000 | outcomes not relevant |

| | |
|---|---|
| Saudi newborn: a pilot study toward newborn screening. Ann. 2019;6(10):2097-103. | |
| Ausems MG, Verbiest J, Hermans MP, Kroos MA, Beemer FA, Wokke JH, et al. Frequency of glycogen storage disease type II in The Netherlands: implications for diagnosis and genetic counselling. Eur J Hum Genet. 1999;7(6):713-6. | not direct sequencing |
| Borges E, Tchonhi C, Couto CSB, Gomes V, Amorim A, Prata MJ, et al. Unusual beta-Globin Haplotype Distribution in Newborns from Bengo, Angola. Hemoglobin. 2019;43(3):149-54. | outcomes not relevant |
| Chen Y, Cao Y, Li HB, Mao J, Liu MJ, Liu YH, et al. SNaPshot reveals high mutation and carrier frequencies of 15 common hearing loss mutants in a Chinese newborn cohort. Clin Genet. 2015;87(5):467-72. | not direct sequencing |
| Chen YJ, Wambach JA, DePass K, Wegner DJ, Chen SK, Zhang QY, et al. Population-based frequency of surfactant dysfunction mutations in a native Chinese cohort. World J Pediatr. 2016;12(2):190-5. | outcomes not relevant |
| Chien YH, Lee NC, Chiang SC, Desnick RJ, Hwu WL. Fabry disease: incidence of the common later-onset alpha-galactosidase A IVS4+919G->A mutation in Taiwanese newborns--superiority of DNA-based to enzyme-based newborn screening for common mutations. Mol Med. 2012;18:780-4. | condition not childhood onset/not monogenic |
| Collins SA, Sinclair G, McIntosh S, Bamforth F, Thompson R, Sobol I, et al. Carnitine palmitoyltransferase 1A (CPT1A) P479L prevalence in live newborns in Yukon, Northwest Territories, and Nunavut. Mol Genet Metab. 2010;101(2-3):200-4. | not direct sequencing |
| Oliveira CA, Pimpinati CJ, Alexandrino F, Magna LA, Maciel-Guerra AT, Sartorato EL. Allelic frequencies of the 35delG mutation of the GJB2 gene in different Brazilian regions. Genet Test. 2007;11(1):1-3. | not direct sequencing |
| Kim J, Field A, Schultz KAP, Hill DA, Stewart DR. The prevalence of DICER1 pathogenic variation in population databases. International Journal of Cancer. 2017 Aug 21;141(10):2030–6. | outcomes not relevant |
| Han SH, Park HJ, Kang EJ, Ryu JS, Lee A, Yang YH, et al. Carrier frequency of GJB2 (connexin-26) mutations causing inherited deafness in the Korean population. Journal of Human Genetics. 2008 Dec;53(11-12):1022–8. | outcomes not relevant |
| Boussaroque A, Audrézet MP ., Raynal C, Sermet-Gaudelus I, Bienvenu T, Férec C, et al. Penetrance is a critical parameter for | outcomes not relevant |

| |
|---|
| assessing the disease liability of CFTR variants. Journal of Cystic Fibrosis. 2020 Apr. |

### Review of cost-effectiveness evaluations of WGS and WES

| Reference | Reason for exclusion |
|---|---|
| Agirrezabal I, Buchanan-Hughes AM, Eddowes LA, Luheshi LM, Sagoo GS, Torok ME. A system dynamics model for the cost-effectiveness evaluation of bacterial whole-genome sequencing for detecting and monitoring outbreaks of MRSA. Value in Health. 2015;18(3):A43-A4. | Conference abstract |
| Alam K, Schofield D. Economic evaluation of genomic sequencing in the paediatric population: a critical review. European journal of human genetics : EJHG. 2018;26(9):1241-7 | Review - checked references (one to add maybe) |
| Alleweldt F, Kara S, Best K, Aarestrup FM, Beer M, Bestebroer TM, et al. Economic evaluation of whole genome sequencing for pathogen identification and surveillance - results of case studies in Europe and the Americas 2016 to 2019. Euro surveillance : bulletin Europeen sur les maladies transmissibles = European communicable disease bulletin. 2021;26(9) | Pathogen surveillance |
| Anonymous. Whole-Exome Sequencing and Chromosomal Microarray Analysis Feasible and Cost-Effective in an Underserved Population: Whole-exome sequencing and chromosomal microarray analysis are more expensive up front but are far more effective in making a genetic diagnosis and much less costly. American journal of medical genetics Part A. 2018;176(5):1044-5 | Not a study (review piece) |
| Arriola E, Bernabe R, Campelo RG, Biscuola M, Enguita AB, Lopez-Rios F, et al. Cost-Effectiveness of Next-Generation Sequencing Versus Single-Gene Testing for the Molecular Diagnosis of Patients With Metastatic Non-Small-Cell Lung Cancer From the Perspective of Spanish Reference Centers. JCO precision oncology. 2023;7:e2200546. | Not WGS/WES |
| Ballesta-Martinez MJ, Perez-Fernandez V, Lopez-Gonzalez V, Sanchez-Soler MJ, Serrano-Anton AT, Rodriguez-Pena LI, et al. Validation of clinical exome sequencing in the diagnostic procedure of patients with intellectual disability in clinical practice. Orphanet journal of rare diseases. 2023;18(1):201 | Not WGS/WES |
| Banerjee S. Cost-Effectiveness and the Economics of Genomic Testing and Molecularly Matched Therapies. Surgical Oncology Clinics of North America. 2024 | Not WGS/WES |
| Bayle A, Droin N, Besse B, Zou Z, Boursin Y, Rissel S, et al. Whole exome sequencing: Evolution of indications and cost to inform public health policy decision-making. Annals of Oncology. 2020;31(Supplement 4):S963 | Conference abstract |
| Bayle A, Marino P, Baffert S, Margier J, Bonastre J. Cost of high-throughput sequencing (NGS) technologies: Literature review and insights. Bulletin du Cancer. 2023 | Non-English |
| Beale S, Sanderson D, Sanniti A, Dundar Y, Boland A. A scoping study to explore the cost-effectiveness of next-generation sequencing compared with traditional genetic testing for the diagnosis of learning disabilities in children. Health technology assessment (Winchester, England). 2015;19(46):1-90. | Review - checked references (none to add) |

| | |
|---|---|
| Bouttell J, Hawkins N, Heggie R, Oien K, Romaniuk A, VanSteenhouse H, et al. Economic evaluation of genomic/genetic tests: a review and future directions. International Journal of Technology Assessment in Health Care. 2022;38(1):e67. | Review - checked references (none to add) |
| Brown B, Allard M, Bazaco MC, Blankenship J, Minor T. An economic evaluation of the Whole Genome Sequencing source tracking program in the U.S. PloS one. 2021;16(10):e0258262. | Pathogen surveillance |
| Buchanan-Hughes AM, Agirrezabal I, Eddowes LA, Luheshi LM, Torok ME, Sagoo GS. A system dynamics model for the cost-effectiveness evaluation of bacterial whole-genome sequencing for monitoring outbreaks of clostridium difficile. Value in Health. 2015;18(3):A44. | Pathogen surveillance |
| Butow P, Davies G, Napier CE, Bartley N, Ballinger ML, Biesecker B, et al. Value of whole-genome sequencing to Australian cancer patients and their first-degree relatives participating in a genomic sequencing study. Journal of genetic counseling. 2022;31(1):96-108 | No costing |
| Cates L, Codreanu A, Ciobanu N, Fosburgh H, Allender CJ, Centner H, et al. Budget impact of next-generation sequencing for diagnosis of TB drug resistance in Moldova. The international journal of tuberculosis and lung disease : the official journal of the International Union against Tuberculosis and Lung Disease. 2022;26(10):963-9. | Pathogen surveillance |
| Chen W, Wong NCB, Wang Y, Zemlyanska Y, Butani D, Teerawattananon Y. Mapping the Value for Money of Precision Medicine: A Systematic Literature Review and Meta-Analysis. Value in Health. 2022;25(12 Supplement):S68. | Review - checked references (none to add) |
| Christensen KD, Phillips KA, Green RC, Dukhovny D. Cost Analyses of Genomic Sequencing: Lessons Learned from the MedSeq Project. Value in health : the journal of the International Society for Pharmacoeconomics and Outcomes Research. 2018;21(9):1054-61. | Not a study (review piece) |
| Christensen KD, Phillips KA, Green RC, Dukhovny D. The cost of integrating whole genome sequencing into the care of cardiomyopathy patients: Sensitivity and Scenario analyses. Value in Health. 2018;21(Supplement 1):S56-S7. | Conference abstract |
| Christensen KD, Vassy JL, Phillips KA, Blout CL, Azzariti DR, Lu CY, et al. Short-term costs of whole genome sequencing in cardiology and primary care: Findings from the medseq project. Value in Health. 2017;20(5):A30. | Conference abstract |
| Christofyllakis K, Bittenbring JT, Thurner L, Ahlgrimm M, Stilgenbauer S, Bewarder M, et al. Cost-effectiveness of precision cancer medicine-current challenges in the use of next generation sequencing for comprehensive tumour genomic profiling and the role of clinical utility frameworks (Review). Molecular and Clinical Oncology. 2022;16(1):21. | Not a study (review piece) |
| Cirillo DM, Cabibbe AM, De Filippo MR, Trovato A, Simonetti T, Rossolini GM, et al. Use of WGS in Mycobacterium tuberculosis routine diagnosis. International journal of mycobacteriology. 2016;5 Suppl 1:S252-S3. | Pathogen surveillance |

| | |
|---|---|
| Cirillo L, Becherucci F, Bellelli S, Mazzinghi B, Raglianti V, Lugli G, et al. COST-ANALYSIS OF A CLINICAL WORKFLOW FOR DIAGNOSIS OF INHERITED KIDNEY DISEASES. Nephrology Dialysis Transplantation. 2022;37(SUPPL 3):i774. | Conference abstract |
| Cirillo L, Francesca B, Mazzierli T, Lomi J, Superchi E, Raglianti V, et al. EARLY IDENTIFICATION OF PODOCYTHOPATHIES AND COLLAGENOPATHIES REDUCE UNNECESSARY MEDICATION FOR PATIENTS AND ECONOMIC BURDEN ON HEALTHCARE SYSTEM. Nephrology Dialysis Transplantation. 2023;38(Supplement 1):i213. | Conference abstract |
| Colman RE, Mace A, Seifert M, Hetzel J, Mshaiel H, Suresh A, et al. Whole-genome and targeted sequencing of drug-resistant Mycobacterium tuberculosis on the iSeq100 and MiSeq: A performance, ease-of-use, and cost evaluation. PLoS medicine. 2019;16(4):e1002794. | Pathogen surveillance |
| Crawford S, Gong C, Randolph LM, Yieh L, Hay JW. PIH27 DIAGNOSING NEWBORNS WITH SUSPECTED SEVERE MITOCHONDRIAL DISORDERS: A COST-EFFECTIVENESS STUDY COMPARING EARLY WHOLE EXOME SEQUENCING TO STANDARD OF CARE. Value in Health. 2020;23(Supplement 1):S156. | Conference abstract |
| Cuckle H, Benn P, Pergament E. Clinical utility and cost of non-invasive prenatal testing. The journal of maternal-fetal & neonatal medicine : the official journal of the European Association of Perinatal Medicine, the Federation of Asia and Oceania Perinatal Societies, the International Society of Perinatal Obstetricians. 2014;27(3):320-1. | Not a study (review piece) |
| de Alava E, Pareja MJ, Carcedo D, Arrabal N, Garcia J-F, Bernabe-Caro R. Cost-effectiveness analysis of molecular diagnosis by next-generation sequencing versus sequential single testing in metastatic non-small cell lung cancer patients from a south Spanish hospital perspective. Expert review of pharmacoeconomics & outcomes research. 2022;22(6):1033-42 | Not WGS/WES |
| Degeling K, Grazziotin Lago L, Hayeems RZ, Boycott KM, Bernier FP, Mendoza-Londono R, et al. Cost-Effectiveness of Alternative Diagnostic Testing Pathways with Whole Exome Sequencing (WES) in a Rare Disease Patient Population: The Canadian Care-for-Rare SOLVE (SOLVE) Multi-Centre Observational Cohort. Value in Health. 2022;25(7 Supplement):S344 | Conference abstract |
| Degeling K, Hayeems RZ, Tagimacruz T, MacDonald KV, Seeger TA, Hartley T, et al. EE171 Time to Diagnosis and Cost Effectiveness of Whole Exome Sequencing (WES) Position in the Diagnostic Pathways of Patients with Suspected Rare Genetic Disease. Value in Health. 2023;26(6 Supplement):S90 | Conference abstract |
| Demougeot L, Houdayer F, Pelissier A, Mohrez F, Thevenon J, Duffourd Y, et al. Changes in clinical practice related to the arrival of next-generation sequencing in the genetic diagnosis of developmental diseases. Archives de Pediatrie. 2018;25(2):77-83 | Non-English |
| Desai K, Hooker G, Gilbert K, Cropper C, Metcalf R, Kachroo S. Real-world trends in costs of next generation sequencing (NGS) testing in U.S. setting. Journal of Clinical Oncology. 2021;39(15 SUPPL) | Conference abstract |

| | |
|---|---|
| Diaby K, Babcock A, Huang Y, Moussa RK, Gupta A, Espinal P, et al. EE144 Real-World Economic Evaluation of Prospective Rapid Whole-Genome Sequencing Compared to a Matched Retrospective Cohort of Critically Ill Pediatric Patients in the United States. Value in Health. 2022;25(7 Supplement):S362 | Conference abstract |
| Ding Z. A cost-effectiveness analysis of a whole genome sequencing test compared to the standard care in patients with hospital-acquired bacteremia | Pathogen surveillance |
| Doble B, Lorgelly P. Clinical players and healthcare payers: aligning perspectives on the cost-effectiveness of next-generation sequencing in oncology. Personalized medicine. 2015;12(1):9-12 | Not a study (review piece) |
| Doble B, Schofield D, Evans C-A, Groza T, Mattick JS, Field M, et al. Impacts of genomics on the health and social costs of intellectual disability. Journal of medical genetics. 2020;57(7):479-86 | Not WGS/WES |
| Douglas MP, Deverka PA, Gelb B, Ferket B, Lich KH, Smith HS, et al. Report - Cost and Clinical Utility of WES and WGS in pediatric patients with suspected genetic disease. medRxiv. 2022.<br>20. Douglas MP, Ladabaum U, Pletcher MJ, Marshall DA, Phillips KA. Economic evidence on identifying clinically actionable findings with whole-genome sequencing: a scoping review. Genetics in medicine : official journal of the American College of Medical Genetics. 2016;18(2):111-6 | Review - checked references (none to add) |
| Douglas MP, Ladabaum U, Pletcher MJ, Marshall DA, Phillips KA. Economic evidence on identifying clinically actionable findings with whole-genome sequencing: a scoping review. Genetics in medicine : official journal of the American College of Medical Genetics. 2016;18(2):111-6 | Review - checked references (none to add) |
| Downie L, Amor D, Halliday J, Lewis S, Martyn M, Goranitis I. Exome sequencing for infants presenting with isolated congenital hearing loss: A cost-effectiveness analysis. Twin Research and Human Genetics. 2021;24(1):74 | Conference abstract |
| Dymond A, Davies H, Mealing S, Pollit V, Coll F, Brown NM, et al. Genomic Surveillance of Methicillin-resistant Staphylococcus aureus: A Mathematical Early Modeling Study of Cost-effectiveness. Clinical infectious diseases : an official publication of the Infectious Diseases Society of America. 2020;70(8):1613-9 | Surveillance |
| Elliott TM, Hare N, Hajkowicz K, Hurst T, Doidge M, Harris PN, et al. Evaluating the economic effects of genomic sequencing of pathogens to prioritise hospital patients competing for isolation beds. Australian health review : a publication of the Australian Hospital Association. 2021;45(1):59-65 | Surveillance |
| Elliott TM, Harris PN, Roberts LW, Doidge M, Hurst T, Hajkowicz K, et al. Cost-effectiveness analysis of whole-genome sequencing during an outbreak of carbapenem-resistant Acinetobacter baumannii. Antimicrobial stewardship & healthcare epidemiology : ASHE. 2021;1(1):e62 | Surveillance |
| Ewans LJ, Schofield D, Shrestha R, Zhu Y, Gayevskiy V, Ying K, et al. Genomic sequencing reanalysis at 12 months boosts mendelian diagnosis and is cost-effectivein intellectual disability. Twin Research and Human Genetics. 2017;20(5):439 | Conference abstract |

| | |
|---|---|
| Fahr P, Buchanan J, Wordsworth S. A Review of Health Economic Studies Comparing Traditional and Massively Parallel Sequencing Diagnostic Pathways for Suspected Genetic Disorders. PharmacoEconomics. 2020;38(2):143-58 | Review - checked references (one to add) |
| Fallah MS, Daneshpour MS, Zeinali S, Bagherian H, Masjoudi S, Zoroufchin Tamiz S. Application of whole-exome sequencing in daily practice: Reducing the cost, diagnostic odyssey, increasing the diagnostic rate. European Journal of Human Genetics. 2019;27(Supplement 2):1818 | Conference abstract |
| Ferket B, Baldwin Z, Murali P, Pai A, Russell H, Lynch F, et al. eP516: Cost-effectiveness frameworks for comparing genome and exome sequencing versus conventional diagnostic pathways. Genetics in Medicine. 2022;24(3 Supplement):S329-S30 | Conference abstract |
| Ferket BS, Baldwin Z, Murali P, Pai A, Mittendorf KF, Russell HV, et al. Cost-effectiveness frameworks for comparing genome and exome sequencing versus conventional diagnostic pathways: A scoping review and recommended methods. Genetics in medicine : official journal of the American College of Medical Genetics. 2022;24(10):2014-27 | Review - checked references (none to add) |
| Ferket BS, Veenstra DL. Economic value of exome sequencing for suspected monogenic disorders. Genetics in medicine : official journal of the American College of Medical Genetics. 2020;22(11):1909 | Not a study (review piece) |
| Ford L, Glass K, Williamson DA, Sintchenko V, Robson JMB, Lancsar E, et al. Cost of whole genome sequencing for non-typhoidal Salmonella enterica. PloS one. 2021;16(3):e0248561 | Surveillance |
| Fox JM, Saunders NJ, Jerwood SH. Economic and health impact modelling of a whole genome sequencing-led intervention strategy for bacterial healthcare-associated infections for England and for the USA. Microbial genomics. 2023;9(8) | Surveillance |
| Gibbs SN, Dalglish H, Campos C, Thakkar S, Palomares M, Yermilov I, et al. Clinical, Humanistic, and Economic Outcomes of Next-Generation Sequencing Tests in Cancer Management of Patients with Advanced Cancer in the United States: Systematic Literature Review. Value in Health. 2022;25(7 Supplement):S536 | Conference abstract |
| Gordon LG, Elliott TM, Forde B, Mitchell B, Russo PL, Paterson DL, et al. Budget impact analysis of routinely using whole-genomic sequencing of six multidrug-resistant bacterial pathogens in Queensland, Australia. BMJ open. 2021;11(2):e041968 | Surveillance |
| Graves JA, Garbett S, Zhou Z, Peterson J. The Value of Pharmacogenomic Information. 2017 | Not WGS/WES |
| Grosse SD, Gudgeon JM. Correspondence on "Cost-effectiveness of exome and genome sequencing for children with rare and undiagnosed conditions" by Lavelle et al. Genetics in medicine : official journal of the American College of Medical Genetics. 2022;24(12):2595-6 | Not a study (review piece) |
| Hart MR, Biesecker BB, Blout CL, Christensen KD, Amendola LM, Bergstrom KL, et al. Secondary findings from clinical genomic sequencing: prevalence, patient perspectives, family history assessment, and health-care costs from a multisite study. Genetics | Not a study (correction) |

| | |
|---|---|
| in medicine : official journal of the American College of Medical Genetics. 2019;21(5):1100-10 | |
| Hart MR, Biesecker BB, Blout CL, Christensen KD, Amendola LM, Bergstrom KL, et al. Correction: Secondary findings from clinical genomic sequencing: prevalence, patient perspectives, family history assessment, and health-care costs from a multisite study. Genetics in medicine : official journal of the American College of Medical Genetics. 2019;21(5):1261-2 | Not WGS/WES |
| Hart MR, Spencer SJ. Consideration for Employer-Based and Geographic Attributes Included in Value Assessment Methods of Next-Generation Sequencing Tests. Journal of managed care & specialty pharmacy. 2019;25(8):936-40 | Review - checked references (none to add) |
| He G, Li Y, Chen X, Chen J, Zhang W. Prediction of treatment outcomes for multidrug-resistant tuberculosis by whole-genome sequencing. International journal of infectious diseases : IJID : official publication of the International Society for Infectious Diseases. 2020;96:68-72 | Pathogen surveillance |
| Howell KB, Eggers S, Dalziel K, Riseley J, Mandelstam S, Carvill GL, et al. Aetiologies and cost-effectiveness of genomic testing in severe epilepsies of infancy: A population-based study. Epilepsia. 2017;58(Supplement 5):S18-S9 | Conference abstract |
| Hwang DY, Hung CC, Chen HC. A cost-effective genetic diagnosis of autosomal dominant polycystic kidney disease and correlation with phenotype. Hong Kong Journal of Nephrology. 2015;17(2 SUPPL. 1):S63 | Conference abstract |
| Jayasinghe K, Wu E, Kerr P, Mallett A, Stark Z, Martyn M, et al. Early genomic sequencing increases diagnostic yield and is cost effective in children. Nephrology. 2020;25(SUPPL 3):31 | Conference abstract |
| Johnson SR, Carter HE, Leo P, Hollingworth SA, Davis EA, Jones TW, et al. Cost-effectiveness Analysis of Routine Screening Using Massively Parallel Sequencing for Maturity-Onset Diabetes of the Young in a Pediatric Diabetes Cohort: Reduced Health System Costs and Improved Patient Quality of Life. Diabetes care. 2019;42(1):69-76 | Not WGS/WES |
| Kang D-YD, Terry SF. Whole Genome Sequencing Will Reduce the Cost of Diagnostic Odyssey. Genetic testing and molecular biomarkers. 2022;26(11):501-2 | Not a study (review piece) |
| Keizer RO, Marouane A, Deden AC, Van Zelst-Stams W, De Boode W, Keusters W, et al. Medical costs of children admitted to the neonatal intensive care unit: the role and possible economic impact of rapid exome sequencing in early diagnosis. European Journal of Human Genetics. 2023;31(Supplement 1):580-1 | Conference abstract |
| Kim H-S, Jeon S, Kim C, Kim YK, Cho YS, Kim J, et al. Chromosome-scale assembly comparison of the Korean Reference Genome KOREF from PromethION and PacBio with Hi-C mapping information. GigaScience. 2019;8(12) | Not costing |
| Kosaki R, Kubota M, Uehara T, Suzuki H, Takenouchi T, Kosaki K. Consecutive medical exome analysis at a tertiary center: Diagnostic and health-economic outcomes. American journal of medical genetics Part A. 2020;182(7):1601-7 | Not WGS/WES |

| | |
|---|---|
| Kossi DS, Touzani R, Borget I, Zhaomin Z, Vaur D, Rouleau E, et al. Cost analysis of human genome sequencing in cancer diagnosis in France: Next generation sequencing. Value in Health. 2016;19(7):A693 | Conference abstract |
| Kumar P, Sundermann AJ, Martin EM, Snyder GM, Harrison LH, Roberts MS. COST-EFFECTIVENESS AND BUDGET IMPACT ANALYSIS OF BACTERIAL WHOLE GENOME SEQUENCING SURVEILLANCE COMPARED TO STANDARD OF CARE IN DETECTING HOSPITAL OUTBREAKS. Med Decis Mak. 2020;40(1):E73-E4 | Conference abstract |
| Kumar P, Sundermann AJ, Martin EM, Snyder GM, Marsh JW, Harrison LH, et al. Method for Economic Evaluation of Bacterial Whole Genome Sequencing Surveillance Compared to Standard of Care in Detecting Hospital Outbreaks. Clinical infectious diseases : an official publication of the Infectious Diseases Society of America. 2021;73(1):e9-e18 | Surveillance |
| Kumar S, Bennett A, Campbell PA, Palidwor G, Lo B, Perkins TJ, et al. Costs of Next-Generation Sequencing Assays in Non-Small Cell Lung Cancer: A Micro-Costing Study. Current oncology (Toronto, Ont). 2022;29(8):5238-46 | Not WGS/WES |
| Lee XJ, Elliott TM, Harris PNA, Douglas J, Henderson B, Watson C, et al. Clinical and Economic Outcomes of Genome Sequencing Availability on Containing a Hospital Outbreak of Resistant Escherichia coli in Australia. Value in health : the journal of the International Society for Pharmacoeconomics and Outcomes Research. 2020;23(8):994-1002 | Surveillance |
| Lemmon CA, Zhou J, Hobbs B, Pennell NA. Modeling Costs and Life-Years Gained by Population-Wide Next-Generation Sequencing or Single-Gene Testing in Nonsquamous Non-Small-Cell Lung Cancer in the United States. JCO precision oncology. 2023;7:e2200294 | Not WGS/WES |
| Levenson D. Benefits of genomic sequencing evident in pediatric diagnoses: recent study finds testing method less costly, more effective than other medical, genetic tests. American journal of medical genetics Part A. 2015;167A(3):vii-viii | Not a study (review piece) |
| Li C, Vandersluis S, Holubowich C, Ungar WJ, Goh ES, Boycott KM, et al. Correspondence on "cost or price of sequencing? implications for economic evaluations in genomic medicine" by Grosse and Gudgeon. Genetics in Medicine. 2022;24(1):251-2 | Not a study (review piece) |
| Loong H, Wong CKH, Leung LKS, Chan CPK, Chang A, Zhou ZY, et al. Economic impact of next-generation sequencing (NGS) versus single-gene testing modalities to detect genomic alterations (GAs) in metastatic non-small cell lung cancer (mNSCLC) in Asia. Annals of Oncology. 2020;31(Supplement 6):S1394-S | Conference abstract |
| Lu CY. Economic evaluation of whole-genome sequencing in healthy individuals: what can we learn from CEAs of whole-body CT screening? Genetics in medicine : official journal of the American College of Medical Genetics. 2016;18(1):103-4 | Not a study (review piece) |
| Lundie B, Patel C, Jeppesen M, Head L, Brion K, McGaughran J, et al. Whole Genome Sequencing Partnership Program: A Risk-Sharing Agreement to Implement WGS as a First Line Test in | Conference abstract |

| | |
|---|---|
| Pediatric Monogenic Disease. Twin Research and Human Genetics. 2023;26(2):62 | |
| Marshall DA, Degeling K, Tagimacruz T, Seeger TA, Boycott KM, Bernier F, et al. Costs And Effectiveness Of Whole Exome Sequencing (WES) In Patients With Unsolved Rare Disease Through The Diagnostic Pathway. International Journal of Technology Assessment in Health Care. 2023;39(Supplement 1):S50 | Conference abstract |
| Mathews C, Niphadkar U, Huffstater T, Andukuri A, Im O, Sheppard E, et al. EE739 Assessing the Value of Next Generation Sequencing in NSCLC. Value in Health. 2023;26(12 Supplement):S197 | Conference abstract |
| Matsuda H, Ogawa T, Sadatsuki Y, Tsujino T, Wada S, Kim S-W, et al. Budget impact analysis of next-generation sequencing versus sequential single-gene testing in Japanese patients with advanced non-small-cell lung cancer. Respiratory investigation. 2023;61(1):61-73. | Not WGS/WES |
| Middelburg P, Monroe G, Van Gassen K, Hovels A, Knoers N, Vrijenhoek T, et al. Impact of whole exome sequencing (WES) on costs and medical decision-making. Value in Health. 2016;19(7):A705-A6 | Conference abstract |
| Milo Rasouly H, Wynn J, Marasa M, Reingold R, Chatterjee D, Kapoor S, et al. Evaluation of the cost and effectiveness of diverse recruitment methods for a genetic screening study. Genetics in medicine : official journal of the American College of Medical Genetics. 2019;21(10):2371-80 | Not WGS/WES |
| Mittal AK, Shekhawat DS, Joshi V, Singh P, Singh K. Next-generation Sequencing in Newborn Screening: A Review on Clinical and Economic Prospects. Open Biotechnology Journal. 2023;17(1):e187407072212120 | Not a study (review piece) |
| Montaut S, Tranchant C, Drouot N, Rudolf G, Guissart C, Tarabeux J, et al. Assessment of a Targeted Gene Panel for Identification of Genes Associated With Movement Disorders. JAMA neurology. 2018;75(10):1234-45 | Not WGS/WES |
| Mugwagwa T, Abubakar I, White PJ. Using molecular testing and whole-genome sequencing for tuberculosis diagnosis in a low-burden setting: a cost-effectiveness analysis using transmission-dynamic modelling. Thorax. 2021;76(3):281-91 | Pathogen surveillance |
| Nahas S, Chowdhury S, Dimmock D. Utilization of Rapid Whole Genomic Sequencing (rWGS) Demonstrates Significant Improvement in Clinical Utility and Cost Effectiveness in Neonatal and Pediatric Hospital Intensive Care Units. Journal of Molecular Diagnostics. 2019;21(6):1119- | Conference Abstract |
| Nahas S, Chowdhury S, Dimmock D, Kingsmore S. Rapid Whole Genome Sequencing Improves Clinical Utility and Cost Effectiveness of Acutely Ill Children admitted to Neonatal Intensive Care Units. European Journal of Human Genetics. 2019;27:801 | Conference Abstract |
| Norris S, Belcher A, Howard K, Ward RL. Evaluating genetic and genomic tests for heritable conditions in Australia: lessons learnt from health technology assessments. Journal of community genetics. 2022;13(5):503-22 | Not costing |
| Nurchis MC, Riccardi MT, Damiani G. Health technology assessment of whole genome sequencing in the diagnosis of | Review - checked references (none to add) |

| | |
|---|---|
| genetic disorders: a scoping review of the literature. International journal of technology assessment in health care. 2022;38(1):e71 | |
| Nurchis MC, Riccardi MT, Radio FC, Chillemi G, Bertini ES, Tartaglia M, et al. Incremental net benefit of whole genome sequencing for newborns and children with suspected genetic disorders: Systematic review and meta-analysis of cost-effectiveness evidence. Health policy (Amsterdam, Netherlands). 2022;126(4):337-45 | Review - checked references (none to add) |
| Oberg JA, Sireci AN, Mansukhani MM, Nagy PL, Glade Bender JL, Kung AL. Clinical Implementation of Genomic Sequencing in Pediatric Oncology: Identification and Valuation of Resources and Costs Associated with Next-Generation Sequencing. Value in health : the journal of the International Society for Pharmacoeconomics and Outcomes Research. 2014;17(7):A645 | Conference Abstract |
| Olde Keizer RACM, Deden AC, Van Zelst-Stams WAG, De Boode WP, Henneman L, Ploos Van Amstel JK, et al. Medical costs of children admitted to the Neonatal Intensive Care Unit; The role and possible value of rapid Whole Exome Sequencing. European Journal of Human Genetics. 2019;27(Supplement 2):1652 | Conference Abstract |
| Olde Keizer RACM, Henneman L, Ploos van Amstel JK, Vissers LELM, Frederix GWJ. Economic evaluations of exome and genome sequencing in pediatric genetics: considerations towards a consensus strategy. Journal of medical economics | Review - checked references (some maybe to add) |
| Panca M, Blackstone J, Stirrup O, Cutino-Moguel MT, Thomson E, Peters C, et al. Evaluating the cost implications of integrating SARS-CoV-2 genome sequencing for infection prevention and control investigation of nosocomial transmission within hospitals. The Journal of hospital infection. 2023;139:23-32 | Surveillance |
| Park SY, Faraci G, Ward PM, Emerson JF, Lee HY. High-precision and cost-efficient sequencing for real-time COVID-19 surveillance. Scientific reports. 2021;11(1):13669. | Surveillance |
| Payne K, Gavan SP, Wright SJ, Thompson AJ. Cost-effectiveness analyses of genetic and genomic diagnostic tests. Nature reviews Genetics. 2018;19(4):235-46. | Not a study (review piece) |
| Pecoraro V, Negro A, Sculco C, Di Brino E, Cicchetti A, Pedrini E. The accuracy and clinical validity of Next Generation Sequencing in the genetic testing of two orthopaedic rare diseases. Biochimica Clinica. 2020;44(SUPPL 2):S38-S9. | Conference abstract |
| Phillips KA, Pletcher MJ, Ladabaum U. Is the ``$1000 Genome'' really $1000? Understanding the full benefits and costs of genomic sequencing. Technology and health care : official journal of the European Society for Engineering and Medicine. 2015;23(3):373-9. | Not costing |
| Planas-Ballve A, Caballol N, Cardona X, Gomez Ruiz I, Balague Marmana M, A'Vila A. Challenges in Diagnosis of Hereditary Ataxia and Spastic Paraplegias. Movement Disorders. 2022;37(Supplement 2):S227. | Conference abstract |
| Pollie R. Genomic Sequencing Costs Set to Head Down Again. Engineering. 2023;23:3-6. | Not a study (review piece) |
| Price V, Ngwira LG, Lewis JM, Baker KS, Peacock SJ, Jauneikaite E, et al. A systematic review of economic evaluations of whole- | Surveillance |

| | |
|---|---|
| genome sequencing for the surveillance of bacterial pathogens. Microbial genomics. 2023;9(2). | |
| Proudman D, DeVito NC, Belinson S, Allo MA, Morris ED, Signorovitch J, et al. Comprehensive genomic profiling in advanced/metastatic colorectal cancer: number needed to test and budget impact of expanded first line use. Journal of Medical Economics. 2022;25(1):817-25. | Not WGS/WES |
| Pruneri G, De Braud F, Sapino A, Aglietta M, Vecchione A, Giusti R, et al. Next-Generation Sequencing in Clinical Practice: Is It a Cost-Saving Alternative to a Single-Gene Testing Approach? PharmacoEconomics - Open. 2021;5(2):285-98. | Not WGS/WES |
| Putra M, Kesavan M, Hackney D. 1106 Cost-Effectiveness analysis of genetic testing for stillbirth: balancing cost and diagnostic rate. American Journal of Obstetrics and Gynecology. 2021;224(2 Supplement):S682. | Conference abstract |
| Radio F, Bartuli A, Novelli A, Tartaglia M, Dallapiccola B. Cost-effectiveness of whole exome sequencing to solve the unsolved: an Italian pilot study. European Journal of Human Genetics. 2019;27:597-. | Conference abstract |
| Rajan V, Terry SF, Green J, Ortega J. Diagnostic Yield and Cost-Benefit When Utilizing Clinical Whole Genome Sequencing. Genetic testing and molecular biomarkers. 2022;26(5):253-4. | Not a study (review piece) |
| Retel V, Mewes J, Wind A, Rocha-Goncalves F, Van Harten W. A general model for cancer centers to conduct health technology assessments; A practical case of next generation sequencing in oncology. European Journal of Cancer. 2017;72(Supplement 1):S118. | Conference abstract |
| Rezapour A, Souresrafil A, Barzegar M, Sheikhy-Chaman M, Tatarpour P. Economic evaluation of next-generation sequencing techniques in diagnosis of genetic disorders: A systematic review. Clinical genetics. 2023;103(5):513-28. | Review - checked references (one to add) |
| Roscioli T, Cliffe C, Elakis G, Zhu Y, Nixon C, Mullan G, et al. The first 500 diagnostic exomes: A demonstration of safety, clinical utility, translation and cost-effectiveness. Twin Research and Human Genetics. 2019;21(5):418-9. | Conference abstract |
| Rosso A, Pitini E, D'Andrea E, Massimi A, De Vito C, Marzuillo C, et al. The Cost-effectiveness of Genetic Screening for Familial Hypercholesterolemia: a Systematic Review. Annali di igiene : medicina preventiva e di comunita. 2017;29(5):464-80. | Not WGS/WES |
| Ruzhnikov MR, Alsadah A, Mendelsohn B, Alhariri A, Cilio MR, Wu Y, et al. Diagnostic outcomes and relative cost of clinical whole exome sequencing. Journal of Investigative Medicine. 2016;64(1):247. | Conference abstract |
| Rynehart L, Schofield D, Shrestha R, Stark Z, White S. Long-term economic impacts of exome sequencing for suspected monogenic disorders. European Journal of Human Genetics. 2019;27(Supplement 2):1812. | Conference abstract |
| Sachdev R, Palmer EE, Schofield D, Shrestha R, Kandula T, Macintosh R, et al. Cost-effectiveness of the diagnostic whole exome sequencing approach in epileptic encephalopathy. Twin Research and Human Genetics. 2016;19(5):544. | Conference abstract |

| | |
|---|---|
| Saini V, Jaber T, Como JD, Abdulmassih R, Min Z, Bhanot N. Clinical and Financial Impact of Next Generation Sequencing (NGS) in addition to Conventional Microbiology Testing in our Urban Referral Health Center. Open Forum Infectious Diseases. 2021;8(SUPPL 1):S434-S5. | Conference abstract |
| Santos Gonzalez F, Mordaunt D, Stark Z, Dalziel K, Christodoulou J, Goranitis I. Microcosting diagnostic genomic sequencing: A systematic review. Genetics in medicine : official journal of the American College of Medical Genetics. 2023;25(6):100829. | Review - checked references (one to add) |
| Sayson B, Popurs MAM, Lafek M, Berkow R, Stockler-Ipsiroglu S, van Karnebeek CDM. Retrospective analysis supports algorithm as efficient diagnostic approach to treatable intellectual developmental disabilities. Molecular Genetics and Metabolism. 2015;115(1):1-9. | Not WGS/WES |
| Schofield D, Alam K, Shrestha R, Rynehart L, Stark Z, White S, et al. Cost-effectiveness of whole exome sequencing for suspected monogenic disorders in in fancy: A follow-up study. Twin Research and Human Genetics. 2017;20(5):445. | Conference abstract |
| Schofield D, Field M, Boyle J, Christie L, Murray L, West S, et al. Evidence of the cost-effectiveness of WGS in familial intellectual disability. Twin Research and Human Genetics. 2017;20(5):478. | Conference abstract |
| Schofield D, Rynehart L, Shrestha R, Stark Z, White S, Gaff C. Estimating the education cost impacts of whole exome sequencing for infants with suspected monogenic disorders. Twin Research and Human Genetics. 2017;20(5):479. | Conference abstract |
| Schofield D, Rynehart L, Shresther R, White S, Stark Z. Long-term economic impacts of whole exome sequencing for suspected monogenic disorders. Twin Research and Human Genetics. 2019;21(5):404. | Conference abstract |
| Schofield D, Tan O, Shrestha R, West S, Boyle J, Christie L, et al. Loosening the purse strings: Influencing public funders about the value of genomic testing for intellectual disability. European Journal of Human Genetics. 2022;30(SUPPL 1):70. | Conference abstract |
| Shrestha R, Schofield D, Ewans L, Field M, Dinger M, Buckley M, et al. What's value for money: Whole-exome sequencing or whole-genome sequencing? comparison from an economic perspective. Twin Research and Human Genetics. 2019;21(5):475-6. | Conference abstract |
| Silas U, Bluher M, Smith AB, Saunders R. Fast In-House Next-Generation Sequencing in the Diagnosis of Metastatic Non-small Cell Lung Cancer: A Hospital Budget Impact Analysis. Journal of Health Economics and Outcomes Research. 2023;10(1):111-8. | Conference Abstract |
| Silas U, M B, Dumanois R, Saunders R. In-House Versus Send-out Next Generation Sequencing Testing for Metastatic Non-Small Cell Lung Cancer Patients. a Budget Impact Analysis. Value in Health. 2023;26(6 Supplement):S299. | Not WGS/WES |
| Simons M, Van De Ven M, Coupe V, Joore M, Ijzerman M, Koffijberg E, et al. Early technology assessment of using whole genome sequencing in personalized oncology. Expert review of pharmacoeconomics & outcomes research. 2021;21(3):343-51. | Not a study (review piece) |
| Smith HS, Swint JM, Lalani SR, Yamal J-M, de Oliveira Otto MC, Castellanos S, et al. Clinical Application of Genome and Exome | Review - checked references (none to add) |

| | |
|---|---|
| Sequencing as a Diagnostic Tool for Pediatric Patients: a Scoping Review of the Literature. Genetics in medicine : official journal of the American College of Medical Genetics. 2019;21(1):3-16. | |
| Soller M, Runheim H, Nordgren A, Pettersson M, Hammarsjo A, Henriksson M, et al. Costs and diagnostic yield of whole genome sequencing in neurodevelopmental disorders. European Journal of Human Genetics. 2023;31(Supplement 1):632. | Conference abstract |
| Spackman E, Hinde S, Bojke L, Payne K, Sculpher M. Using Cost-Effectiveness Analysis to Quantify the Value of Genomic-Based Diagnostic Tests: Recommendations for Practice and Research. Genetic testing and molecular biomarkers. 2017;21(12):705-16. | Review - checked references (none to add) |
| Spackman E, Hinde S, Bojke L, Payne K, Sculpher MJ. Establshing the cost-effectiveness of genomic-based diagnostic tests: Are current methods sufficient and appropriate? Value in Health. 2015;18(7):A727. | Conference abstract |
| Spencer S, Veenstra DL, Guzauskas G. PCV58 COST-EFFECTIVENESS OF POPULATION-WIDE GENOMIC SCREENING FOR FAMILIAL HYPERCHOLESTEROLEMIA. Value in Health. 2019;22(Supplement 2):S129. | Conference abstract |
| Stark Z, Schofield D, Alam K, Wilson W, Mupfeki N, Macciocca I, et al. Cost-effectiveness of singleton whole exome sequencing compared with standard diagnostic care. Twin Research and Human Genetics. 2016;19(5):539. | Conference abstract |
| Stark Z, Schofield D, Martyn M, Rynehart L, Shrestha R, Alam K, et al. Correction: Does genomic sequencing early in the diagnostic trajectory make a difference? A follow-up study of clinical outcomes and cost-effectiveness. Genetics in medicine : official journal of the American College of Medical Genetics. 2019;21(2):516. | Not a study (correction) |
| Stenzinger A, Cuffel B, Paracha N, Vail E, Garcia-Foncillas J, Goodman C, et al. Supporting Biomarker-Driven Therapies in Oncology: A Genomic Testing Cost Calculator. The oncologist. 2023;28(5):e242-e53. | Not WGS/WES |
| Suokas A, Yi Y, Hirst A. Cost-Effectiveness of Earlier Whole-Genome Sequencing in Rare Genetic Childhood Epilepsy. Value in Health. 2022;25(12 Supplement):S68. | Conference abstract |
| Swaggart KA, Swarr DT, Tolusso LK, He H, Dawson DB, Suhrie KR. Making a Genetic Diagnosis in a Level IV Neonatal Intensive Care Unit Population: Who, When, How, and at What Cost? The Journal of pediatrics. 2019;213:211-7.e4. | Not WGS/WES |
| Tan O, Shrestha R, Cunich M, Schofield DJ. Application of next-generation sequencing to improve cancer management: A review of the clinical effectiveness and cost-effectiveness. Clinical genetics. 2018;93(3):533-44. | Review - checked references (two to add) |
| Tran M, Smurthwaite KS, Nghiem S, Cribb DM, Zahedi A, Ferdinand AD, et al. Economic evaluations of whole-genome sequencing for pathogen identification in public health surveillance and health-care-associated infections: a systematic review. The Lancet Microbe. 2023;4(11):e953-e62. | Surveillance |
| Tutera S, Olson M, Williams C, Leyland-Jones B, Huber M, McKean H, et al. Cost analysis of genomic-directed therapy in patients with | Conference abstract |

| | |
|---|---|
| metastatic lung and colorectal cancers. Cancer Research. 2015;75(15 SUPPL. 1). | |
| Van Nimwegen K, Vissers L, Willemsen M, Schieving J, Veltman J, Van Der Wilt G, et al. The cost-effectiveness of whole-exome sequencing in complex paediatric neurology. Value in Health. 2016;19(7):A695. | Not WGS/WES |
| Van Nimwegen K, Van Soest R, Grutters J, Veltman J, Vissers L, Van Der Wilt G. A next-generation framework: Deciding on the role of costs in the clinical use of targeted gene panels, exome and genome sequencing. Value in Health. 2015;18(7):A352. | Conference abstract |
| Van Nimwegen KJ. Feasibility of the headroom analysis in early economic evaluation of innovative diagnostic technologies with no immediate treatment implications. Value in Health. 2014;17(7):A550. | Conference abstract |
| Varesio C, Gana S, Asaro A, Ballante E, Tartara E, Pasca L, et al. Diagnostic yield and cost-effectiveness of "dynamic exome-based approach" in epilepsy with neurodevelopmental disorders: A clinical experience. Epilepsia. 2021;62(SUPPL 3):207-8. | Conference abstract |
| Vogel M, Utpatel C, Corbett C, Kohl TA, Iskakova A, Ahmedov S, et al. Implementation of whole genome sequencing for tuberculosis diagnostics in a low-middle income, high MDR-TB burden country. Scientific reports. 2021;11(1):15333. | Surveillance |
| Wallbillich JJ, Forde B, Havrilesky LJ, Cohn DE. A personalized paradigm in the treatment of platinum-resistant ovarian cancer - A cost utility analysis of genomic-based versus cytotoxic therapy. Gynecologic oncology. 2016;142(1):144-9. | Not WGS/WES |
| Weymann D, Dragojlovic N, Pollard S, Regier DA. Allocating healthcare resources to genomic testing in Canada: latest evidence and current challenges. Journal of community genetics. 2022;13(5):467-76. | Review - checked references (none to add) |
| Wilsdon T, Mirza M, Goerke L. Evaluating the Cost-Effectiveness of Next-Generation Sequencing as a Biomarker Testing Strategy in Oncology and Implications for Policy: A Literature Review. Value in Health. 2023;26(12 Supplement):S62. | Conference abstract |
| Wong BE, Carmona JJ, Fortunato-habib MM, Van Aggelen HC, Doty AJ, Gross BD. A data-driven model of the economic burden of healthcare-associated infec-tions as impacted by use of comprehensive genomic analysis of bacteria. Open Forum Infectious Diseases. 2019;6(Supplement 2):S847. | Surveillance |
| Yang S-C, Yeh Y-C, Chen Y-L, Chiu C-H. Economic Analysis of Exclusionary EGFR Test Versus Up-Front NGS for Lung Adenocarcinoma in High EGFR Mutation Prevalence Areas. Journal of the National Comprehensive Cancer Network : JNCCN. 2022;20(7):774-82.e4. | Not WGS/WES |
| Yenamandra VK. Establishing a cost-effective strategy for diagnosis of inherited epidermolysis bullosa in resource-limited settings. Journal of Investigative Dermatology. 2017;137(10 Supplement 2):S228. | Conference abstract |
| Yeung A, Tan N, Tan T, Stark Z, Brown N, Delatycki M, et al. Evaluating cost-effectiveness of exome sequencing in a prospective | Conference abstract |

| | |
|---|---|
| versus historical cohort of complex pediatric patients. Twin Research and Human Genetics. 2019;22(5):338. | |
| Yeung A, Tan NB, Tan TY, Stark Z, Brown N, Hunter MJ, et al. A cost-effectiveness and utility analysis of genomic sequencing in a prospective versus historical cohort of complex pediatric patients. European Journal of Human Genetics. 2020;28(SUPPL 1):54. | Conference abstract |
| Yu TM, Morrison C, Gold EJ, Tradonsky A, Arnold RJG. Budget Impact of Next-Generation Sequencing for Molecular Assessment of Advanced Non-Small Cell Lung Cancer. Value in health : the journal of the International Society for Pharmacoeconomics and Outcomes Research. 2018;21(11):1278-85. | Not WGS/WES |
| Zeppel M, Schofield D, Shrestha R, Staffieri S, Jamieson R, Jelovic D, et al. Cost-effectiveness in retinoblastoma in relation to reproductive technologies and preimplantation genetic diagnosis (PGD). Twin Research and Human Genetics. 2019;21(5):457. | Conference abstract |
| Zheng Y, Vioix H, Liu FX, Singh B, Sharma S, Sharda D. Diagnostic and economic value of biomarker testing for targetable mutations in non-small-cell lung cancer: a literature review. Future oncology (London, England). 2022;18(4):505-18. | Not WGS/WES |
| Zischke J, White N, Gordon L. Accounting for Intergenerational Cascade Testing in Economic Evaluations of Clinical Genomics: A Scoping Review. Value in health : the journal of the International Society for Pharmacoeconomics and Outcomes Research. 2022;25(6):944-53. | Review |
| Zou D, Ye W, Hess LM, Bhandari NR, Ale-Ali A, Foster J, et al. Diagnostic Value and Cost-Effectiveness of Next-Generation Sequencing-Based Testing for Treatment of Patients with Advanced/Metastatic Non-Squamous Non-Small-Cell Lung Cancer in the United States. The Journal of molecular diagnostics : JMD. 2022;24(8):901-14. | Not WGS/WES |
| Richards J, Korgenski EK, Taft RJ, Vanderver A, Bonkowsky JL. Targeted leukodystrophy diagnosis based on charges and yields for testing. Am J Med Genet A. 2015;167a(11):2541-3. | Not WGS/WES |
| Dillon OJ, Lunke S, Stark Z, Yeung A, Thorne N, Gaff C, et al. Exome sequencing has higher diagnostic yield compared to simulated disease-specific panels in children with suspected monogenic disorders. European Journal of Human Genetics. 2018;26(5):644-51. | Not costing |
| Demos M, Guella I, DeGuzman C, McKenzie MB, Buerki SE, Evans DM, et al. Diagnostic Yield and Treatment Impact of Targeted Exome Sequencing in Early-Onset Epilepsy. Frontiers in neurology. 2019;10:434. | Not WGS/WES |
| Yokoi T, Enomoto Y, Tsurusaki Y, Harada N, Saito T, Nagai J-I, et al. An efficient genetic test flow for multiple congenital anomalies and intellectual disability. Pediatrics international : official journal of the Japan Pediatric Society. 2020;62(5):556-61 | Not WGS/WES |
| Mody RJ, Wu YM, Lonigro RJ, Cao X, Roychowdhury S, Vats P, et al. Integrative Clinical Sequencing in the Management of Refractory or Relapsed Cancer in Youth. Jama. 2015;314(9):913-25. | Not WGS/WES |
| Oberg JA, Glade Bender JL, Sulis ML, Pendrick D, Sireci AN, Hsiao SJ, et al. Implementation of next generation sequencing into | Not costing |

| | |
|---|---|
| pediatric hematology-oncology practice: moving beyond actionable alterations. Genome Med. 2016;8(1):133. | |
| Catchpool M, Ramchand J, Martyn M, Hare DL, James PA, Trainer AH, et al. A cost-effectiveness model of genetic testing and periodical clinical screening for the evaluation of families with dilated cardiomyopathy. Genet Med. 2019;21(12):2815-22. | Not WGS/WES |
| Bennette CS, Gallego CJ, Burke W, Jarvik GP, Veenstra DL. The cost-effectiveness of returning incidental findings from next-generation genomic sequencing. Genetics in medicine : official journal of the American College of Medical Genetics. 2015;17(7):587-95. | Not WGS/WES |
| Van Nimwegen K, Vissers L, Willemsen M, Schieving J, Veltman J, Van Der Wilt G, et al. The cost-effectiveness of whole-exome sequencing in complex paediatric neurology. Value in Health. 2016;19(7):A695. | Conference abstract |
| Pankhurst LJ, Del Ojo Elias C, Votintseva AA, Walker TM, Cole K, Davies J, et al. Rapid, comprehensive, and affordable mycobacterial diagnosis with whole-genome sequencing: a prospective study. The Lancet Respiratory medicine. 2016;4(1):49-58. | Surveillance |
| Buchanan-Hughes AM, Griffiths A, Evans J, Slater D, Eddowes LA. Investigating the cost-effectiveness of bacterial whole-genome sequencing for enabling targeted antibiotic selection in urinary tract infections. Value in Health. 2015;18(7):A510. | Conference abstract |
| Bonnefond A, Philippe J, Durand E, Muller J, Saeed S, Arslan M, et al. Highly sensitive diagnosis of 43 monogenic forms of diabetes or obesity through one-step PCR-based enrichment in combination with next-generation sequencing. Diabetes care. 2014;37(2):460-7. | Not economic evaluation (outcome study) |
| Ghaoui R, Cooper ST, Lek M, Jones K, Corbett A, Reddel SW, et al. Use of Whole-Exome Sequencing for Diagnosis of Limb-Girdle Muscular Dystrophy: Outcomes and Lessons Learned. JAMA Neurology. 2015;72(12):1424-32. | Not economic evaluation (just mentions cost of WES) |
| Lee E-J, Dykas D, Bale A, Cromwell C, Parker TL, Halene S, et al. Whole Exome Sequencing in Evaluation of Thrombophilia: A Novel 33-Gene Panel. Blood. 2015;126(23):3529. | Not economic evaluation (outcome study) |
| McDonell LM, Warman Chardon J, Schwartzentruber J, Foster D, Beaulieu CL, Majewski J, et al. The utility of exome sequencing for genetic diagnosis in a familial microcephaly epilepsy syndrome. BMC Neurol. 2014;14:22. | Not economic evaluation (outcome study) |
| Neveling K, Feenstra I, Gilissen C, Hoefsloot LH, Kamsteeg E-J, Mensenkamp AR, et al. A Post-Hoc Comparison of the Utility of Sanger Sequencing and Exome Sequencing for the Diagnosis of Heterogeneous Diseases. Human Mutation. 2013;34(12):1721-6. | Not an economic evaluation (outcomes) |
| Sawyer SL, Schwartzentruber J, Beaulieu CL, Dyment D, Smith A, Warman Chardon J, et al. Exome sequencing as a diagnostic tool for pediatric-onset ataxia. Hum Mutat. 2014;35(1):45-9. | Not an economic evaluation (outcomes) |
| Schieving JH. PP05.5 – 3064: The role of exome sequencing in daily pediatric neurology practice. European Journal of Paediatric Neurology. 2015;19:S47. | |
| Chrystoja CC, Diamandis EP. Whole Genome Sequencing as a Diagnostic Test: Challenges and Opportunities. Clinical Chemistry. 2014;60(5):724-33. | Schofield D, Alam K, Shrestha R, Rynehart L, Stark Z, White S, et al. Cost-effectiveness of |

| | |
|---|---|
| | whole exome sequencing for suspected monogenic disorders in in fancy: A follow-up study. Twin Research and Human Genetics. 2017;20(5):445. |
| Dewey FE, Grove ME, Pan C, Goldstein BA, Bernstein JA, Chaib H, et al. Clinical Interpretation and Implications of Whole-Genome Sequencing. JAMA. 2014;311(10):1035-45. | Not an economic evaluation (outcomes) |
| Wright C, Burton H, Hall A, Moorthie S, Pokorska-Bocci‡ DA, Coordinator P, et al. Next steps in the sequence The implications of whole genome sequencing for health in the UK. Cambridge: PHG Foundation; 2011. | Review |
| Towne MC, Beggs AH, Agrawal PB, editors. Efficiency of whole exome/genome sequencing for achieving a diagnosis in rare presentations. Annual meeting of the American Society of Human Genetics 22–26 October 2013; 2013; Boston, MA, . | Conference Abstract |
| Young C, Argáez C. CADTH Rapid Response Reports. Rapid Genome-wide Testing: A Review of Clinical Utility, Cost-Effectiveness, and Guidelines. Ottawa (ON): Canadian Agency for Drugs and Technologies in Health<br>Copyright © 2019 Canadian Agency for Drugs and Technologies in Health.; 2019. | Review |
| Hanquet G, Vinck I, Thiry N. THE USE OF WHOLE GENOME SEQUENCING IN CLINICAL PRACTICE: CHALLENGES AND ORGANISATIONAL CONSIDERATIONS FOR BELGIUM Belgium: Belgian Health Care Knowledge Centre (KCE); 2018. Contract No.: KCE Reports 300 | Review |
