## Supplement 3 for "Evaluating whole genome sequencing for rare diseases in newborn screening: evidence synthesis from a series of systematic reviews"

### Included studies for Q2 (prevalence of genetic variants in those with biochemical or biochemical and clinical features of each condition)

Overview and characteristics of included studies for Q2 (prevalence of genetic variants in those with biochemical or biochemical and clinical features of each condition) of the review of five conditions including study aim, population, total population number in study, eligible population number for the review, study outcome and disease category of included population.

### PDE

| Reference | Study aim | Population | Total population N in study | Eligible population for review | Study outcome | Disease category of included population |
|---|---|---|---|---|---|---|
| Akiyama T, Hyodo Y, Hasegawa K, Oboshi T, Imai K, Ishihara N, et al. Pyridoxal in the Cerebrospinal Fluid May Be a Better Indicator of Vitamin B6-dependent Epilepsy Than Pyridoxal 5'-Phosphate. Pediatric neurology. 2020;113:33-41. | To demonstrate the biochemical characteristics of vitamin B6-dependent epilepsy, with a particular focus on pyridoxal 5'-phosphate and pyridoxal in the cerebrospinal fluid | Patients with PDE or PLPHP deficiency | 9 | 7 patients with PDE | Variant frequency in ALDH7A1gene | Gene (ALDH7A1) positive PDE |
| Baumgart A, Spiczak Sv, Verhoeven-Duif NM, Moller RS, Boor R, Muhle H, et al. Atypical vitamin B6 deficiency: a rare cause of unexplained neonatal and infantile epilepsies. Journal of child neurology. 2014;29(5):704-7. | To assess the frequency of (ALDH7A1 and PNPO deficiencies) in unexplained infantile epilepsy | Patients with unexplained seizure disorders starting during the neonatal or infantile period | 113 | 113 | Variant frequency in ALDH7A1 gene | Gene (ALDH7A1) positive PDE |

| | | | | | | |
|---|---|---|---|---|---|---|
| Bennett CL, Chen Y, Hahn S, Glass IA, Gospe SM, Jr. Prevalence of ALDH7A1 mutations in 18 North American pyridoxine-dependent seizure (PDS) patients. Epilepsia. 2009;50(5):1167-75. | To evaluate the hypothesis that the prevalence of ALDH7A1 mutations is discordant between early (neonatal) and later-onset cases of PDS | Patients with pyridoxine-dependent seizure | 18 | 18 | Variant frequency in ALDH7A1 gene | Clinical-biochemical PDE |
| Boonsimma P, Ittiwut C, Kamolvisit W, Ittiwut R, Chetruengchai W, Phokaew C, et al. Exome sequencing as first-tier genetic testing in infantile-onset pharmacoresistant epilepsy: diagnostic yield and treatment impact. European journal of human genetics : EJHG. 2023;31(2):179-87. | To identify yield and treatment implications of exome sequencing (ES) as first-tier genetic testing for infantile-onset pharmacoresistant epilepsy | Children with infantile-onset (age <= 12 months) pharmacoresistant epilepsy | 103 | 103 | Variant frequency in ALDH7A1 gene | Childhood onset pharmaco-resistant seizures |
| Costain G, Cordeiro D, Matviychuk D, Mercimek-Andrews S. Clinical Application of | To identify underlying genetic diagnoses | Children seen in a single Paediatric Epilepsy Genetics Clinic | 197 | 197 | Variant frequency in ALDH7A1 gene | Childhood onset pharmaco-resistant seizures |

| | | | | | | |
|---|---|---|---|---|---|---|
| Targeted Next-Generation Sequencing Panels and Whole Exome Sequencing in Childhood Epilepsy. Neuroscience. 2019;418:291-310. | | | | | | |
| Coughlin CR, II, van Karnebeek CDM, Al-Hertani W, Shuen AY, Jaggumantri S, Jack RM, et al. Triple therapy with pyridoxine, arginine supplementation and dietary lysine restriction in pyridoxine-dependent epilepsy: Neurodevelopmental outcome. Mol Genet Metab. 2015;116(1-2):35-43. | To report the developmental and biochemical outcome of six subjects who were treated with this triple therapy | Patients who were genetically positive for PDE | 6 | 6 | Variant frequency in ALDH7A1 gene | Gene (ALDH7A1) positive PDE |
| Costain G, Cordeiro D, Matviychuk D, Mercimek-Andrews S. Clinical Application of Targeted Next-Generation Sequencing Panels and Whole Exome | To provide a comprehensive overview of ALDH7A1 mutations that cause PDE | Patients with PDE-ALDH7A1 | 185 | 185 | Variant frequency in ALDH7A1 gene | Gene (ALDH7A1) positive PDE |

| | | | | | | |
|---|---|---|---|---|---|---|
| Sequencing in Childhood Epilepsy. Neuroscience. 2019;418:291-310. | | | | | | |
| Gallagher RC, Van Hove JLK, Scharer G, Hyland K, Plecko B, Waters PJ, et al. Folinic acid-responsive seizures are identical to pyridoxine-dependent epilepsy. Annals of neurology. 2009;65(5):550-6. | To perform genetic and biochemical testing and to determine the relation between folinic acid-responsive seizures and pyridoxine-dependent epilepsy | Patients with Folinic acid–responsive seizures and pyridoxine-dependent epilepsy | 9 | 9 | Variant frequency in ALDH7A1 gene | Clinical-biochemical PDE |
| Hu C, Liu D, Luo T, Wang Y, Liu Z. Phenotypic spectrum and long-term outcome of children with genetic early-infantile-onset developmental and epileptic encephalopathy. Epileptic Disorders. 2022;24(2):343-52. | To explore the clinical phenotype and long-term outcome in children with genetic early-infantile-onset DEEs (developmental and epileptic encephalopathy) | Patients with early onset pharmacoresistant seizures and developmental delay | 470 | 470 | Variant frequency in ALDH7A1 gene | Childhood onset pharmaco-resistant seizures |
| Jamali A, Kristensen E, Tangeraas T, Arntsen V, Sikiric A, Kupliauskiene G, et al. Corrigendum to "The spectrum of | To study all available Norwegian subjects with PDE to characterise the phenotypic spectrum and treatment response across the age span to address factors that might | Cases of PDE based on current treatment with pyridoxine | 15 | 15 | Variant frequency in ALDH7A1 gene | Clinical-biochemical PDE |

| | | | | | | |
|---|---|---|---|---|---|---|
| pyridoxine dependent epilepsy across the age span: A nationwide retrospective observational study" [Epilepsy Res. 190 (2023) 107099]. Epilepsy research. 2023;198:107113. | determine the overall prognosis of the disorder. | | | | | |
| Jiao X, Xue J, Gong P, Wu Y, Zhang Y, Jiang Y, et al. Clinical and genetic features in pyridoxine-dependent epilepsy: a Chinese cohort study. Developmental medicine and child neurology. 2020;62(3):315-21. | To characterize the clinical and genetic characteristics of a large cohort of patients with PDE. | Patients with PDE | 33 | 33 | Variant frequency in ALDH7A1 gene | Clinical-biochemical PDE |
| Koul R, Alfutaisi A, Abdelrahim R, Altihilli K. Pyridoxine Responsive Seizures: Beyond Aldehyde Dehydrogenase 7A1. Journal of neurosciences in rural practice. 2019;10(4):613-6. | To report the profile of children with pyridoxine responsive seizures | Children with neonatal and infantile seizures responding to pyridoxine | 35 | 35 | Variant frequency in ALDH7A1 gene | Clinical-biochemical PDE |

| | | | | | | |
|---|---|---|---|---|---|---|
| Mefford HC, Zemel M, Geraghty E, Cook J, Clayton PT, Paul K, et al. Intragenic deletions of ALDH7A1 in pyridoxine-dependent epilepsy caused by Alu-Alu recombination. Neurology. 2015;85(9):756-62. | To investigate the role of intragenic deletions of ALDH7A1 in patients with clinical and biochemical evidence of PDE | Patients with clinical and biochemical evidence of PDE | 6 | 6 | Variant frequency in ALDH7A1 gene | Clinical-biochemical PDE |
| Mercimek-Mahmutoglu S, Patel J, Cordeiro D, Hewson S, Callen D, Donner EJ, et al. Diagnostic yield of genetic testing in epileptic encephalopathy in childhood. Epilepsia. 2015;56(5):707-16. | To determine the genetic diagnostic yield in epileptic encephalopathy, we performed a retrospective cohort study in a single epilepsy genetics clinic | Patients with intractable epilepsy, global developmental delay, and cognitive dysfunction | 110 | 110 | Variant frequency in ALDH7A1 gene | Childhood onset pharmaco-resistant seizures |
| Mills PB, Footitt EJ, Mills KA, Tuschl K, Aylett S, Varadkar S, et al. Genotypic and phenotypic spectrum of pyridoxine-dependent epilepsy (ALDH7A1 deficiency). Brain : a | To evaluate the use of measurement of urine L-alpha-aminoadipic semialdehyde/creatinine ratio and mutation analysis of ALDH7A1 (antiquitin) in investigation of patients with suspected or clinically proven PDE and to characterize further the phenotypic | Children with a seizure disorder; those with elevated alpha-AASA/creatinine ratio had repeat urine analysis or offered sequence analysis of ALDH7A1 | 243 | 37 underwent genetic testing | Variant frequency in ALDH7A1 gene | Gene (ALDH7A1) positive PDE |

| | | | | | | |
|---|---|---|---|---|---|---|
| journal of neurology. 2010;133(Pt 7):2148-59. | spectrum of antiquitin deficiency | | | | | |
| Della Mina E, Ciccone R, Brustia F, Bayindir B, Limongelli I, Vetro A, et al. Improving molecular diagnosis in epilepsy by a dedicated high-throughput sequencing platform. Eur J Hum Genet. 2015;23(3):354-62. | To investigate whether a quick and cheap molecular diagnosis could be provided | Patients suffering from a range of seizures, either familial or sporadic; randomly selected from patients' afferent to epileptic centre for children and adolescents. | 19 | 19 | Variant frequency in ALDH7A1 gene | Childhood onset pharmaco-resistant seizures |
| Perez B, Gutierrez-Solana LG, Verdu A, Merinero B, Yuste-Checa P, Ruiz-Sala P, et al. Clinical, biochemical, and molecular studies in pyridoxine-dependent epilepsy. Antisense therapy as possible new therapeutic option. Epilepsia. 2013;54(2):239-48. | To report the clinical, biochemical, and mutational spectrum of 12 patients with clinically proven PDE. | Patients suspected with PDE (clinically confirmed) | 12 | 12 | Variant frequency in ALDH7A1 gene | Gene (ALDH7A1) positive PDE |
| Plecko B, Paul K, Paschke E, Stoeckler-Ipsiroglu S, Struys E, Jakobs C, et al. | To provide biochemical and molecular characterization of 18 patients with PDE in 16 unrelated families | Patients with neonatal seizure onset | 18 | 18 | Variant frequency in ALDH7A1 gene | Gene (ALDH7A1) positive PDE |

| | | | | | | |
|---|---|---|---|---|---|---|
| Biochemical and molecular characterization of 18 patients with pyridoxine-dependent epilepsy and mutations of the antiquitin (ALDH7A1) gene. Human mutation. 2007;28(1):19-26. | | | | | | |
| Salomons GS, Bok LA, Struys EA, Pope LL, Darmin PS, Mills PB, et al. An intriguing "silent" mutation and a founder effect in antiquitin (ALDH7A1). Annals of neurology. 2007;62(4):414-8. | To report an intriguing “silent” mutation in ALDH7A1, a novel missense mutation and a founder mutation in a Dutch cohort (10 patients) with alpha-AASA dehydrogenase deficiency. | Patients with alpha-AASA deficiency & ALDH7A1 | 10 | 10 | Variant frequency in ALDH7A1 gene | Gene (ALDH7A1) positive PDE |
| Scharer G, Brocker C, Vasiliou V, Creadon-Swindell G, Gallagher RC, Spector E, et al. The genotypic and phenotypic spectrum of pyridoxine-dependent epilepsy due to mutations in | To report three patients with PDE presenting with variable phenotypes and responses to pyridoxine supplementation. | Patients with PDE-ALDH7A1 | 15 | 15 | Variant frequency in ALDH7A1 gene | Gene (ALDH7A1) positive PDE |

| | | | | | | |
|---|---|---|---|---|---|---|
| ALDH7A1. Journal of inherited metabolic disease. 2010;33(5):571-81. | | | | | | |
| Tlili A, Hamida Hentati N, Chaabane R, Gargouri A, Fakhfakh F. Pyridoxine-dependent epilepsy in Tunisia is caused by a founder missense mutation of the ALDH7A1 gene. Gene. 2013;518(2):242-5. | To report the first evidence for a founder mutation responsible for PDE | Unrelated Tunisian families | 25 | 7 patients with PDE | Variant frequency in ALDH7A1 gene | Clinical-biochemical PDE |
| Tincheva S, Todorov T, Todorova A, Georgieva R, Stamatov D, Yordanova I, et al. First cases of pyridoxine-dependent epilepsy in Bulgaria: novel mutation in the ALDH7A1 gene. Neurological sciences : official journal of the Italian Neurological Society and of the Italian Society of Clinical | To report the first cases of molecular genetic diagnostics of PDE in Bulgaria: a dizygotic twin pair and one non-related child with classical PDE caused by one novel and two recurrent mutations in the ALDH7A1 gene | Families referred for genetic testing with because of intractable seizures (all classical clinical PDE) | 7 | 5 patients underwent testing | Variant frequency in ALDH7A1 gene | Clinical-biochemical PDE |

| | | | | | | |
|---|---|---|---|---|---|---|
| Neurophysiology. 2015;36(12):2209-12. | | | | | | |
| Tlili A, Hamida Hentati N, Gargouri A, Fakhfakh F. Identification of a novel missense mutation in the ALDH7A1 gene in two unrelated Tunisian families with pyridoxine-dependent epilepsy. Molecular biology reports. 2013;40(1):487-90. | To report a novel ALDH7A1 homozygous missense mutation identified in two Tunisian families with PDE | Patients from two unrelated Tunisian families with PDE | 10 | 2 patients with PDE | Variant frequency in ALDH7A1 gene | Clinical-biochemical PDE |
| van Karnebeek CDM, Stockler S. Treatable inborn errors of metabolism causing intellectual disability: A systematic literature review. Molecular Genetics and Metabolism. 2012;105(3):368-81. | To test the effectiveness and safety of dietary lysine restriction as an adjunct to pyridoxine therapy on chemical biomarkers, seizure control, and developmental or cognitive outcomes in 7 children with confirmed ATQ deficiency | Children with PDE caused by antiquitin (ATQ) deficiency | 7 | 7 | Variant frequency in ALDH7A1 gene | Gene (ALDH7A1) positive PDE |

### hRB

| Reference | Study aim | Population | Total population N in study | Eligible population for review | Study outcome | Disease category of included population |
|---|---|---|---|---|---|---|
| Abidi O, Knari S, Sefri H, Charif M, Senechal A, Hamel C, et al. Mutational analysis of the RB1 gene in Moroccan patients with retinoblastoma. Molecular vision. 2011;17:3541-7. | To identify the spectrum and the effect of germline mutations | Unrelated cases of RB with different clinical presentations | 41 | 41 | Variant frequency in RB1 gene | Any RB |
| Abouzeid H, Munier FL, Thonney F, Schorderet DF. Ten novel RB1 gene mutations in patients with retinoblastoma. Molecular vision. 2007;13:1740-5. | To study phenotype-genotype correlations in RB patients | Patients with RB | 65 | 65 | Variant frequency in RB1 gene | Any RB |
| Abraham A, Thirumalairaj K, Gaikwad N, Muthukkaruppan V, Reddy AG, Thangaraj K, et al. Retinoblastoma discordance in families with twins. Indian journal of ophthalmology. 2019;67(3):436-9. | To describe twins (including genetic testing) where one is affected by RB and the other is unaffected | One twin in each family affected by bilateral RB, other twin unaffected | 17 | 9<br>Three sets of twins from three families | Variant frequency in RB1 gene | Familial/ germline / bilateral RB |

| | | | | | | |
|---|---|---|---|---|---|---|
| 4. Afshar AR, Pekmezci M, Bloomer MM, Cadenas NJ, Stevers M, Banerjee A, et al. Next-Generation Sequencing of Retinoblastoma Identifies Pathogenic Alterations beyond RB1 Inactivation That Correlate with Aggressive Histopathologic Features. Ophthalmology. 2020;127(6):804-13. | | | | | | |
| Afshar AR, Pekmezci M, Bloomer MM, Cadenas NJ, Stevers M, Banerjee A, et al. Next-Generation Sequencing of Retinoblastoma Identifies Pathogenic Alterations beyond RB1 Inactivation That Correlate with Aggressive Histopathologic Features. Ophthalmology. 2020;127(6):804-13. | To determine the utility of next-generation sequencing for the clinical management of children undergoing enucleation for RB | Patients with RB | 32 | 32 | Variant frequency in RB1 gene | Any RB |

| | | | | | | |
|---|---|---|---|---|---|---|
| Ahani A, Behnam B, Khorshid HRK, Akbari MT. RB1 gene mutations in Iranian patients with retinoblastoma: report of four novel mutations. Cancer genetics. 2011;204(6):316-22. | To perform mutation analysis of Iranian RB patients | Children with RB | 18 | 18 | Variant frequency in RB1 gene | Any RB |
| Ahani A, Akbari MT, Saliminejad K, Behnam B, Akhondi MM, Vosoogh P, et al. Screening for large rearrangements of the RB1 gene in Iranian patients with retinoblastoma using multiplex ligation-dependent probe amplification. Molecular vision. 2013;19:454-62. | To identify deletions/duplications of RB1 gene in a cohort of Iranian patients | Patients with RB who were referred to Mahak, Farabi, and Rasoul Akram hospitals | 121 | 121 | Variant frequency in RB1 gene | Any RB |
| Akdeniz Odemis D, Kebudi R, Bayramova J, Kilic Erciyas S, Kuru Turkcan G, Tuncer SB, et al. RB1 gene mutations and genetic spectrum in retinoblastoma cases. | To investigate the frequency and types of mutations in the RB1 gene in a Turkish population | Patients with RB in Turkey | 136 | 122 probands | Variant frequency in RB1 gene | Any RB |

| | | | | | | |
|---|---|---|---|---|---|---|
| Medicine. 2023;102(36):e35068. | | | | | | |
| Albrecht P, Ansperger-Rescher B, Schuler A, Zeschnigk M, Gallie B, Lohmann DR. Spectrum of gross deletions and insertions in the RB1 gene in patients with retinoblastoma and association with phenotypic expression. Human mutation. 2005;26(5):437-45. | To use different sequencing techniques to identify mutations (gross deletions or insertions mainly) in RB patients where no mutation was previously found | Patients with RB | 129 | 129 | Variant frequency in RB1 gene | Any RB |
| Alekseeva EA, Babenko OV, Kozlova VM, Ushakova TL, Kazubskaya TP, Nemtsova MV, et al. Parental origin of the RB1 gene mutations in families with low penetrance hereditary retinoblastoma. Cancers. 2021;13(20):5068. | To identify RB1 alterations causing hereditary low penetrance RB and to evaluate how the parental origin of an RB1 mutation affects its phenotypic expression | Patients with RB | 332 | 332 | Variant frequency in RB1 gene | Any RB |
| Ali MJ, Parsam VL, Honavar SG, Kannabiran C, Vemuganti GK, Reddy VAP. RB1 gene mutations in | To identify correlations between the type of mutations observed and the severity of the disease using multiple sequencing techniques | Patients with RB | 74 | 74 | Variant frequency in RB1 gene | Any RB |

| | | | | | | |
|---|---|---|---|---|---|---|
| retinoblastoma and its clinical correlation. Saudi Journal of Ophthalmology. 2010;24(4):119-23. | | | | | | |
| Alonso J, Moreno C, Lopez A, Mendiola M, Garcia-Miguel P, Abelairas J, et al. Five novel single nucleotide polymorphisms of the RB1 gene (g.5625T>C, g.70169T>G, g.76875A>T, g.78026delA, and g.150072T>C) in retinoblastoma patients. Human mutation. 2001;17(5):437. | To perform mutation analysis of RB patients | Patients with RB | 43 | 43 | Variant frequency in RB1 gene | Any RB |
| Arbetman A, Abdala M, Fandino A, Herrera J, Baranzini S, Borelina D, et al. Clinical, cytogenetic, and molecular testing of Argentine patients with retinoblastoma. Journal of AAPOS : the official publication of the American Association for Pediatric | To determine clinical, chromosomal, and molecular characteristics of Argentine patients with unilateral and bilateral RB | Patients with RB | 86 | 30 received genetic testing | Variant frequency in RB1 gene | Any RB |

| | | | | | | |
|---|---|---|---|---|---|---|
| Ophthalmology and Strabismus. 1998;2(2):102-7. | | | | | | |
| Ata-ur-Rasheed M, Vemuganti Gk, Honavar Sg, Ahmed N, Hasnain Se, Kannabiran C. Mutational analysis of the RB1 gene in Indian patients with retinoblastoma. Ophthalmic genetics. 2002;23(2):121-8. | To perform mutation analysis of RB patients | Patients with RB | 21 | 21 | Variant frequency in RB1 gene | Any RB |
| Ayari Jeridi H, Bouguila H, Ansperger-Rescher B, Baroudi O, Mdimegh I, Omran I, et al. Genetic testing in Tunisian families with heritable retinoblastoma using a low cost approach permits accurate risk prediction in relatives and reveals incomplete penetrance in adults. Experimental eye research. 2014;124:48-55. | To provide stepwise strategy for mutation analysis of Tunisian RB patients | Patients with RB | 20 | 20 | Variant frequency in RB1 gene | Familial/ germline / bilateral RB |
| Bamne MN, Ghule PN, Jose J, Banavali SD, Kurkure PA, Amare | To identify and characterize constitutional and | Patients with RB | 34 | 34 | Variant frequency in RB1 gene | Sporadic RB |

| | | | | | | |
|---|---|---|---|---|---|---|
| Kadam PS. Constitutional and somatic RB1 mutation spectrum in nonfamilial unilateral and bilateral retinoblastoma in India. Genetic testing. 2005;9(3):200-11. | somatic RB1 gene mutations | | | | | |
| Barbosa RH, Aguiar FCC, Silva MFL, Costa RA, Vargas FR, Lucena E, et al. Screening of RB1 alterations in Brazilian patients with retinoblastoma and relatives with retinoma: phenotypic and genotypic associations. Investigative ophthalmology & visual science. 2013;54(5):3184-94. | To perform mutation analysis of Brazilian RB patients and analysis of genotype-phenotype associations | Patients with RB | 75 | 71 patients with RB | Variant frequency in RB1 gene | Any RB |
| Barbosa RH, Vargas FR, Aguiar FCC, Ferman S, Lucena E, Bonvicino CR, et al. Hereditary retinoblastoma transmitted by maternal germline mosaicism. Pediatric | To report a family with transmission of a germline RB1 mutation | A family with affected and healthy offspring | 10 | 10 | Variant frequency in RB1 gene | Familial/ germline / bilateral RB |

| | | | | | | |
|---|---|---|---|---|---|---|
| blood & cancer. 2008;51(5):598-602. | | | | | | |
| Bisht S, Chawla B, Kumar A, Vijayan V, Kumar M, Sharma P, et al. Identification of novel genes by targeted exome sequencing in Retinoblastoma. Ophthalmic genetics. 2022;43(6):771-88. | To perform targeted exome sequencing approach to find novel mutations in Indian RB patients | Patients with RB | 75 | 75 | Variant frequency in RB1 gene | Sporadic RB |
| Blanquet V, Turleau C, Gross-Morand MS, Senamaud-Beaufort C, Doz F, Besmond C. Spectrum of germline mutations in the RB1 gene: a study of 232 patients with hereditary and non hereditary retinoblastoma. Human molecular genetics. 1995;4(3):383-8. | To perform mutation analysis of all 27 exons of the RB1 | Patients affected with familial/ bilateral sporadic or unilateral multifocal, unilateral sporadic RB | 232 | 232 | Variant frequency in RB1 gene | Any RB |
| Braggio E, Bonvicino CR, Vargas FR, Ferman S, Eisenberg ALA, Seuanez HN. Identification of three novel RB1 mutations in Brazilian patients with | To perform mutation analysis of RB1 gene using different techniques | Patients with sporadic bilateral/unilateral. Patients with familial RB | 28 | 28 | Variant frequency in RB1 gene | Any RB |

| | | | | | | |
|---|---|---|---|---|---|---|
| retinoblastoma by "exon by exon" PCR mediated SSCP analysis. Journal of clinical pathology. 2004;57(6):585-90. | | | | | | |
| Chai P, Luo Y, Yu J, Li Y, Yang J, Zhuang A, et al. Clinical characteristics and germline mutation spectrum of RB1 in Chinese patients with retinoblastoma: A dual-center study of 145 patients. Experimental eye research. 2021;205:108456. | To compare patient demographics and clinical characteristics between patients with or without RB1 mutations | Consecutive patients with RB | 145 | 145 | Variant frequency in RB1 gene | Any RB |
| Dalamon V, Surace E, Borelina D, Ziembar M, Esperante S, Francipane L, et al. Detection of mutations in argentine retinoblastoma patients by segregation of polymorphisms, exon analysis and cytogenetic test. Ophthalmic research. 2001;33(6):336-9. | To detect chromosome and molecular abnormalities in 16 Argentine families with RB | Families with RB | 16 | 16 | Variant frequency in RB1 gene | Sporadic RB |

| | | | | | | |
|---|---|---|---|---|---|---|
| Dalamon V, Surace E, Giliberto F, Ferreiro V, Fernandez C, Szijan I. Detection of germline mutations in argentine retinoblastoma patients: low and full penetrance retinoblastoma caused by the same germline truncating mutation. Journal of biochemistry and molecular biology. 2004;37(2):246-53. | To perform mutation analysis in Argentinian RB patients | Patients with RB in Argentina | 21 | 21 | Variant frequency in RB1 gene | Any RB |
| Davies HR, Broad KD, Onadim Z, Price EA, Zou X, Sheriff I, et al. Whole-genome sequencing of retinoblastoma reveals the diversity of rearrangements disrupting RB1 and uncovers a treatment-related mutational signature. Cancers. 2021;13(4):1-19. | To identify new mutations in RB1 gene using tumour samples from 20 patients with sporadic RB | Individuals with no previous family history | 20 | 20 | Variant frequency in RB1 gene | Sporadic RB |
| Devarajan B, Prakash L, Kannan TR, Abraham AA, Kim U, Muthukkaruppan V, et al. Targeted next | To use targeted next generation sequencing with in-house bioinformatics pipeline for the molecular | Patients with RB | 33 | 33 | Variant frequency in RB1 gene | Any RB |

| | | | | | | |
|---|---|---|---|---|---|---|
| generation sequencing of RB1 gene for the molecular diagnosis of Retinoblastoma. BMC cancer. 2015;15:320. | diagnosis of RB in families | | | | | |
| Dommering CJ, Mol BM, Moll AC, Burton M, Cloos J, Dorsman JC, et al. RB1 mutation spectrum in a comprehensive nationwide cohort of retinoblastoma patients. Journal of medical genetics. 2014;51(6):366-74. | To describe RB1 mutations in a Dutch cohort | Patients registered in the Dutch Retinoblastoma Registry | 1173 | 529 underwent genetic testing | Variant frequency in RB1 gene | Any RB |
| Frenkel S, Pe'er J. The clinical presentation of retinoblastoma patients with mosaics. Investigative Ophthalmology and Visual Science. 2016;57(12):3672. | To describe the association between RB1 mutations and disease characteristics in RB patients | Patients with RB | 295 | 149 received genetic testing | Variant frequency in RB1 gene | Any RB |
| Gargallo P, Oltra JS, Yanez Y, Segura V, Balaguer J, Canete A. Retinoblastoma: towards an earlier diagnosis. Retinoblastoma: hacia un diagnostico mas | To evaluate the different diagnostic aspects and clinical features of RB patients, and propose strategies that might improve their clinical management | Patients with RB | 38 | 38 | Variant frequency in RB1 gene | Any RB |

| | | | | | | |
|---|---|---|---|---|---|---|
| precoz. 2018;93(9):439-43. | | | | | | |
| Genuardi M, Klutz M, Devriendt K, Caruso D, Stirpe M, Lohmann DR. Multiple lipomas linked to an RB1 gene mutation in a large pedigree with low penetrance retinoblastoma. European journal of human genetics : EJHG. 2001;9(9):690-4. | To describe large pedigree in which lipoma predisposition is linked to a mutant RB1 allele that causes low-penetrance RB | Family with individuals affected by bilateral RB | 28 | 28 | Variant frequency in RB1 gene | Familial/ germline / bilateral RB |
| Grotta S, D'Elia G, Scavelli R, Genovese S, Surace C, Sirleto P, et al. Advantages of a next generation sequencing targeted approach for the molecular diagnosis of retinoblastoma. BMC cancer. 2015;15:841. | To report experience on a cohort of RB patients using a combined approach of NGS and RB1 custom array-Comparative Genomic Hybridization | Patients affected by RB in Rome | 65 | 65 | Variant frequency in RB1 gene | Any RB |
| Gupta H, Malaichamy S, Mallipatna A, Murugan S, Jeyabalan N, Suresh Babu V, et al. Retinoblastoma genetics screening and clinical management. BMC medical | To describe and identify associations between genetic and clinical parameters of 50 RB patients from India | Patients with RB | 50 | 50 | Variant frequency in RB1 gene | Any RB |

| | | | | | | |
|---|---|---|---|---|---|---|
| genomics. 2021;14(1):188. | | | | | | |
| He M-y, An Y, Gao Y-j, Qian X-w, Li G, Qian J. Screening of RB1 gene mutations in Chinese patients with retinoblastoma and preliminary exploration of genotype-phenotype correlations. Molecular vision. 2014;20:545-52. | To screen constitutional mutations in the RB1 gene via a method combining DNA sequencing and multiplex ligation-dependent probe amplification, and a preliminary exploration of genotype–phenotype correlations | Children diagnosed with RB | 85 | 85 | Variant frequency in RB1 gene | Sporadic RB |
| Hulsenbeck I, Frank M, Biewald E, Kanber D, Lohmann DR, Ketteler P. Introduction of a variant classification system for analysis of genotype-phenotype relationships in heritable retinoblastoma. Cancers. 2021;13(7):1605. | To study the retinoblastoma variant effect classification which considers each variant's predicted effects on the common causal mediator | Patients with germline mutation who did not undergo screening | 287 | 287 | Variant frequency in RB1 gene | Familial/ germline / bilateral RB |
| Hung C-C, Lin S-Y, Lee C-N, Chen C-P, Lin S-P, Chao M-C, et al. Low penetrance of retinoblastoma for p.V654L mutation of the RB1 gene. BMC | To perform clinical assessments and molecular analyses in a large Taiwanese family with RB | Taiwanese family with RB | 30 | 30 | Variant frequency in RB1 gene | Familial/ germline / bilateral RB |

| | | | | | | |
|---|---|---|---|---|---|---|
| medical genetics. 2011;12:76. | | | | | | |
| Jakubowska A, Zajaczek S, Haus O, Limon J, Kostyk E, Krzystolik Z, et al. Novel RB1 gene constitutional mutations found in Polish patients with familial and/or bilateral retinoblastoma. Human mutation. 2001;18(5):459. | To report three novel constitutional RB1 gene mutations in patients with retinoblastoma. | Patients with RB | 8 | 8 | Variant frequency in RB1 gene | Familial/ germline / bilateral RB |
| Kalsoom S, Wasim M, Afzal S, Shahzad MS, Ramzan S, Awan AR, et al. Alterations in the RB1 gene in Pakistani patients with retinoblastoma using direct sequencing analysis. Molecular vision. 2015;21:1085-92. | To identify mutational alterations in the RB1 gene in Pakistani patients with RB | Clinically evaluated patients with RB from different regions of Pakistan | 70 | 70 | Variant frequency in RB1 gene | Any RB |
| Kiet NC, Khuong LT, Minh DD, Quan NHM, Xinh PT, Trang NNC, et al. Spectrum of mutations in the RB1 gene in Vietnamese patients with | To present the spectrum of mutations in the RB1 gene in Vietnamese patients with RB. | Probands with advanced stage (group D or E) RB | 50 | 41 underwent genetic testing | Variant frequency in RB1 gene | Any RB |

| | | | | | | |
|---|---|---|---|---|---|---|
| retinoblastoma. Molecular vision. 2019;25:215-21. | | | | | | |
| Kiran VS, Kannabiran C, Chakravarthi K, Vemuganti GK, Honavar SG. Mutational screening of the RB1 gene in Indian patients with retinoblastoma reveals eight novel and several recurrent mutations. Human mutation. 2003;22(4):339. | To perform mutational screen of the exons and promoter of the RB1 gene in Indian patients with retinoblastoma to determine the range of mutations giving rise to disease | Probands with RB | 47 | 20 with peripheral blood only | Variant frequency in RB1 gene | Any RB |
| Lan X, Xu W, Tang X, Ye H, Song X, Lin L, et al. Spectrum of RB1 Germline Mutations and Clinical Features in Unrelated Chinese Patients With Retinoblastoma. Frontiers in Genetics. 2020;11:142. | To identify germline RB1 mutations and correlate the identified mutations with the clinical features of Rb patients. | Unrelated Rb patients and their parents | 118 | 118 | Variant frequency in RB1 gene | Any RB |
| Li L, Li H, Zhang J, Gan H, Liu R, Hu X, et al. Five novel RB1 gene mutations and genotype-phenotype correlations in Chinese | To identify the spectrum of RB1 gene mutations in 114 Chinese patients with retinoblastoma. | Patients with retinoblastoma from Southern China | 114 | 114 | Variant frequency in RB1 gene | Any RB |

| | | | | | | |
|---|---|---|---|---|---|---|
| children with retinoblastoma. International ophthalmology. 2022;42(11):3421-30. | | | | | | |
| Linh DNH, Van Huy N, Nguyen PD, Le Thi P, Tuan HA, Van Nguyen T, et al. Mutation spectrum of retinoblastoma patients in Vietnam. Molecular genetics & genomic medicine. 2023;11(11):e2244. | To report mutations (both germline and somatic) found in (RB) patients … (in stage B–E) … , which would then aid the genetic counselling and detection of RB patients at earlier stages, reducing the mortality rate | Patients with RB, diagnosed by standard ophthalmologic and histological criteria in the 2020–2022 period, at the National Institute of Ophthalmology | 42 | 42 | Variant frequency in RB1 gene | Any RB |
| Lohmann D, Horsthemke B, Gillessen-Kaesbach G, Stefani FH, Hofler H. Detection of small RB1 gene deletions in retinoblastoma by multiplex PCR and high-resolution gel electrophoresis. Human genetics. 1992;89(1):49-53. | To screen all the 27 exons and adjacent intronic sequences. To reduce the labor involved, the different regions of RB1 gene were simultaneously amplified n a single polymerase chain reaction (multiplex PCR). | Unrelated patients with hereditary retinoblastoma | 24 | 24 | Variant frequency in RB1 gene | Familial/ germline / bilateral RB |
| Lohmann DR, Brandt B, Hopping W, Passarge E, Horsthemke B. Spectrum of small length germline | To identify germline mutations in the RB1 gene | Patients with hRB | 106 | 106 | Variant frequency in RB1 gene | Familial/ germline / bilateral RB |

| | | | | | | |
|---|---|---|---|---|---|---|
| mutations in the RB1 gene. Human molecular genetics. 1994;3(12):2187-93. | | | | | | |
| Lohmann DR, Brandt B, Hopping W, Passarge E, Horsthemke B. The spectrum of RB1 germ-line mutations in hereditary retinoblastoma. American journal of human genetics. 1996;58(5):940-9. | To delineate the spectrum of RB1 germ-line mutations and to search for genotype-phenotype correlations | Patients with RB | 71 | 71 | Variant frequency in RB1 gene | Familial/ germline / bilateral RB |
| Manukonda R, Pujar A, Ramappa G, Vemuganti GK, Kaliki S. Identification of novel RB1 genetic variants in Retinoblastoma patients and their impact on clinical outcome. Ophthalmic genetics. 2022;43(1):64-72. | To study the clinical outcomes in all RB patients who underwent genetic testing and attempted to understand the association between genetic mutations and clinical outcomes in Asian Indian RB patients | Patients diagnosed with RB and their parents/family members who attended genetic counselling followed by genetic testing were included in the study | 62 | 62 | Variant frequency in RB1 gene | Any RB |
| Mehyar M, Mosallam M, Tbakhi A, Saab A, Sultan I, Deebajah R, et al. Impact of RB1 gene mutation type in retinoblastoma patients on clinical presentation and | To evaluate the impact of the type of RB1 gene mutation on clinical presentation and management outcome | Children with RB | 50 | 50 | Variant frequency in RB1 gene | Any RB |

| | | | | | | |
|---|---|---|---|---|---|---|
| management outcome. Hematol Oncol Stem Cell Ther. 2020;13(3):152-9. | | | | | | |
| Mendonca V, Evangelista AC, P Matta B, M Moreira MA, Faria P, Lucena E, et al. Molecular alterations in retinoblastoma beyond RB1. Experimental eye research. 2021;211:108753. | To identify novel genomic alterations in non-conventional RB (with apparent wild-type RB1) which could contribute to initiation or progression in this malignancy | Children with RB | 96 | 96 | Variant frequency in RB1 gene | Any RB |
| Mohd Khalid MKN, Yakob Y, Md Yasin R, Wee Teik K, Siew CnG, Rahmat J, et al. Spectrum of germ-line RB1 gene mutations in Malaysian patients with retinoblastoma. Molecular vision. 2015;21:1185-90. | To perform molecular genetic testing of the RB1 gene | Children with RB | 19 | 19 | Variant frequency in RB1 gene | Any RB |
| Nguyen HH, Nguyen HTT, Vu NP, Le QT, Pham CM, Huyen TT, et al. Mutational screening of germline RB1 gene in Vietnamese patients with retinoblastoma reveals three novel | To identify germline mutations in RB1 in a cohort of patients with Rb from northern Vietnam. | Children with RB | 34 | 34 | Variant frequency in RB1 gene | Any RB |

| | | | | | | |
|---|---|---|---|---|---|---|
| mutations. Molecular vision. 2018;24:231-8. | | | | | | |
| Nichols KE, Houseknecht MD, Godmilow L, Bunin G, Shields C, Meadows A, et al. Sensitive multistep clinical molecular screening of 180 unrelated individuals with retinoblastoma detects 36 novel mutations in the RB1 gene. Human mutation. 2005;25(6):566-74. | To facilitate genetics counselling and patient management, we adopted a multistep molecular screening assay for detecting RB1 mutations | Patients with RB | 180 | 85 patients underwent genetic testing | Variant frequency in RB1 gene | Any RB |
| Ottaviani D, Parma D, Giliberto F, Ferrer M, Fandino A, Davila MT, et al. Spectrum of RB1 mutations in argentine patients: 20-years experience in the molecular diagnosis of retinoblastoma. Ophthalmic genetics. 2013;34(4):189-98. | To identify RB1 mutations in as many RB patients as possible and to correlate them to the patient phenotype, which allows better genetic counselling and clinical management of affected families. | Families with RB in Argentina | 144 | 144 | Variant frequency in RB1 gene | Any RB |
| Parma D, Ferrer M, Luce L, Giliberto F, Szijan I. RB1 gene mutations in Argentine retinoblastoma patients. Implications | To identify causative RB1 mutations in RB patients with different clinical presentations | Children with RB | 34 | 34 | Variant frequency in RB1 gene | Any RB |

| | | | | | | |
|---|---|---|---|---|---|---|
| for genetic counseling. PloS one. 2017;12(12):e0189736. | | | | | | |
| Parsam VL, Kannabiran C, Honavar S, Vemuganti GK, Ali MJ. A comprehensive, sensitive and economical approach for the detection of mutations in the RB1 gene in retinoblastoma. Journal of genetics. 2009;88(4):517-27. | To identify the RB1 mutations, we analysed 74 retinoblastoma patients by screening the exons and the promoter region of RB1 | Children with RB | 74 | 74 | Variant frequency in RB1 gene | Any RB |
| Pradhan MA, Ng Y, Strickland A, George PM, Raizis A, Warrington J, et al. Role of genetic testing in retinoblastoma management at a tertiary referral centre. Clinical & experimental ophthalmology. 2010;38(3):231-6. | To investigate the results of RB1 testing in retinoblastoma management in a tertiary referral centre | RB probands and their families | 20 | 20 | Variant frequency in RB1 gene | Any RB |
| Price EA, Price K, Kolkiewicz K, Hack S, Reddy MA, Hungerford JL, et al. Spectrum of RB1 mutations identified in 403 retinoblastoma | To present the spectrum of genetic and epigenetic changes identified in 194 tumours | Patients who underwent screening for RB | 209 | 209 | Variant frequency in RB1 gene | Any RB |

| | | | | | | |
|---|---|---|---|---|---|---|
| patients. Journal of medical genetics. 2014;51(3):208-14. | and 209 blood samples, from 403 unrelated RB patients. | | | | | |
| Reddy MA, Butt M, Hinds A-M, Duncan C, Price EA, Sagoo MS, et al. Prognostic Information for Known Genetic Carriers of RB1 Pathogenic Variants (Germline and Mosaic). Ophthalmology Retina. 2021;5(4):381-7. | To provide prognostic information from the identification of different genetic categories of potentially heritable retinoblastoma | Children with RB | 111 | 111 | Variant frequency in RB1 gene | Familial/ germline / bilateral RB |
| Richter S, Vandezande K, Chen N, Zhang K, Sutherland J, Anderson J, et al. Sensitive and efficient detection of RB1 gene mutations enhances care for families with retinoblastoma. American journal of human genetics. 2003;72(2):253-69. | To present a sensitive and efficient strategy to screen probands with retinoblastoma for RB1 mutations | Probands with RB | 378 | 378 | Variant frequency in RB1 gene | Any RB |
| Rojanaporn D, Boontawon T, Chareonsirisuthigul T, Thanapanpanich O, Attaseth T, Saengwimol D, et al. Spectrum of germline RB1 | To identify germline RB1 mutations and to correlate the mutations with clinical phenotypes of RB patients. | Children with RB | 52 | 52 | Variant frequency in RB1 gene | Any RB |

| | | | | | | |
|---|---|---|---|---|---|---|
| mutations and clinical manifestations in retinoblastoma patients from Thailand. Molecular vision. 2018;24:778-88. | | | | | | |
| Rojanaporn D, Chitphuk S, Iemwimangsa N, Chareonsirisuthigul T, Saengwimol D, Aroonroch R, et al. Germline RB1 Mutation in Retinoblastoma Patients: Detection Methods and Implication in Tumor Focality. Translational vision science & technology. 2022;11(9):30. | To generate a stepwise method to reduce the workload of full scale RB1 sequencing for germline mutation screening in RB patients | Children with RB | 42 | 42 | Variant frequency in RB1 gene | Any RB |
| Rushlow D, Piovesan B, Zhang K, Prigoda-Lee NL, Marchong MN, Clark RD, et al. Detection of mosaic RB1 mutations in families with retinoblastoma. Human mutation. 2009;30(5):842-51. | To detect low-level mosaicism for 11 recurrent RB1 CGA4TGA nonsense mutations | Probands with RB | 1020 | 1020 | Variant frequency in RB1 gene | Any RB |

| | | | | | | |
|---|---|---|---|---|---|---|
| Sagi M, Frenkel A, Eilat A, Weinberg N, Frenkel S, Pe'er J, et al. Genetic screening in patients with Retinoblastoma in Israel. Familial cancer. 2015;14(3):471-80. | To present experience in detecting the pathogenic mutations in blood samples, from 150 unrelated Rb patients and highlight the relevant counselling issues | Patients with RB | 150 | 150 | Variant frequency in RB1 gene | Any RB |
| Saliminejad K, Behnam B, Akbari MT, Khorshid HRK, Ghassemi F, Amoli FA, et al. Rapid detection of RB1 recurrent mutations in retinoblastoma by ARMS-PCR. Journal of genetics. 2013;92(2):e36-40. | To develop a multiplex ARMS-PCR method to screen for the most recurrent mutations in RB1 in patients with RB. | Children with RB | 121 | 121 | Variant frequency in RB1 gene | Any RB |
| Salviat F, Gauthier-Villars M, Carton M, Cassoux N, Lumbroso-Le Rouic L, Dehainault C, et al. Association Between Genotype and Phenotype in Consecutive Unrelated Individuals With Retinoblastoma. JAMA ophthalmology. 2020;138(8):843-50. | To assess the association between genotype and phenotype in patients with RB | Patients with RB | 1404 | 1404 | Variant frequency in RB1 gene | Any RB |
| Sampieri K, Hadjistilianou T, Mari F, Speciale C, Mencarelli | To perform a | Patients with RB | 35 | 35 | Variant frequency in RB1 gene | Any RB |

| | | | | | | |
|---|---|---|---|---|---|---|
| MA, Cetta F, et al. Mutational screening of the RB1 gene in Italian patients with retinoblastoma reveals 11 novel mutations. Journal of human genetics. 2006;51(3):209-16. | mutational screening of the RB1 gene in Italian patients affected by RB referred to the University of Siena | | | | | |
| Sanchez-Sanchez F, Ramirez-Castillejo C, Weekes DB, Beneyto M, Prieto F, Najera C, et al. Attenuation of disease phenotype through alternative translation initiation in low-penetrance retinoblastoma. Human mutation. 2007;28(2):159-67. | To report a novel mutation detected in 10 individuals of an extended family, only three of whom are affected by RB disease | Patients with RB | 10 | 10 | Variant frequency in RB1 gene | Familial/ germline / bilateral RB |
| Shahraki K, Ahani A, Sharma P, Faranoush M, Bahoush G, Torktaz I, et al. Genetic screening in Iranian patients with retinoblastoma. Eye (London, England). 2017;31(4):620-7. | To evaluate the RB1 mutations in 106 patients with retinoblastoma | Children with RB | 106 | 106 | Variant frequency in RB1 gene | Any RB |
| Singh J, Mishra A, Pandian AJ, Mallipatna AC, Khetan V, Sripriya | To use an improved NGS-based method to screen the RB1 gene in | Children with RB | 50 | 50 | Variant frequency in RB1 gene | Any RB |

| | | | | | | |
|---|---|---|---|---|---|---|
| S, et al. Next-generation sequencing-based method shows increased mutation detection sensitivity in an Indian retinoblastoma cohort. Molecular vision. 2016;22:1036-47. | the DNA isolated from blood or saliva samples from an Indian Rb cohort (50 cases) and detected all types of germline mutations | | | | | |
| Sippel KC, Fraioli RE, Smith GD, Schalkoff ME, Sutherland J, Gallie BL, et al. Frequency of somatic and germ-line mosaicism in retinoblastoma: implications for genetic counseling. American journal of human genetics. 1998;62(3):610-9. | To determine how often mosaicism could be documented in a group of families that included one or more individuals affected with retinoblastoma | Families with one family member having RB | 405 | 405 | Variant frequency in RB1 gene | Familial/ germline / bilateral RB |
| Siti-Norulhuda H, Hanani H, Siti-Raihan I, Jamalia R, Ariffin N, Shatriah I, et al. Mutational analysis in N- and C- termini of RB1 gene among sporadic retinoblastoma patients in Malaysia. | To detect mutations and single nucleotide polymorphisms (SNPs) in N- and C-termini of RB1 and to determine the association of these genetic variations with laterality and staging of retinoblastoma in | Children with RB and matched controls | 132 | 132 | Variant frequency in RB1 gene | Any RB |

| | | | | | | |
|---|---|---|---|---|---|---|
| International Medical Journal. 2012;19(4):369-72. | Malaysian children with RB. | | | | | |
| Sugano K, Yoshida T, Izumi H, Umezawa S, Ushiama M, Ichikawa A, et al. Outpatient clinic for genetic counseling and gene testing of retinoblastoma. International journal of clinical oncology. 2004;9(1):25-30. | To report experience in genetic counselling and gene testing for RB | Patients with RB | 51 | 51 | Variant frequency in RB1 gene | Any RB |
| Szijan I, Lohmann DR, Parma DL, Brandt B, Horsthemke B. Identification of RB1 germline mutations in Argentinian families with sporadic bilateral retinoblastoma. Journal of medical genetics. 1995;32(6):475-9. | To use two step strategy and detected the gene defect in six of 10 Argentinian patients | Families with one child affected by bilateral RB | 10 | 10` | Variant frequency in RB1 gene | Familial/ germline / bilateral RB |
| Taylor M, Dehainault C, Desjardins L, Doz F, Levy C, Sastre X, et al. Genotype-phenotype correlations in hereditary familial retinoblastoma. | To delineate the spectrum of RB1 germline mutations in familial Rb and to identify genotype–phenotype correlations as well as putative modifier | Pedigrees with a family history of RB | 165 | 165 | Variant frequency in RB1 gene | Familial/ germline / bilateral RB |

| | | | | | | |
|---|---|---|---|---|---|---|
| Human mutation. 2007;28(3):284-93. | | | | | | |
| Temming P, Viehmann A, Biewald E, Lohmann DR. Sporadic unilateral retinoblastoma or first sign of bilateral disease? The British journal of ophthalmology. 2013;97(4):475-80. | To identify clinical and genetic characteristics of children with sporadic unilateral retinoblastoma | Children with sporadic unilateral RB | 480 | 195 patients with genetic data | Variant frequency in RB1 gene | Sporadic RB |
| Tomar S, Sethi R, Sundar G, Quah TC, Quah BL, Lai PS. Mutation spectrum of RB1 mutations in retinoblastoma cases from Singapore with implications for genetic management and counselling. PloS one. 2017;12(6):e0178776. | To characterize the spectrum of RB1 mutations in RB cases seen among patients in Singapore to aid disease management | Infants with RB | 59 | 59 | Variant frequency in RB1 gene | Any RB |
| Tsai T, Fulton L, Smith BJ, Mueller RL, Gonzalez GA, Uusitalo MS, et al. Rapid identification of germline mutations in retinoblastoma by protein truncation testing. Archives of ophthalmology | To demonstrate the utility of protein truncation testing (PTT) for rapid detection and sequencing of germline mutations in the retinoblastoma tumour suppressor gene (RB1) | Probands with RB | 27 | 27 | Variant frequency in RB1 gene | Familial/ germline / bilateral RB |

| | | | | | | |
|---|---|---|---|---|---|---|
| (Chicago, Ill : 1960). 2004;122(2):239-48. | | | | | | |
| Umar BT, Rimayanti U, Pagarra H, Budu, Massi N, Muhiddin HS. Novel point mutation and intronic mutations of RB1 gene in retinoblastoma patients in Indonesia. Medical Journal of Indonesia. 2022;31(4):218-24. | To identify the germline mutation in the RB1 gene in patients with RB and their parents from the eastern part of Indonesia | Patients with RB and their parents and normal control subjects | 35 | 21 patients with RB | Variant frequency in RB1 gene | Any RB |
| Vural O, Atalay HT, Kayhan G, Tarlan B, Oral M, Okur A, et al. Clinical and genetic characteristics of retinoblastoma patients in a single center with four novel RB1 variants. International Journal of Ophthalmology. 2023;16(8):1274-9. | To assess the clinical and genetic characteristics of children diagnosed with retinoblastoma (RB) at Gazi University Faculty of Medicine's Department of Pediatric Oncology | Children with RB | 53 | 19 patients underwent NGS | Variant frequency in RB1 gene | Any RB |
| Xie Y, Xu XL, Wei WB. The rb1 mutation spectrum and genetic management consultation in pediatric patients with retinoblastoma in Beijing, China. Risk | To screen the structural mutations of the RB1 gene using gene capture and a preliminary exploration of the correlation between the genotypes and phenotypes | Children with RB | 12 | 12 | Variant frequency in RB1 gene | Sporadic RB |

| | | | | | | |
|---|---|---|---|---|---|---|
| Management and Healthcare Policy. 2021;14:3453-63. | | | | | | |
| Yousef YA, Tbakhi A, Al-Hussaini M, AlNawaiseh I, Saab A, Afifi A, et al. Mutational analysis of the RB1 gene and the inheritance patterns of retinoblastoma in Jordan. Familial cancer. 2018;17(2):261-8. | To evaluate the oncogenic mutations in the RB1 gene and the inheritance patterns of RB in the Jordanian patients | Children with RB and their parents | 50 | 40 probands | Variant frequency in RB1 gene | Any RB |
| Zajaczek S, Jakubowska A, Gorski B, Kurzawski G, Krzystolik Z, Lubinski J. Frequency and nature of germline Rb-1 gene mutations in a series of patients with sporadic unilateral retinoblastoma. European journal of cancer (Oxford, England : 1990). 1999;35(13):1824-7. | To study constitutional RB1 gene mutations in a series of 17 families with isolated unilateral retinoblastoma patients | Children with RB | 17 | 17 | Variant frequency in RB1 gene | Sporadic RB |
| Zhang L, Jia R, Zhao J, Fan J, Zhou Y, Han B, et al. Novel mutations in the RB1 gene from Chinese families with a history of retinoblastoma. | To perform genetic screening of Chinese RB patients and their family members for heritable RB1 mutations | Children with RB | 117 | 17 with RB | Variant frequency in RB1 gene | Any RB |

| | | | | | | |
|---|---|---|---|---|---|---|
| Tumour biology : the journal of the International Society for Oncodevelopmental Biology and Medicine. 2015;36(4):2409-20. | | | | | | |
| Zhang Z, Xiao Y-S, Shen R, Jiang H-C, Tan L, Li R-Q, et al. Next generation sequencing of RB1gene for the molecular diagnosis of ethnic minority with retinoblastoma in Yunnan. BMC medical genetics. 2020;21(1):230. | To use targeted NGS to screen probands and then Sanger sequencing was used to identify variants in other members of pedigrees | Non-consanguineous families with RB | 12 | 12 | Variant frequency in RB1 gene | Sporadic RB |
| Zhang Y, Wang Y, Huang D, Ma J, Zhang W, Gu H, et al. Correlation between Family RB1 Gene Pathogenic Variant with Clinical Features and Prognosis of Retinoblastoma under 5 Years Old. Disease markers. 2021;2021:9981028. | To analyze the impact of RB1 gene polymorphism on morbidity of RB and provide a molecular level diagnosis information | Children with RB | 40 | 40 | Variant frequency in RB1 gene | Any RB |

### XLHR

| Reference | Study aim | Population | Total population N in study | Eligible population for review | Study outcome | Disease category of included population |
|---|---|---|---|---|---|---|
| Acar S, BinEssa HA, Demir K, Al-Rijjal RA, Zou M, Catli G, et al. Clinical and genetic characteristics of 15 families with hereditary hypophosphatemia: Novel Mutations in PHEX and SLC34A3. PloS one. 2018;13(3):e0193388. | To analyse clinical and genetic characteristics of patients from 15 unrelated families from a different region of Turkey | 15 unrelated Turkish families with hereditary hypophosphatemia diagnosed based on clinical and laboratory assessment | 51 | 15 probands | Variant frequency in PHEX gene | Hereditary HR |
| Alikasifoglu A, Unsal Y, Gonc EN, Ozon ZA, Kandemir N, Alikasifoglu M. Long-term effect of conventional phosphate and calcitriol treatment on metabolic recovery and catch-up growth in children with PHEX mutation. Journal of pediatric endocrinology & | To investigate the genetic and clinical variability of XLHR in paediatric patients and analyse genotype–phenotype correlations focusing on short term, long term and pubertal impact of conventional treatment, being on the verge of an advance in treatment such as burosumab | 12 unrelated families diagnosed with HR, based on clinical, laboratory and radiologic investigations | 16 | 11 probands | Gene frequency and variant frequency in PHEX, FGF23 and CLCN5 | XLH |

| | | | | | | |
|---|---|---|---|---|---|---|
| metabolism : JPEM. 2021;34(12):1573-84. | | | | | | |
| Ariceta G, Beck-Nielsen SS, Boot AM, Brandi ML, Briot K, de Lucas Collantes C, et al. The International X-Linked Hypophosphatemia (XLH) Registry: first interim analysis of baseline demographic, genetic and clinical data. Orphanet journal of rare diseases. 2023;18(1):304. | To present the findings of the first interim analysis of the International X-Linked Hypophosphatemia registry | Patients with diagnosis of XLH based on the clinical judgement of an XLH-treating expert physician, using information such as family history and clinical, radiological and biochemical findings | 579 | 282 children with genetic test results | Gene frequency and variant frequency in PHEX, FGF23 and SLC34A3 | XLHR |
| Cao Y, You Y, Wang Q, Ren X, Li S, Li L, et al. Identification of six novel variants from nine Chinese families with hypophosphatemic rickets. BMC medical genomics. 2022;15(1):161. | To identify causative variants in nine unrelated Chinese families associated with HR, and to determine potential pathogenicity of the identified variants | Nine unrelated Chinese families with HR | 56 | 3 patients with age ≤18 yrs | Gene frequency and variant frequency in PHEX and SLC34A3 | HR |
| Capelli S, Donghi V, Maruca K, Vezzoli G, Corbetta S, Brandi ML, et al. Clinical and molecular heterogeneity in a | To describe the clinical features of patients with HR and the results of the genetic analyses in the family members | Patients with clinical and biochemical features of HR | 26 | 25 with age ≤18 yrs | Gene frequency and variant frequency in PHEX, FGF23, DMP1, MEPE, ENPP1 | HR |

| | | | | | | |
|---|---|---|---|---|---|---|
| large series of patients with hypophosphatemic rickets. Bone. 2015;79:143-9. | | | | | | |
| Cho HY, Lee BH, Kang JH, Ha IS, Cheong HI, Choi Y. A clinical and molecular genetic study of hypophosphatemic rickets in children. Pediatric research. 2005;58(2):329-33. | To perform a clinical and molecular genetic analysis in children with a clinical diagnosis of HR | Unrelated Korean children, diagnosed with HR by the Department of Pediatrics, Seoul National University Children's Hospital | 17 | 17 | Gene frequency and variant frequency in PHEX and FGF23 | HR |
| Del Pino M, Viterbo GL, Arenas MA, Perez Garrido N, Ramirez P, Marino R, et al. Growth in height and body proportion from birth to adulthood in hereditary hypophosphatemic rickets: a retrospective cohort study. Journal of endocrinological investigation. 2022;45(7):1349-58. | To analyse the long-term growth pattern, growth velocity, body disproportion and pubertal development in children with HHR under conventional treatment | Patients diagnosed with Hereditary HR | 96 | 42 | Variant frequency in PHEX gene | Hereditary HR |
| Durmaz E, Zou M, Al-Rijjal RA, Baitei EY, Hammami S, Bircan I, et al. Novel and de | To report 10 Turkish patients from 6 unrelated families. Four novel mutations were found in the PHEX gene and de novo | Patients with XLHR and their parents from 6 unrelated Turkish families | 10 from 6 families | 6 probands with age at diagnosis ≤18 yrs | Variant frequency in PHEX gene | HR |

| | | | | | | |
|---|---|---|---|---|---|---|
| novo PHEX mutations in patients with hypophosphatemic rickets. Bone. 2013;52(1):286-91. | mutations were exclusively present in female patients | | | | | |
| Gao Y, Wang Z-M, Li X-L. Analysis of 2 novel mutations of PHEX gene inducing X-linked dominant hypophosphatemia rickets in 2 families: Two case reports. Medicine. 2018;97(31):e11453. | To report two novel mutations of PHEX gene: c.497delG and c.388G>T found by Sanger sequencing in 2 families manifesting growth retardation, skeletal malformations, dental dysplasia | Family members affected with XLHR showing skeletal malformations, bowing of legs, and dental dysplasia | 2 | 2 | Variant frequency in PHEX gene | XLHR |
| Gaucher C, Walrant-Debray O, Nguyen T-M, Esterle L, Garabedian M, Jehan F. PHEX analysis in 118 pedigrees reveals new genetic clues in hypophosphatemic rickets. Human genetics. 2009;125(4):401-11. | To analyse the PHEX gene using classical sequencing in 209 patients with hypophosphatemic rickets, representing 118 families, and high-resolution melting curves analysis was evaluated as a possible screening method | Patients with HR (hypophosphatemia associated with tubular phosphate wasting, bone deformities and radiological signs of rickets) | 209 | 118 probands | Variant frequency in PHEX gene | HR |
| Guven A, Al-Rijjal RA, BinEssa HA, Dogan D, Kor Y, Zou M, et al. Mutational analysis of PHEX, FGF23 and CLCN5 in patients with | To investigate additional 14 Turkish patients with hypophosphataemic rickets from nine unrelated families and identified seven novel mutations | Patients with HR and their parents from three different regional hospitals in Turkey | 14 from 9 families | 9 probands | Gene frequency and variant frequency in PHEX, CLCN5, FGF23 | HR |

| | | | | | | |
|---|---|---|---|---|---|---|
| hypophosphataemic rickets. Clinical endocrinology. 2017;87(1):103-12. | | | | | | |
| Holm IA, Nelson AE, Robinson BG, Mason RS, Marsh DJ, Cowell CT, et al. Mutational analysis and genotype-phenotype correlation of the PHEX gene in X-linked hypophosphatemic rickets. The Journal of clinical endocrinology and metabolism. 2001;86(8):3889-99. | To report our mutational analysis of all 22 PHEX exons in 28 individuals with HYP and 5 PHEX exons in 13 individuals with HYP (familial and sporadic cases). To present phenotype analysis of all patients in whom a mutation was detected in this and our previous study and all available affected family members for familial cases. To report our analysis of potential correlation between phenotype and genotype and our investigation of reverse anticipation and gene dosage effect | Unrelated patients with HR | 50 | 50 | Variant frequency in PHEX gene | XLHR |
| Huang Y, Mei L, Pan Q, Tan H, Quan Y, Gui B, et al. Novel de novo nonsense mutation of the PHEX gene (p.Lys50Ter) in a Chinese patient with hypophosphatemic rickets. Gene. 2015;565(1):150-4. | To clinically characterized four unrelated families with hypophosphatemia, bone abnormalities, short stature, and dentin malformation | Four Chinese families with XLHR based on clinical studies and molecular diagnostics | 7 | 4 probands with age at diagnosis ≤18 yrs | Variant frequency in PHEX gene | HR |
| Jacob P, Bhavani GS, Udupa P, Wang Z, | To understand the phenotypic and genotypic spectrum of | Unrelated individuals from different families | 10 | 8 individuals with age at | Gene frequency and variant | Hereditary rickets |

| | | | | | | |
|---|---|---|---|---|---|---|
| Hariharan SV, Delampady K, et al. Exome Sequencing in Monogenic Forms of Rickets. Indian journal of pediatrics. 2023. | genetic forms of rickets in 10 families | with suspected hereditary rickets. | | diagnosis ≤18 yrs | frequency in CYP27B1, VDR, PHEX, SLC2A2, SLC34A3 | |
| Jap T-S, Chiu C-Y, Niu D-M, Levine MA. Three novel mutations in the PHEX gene in Chinese subjects with hypophosphatemic rickets extends genotypic variability. Calcified tissue international. 2011;88(5):370-7. | To identify PHEX gene mutations in Chinese patients with hypophosphatemic rickets who are living in Taiwan | Unrelated Chinese patients with clinical and biochemical manifestations of early-onset or congenital hypophosphatemic rickets, including hypophosphatemia, short stature, and lower extremity deformities | 9 | 9 patients with age at time of testing ≤18 yrs | Variant frequency in PHEX gene | HR |
| Jimenez M, Ivanovic-Zuvic D, Loureiro C, Carvajal CA, Cavada G, Schneider P, et al. Clinical and molecular characterization of Chilean patients with X-linked hypophosphatemia. Osteoporosis international : a journal established as result of cooperation between the European Foundation | To describe the clinical, biochemical, and molecular presentation of patients with XLH in Chile, with the purpose of increasing awareness in our medical community about the importance of early diagnosis, pertinent treatment, and prevailing unmet needs in care | Patients with hereditary hypophosphatemic rickets from different centres throughout Chile | 26 | 17 probands | Gene frequency and variant frequency in CLCN5, CYP27B1, CYP2R1, DMP1, ENPP1, FGF23, PHEX, SLC34A4, VDR, ALPL, FAH, FAM20C, FGFR1 | Hereditary HR |

| | | | | | | |
|---|---|---|---|---|---|---|
| for Osteoporosis and the National Osteoporosis Foundation of the USA. 2021;32(9):1825-36. | | | | | | |
| Kinoshita Y, Saito T, Shimizu Y, Hori M, Taguchi M, Igarashi T, et al. Mutational analysis of patients with FGF23-related hypophosphatemic rickets. European journal of endocrinology. 2012;167(2):165-72. | To conduct a detailed analysis to identify the aetiology of FGF23-related hypophosphatemic rickets in 27 consecutive Japanese patients | Patients with clinical and biochemical manifestations of congenital hypophosphatemic rickets with FGF23 above 30 pg/ml | 27 | 9 patients with age ≤18 yrs | Gene frequency of PHEX, FGF23, DMP1, ENPP1, and variant frequencies in PHEX gene | FGF23-related HR |
| Li S-S, Gu J-M, Yu W-J, He J-W, Fu W-Z, Zhang Z-L. Seven novel and six de novo PHEX gene mutations in patients with hypophosphatemic rickets. International journal of molecular medicine. 2016;38(6):1703-14. | To screen a total of 18 affected families for mutations in the PHEX gene in order to interpret the mutation traits in Chinese patients and potentially provide evidence of a critical domain in PHEX protein | Unrelated Chinese families with hypophosphatemic rickets of Han ethnicity | 43 | 8 probands | Variant frequency in PHEX gene | HR |
| Liao H, Zhu H-M, Liu H-Q, Li L-P, Liu S-L, Wang H. Two novel variants of the PHEX gene in patients with | To report on two variants of the PHEX gene in two Chinese families affected by XLHR. The primary aim was to provide a genetic diagnosis for the | Two Chinese families affected by XLHR | 23 | 2 probands | Variant frequency in PHEX gene | XLHR |

| | | | | | | |
|---|---|---|---|---|---|---|
| X-linked dominant hypophosphatemic rickets and prenatal diagnosis for fetuses in these families. International journal of molecular medicine. 2018;41(4):2012-20. | affected family members and to analyse the underlying genotype-phenotype correlations | | | | | |
| Lin X, Zhu Y, Luo J, Huang J. Genetic analysis of three families with X-linked dominant hypophosphatemic rickets. Journal of pediatric endocrinology & metabolism : JPEM. 2018;31(7):789-97. | To identify the gene mutations responsible for three cases of XLH and its clinical phenotype | Three children with XLHR with XLH-affected mothers | 3 | 3 | Variant frequency in PHEX gene | XLHR |
| Lin Y, Xu J, Li X, Sheng H, Su, L, Wu M, Cheng J, Huang Y, Mao X, Zhou Z, Zhang W, Li C, Cai Y, Wu D, Lu Z, Yin X, Zeng C, Liu L. Novel variants and uncommon cases among southern Chinese children with X-linked hypophosphatemia. Journal of | To provide clinical and mutational characteristics of 65 XLH paediatric patients in southern China | Paediatric hypophosphatemia patients and 123 relatives from 76 unrelated families in Guangzhou Women and Children's Medical Center | 80 | 80 | Variant frequency in PHEX gene | HR |

| | | | | | | |
|---|---|---|---|---|---|---|
| Endocrinological Investigation. 2020;43(11):p1577-90. | | | | | | |
| Lo F-S, Kuo M-T, Wang C-J, Chang C-H, Lee Z-L, Van Y-H. Two novel PHEX mutations in Taiwanese patients with X-linked hypophosphatemic rickets. Nephron Physiology. 2006;103(4):p157-63. | To describe two novel mutations of the PHEX gene, a missense mutation (L206W) in exon 5, and a frameshift mutation (nucleotide 1826/1830delAAAAG, stop after codon 610) in exon 18, in two unrelated Taiwanese families with XLH. To analyse the laboratory and radiographic findings for these patients | Two unrelated Taiwanese families with XLHR in Taiwan | 6 | 2 probands | Variant frequency in PHEX gene | XLHR |
| Marik B, Bagga A, Sinha A, Hari P, Sharma A. Genetics of Refractory Rickets: Identification of Novel PHEX Mutations in Indian Patients and a Literature Update. Journal of pediatric genetics. 2018;7(2):47-59. | To identify mutations responsible for the pathogenesis of refractory rickets | Patients with refractory rickets | 37 | 32 probands | Variant frequency in PHEX gene | HR |
| Marik B, Bagga A, Sinha A, Khandelwal P, Hari P, Sharma A. Genetic and clinical profile of patients with | To prospectively examined the aetiology of hypophosphatemic rickets in consecutive patients referred to this tertiary care centre, by testing for presence of disease-causing variations to | Patients with hypophosphatemic rickets | 66 | 65 individuals with age ≤18 yrs | Gene frequency and variant frequency in PHEX, FGF23, DMP1, ENPP1, CLCN5, CTNS, | HR |

| | | | | | | |
|---|---|---|---|---|---|---|
| hypophosphatemic rickets. European journal of medical genetics. 2022;65(8):104540. | enable molecular diagnosis and appropriate therapy | | | | SLC2A2, GATM, SLC34A1, EHHADH, SLC4A1, ATP6V1B1, ATP6V0A4, CYP27B1, VDR and FGFR1 | |
| Morey M, Castro-Feijoo L, Barreiro J, Cabanas P, Pombo M, Gil M, et al. Genetic diagnosis of X-linked dominant Hypophosphatemic Rickets in a cohort study: tubular reabsorption of phosphate and 1,25(OH)2D serum levels are associated with PHEX mutation type. BMC medical genetics. 2011;12:116. | To perform genetic diagnosis in a cohort of patients with clinical diagnosis of HR | Patients clinically diagnosed with HR and one pre-symptomatic infant belonging to 36 unrelated families | 43 | 36 probands | Variant frequency in PHEX gene | HR |
| Obara-Moszynska M, Rojek A, Kolesinska Z, Jurkiewicz D, Chrzanowska KH, Niedziela M. X-linked hypophosphataemic rickets in children: clinical phenotype, therapeutic | To investigate the clinical phenotype, therapeutic strategies, and molecular background of HR in children hospitalised in our clinic | Children diagnosed with HR due to their clinical and biochemical profiles | 11 | 11 | Gene frequency and variant frequency in PHEX and FGF23 | HR |

| | | | | | | |
|---|---|---|---|---|---|---|
| strategies, and molecular background. Endokrynologia Polska. 2021;72(2):108-19. | | | | | | |
| Pecoraro C, Fioretti T, Perruno A, Klain A, Cioffi D, Ambrosio A, et al. De Novo Large Deletions in the PHEX Gene Caused X-Linked Hypophosphataemic Rickets in Two Italian Female Infants Successfully Treated with Burosumab. Diagnostics (Basel, Switzerland). 2023;13(15). | To report the clinical and genetic features of two Italian female infants with sporadic HR who successfully responded to Burosumab | Two Italian female infants with sporadic HR who successfully responded to Burosumab | 2 | 2 | Variant frequency in PHEX gene | HR |
| Popowska E, Pronicka E, Sulek A, Jurkiewicz D, Rowinska E, Sykut-Cegielska J, et al. X-linked hypophosphatemia in Polish patients. 2. Analysis of clinical features and genotype-phenotype correlation. Journal of applied genetics. 2001;42(1):73-88. | To present biochemical, stomatological, audiological and anthropometrical characteristics of patients with established genotypes and analyse correlations between clinical symptoms and the type and localisation of the PHEX gene mutations | Individuals from 36 unrelated families with XLH | 59 | 31 probands | Variant frequency in PHEX gene | XLHR |

| | | | | | | |
|---|---|---|---|---|---|---|
| Quinlan C, Guegan K, Offiah A, Neill RO, Hiorns MP, Ellard S, et al. Growth in PHEX-associated X-linked hypophosphatemic rickets: the importance of early treatment. Pediatric nephrology (Berlin, Germany). 2012;27(4):581-8. | Secondary goals were to reports results of mutation screening and genotype-phenotype relationships | Children with XLHR | 61 | 46 who were screened for PHEX mutation | Gene frequency and variant frequency in PHEX | XLHR |
| Rafaelsen S, Johansson S, Raeder H, Bjerknes R. Hereditary hypophosphatemia in Norway: a retrospective population-based study of genotypes, phenotypes, and treatment complications. European journal of endocrinology. 2016;174(2):125-36. | To explore the prevalence, genotypes, phenotypic spectrum, treatment response, and complications of treatment in the Norwegian population of children with HH | Children with HH based on biochemical markers +/- FH or genetic diagnosis from all paediatric hospital departments in Norway | 28 | 19 probands | Gene frequency and variant frequency in PHEX, FGF23, DMP1, ENPP1, KL, FAM20C | HR |
| Sant' Ana I, Torrini R, Alves Coelho MC, Cantoni J, Madeira M, Ribeiro M. X-linked hypophosphatemic rickets: Description of | To describe new mutations to improve our knowledge about this rare disease, its genetic bases and possible associated phenotypes, laboratory changes, and family history | Patients with a previously presumptive diagnosis (based on clinical history, laboratory findings, and | 26 | 15 patients with age at diagnosis ≤18 yrs and underwent | Variant frequency in PHEX gene | XLH |

| | | | | | | |
|---|---|---|---|---|---|---|
| seven new variants in patients followed up in reference hospitals in Rio de Janeiro. Molecular genetics & genomic medicine. 2022;10(6):e1941. | | physical examination) of XLHR | | sequence analysis | | |
| Song HR, Park JW, Cho DY, Yang JH, Yoon HR, Jung SC. PHEX gene mutations and genotype-phenotype analysis of Korean patients with hypophosphatemic rickets. Journal of Korean medical science. 2007;22(6):981-6. | To describe eight different PHEX mutations identified in 15 unrelated Korean patients with hypophosphatemic rickets, including five novel mutations. To analyse the correlation between genotype and phenotype, phenotypes were compared between groups with and without a mutation, in terms of mutation location, mutation type, and sex | Patients with a diagnosis of XLHR based on clinical, radiological, and laboratory findings by specialists at the Korea University Guro Hospital | 15 | 13 patients with age at diagnosis ≤18 yrs | Variant frequency in PHEX gene | HR |
| Tavana N, Ting TH, Lai K, Kennerson ML, Thilakavathy K. Whole exome sequencing identifies two novel variants in PHEX and DMP1 in Malaysian children with hypophosphatemic rickets. Italian journal of pediatrics. 2022;48(1):193. | To identify the gene variants responsible for HR in three cases of Malaysian origin from three independent families and to describe their clinical, biochemical, and radiological features | Paediatric patients with HR in Malayasia presenting with clinical and biochemical features of HR including lower limb bowing deformity, short stature, low serum phosphate, elevated alkaline phosphatase, normal serum calcium, and normal 25-hydroxyvitamin D | 3 | 3 | Gene frequency and variant frequency in PHEX, DMP1, FGF23, SLC34A3, CLCN5 | HR |

| | | | | | | |
|---|---|---|---|---|---|---|
| Xu X, Tang Y, Luo Z, Song H, Xu W. Role of serum cytokines in the diagnosis and treatment of pediatric hemophagocytic lymphohistiocytosis. Pediatric Blood and Cancer. 2022;69(SUPPL 1). | To describe the clinical features, PHEX gene mutation sites, and iFGF23 levels of 29 patients with XLH as well as recommend optimal operation timing | Chinese patients with XLHR | 29 | 20 probands | Variant frequency in PHEX gene | XLHR |
| Turan I, Erdem S, Kotan LD, Ozdemir Dilek S, Tastan M, Gurbuz F, et al. Experience with the targeted next-generation sequencing in the diagnosis of hereditary hypophosphatemic rickets. Journal of pediatric endocrinology & metabolism : JPEM. 2021;34(5):639-48. | To present clinical and molecular genetic features of 35 consecutive patients diagnosed at one tertiary care centre | 18 consecutive probands and their 17 affected family members with HHR who were admitted to the Cukurova University | 35 | 18 probands | Gene frequency and variant frequency in PHEX, SLC34A3, ENPP1, SLC34A1, CLCN5, DMP1 | Hereditary HR |
| Tyynismaa H, Kaitila I, Nanto-Salonen K, Ala-Houhala M, Alitalo T. Identification of fifteen novel PHEX gene mutations in Finnish patients with | To screen the PHEX gene in Finnish patients with hypophosphatemia | Finish patients with early onset clinical, radiographic, and laboratory hypophosphatemic rickets with favourable outcome of oral | 23 | 23 | Variant frequency in PHEX gene | HR |

| | | | | | | |
|---|---|---|---|---|---|---|
| hypophosphatemic rickets. Human mutation. 2000;15(4):383-4. | | phosphate and alfa-calcidol treatment, and knowledge of their family history | | | | |
| Xia WB, Meng XW, Jiang Y, Li M, Xing XP, Pang L, et al. Three novel mutations of the PHEX gene in three Chinese families with X-linked dominant hypophosphatemic rickets. Calcified Tissue International. 2007;81(6):415-20. | To analyse the molecular basis in three unrelated Chinese families with XLH, we determined the nucleotide sequence of the PHEX gene and fibroblast growth factor 23 (FGF23) gene of affected members | Patients with XLHR according to the evaluation of their medical history, physical examination, radiologic evidence of rachitic disease, unremarkable serum calcium, electrolyte concentrations, hypophosphatemia, and X-linked dominant transmission | 8 | 3 patients with age at diagnosis ≤18 yrs | Gene frequency and variant frequency in PHEX and FGF23 | HR |
| Yue H, Yu JB, He JW, Zhang Z, Fu WZ, Zhang H, et al. Identification of two novel mutations in the PHEX gene in Chinese patients with hypophosphatemic rickets/osteomalacia. Plos One. 2014;9(5):9. | To identify PHEX gene mutations and describe the clinical features observed in 6 unrelated Chinese families and 3 sporadic patients with hypophosphatemic rickets/osteomalacia | Patients from Chinese Han ethnicity with a diagnosis of XLH based on clinical manifestations, radiology results, skeletal deformities, growth impairment, and laboratory results that indicated the occurrence of hypophosphatemia and renal phosphate wasting | 45 | 8 individuals with age ≤18 yrs | Variant frequency in PHEX gene | XLHR |
| Zheng B, Wang C, Chen Q, Che R, Sha Y, Zhao F, et al. | To investigate the mutational spectrum of PHEX in 53 patients who received a clinical | Unrelated patients with XLHR with confirmed pathogenic or likely | 53 | 53 | Variant frequency in PHEX gene | XLHR |

| | | |
|---|---|---|
| Functional Characterization of PHEX Gene Variants in Children With X-Linked Hypophosphatemic Rickets Shows No Evidence of Genotype-Phenotype Correlation. Journal of bone and mineral research : the official journal of the American Society for Bone and Mineral Research. 2020;35(9):1718-25. | and biochemical diagnosis of HR | pathogenetic variations in PHEX gene and presence of hypophosphatemia and rickets |

### fHLH

| Reference | Study aim | Population | Total population N in study | Eligible population for review | Study outcome | Disease category of included population |
|---|---|---|---|---|---|---|
| Ahmari AA, Alsmadi O, Sheereen A, Elamin T, Jabr A, El-Baik L, et al. Genetic and clinical characteristics of pediatric patients with familial hemophagocytic | Explore spectrum of inherited variants of the eight FHL genes in a large cohort of Saudi FHL patients | Saudi patients diagnosed with HLH | 87 | 87 | Gene and variant frequency in PRF1, UNC13D, STXBP2, STX11, LYST, RAB27A and XIAP | HLH |

| | | | | | | |
|---|---|---|---|---|---|---|
| lymphohistiocytosis. Blood research. 2021;56(2):86-101. | | | | | | |
| Almalky MA, Saleh SHA, Baz EG, Fakhr AE. Clinico-laboratory profile and perforin gene mutations of pediatric hemophagocytic lymphohistiocytosis cases: a five-year single center study. The Pan African medical journal. 2020;36:354. | Illustrate clinical and laboratory features, management outcomes and PRF1 gene mutation of HLH patients | Paediatric patients diagnosed with the HLH 2004 criteria | 18 | 18 | Variant frequency in PRF1 | HLH |
| Amirifar P, Ranjouri MR, Abolhassani H, Moeini Shad T, Almasi-Hashiani A, Azizi G, et al. Clinical, immunological and genetic findings in patients with UNC13D deficiency (FHL3): A systematic review. Pediatric allergy and immunology : official publication of the European Society of Pediatric Allergy and Immunology. 2021;32(1):186-97. | Systematically review clinical features, immunologic data and genetic findings of confirmed patients with fhlh3 and determine genotype-phenotype relationship | Patients with genetically confirmed UNC13D mutation | 322 | 322 | Variant type frequency in UNC13D | single gene/variant |

| | | | | | | |
|---|---|---|---|---|---|---|
| Ammann S, Lehmberg K, Zur Stadt U, Klemann C, Bode SFN, Speckmann C, et al. Effective Immunological Guidance of Genetic Analyses Including Exome Sequencing in Patients Evaluated for Hemophagocytic Lymphohistiocytosis. Journal of clinical immunology. 2017;37(8):770-80. | Establish whether immunological studies can be used as a decision tool for the need and extent of genetic studies in children referred for suspected HLH | Children referred with suspected HLH | 290 | 290 | Gene frequency of PRF1, SH2D1A, BIRC4A, UNC13D, STXBP2, STX11, RAB27A, LYST and AP3B1 | HLH |
| Babol-Pokora K, Wolowiec M, Popko K, Jaworowska A, Bryceson YT, Tesi B, et al. Molecular Genetics Diversity of Primary Hemophagocytic Lymphohistiocytosis among Polish Pediatric Patients. Archivum immunologiae et therapiae experimentalis. 2021;69(1):31. | Present molecular genetics among paediatric HLH patients | Patients with HLH based on clinical + biochemical criteria and/or presence of relevant genetic mutations | 54 | 36 with genetic studies | Gene and variant frequency in PRF1, UNC13D, STX11, STXBP2, RAB27A, SH2D1A, XIAP, MAGT1, AP3B2, ITK, CD27, LYST and NLRC4 | HLH |
| Bayram C, Tahtakesen TN, Arslantaş E, Yilmaz E, Özdemir GN, | Evaluate the clinical and laboratory characteristics that impact prognosis and | Patients with primary HLH | 41 | 23 who had HLH mutation analysis performed | Gene frequency of PRF1, UNC13D and STX11 | HLH |

| | | | | | | |
|---|---|---|---|---|---|---|
| Pasli Uysalol E, et al. Prognostic factors and long-term outcomes in 41 children with primary hemophagocytic lymphohistiocytosis: Report of a single-center experience and review of the literature. J Pediatr Hematol Oncol. 2023;45(5):262-6. | long term outcomes in patients with primary HLH | | | | | |
| Beken B, Aytac S, Balta G, Kuskonmaz B, Uckan D, Unal S, et al. The clinical and laboratory evaluation of familial hemophagocytic lymphohistiocytosis and the importance of hepatic and spinal cord involvement: a single center experience. Haematologica. 2018;103(2):231-6. | Evaluate the demographic, clinical and laboratory findings of FHL patients | Patients with primary HLH | 37 | 37 | Gene frequency of PRF1, UNC13D and STX11 | HLH |
| Cetica V, Sieni E, Pende D, Danesino C, De Fusco C, Locatelli F, et al. Genetic predisposition to | Describe the genetic basis of disease for 500 patients with HLH in the national registry | Clinical diagnosis of HLH | 500 | 426 with DNA studies carried out | Gene and variant frequency in PRF1, UNC13D, STXBP2, STX11 and RAB27A | HLH |

| | | | | | | |
|---|---|---|---|---|---|---|
| hemophagocytic lymphohistiocytosis: Report on 500 patients from the Italian registry. The Journal of allergy and clinical immunology. 2016;137(1):188-96.e4. | | | | | | |
| Chen X, Wang F, Zhang Y, Teng W, Wang M, Nie D, et al. Genetic variant spectrum in 265 Chinese patients with hemophagocytic lymphohistiocytosis: Molecular analyses of PRF1, UNC13D, STX11, STXBP2, SH2D1A, and XIAP. Clinical genetics. 2018;94(2):200-12. | Inspect the inherited variant spectrum of 6 FHL genes in a large cohort of chinese HLH patients and compare with other ethnic and regional populations | Patients with HLH meeting the HLH-2004 criteria admitted to hospital | 283 | 265 with DNA sample of sufficient quality for analysis | Gene and variant frequency in PRF1, UNC13D, STXBP2, STX11, SH2D1A and XIAP | HLH |
| Chinn IK, Eckstein OS, Peckham-Gregory EC, Goldberg BR, Forbes LR, Nicholas SK, et al. Genetic and mechanistic diversity in pediatric hemophagocytic lymphohistiocytosis. | Evaluate the genomic spectrum and associated outcomes of children with HLH | Children diagnosed with HLH using the HLH 2004 criteria | 122 | 101 with genetic testing performed | Gene frequency in PRF1, RAB27A, STX11, UNC13D, CARMIL2, CASP10, CYBB, DOCK8, LRBA, MCM3AP, MCM9, NCF1, PIK3CD, RAG1, RAG2, STAT1, STAT2, STAT3, TTC7A, WAS, NLRC3, NLRC4, NLRP12, NLRP13, NLRP4, NRAS, | HLH |

| | | | | | | |
|---|---|---|---|---|---|---|
| Blood. 2018;132(1):89-100. | | | | | ARHGEF6, ERCC4, G3BP1, IL16, RASGRP3, STAT4, TREM2 and variant frequency in PRF1, RAB27A, STX11, UNC13D and LYST | |
| Clementi R, zur Stadt U, Savoldi G, Varoitto S, Conter V, De Fusco C, et al. Six novel mutations in the PRF1 gene in children with haemophagocytic lymphohistiocytosis. Journal of medical genetics. 2001;38(9):643-6. | Report 6 novel PRF1 mutations in children with HLH | Families in which index case fulfilled diagnostic criteria | 10 | 10 | Gene and variant frequency in PRF1 | HLH |
| Cleves D, Lotero V, Medina D, Perez PM, Patino JA, Torres-Canchala L, et al. Pediatric hemophagocytic lymphohistiocytosis: A rarely diagnosed entity in a developing country. BMC pediatrics. 2021;21(1):411. | Determine the frequency of haemophagocytic syndrome and describe demographic, clinical and outcome characteristics | Hospitalised children diagnosed with HLH based in HLH-2004 criteria | 21 | 3 with genetic testing performed | Gene and variant frequency in G6PC3, LYST, XIAP and UNC13D | HLH |
| Elsharkawy A, Assem H, Salama M, Mikhael N, Zeid MY, El Chazli Y. Clinical Characteristics | Epidemiological study if Egyptian children with HLH | Children <16y fulfilling the HLH-2004 criteria | 101 | 57 with genetic testing performed | Gene frequency of PRF1, UNC13D, STXBP2, LYST, RAB27A, SH2D1A | HLH |

| | | | | | | |
|---|---|---|---|---|---|---|
| and Outcomes of 101 Children with Hemophagocytic Lymphohistiocytosis: A Four-Year Single-Center Experience from Egypt. Pediatric hematology and oncology. 2021;38(3):194-207. | | | | | | |
| Elstak ED, te Loo M, Tesselaar K, van Kerkhof P, Loeffen J, Grivas D, et al. A novel Dutch mutation in UNC13D reveals an essential role of the C2B domain in munc13-4 function. Pediatric blood & cancer. 2012;58(4):598-605. | Describe seven patients with UNC13D mutations | Patients fulfilling HLH criteria | 7 | 7 | Variant frequency in UNC13D | single gene/variant |
| Elyamany G, Alzahrani A, Elfaraidi H, Alsuhaibani O, Othman N, Al Mussaed E, et al. Hemophagocytic Lymphohistiocytosis: Single-Center Series of 12 Cases from Saudi Arabia. Clinical medicine insights | Present 12 cases of HLH | Patients from Saudi with HLH using HLH-2004 criteria | 12 | 10 with genetic testing performed | Gene frequency of PRF1, UNC13D, STXBP2 and STX11 | HLH |

| | | | | | | |
|---|---|---|---|---|---|---|
| Pediatrics. 2016;10:21-6. | | | | | | |
| Feldmann J, Le Deist F, Ouachee-Chardin M, Certain S, Alexander S, Quartier P, et al. Functional consequences of perforin gene mutations in 22 patients with familial haemophagocytic lymphohistiocytosis. British journal of haematology. 2002;117(4):965-72. | Describe the spectrum of clinical, immunological and genetic features in patients whose FHL results from PRF1 deficiency | Patients with FHL cause by PRF1 deficiency | 14 | 14 | Variant frequency in PRF1 | single gene/variant |
| Ferreira M, Martins J, Silvestre C, Abadesso C, Matias E, Loureiro H, et al. Familial haemophagocytic lymphohistiocytosis: two case reports. BMJ case reports. 2010;2010. | Describe two cases of primary HLH with different clinical presentation and evolution | Children presenting to A&E following prolonged fever without accompanying symptoms | 2 | 2 | Gene and variant frequency in PRF1, UNC13D and STX11 | HLH |
| Goransdotter Ericson K, Fadeel B, Nilsson-Ardnor S, Soderhall C, Samuelsson A, Janka G, et al. Spectrum of perforin gene mutations in familial hemophagocytic | Determine the frequency and type of mutations in the perforin gene by direct sequencing of DNA from a large set of well-defined families affected by FHL | Unrelated families with one or more children affected with FHL -- 1 child per family investigated in case where multiple affected siblings | 34 | 34 | Variant frequency in PRF1 | HLH |

| | | | | | | |
|---|---|---|---|---|---|---|
| lymphohistiocytosis. American journal of human genetics. 2001;68(3):590-7. | | | | | | |
| Gurgey A, Unal S, Okur H, Orhan D, Yurdakok M. Neonatal primary hemophagocytic lymphohistiocytosis in Turkish children. Journal of pediatric hematology/oncology. 2008;30(12):871-6. | Present 8 cases of neonatal onset primary HLH | Neonates with HLH | 8 | 7 probands | Gene and variant frequency in PRF1 and UCN13D | HLH |
| Horne A, Ramme KG, Rudd E, Zheng C, Wali Y, al-Lamki Z, et al. Characterization of PRF1, STX11 and UNC13D genotype-phenotype correlations in familial hemophagocytic lymphohistiocytosis. British journal of haematology. 2008;143(1):75-83. | Characterise the clinical phenotype in patients with FHL and analyse whether a correlation could be found to patients with the three genotypes prf1, UNC13D, STX11 and the no known gene defect. | Patients treated for FHL | 76 | 65 probands | Gene frequency of PRF1, UNC13D and STX11 | fHLH (4 genes only) |
| Imashuku S, Ueda I, Teramura T, Mori K, Morimoto A, Sako M, et al. Occurrence of haemophagocytic lymphohistiocytosis at | Determine correlations of onset age, clinical characteristics, underlying disease and outcomes | Patients under 1 registered to the HLH study centre in Kyoto | 96 | 28 sequenced | Gene frequency of PRF1 and UNC13D | HLH |

| | | | | | | |
|---|---|---|---|---|---|---|
| less than 1 year of age: analysis of 96 patients. European journal of pediatrics. 2005;164(5):315-9. | | | | | | |
| Ishii E, Ueda I, Shirakawa R, Yamamoto K, Horiuchi H, Ohga S, et al. Genetic subtypes of familial hemophagocytic lymphohistiocytosis: correlations with clinical features and cytotoxic T lymphocyte/natural killer cell functions. Blood. 2005;105(9):3442-8. | Analyse relationships among the clinical features, genetic defects and CTL/NK cell functions of FHL patients with different molecular subtypes | Patients meeting diagnostic criteria for FHL | 57 | 57 | Gene and variant frequency in PRF1 and UNC13D | HLH |
| Kaya Z, Bay A, Albayrak M, Kocak U, Yenicesu I, Gursel T. Prognostic Factors and Long-Term Outcome in 52 Turkish Children With Hemophagocytic Lymphohistiocytosis. Pediatric critical care medicine : a journal of the Society of Critical Care Medicine | Investigate the aetiology, clinical characteristics, prognostic risk factors and long-term outcome in a large group of patients with HLH | Patients with HLH | 52 | 25 underwent genetic analysis | Gene and variant frequency in PRF1, UNC13D, STX11, STXBP2, RAB27A and LYST | HLH |

| | | | | | | |
|---|---|---|---|---|---|---|
| and the World Federation of Pediatric Intensive and Critical Care Societies. 2015;16(6):e165-73. | | | | | | |
| Koh K-N, Im HJ, Chung N-G, Cho B, Kang HJ, Shin HY, et al. Clinical features, genetics, and outcome of pediatric patients with hemophagocytic lymphohistiocytosis in Korea: report of a nationwide survey from Korea Histiocytosis Working Party. European journal of haematology. 2015;94(1):51-9. | Investigate the epidemiological features and ethnic characteristics of Korean paediatric patients with HLH, as well as the general clinical features and prognostic factors of HLH | Patients diagnosed with HLH | 251 | 85 underwent genetic testing | Gene frequency of PRF1 and UNC13D | HLH |
| Lee SM, Sumegi J, Villanueva J, Tabata Y, Zhang K, Chakraborty R, et al. Patients of African ancestry with hemophagocytic lymphohistiocytosis share a common haplotype of PRF1 with a 50delT mutation. The Journal | Explore the links between a specific PRF1 mutation and ethnic and disease characteristics | Patients with HLH referred for genetic testing and who harboured monoallelic or biallelic 50delt mutation on PRF1 | 23 | 23 | Variant frequency in PRF1 | single gene/variant |

| | | | | | | |
|---|---|---|---|---|---|---|
| of pediatrics. 2006;149(1):134-7. | | | | | | |
| Lee SM, Villanueva J, Sumegi J, Zhang K, Kogawa K, Davis J, et al. Characterisation of diverse PRF1 mutations leading to decreased natural killer cell activity in North American families with haemophagocytic lymphohistiocytosis. Journal of Medical Genetics. 2004;41(2):137-44. | Investigate the immunophenotypes of HLH in relation to the presence or absence of disease-causing mutations in PRF1 | North American families with children diagnosed with primary HLH | 50 | 43 sequenced | Variant frequency in PRF1 | HLH |
| Lee W-I, Chen S-H, Hung I-J, Yang C-P, Jaing T-H, Chen C-J, et al. Clinical aspects, immunologic assessment, and genetic analysis in Taiwanese children with hemophagocytic lymphohistiocytosis. The Pediatric infectious disease journal. 2009;28(1):30-4. | Define the clinical spectrum, immunology and candidate genes in HLH in Taiwanese patients | Patients diagnosed with HLH | 32 | 26 underwent genetic analysis | Gene frequency of PRF1, UNC13S, STX11 and SH2D1A | HLH |
| Ma H, Zhang R, Zhang L, Wei A, Zhao X, Yang | Analyse the clinical characteristics, prognostic | Paediatric patients with primary HLH | 38 | 38 | Gene frequency of PRF1, UNC13D, STX11, | HLH |

| | | | | | | |
|---|---|---|---|---|---|---|
| Y, et al. Treatment of pediatric primary hemophagocytic lymphohistiocytosis with the HLH-94/2004 regimens and hematopoietic stem cell transplantation in China. Annals of hematology. 2020;99(10):2255-63. | factors, and effectiveness of HLH 94 and HLH 04 regimens and HSCT in paediatric patients with pHLH | | | | STXBP2, LYST, RAB27A, ITK, AP3B1, CD27, MAGT1 | |
| Macartney CA, Weitzman S, Wood SM, Bansal D, Steele M, Meeths M, et al. Unusual functional manifestations of a novel STX11 frameshift mutation in two infants with familial hemophagocytic lymphohistiocytosis type 4 (FHL4). Pediatric blood & cancer. 2011;56(4):654-7. | Describe two unrelated cases with same novel STX11 mutation | Paediatric cases of fHLH with novel STX11 mutation | 2 | 2 | Gene and variant frequency in PRF1, UNC13D and STX11 | HLH |
| Meeths M, Chiang SCC, Wood SM, Entesarian M, Schlums H, Bang B, et al. Familial hemophagocytic | Describe two UNC13D mutations that can explain the majority of FHL cases in Scandinavia and several cases across Europe | Swedish infants presenting with a known family history of HLH/defective cytotoxicity/degranulation + all infants identified with | 13 | 13 | Variant frequency in UNC13D | HLH |

| | | | | | | |
|---|---|---|---|---|---|---|
| lymphohistiocytosis type 3 (FHL3) caused by deep intronic mutation and inversion in UNC13D. Blood. 2011;118(22):5783-93. | | UNC13D mutation in their unit | | | | |
| Meeths M, Entesarian M, Al-Herz W, Chiang SCC, Wood SM, Al-Ateeqi W, et al. Spectrum of clinical presentations in familial hemophagocytic lymphohistiocytosis type 5 patients with mutations in STXBP2. Blood. 2010;116(15):2635-43. | Present patients with biallelic STXBP2 mutations | Families with biallelic STXBP2 mutations identified from a cohort of patients without identifiable mutations in PRF1, UNC13D or STX11 | 11 | 8 probands | Variant frequency in STXBP2 | single gene/variant |
| Mhatre S, Madkaikar M, Desai M, Ghosh K. Spectrum of perforin gene mutations in familial hemophagocytic lymphohistiocytosis (FHL) patients in India. Blood cells, molecules & diseases. 2015;54(3):250-7. | Elucidating the molecular pathology of a series of patients from India | Patients with HLH | 121 | 13 with samples available for mutation studies | Variant frequency in PRF1 | single gene/variant |

| | | | | | | |
|---|---|---|---|---|---|---|
| Mukda E, Trachoo O, Pasomsub E, Tiyasirichokchai R, Iemwimangsa N, Sosothikul D, et al. Exome sequencing for simultaneous mutation screening in children with hemophagocytic lymphohistiocytosis. International journal of hematology. 2017;106(2):282-90. | Analyse PRF1, UNC13D, STX11, and STXBP2 mutations using WES. (and also 8 other genes for inflammatory conditions) | Thai children with HLH diagnosis | 22 | 22 | Gene and variant frequency in PRF1, UNC13D, STX11, STXBP2, RAB27A, LYST, AP3B1, SH2D13, XIAP, ITK, CD27, and MAGT1 | HLH |
| Muralitharan S, Wali YA, Dennison D, Lamki ZA, Zachariah M, Nagwa EB, et al. Novel spectrum of perforin gene mutations in familial hemophagocytic lymphohistiocytosis in ethnic Omani patients. American journal of hematology. 2007;82(12):1099-102. | Describe the genetic screening for PRF1 mutation in unrelated ethnic Omani families | Patients with HLH | 16 | 16 | Gene and variant frequency in PRF1 | HLH |
| My LT, Lien LB, Hsieh W-C, Imamura T, Anh TNK, Anh PNL, et al. Comprehensive | Analyse the clinical assessment, cytokine profiles, virological aetiologies, family studies | Children with HLH (clinical or genetically confirmed) | 33 | 33 | Gene frequency of PRF1 and SH2D1A | HLH |

| | | | | | | |
|---|---|---|---|---|---|---|
| analyses and characterization of haemophagocytic lymphohistiocytosis in Vietnamese children. British journal of haematology. 2010;148(2):301-10. | and genetic analyses and treatment outcome of HLH in Vietnamese children | | | | | |
| Nepesov S, Yaman Y, Elli M, Bayram N, Ozdilli K, Kiykim A, et al. Clinical, Genetic, and Outcome Characteristics of Pediatric Patients with Primary Hemophagocytic Lymphohistiocytosis. Turkish archives of pediatrics. 2022;57(4):398-405. | Define the clinical, laboratory and genetic findings of people with familial HLH | Patients with familial HLH | 9 | 8 probands | Gene frequency of UNC13D, SH2D1A, PRF1, RAB27A and LYST | HLH |
| Okur H, Balta G, Akarsu N, Oner A, Patiroglu T, Bay A, et al. Clinical and molecular aspects of Turkish familial hemophagocytic lymphohistiocytosis patients with perforin mutations. Leukemia research. 2008;32(6):972-5. | Present the molecular defects and associated clinical phenotypes of FHL patients showing homozygosity for the perforin gene | Patients with primary HLH | 37 | 9 patients with perforin gene homozygosity (studied by linkage analysis) who were then direct sequenced | Variant frequency in PRF1 | single gene/variant |

| | | | | | | |
|---|---|---|---|---|---|---|
| Pagel J, Beutel K, Lehmberg K, Koch F, Maul-Pavicic A, Rohlfs A-K, et al. Distinct mutations in STXBP2 are associated with variable clinical presentations in patients with familial hemophagocytic lymphohistiocytosis type 5 (FHL5). Blood. 2012;119(25):6016-24. | Present detailed genetic and clinical analyses of a large cohort of patients with FHL with biallelic STXBP2 mutations from diverse ethnic origins and describe an extended clinical spectrum of the disease together with a clear genotype-phenotype correlation | Patients with suspected FHL | 185 | 28 probands | Variant frequency in STXBP2 | single gene/variant |
| Ramzan M, Yadav SP, Kharya G, Chinnabhandar V, Enteserian M, Henter JI, et al. Hemophagocytic lymphohistiocytosis in infants: a single center experience from India. Pediatric hematology and oncology. 2014;31(3):285-92. | Report the clinical profiles and outcomes of eight infants diagnoses with HLH | Infants with confirmed diagnosis of HLH as per HLH-2004 guidelines | 8 | 7 underwent mutation analysis | Gene and variant frequency in RAB27A, UNC13D, PRF1 and STX11 | HLH |
| Sanchez IP, Leal-Esteban LC, Alvarez-Alvarez JA, Perez-Romero CA, Orrego JC, Serna ML, et al. Analyses of the PRF1 | Present common PRF1 haplotype in unrelated families with atypical FHL2 | Patients with FHL2 | 4 | 4 | Variant frequency in PRF1 | single gene/variant |

| | | | | | | |
|---|---|---|---|---|---|---|
| gene in individuals with hemophagocytic lymphohystiocytosis reveal the common haplotype R54C/A91V in Colombian unrelated families associated with late onset disease. Journal of clinical immunology. 2012;32(4):670-80. | | | | | | |
| Sato H, Kawasaki N, Kawasaki M, Abiko Y, Meguro T, Takahashi N, et al. Three Consecutive Cases of Familial Hemophagocytic Lymphohistiocytosis, Including a Case Due to Maternal Uniparental Disomy. Journal of pediatric hematology/oncology. 2020;42(8):e819-e21. | Present consecutive cases of FHLH | Patients with fHLH | 3 | 3 | Variant frequency in PRF1 | HLH |
| Shabrish S, Kelkar M, Yadav RM, Bargir UA, Gupta M, Dalvi A, et al. The Spectrum of Clinical, Immunological, and Molecular Findings in | Report general clinical features, immunological and molecular findings and outcomes of FHL | Patients fulfilling HLH criteria | 101 | 98 <18y | Gene and variant frequency in PRF1, UNC13D, STXBP2 and STX11 | fHLH (4 genes only) |

| | | | | | | |
|---|---|---|---|---|---|---|
| Familial Hemophagocytic Lymphohistiocytosis: Experience From India. Frontiers in immunology. 2021;12:612583. | | | | | | |
| Sieni E, Cetica V, Santoro A, Beutel K, Mastrodicasa E, Meeths M, et al. Genotype-phenotype study of familial haemophagocytic lymphohistiocytosis type 3. Journal of medical genetics. 2011;48(5):343-52. | Genotype phenotype correlation analysis for fHLH3 | Cases with genetically diagnosed fHLH3 (presence of biallelic UNC13D mutations) extracted from consortium database | 84 | 69 probands | Variant frequency in UNC13D | single gene/variant |
| Suga N, Takada H, Nomura A, Ohga S, Ishii E, Ihara K, et al. Perforin defects of primary haemophagocytic lymphohistiocytosis in Japan. British journal of haematology. 2002;116(2):346-9. | Investigate perforin gene in Japanese HLH cases with or without family history | Patients with FHL or sporadic primary HLH | 16 | 15 probands | Variant frequency in PRF1 | HLH |
| Trizzino A, zur Stadt U, Ueda I, Risma K, Janka G, Ishii E, et al. Genotype-phenotype study of familial | Explore the contribution of various PRF1 mutations in FHL2 patients | Patients with fHLH2 pooled from common database | 124 | 124 | Variant frequency in PRF1 | single gene/variant |

| | | | | | | |
|---|---|---|---|---|---|---|
| haemophagocytic lymphohistiocytosis due to perforin mutations. Journal of medical genetics. 2008;45(1):15-21. | | | | | | |
| Ueda I, Ishii E, Morimoto A, Ohga S, Sako M, Imashuku S. Correlation between phenotypic heterogeneity and gene mutational characteristics in familial hemophagocytic lymphohistiocytosis (FHL). Pediatric blood & cancer. 2006;46(4):482-8. | Examine PRF1 and UNC13D abnormalities in a large group of patients with FHL (defined clinically and/or family history) | Patients with clinical FHL (at least one of: positive family history, deficient NK activity, CNS disease) | 40 | 40 | Gene and variant frequency in PRF1 and UNC13D | HLH |
| Woon S-T, Ameratunga R. Comprehensive genetic testing for primary immunodeficiency disorders in a tertiary hospital: 10-year experience in Auckland, New Zealand. Allergy, asthma, and clinical immunology : official | Review results of patients referred to comprehensive customised genetic testing service | Patients with PID referred for genetic testing | 228 | 20 with genetically confirmed fHLH | Gene frequency in PRF1, UNC13D, DTX11, STXBP2 and variant frequency in PRF1 and UNC13D. Additional genes/variant investigated: CD46, CFH, CFI, CD95, AIRE, NCF1, CYBB, CHD7, C2, NLRP3/CIAS1, DOCK8, NEMO, RAB27A, SERPING1, CD40L, AICDA, UNG, AUG, CD40, STAT3, FoxP3, ITK, | HLH |

| | | | | | | |
|---|---|---|---|---|---|---|
| journal of the Canadian Society of Allergy and Clinical Immunology. 2016;12:65. | | | | | SH2D1A BIRC4, SPINK5, MEFV, properdin, JAK3, RAG1, RAG2, ADA, LIG4, artemis, cernunnos factor, IL-7R, IL2-RG, SBDS, TNFRSF1A, UNC93b, WASP, CXCR4, BTK | |
| Xu XJ, Wang HS, Ju XL, Hu SY, Xiao Y, Xue HM, et al. Clinical presentation and outcome of pediatric patients with hemophagocytic lymphohistiocytosis in china: A retrospective multicenter study. Pediatric Blood and Cancer. 2016;63(Supplement 2):S55-S6. | Investigate the epidemiologic and general clinical features and outcome of HLH | Patients diagnosed with HLH according to the 2004 criteria, all children | 323 | 86 underwent genetic testing | Gene frequency of PRF1, UNC13D, STX11, STXBP2, SH2D1A, BIRC4, RAB27A, AP3B1 and LYST | HLH |
| Yoon HS, Kim HJ, Yoo KH, Sung KW, Koo HH, Kang HJ, et al. UNC13D is the predominant causative gene with recurrent splicing mutations in Korean patients with familial hemophagocytic lymphohistiocytosis. | Explore the genetic background of HLH in Korea | Patients with FHL | 40 | 40 | Gene and variant frequency in PRF1 and UNC13D | HLH |

| | | | | | | |
|---|---|---|---|---|---|---|
| Haematol-Hematol J. 2010;95(4):622-6. | | | | | | |
| Zhang L, Li Z, Liu W, Ma H, Wang T, Zhang R. Genetic characterization of pediatric primary hemophagocytic lymphohistiocytosis in China: a single-center study. Annals of hematology. 2019;98(10):2303-10. | Clarify genetics of Chinese paediatric primary HLH patients | Patients with HLH (clinical and genetic diagnosis/family history) | 26 | 26 | Gene and variant frequency in PRF1, UNC13D, STX11, STXBP2, LYST, AP3B1, RAB27A, SH2D1A, BIRC4, CD27, ITK, MAGT1 | HLH |
| Zur Stadt U, Beutel K, Kolberg S, Schneppenheim R, Kabisch H, Janka G, et al. Mutation spectrum in children with primary hemophagocytic lymphohistiocytosis: molecular and functional analyses of PRF1, UNC13D, STX11, and RAB27A. Human mutation. 2006;27(1):62-8. | Provide an overview of mutations involved in FHL | Patients with FHL | 63 | 63 | Gene and variant frequency in PRF1, UNC13D, STX11 and RAB27A | HLH |

### MCADD

| Reference | Study aim | Population | Total population N in study | Eligibile population for review | Study outcome | Disease category of included population |
|---|---|---|---|---|---|---|
| Martin-Rivada A, Palomino Perez L, Ruiz-Sala P, Navarrete R, Cambra Conejero A, Quijada Fraile P, et al. Diagnosis of inborn errors of metabolism within the expanded newborn screening in the Madrid region. JIMD reports. 2022;63(2):146-61. | To report experience of IEM diagnosis after implementation of expanded newborn screening programme | Newborns screened positive for IEMs through NBS programme | 902 | 222 | Gene frequency and variant frequency in ACADM, PAH, DNAJC12, PCBD1, GCDH, MCC1, MCC2, PCCB, MAT1A, FAH, HPD, BCKDHB, BCKDHA, DBT, BCAT2, CBs, SLC3A!, OTC, ASS1, HMGCL, ACADVL, HADHA, SLC22A5, CPT2, ETFB, CPT!A, MMACHC, MMADHC, MMUT, MMAB | MCADD |
| Nichols MJ, Saavedra-Matiz CA, Pass KA, Caggana M. Novel mutations causing medium chain acyl-CoA dehydrogenase deficiency: under-representation of the common c.985 A > G mutation in the New York state population. American journal of | To explore molecular basis for MCADD in New York state newborns | Newborns screened positive for MCADD | 511 | 511 | Variant frequency in ACADM | MCADD |

| | | | | | | |
|---|---|---|---|---|---|---|
| medical genetics Part A. 2008;146A(5):610-9. | | | | | | |
| Wang B, Zhang Q, Gao A, Wang Q, Ma J, Li H, et al. New Ratios for Performance Improvement for Identifying Acyl-CoA Dehydrogenase Deficiencies in Expanded Newborn Screening: A Retrospective Study. Frontiers in genetics. 2019;10:811. | To select optimal biochemical indicators (aka cutoffs) to maximise test accuracy in biochemical screening of ACAD disorders | Patients with suspected ACAD deficiencies | 673 | 81 | Gene frequency and variant frequency in ACADS, ACADM, and ACADVL | MCADD |
| Touw CML, Smit GPA, de Vries M, de Klerk JBC, Bosch AM, Visser G, et al. Risk stratification by residual enzyme activity after newborn screening for medium-chain acyl-CoA dehyrogenase deficiency: data from a cohort study. Orphanet journal of rare diseases. 2012;7:30. | To explore relationship between genotype and residual enzyme activity | Patients diagnosed with MCADD following positive NBS | 84 | 68 | Variant frequency in ACADM | MCADD |
| Maguolo A, Rodella G, Dianin A, Nurti R, Monge I, Rigotti E, et al. Diagnosis, genetic characterization and clinical follow up of mitochondrial fatty acid oxidation disorders in the | To evaluate the complexity of of genotype-phenotype correlation across range of FAO disorders | Patients diagnosed with FAODs | 34 | 20 | Gene frequency and variant frequency in ACADS, ACADM, ACADVL, ETFDH, SCL22A5, CPT2 | MCADD |

| | | | | | | |
|---|---|---|---|---|---|---|
| new era of expanded newborn screening: A single centre experience. Molecular genetics and metabolism reports. 2020;24:100632. | | | | | | |
| Mesbah Z, Sing Ho K, Fitzsimons P, Monavari AA, Crushell E, Mayne PD. Medium Chain Acyl-CoA Dehydrogenase Deficiency (MCADD) in the Irish Paediatric Population. Irish medical journal. 2020;112(10):1016. | To characterise the epidemiology and clinical features of MCADD in the Irish population | Children under 18 yrs diagnosed with MCADD in Ireland | 17 | 17 | Variant frequency in ACADM | MCADD |
| Carpenter K, Wiley V, Sim KG, Heath D, Wilcken B. Evaluation of newborn screening for medium chain acyl-CoA dehydrogenase deficiency in 275 000 babies. Archives of disease in childhood Fetal and neonatal edition. 2001;85(2):F105-9. | To report screening results for MCADD in over 250,000 neonates and retrospective review of samples from patietns now diagnosed with MCADD | Neonates as part of newborn screening, those with octanoylcarnitine >= 1umol/L were given genetic testing; clinically detected MCADD with a newborn screening sample available | 275,666 | 36 | Gene frequency in ACADM | MCADD |
| Li Y-Y, Xu J, Sun X-C, Li H-Y, Mu K. Newborn screening and genetic variation of medium chain acyl-CoA dehydrogenase deficiency in the Chinese population. Journal of pediatric | To explore prevalence of MCADD in the Chinese population and explore the characteristics of genetic variation | Neonates with positive screening for MCADD | 183,082 | 6 | Variant frequency in ACADM | MCADD |

| | | | | | | |
|---|---|---|---|---|---|---|
| endocrinology & metabolism : JPEM. 2022;35(10):1264-71. | | | | | | |
| Oerton J, Khalid JM, Besley G, Dalton RN, Downing M, Green A, et al. Newborn screening for medium chain acyl-CoA dehydrogenase deficiency in England: Prevalence, predictive value and test validity based on 1.5 million screened babies. J Med Screen. 2011;18(4):173-81. | To evaluate pilot screening service for MCADD | Newborns screened positive for MCADD | 190 | 147 | Variant frequency in ACADM | MCADD |
| Bentler K, Zhai S, Elsbecker SA, Arnold GL, Burton BK, Vockley J, et al. 221 newborn-screened neonates with medium-chain acyl-coenzyme A dehydrogenase deficiency: Findings from the Inborn Errors of Metabolism Collaborative. Molecular genetics and metabolism. 2016;119(1-2):75-82. | To investigate whether first newborn screen C8 values are related to gender, genotype, birth weight, or initial food source | Patients with MCADD, an abnormal newborn screening result and an available first newborn screen C8 value | 221 | 186 | Variant frequency in ACADM | MCADD |
| Tucci S, Wagner C, Grunert SC, Matysiak U, Weinhold N, Klein J, et al. Genotype and residual enzyme activity in medium-chain acyl-CoA dehydrogenase | To report results on confirmation testing after positive NBS for MCADD | Newborns with positive NBS results suggestive of MCADD | 170 | 170 | Variant frequency in ACADM | MCADD |

| | | | | | | |
|---|---|---|---|---|---|---|
| (MCAD) deficiency: Are predictions possible? Journal of inherited metabolic disease. 2021;44(4):916-25. | | | | | | |
| Jager EA, Kuijpers MM, Bosch AM, Mulder MF, Gozalbo ER, Visser G, et al. A nationwide retrospective observational study of population newborn screening for medium-chain acyl-CoA dehydrogenase (MCAD) deficiency in the Netherlands. Journal of inherited metabolic disease. 2019;42(5):890-7. | To evaluate the Dutch population NBS for MCADD through a nationwide retrospective observational study of clinical, laboratory and epidemiological parameters | Screen positive MCADD patients | 191 | 167 | Variant frequency in ACADM | MCADD |
| Gregersen N, Winter V, Curtis D, Deufel T, Mack M, Hendrickx J, et al. Medium-chain acyl-CoA dehydrogenase (MCAD) deficiency: the prevalent mutation G985 (K304E) is subject to a strong founder effect from northwestern Europe. Human heredity. 1993;43(6):342-50. | To study the evolution of the 985A>G mutation from European data | European patients with proven or suspected MCADD | 109 | 109 | Single variant (985A>G) frequency in ACADM | MCADD |

| | | | | | | |
|---|---|---|---|---|---|---|
| Ventura FV, Leandro P, Luz A, Rivera IA, Silva MFB, Ramos R, et al. Retrospective study of the medium-chain acyl-CoA dehydrogenase deficiency in Portugal. Clinical genetics. 2014;85(6):555-61. | To present first report of Portuguese MCADD population | Patients diagnosed with MCADD either by NBS or at clinical onset or through family studies | 109 | 109 | Variant frequency in ACADM | MCADD |
| Weiss KJ, Berger U, Haider M, Wagner M, Martner EMC, Regenauer-Vandewiele S, et al. Free carnitine concentrations and biochemical parameters in medium-chain acyl-CoA dehydrogenase deficiency: Genotype-phenotype correlation. Clinical genetics. 2023;103(6):644-54. | To report on biochemical phenotypes, secondary carnitine deficiency and carnitine supplementation | MCADD patients, most detected through NBS, a handful through targeted screening or clinically | 109 | 109 | Variant frequency in ACADM | MCADD |
| Anderson S, Botti C, Li B, Millonig JH, Lyon E, Millson A, et al. Medium chain acyl-CoA dehydrogenase deficiency detected among Hispanics by New Jersey newborn screening. American journal of medical genetics Part A. 2012;158A(9):2100-5. | To report incidence of clinically manifested MCADD and frequency of MCADD detected by NBS to estimate risk of clinical manifestation in undiagnosed children and evaluate effectiveness of screening. To report the mutational spectrum underlying MCAD | Patients either clinically detected or MS/MS NBS detected | 81 | 81 | Variant frequency in ACADM | MCADD |

| | | | | | | |
|---|---|---|---|---|---|---|
| | deficiency in Denmark and how it correlates with the clinical genotypes | | | | | |
| Nennstiel-Ratzel U, Arenz S, Maier EM, Knerr I, Baumkotter J, Roschinger W, et al. Reduced incidence of severe metabolic crisis or death in children with medium chain acyl-CoA dehydrogenase deficiency homozygous for c.985A>G identified by neonatal screening. Molecular genetics and metabolism. 2005;85(2):157-9. | To assess the outcome of MCADD in screened and unscreened populations by genotype | Children who screen positive for MCADD | 74 | 74 | Variant frequency in ACADM | MCADD |
| Anderson DR, Viau K, Botto LD, Pasquali M, Longo N. Clinical and biochemical outcomes of patients with medium-chain acyl-CoA dehydrogenase deficiency. Molecular genetics and metabolism. 2020;129(1):13-9. | To report on five Hispanic MCADD patients identified through New Jersey newborn screening and the cumulative incidence of MCADD among Hispanic and non-Hispanic infants identified through newborn screening during 30-month period | Hispanic infants with MCADD based on neonatal screening | 90 | 79 | Variant frequency in ACADM | MCADD |

| | | | | | | |
|---|---|---|---|---|---|---|
| Janeiro P, Jotta R, Ramos R, Florindo C, Ventura FV, Vilarinho L, et al. Follow-up of fatty acid beta-oxidation disorders in expanded newborn screening era. European journal of pediatrics. 2019;178(3):387-94. | To evaluate the effects of expanded NBS on the outcome of FAO disorder patients - analysis of 83 patient charts | Patients with FAODs positive on NBS screen | 83 | 66 | Variant frequency in ACADM | MCADD |
| Sturm M, Herebian D, Mueller M, Laryea MD, Spiekerkoetter U. Functional effects of different medium-chain acyl-CoA dehydrogenase genotypes and identification of asymptomatic variants. PloS one. 2012;7(9):e45110. | To analyse the genetic basis for the increase in C8 concentration in NBS-detected MCADD patients | Patients who screen positive for MCADD | 65 | 65 | Variant frequency in ACADM | MCADD |
| Andresen BS, Dobrowolski SF, O'Reilly L, Muenzer J, McCandless SE, Frazier DM, et al. Medium-chain acyl-CoA dehydrogenase (MCAD) mutations identified by MS/MS-based prospective screening of newborns differ from those observed in patients with clinical symptoms: identification and | To investigate the spectrum of mutations in the MCAD gene in newborns identified by observation of a diagnostic acylcarnitine profile by prospective MS/MS-based screening | Infants identified having MCAD | 62 | 62 | Variant frequency in ACADM | MCADD |

| | | | | | | |
|---|---|---|---|---|---|---|
| characterization of a new, prevalent mutation that results in mild MCAD deficiency. American journal of human genetics. 2001;68(6):1408-18. | | | | | | |
| Maier EM, Liebl B, Roschinger W, Nennstiel-Ratzel U, Fingerhut R, Olgemoller B, et al. Population spectrum of ACADM genotypes correlated to biochemical phenotypes in newborn screening for medium-chain acyl-CoA dehydrogenase deficiency. Human mutation. 2005;25(5):443-52. | To describe the spectrum of ACADM mutations from NBS | Patients who are biochemically confirmed of MCADD | 57 | 57 | Variant frequency in ACADM | MCADD |
| Yokota I, Coates PM, Hale DE, Rinaldo P, Tanaka K. Molecular survey of a prevalent mutation, 985A-to-G transition, and identification of five infrequent mutations in the medium-chain Acyl-CoA dehydrogenase (MCAD) gene in 55 patients with MCAD deficiency. Am J Hum | To evaluate prevalence of 985A>G mutation in MCADD patients | Patients with MCADD | 55 | 55 | Frequency of MCAD G985 gene mutation | MCADD |

| | | | | | | |
|---|---|---|---|---|---|---|
| Genet. 1991;49(6):1280-91. | | | | | | |
| Arnold GL, Saavedra-Matiz CA, Galvin-Parton PA, Erbe R, Devincentis E, Kronn D, et al. Lack of genotype-phenotype correlations and outcome in MCAD deficiency diagnosed by newborn screening in New York State. Molecular genetics and metabolism. 2010;99(3):263-8. | To identify patients with MCADD to identify biochemical or genotypic markers which might predict outcomes | Infants with a positive state screen for MCADD and a formal diagnosis by metabolite or DNA criteria and follow-up care | 53 | 53 | Frequency of MCAD G985 gene mutation | MCADD |
| Touw CML, Smit GPA, Niezen-Koning KE, Bosgraaf-de Boer C, Gerding A, Reijngoud D-J, et al. In vitro and in vivo consequences of variant medium-chain acyl-CoA dehydrogenase genotypes. Orphanet journal of rare diseases. 2013;8:43. | To evaluate fasting tolerance and enzyme assays in different MCADD genotypes | Newborns diagnosed with MCADD following NBS | 50 | 50 | Variant frequency in ACADM | MCADD |
| Hsu H-W, Zytkovicz TH, Comeau AM, Strauss AW, Marsden D, Shih VE, et al. Spectrum of medium-chain acyl-CoA dehydrogenase deficiency detected by newborn screening. Pediatrics. 2008;121(5):e1108-14. | To summarise results of first 6 years of screening programme | Cases of MCADD diagnosed using a combination of genetic and biochemical testing following screen positive | 47 | 47 | Frequency of MCAD G985 gene mutation | MCADD |

| | | | | | | |
|---|---|---|---|---|---|---|
| Couce ML, Sanchez-Pintos P, Diogo L, Leao-Teles E, Martins E, Santos H, et al. Newborn screening for medium-chain acyl-CoA dehydrogenase deficiency: regional experience and high incidence of carnitine deficiency. Orphanet journal of rare diseases. 2013;8:102. | To evaluate any relationships between biochemical findings at diagnosis, genotype, free carnitine levels during follow-up, and clinical outcome, in patients with MCADD detected by newborn screening | Patients with MCADD diagnosed by two newborn screening programmes | 45 | 45 | Variant frequency in ACADM | MCADD |
| Al-Jasmi FA, Al-Shamsi A, Hertecant JL, Al-Hamad SM, Souid A-K. Inborn Errors of Metabolism in the United Arab Emirates: Disorders Detected by Newborn Screening (2011-2014). JIMD reports. 2016;28:127-35. | To endorse preventive endeavours such as premarital counselling and genetic screening in order to mitigate these diseases in the community | Patients with IEM that were detected by newborn screening | 114 | 48 | Gene frequency and variant frequency in ACADM, PAH, QDPR, DBT, ASL, PCCB, MMAB, MMACHC, MCCC1, MCCC2, ACAT1, GCDH, IVD, ETFDH,ACADM, SLC22A5, BTD | MCADD |
| Wilcken B, Haas M, Joy P, Wiley V, Chaplin M, Black C, et al. Outcome of neonatal screening for medium-chain acyl-CoA dehydrogenase deficiency in Australia: a cohort study. Lancet (London, England). 2007;369(9555):37-42. | To evaluate the overall effectiveness of neonatal screening by MS/MS | Patients with MCADD (either diagnosed clinically or through NBS) | 41 | 41 | Single variant (985A>G) frequency in ACADM | MCADD |
| Tajima G, Hara K, Tsumura M, Kagawa R, Okada S, Sakura N, et al. Screening | To present characteristics of Japanese patients with MCADD | Japanese patients diagnosed with MCADD | 40 | 40 | Variant frequency in ACADM | MCADD |

| | | | | | | |
|---|---|---|---|---|---|---|
| of MCAD deficiency in Japan: 16years' experience of enzymatic and genetic evaluation. Mol Genet Metab. 2016;119(4):322-8. | | | | | | |
| Prasad C, Speechley KN, Dyack S, Rupar CA, Chakraborty P, Kronick JB. Incidence of medium-chain acyl-CoA dehydrogenase deficiency in Canada using the Canadian Paediatric Surveillance Program: Role of newborn screening. Paediatrics & child health. 2012;17(4):185-9. | To study incidence of MCADD, describe health status of children with MCADD, evaluate the impact of diagnosing method (clinically or NBS) on health outcomes, and identify mutation associated with MCADD cases and their relationship to phenotype | Children with a diagnosis of MCADD | 37 | 37 | Variant frequency in ACADM | MCADD |
| Gramer G, Haege G, Fang-Hoffmann J, Hoffmann GF, Bartram CR, Hinderhofer K, et al. Medium-Chain Acyl-CoA Dehydrogenase Deficiency: Evaluation of Genotype-Phenotype Correlation in Patients Detected by Newborn Screening. JIMD reports. 2015;23:101-12. | To evaluate the genotype-phenotype correlation in MCADD patients detected by NBS | Newborns detected with MCADD | 37 | 37 | Variant frequency in ACADM | MCADD |
| Waddell L, Wiley V, Carpenter K, Bennetts B, Angel L, Andresen BS, et | To explore genotypes of screen positive MCAD patients | Patients with biochemical evidence of MCADD | 36 | 36 | Variant frequency in ACADM | MCADD |

| | | | | | | |
|---|---|---|---|---|---|---|
| al. Medium-chain acyl-CoA dehydrogenase deficiency: genotype-biochemical phenotype correlations. Molecular genetics and metabolism. 2006;87(1):32-9. | | | | | | |
| Clayton PT, Doig M, Ghafari S, Meaney C, Taylor C, Leonard JV, et al. Screening for medium chain acyl-CoA dehydrogenase deficiency using electrospray ionisation tandem mass spectrometry. Archives of disease in childhood. 1998;79(2):109-15. | To describe experience of diagnosing MCADD using ESI-MS/MS | Children and neonates with a diagnosis of MCADD | 482 | 33 | Frequency of MCAD A985G gene mutation | MCADD |
| Clayton PT, Doig M, Ghafari S, Meaney C, Taylor C, Leonard JV, et al. Screening for medium chain acyl-CoA dehydrogenase deficiency using electrospray ionisation tandem mass spectrometry. Archives of disease in childhood. 1998;79(2):109-15. | To review both prevalence of MCADD in the Ontario population along with biochemical and molecular characteristics of confirmed cases + predictive value of screening test | Patients with MCADD, biochemically diagnosed after positive screening | 29 | 29 | Variant frequency in ACADM | MCADD |
| Janzen N, Hofmann AD, Schmidt G, Das AM, Illsinger S. Non-invasive test using palmitate in | To explore disease specific acylcarnitine patterns to help establish the FAOD diagnosis | Patients with FAOD confirmed after positive NBS | 47 | 30 | Variant frequency in ACADM | MCADD |

| | | | | | | |
|---|---|---|---|---|---|---|
| patients with suspected fatty acid oxidation defects: disease-specific acylcarnitine patterns can help to establish the diagnosis. Orphanet journal of rare diseases. 2017;12(1):187. | | | | | | |
| Thodi G, Georgiou V, Molou E, Loukas YL, Dotsikas Y, Biti S, et al. Characterization of the molecular spectrum of Medium-Chain Acyl-CoA Dehydrogenase Deficiency in a Greek newborns cohort: identification of a novel variant. Clinical biochemistry. 2012;45(15):1167-72. | To assess MCADD prevalence in Greece and determine the prevalent alleles in suspected infants | Infants screening positive for MCADD on MS/MS | 24 | 24 | Variant frequency in ACADM | MCADD |
| Gong Z, Liang L, Qiu W, Zhang H, Ye J, Wang Y, et al. Clinical, Biochemical, and Molecular Analyses of Medium-Chain Acyl-CoA Dehydrogenase Deficiency in Chinese Patients. Frontiers in genetics. 2021;12:577046. | To examine clinical biochemical and genotype characteristics of MCADD | Chinese MCADD patients | 24 | 23 | Variant frequency in ACADM | MCADD |
| Ding JH, Yang BZ, Bao Y, Roe CR, Chen YT. Identification of a new mutation in medium-chain | To molecular investigation of G985 in 22 unrelated MCADD families | Patients with MCADD | 22 | 22 | Single variant (985A>G) frequency in ACADM | MCADD |

| | | | | | | |
|---|---|---|---|---|---|---|
| acyl-CoA dehydrogenase (MCAD) deficiency. American journal of human genetics. 1992;50(1):229-33. | | | | | | |
| Al-Hassnan ZN, Imtiaz F, Al-Amoudi M, Rahbeeni Z, Al-Sayed M, Al-Owain M, et al. Medium-chain acyl-CoA dehydrogenase deficiency in Saudi Arabia: incidence, genotype, and preventive implications. Journal of inherited metabolic disease. 2010;33 Suppl 3:S263-7. | To present the biochemical phenotype, and molecular findings of MCADD in the Saudi population | Patients with MCADD that have been diagnosed by MS-MS using either dried blood spots (DBS) for newborns or blood samples from clinically suspected or at-risk infants | 30 | 20 | Variant frequency in ACADM | MCADD |
| ter Veld F, Mueller M, Kramer S, Haussmann U, Herebian D, Mayatepek E, et al. A novel tandem mass spectrometry method for rapid confirmation of medium- and very long-chain acyl-CoA dehydrogenase deficiency in newborns. PloS one. 2009;4(7):e6449. | To present new method for measurement of MCAD and VLCAD activity and correlate residual MCAD activities with genotype in MCADD | Patients who screen positive for MCADD | 18 | 18 | Variant frequency in ACADM | MCADD |
| Purevsuren J, Hasegawa Y, Fukuda S, Kobayashi H, Mushimoto Y, Yamada K, et al. Clinical and molecular aspects of | To present clinical onset, genotypes, and outcomes of Japanese children with MCADD | Patients diagnosed with MCADD through NBS, after clinical metabolic crisis or through sibling detection | 16 | 15 | Variant frequency in ACADM | MCADD |

| | | | | | | |
|---|---|---|---|---|---|---|
| Japanese children with medium chain acyl-CoA dehydrogenase deficiency. Molecular genetics and metabolism. 2012;107(1-2):237-40. | | | | | | |
| Catarzi S, Caciotti A, Thusberg J, Tonin R, Malvagia S, la Marca G, et al. Medium-chain acyl-CoA deficiency: outlines from newborn screening, in silico predictions, and molecular studies. TheScientificWorldJournal. 2013;2013:625824. | To report of biochemical and genetic studies on MCADD neonates identified through newborn screening | Patients with a clinical suspicion of MCADD following newborn screening | 14 | 14 | Variant frequency in ACADM | MCADD |
| Couce ML, Castineiras DE, Moure JD, Cocho JA, Sanchez-Pintos P, Garcia-Villoria J, et al. Relevance of expanded neonatal screening of medium-chain acyl co-a dehydrogenase deficiency: outcome of a decade in galicia (Spain). JIMD reports. 2011;1:131-6. | To evaluate the diagnostic results as well as the outcome of the MCADD deficient individuals after 10 years of screening | Cases of MCADD diagnosed with a newborn screening programme | 11 | 11 | Variant frequency in ACADM | MCADD |
| Howard C, Gorman I, Crushell E, Knerr I, Hughes J, Boruah R, et al. Medium Chain Acyl-CoA Dehydrogenase Deficiency: 3 years of | To review data since the launch of the MCADD NBS in Ireland to characterise the confirmed cases biochemically, clinically and genetically and | Patients who screen positive for MCADD | 11 | 11 | Variant frequency in ACADM | MCADD |

| | | | | | | |
|---|---|---|---|---|---|---|
| Newborn Screening. Irish medical journal. 2023;116(3):743. | reassess the incidence of MCADD in Ireland | | | | | |
| Purevsuren J, Kobayashi H, Hasegawa Y, Mushimoto Y, Li H, Fukuda S, et al. A novel molecular aspect of Japanese patients with medium-chain acyl-CoA dehydrogenase deficiency (MCADD): c.449-452delCTGA is a common mutation in Japanese patients with MCADD. Molecular genetics and metabolism. 2009;96(2):77-9. | To report genetic aspects of 11 Japanese patients with MCADD | Japanese patients diagnosed with MCADD | 11 | 10 | Variant frequency in ACADM | MCADD |
| Horvath GA, Davidson AGF, Stockler-Ipsiroglu SG, Lillquist YP, Waters PJ, Olpin S, et al. Newborn screening for MCAD deficiency - Experience of the first three years in British Columbia, Canada. Can J Public Health-Rev Can Sante Publ. 2008;99(4):276-80. | To evaluate incidence, clinical outcome. Biochemical and molecular phenotype of MCADD cases detected in the first 3 years of screening programme in BC Canada | Patients diagnosed with MCADD following positive NBS | 10 | 10 | Frequency of MCAD A985G gene mutation | MCADD |
| Balci MC, Karaca M, Ergul Y, Omeroglu RE, Demirkol M, Gokcay GF. Cardiologic evaluation of Turkish | To investigate type and frequency of cardiac pathologies and response | Children followed up with a diagnosis of FAOD divided into 7 subgroups based on types of FAOD | 68 | 63 | Variant frequency in ACADM | MCADD |

| | | | | | | |
|---|---|---|---|---|---|---|
| mitochondrial fatty acid oxidation disorders. Pediatrics international : official journal of the Japan Pediatric Society. 2022;64(1):e15317. | to recommended treatment in FAODs | | | | | |
| Li Y, Zhu R, Liu Y, Song J, Xu J, Yang Y. Medium-chain acyl-coenzyme A dehydrogenase deficiency: Six cases in the Chinese population. Pediatrics international : official journal of the Japan Pediatric Society. 2019;61(6):551-7. | To report the clinical, biochemical and mutation spectrum of Chinese MCADD patients | Patients with MCADD: identified through NBS or clinically | 6 | 6 | Variant frequency in ACADM | MCADD |
| Tian Y, Zhu X, Lv S, Jia C, Zhang L, Ni M, et al. Analysis of gene mutations of medium-chain acyl-coenzyme a dehydrogenase deficiency (MCADD) by next-generation sequencing in Henan, China. Clinica chimica acta; international journal of clinical chemistry. 2022;536:155-61. | To find out the novel pathogenic variants in the patients and clarify the detection rate of the disease of high frequency ACADM pathogenic variants in this population | Infants with MS/MS screen positive MCADD in Henan, China | 6 | 6 | Variant frequency in ACADM | MCADD |

| | | | | | | |
|---|---|---|---|---|---|---|
| Anderson S, Botti C, Li B, Millonig JH, Lyon E, Millson A, et al. Medium chain acyl-CoA dehydrogenase deficiency detected among Hispanics by New Jersey newborn screening. American journal of medical genetics Part A. 2012;158A(9):2100-5. | To report on five Hispanic MCADD patients identified through New Jersey newborn screening and the cumulative incidence of MCADD among Hispanic and non-Hispanic infants identified through newborn screening | Patients with elevated octanoylcarnitine identified through NBS | 20 | 5 | Variant frequency in ACADM | MCADD |
| Kim MJ, Kim SY, Lee JS, Kang S, Park L-J, Choi W, et al. Rapid Targeted Sequencing Using Dried Blood Spot Samples for Patients With Suspected Actionable Genetic Diseases. Annals of laboratory medicine. 2023;43(3):280-9. | To evaluate a rapid genomic testing platform for 254 genetic diseases | Patients with abnormalities in neonatal screening, or unexplained neonatal hypotonia/neonates-onset seizure, unexplained abnormal lab findings, skeletal dysplasia or joint problems, neurodevelopmental delay with abnormal metabolic screening test | 111 | 15 | Gene and variant frequency in ACADM, ACADS, PAH, PREPL, PHKA2, MCCC2, PCCA, SLC25A13 | MCADD |
| Casey JL. MCAD deficiency in the Holderman Mennonite population in central Kansas. Kansas medicine : the journal of the Kansas Medical Society. 1992;93(11):306-8. | To diagnose MCADD in Holderman Mennonite children | Children ≤5yrs | 147 | 12 | Frequency of MCAD A985G gene mutation | MCADD |
| Gregersen N, Winter V, Lyonnet S, Saudubray JM, Wendel U, Jensen TG, et al. MOLECULAR-GENETIC | To characterize, clinically, biochemically, and genetically, two families with a 13 base pair repeat | Two families with medium-chain acyl-CoA dehydrogenase deficiency due to compound heterozygosity | 9 | 2 | Frequency of MCAD A985G gene mutation | MCADD |

| | | | | | | |
|---|---|---|---|---|---|---|
| CHARACTERIZATION AND URINARY-EXCRETION PATTERN OF METABOLITES IN 2 FAMILIES WITH MCAD DEFICIENCY DUE TO COMPOUND HETEROZYGOSITY WITH A 13 BASE-PAIR INSERTION IN ONE ALLELE. Journal of Inherited Metabolic Disease. 1994;17(2):169-84. | insertion in one allele, resulting in reduced level of MCAD mRNA | | | | | |
| Kirk JM, Laing IA, Smith N, Uttley WS. Neonatal presentation of medium-chain acyl-CoA dehydrogenase deficiency in two families. Journal of inherited metabolic disease. 1996;19(3):370-1. | To present two non-consanguineous Scottish families in whom definitive diagnosis of medium-chain acyl-CoA dehydrogenase (MCAD) deficiency (MCADD; McKusick 201450), with homozygosity for the G985A mutation | Two Scottish families diagnosed with MCADD in one surviving child | 2 | 2 | Frequency of MCAD G985A gene mutation | MCADD |
| Woo HI, Park H-D, Lee Y-W, Lee DH, Ki C-S, Lee S-Y, et al. Clinical, biochemical and genetic analyses in two Korean patients with medium-chain acyl-CoA dehydrogenase deficiency. The Korean journal of laboratory medicine. 2011;31(1):54-60. | To describe mutations observed in Asian patients with MCADD | Korean patients screened positive for MCADD through NBS MS/MS | 2 | 2 | Variant frequency in ACADM | MCADD |
